# Live biotherapeutic product exhibits similar efficacy and superior engraftment to same donor fecal microbiota transplant for recurrent Clostridioides difficile infection

**DOI:** 10.1101/2025.09.08.25335342

**Authors:** Lukas Bethlehem, Lorenza Bartu, Gina Marke, Phyu Mar, Sari Feldman, Joseph Eggers, Constantin Ruprecht, Graham Britton, Varun Aggarwala, Gerold Bongers, Zhihua Li, Nancy Yang, Elizabeth Hohmann, Ilaria Mogno, Jeremiah J Faith, Ari Grinspan

## Abstract

Fecal microbiota transplantation (FMT) is an effective therapy for recurrent *Clostridioides difficile* infection (rCDI) but has undefined composition and poor scalability. *In vitro* manufactured live biotherapeutic products (LBP) enable both scalability and defined strain composition but with higher manufacturing complexity, resulting in few LBP trials. We developed an accessible platform to produce human-grade LBPs. We provide regulatory documentation and manufacturing protocols to facilitate translating microbiome advances to human trials. With this platform, we conduct the first direct comparison of the same bacterial strains administered after *in vitro* manufacturing (LBP) compared to donor sourced (FMT) across two doses. In a phase 1b trial (n=18), an endoscopic dose of the 15-strain consortium MTC01 was safe with rCDI prevention eight weeks after dosing in seven out of nine LBP patients, similar to eight out of nine FMT patients. Notably, MTC01 strain engraftment was superior to FMT at higher doses.

## Introduction

The approvals of two stool-derived products, fecal microbiota, live-jslm (Rebyota)^1^ and fecal microbiota spores, live-brpk (Vowst)^2^, for the treatment of recurrent *C. difficile* infection (rCDI) provide a key milestone in the development of live microbial therapeutics. However, the undefined composition of these products also highlights our limited understanding of what is necessary and sufficient for success. Rebyota demonstrates that a complete fecal slurry is an effective rCDI therapy, as supported by numerous prior fecal microbiota transplantation trials^3–6^, while Vowst demonstrates that the bacterial spore fraction remaining after ethanol treatment of stool is sufficient as an effective rCDI treatment. However, fecal-based drugs have little to no overlap in strain composition across the varied donors used in their production^7–9^, and therefore the efficacy and safety of such products represent an average across many microbial compositions resulting in uncertainty for individual product batches.

Defined in vitro manufactured bacterial strain communities, known as live biotherapeutic products (LBPs), provide the potential to identify sets of bacterial strains that are sufficient to treat a given disease with a fixed *in vitro* manufactured composition. LBPs have the clear safety and efficacy profile of a single defined product^10–12^ and enable biomanufacturing scaling rather than a linear scaling by the number of donor samples for stool-derived products. Five LBPs have been tested for rCDI to date including one positive phase 2 trial of an 8 strain consortium^13^, one negative phase 2 trial of a 12 strain consortium^14^, and three promising smaller trials with larger defined communities^15–17^. Despite the inherent advantages of LBPs, FMT trials still far outnumber LBP trials as FMTs require only simple equipment to manufacture and have published manufacturing^18,19^ and regulatory^20^ know-how. As a result, we lack advancement on several fundamental principles for designing LBPs such as 1) how to select the best engrafting strains, 2) do in vitro manufactured strains engraft as well as donor-sourced materials, and 3) how to run safe, low-costLBPs and mechanistic understanding.

We developed a novel cost- and space-efficient platform for manufacturing human-grade LPBs for clinical trials. To maximize community access and the therapeutic potential of the microbiome for human health, we share the regulatory documentation and protocols, as a blueprint for other manufacturing efforts. The manufacturing platform introduces the concept of sterilizable anaerobic chambers with disposable culture flasks as an alternative to space-intensive clean rooms and costly bioreactors. Using our LBP platform, we manufactured two doses of a 15 strain obligate anaerobe defined consortium to address fundamental questions about LBP strain selection, engraftment, and dosing in a phase 1b clinical trial. The strains were isolated from a frequently successful rCDI FMT donor^8,21^, based on engraftment frequency in recurrency free rCDI FMT patients. In parallel, we produced FMT, at the same doses as the LBP, from the same donor used to isolate the LBP strains. These shared donor-derived FMT and donor-isolated LBP drug products allowed us to compare the safety, efficacy, and engraftment of the same strains manufactured *in vitro* with the full spectrum donor sourced FMT fecal slurry at two matched doses. This first of its kind head-to-head comparison in eighteen subjects with rCDI treated with either FMT or LBP demonstrates that *in vitro* manufactured bacteria have similar efficacy and higher, dose-dependent engraftment to the same microbes derived from their native human colon environment for FMT administration.

## Results

### Manufacturing of LBP MTC01 and an FMT product from the same donor

Drug candidate “Microbiome Translational Center drug 01” (MTC01) is composed of 15 obligate anaerobe bacterial strains isolated from a single FMT donor (D283) whose stool was used in 28 fecal transplants to treat rCDI from 2014 to 2017 with 89% prevention of relapse at 8 weeks across all transplants^21^. From five donor stool samples, we isolated and sequenced bacterial strains using a high throughput culturing platform with selective media to enrich different bacterial groups yielding 67 strains from 1387 isolates across 49 species as potential candidates for a defined alternative to D283 FMT^7,8^ (Figure 1A).

**Figure 1.**
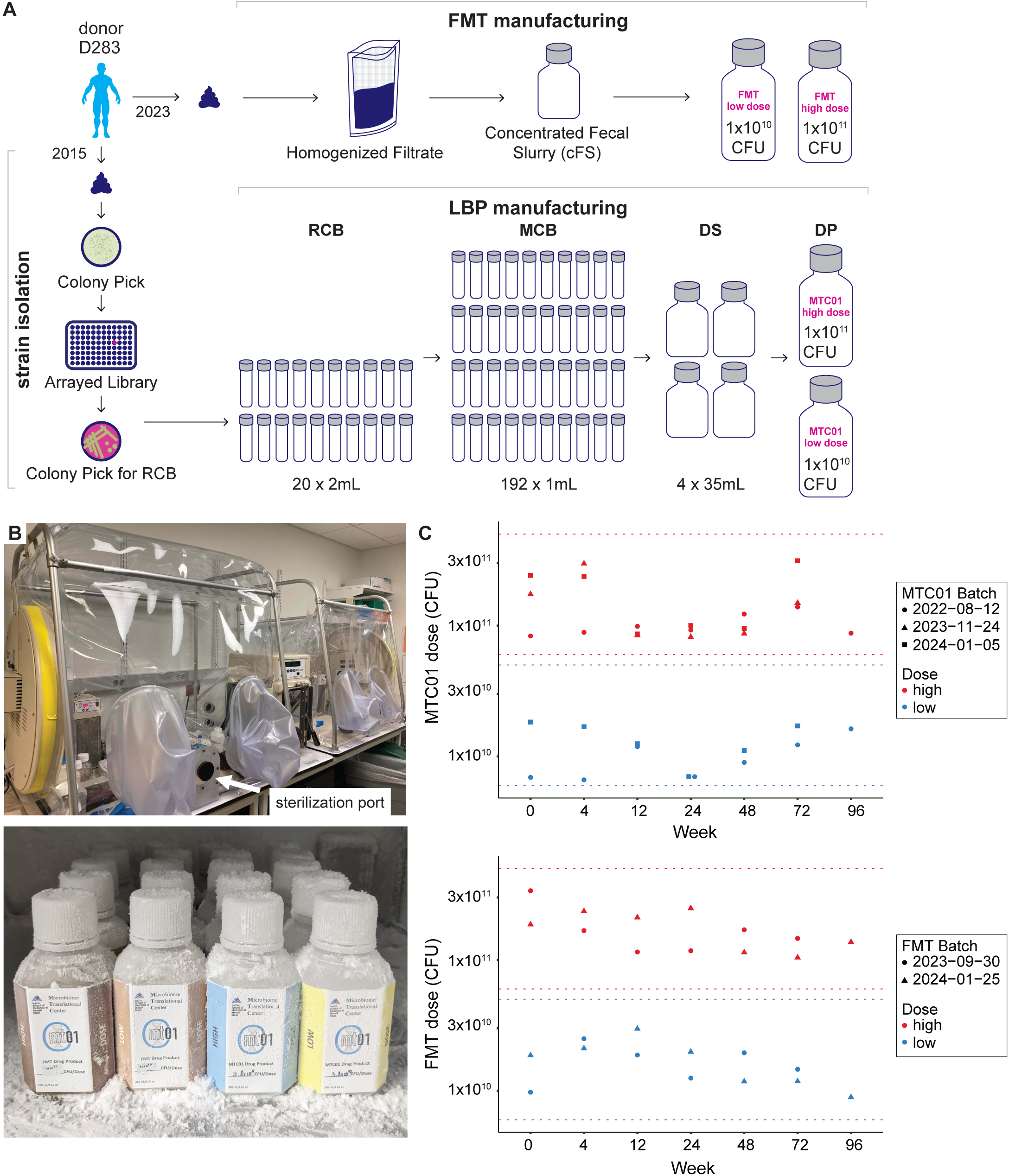
Manufacturing of FMT and LBP from the same donor’s microbiome. (A) Bacterial strains were isolated from FMT donor D283 stool samples collected in 2015 by plating on solid media and picking single colonies for cultivation and storage in 96-well plates with each well containing a single isolate confirmed by MALDI-TOF and genome sequencing. To manufacture MTC01 a Research Cell Bank (RCB, 20 vials per strain) was generated from a single colony isolated from plating each MTC01 strain. A Master Cell Bank (MCB, up to 192 vials per strain) was generated from each RCB strain, and Drug Substance (DS, 4 x35 mL per strain) was generated from each MCB. The Drug Product (DP) was generated from the DS by combining all strains at even CFU per dose, where possible, for a total dose of approximately 1×10^11^ CFU (high dose) or 1×10^10^ CFU (low dose) in a volume of 250 mL. In 2023, additional stools were collected from donor D283, homogenized, coarse filtered, and used to generate FMT DPs at the same high and low dose potencies as MTC01. (B) Manufacturing was performed in a dedicated flexible film anaerobic chamber equipped with a sterilization port to easily fog the chamber with sterilant. Final DPs were stored in a volume of 250 mL at −80°C until the time of colonoscopic delivery. (C) MTC01 and FMT DPs were stable within the specification bounds (indicated with horizontal dotted lines) of high dose and low dose for the duration of at least 72 weeks.

Given prior observations that higher engraftment of FMT and LBP donor strains is predictive of relapse prevention in rCDI trials of microbial therapeutics^8,13,22–25^, we hypothesized that an LBP comprised of strains with high FMT engraftment frequencies would facilitate high LBP engraftment and protection from relapse, while avoiding the manufacture and administration of non-engrafting strains that would not prevent relapse. Using the empirical engraftment data from recipients of D283 strains administered by FMT^8^, we identified a drug candidate MTC01 – comprised of 15 phylogenetically diverse strains from commonly isolated gut species that frequently engraft in non-relapsing rCDI recipients (Table S1). During the design of the consortium, we performed several filtering steps to improve the safety profile and bring the defined consortium to an easier size for manufacturing. For species with multiple strains isolated from D283 stool, we selected only a single strain to maximize species diversity. We excluded *Escherichia coli* based on its previous association with FMT adverse events^11,26,27^, and we eliminated a strain of *Bacteroides fragilis* as a result of its antibiotic resistance profile. Last, we eliminated rare species, as we hypothesized that commonly occurring strains of bacteria are sufficient to treat rCDI given that many different donors have been successful in rCDI treatment with FMT.

Prior to GMP manufacturing, we plated each MTC01 strain from the culture libraries onto solid agar to obtain individual bacterial colonies (Figure 1A). We inoculated a liquid culture with a single colony of each strain to generate research cell banks (RCB) for strain characterization. RCB vials were used to identify that all 15 strains were strict anaerobes and did not interfere with microbiological test methods for non-sterile products, defined by the United States Pharmacopeia (USP), USP<61> (enumeration of total aerobic microbial, yeast and mold) or USP<62> (detection of specific pathogens) assays. Each strain has at least seven antibiotic susceptibilities, and all strains share a susceptibility to three of the 12 tested antibiotics, allowing us to control MTC01 in case of adverse events in the clinical trial (Table S1). We used an established manufacturing and banking system for producing MTC01, generating Master Cell Banks (MCB), Drug Substance (DS), and Drug Product (DP), to ensure traceability, genetic stability, and quality control following regulatory guidance^28^ (Figure 1A). To shorten the time to the clinic, we did not generate Working Cell Banks for this production of MTC01. MCB, DS, and DP manufacturing was performed according to GMP standards in a dedicated vinyl anaerobic chamber with an integrated port to sterilize the chamber environment and the equipment with ionized hydrogen peroxide before each production run (Figure 1B). Strains were grown individually in either side of the two-sided anaerobic chamber, to prevent internal cross-contamination, utilizing single-use, sterile consumables. The production of MCB, DS and DP were documented in batch records (see Supplemental Materials) and concluded within 9 months (Table S2). All GMP cell banks were stored in a dedicated −80°C freezer (Figure 1B). Since rCDI FMT trials have used widely varied doses, typically defined in grams of stool rather than a direct measure of microbial potency, a high dose and a low dose drug product were designed for both the LBP and FMT with colony forming units (CFU) as the potency measure.

Each MCB and DS strain was batch fermented in an animal product free complex medium for 12-54 hours at 37°C. After fermentation, cells were washed with USP-grade phosphate buffered saline before resuspension in USP-grade formulation buffer. To generate MTC01 DP, DS aliquots from each of the 15 strains were thawed and combined in a sterilized anaerobic chamber to reach a target cumulative high dose of 1×10^11^ colony forming units (CFU) or a low dose of 1×10^10^ CFU. For twelve of the fifteen MTC01 strains, an equal potency of viable cells (CFU) was added to the DP to reach the desired cumulative target dose in a total volume of 250 mL. For *Bacteroides caccae*, *Eubacterium rectale* and *Dorea longicatena* with insufficient viable cells after DS production, the highest strain dose possible was used (Table S3). Identity criteria of MCB and DS were met with over 48 high quality MALDI-TOF spectra (Bruker Biotyper, confidence score >1.8) from independent bacterial colonies matching the correct Bruker library entry for the MTC01 strain produced. DP identity release was met with over 200 high quality spectra matching to the 15 MTC01 strains. Potency was within range for high dose (5×10^11^ - 6×10^10^ CFU) or low dose (5×10^10^ - 6×10^9^ CFU). All MCB, DS, and DP also passed USP<61> and USP<62> testing (Table 1). The Right First Time manufacturing percentage was 53%, 80%, and 100% for the MCB, DS, and DP respectively.

**Table 1.** Drug product release results for (A) MTC01 and (B) FMT.

To generate FMT drug product, two fecal samples from D283 were individually homogenized and filtered (Seward Stomacher EVO 400) to generate two batches of concentrated fecal slurry that were frozen until FMT DP dilution. An aliquot of each concentrated fecal slurry was treated likewise, frozen and thawed, whereupon its potency was assessed by CFU. This potency was then used to dilute the concentrated fecal slurries into two FMT DP batches that met the same high dose and low dose CFU release criteria as MTC01 (Table 1). Both the FMT and MTC01 DPs used the same formulation buffer in a volume of 250 mL, and were stored at −80°C until the day of colonoscopic delivery. We found robust stability of cumulative DP potency at −80°C (Figure 1 and Figure S1A). We also found MTC01 cumulative potency to be stable under the clinical application conditions that included up to 4 hours thaw time and up to 1 hour time in ambient aerobic conditions outside of the storage bottle prior to colonic infusion (Figure S1B). We did, however, find that strains *D. longicatena* and *E. rectale* might decrease in potency after longer storage periods (Figure S1A).

Regulatory considerations were discussed with the FDA via a pre-investigational new drug (IND) application with an FDA written response. An IND application was subsequently submitted with all manufacturing details and a clinical study protocol with no hold. Pre-IND communications, the IND (29067), and the master batch records (MBRs) are provided to facilitate other manufacturing efforts (Supplemental Materials). Institutional research board (IRB,STUDY-23-00563) approval was obtained for a single site trial at Mount Sinai Hospital registered at clinicaltrials.gov (NCT05911997).

### Phase-1b clinical trial of MTC01 and FMT for rCDI

MTC01 and FMT drug products from the same human donor were tested in a phase 1b clinical trial at Mount Sinai Hospital enrolling individuals ≥18 years old with 2 or more qualifying CDI episodes occurring within the prior 6 months, inclusive of the current episode. A qualifying episode was defined as CDI symptoms starting within 60 days of randomization with ≥3 unformed stools/day for ≥2 days that is clinically consistent with CDI, documented positive stool test for toxigenic *C. difficile* (toxin EIA or PCR) for the current episode within 60 days of randomization, and received standard-of-care CDI antibiotics for the current episode for 10-42 days with adequate clinical response to the antibiotics defined as ≤3 unformed stools/day for ≥2 days. Subjects with LBP or FMT in the prior 6 months were excluded.

Patients enrolled into the study were block randomized (block size = 4) into either low-dose FMT, high-dose FMT, low-dose MTC01, or high-dose MTC01 (Figure 2A). Prior to colonoscopic drug administration, subjects were taken off all antibiotics for 48hr and underwent a standard colonoscopy bowel preparation within 12 hours of their procedure (Figure 2B). The microbial therapeutic was administered to the right colon by colonoscopy. The primary endpoint of safety was determined by the incidence of serious adverse events (SAE), adverse events (AE), vital signs, and physical examination up to 24 weeks. Secondary endpoints were 1) recurrence of CDI up to 8 weeks after treatment with recurrence defined as positive *C. difficile* toxin EIA as well as diarrhea (≥3 loose stools/day for 2 days), 2) the engraftment of MTC01 and FMT in the fecal microbiome 2 weeks after treatment, and MTC01 and FMT related taxonomic changes in the fecal microbiome 2 weeks after treatment. Other outcome measures were time to recurrence of CDI up to 24 weeks and microbiome changes up to 26 weeks.

**Figure 2.**
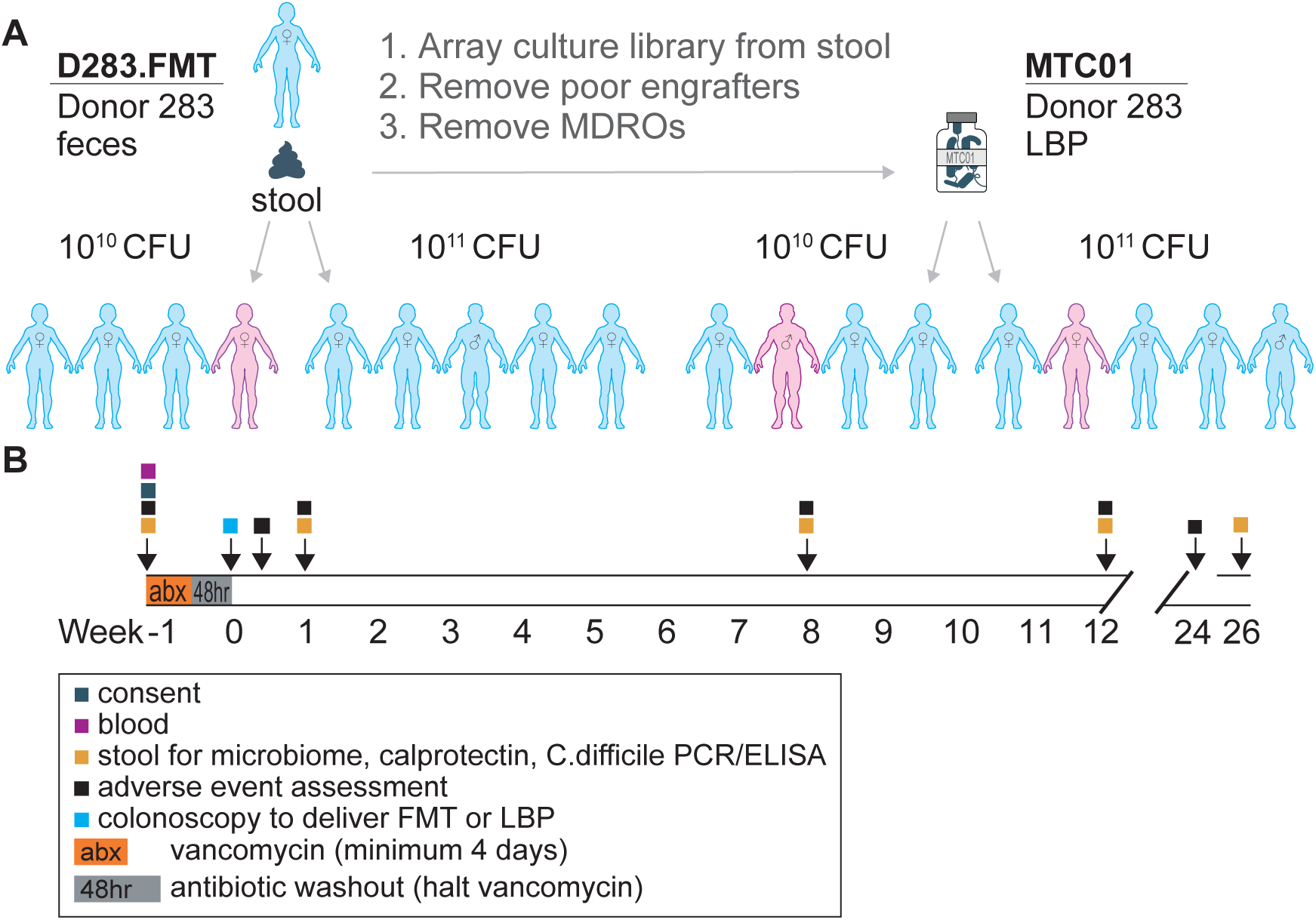
Clinical trial design and outcomes. (A) Patients were randomized to receive either low dose FMT, high dose FMT, low dose MTC01, or high dose MTC01. Patient sex is indicated by the symbol on each individual. Patients that relapsed are coloured magenta. (B) Adverse events were assessed for up to 24 weeks. Stool samples were collected for up to 26 weeks for metagenomic sequencing to track FMT donor strains and MTC01 strains in recipients over time.

Twenty subjects were assessed for eligibility with 18 patients dosed (5 high dose FMT, 4 low dose FMT, 5 high dose MTC01, 4 low dose MTC01) and 2 screen failures (Table 2). The average patient age was 58 with 15 females and 3 males (Figure 2A). For the primary endpoint of safety, one patient had a severe AE due to underlying disease with the patient not responding to treatment, and two patients had moderate AEs. In addition, there were 7 mild AE with AE observed in 6 total patients (Table 2). The 10 adverse events across 8 patients were evenly spread across MTC01 (5 events) and FMT (5 events) recipients and were deemed not related to the study interventions.

**Table 2.** Clinical (A) recruitment, (B) safety, and (C) outcomes.

For the secondary endpoint of recurrence of CDI up to 8 weeks after treatment, three of the eighteen subjects had a recurrence with success rates of 100%, 75%, 80%, and 75% across high dose FMT (5 of 5), low dose FMT (3 of 4), high dose MTC01 (4 of 5), and low dose MTC01 (3 of 4) respectively (Figure 2A; Table 2). Among the recurrences, one occurred in a high dose patient and two occurred in low dose patients for success rates of 90% and 75% in the high dose and low dose recipients respectively. The mean age of the three patients that recurred was 66 compared to a mean age of 56 in non-recurring patients. Both recurrences to MTC01 occurred in the first week, while the FMT recurrence occurred at week 8. Although this phase 1b trial was not powered or placebo controlled to assess efficacy or non-inferiority, all four groups had success rates above historic placebo results from the two phase 3 trials of stool based live microbial therapeutics (SER-109 = 60%, RBX2660 = 57.5%)^1,2^.

### Engraftment of MTC01 and FMT strains in rCDI recipients

To assess the engraftment of strains from D283 FMT product and the *in vitro* manufactured 15 strain consortium isolated from D283 in rCDI recipients, we collected recipient stool samples prior to drug administration and at 1, 8, 12, and 26 weeks after the single endoscopic dose (Figure 2B). Metagenomics sequencing was performed on both patient and donor samples. We assessed the presence or absence of all 15 MTC01 strains in each recipient and donor sample with the Strainer algorithm that identifies strain-specific kmers in each strain and assesses their abundance in patient and donor stool metagenomes relative to a negative control database of unrelated metagenomes^8,29^. For FMT recipients and the donor, we also used Strainer to assess colonization of an additional 115 strains that were not in MTC01 but that were previously isolated and sequenced from donor D283, including strains cultured from donor D283 after the original isolation of MTC01 strains and from the new samples collected to produce the FMT drug product used in this trial. We used linear mixed effects models to assess differences in engraftment between groups using all samples and incorporating random effects to account for the multiple measures for each participant.

As expected, we detect MTC01 strains far more often in recipients after dosing than prior to dosing (p<1.0×10^−5^ for all groups) – with the few detections prior to dosing likely representing false positive detections of genetically similar strains (Figure 3A). High dose MTC01 had the highest overall number of the 15 strains colonized even when compared with high dose FMT (p=0.03; Figure 3A). These differences in colonization between high dose FMT and high dose MTC01 were largely driven by lower colonization of *Bifidobacterium longum*, *Bifidobacterium adolescentis*, *Eubacterium rectale*, and *Bifididobacterium pseudocatenulatum* suggesting that, between their initial isolation in 2015 and the resampling of the donor to produce the FMT product, these strains were lost from the donor microbiome and therefore unable to be transmitted to recipients or alternatively that they are too low in abundance in the stool of the donor for effective transmission compared to the higher doses attainable in MTC01 where each strain dose can be individually controlled. Interestingly, *Bifidobacterium longum*, *Bifidobacterium adolescentis*, and *Eubacterium rectale* had one or more additional non-MTC01 strains in the donor microbiota that preferentially transmitted to FMT recipients^7^.

**Figure 3.**
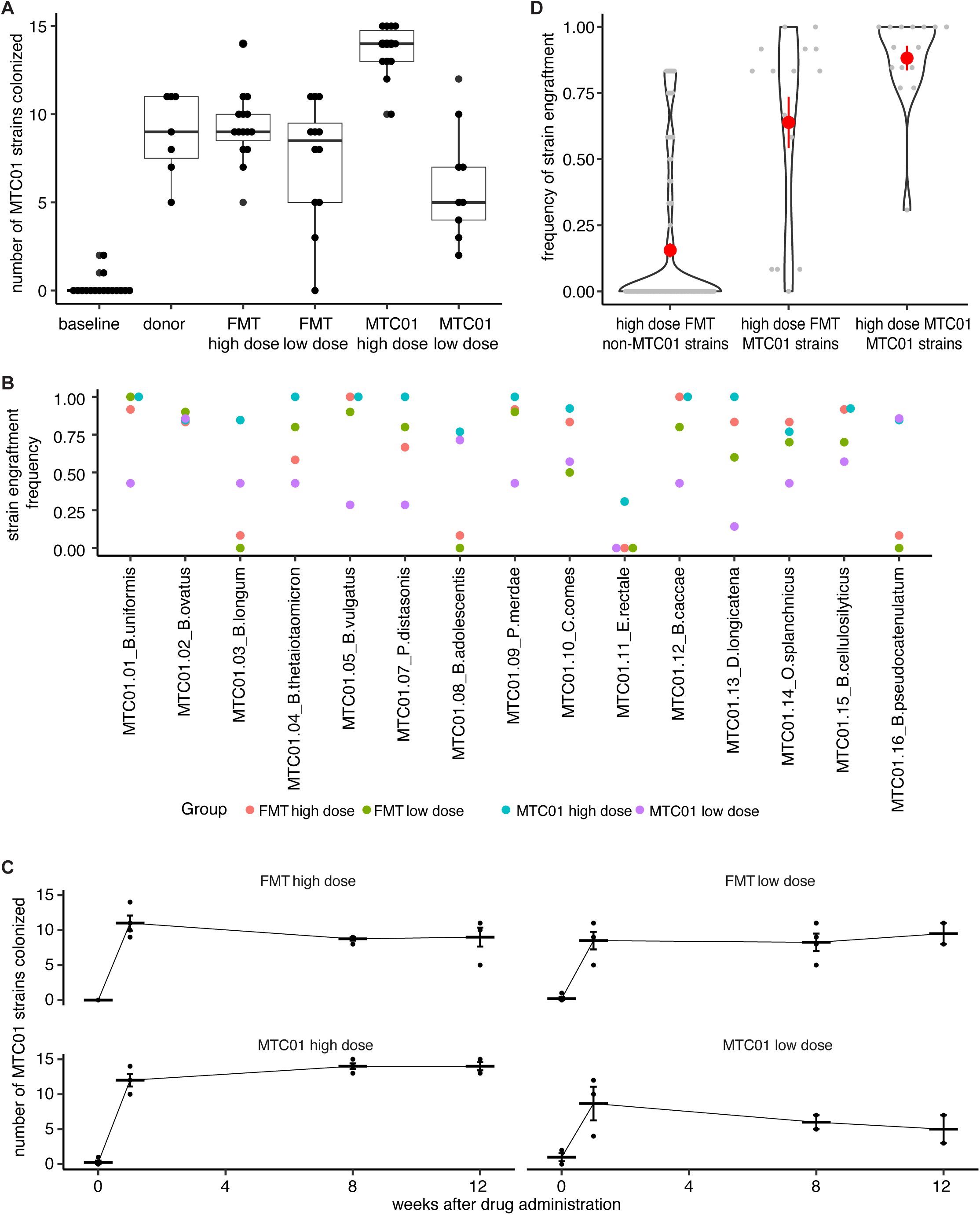
Engraftment of strains in FMT and MTC01 recipients. (A) Few of the MTC01 strains were detected in baseline samples from all recipients prior to drug administration. After administration up to all 15 MTC01 strains were detected in recipients with the most strains detected in recipients of the high dose MTC01. (B) Individual strains generally had high engraftment frequencies when administered in the high dose MTC01, lower engraftment frequencies when administered in the low dose MTC01, and varied engraftment when administered in the FMT drug products. (C) Strain engraftment was stable after the single dose administration of live microbial therapeutic. (D) The 15 MTC01 strains had higher engraftment frequencies than the non-MTC01 strains in donor D283 both when administered via FMT or LBP. Each gray dot represents the engraftment frequency of an individual bacterial strain across all post-dosing recipient samples. The red dots and error bars represent the mean ± stderr of the engraftment frequency of all strains in a group.

Low dose MTC01 had the fewest number of strains colonized suggesting a likely dose effect in the LBP recipients. This dose effect was consistent across strains except for *Bacteroides ovatus*, *B. adolescentis*, and *B. pseudocatenulatum* that colonized with similar frequencies in both high and low dose MTC01 (Figure 3B). As we had previously observed for FMT^8^, we find that once colonized in recipients the strains dosed either via FMT or LBP remained durably colonized for the duration of the sampling period (Figure 3C).

To test our hypothesis that selection of frequently engrafting strains via empirical FMT results could focus LBP manufacturing on strains that engraft well rather than spending resources on strains that poorly engraft, we assessed that engraftment frequency of MTC01 strains compared with the engraftment of strains from D283 that are not in the 15 MTC01 strains. We find the 15 MTC01 strains have significantly higher engraftment frequencies compared to non-MTC01 strains both when administered by FMT or by LBP (p = 4.1×10^−8^ and 4.8×10^−12^ respectively; Wilcoxon signed-rank test; Figure 3D). Many of the non-MTC01 strains from D283 that were detected in ≥50% of samples were strains of the same species that were not included due to our design criterion of including only one strain per species in the LBP. The only species of high engrafting strains without a representative in MTC01 were *Bifidobacterium bifidum*, *Bacteroides fragilis* (not included in MTC01 as it had multiple antibiotic resistances), *Barnesiella intestinihominis*, *Bacteroides stercoris*, *Blautia massiliensis*, *Blautia wexlerae*, *Collinsella aerofaciens*, *Catenibacterium mitsuokai*, *Ruminococcus bicirculans*, and *Ruminococcus torques*.

## Discussion

The strong safety profile of LBPs^14,16,30,31^, the growing set of tools to track their engraftment^8,22,30,32–35^, and their promising results in clinical studies make them a compelling drug modality for future trials^36^. Unlike FMT based discovery trials, LBPs allow testing of clearly defined hypotheses^37–45^ regarding the microbiome’s influence on disease with tracking of each strain to identify associations with markers of disease response. Although numerous academic and commercial laboratories have the capacity to grow enough anaerobic bacteria for clinical trials with LBPs, the lack of know-how and added quality and regulatory burden for safely manufacturing them for human trials is a barrier that has limited the translation of numerous promising discoveries in animal models and human observational studies. To alleviate this barrier, we developed a manufacturing platform requiring only a hydrogen peroxide ionizer and a dedicated anaerobic chamber – both of which are straightforward to use and practical with most laboratory budgets. We provide regulatory documentation and manufacturing batch records as blueprints to accelerate future exploratory trials.

A key limitation to the manufacturing process used here is the time restriction of only manufacturing two strains per week, which could be prohibitive to produce very large consortia. However, there is ample space in the anaerobic chamber to grow far more strains in parallel if adequate controls were established. A second limitation is the endoscopic formulation, where most patients would prefer an oral delivery. Although we chose a colonoscopic delivery in this study to compare FMT and LBP without confounding variables of enteric capsule delivery that might cloud interpretation, the drug substance could be formulated for oral delivery.

Manipulation of living cells or their protein constituents *in vitro* or *ex vivo* is fundamental to numerous therapies including *in vitro* fertilization, biologics, and CAR-T cells. These *ex vivo* and *in vitro* processes risk altering the chemical and structural composition in ways that reduce their efficacy relative to the same components generated in their native environment. Although the *in vitro* manufactured strains in MTC01 are grown in monoculture without many of the environmental constitutes they would receive from a diverse microbiome and human host, we find *in vitro* manufactured monocultures colonize human recipients as well or better than the same bacterial strains cultivated in a human gut with similar efficacy and safety in the treatment of rCDI, supporting the future use of *in vitro* manufactured LBPs. Finally, we demonstrate that isolating bacterial strains from FMT donors and using those with the highest empirical engraftment frequency in human recipients provides a strong selection for strains that will engraft when administered as an LBP for all therapeutic strains whose functional benefit requires durable colonization. Selection of consistent engrafting strains reduces the burden of manufacturing and clinically evaluating strains that poorly colonize the host and thus cannot realize their therapeutic potential.

## Supporting information

Bethlehem etal MTC01 Master Batch Records

Bethlehem etal MTC01 preIND

Bethlehem etal MTC01 IND

Bethlehem etal MTC01 tables and supp tables

## Data Availability

All data produced in the present study are available upon reasonable request to the authors.

## Acknowledgements

This work was performed with the support of staff and facilities of the Icahn School of Medicine Gnotobiotic Facility, and the Microbiome Translation Center at Mount Sinai. This work was supported in part through the Minerva computational and data resources and staff expertise provided by Scientific Computing and Data at the Icahn School of Medicine at Mount Sinai and supported by the Clinical and Translational Science Awards (CTSA) grant UL1TR004419 from the National Center for Advancing Translational Sciences. Research reported in this publication was also supported by the Office of Research Infrastructure of the National Institutes of Health under award number S10OD026880 and S10OD030463.

---

**Figure S1.**
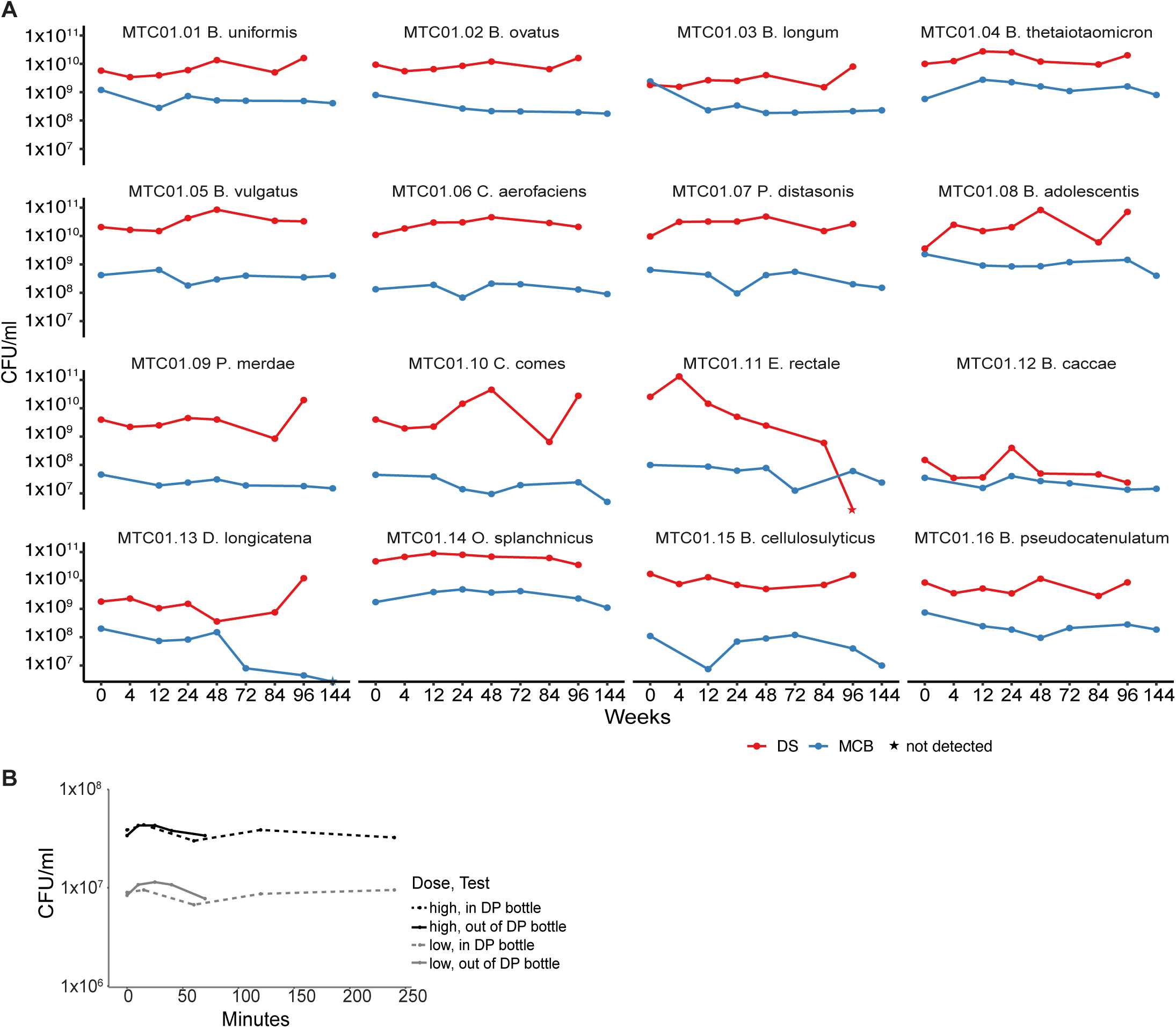
Strain specific stability. (A) The MCB and DS cell banks were largely stable over the sampling period except for *Eubacterium rectale* and *Dorea longicatena* which showed a reduction in potency over time. (B) DP potency was stable in the sealed bottle for at least 4 hours (typical thawing-time in clinic is about 1 hour) and outside the bottle for at least 1 hour (typical time outside the bottle in clinic is about 5 minutes).

**Table S1.** In vitro antibiotic minimum inhibitory concentrations assessed by CLSI and e-test.

**Table S2.** Manufacturing and trial timelines.

**Table S3.** Strain specific dosing in FMT and MTC01 batches.

## Online Methods

### Strain Isolation

Clarified and diluted donor stool was plated onto a variety of solid selective and non-selective media under anaerobic, microaerophilic and aerobic conditions. Plates were incubated for 48-72 hours at 37°C. 384 single colonies from each donor microbiota were individually picked and regrown in liquid LYHBHIv4 media for 48 hours under anaerobic conditions (see supplemental materials; IND Appendix 9 for growth media details) ^7,8,46^. Regrown isolates were identified at the species level using a combination of MALDI-TOF mass spectrometry (Bruker Biotyper) and 16S rDNA amplicon sequencing. All regrown isolates were stored in LYHBHIv4 with 15% glycerol at −80 °C and strains for use in MTC01 were genome sequenced with paired-end 150nt reads on an Illumina platform.

### Manufacturing of MTC01

In addition to the methods described here, a detailed description can be found in the IND and MBRs (supplemental materials). To produce RCBs, single colonies were selected after plating the relevant strains from the donor D283 culture library on rich solid agar (chocolate agar). Up to 20 freezer stocks were made of each strain (LYHBHIv4 with 15% glycerol) for use in strain characterization and the generation of master cell banks. The test for antibiotic resistance of each strain was performed in-house by e-test on solid agar and through an external CRO (Micromyx) according to CLSI standards (see Table S1 and supplemental materials IND Appendix 3 for full report).

A dedicated vinyl flexible film two-sided anaerobic chamber with a sterilization port on each side of the chamber was used for the manufacture of MCB, DS, and DP (see supplemental materials IND Appendix 5 for facility design). Compared to a bioreactor, the chambers are relatively robust in operation, require little training and are quickly operational after installation. The confined space of the chamber allowed us to design a unique and simple sterilization process that grants easier microbial control than a more typical setup of gowned staff in a clean room. Prior to manufacturing of MCB, DS, or DP, almost all equipment and consumables are sterilized in place with ionized hydrogen peroxide. The few items that must be imported (e.g., the strains to be cultured) are surface sterilized with ethanol and imported through the gas exchange port that uses a HEPA filtered air supply.

All fermentations and culture manipulations were performed in single-use, disposable, sterilized plasticware with animal product free LYH_VIB media (vegitone infusion broth [Sigma-Aldrich], yeast extract, monosaccharide mix, disaccharide mix, cysteine, malic acid, sodium sulfate, MOPS, menadione, tween 80, vitamin-K; see supplemental materials IND Appendix 9 for growth media details), and each strain was manufactured in a dedicated side of the anaerobic chamber with at most two strains manufactured in parallel. OD600 was used for in process monitoring of cell density. Colony forming units was the primary potency assessment for the cell banks and products. For MCBs, a 10 mL starter culture in a conical tube was inoculated with the total volume of the 2 mL RCB glycerol stock and varying potency of 2.0×10^4^ - 2.0×10^9^ CFU/mL. After reaching sufficient cell density, the starter culture was used to inoculate the primary MCB fermentation volume of 100 mL to OD600 0.1-0.2 in a single-use shake flask. In late exponential to early stationary growth phase, the 100 mL culture was mixed 1:1 with cryo-buffer (PBS pH 7.5, 0.5 g/L cysteine, 30% glycerol). Up to 192×1 mL MCB cryo-tubes were generated for each strain. The timepoint of harvest for each strain was estimated from liquid batch fermentations of RCB cultures utilizing the same growth medium.

For DS generation, three 100 mL starter cultures were each inoculated with 1-3 mL MCB glycerol stock depending on the stock potency (3.5×10^7^ - 2.4×10^9^ CFU/mL). After reaching sufficient cell density, starter cultures were used to inoculate 1.4 L DS primary fermentations to a target OD600 of 0.1-0.2 in a single-use shake flask.

After DS primary fermentation reached late exponential to early stationary growth phase, strains were centrifuged and washed with United States Pharmacopeia (USP) grade phosphate buffered saline (PBS, pH 7.5, 0.5 g/L cysteine) to remove non-USP grade components of the growth media and bacterial fermentation products.

Centrifugation to pellet the bacterial cells was performed in sterile single-use 250 mL centrifuge tubes spun at 4000 RCF for 15 mins. After washing, each DS was centrifuged again and resuspended at approximately 1:10 the original volume (140ml) in USP grade formulation buffer (PBS, 0.5 g/L cysteine, 30% glycerol) to facilitate mixing of all 15 strains into a 250 mL DP. The DS was stored in 4 x 35 mL aliquots at −80°C in sterile 60 mL cryo-bottles along with 0.25 mL QC-aliquots in 10 cryo-tubes for downstream stability monitoring, although in retrospect 20 cryo-tubes would have been better to allow for retests and extended stability.

The DP was produced by simultaneously thawing one 35 mL DS aliquot for each strain in a sterile room temperature water bath in the anaerobic chamber. We generated DP at a high dose (1×10^11^ CFU) and a low dose (1×10^10^ CFU), with the high dose as close as practical to the average dose of FMT previously used with D283 (4.95×10^11^) and the low dose targeting 10-fold lower than the high dose. DP was made in batches of 5-6 high doses per batch (or an equivalent CFU combination of high doses and low doses; e.g., 5 high doses and 10 low doses). For strains with insufficient viable cells to provide 1:15 of the target dose for at least 15 high and 15 low doses using a single 35 mL DS aliquot, the full 35 mL was used to reach the highest strain dose possible. For the remaining strains, an equal potency of cells was added to the DP to reach the target dose in a total volume of 250 mL. The resulting strain-specific doses are in Supplementary Table 3. Three rounds of DP production (Table 1) were sufficient to reach our target of 15 high doses and 15 low doses, leaving one aliquot of DS per strain for future use.

### Quality assessment and stability of MTC01

The characterization of the MCB, DS and DP used colony forming units (CFU) as the primary potency metric. CFU was measured by spot-plating ten times 1 uL of four dilutions of an aliquot of a bacterial culture on at least 2 chocolate agar plates. MALDI-TOF biotyping (Bruker) was used as the primary identity metric for strain manufacturing characterization. The composition of the MCB, DS and DP, was analysed by spread plating dilutions from an aliquot of the individual strain (MCB and DS) or the mixture of all strains (DP) on chocolate agar plates followed by picking individual colonies for Biotyping. For MCB and DS at least 48 high quality spectra reads (confidence score >1.8) were analysed, and for the DP at least 200 high quality spectra reads were analysed. All high quality MALDI-TOF spectra had to match the correct MTC01 strain or strain mixture (i.e. no single colony mismatch was allowed). In the identification of strains in the DP mixture, 14 out of 15 strains of MTC01 were identified. Among the 1440 colonies picked for all three DP production runs, only MTC01.11 *E. rectale* was not detected by Biotyping. In monoculture, *E. rectale* has slower growth on chocolate agar compared to the remaining 14 bacterial strains, reducing chances of identification. For this DP produced for a phase 1b clinical trial the quantification of all viable bacterial strains within the mixed DP was not a release criterion, and it remains a challenge in the field of defined microbiota therapeutics^47^.

For the microbiological examination of nonsterile products, USP<61> (microbial enumeration tests) and USP<62> (tests for specific microorganisms) are frequently used bioburden assays. Except for the specific test for *Clostridia* in USP<62>, all culture conditions for USP61 and USP62 use aerobic growth on rich agar. We cultured all individual MTC01 strains aerobically on rich agar and found all strains were strict anaerobes with no aerobic growth. An external CRO (Focus Laboratories, Allentown, PA), subsequently performed USP<61> and USP<62> suitability assays on all strains individually as well as on the combined MTC01 community, demonstrating that none of the strains or communities inhibit the target bacteria and fungi in the USP<61> and USP<62> assays (including *Clostridium sporogenes* in anaerobic conditions). All MCB, DS, and DP passed the USP bioburden assessments.

Since clinical trials take months or years to recruit and drugs need a defined shelf-life, monitoring the stability of the MCB, DS, and DP in long-term storage is critical. Based on the common storage duration of FMT at −80 °C for up to two years^19^, we would expect individual bacterial strains and the defined LBP would likewise remain stable for extended periods. The CFU of the 15 individual strains (MCB, DS) and the 15 mixed strains (DP) stored at −80°C in cryo-buffer (PBS, 0.5 g/L cysteine, 15-30% glycerol) has been measured for at least 96wks (Figure S1).

Given the need for flexible time windows when performing the clinical procedure of thawing and application of the DP, we quantified a range of times for these two clinical steps and found the total potency of DP is stable under aerobic conditions in the cryogenic storage bottle post thawing for at least 4 hours and in the clinical tray for at least 1 hour (Figure S1).

### Manufacturing of donor D283 FMT drug product

Fecal samples from D283 were collected, weighed, and split into 40-80 g subsamples in Seward Stomacher filter bags. Each sample was treated separately; the subsamples were weighed and stored at −80°C until production of concentrated fecal slurry (cFS). Each stool’s subsamples were homogenized with 6mL/g cryogenic media (PBS, 0.5 g/L cysteine, 30% glycerol) through the Seward Stomacher EVO 400. The homogenized cFS from the filtered side of the stomacher bags was combined into 1 L collection bottles(s) and weighed. After combining all cFS from a stool’s subsamples, the containers were weighed and stored at −80°C along with three 100 µl QC aliquots. QC vials were freeze-thawed after ≥24 hours, returned to −80 °C for another 24 hours, then spot-plated ten times 1 µl from 10^−3^-10^−6^ dilutions across two chocolate agar plates to determine cFS CFU/ml potency.

The volume of cFS needed for high or low dose was calculated based on CFU potency of the QC vials. Cryo-media was prepared to normalize the cFS to 1 × 10^10^ for low doses and 1 × 10^11^ for high doses in a final volume of 250ml FMT (Figure 1B). Ten 100 µl QC vials were each taken from a low and high dose product container for stability assessment (Figure 1C). The QC vials were spot-plated as the cFS QC with ten times 1 µl from 10^−3^-10^−6^ dilutions across two chocolate agar plates to determine FMT CFU/total 250ml dose potency (Table 1).

### Recruitment and administration of drug to rCDI patients

Patients (≥18 years old) with 2 or more qualifying CDI episodes occurring within the prior 6 months, inclusive of the current episode were consented at Mount Sinai Hospital under IRB STUDY-23-00563 (NCT05911997). Prior to colonoscopic drug administration, subjects were taken off all antibiotics for 48hr and underwent a standard colonoscopy bowel preparation within 12 hours of their procedure. Patients were block randomized (block size = 4) to receive either high dose FMT, low dose FMT, high dose MTC01, or low dose MTC01. The relevant microbial therapeutic was administered to the right colon by colonoscopy in a total volume of 250 mL.

### Detection of MTC01 and donor D283 FMT bacterial strain colonization

All sequenced bacterial strains from donor D283 were tracked using the open source strainer algorithm^8,29^. Briefly the algorithm uses a database of bacterial strains and metagenomes not isolated from donor D283 to identify unique regions of the D283 strains. These unique regions are quantified in metagenomic samples of the donor and the recipients of FMT or MTC01. A strain is designated as detected if the informative kmers is above that estimated in a background database of unrelated metagenomes. Since the original publication, we have slightly modified the algorithm to 1) make it more streamlined for automation in high performance computing environments and 2) used the breadth of informative kmers covered in the metagenomes rather than the depth as the metric for scoring when comparing with the background database of metagenomes.

Strain-specific potencies in the FMT drug product batches (Table S3) were estimated from the total product potency multiplied by the strain relative abundance determined by the strainer algorithm.

### Statistical analyses

Linear mixed effects models, using the lme4 R package^48^, were used to assess differences in engraftment before and after treatment with microbial therapeutics as well as between treatment modalities/doses, while accounting for non-independence between samples from the same subject. For comparisons between strain engraftment with MTC01 strains pre- vs post-dosing strains for all drugs and doses (FMT high dose, FMT low dose, MTC01 high dose, MTC01 low dose), we used the following model:

*number_MTC01_strains_detected ∼ drug_and_dose + 1|patient_id*

that accounts for the non-independence between samples collected over time from the same individual, post-dosing. Baseline samples were pooled across all drugs and doses.

The same model model structure was used to compare the number of MTC01 strains engrafted post-dosing between FMT high dose vs the three remaining drug/dose combinations (FMT low dose, MTC01 high dose, MTC01 low dose) – again accounting for non-independence between between samples collected over time from the same patient.

## References

1. Khanna, S. et al. Efficacy and Safety of RBX2660 in PUNCH CD3, a Phase III, Randomized, Double-Blind, Placebo-Controlled Trial with a Bayesian Primary Analysis for the Prevention of Recurrent Clostridioides difficile Infection. Drugs 82, 1527–1538 (2022).

2. Feuerstadt, P. et al. SER-109, an Oral Microbiome Therapy for Recurrent *Clostridioides difficile* Infection. N Engl J Med 386, 220–229 (2022).

3. van Nood, E. et al. Duodenal infusion of donor feces for recurrent Clostridium difficile. N Engl J Med 368, 407–415 (2013).

4. Kao, D. et al. Effect of Oral Capsule– vs Colonoscopy-Delivered Fecal Microbiota Transplantation on Recurrent *Clostridium difficile* Infection: A Randomized Clinical Trial. JAMA 318, 1985 (2017).

5. Allegretti, J. R. et al. Inflammatory Bowel Disease Outcomes Following Fecal Microbiota Transplantation for Recurrent C. difficile Infection. Inflamm Bowel Dis 27, 1371–1378 (2021).

6. Youngster, I. et al. Oral, capsulized, frozen fecal microbiota transplantation for relapsing Clostridium difficile infection. JAMA 312, 1772–1778 (2014).

7. Chen-Liaw, A. et al. Gut microbiota strain richness is species specific and affects engraftment. Nature (2024) doi:10.1038/s41586-024-08242-x.

8. Aggarwala, V. et al. Precise quantification of bacterial strains after fecal microbiota transplantation delineates long-term engraftment and explains outcomes. Nat Microbiol 6, 1309–1318 (2021).

9. Faith, J. J., et al. Strain Population Structure Varies Widely across Bacterial Species and Predicts Strain Colonization in Unrelated Individuals. http://biorxiv.org/lookup/doi/10.1101/2020.10.17.343640 (2020) doi:10.1101/2020.10.17.343640.

10. Drewes, J. L. et al. Transmission and clearance of potential procarcinogenic bacteria during fecal microbiota transplantation for recurrent Clostridioides difficile. JCI Insight 4, 130848 (2019).

11. DeFilipp, Z. et al. Drug-Resistant *E. coli* Bacteremia Transmitted by Fecal Microbiota Transplant. N Engl J Med 381, 2043–2050 (2019).

12. Zellmer, C. et al. Shiga Toxin-Producing Escherichia coli Transmission via Fecal Microbiota Transplant. Clin Infect Dis 72, e876–e880 (2021).

13. Louie, T. et al. VE303, a Defined Bacterial Consortium, for Prevention of Recurrent Clostridioides difficile Infection: A Randomized Clinical Trial. JAMA 329, 1356–1366 (2023).

14. Rode, A. A. et al. Randomised clinical trial: a 12-strain bacterial mixture versus faecal microbiota transplantation versus vancomycin for recurrent Clostridioides difficile infections. Aliment Pharmacol Ther 53, 999–1009 (2021).

15. Petrof, E. O. et al. Stool substitute transplant therapy for the eradication of Clostridium difficile infection: ‘RePOOPulating’ the gut. Microbiome 1, 3 (2013).

16. Kao, D. et al. The effect of a microbial ecosystem therapeutic (MET-2) on recurrent Clostridioides difficile infection: a phase 1, open-label, single-group trial. Lancet Gastroenterol Hepatol 6, 282–291 (2021).

17. Tvede, M. & Rask-Madsen, J. Bacteriotherapy for chronic relapsing Clostridium difficile diarrhoea in six patients. Lancet 1, 1156–1160 (1989).

18. Varga, A. et al. How to Apply FMT More Effectively, Conveniently and Flexible – A Comparison of FMT Methods. Front. Cell. Infect. Microbiol. 11, 657320 (2021).

19. Terveer, E. M. et al. How to: Establish and run a stool bank. Clin Microbiol Infect 23, 924–930 (2017).

20. Kelly, C. R., Kunde, S. S. & Khoruts, A. Guidance on preparing an investigational new drug application for fecal microbiota transplantation studies. Clin Gastroenterol Hepatol 12, 283–288 (2014).

21. Hirten, R. P. et al. Microbial Engraftment and Efficacy of Fecal Microbiota Transplant for Clostridium Difficile in Patients With and Without Inflammatory Bowel Disease. Inflamm. Bowel Dis. 25, 969–979 (2019).

22. Ianiro, G. et al. Variability of strain engraftment and predictability of microbiome composition after fecal microbiota transplantation across different diseases. Nat Med 28, 1913–1923 (2022).

23. Claypool, J. et al. Microbiome compositional changes and clonal engraftment in a phase 3 trial of fecal microbiota, live-jslm for recurrent Clostridioides difficile infection. Gut Microbes 17, 2520412 (2025).

24. Seekatz, A. M. et al. Recovery of the gut microbiome following fecal microbiota transplantation. mBio 5, e00893–00814 (2014).

25. Hamilton, M. J., Weingarden, A. R., Unno, T., Khoruts, A. & Sadowsky, M. J. High-throughput DNA sequence analysis reveals stable engraftment of gut microbiota following transplantation of previously frozen fecal bacteria. Gut Microbes 4, 125–135 (2013).

26. FDA. Fecal Microbiota for Transplantation: Safety Alert - Risk of Serious Adverse Events Likely Due to Transmission of Pathogenic Organisms. https://www.fda.gov/safety/medical-product-safety-information/fecal-microbiota-transplantation-safety-alert-risk-serious-adverse-events-likely-due-transmission (2020).

27. FDA. Fecal Microbiota for Transplantation: Safety Communication-Risk of Serious Adverse Reactions Due to Transmission of Multi-Drug Resistant Organisms. https://www.fda.gov/safety/medical-product-safety-information/fecal-microbiota-transplantation-safety-communication-risk-serious-adverse-reactions-due (2019).

28. FDA. Early Clinical Trials with Live Biotherapeutic Products: Chemistry, Manufacturing, and Control Information.

29. strainer2 algorithm for detecting bacterial strains inside metagenomes.

30. Dsouza, M. et al. Colonization of the live biotherapeutic product VE303 and modulation of the microbiota and metabolites in healthy volunteers. Cell Host Microbe 30, 583–598.e8 (2022).

31. Glitza, I. C. et al. Randomized Placebo-Controlled, Biomarker-Stratified Phase Ib Microbiome Modulation in Melanoma: Impact of Antibiotic Preconditioning on Microbiome and Immunity. Cancer Discov 14, 1161–1175 (2024).

32. Valles-Colomer, M. et al. The person-to-person transmission landscape of the gut and oral microbiomes. Nature 614, 125–135 (2023).

33. Faith, J. J. Assessing live microbial therapeutic transmission. Gut Microbes 17, 2447836 (2025).

34. Olm, M. R. et al. inStrain profiles population microdiversity from metagenomic data and sensitively detects shared microbial strains. Nat Biotechnol 39, 727–736 (2021).

35. Smillie, C. S. et al. Strain Tracking Reveals the Determinants of Bacterial Engraftment in the Human Gut Following Fecal Microbiota Transplantation. Cell Host Microbe 23, 229–240.e5 (2018).

36. Sorbara, M. T. & Pamer, E. G. Microbiome-based therapeutics. Nat Rev Microbiol 20, 365–380 (2022).

37. Buffie, C. G. et al. Precision microbiome reconstitution restores bile acid mediated resistance to Clostridium difficile. Nature 517, 205–208 (2015).

38. Lawley, T. D. et al. Targeted restoration of the intestinal microbiota with a simple, defined bacteriotherapy resolves relapsing Clostridium difficile disease in mice. PLoS Pathog 8, e1002995 (2012).

39. Battaglioli, E. J. et al. Clostridioides difficile uses amino acids associated with gut microbial dysbiosis in a subset of patients with diarrhea. Sci Transl Med 10, eaam7019 (2018).

40. McCulloch, J. A. et al. Intestinal microbiota signatures of clinical response and immune-related adverse events in melanoma patients treated with anti-PD-1. Nat Med 28, 545–556 (2022).

41. Hayase, E. et al. Bacteroides ovatus alleviates dysbiotic microbiota-induced graft-versus-host disease. Cell Host Microbe 32, 1621–1636.e6 (2024).

42. Bredon, M. et al. Faecalibaterium prausnitzii strain EXL01 boosts efficacy of immune checkpoint inhibitors. Oncoimmunology 13, 2374954 (2024).

43. Seekatz, A. M. Straining to define a healthy microbiome. mSphere 10, e0079724 (2025).

44. Theriot, C. M., Bowman, A. A. & Young, V. B. Antibiotic-Induced Alterations of the Gut Microbiota Alter Secondary Bile Acid Production and Allow for Clostridium difficile Spore Germination and Outgrowth in the Large Intestine. mSphere 1, e00045–15 (2016).

45. Smith, A. B. et al. Enterococci enhance Clostridioides difficile pathogenesis. Nature 611, 780–786 (2022).

46. Britton, G. J. et al. Defined microbiota transplant restores Th17/RORγt+ regulatory T cell balance in mice colonized with inflammatory bowel disease microbiotas. Proc. Natl. Acad. Sci. U.S.A. 117, 21536–21545 (2020).

47. Coryell, M. P., Sava, R. L., Hastie, J. L. & Carlson, P. E. Application of MALDI-TOF MS for enumerating bacterial constituents of defined consortia. Appl Microbiol Biotechnol 107, 4069–4077 (2023).

48. Bates, D., Mächler, M., Bolker, B. & Walker, S. Fitting Linear Mixed-Effects Models Using lme4. J. Stat. Soft. 67, (2015).

