## Supplementary material for "Live biotherapeutic product exhibits similar efficacy and superior engraftment to same donor fecal microbiota transplant for recurrent Clostridioides difficile infection": Bethlehem etal MTC01 Master Batch Records

[*] Co-corresponding authors

### Master Batch Records

for

MTC01 Master Cell Banks

MTC01 Drug Substance

MTC01 Drug Product

FMT Drug Product

Bethlehem et.al., 2025

### MTC01

Master Cell Banks

Master Batch Records

Bethlehem et.al., 2025

| **Reviewed By:** | | **Date:** |
|---|---|---|
| Jeremiah Faith, PhD<br>Faculty Director | | |
| **Approved By:** | | **Date:** |
| Ilaria Mogno, PhD<br>Quality Assurance Specialist | | |
| Control No: | | |

| |
|---|
| **STUDY ID** |
| **Batch number**<br>***(yyyy-mm-dd-MTC01.Strain#)*** |

#### 1. REFERENCE SOP(s)

- Generation of Master/Working Cell Banks (MCB/WCB) for Live Biotherapeutic Products (LBP), MTC-SOP-0001
- Culture Media for Live Biotherapeutic Products (LBP), MTC-SOP-0002
- CFU Count for Live Biotherapeutic Products (LBP), MTC-SOP-0003
- Biotyping for Live Biotherapeutic Products (LBP), MTC-SOP-0004
- Sterilization of Anaerobic Chambers for Live Biotherapeutic Products (LBP), MTC-SOP-0005

#### 2. ADDITIONAL DOCUMENTS NEEDED

- Reference cultivations for MTC01.docx

| Performed by (Init/Date) | Comments: |
|---|---|

#### 3. EQUIPMENT CHECKLIST

| | | | |
|---|---|---|---|
| **Anaerobic Chamber for this production run:** | | | |
| **Equipment Type** | **Serial Number** | **Post-Production Cleaning** | |
| | | **Initials** | **Date** |
| Anaerobic Chamber | AC21-092 | | |
| Biological Safety Cabinet | 1309819038 | | |
| Cell Density Meter | 1601 (Chamber 1), 1602 (Chamber 2) | | |
| Freezer -80 °C | | Not applicable | Not applicable |
| Refrigerator | Not applicable | Not applicable | Not applicable |
| Laboratory Oven | G1-010356 | Not applicable | Not applicable |
| Micropipette (10μL) | G42248J (Chamber 1), G42132J (Chamber 2) | | |
| Micropipette (100μL) | J48012J (Chamber 1), J48210J (Chamber 2) | | |
| Micropipette (1000μL) | J45407J (Chamber 1), J45421J (Chamber 2) | | |
| Multichannel Pipette | SH25393 (Chamber 1), SH35385 (Chamber 2) | | |
| Pipetting Controller | B03320691 (Chamber 1), B03320694 (Chamber 2) | | |
| Repeater Pipette | P38164J (Chamber 1), M48855G (Chamber 2) | | |
| Cuvette Rack | Not applicable | | |
| Cryogenic Vial Rack | Not applicable | | |
| Matrix Tube Decapper | | | |
| Tweezer | Not applicable | | |
| Vortex Mixer | 20060874 (Chamber 1), 21011317 (Chamber 2) | | |
| Waste Pouch Holder | Not applicable | | |
| Thermometer | Not applicable | | |
| *Other:* | | | |
| *Other:* | | | |

| Performed by (Init/Date) | Comments: |
|---|---|

#### 4. REAGENTS AND SUPPLIES

| Item | Manufacturer | Catalog No. | Lot/Batch No. | Exp. Date |
|---|---|---|---|---|
| Sampling Tubes, 15 mL | | | | |
| Sampling Tubes, 50 mL | | | | |
| Sampling Tubes, 1.5 mL | Thermo Scientific | Not applicable | Not applicable | Not applicable |
| UV Cuvettes | Thomas Scientific | Not applicable | Not applicable | Not applicable |
| Serological Pipets, 10 mL | Corning | | | |
| Serological Pipets, 50 mL | Corning | | | |
| Production Flasks, 125 mL | Thomson Instrument | | | |
| Glass Beads | Midland Scientific | 100B | Not applicable | Not applicable |
| Pipet Tips, 10µL | USA Scientific | Not applicable | Not applicable | Not applicable |
| Pipet Tips, 100µL | USA Scientific | Not applicable | Not applicable | Not applicable |
| Pipet Tips, 1000µL | USA Scientific | Not applicable | Not applicable | Not applicable |
| Multichannel Tips | Thermo Scientific | 94420043 | Not applicable | Not applicable |
| Combitips Advanced, 10 mL | Eppendorf | | | |
| Syringe, 1 mL | | | | |
| Syringe Needle | | | | |
| Matrix Tubes | Thermo Scientific | | | Not applicable |
| Matrix Tube Rack | Thermo Scientific | | | Not applicable |
| 96 well plates | Corning | 3697 | Not applicable | Not applicable |
| Sterile Towelettes | Medline | MDS094181 | Not applicable | Not applicable |
| Ziploc Bag | Office Depot | Not applicable | Not applicable | Not applicable |
| Chocolate Agar Plates | BD Biosciences | 221169 | | |
| Ethanol (70%) | Fisher Scientific | 25-467-01 | | |
| Phosphate Buffer Solution | Caisson Labs | PBL06 | | |
| *Other:* | | | | |

| Performed by (Init/Date) | Comments: |
|---|---|

#### 5. Preparation

| **Initial Preparation**<br>**(Can be done one to four days before the cells are thawed)** | | **Done (√)** |
|---|---|---|
| Print and attach *Reference cultivations for MTC01* to this batch record. | | |
| Scan two racks full of Matrix tubes (bottom label) and generate Freezerworks entries.<br>*Note: Label racks temporarily with Strain Name and Box number.* | | |
| Print 192 labels for Matrix tubes and one extra label with the following information:<br>**MTC01.*StrainID*-MCB *or* WCB** **BARCODE**<br>***Strain Name + Vial Number (1-192)***<br>***Date printed***<br>*Note: The barcode is the samples global unique identifier in the Freezerworks.* | Stick extra label here: | |
| Print 2 labels for Matrix tube racks and one extra label with the following information:<br>***Strain Name***<br>**MTC01.*StrainID***<br>**MCB Box *Number*** **BARCODE**<br>***Date printed***<br>*Note: The barcode is the parent global unique identifier in the Freezerworks.* | Stick extra label here: | |
| Attach the labels to the Matrix tubes and racks.<br>*Note: Make sure to attach the labels to the correct racks and tubes, then remove temporary label on racks.* | | |
| Regenerate catalyst plates:<br>☐ Set the laboratory oven to 200°C (level 4)<br>☐ place catalyst plates in the oven<br>☐ Remove catalyst plates after 1 hour and install plates in anaerobic chamber fan | | |
| Print and attach *MTC-BR-0002_Culture Media for Live Biotherapeutic Products (LBP)* to this batch record and prepare culture media and cryogenic media accordingly. | | |
| Aliquot the following supplies under the biosafety cabinet:<br>☐ 20 mL sterile PBS into a sterile sampling tube (50 mL)<br>☐ Sterile glass beads into a sterile sampling tube (15 or 50 mL, until the 10 mL mark) | | |
| Place equipment and supplies in the anaerobic chamber:<br>☐ Sterile culture media<br>☐ Sterile cryogenic media<br>☐ 2 labeled Matrix tube Racks with 96 labeled Matrix tubes each<br>☐ 1 bag of additional Matrix tubes (48 tubes)<br>☐ Chocolate agar plates (2 stacks of 10 plates)<br>☐ 1 Sterile PBS aliquot<br>☐ 1 cuvette-rack with 12 UV cuvettes<br>☐ 1 production flask | | |

| Performed by (Init/Date) | Comments: |
|---|---|

| |
|---|
| ☐ 7 sampling tubes (50 mL) in sampling tube rack (50 mL)<br>☐ 1 box of each micropipette tips (10 µL, 100 µL, 1000 µL)<br>☐ Pipette controller (recharged)<br>☐ 5 10 mL and 2 10 mL serological pipettes<br>☐ Electronic multichannel pipette (recharged)<br>☐ 1 box multichannel pipette tips (12.5 µL)<br>☐ Electronic repeater pipette (recharged)<br>☐ 4 Biopur Combitips (10 mL)<br>☐ Matrix tube decapper and charger (recharged)<br>☐ 15 syringes (1 mL) and 5 needles<br>☐ 10 sampling tubes (1.5 mL, open in vial rack)<br>☐ Sterile glass beads<br>☐ 1 squeeze bottle ethanol (70 %)<br>☐ 3 Biohazard waste bags<br>☐ 2 Ziploc bags<br>☐ 1 sterile 96 well plate<br>☐ 5 sterile Towelettes |
| Print and attach *MTC-MBR-0005_Sterilization of Anaerobic Chamber for Live Biotherapeutic Products (LBP)* to this batch record and perform the sterilization accordingly, at least 24hours before the start of cultivation. |
| Tear open the chocolate agar plate bags, place in Ziploc bag and leave about 1 inch of the Ziploc bag open. |

| Performed by (Init/Date) | Comments: |
|---|---|

#### 6. Cultivation – Starter Culture

| If hydrogen level <2.0% and/or oxygen level >50 ppm, exchange chamber air with new gas mix. | Hydrogen level [%] | Oxygen level [ppm] |
|---|---|---|

| Preparation (In the anaerobic chamber) | Done (√) |
|---|---|
| Fill 1 sampling tube (50 mL) with 20 mL culture media using the pipette controller with a sterile serological pipette (50 mL). | |
| Label the sampling tube with the following information: *strain name*, Starter Culture | |
| Fill the production flask with 80 mL culture media using the pipette controller and a sterile serological pipette (50 mL). | |

| Thaw Cells | Done (√) |
|---|---|
| Remove 2 cryogenic vials from -80°C freezer, wipe all sides thoroughly with ethanol, place them into the anaerobic chamber via the airlock and thaw the vials for 5-10 minutes. | |

| Storage Location | Freezer | Shelf | Rack | Box |
|---|---|---|---|---|

| Inoculate Starter Culture | | Done (√) |
|---|---|---|
| Sterilize the rubber stopper of the cryogenic vials with ethanol.<br>*Note: Allow for 5 minutes contact time!* | | |
| Vortex cryogenic vials briefly and transfer 0.8 mL of each cryogenic vial into the starter culture, using a sterile syringe (1 mL).<br>*Note: Two cryogenic vials are used to inoculate one 20 mL starter culture.* | Date/Time: | |
| Incubate the starter culture at 37°C for X hours, where X can be estimated from the attached document *Reference Cultivations for MTC01*. | Expected Cultivation Time: | |

| CFU Count – Cryogenic Vials |
|---|
| Print and attach *MTC-BR-0003_CFU Count for Live Biotherapeutic Products (LBP)* to this batch record and perform CFU spot plating for the cryogenic vials accordingly. |

| Biotyping – Cryogenic Vials |
|---|
| Print and attach *MTC-BR-0004_Biotyping for Live Biotherapeutic Products (LBP)* to this batch record and perform biotyping of 24 unique colonies accordingly. |

| Performed by (Init/Date) | Comments: |
|---|---|

#### 7. Cultivation – Production Culture

| If hydrogen level <1.7% and/or oxygen level >50 ppm, exchange chamber air with new gas mix. | **Hydrogen level [%]** | **Oxygen level [ppm]** |
|---|---|---|

| **Measure $OD_{600}$ - Starter Culture** | **Done (√)** |
|---|---|
| Reset the memory of the cell density meter by pressing the 'reset button' twice.<br>*Note: Make sure MEM is displayed, if not press 'memory button'.* | |
| Fill a UV cuvette with 750 µL PBS and 250 µL culture medium.<br>*Note: This is the blank sample.* | |
| Measure blank sample in cell density meter (press 'blank button').<br>*Note: Repeat anytime if blank value gets deleted from cell density meter.* | |
| For every $OD_{600}$ measurement:<br>• Fill a UV cuvette with 750 µL sterile PBS<br>• Vortex the starter culture briefly<br>• Transfer 250 µL starter culture to the UV cuvette with repeater pipette and a sterile Combitip (10 mL)<br>• Mix with micropipette (1000 µL) and place UV cuvette in cell density meter<br>• Press 'sample button' and note value | |

| **Date/Time** | **$OD_{600}$ Starter Culture** | **Comment** |
|---|---|---|

Calculate the volume of inoculation ($V_{inoculation}$) with the following formula:

$$V_{inoculation} = (0.2 \div OD_{600}) \times 100$$

*Note: Before inoculation inform supervisor (Lukas Bethlehem or Ilaria Mogno or Jeremiah Faith). If $V_{inoculation}$ is >40 mL, after 96 hours incubation, terminate production. If $V_{inoculation}$ >20 mL, but <40 mL, add the full volume of the starter culture (20 mL) to the production culture. The target $OD_{600}$ for production culture is 0.1 - 0.2.*

| **$V_{inoculation}$ =** |
|---|

| **Inoculate Production Culture** | **Date/Time** | **Done (√)** |
|---|---|---|
| Vortex the starter culture tube briefly and transfer the inoculation volume into the production flask with a sterile serological pipette (10 mL). | | |
| Incubate the production culture at 37 °C in the anaerobic chamber and measure the $OD_{600}$ at least every three hours (except overnight), beginning with inoculation. | | |

| Performed by (Init/Date) | Comments: |
|---|---|

| Measure $OD_{600}$ - Production Culture | Done (√) |
|---|---|
| Prepare a waste disposal tube (sampling tube 50 mL) and label it: Waste | |
| For every $OD_{600}$ measurement:<br>• Fill a UV cuvette with 750 µL sterile PBS<br>• Swirl the production flask briefly<br>• Withdraw 0.3 mL from the unidirectional port with a sterile syringe and <u>discard</u> into waste<br>• disposal tube<br>• Withdraw 0.3 mL from the unidirectional port with a sterile syringe and <u>transfer</u> into a<br>• sampling tube (1.5 mL)<br>• Mix and transfer 250 µL of the bacterial solution from the sampling tube into the prepared<br>• UV cuvette with the micropipette (1000 µL)<br>• Mix with micropipette (1000 µL) and place UV cuvette in cell density meter<br>• Press ‘sample button’ and note value<br>*Note: If blank value lost, measure UV cuvette with 750 µL PBS and 250 µL culture medium (press ‘blank button’).* | |

| Date/Time | $OD_{600}$ | Comment |
|---|---|---|

Stop production culture when harvest time-point is reached. *Note: Harvest time-point is defined as 80% (± 20%) of final $OD_{600}$ as determined in previous cultivations (see attached document: Reference cultivations for MTC01).*

| Target $OD_{600}$ range | Expected Cultivation Time | Actual Cultivation Time |
|---|---|---|

Before ending production-run, inform supervisor (*Lukas Bethlehem* or *Ilaria Mogno* or *Jeremiah Faith*).

| Performed by (Init/Date) | Comments: |
|---|---|

#### 8. Harvest

| Aliquot Production Culture | | Done (√) |
|---|---|---|
| Repeat this step, into individual sterile sampling tubes, until the bacterial culture is depleted:<br>☐ Swirl the production culture briefly and transfer 20 mL of the bacterial culture into a sterile sampling tube (50 mL), using a sterile serological pipette (50 mL).<br>*Note: The 100 mL production culture should fit into 5 sampling tubes. The last tube will probably be less than 20 mL (note the total volume).* | **Total Volume [mL]** | |
| Add the same volume cryogenic medium into the sampling tubes with a sterile serological pipette (50 mL) and close the tubes. | | |
| Attach a sterile Combitip (10 mL) to the repeater pipette and set to 1 mL in 8 steps with speed 5.<br>*Note: Combitip must stay sterile during the entire process. If Combitip touches anything except the bacterial culture and/or the sterile tubes, replace with a sterile Combitip.* | | |
| ☐ Vortex the sampling tube with bacterial culture briefly and open the tube<br>Repeat the following steps until the bacterial culture in the sampling tube is depleted (then proceed with a new sampling tube):<br>• Open 8 Matrix tubes with the Matrix tube decapper<br>• Aliquot 1 mL bacterial culture into each Matrix tube using the repeater pipette with the sterile Combitip<br>• Close the 8 Matrix tubes with the Matrix tube decapper<br>*Note: If possible fill all vials in the two Matrix tube racks (192 tubes), else fill as much as possible and remove empty tubes. Do not fill tubes with <1 mL.* | **Total Number of Tubes** | |
| Keep one Matrix tube with bacterial culture in the anaerobic chamber for CFU count, then remove both cryogenic storage boxes from the anaerobic chamber and place into -80 °C freezer, immediately. | | |

| | Freezer | Shelf | Rack | Box |
|---|---|---|---|---|
| **Cell Bank Box Storage Location** | | | | |
| | **Freezer** | **Shelf** | **Rack** | **Box** |
| **Cell Bank Box Storage Location** | | | | |

| CFU Count – Harvest |
|---|
| Perform CFU spot plating for the harvest according to the attached batch record *MTC-MBR-0003_CFU Count for Live Biotherapeutic Products (LBP)*. |

| Performed by (Init/Date) | Comments: |
|---|---|

| Spread Plating for Biotyping - Harvest |
|---|
| Label three chocolate agar plates with:<br>*Strain Name*, Harvest-Biotyping, *Dilution* ($10^{-5}$, $10^{-6}$, $10^{-7}$) and *Date*. |
| Transfer 270 µL sterile PBS to three sample tubes (1.5 mL) and label with $10^{-5}$, $10^{-6}$, $10^{-7}$. |
| From the dilution series in the 96 well plate (prepared in *MTC-SOP-0003_CFU Count for Live Biotherapeutic Products (LBP)*), mix and transfer 30 µL bacterial suspension from well E5 to sample tube $10^{-5}$, from well F5 to sample tube $10^{-6}$ and from well G5 to sample tube $10^{-7}$. |
| Add 5-15 sterile glass beads per chocolate agar plate, vortex the sample tubes briefly and transfer 250 µL bacterial suspension to the corresponding agar plate. |
| Close the plates and shake vigorously in horizontal plane for 10-15 seconds. |
| Discard the glass beads, place the spread plates in a Ziploc bag and incubate in the anaerobic chamber until single colonies are clearly visible by eye.<br>*Note: Closed Ziploc bags can be transferred into any other anaerobic chamber for incubation, if necessary. Transfer bag as swift as possible to the new chamber to prevent oxygen damage.* |
| Perform biotyping for the harvest according to the attached batch record *MTC-MBR-0004_Biotyping for Live Biotherapeutic Products (LBP)*. |

| USP<61> Testing | |
|---|---|
| Send one Matrix tube for USP<61> testing to FOCUS Laboratories (177 N. Commerce Way, Bethlehem, PA 18017).<br>*Note: Tubes can be shipped in bulk with other strains (note shipping date).* | Shipping Date |
| Attach the results from USP<61> testing to this batch record.<br>*Note: If USP<61> testing fails, destroy all cryogenic vials. Specificity: TAMC ($10^3$ CFU/mL) TYMC ($10^2$ CFU/mL); while up to 200% specificity acceptable.* | |

| Performed by (Init/Date) | Comments: |
|---|---|

#### 9. POST-PRODUCTION REVIEW

| End-of-Production Cleaning | Done (√) |
|---|---|
| When the current production run is completely finished, discard all remaining flasks and tubes used for cultivation and any leftover medium and PBS. | |
| Remove all reagents and supplies used during production. | |
| The only equipment remaining in the anaerobic chamber is listed below:<br>☐ Vortex mixer<br>☐ Cell density meter<br>☐ Micropipettes (10 μL, 100 μL, 1000 μL)<br>☐ Waste pouch holder<br>☐ Tweezers<br>☐ Cryogenic Vial Rack<br>☐ Marker<br>☐ Thermometer<br>☐ Scissor<br>*Note: Clean the entire surface of the equipment, including below and standing surface with ethanol (70%).* | |
| Clean all non-single use equipment removed from the chamber with ethanol (70%). | |
| Recharge the electronic equipment. | |

| Name | Signature | Date |
|---|---|---|
| Jeremiah Faith, PhD<br>Faculty Director | | |
| Ilaria Mogno, PhD<br>Quality Assurance Specialist | | |

| Performed by (Init/Date) | Comments: |
|---|---|

| Title: Culture Media for Live Biotherapeutic Products (LBP) | MBR No: MTC-MBR-0002 |
|---|---|
| | Batch No. |
| | Effective Date: 08/06/21<br>Supersedes Date: 07/28/21 |

| Reviewed By: | | Date: |
|---|---|---|
| Jeremiah Faith, PhD<br>Faculty Director | | |
| **Approved By:** | | **Date:** |
| Ilaria Mogno, PhD<br>Quality Assurance Specialist | | |
| Control No: | | |

| |
|---|
| **STUDY ID** |
| **Batch number**<br>***(yyyy-mm-dd-MTC01.Strain#)*** |

#### 1. REFERENCE SOP(s)

- Generation of Master/Working Cell Banks (MCB/WCB) for Live Biotherapeutic Products (LBP), MTC-SOP-0001
- Culture Media for Live Biotherapeutic Products (LBP), MTC-SOP-0002

| Performed by (Init/Date) | Comments: |
|---|---|

#### 2. EQUIPMENT CHECKLIST

| Equipment Type | Serial Number | Post-Production Cleaning | |
|---|---|---|---|
| | | Initials | Date |
| Biological Safety Cabinet | 1309819038 | | |
| Refrigerator | Not applicable | Not applicable | Not applicable |
| Micropipette (1000μL) | | | |
| Magnetic Stirrer | Not applicable | | |
| Balance | B323415058 | | |
| Fine Balance | B324443441 | | |
| pH Meter | B322383205 | | |
| *Other:* | | | |
| *Other:* | | | |

| Performed by (Init/Date) | Comments: |
|---|---|

#### 3. REAGENTS AND SUPPLIES

| Item | Manufacturer | Catalog No. | Lot/Batch No. | Exp. Date |
|---|---|---|---|---|
| Sampling Tubes, 50 mL | | | | |
| Sampling Tubes, 15 mL | | | | |
| Pipet Tips, 1000μL | USA Scientific | Not applicable | Not applicable | Not applicable |
| Media Bottle | | | | |
| Filtration Flask | | | | |
| Aluminum Foil | Office Depot | Not applicable | Not applicable | Not applicable |
| Ethanol (190 Proof) | Fisher Scientific | 04-355-225 | | |
| Phosphate Buffered Saline (PBS) | Caisson Labs | PBL06 | | |
| Water for Injection (WFI) | Cytiva | SH30221.17 | | |
| Glycerol (>99.5%) | Fisher Scientific | BP229 | 207205 | 04/01/2026 |
| L-cysteine | Sigma Aldrich | C1276 | BCCC6208 | 12/01/2023 |
| Vegitone Infusion Broth | Sigma Aldrich | 41960 | BCCC3671 | 10/16/2024 |
| Yeast Extract | Fisher Scientific | DF0127179 | 2316459 | 01/31/2026 |
| D-xylose | Sigma Aldrich | X1500 | WXBD2058V | 04/01/2025 |
| D-fructose | Sigma Aldrich | F0127 | SLCD6611 | 12/01/2024 |
| D-glucose | Sigma Aldrich | G8270 | SLCH8955 | 01/01/2027 |
| D-galactose | Sigma Aldrich | G5388 | BCCF2804 | 12/01/2025 |
| N-acetyl-D-glucosamine | Sigma Aldrich | A3286 | SLCB8620 | 02/01/2023 |
| L-arabinose | Sigma Aldrich | W325501 | MKBP1723V | Not applicable |
| D-cellobiose | Sigma Aldrich | C7252 | BCCF4581 | 02/01/2023 |
| D-maltose | Sigma Aldrich | M5885 | SLCC1608 | 02/01/2025 |
| Sucrose | Sigma Aldrich | S0389 | SLCJ6927 | 03/01/2026 |
| L-malic Acid | Sigma Aldrich | M6413 | BCBJ3883V | Not applicable |
| Sodium Sulfate | Sigma Aldrich | S5640 | SLCB8987 | Not applicable |
| Tween 80 | Sigma Aldrich | P1754 | BCCF4367 | 01/01/2026 |
| Menadione | Sigma Aldrich | M5625 | WXBD3522V | 09/01/2022 |
| MOPS | Sigma Aldrich | M1254 | SLCF5165 | 03/01/2022 |
| Sodium Hydroxide | Fisher Scientific | S318 | 200839 | 11/01/2025 |
| *Other:* | | | | |
| *Other:* | | | | |

| Performed by (Init/Date) | Comments: |
|---|---|

#### 4. Preparation

| Cleaning | Done (√) |
|---|---|
| Clean balances before use with ethanol (70%).<br>*Note: <u>Before</u> cleaning turn balances off.* | |

<table>
<tr><th colspan="3">Solutions and Mixes</th><th>Done (√)</th></tr>
<tr><td colspan="3">If not available, prepare monosaccharide and disaccharide mix in sterile sample tubes (50 mL).<br>Note: Mono- and disaccharide mix are stored at room temperature and expire after 6 months.</td><td></td></tr>
<tr><td colspan="2">Monosaccharide Mix available: ☐ yes ☐ no<br>Note: If available, give reference batch number.</td><td>Reference Batch Number<br></td><td></td></tr>
<tr><td colspan="3">Label the sample tube with: Monosaccharide Mix, Date</td><td></td></tr>
<tr><td>Component</td><td colspan="2">Amount [g]</td><td></td></tr>
<tr><td>D-xylose</td><td colspan="2">4</td><td></td></tr>
<tr><td>D-fructose</td><td colspan="2">4</td><td></td></tr>
<tr><td>D-glucose</td><td colspan="2">4</td><td></td></tr>
<tr><td>D-galactose</td><td colspan="2">4</td><td></td></tr>
<tr><td>N-acetylglucosamine</td><td colspan="2">2</td><td></td></tr>
<tr><td>L-arabinose</td><td colspan="2">2</td><td></td></tr>
<tr><td colspan="2">Disaccharide Mix available: ☐ yes ☐ no<br>Note: If available, give reference batch number.</td><td>Reference Batch Number<br></td><td></td></tr>
<tr><td colspan="3">Label the sample tube with: Disaccharide Mix, Date</td><td></td></tr>
<tr><td>Component</td><td colspan="2">Amount [g]</td><td></td></tr>
<tr><td>D-Cellobiose</td><td colspan="2">5</td><td></td></tr>
<tr><td>D-Maltose</td><td colspan="2">5</td><td></td></tr>
<tr><td>Sucrose</td><td colspan="2">5</td><td></td></tr>
</table>

| Performed by (Init/Date) | Comments: |
|---|---|

| Solutions and Mixes (continued) | | Done (√) |
|---|---|---|
| If not available, prepare vitamin-K solution in a sterile sample tube (15 mL). Use the fine balance to weigh menadione.<br>*Note: Vitamin-K solution is stored at 4 °C and expires after 2 weeks.* | | |
| **Vitamin-K Solution** available: ☐ yes ☐ no<br>*Note: If available, give reference batch number.* | **Reference Batch Number**<br> | |
| Wrap the tube with aluminum foil and label it with: Vitamin K, *Date* | | |
| **Component** | **Amount** | |
| Menadione | 5 mg | |
| Ethanol (190 Proof) | 5 mL | |

#### 5. Media Formulation

| Culture Media | | | Done (√) |
|---|---|---|---|
| Prepare culture media up to 72 hours before the start of cultivation.<br>*Note: Culture media expires after one week and should be prepared fresh for every production.* | | | |
| Prepare **200 mL** LYH_VIB in a sterile media bottle (250 mL). | | | |
| Label the media bottle with: LYH_VIB | | | |
| **Component** | **Amount** | | |
| Vegitone infusion broth | 7.4 g | | |
| Yeast extract | 1 g | | |
| Monosaccharide mix (invert 3x) | 0.8 g | | |
| Disaccharide mix (invert 3x) | 0.6 g | | |
| L-cysteine hydrochloride | 0.1 g | | |
| L-malic acid | 0.2 g | | |
| Sodium sulfate | 0.4 g | | |
| MOPS | 4.18 g | | |
| Vitamin-K solution | 0.2 mL | | |
| Tween 80 | 0.1 mL | | |
| WFI water | Adjust to 200 mL | | |
| Adjust the pH to 7.1-7.2 with sodium hydroxide, by adding one pellet after another.<br>*Note: Clean pH electrode with distilled water, before use. Check the accuracy of the pH electrode with prepared buffer (pH7).* | | | |
| **Starting pH** | **Final pH** | **NaOH Pellets Added** | |

| Performed by (Init/Date) | Comments: |
|---|---|

| Cryogenic Media |
|---|
| Label a sterile media bottle (250 mL) with: Cryogenic Media |
| Transfer 56.7 g of glycerol into the media bottle (250 mL), with a sterile serological pipette (50 mL). |
| Add 105 mL sterile PBS and 0.1 g L-cysteine hydrochloride to the glycerol and mix with a magnetic stirrer bar. |

| Filter Sterilization<br>(Perform under BSC) |
|---|
| Fill the culture media into a filtration flask, filter and close the flask aseptically with the included sterile cap. |
| Label the flask with: LYH_VIB, *Date* |
| Fill the cryogenic media into a filtration flask, filter and close the flask aseptically with the included sterile cap. |
| Label the flask with: Cryogenic Media, *Date* |

| Performed by (Init/Date) | Comments: |
|---|---|

#### 6. POST-PRODUCTION REVIEW

| End-of-Production Cleaning | Done (√) |
|---|---|
| Stirrer bars are thoroughly cleaned with distilled water. | |
| All the dedicated ingredients and the stirrer bars are placed back on the dedicated laboratory bench.<br>*Note: Dedicated equipment and ingredients must not be mixed with regular laboratory supplies.* | |
| Clean the balances and the BSC with ethanol (70%).<br>*Note: <u>Before</u> cleaning turn balances off.* | |

| Name | Signature | Date |
|---|---|---|
| Jeremiah Faith, PhD<br>Faculty Director | | |
| Ilaria Mogno, PhD<br>Quality Assurance Specialist | | |

| Performed by (Init/Date) | Comments: |
|---|---|

| | |
|---|---|
| **Title: CFU Count for Live Biotherapeutic Products (LBP)** | **MBR No: MTC-MBR-0003** |
| | **Batch No.** |
| | **Effective Date: 8/27/21**<br>**Supersedes Date: 7/28/21** |

| **Reviewed By:** | | **Date:** |
|---|---|---|
| Jeremiah Faith, PhD<br>Faculty Director | | |
| **Approved By:** | | **Date:** |
| Ilaria Mogno, PhD<br>Quality Assurance Specialist | | |
| Control No: | | |

| **STUDY ID** |
|---|
| **Batch number** ***(yyyy-mm-dd-MTC01.Strain#)*** |

#### 1. REFERENCE SOP(s)

- Generation of Master/Working Cell Banks (MCB/WCB) for Live Biotherapeutic Products (LBP), MTC-SOP-0001
- CFU Count for Live Biotherapeutic Products (LBP), MTC-SOP-0003
- Biotyping for Live Biotherapeutic Products (LBP), MTC-SOP-0004

| Performed by (Init/Date) | Comments: |
|---|---|

#### 2. EQUIPMENT CHECKLIST

| Equipment Type | Serial Number | Post-Production Cleaning | |
|---|---|---|---|
| | | Initials | Date |
| Anaerobic Chamber | AC21-092 | Not applicable | Not applicable |
| Micropipette (10μL) | G42248J (Chamber 1), G42132J (Chamber 2) | | |
| Micropipette (100μL) | J48012J (Chamber 1), J48210J (Chamber 2) | | |
| Micropipette (1000μL) | J45407J (Chamber 1), J45421J (Chamber 2) | | |
| Multichannel Micropipette | SH25393 (Chamber 1), SH35385 (Chamber 2) | | |
| Vortex Mixer | Not specified | Not applicable | Not applicable |
| *Other:* | | | |

#### 3. REAGENTS AND SUPPLIES

| Item | Manufacturer | Catalog No. | Lot/Batch No. | Exp. Date |
|---|---|---|---|---|
| Pipet Tips, 10μL | USA Scientific | Not applicable | Not applicable | Not applicable |
| Pipet Tips, 100μL | USA Scientific | Not applicable | Not applicable | Not applicable |
| Pipet Tips, 1000μL | USA Scientific | Not applicable | Not applicable | Not applicable |
| Multichannel Tips | Thermo Scientific | 94420043 | Not applicable | Not applicable |
| Syringe, 1 mL | | | | |
| Syringe Needle | | | | |
| Ziploc Bag | Office Depot | Not applicable | Not applicable | Not applicable |
| Chocolate Agar Plates | BD Biosciences | 221169 | | |
| Ethanol (70%) | Fisher Scientific | 25-467-01 | | |
| Phosphate Buffer Solution | Caisson Labs | PBL06 | | |
| *Other:* | | | | |

| Performed by (Init/Date) | Comments: |
|---|---|

#### 4. CFU Count – Cryogenic Vial

| **Preparation (In the anaerobic chamber)** | **Done (√)** |
|---|---|
| Label four chocolate agar plates with:<br>*Strain Name, Plate Number (1.1, 1.2, 2.1 or 2.2), Date*<br>*Note: Two plates for each of the two cryogenic vials used for inoculation of the starter culture.* | |
| Transfer 90 µL of sterile PBS into positions B1-G1 and B3-G3 in one sterile 96 well plate.<br>*Note: Leave column 2 empty.* | |

| **Dilution series (In the anaerobic chamber)** | **Done (√)** |
|---|---|
| Sterilize the rubber stopper of the cryogenic vials with ethanol (70%).<br>*Note: Allow for 5 minutes contact time!* | |
| Transfer 10-100 µL of bacterial culture (with a sterile syringe) from the cryogenic vials to position A1 or position A3. | |
| Mix the bacterial culture 3-5 times, starting in position A1, using a micropipette (10 µL) and transfer 10 µL to the next position (B1). Dispose the pipette tip and repeat the process with a new pipette tip, until reaching position G1. | |
| Repeat this process for positions A3 to G3. | |

| **Plating (In the anaerobic chamber)** | | **Done (√)** |
|---|---|---|
| Use the multichannel micropipette (program CFU SPOT PLATE), with 4 multichannel pipette tips, to transfer ten-times 1 µL bacterial solution, from positions D-G (dilutions $10^{-3}$-$10^{-6}$), to the respective chocolate agar plates.<br>*Note: If water in the agar plate lids, knock lid on sterile towelette to remove water. Change tips for each agar plate. Decrease spacing on multichannel pipette to second stopper for plating.* | **Date/Time** | |
| Leave the plates sitting, lid side on top, for 5-10 minutes. | | |
| Incubate the plates at 37°C outside the Ziploc bag until next day (~24 hours)<br>Note: If colonies are not clearly visible at the end of next day (~24 hours), place the plates in a Ziploc bag (leave bag half open) and continue to incubate at 37°C in the anaerobic chamber until colonies are visible. | | |

| Performed by (Init/Date) | Comments: |
|---|---|

| Counting<br>(Outside the anaerobic chamber) | Done (√) |
|---|---|
| Check the colony growth at least every 24 hours and count colonies as soon as they become clearly visible by eye.<br>*Note: Do not let colonies grow too long to prevent inaccurate counts.* | |
| The countable range of colonies is between 10-100 CFU for the total of 10 spots of one dilution. Everything above is considered too numerous to count (TNTC). | |

| Results for Cryogenic vials<br>(Outside the anaerobic chamber) | | | | | |
|---|---|---|---|---|---|
| Calculate the CFU/mL for the lowest dilution in the countable range according to:<br>$CFU/mL = (colonies * 100) * dilution$ | | | | | Date/Time |
| **Vial** | **Cryogenic vial #1** | | | | |
| Dilution | $10^{-3}$ (D1) | $10^{-4}$ (E1) | $10^{-5}$ (F1) | $10^{-6}$ (G1) | Comment |
| Plate 1.1 | | | | | |
| Plate 1.2 | | | | | |
| CFU/mL | | | | | |
| **Vial** | **Cryogenic vial #2** | | | | |
| Dilution | $10^{-3}$ (D3) | $10^{-4}$ (E3) | $10^{-5}$ (F3) | $10^{-6}$ (G3) | Comment |
| Plate 2.1 | | | | | |
| Plate 2.2 | | | | | |
| CFU/mL | | | | | |

| Performed by (Init/Date) | Comments: |
|---|---|

#### 5. CFU Count – Harvest

| Preparation<br>(In the anaerobic chamber) | Done<br>(√) |
| --- | --- |
| Label three chocolate agar plates with:<br>*Strain Name,* Harvest*, Plate Number, Date* | |
| Transfer 90 µL of sterile PBS into positions B5-G5 in the 96 well plate. | |

| Dilution series<br>(In the anaerobic chamber) | Done<br>(√) |
| --- | --- |
| Vortex Matrix tube briefly and transfer 10-100 µL of bacterial solution to position A5 of the 96 well plate. | |
| Mix the bacterial solution 3-5 times, starting in position A5, using a micropipette (10 µL) and transfer 10 µL to the next position (B5). Dispose the pipette tip and repeat the process with a fresh pipette tip, until reaching position G5. | |

| Plating<br>(In the anaerobic chamber) | | Done<br>(√) |
| --- | --- | --- |
| Use the multichannel micropipette (program CFU SPOT PLATE), with 4 multichannel pipette tips, to transfer ten-times 1 µL bacterial solution, from positions D-G (dilutions $10^{-3}$-$10^{-6}$), to the respective chocolate agar plates.<br>*Note: If water in the agar plate lids, knock lid on sterile paper to remove water. Change tips for each agar plate. Decrease spacing on multichannel pipette to second stopper for plating.* | Date/Time | |
| Leave the plates sitting, lid side on top, for 5-10 minutes. | | |
| Place the plates in a Ziploc bag, close tightly and transfer the bag into the QC-anaerobic chamber for incubation at 37°C until colonies are visible (24-72 hours).<br>*Note: Perform transfer as swiftly as possible to prevent cell damage by oxygen.* | | |

| Performed by (Init/Date) | Comments: |
| --- | --- |

| **Counting (Outside the anaerobic chamber)** | **Done (√)** |
|---|---|
| Check the colony growth at least every 24 hours and count colonies as soon as they become clearly visible by eye.<br>*Note: Do not let colonies grow too long to prevent inaccurate counts.* | |
| The countable range of colonies is between 10-100 CFU for the total of 10 spots of one dilution. Everything above is considered too numerous to count (TNTC). | |

| **Results for Harvest (Outside the anaerobic chamber)** | | | | | |
|---|---|---|---|---|---|
| Calculate the CFU/mL for the lowest dilution in the countable range according to: $CFU/mL = (colonies * 100) * dilution$ | | | | | **Date/Time** |
| **Vial** | **Harvest** | | | | |
| Dilution | $10^{-3}$ (D5) | $10^{-4}$ (E5) | $10^{-5}$ (F5) | $10^{-6}$ (G5) | Comment |
| Plate 1 | | | | | |
| Plate 2 | | | | | |
| Plate 3 | | | | | |
| CFU/mL | | | | | |
| *Note: CFU count is documented to assess potency of the cell bank. If CFU count is >2 logs below target CFU, determined during RCB characterization, terminate production and destroy cryogenic vials. Target CFU can be derived from the document Reference cultivations for MTC01, attached to the batch redords.* | | | | | |
| Target CFU/mL = | | | Cryogenic vials terminated? ☐ yes ☐ no | | |

| Performed by (Init/Date) | Comments: |
|---|---|

#### 6. POST-PRODUCTION REVIEW

| End-of-Production Cleaning | Done (√) |
|---|---|
| Clean all equipment after use with ethanol (70%). | |

| Name | Signature | Date |
|---|---|---|
| Jeremiah Faith, PhD<br>Faculty Director | | |
| Ilaria Mogno, PhD<br>Quality Assurance Specialist | | |

| Performed by (Init/Date) | Comments: |
|---|---|

| Microbiome Translational Center (Icahn School of Medicine at Mount Sinai) | Microbiome Translational Center Master Batch Record |
|---|---|

| | |
|---|---|
| **Title: Biotyping for Live Biotherapeutic Products (LBP)** | **MBR No: MTC-MBR-0004** |
| | **Batch No.** |
| | **Effective Date: 7/28/21**<br>**Supersedes Date: New** |

| **Reviewed By:** | | **Date:** |
|---|---|---|
| Jeremiah Faith, PhD<br>Faculty Director | | |
| **Approved By:** | | **Date:** |
| Ilaria Mogno, PhD<br>Quality Assurance Specialist | | |
| Control No: | | |

| |
|---|
| **STUDY ID** |
| **Batch number *(yyyy-mm-dd-MTC01.Strain#)*** |

#### 1. REFERENCE SOP(s)

- Generation of Master/Working Cell Banks (MCB/WCB) for Live Biotherapeutic Products (LBP), MTC-SOP-0001
- CFU Count for Live Biotherapeutic Products (LBP), MTC-SOP-0003
- Biotyping for Live Biotherapeutic Products (LBP), MTC-SOP-0004

| Performed by (Init/Date) | Comments: |
|---|---|

#### 2. EQUIPMENT CHECKLIST

| Equipment Type | Serial Number | Post-Production Cleaning | |
|---|---|---|---|
| | | Initials | Date |
| Fume Hood | Not specified | Not applicable | Not applicable |
| Fine Balance | B324443441 | | |
| Micropipette (10µL) | Not specified | | |
| Micropipette (100µL) | Not specified | | |
| Micropipette (1000µL) | Not specified | | |
| Multichannel Micropipette | Not specified | | |
| MALDI Biotyper | 8269944.00964 | Not applicable | Not applicable |
| Target Plate | 1011022274 | | |
| *Other:* | | | |
| *Other:* | | | |

#### 3. REAGENTS AND SUPPLIES

| Item | Manufacturer | Catalog No. | Lot/Batch No. | Exp. Date |
|---|---|---|---|---|
| Sampling Tubes, 15 mL | Thermo Scientific | Not applicable | Not applicable | Not applicable |
| Sampling Tubes, 1.5 mL | Thermo Scientific | Not applicable | Not applicable | Not applicable |
| Pipet Tips, 10µL | USA Scientific | Not applicable | Not applicable | Not applicable |
| Pipet Tips, 100µL | USA Scientific | Not applicable | Not applicable | Not applicable |
| Pipet Tips, 1000µL | USA Scientific | Not applicable | Not applicable | Not applicable |
| Multichannel Tips | Thermo Scientific | 94420043 | Not applicable | Not applicable |
| Aluminum Foil | Office Depot | Not applicable | Not applicable | Not applicable |
| Ethanol (70%) | Fisher Scientific | 25-467-01 | | |
| Formic Acid (>98%) | Honeywell Fluka | 94318 | | |
| Acetonitrile (>99%) | Jade Scientific | HBJ34967 | | |
| Matrix (alpha-Cyano-4-hydroxycinnamic acid) | Bruker Daltonics | 8201344 | | |

| Performed by (Init/Date) | Comments: |
|---|---|

| | | |
|---|---|---|
| BTS | Bruker Daltonics | 8255343 |
| Trifluoracetic Acid (TFA) | Jade Scientific | SIGT6508 |
| *Other:* | | |
| *Other:* | | |

| Performed by (Init/Date) | Comments: |
|---|---|

#### 4. Colony Biotyping – Cryogenic Vials (performed outside the anaerobic chambers)

| Preparation | Done (√) |
|---|---|
| If not available, prepare 10 mL formic acid (70%) solution by mixing 3 mL distilled water with 7 mL formic acid (>98%), in a sterile sample tube (15 mL).<br>☐ Label the tube with: Formic Acid (70%), *Date*<br>*Note: You have to work under the fume hood when aliquoting formic acid (>98%). The formic acid solution (70%) can be used for multiple production runs and does not expire.* | |
| If not available, transfer 10 mL acetonitrile (>99%) into a sterile sample tube (15 mL).<br>☐ Label the tube with: Acetonitrile, *Date*<br>*Note: You have to work under the fume hood when aliquoting acetonitrile (>99%). The acetonitrile aliquot can be used for multiple production runs and does not expire.* | |
| Prepare 1 mL matrix solution by mixing 10 mg matrix with 1 mL acetonitrile (>99%) in a sample tube (1.5 mL).<br>☐ Label the tube with: Matrix, *Date*<br>☐ Wrap the sample tube with aluminum foil<br>*Note: Use the fine balance to weigh the matrix. The matrix solution expires after two weeks.* | |

| Colony Biotyping – Cryogenic Vials | | | Done (√) |
|---|---|---|---|
| Spot 1 µL BTS on a target plate and let dry for calibration of MALDI Biotyper.<br>☐ Add 1 µL matrix solution and let dry | | | |
| Pick 24 unique colonies, (for each of the 2 cryogenic vials) from the spot plates prepared for CFU counting of the cryogenic vials, and transfer to a clean spot on a target plate.<br>*Note: Make sure CFU count is completed.*<br>☐ Add 1 µL formic acid solution (70%) per spot and let dry<br>☐ Add 1 µL acetonitrile solution per spot and let dry<br>☐ Add 1 µL matrix solution per spot and let dry | | | |
| Perform calibration with BTS spot first, then strain identification by MALDI Biotyper.<br>*Note: Put the batch number as comment and 'cryogenic vial 1' or '2' for the respective sample IDs.* | | | |
| | Cryogenic vial# 1 | Cryogenic vial# 2 | |
| Number correct identifications | | | |
| Number false identifications | | | |
| Total number of spots | | | |
| *Note: ≥50% of the spectra must be high quality identifications (score >1.8) of the target strain. If <50% of the spectra are high quality identifications (score >1.8), prepare new samples until ≥12 are high-quality identifications. If any high-quality identification (score >1.8) of a non-target strain occurs terminate production.* | | | |

| Performed by (Init/Date) | Comments: |
|---|---|

| |
|---|
| The printout of the MALDI-TOF identification overview table must be attached to this batch record. |

#### 5. Colony Biotyping – Harvest

| Colony Biotyping - Harvest | | Done (√) |
|---|---|---|
| Spot 1 µL BTS on a target plate and let dry for calibration of MALDI Biotyper. ☐ NA<br>☐ Add 1 µL matrix solution and let dry<br>*Note: Calibration has to be performed only once a day.* | | |
| Pick 96 unique colonies from the spread plates prepared in *MTC-SOP-0001_Generation of Master or Working Cell Banks (MCB, WCB) for Live Biotherapeutic Products (LBP)* and transfer to clean spots on a target plate.<br>☐ Add 1 µL formic acid solution (70%) per spot and let dry<br>☐ Add 1 µL acetonitrile solution per spot and let dry<br>☐ Add 1 µL matrix solution per spot and let dry | | |
| Perform calibration with BTS spot first, then strain identification by MALDI Biotyper.<br>*Note: Put the batch number as comment and 'harvest' for the respective sample IDs.* | | |
| | Harvest | |
| Number correct identifications | | |
| Number false identifications | | |
| Total number of spots | | |
| *Note: ≥50% of the spectra must be high quality identifications (score >1.8) of the target strain.*<br>*If <50% of the spectra are high quality identifications (score >1.8), prepare new samples until ≥48 are high-quality identifications.*<br>*If any high-quality identification (score >1.8) of a non-target strain occurs destroy all cryogenic vials.* | | |
| The printout of the MALDI-TOF identification overview table must be attached to this batch record. | | |

| Performed by (Init/Date) | Comments: |
|---|---|

#### 6. POST-PRODUCTION REVIEW

| End-of-Production Cleaning | Done (√) |
|---|---|
| Clean target plates after use (work under the fume hood):<br>☐ Add ethanol (70%) and incubate for 5-10 minutes<br>☐ Wipe ethanol with tissue, dip in distilled water and wipe again<br>☐ Add ethanol (70%), wipe with tissue, dip in water and wipe again<br>☐ Add TFA (80 µL) until plate is covered<br>☐ Wipe TFA with tissue, dip in distilled water and wipe again<br>*Note: Discard everything which was in contact with TFA in the appropriate waste container.* | |
| Clean all equipment with ethanol (70%). | |

| Name | Signature | Date |
|---|---|---|
| Jeremiah Faith, PhD<br>Faculty Director | | |
| Ilaria Mogno, PhD<br>Quality Assurance Specialist | | |

| Performed by (Init/Date) | Comments: |
|---|---|

| Microbiome Translational Center (Icahn School of Medicine at Mount Sinai) | Microbiome Translational Center Master Batch Record |
|---|---|
| **Title: Chamber Sterilization for Live Biotherapeutic Product (LBP) MTC01** | **MBR No: MTC-MBR-0005** |
| | **Batch No.** |
| | **Effective Date: 7/28/21**<br>**Supersedes Date: New** |

| **Reviewed By:** | | **Date:** |
|---|---|---|
| Jeremiah Faith, PhD<br>Faculty Director | | |
| **Approved By:** | | **Date:** |
| Ilaria Mogno, PhD<br>Quality Assurance Specialist | | |
| Control No: | | |

| **STUDY ID** |
|---|
| **Batch number**<br>***(yyyy-mm-dd-MTC01.Strain#)*** |

#### 1. REFERENCE SOP(s)

- Generation of Master/Working Cell Banks (MCB/WCB) for Live Biotherapeutic Products (LBP), MTC-SOP-0001
- Sterilization of Anaerobic Chambers for Live Biotherapeutic Products (LBP), MTC-SOP-0005

| Performed by (Init/Date) | Comments: |
|---|---|

#### 2. EQUIPMENT CHECKLIST

| Anaerobic Chamber for this production run: | | | |
|---|---|---|---|
| **Equipment Type** | **Serial Number** | **Post-Production Cleaning** | |
| | | **Initials** | **Date** |
| Anaerobic Chamber | AC21-092 | | |
| Surface Unit | 102-2-110-SS-0032 | | |
| Dräger X-am 5100 hydrazine gas monitor | | Not applicable | Not applicable |
| *Other:* | | | |
| *Other:* | | | |

#### 3. REAGENTS AND SUPPLIES

| Item | Manufacturer | Catalog No. | Lot/Batch No. | Exp. Date |
|---|---|---|---|---|
| Ziploc Bag | Office Depot | Not applicable | Not applicable | Not applicable |
| Medium Bottle | Corning | Not applicable | Not applicable | Not applicable |
| Iodine Test Paper | La Motte | 2948-BJ | | |
| Distilled Water (Milli-Q) | Millipore-Sigma | Not applicable | Not applicable | Not applicable |
| Hydrogen Peroxide solution (BIT) | TOMI | BIT 400 | | |
| Ethanol (70%) | Fisher Scientific | 25-467-01 | | |
| *Other:*<br>☐ N/A | | | | |
| *Other:*<br>☐ N/A | | | | |
| *Other:*<br>☐ N/A | | | | |

| Performed by (Init/Date) | Comments: |
|---|---|

#### 4. Preparation

| Chamber Preparation | Done (√) |
|---|---|
| Chamber preparation according to *MTC-SOP-0001_Generation of Master/Working Cell Banks (MCB/WCB) for Live Biotherapeutic Products (LBP)* needs to be finished before the chamber sterilization process. | |
| Turn off the catalytic fan and dehumidifier in the anaerobic chamber. | |
| Remove the rubber plug from the sterilization port and open both airlock doors for 2-5 minutes. | |
| Open the Zip-Loc bags and put one stack of chocolate agar plates in to hold them open. | |
| Stick one iodine test paper to the upper left corner and upper right corner of the anaerobic chamber. | |
| Close the outer airlock door (only the inner door stays open). | |

| Surface Unit Preparation | Done (√) |
|---|---|
| Wear protective lab glasses and lab coat. Turn on Dräger X-am 5100 hydrazine gas monitor and attach to your upper body.<br>*Note: Whenever the gas monitor shows a value above 1 ppm for more than 5 minutes turn-off the mister and leave the room until the value is below 1 ppm. Make sure no one can enter the room in the meantime.* | |
| Prepare the tripod stand in front of the chamber. | |
| Plug-in the surface unit and connect the mister to the hydrogen peroxide (HP) solution.<br>*Note: Make sure there is a small air bubble between the remaining water and the HP solution. This step can be skipped for the second chamber if both chambers are sterilized sequentially.* | |
| Set the surface unit to Prime mode, set the liquid flow (digital pad) to 20 and empty the remaining water out of the system and into the sink (2 minutes).<br>*Note: There should be a short halt in the stream when changing from water to HP solution. This step can be skipped for the second chamber if both chambers are sterilized sequentially.* | |
| Place the mister on the tripod stand and insert the nozzle into the sterilization port in the center of the anaerobic chamber.<br>*Note: Make sure the nozzle sits as tight as possible in the sterilization port and points slightly upwards.* | |

| Performed by (Init/Date) | Comments: |
|---|---|

#### 5. Sterilization Process

| Sterilization | Done (√) |
|---|---|
| Set the surface unit to <u>spray mode</u>, the liquid flow (digital pad) to 10 and spray the chamber for 3-4 minutes.<br>*Note: The air pressure should be 13-15 and the liquid flow = 10. If necessary, adjust airflow with yellow knob and liquid flow with digital pad on the surface unit.* | |
| Turn on the catalyst fan and let the iHP dwell for 5 minutes. | |
| After the dwell time, turn off the catalyst fan and spray the chamber again for 3-4 minutes. | |
| Both iodine test papers must change color to dark purple to indicate successful sterilization.<br>*Note: If sterilization not successful, repeat spraying.* | |
| Remove the mister swiftly and plug the rubber stopper back into the sterilization port immediately. | |
| Let the iHP dwell for 10 minutes (keep catalyst fan turned off) and try to make sure all the equipment in the chamber comes in contact with the iHP, by moving it through the mist. | |
| After the dwell time turn on the catalyst fan, remove the front horizontal chamber bar of the chamber, evacuate the iHP air and fill with nitrogen. | |
| Repeat this process once with nitrogen and three times with gas mixture. After filling with gas mixture, wait 5-10 minutes for catalyst reaction (Hydrogen level <2%).<br>*Note: After third gas mix filling hydrogen levels should be between 2-3% and oxygen levels ≤300 ppm, otherwise repeat last step.* | |
| Turn on the dehumidifier on low setting. | |
| After 12-72 hours the oxygen level must be <50 ppm and the hydrogen level must be >2%.<br>*Note: If oxygen level >50 ppm and/or hydrogen level <2 ppm, evacuate chamber and fill with gas mixture until values are in range.* | |

| Performed by (Init/Date) | Comments: |
|---|---|

#### 6. POST-PRODUCTION REVIEW

| End-of-Process Cleaning | Done (√) |
|---|---|
| Disconnect the HP solution from the surface unit and connect a medium bottle with 0.5 liter of distilled water.<br>*Note: Make sure there is a <u>small</u> air bubble between the remaining HP solution and the distilled water.* | |
| Set the surface unit to <u>Prime mode</u>, set the liquid flow (digital pad) to 20 and empty the remaining HP solution out of the system and into the sink (2 minutes).<br>*Note: The system should always have remaining liquid inside.* | |
| Clean the electrodes of the mister with ethanol (70%). | |
| Disconnect the media bottle, unplug the surface unit and store the mister, the cable and the HP solution in the surface unit case. | |
| Recharge the Dräger X-am 5100 hydrazine gas monitor. | |

| Name | Signature | Date |
|---|---|---|
| Jeremiah Faith, PhD<br>Faculty Director | | |
| Ilaria Mogno, PhD<br>Quality Assurance Specialist | | |

| Performed by (Init/Date) | Comments: |
|---|---|

### MTC01

Drug Substance

Master Batch Records

Bethlehem et.al., 2025

| Title: Generation of Drug Substance (DS) for Live Biotherapeutic Products (LBP) | MBR No: MTC-MBR-0006 Rev 1 |
|---|---|
| | Batch No. |
| | Effective Date: 03/17/2022<br>Supersedes Date: 02/02/2022 |

| Reviewed By: | | Date: |
|---|---|---|
| Jeremiah Faith, PhD<br>Faculty Director | | |
| **Approved By:** | | **Date:** |
| Ilaria Mogno, PhD<br>Quality Assurance Specialist | | |
| Control No: | | |

| STUDY ID |
|---|
| **Batch number** ***(yyyy-mm-dd-MTC01.Strain#)*** |

#### 1. REFERENCE SOP(s)

- Generation of Drug Substance (DS) for Live Biotherapeutic Products (LBP), MTC-SOP-0006
- DS Culture Media for Live Biotherapeutic Products (LBP), MTC-SOP-0007
- CFU Count of the Drug Substance (DS) for Live Biotherapeutic Products (LBP), MTC-SOP-0008
- Biotyping of the Drug Substance (DS) for Live Biotherapeutic Products (LBP), MTC-SOP-0009
- Sterilization of Anaerobic Chambers for the Drug Substance (DS) of Live Biotherapeutic Products (LBP, DS), MTC-SOP-0010

#### 2. ADDITIONAL DOCUMENTS NEEDED

- Reference cultivations for MTC01.docx
- MTC01_Strains_Growth_Plots.pptx

| Performed by (Init/Date) | Comments: |
|---|---|

#### 3. EQUIPMENT CHECKLIST

| Anaerobic Chamber for this production run: | | | |
|---|---|---|---|
| **Equipment Type** | **Serial Number** | **Post-Production Cleaning** | |
| | | **Initials** | **Date** |
| Anaerobic Chamber | AC21-092 | | |
| Biological Safety Cabinet | 1309819038 | | |
| Cell Density Meter | 1601 (Chamber 1), 1602 (Chamber 2) | | |
| Freezer -80 °C | | Not applicable | Not applicable |
| Refrigerator | Not applicable | Not applicable | Not applicable |
| Laboratory Oven | G1-010356 | Not applicable | Not applicable |
| Micropipette (10μL) | G42248J (Chamber 1), G42132J (Chamber 2) | | |
| Micropipette (100μL) | J48012J (Chamber 1), J48210J (Chamber 2) | | |
| Micropipette (1000μL) | J45407J (Chamber 1), J45421J (Chamber 2) | | |
| Multichannel Pipette | SH25393 (Chamber 1), SH35385 (Chamber 2) | | |
| Pipetting Controller | B03320691 (Chamber 1), B03320694 (Chamber 2) | | |
| Repeater Pipette | P38164J (Chamber 1), M48855G (Chamber 2) | | |
| Cuvette Rack | Not applicable | | |
| Tube Rack | Not applicable | | |
| Matrix Tube Decapper | | | |
| Tweezer | Not applicable | | |
| Scissor | Not applicable | | |
| Vortex Mixer | 20060874 (Chamber 1), 21011317 (Chamber 2) | | |
| Waste Pouch Holder | Not applicable | | |
| Thermometer | Not applicable | | |
| Balance | 8342297383 (Chamber 1) 8342105042 (Chamber 2) | | |
| Centrifuge Tube Adapter | Not applicable | | |
| Centrifuge | 41508814 | | |
| *Other:* | | | |

| Performed by (Init/Date) | Comments: |
|---|---|

#### 4. REAGENTS AND SUPPLIES

| Item | Manufacturer | Catalog No. | Lot/Batch No. | Exp. Date |
|---|---|---|---|---|
| Sampling Tubes, 15 mL | | | | |
| Sampling Tubes, 50 mL | | | | |
| Sampling Tubes, 1.5 mL | | | | Not applicable |
| UV Cuvettes | | | | Not applicable |
| Serological Pipets, 5 mL | | | | |
| Serological Pipets, 10 mL | | | | |
| Serological Pipets, 50 mL | | | | |
| Starter Flask | | | | |
| Production Flask | | | | |
| Glass Beads | | | | Not applicable |
| Pipet Tips, 10µL | | | | Not applicable |
| Pipet Tips, 100µL | | | | Not applicable |
| Pipet Tips, 1000µL | | | | Not applicable |
| Multichannel Tips | Thermo Scientific | 94420043 | | |
| Combitips Advanced, 10 mL | Eppendorf | 0030089677 | | |
| Combitips Advanced, 0.1 mL | Eppendorf | 0030089618 | | |
| Syringe, 1 mL | | | | |
| Matrix Tubes | Thermo Scientific | | | Not applicable |
| 96 well plates | | | | |
| Biohazard Waste Bag | Bel-Art Prod. | 8945C02 | | Not applicable |
| Centrifuge Tubes | | | | |
| Cryogenic Storage Container | | | | |
| Cryogenic Storage Box | | | Not applicable | Not applicable |
| Sterile Towelettes | | | | Not applicable |
| Ziploc Bag | Office Depot | SJN682253 | Not applicable | Not applicable |
| Chocolate Agar Plates | BD Biosciences | 221169 | | |
| Ethanol (70%) | Fisher Scientific | 25-467-01 | | |
| Phosphate Buffered Saline | | | | |
| *Other:* | | | | |

| Performed by (Init/Date) | Comments: |
|---|---|

#### 5. Preparation

| Initial Preparation (Can be done one to four days before the cells are thawed) | | Done (√) |
|---|---|---|
| Print *Reference cultivations for MTC01* plus the growth curve of the strain in this production run from *MTC01_Strains_Growth_Plots* and attach them to this batch record. | | |
| Generate the Freezerworks entry for the cryogenic storage containers and DS-QC vials.<br>*Note: The barcode on the labels is the samples global unique identifier in Freezerworks.* | | |
| Print and attach 4 labels for the DS cryogenic storage containers, with the following information:<br>Print one extra label and put in box to the left.<br>***Strain Name***<br>**MTC01.*StrainID*** **BARCODE**<br>**DS *1-4***<br>***Date printed*** | Stick extra label here: | |
| Print and attach 1 label for the DS cryogenic storage box, with the following information:<br>Print one extra label and put in box to the left.<br>***Strain Name***<br>**MTC01.*StrainID*** **BARCODE**<br>**DS Box**<br>***Date printed*** | Stick extra label here: | |
| Print and attach 10 labels for the DS-QC vials, with the following information:<br>Print one extra label and put in box to the left.<br>**MTC01.*StrainID* DS-QC** **BARCODE**<br>***Strain Name 1-10***<br>***Date printed*** | Stick extra label here: | |
| Print and attach 1 label for the DS-QC Box, with the following information:<br>**MTC01 DS-QC Box *1 or 2*** **BARCODE**<br>***Date printed***<br>*Note: DS-QC Box needs only to be created once per 8 strains and is stored in the last position of the respective DS freezer rack. Put extra label in box to the left or note Batch Record Number when DS-QC Box was generated.* | Stick extra label here *or* note BR# of DS-QC Box generation: | |
| Regenerate catalyst plates:<br>☐ Set the laboratory oven to 200 °C<br>☐ place catalyst plates in the oven<br>☐ Remove catalyst plates after 1 hour and install plates in anaerobic chamber fan<br>*Note: Wear heat resistant gloves when operating the oven and handling hot catalyst plates.* | | |

| Performed by (Init/Date) | Comments: |
|---|---|

| Initial Preparation (Continued)<br>(Can be done one to four days before the cells are thawed) | Done (√) |
| --- | --- |
| Print and attach *MTC-BR-0007_DS Culture Media for Live Biotherapeutic Products (LBP)* to this batch record and prepare culture media, cryogenic media and wash media accordingly. | |
| Aliquot the following supplies under the biosafety cabinet:<br>☐ Sterile glass beads into a sterile sampling tube (15 mL, until the 10 mL mark)<br>☐ 30 mL sterile PBS into a sterile sampling tube (50 mL) | |
| Place equipment and supplies in the anaerobic chamber:<br>☐ Sterile culture media<br>☐ Sterile cryogenic media<br>☐ Sterile wash media<br>☐ Chocolate agar plates (2 stacks of 10 plates each)<br>☐ Sterile PBS (30 mL)<br>☐ 2 cuvette-racks with each 12 UV cuvettes<br>☐ 1 production flask (1.5-2 L)<br>☐ 3 starter flasks (100-200 mL)<br>☐ 2 sampling tubes (50 mL) in sampling tube rack<br>☐ 1 box of each micropipette tips (10 µL, 100 µL, 1000 µL)<br>☐ Pipette controller (<u>recharged</u>)<br>☐ 7x50 mL, 2x10 mL and 15x5 mL serological pipettes<br>☐ Electronic multichannel pipette (<u>recharged</u>)<br>☐ 1 box multichannel pipette tips (12.5 µL)<br>☐ Electronic repeater pipette (<u>recharged</u>)<br>☐ 4 Biopur Combitips (10 mL) + 2 Biopur Combitips (0.1 mL)<br>☐ Matrix Tube Decapper (<u>recharged</u>)<br>☐ 12 syringes (1 mL)<br>☐ 25 sampling tubes (1-2 mL, open in rack)<br>☐ Sterile glass beads<br>☐ 1 squeeze bottle ethanol (70 %) (half full)<br>☐ 3 Biohazard waste bags<br>☐ 2 Ziploc bags<br>☐ 1 sterile 96 well plate<br>☐ 5 sterile Towelettes<br>☐ 6 sterile centrifuge tubes + adapter<br>☐ 1 labelled DS cryogenic storage box<br>☐ 4 labelled Cryogenic storage containers<br>☐ 10 labelled DS-QC Matrix tubes (in Matrix Tube Rack) | |
| Print and attach *MTC-MBR-0010_Sterilization of Anaerobic Chambers for Drug Substance (DS) of Live Biotherapeutic Products (LBP)* to this batch record and perform the sterilization accordingly, at least 24 hours before the start of cultivation. | |
| Tear open the chocolate agar plate bags, place in Ziploc bag and leave about 1 inch of the Ziploc bag open (to prevent plates collecting water and drying out). | |

| Performed by (Init/Date) | Comments: |
| --- | --- |

#### 6. Cultivation – Starter Culture

| If hydrogen level <2.0% and/or oxygen level >50 ppm, exchange chamber air with new gas mix. | Hydrogen level [%] | Oxygen level [ppm] |
|---|---|---|

| Preparation (In the anaerobic chamber) | | | Done (√) |
|---|---|---|---|
| Count number of colonies on agar plates (<u>without opening</u> the plates) from *Media Sterility Test* (*MTC-MBR-007*), note colony count and put plates <u>back into the incubator</u>. Stop production if any colonies are visible. | TSA: | Chocolate: | |
| Fill the three starter flasks each with 100 mL culture media using a sterile serological pipette. | | | |
| Label the starter flasks with the following information: *strain name*, Starter 1-3 | | | |
| Fill the production flask with 1.1 L culture media by pouring the media.<br>*Note: Use aseptic technique! Close flasks directly after pouring and clean any spills.* | | | |

| Thaw Cells | Done (√) |
|---|---|
| Remove 3 cryogenic vials from -80°C freezer, wipe all sides thoroughly with ethanol, transfer them into the anaerobic chamber via the airlock and thaw the vials for 5-10 minutes.<br>*Note: Remove vials from Freezerworks. Storage locations for cryogenic vials are given in the attached document Reference Cultivations for MTC01.* | |

| | Freezer | Shelf | Rack | Box | Vial Numbers |
|---|---|---|---|---|---|
| Storage Location | | | | | |

| Inoculate Starter Culture | | Done (√) |
|---|---|---|
| Vortex cryogenic vials and pool 5 µL of each cryogenic vial into position A1 of the sterile 96 well plate for *CFU Count-Cryogenic Vials*, using the repeater pipette and a sterile Combitip (0.1 mL). | | |
| Transfer 1 mL bacterial culture from cryogenic vials into each starter culture, using the repeater pipette with a sterile Combitip (10 mL). | Date/Time: | |
| Incubate the starter culture at 37 °C for X hours, where X can be estimated from the attached document *Reference Cultivations for MTC01*. | Expected Cultivation Time: | |

| CFU Count – Cryogenic Vials | Done (√) |
|---|---|
| Print and attach *MTC-BR-0008_CFU Count for Live Biotherapeutic Products (LBP)* to this batch record and perform CFU spot plating for the cryogenic vials accordingly. | |

| Biotyping – Cryogenic Vials | Done (√) |
|---|---|
| Print and attach *MTC-BR-0009_Biotyping for Live Biotherapeutic Products (LBP)* to this batch record and perform biotyping for the cryogenic vials accordingly. | |

| Performed by (Init/Date) | Comments: |
|---|---|

#### 7. Cultivation – Production Culture

| | | |
|---|---|---|
| If hydrogen level <2.0% and/or oxygen level >50 ppm, exchange chamber air with new gas mix. | **Hydrogen level [%]** | **Oxygen level [ppm]** |

| **Measure $OD_{600}$ - Starter Culture** | **Done (√)** |
|---|---|
| Reset the memory of the cell density meter by pressing the 'reset button' twice.<br>*Note: Make sure MEM is displayed, if not press 'memory button'.* | |
| Fill a UV cuvette with 750 µL PBS and 250 µL culture medium.<br>*Note: This is the blank sample.* | |
| Measure blank sample in cell density meter (press '*blank button*').<br>*Note: Repeat anytime if blank value gets deleted from cell density meter.* | |
| Prepare a waste disposal tube (sampling tube 50 mL) and label it: Waste | |
| For every $OD_{600}$ measurement:<br>• Fill a UV cuvette with 750 µL sterile PBS<br>• Swirl the starter culture<br>• Withdraw 0.3 mL from the sampling port with a sterile syringe and discard into waste (if no sampling port available, skip this step)<br>• Withdraw 0.5 mL from the sampling port and transfer into a sampling tube (1-2 mL) (if no sampling port available, aseptically sample with a sterile serological pipette)<br>• Mix and transfer 250 µL of the bacterial solution from the sampling tube into the prepared UV cuvette using a micropipette (1000 µL)<br>• Mix with micropipette (1000 µL) and place UV cuvette in cell density meter<br>• Press '*sample button*' and note value below (multiply value by 4, according to dilution)<br>• Measure the temperature of cultures, note value and adjust temperature via catalytic fan if necessary (Temperature of flasks should be 36-38 °C) | |

| **Date/Time** | **$OD_{600}$ Starter Culture** | | | **Temperature/ Comment** | **Done (√)** |
|---|---|---|---|---|---|
| | **#1** | **#2** | **#3** | | |

Calculate the volume of inoculation ($V_{inoculation}$) from the average $OD_{600}$ of the three starter cultures with the following formula:

$$V_{inoculation} = (0.2 \div AverageOD_{600}) \times 1400$$

*Note: Before inoculation inform supervisor. If $V_{inoculation}$ is >600 mL, after 96 hours incubation, terminate production. If $V_{inoculation}$ >300 mL, but <600 mL, add the full volume of the starter culture (300 mL) to the production culture. The target $OD_{600}$ for production culture is 0.1 - 0.2.*

**$V_{inoculation}$ =**

| Performed by (Init/Date) | Comments: |
|---|---|

| Inoculate Production Culture | | Done (√) |
|---|---|---|
| Swirl the starter flask(s), transfer the inoculation volume into the production flask using a sterile serological pipette (50 mL) and fill the production flask with sterile culture medium to 1.4 L. | Date/Time | |
| Incubate the production culture at 37 °C in the anaerobic chamber and measure the $OD_{600}$ at least every three hours (except overnight), beginning with inoculation. | | |
| **Measure $OD_{600}$ - Production Culture** | | |

For every $OD_{600}$ measurement:

- Fill a UV cuvette with 750 µL sterile PBS
- Swirl the production flask, withdraw 1 mL from the sampling port with a sterile syringe and discard into waste disposal tube (if no sampling port available, skip this step)
- Withdraw 0.5 mL from the sampling port and transfer into a sampling tube (1-2 mL) (if no sampling port available, aseptically sample with a sterile serological pipette)
- Mix and transfer 250 µL of the bacterial solution from the sampling tube into the prepared UV cuvette using a micropipette (1000 µL)
- Mix with micropipette (1000 µL) and place UV cuvette in cell density meter
- Press '*sample button*' and note value below (multiply value by 4, according to dilution)
- Measure the temperature of culture, note value and adjust temperature if necessary

| Date/Time | $OD_{600}$ | Temperature/ Comment |
|---|---|---|

| Stop production culture when harvest time-point is reached. *Note: Harvest time-point is defined as 80% (± 20%) of final $OD_{600}$ as determined in previous cultivations (see attached document: Reference cultivations for MTC01). Before ending production-run inform supervisor.* | Target $OD_{600}$ range | Expected Cultivation Time | Actual Cultivation Time |
|---|---|---|---|

| Performed by (Init/Date) | Comments: |
|---|---|

#### 8. Harvest

| Centrifuge Production Culture | Done (√) |
|---|---|
| Swirl the production culture and transfer 100 µL into a sampling tube (1-2 mL) for *CFU Count Harvest* using the repeater pipette and a sterile Combitip (1 mL). Label tube with Harvest-CFU. | |
| Repeat the following step for 6 sterile centrifuge tubes:<br>• Swirl the production culture and transfer 225 mL of the bacterial culture into a sterile centrifuge tube, using a sterile serological pipette<br>**Total Culture Volume [mL]**: | |
| Place tubes into centrifuge tube adapters (lower and top part) and balance the weight of assembled centrifuge tubes (<u>weight difference must not be >1 g, adjust with sterile culture media using a sterile serological pipette</u>)<br>*Note: If the last tube is less than 225 mL, adjust with sterile culture media to 225 mL.*<br>*Note total bacterial culture volume harvested (without PBS).* | |
| Remove centrifuge tubes from anaerobic chamber and proceed to centrifugation immediately.<br>• Centrifuge cells at 4,000 RCF for 15 minutes<br>• Return cells to the anaerobic chamber immediately and discard the supernatant carefully into the empty production flask<br>• Add 80 mL wash media per centrifuge tube with a sterile serological pipette and mix cells by vortexing until cells are resuspended homogeneously (mix by pipetting with sterile serological pipette if necessary)<br>• Close centrifuge tubes tightly, place into adapters (lower and top part) and balance the weight of assembled centrifuge tubes (<u>weight difference must not be >1 g, adjust with sterile wash media using a sterile serological pipette</u>)<br>• Remove tubes from the anaerobic chamber and proceed to centrifugation immediately<br>• Centrifuge cells at 4,000 RCF for 15 minutes<br>• Return cells to the anaerobic chamber immediately and discard the supernatant carefully into the empty wash media bottle(s)<br>• Add 20 mL cryogenic media per centrifuge tube with a sterile serological pipette<br>• Mix cells by vortexing and by pipetting with a sterile serological pipette (5-10 mL) until cells are <u>homogeneously</u> resuspended (keep volume as low as possible, but <u>if cells do not mix fully</u>, add up to 10 mL cryogenic media per tube in steps of 5 mL)<br>• Pool cells into one centrifuge tube using a sterile serological pipette<br>• Aliquot ten-times 100 µL of <u>homogenized</u> cells into labelled Matrix tubes for quality control (QC-vials), using the repeater pipette and a sterile Combitip (10 mL)<br>• Mix and aliquot equal volumes (± 2 mL) of <u>homogenized</u> cells into the 4 cryogenic storage containers using a sterile serological pipette and close containers tightly | |
| Keep one DS-QC vial in the anaerobic chamber for *CFU Count Harvest*. Transfer the remaining QC vials and DS cryogenic storage containers (in cryogenic storage box) immediately into -80 °C freezer. Note if additional cryogenic media was added and note the volume aliquoted into each cryogenic storage container.<br>**Volume added [mL]**: **Volume aliquoted [mL]**: |  |

| Performed by (Init/Date) | Comments: |
|---|---|

| DS Storage Location | Freezer | Shelf | Rack | Box |
|---|---|---|---|---|
| **DS-QC Storage Location** | **Freezer** | **Shelf** | **Rack** | **Box** |
| *Note: DS-QC Box is only created once per 8 strains and is stored in the last position of the respective DS freezer rack.* | | | | |

| CFU Count – Harvest | Done (√) |
|---|---|
| Perform CFU spot plating for the harvest according to the attached batch record *MTC-BR-0008_CFU Count of the Drug Substance (DS) for Live Biotherapeutic Products (LBP)*. | |

| Spread Plating for Biotyping - Harvest | Done (√) |
|---|---|
| Label three chocolate agar plates with:<br>*Reference Batch Number*, Harvest-Biotyping and *Dilution* ($10^{-6}$, $10^{-7}$ or $10^{-8}$). | |
| Transfer 270 µL sterile PBS to three sample tubes (1.5 mL) and label with $10^{-6}$, $10^{-7}$ or $10^{-8}$. | |
| From the dilution series in the 96 well plate (prepared in *MTC-BR-0008*), mix and transfer 30 µL bacterial suspension from well F3 to sample tube $10^{-6}$, from well G3 to sample tube $10^{-7}$ and from well H3 to sample tube $10^{-8}$. | |
| Add 5-15 sterile glass beads per chocolate agar plate, vortex the sample tubes briefly and transfer 250 µL bacterial suspension to the corresponding agar plates. | |
| Close the plates and shake in horizontal plane for 10-15 seconds. | |
| Discard the glass beads, place the spread plates in a Ziploc bag and incubate in an anaerobic chamber at 37 °C until single colonies are clearly visible.<br>*Note: Closed Ziploc bags can be transferred into any other anaerobic chamber for incubation, if necessary. Transfer bag as swift as possible to the new chamber to prevent oxygen damage.* | |
| Perform biotyping for the harvest according to the attached batch record *MTC-MBR-0009*. | |

| USP<61> Testing | | Done (√) |
|---|---|---|
| Send two DS-QC vials (200 µL total) for USP<61> testing to FOCUS Laboratories (177 N. Commerce Way, Bethlehem, PA 18017).<br>*Note: Tubes can be shipped in bulk with other strains (note shipping date).* | **Shipping Date**<br> | |
| Attach the results from USP<61> testing to this batch record.<br>*Note: If USP<61> testing fails, destroy production. Specificity: TAMC ($10^3$ CFU/mL), TYMC ($10^2$ CFU/mL); while up to 200% specificity acceptable.* | | |

| Performed by (Init/Date) | Comments: |
|---|---|

#### 9. POST-PRODUCTION REVIEW

| End-of-Production Cleaning | Done (√) |
| --- | --- |
| When the current production run is completely finished, discard all remaining flasks and tubes used for cultivation and any leftover medium and PBS. | |
| Remove all reagents and supplies used during production. | |
| The only equipment remaining in the anaerobic chamber is listed below:<br>☐ Vortex mixer<br>☐ Cell density meter<br>☐ Micropipettes (10 µL, 100 µL, 1000 µL)<br>☐ Waste pouch holder<br>☐ Tweezers<br>☐ Marker<br>☐ Thermometer (exchange batteries if necessary)<br>☐ Scissor<br>☐ Balance<br>*Note: Clean the entire surface of the equipment, including below with ethanol (70%).* | |
| Clean all non-single use equipment removed from the chamber with ethanol (70%). | |
| Recharge the electronic equipment. | |

| Name | Signature | Date |
| --- | --- | --- |
| Jeremiah Faith, PhD<br>Faculty Director | | |
| Ilaria Mogno, PhD<br>Quality Assurance Specialist | | |

| Performed by (Init/Date) | Comments: |
| --- | --- |

| **Reviewed By:** | | **Date:** |
|---|---|---|
| Jeremiah Faith, PhD<br>Faculty Director | | |
| **Approved By:** | | **Date:** |
| Ilaria Mogno, PhD<br>Quality Assurance Specialist | | |
| Control No: | | |

| |
|---|
| **STUDY ID** |
| **Batch number**<br>***(yyyy-mm-dd-MTC01.Strain#)*** |

#### 1. REFERENCE SOP(s)

- Generation of Drug Substance (DS) for Live Biotherapeutic Products (LBP), MTC-SOP-0006
- DS Culture Media for Live Biotherapeutic Products (LBP), MTC-SOP-0007

#### 2. EQUIPMENT CHECKLIST

| Equipment Type | Serial Number | Post-Production Cleaning | |
|---|---|---|---|
| | | Initials | Date |
| Biological Safety Cabinet | 1309819038 | | |
| Anaerobic Chamber | | | |
| Aerobic Incubator | | | |
| Refrigerator | Not applicable | Not applicable | Not applicable |
| Micropipette (1000µL) | | | |
| Repeater Pipette | | | |
| Magnetic Stirrer | Not applicable | | |
| Balance | B323415058 | | |
| Fine Balance | B324443441 | | |
| Balance (BSC) | C132329532 | | |
| pH Meter | B322383205 | | |
| *Other:* | | | |
| *Other:* | | | |

#### 3. REAGENTS AND SUPPLIES

| Item | Manufacturer | Catalog No. | Lot/Batch No. | Exp. Date |
|---|---|---|---|---|
| Sampling Tubes, 50 mL | | | | |
| Sampling Tubes, 15 mL | | | | |
| Pipet Tips, 1000µL | | | | Not applicable |
| Media Bottle (1 L) | | | | |
| Media Bottle (250 mL) | | | | |
| Filter Unit (1 L) | | | | |
| Filter Unit (0.5 L) | | | | |
| Filter Unit (0.25 L) | | | | |
| Matrix Tubes | Thermo Scientific | | | Not applicable |
| Combitips Advanced, 10 mL | Eppendorf | 0030089677 | | |
| Aluminum Foil | Office Depot | Not applicable | Not applicable | Not applicable |
| Ziploc Bag | Office Depot | SJN682253 | Not applicable | Not applicable |
| Sterile Spatula | | | | Not applicable |

| Performed by (Init/Date) | Comments: |
|---|---|

| Item | Manufacturer | Catalog No. | Lot/Batch No. | Exp. Date |
|---|---|---|---|---|
| Glass Beads | | | | Not applicable |
| Chocolate Agar Plates | BD Biosciences | 221169 | | |
| Tryptic Soy Agar Plates | | | | |
| Ethanol (190 Proof) | | | | |
| USP Grade PBS | | | | |
| USP Grade Glycerol | | | | |
| L-cysteine | | | | |
| USP Grade L-cysteine | | | | |
| Water for Injection (WFI) | | | | |
| Vegitone Infusion Broth | | | | |
| Yeast Extract | | | | |
| D-fructose | | | | |
| D-xylose | | | | |
| D-glucose | | | | |
| D-galactose | | | | |
| N-acetyl-D-glucosamine | | | | |
| L-arabinose | | | | |
| D-cellobiose | | | | |
| D-maltose | | | | |
| Sucrose | | | | |
| L-malic Acid | | | | |
| Sodium Sulfate | | | | |
| Tween 80 | | | | |
| Menadione | | | | |
| MOPS | | | | |
| USP Grade Sodium Chloride | | | | |
| Sodium Hydroxide | | | | |
| *Other:* | | | | |

| Performed by (Init/Date) | Comments: |
|---|---|

#### 4. Preparation

| Cleaning | Done (√) |
|---|---|
| Clean balances before use with ethanol (70%).<br>*Note: Before cleaning turn balances off.* | |

| Solutions and Mixes | | Done (√) |
|---|---|---|
| If not available, prepare monosaccharide and disaccharide mix in sterile sample tubes (50 mL).<br>*Note: Mono- and disaccharide mix are stored at room temperature and expire after 6 months.* | | |
| **Monosaccharide Mix** available: ☐ yes ☐ no<br>*Note: If available, give reference batch number.* | **Reference Batch Number** | |
| Label the sample tube with: Monosaccharide Mix, *Reference Batch Number* | | |
| **Component** | **Amount [g]** | |
| D-xylose | 6 | |
| D-fructose | 6 | |
| D-glucose | 6 | |
| D-galactose | 6 | |
| N-acetylglucosamine | 3 | |
| L-arabinose | 3 | |
| **Disaccharide Mix** available: ☐ yes ☐ no<br>*Note: If available, give reference batch number.* | **Reference Batch Number** | |
| Label the sample tube with: Disaccharide Mix, *Reference Batch Number* | | |
| **Component** | **Amount [g]** | |
| D-Cellobiose | 10 | |
| D-Maltose | 10 | |
| Sucrose | 10 | |

| Performed by (Init/Date) | Comments: |
|---|---|

| Solutions and Mixes (continued) | | Done (√) |
|---|---|---|
| If not available, prepare vitamin-K solution in a sterile sample tube (15 mL). Use the fine balance to weigh menadione.<br>*Note: Vitamin-K solution is stored at 4 °C and expires after 2 weeks.* | | |
| **Vitamin-K Solution** available: ☐ yes ☐ no<br>*Note: If available, give reference batch number.* | **Reference Batch Number** | |
| Wrap the tube with aluminum foil (to protect from light) and label it with:<br>Vitamin K, *Reference Batch Number* | | |
| **Component** | **Amount** | |
| Menadione | 10 mg | |
| Ethanol (190 Proof) | 10 mL | |

#### 5. Media Formulation

| Culture Media | | | Done (√) |
|---|---|---|---|
| Prepare culture media at least 24 hours before the start of cultivation and transfer immediately into anaerobic chamber (Media must not be stored at ambient air >1 h).<br>*Note: Culture media must be prepared fresh for every production.* | | | |
| Prepare **2 L** LYH_VIB in a sterile media bottle (2 L). | | | |
| Label the media bottle with: LYH_VIB, *Reference Batch Number* | | | |
| **Component** | **Amount** | | |
| Vegitone infusion broth | 74 g | | |
| Yeast extract | 10 g | | |
| Monosaccharide mix (invert 3x) | 8 g | | |
| Disaccharide mix (invert 3x) | 6 g | | |
| L-cysteine hydrochloride | 1 g | | |
| L-malic acid | 2 g | | |
| Sodium sulfate | 4 g | | |
| MOPS | 41.8 g | | |
| Vitamin-K solution | 2 mL | | |
| Tween 80 | 1 mL | | |
| WFI water | Adjust to 2 L | | |
| Adjust the pH to 7.1-7.2 with sodium hydroxide.<br>*Note: Clean pH electrode with distilled water, before use. Check the accuracy of the pH electrode with control buffer (pH7).* | | | |
| **Starting pH** | **Final pH** | **NaOH Pellets Added** | |

| Performed by (Init/Date) | Comments: |
|---|---|

| Cryogenic Media (USP Grade)<br>(Use only USP Grade chemicals for this step, perform under BSC and use aseptic technique) | Done (√) |
|---|---|
| Prepare cryogenic media at least 24 hours before the start of cultivation and transfer immediately into anaerobic chamber (Media must not be stored at ambient air >1 h).<br>*Note: Cryogenic media must be prepared fresh for every production.* | |
| Label a sterile media bottle (250 mL) with: Cryogenic Media, *Reference Batch Number* | |
| Add 0.1 g ***USP Grade*** L-cysteine hydrochloride and 1.8 g ***USP Grade*** NaCl using a sterile spatula. | |
| Transfer 60 mL of ***USP Grade*** glycerol (with a sterile serological pipette) and 140 mL sterile ***USP Grade*** **PBS** into the media bottle (250 mL) and mix by inverting until components are dissolved completely (≥ 10 times). | |

| Wash Media (USP Grade)<br>(Use only USP Grade chemicals for this step, perform under BSC and use aseptic technique) | Done (√) |
|---|---|
| Prepare wash media at least 24 hours before the start of cultivation and transfer immediately into anaerobic chamber (Media must not be stored at ambient air >1 h).<br>*Note: Wash media must be prepared fresh for every production.* | |
| Label two sterile media bottles (250 mL) with: Wash Media, *Reference Batch Number* | |
| Add 0.13 g ***USP Grade*** L-cysteine hydrochloride and 2.25 g ***USP Grade*** NaCl per bottle using a sterile spatula. | |
| Transfer 250 mL of sterile ***USP Grade*** **PBS** into each media bottle and mix by inverting until components are dissolved completely (≥ 10 times). | |

| Filter Sterilization<br>(Perform under BSC and use aseptic technique) | Done (√) |
|---|---|
| Fill the culture media into 2 filter units (1 L), filter using the vacuum pump in the BSC and close the flask aseptically with the included sterile cap. | |
| Label the flask with: LYH_VIB*, Reference Batch Number* | |
| Fill the cryogenic media into a filter unit (250 mL), filter using the vacuum pump in the BSC and close the flask aseptically with the included sterile cap. | |
| Label the flask with: Cryogenic Media, *Reference Batch Number* | |
| Fill the wash media into a filter unit (500 mL), filter using the vacuum pump in the BSC and close the flask aseptically with the included sterile cap. | |
| Label the flask with: Wash Media, *Reference Batch Number* | |

| Performed by (Init/Date) | Comments: |
|---|---|

| Media Sterility Test<br>(Perform under BSC and Use aseptic technique) | | Done (√) |
|---|---|---|
| Print and attach 4 labels for the DS-Culture Media-QC vials, DS-Cryo Media-QC vials and DS-Wash Media-QC vials, with the following information:<br>Print one extra label each and put in the box to the left.<br>**MTC01.*StrainID* Culture-QC *or* Wash-QC *or* Cryo-QC**<br>***Strain Name 1-4*** BARCODE<br>***Date printed***<br>*Note: The barcode is the samples global unique identifier in Freezerworks.* | Stick extra label here: | |
| Print and attach 1 label for the DS-Media-QC Box, with the following information:<br>**MTC01 DS-Media-QC Box *1 or 2*** BARCODE<br>***Date printed***<br>*Note: The barcode is the samples global unique identifier in Freezerworks. DS-Media-QC Box needs only to be created once per 8 strains and is stored in the second last position of the DS freezer rack. Put extra label in box to the left or note Batch Record # when DS-Media-QC Box was created.* | Stick extra label here *or* note BR# of DS-Media-QC Box creation: | |
| Label three chocolate agar plates and TSA plates with one of the following information:<br>Culture Media *or* Wash Media *or* Cryo Media, Reference *Batch Number* | | |
| Add 5-15 sterile glass beads per agar plate. | | |
| Transfer four times 500 µL culture media, cryogenic media and wash media into the respective labelled Matrix tubes using a repeater pipette and a sterile Combitip.<br>*Note: Store DS-QC vials in DS-Media-QC Box at -80 °C.* | | |
| Transfer 250 µL culture medial (125 µL from each bottle), cryogenic media or wash media to the respective chocolate and TSA agar plates using a repeater pipette and sterile Combitip. | | |
| Close the plates and shake in horizontal plane for 10-15 seconds. | | |
| Discard the glass beads, place the chocolate plates in a Ziploc bag and incubate in an anaerobic chamber at 37 °C. Incubate the TSA plates in an aerobic incubator at 37 °C. | | |
| Note number of colonies formed per plate after 14 days.<br>*Note: If colonies form, identify via biotyping (attach documents) and repeat sterility test with a respective Media-QC-vial. If contamination can be re-isolated from the second cultivation, eliminate production run.* | **Date/Time** | |

| **Culture Media** | | **Wash Media** | | **Cryogenic Media** | |
|---|---|---|---|---|---|
| **TSA** | **Chocolate** | **TSA** | **Chocolate** | **TSA** | **Chocolate** |

| **DS-Media-QC Storage Location** | **Freezer** | **Shelf** | **Rack** | **Box** |
|---|---|---|---|---|

| Performed by (Init/Date) | Comments: |
|---|---|

#### 6. POST-PRODUCTION REVIEW

| End-of-Production Cleaning | Done (√) |
| --- | --- |
| Stirrer bars are thoroughly cleaned with distilled water. | |
| All dedicated ingredients and stirrer bars are placed back on the dedicated laboratory bench.<br>*Note: Dedicated equipment and ingredients must not be mixed with regular laboratory supplies.* | |
| Clean the balances and the BSC with ethanol (70%).<br>*Note: Before cleaning turn balances off.* | |

| Name | Signature | Date |
| --- | --- | --- |
| Jeremiah Faith, PhD<br>Faculty Director | | |
| Ilaria Mogno, PhD<br>Quality Assurance Specialist | | |

| Performed by (Init/Date) | Comments: |
| --- | --- |

| | |
|---|---|
| **Title: CFU Count of the Drug Substance (DS) for Live Biotherapeutic Products (LBP)** | **MBR No: MTC-MBR-0008 Rev 1** |
| | **Batch No.** |
| | **Effective Date: 03/17/2022**<br>**Supersedes Date: 02/02/2022** |

| **Reviewed By:** | | **Date:** |
|---|---|---|
| Jeremiah Faith, PhD<br>Faculty Director | | |
| **Approved By:** | | **Date:** |
| Ilaria Mogno, PhD<br>Quality Assurance Specialist | | |
| Control No: | | |

| |
|---|
| **STUDY ID** |
| **Batch number**<br>***(yyyy-mm-dd-MTC01.Strain#)*** |

#### 1. REFERENCE SOP(s)

- Generation of Drug Substance (DS) for Live Biotherapeutic Products (LBP), MTC-SOP-0006
- CFU Count of the Drug Substance (DS) for Live Biotherapeutic Products (LBP), MTC-SOP-0008
- Biotyping of the Drug Substance (DS) for Live Biotherapeutic Products (LBP), MTC-SOP-0009

| Performed by (Init/Date) | Comments: |
|---|---|

#### 2. EQUIPMENT CHECKLIST

| Equipment Type | Serial Number | Post-Production Cleaning | |
|---|---|---|---|
| | | Initials | Date |
| Anaerobic Chamber | AC21-092 | Not applicable | Not applicable |
| Micropipette (10µL) | G42248J (Chamber 1), G42132J (Chamber 2) | | |
| Micropipette (100µL) | J48012J (Chamber 1), J48210J (Chamber 2) | | |
| Micropipette (1000µL) | J45407J (Chamber 1), J45421J (Chamber 2) | | |
| Multichannel Micropipette | SH25393 (Chamber 1), SH35385 (Chamber 2) | | |
| Vortex Mixer | Not specified | Not applicable | Not applicable |
| *Other:* | | | |

#### 3. REAGENTS AND SUPPLIES

| Item | Manufacturer | Catalog No. | Lot/Batch No. | Exp. Date |
|---|---|---|---|---|
| Pipet Tips, 10µL | | | | Not applicable |
| Pipet Tips, 100µL | | | | Not applicable |
| Pipet Tips, 1000µL | | | | Not applicable |
| Multichannel Tips | Thermo Scientific | 94420043 | | Not applicable |
| Ziploc Bag | Office Depot | Not applicable | Not applicable | Not applicable |
| Chocolate Agar Plates | BD Biosciences | 221169 | | |
| Ethanol (70%) | Fisher Scientific | 25-467-01 | | |
| Phosphate Buffer Solution | | | | |
| *Other:* | | | | |

| Performed by (Init/Date) | Comments: |
|---|---|

#### 4. CFU Count – Cryogenic Vials

| **Preparation (In the anaerobic chamber)** | **Done (√)** |
|---|---|
| Label two chocolate agar plates with:<br>*Reference Batch Number,* Cryogenic Vials, Plate 1 or 2<br>*Note: Two plates as duplicates for the pool of 3 cryogenic vials used for inoculation of the starter culture in MTC-BR-0006.* | |
| Transfer 90 µL of sterile PBS into positions B1-G1 in the sterile 96 well plate. | |

| **Dilution series (In the anaerobic chamber)** | **Done (√)** |
|---|---|
| Mix the bacterial culture 3-5 times, starting in position A1, using a micropipette (10 µL) and transfer 10 µL to the next position (B1). | |
| Dispose the pipette tip and repeat the process from the next well (B1) with a new pipette tip, until reaching position G1. | |

| **Plating (In the anaerobic chamber)** | | **Done (√)** |
|---|---|---|
| Use the multichannel micropipette (program CFU SPOT PLATE), with 4 multichannel pipette tips, to transfer ten-times 1 µL bacterial solution, from positions D-G (dilutions $10^{-3}$-$10^{-6}$), to the respective chocolate agar plates.<br>*Note: Agar plates need to be dry. If there is water in the agar plate lids, knock lid on sterile towelette to remove water. Change tips for each agar plate. Decrease spacing on multichannel pipette to second stopper position for plating.* | **Date/Time**<br><br> | |
| Leave the plates sitting, lid side up, for 5-10 minutes. | | |
| Incubate the plates at 37 °C, lid side down, in a Ziploc bag until colonies are visible.<br>*Note: Leave Ziploc bag half open (to allow for condensation water to leave the bag). Check the colony growth at least every 24 hours and count colonies as soon as they become clearly visible. <u>Do not let colonies grow too long</u> to prevent inaccurate counts.* | | |

| Performed by (Init/Date) | Comments: |
|---|---|

| **Counting (Outside the anaerobic chamber)** | **Done (√)** |
|---|---|
| The countable range of colonies is between 10-100 CFU for the total of 10 spots of one dilution. Everything above is considered too numerous to count (TNTC). | |
| **Results for Cryogenic Vials** | |
| Calculate the CFU/mL for the lowest dilution in the countable range according to: $CFU/mL = (colonies * 100) * dilution$ — **Date/Time:** | |

| **Vial** | **Cryogenic Vials** | | | | |
|---|---|---|---|---|---|
| Dilution | $10^{-3}$ (D1) | $10^{-4}$ (E1) | $10^{-5}$ (F1) | $10^{-6}$ (G1) | Comment |
| Plate 1 | | | | | |
| Plate 2 | | | | | |
| CFU/mL | | | | | |

| Performed by (Init/Date) | Comments: |
|---|---|

#### 5. CFU Count – Harvest

| Preparation<br>(In the anaerobic chamber) | Done (√) |
| --- | --- |
| Label three chocolate agar plates with:<br>*Reference Batch Number,* Harvest-Culture *1-3*<br>Label three additional chocolate agar plates with:<br>*Reference Batch Number,* Harvest-Centrifuged *1-3* | |
| Transfer 90 µL of sterile PBS into positions B3-H3 and B5-H5 in the 96 well plate.<br>*Note: There is <u>one more dilution</u> for the CFU Count – Harvest (B-H, $10^{-1}$-$10^{-7}$), than for the CFU Count -Cryogenic Vials (B-G, $10^{-1}$-$10^{-6}$), because of higher CFU expected.* | |

| Dilution series<br>(In the anaerobic chamber) | Done (√) |
| --- | --- |
| Vortex the sample tube (= Harvest-Culture) and the Matrix tube (= Harvest-Centrifuged; DS-QC vial) and transfer 50 µL of the first one to position A3 and 50 µL of the second one to position A5 of the 96 well plate. | |
| Mix the bacterial solution 3-5 times, starting in position A3, using a micropipette (10 µL) and transfer 10 µL to the next position (B3). | |
| Dispose the pipette tip and repeat the process from the next well (B3) with a new pipette tip, until reaching position H3. | |
| Repeat the same steps for the dilution series of the bacterial culture in position A5. | |

| Plating<br>(In the anaerobic chamber) | | Done (√) |
| --- | --- | --- |
| Use the multichannel micropipette (program CFU SPOT PLATE), with 4 multichannel pipette tips, to transfer ten-times 1 µL bacterial solution, from positions <u>E-H (dilutions $10^{-4}$-$10^{-7}$)</u>, to the respective chocolate agar plates.<br>*Note: Agar plates need to be dry. If there is water in the agar plate lids, knock lid on sterile towelette to remove water. Change tips for each agar plate. Decrease spacing on multichannel pipette to second stopper position for plating.* | **Date/Time** | |
| Leave the plates sitting, lid side up, for 5-10 minutes. | | |
| Place the plates, lid side down, in a Ziploc bag, close tightly and transfer the bag into the QC-anaerobic chamber for incubation at 37 °C until colonies are visible (24-72 hours).<br>*Note: Perform transfer as swiftly as possible to prevent cell damage by oxygen. Check the colony growth at least every 24 hours and count colonies as soon as they become clearly visible. <u>Do not let colonies grow too long</u> to prevent inaccurate counts.* | | |

| Performed by (Init/Date) | Comments: |
| --- | --- |

| Counting (Outside the anaerobic chamber) | | | | | | Done (√) |
|---|---|---|---|---|---|---|
| The countable range of colonies is between 10-100 CFU for the total of 10 spots of one dilution. Everything above is considered too numerous to count (TNTC). | | | | | | |
| **Results for Harvest** | | | | | | |
| Calculate the CFU/mL for the lowest dilution in the countable range according to: $CFU/mL = (colonies * 100) * dilution$ | | | | | Date/Time | |
| **Vial** | **Harvest – Culture** | | | | | |
| Dilution | $10^{-4}$ (D3) | $10^{-5}$ (E3) | $10^{-6}$ (F3) | $10^{-7}$ (G3) | Comment | |
| Plate 1 | | | | | | |
| Plate 2 | | | | | | |
| Plate 3 | | | | | | |
| CFU/mL | | | | | | |
| **Vial** | **Harvest - Centrifuged** | | | | | |
| Dilution | $10^{-4}$ (D3) | $10^{-5}$ (E3) | $10^{-6}$ (F3) | $10^{-7}$ (G3) | Comment | |
| Plate 1 | | | | | | |
| Plate 2 | | | | | | |
| Plate 3 | | | | | | |
| CFU/mL | | | | | | |
| *Note: CFU count is documented to assess potency of the DS. If CFU count of Harvest - Centrifuged is >2 logs below target CFU, determined during MCB characterization, terminate production and destroy product. Target CFU can be derived from the document Reference cultivations for MTC01, attached to the batch records.* | | | | | | |
| Target CFU/mL = | | | Cryogenic vials terminated? ☐ yes ☐ no | | | |

| Performed by (Init/Date) | Comments: |
|---|---|

#### 6. POST-PRODUCTION REVIEW

| End-of-Production Cleaning | Done (√) |
|---|---|
| Clean all equipment after use with ethanol (70%). | |

| Name | Signature | Date |
|---|---|---|
| Jeremiah Faith, PhD<br>Faculty Director | | |
| Ilaria Mogno, PhD<br>Quality Assurance Specialist | | |

### MTC01

Drug Product

Master Batch Records

Bethlehem et.al., 2025

| Reviewed By: | | Date: |
|---|---|---|
| Jeremiah Faith, PhD<br>Faculty Director | | |
| **Approved By:** | | **Date:** |
| Ilaria Mogno, PhD<br>Quality Assurance Specialist | | |
| Control No: | | |

| |
|---|
| **STUDY ID** |
| **Batch number**<br>***(yyyy-mm-dd*-MTC*XX*-DP*)*** |

#### 1. REFERENCE SOP(s)

- Generation of Drug Substance (DP) for Live Biotherapeutic Products (LBP), MTC-SOP-0011
- DP Buffer for Live Biotherapeutic Products (LBP), MTC-SOP-0012
- CFU Count of the Drug Product (DP) for Live Biotherapeutic Products (LBP), MTC-SOP-0013
- Biotyping of the Drug Product (DP) for Live Biotherapeutic Products (LBP), MTC-SOP-0014
- Sterilization of Anaerobic Chambers for the Drug Product (DP) of Live Biotherapeutic Products (LBP), MTC-SOP-0015

#### 2. ADDITIONAL DOCUMENTS NEEDED

- DP Mixing.xlsx

| Performed by (Init/Date) | Comments: |
|---|---|

#### 3. EQUIPMENT CHECKLIST

| Anaerobic Chamber for this production run: | | | |
|---|---|---|---|
| **Equipment Type** | **Serial Number** | **Post-Production Cleaning** | |
| | | **Initials** | **Date** |
| Anaerobic Chamber | AC21-092 | | |
| Biological Safety Cabinet | 1309819038 | | |
| Freezer -80 °C | | Not applicable | Not applicable |
| Laboratory Oven | G1-010356 | Not applicable | Not applicable |
| Micropipette (10μL) | G42248J (Chamber 1), G42132J (Chamber 2) | | |
| Micropipette (100μL) | J48012J (Chamber 1), J48210J (Chamber 2) | | |
| Micropipette (1000μL) | J45407J (Chamber 1), J45421J (Chamber 2) | | |
| Pipetting Controller | B03320691 (Chamber 1), B03320694 (Chamber 2) | | |
| Repeater Pipette | P38164J (Chamber 1), M48855G (Chamber 2) | | |
| Tube Rack | Not applicable | | |
| Matrix Tube Decapper | 050721099 (Chamber 1), 050721117 (Chamber 2) | | |
| Tweezer | Not applicable | | |
| Scissor | Not applicable | | |
| Vortex Mixer | 20060874 (Chamber 1), 21011317 (Chamber 2) | | |
| Laboratory Tray (Water Bath) | Not applicable | | |
| Rack for DS/DP | Not applicable | | |
| Waste Pouch Holder | Not applicable | | |
| *Other:* | | | |

| Performed by (Init/Date) | Comments: |
|---|---|

#### 4. REAGENTS AND SUPPLIES

| Item | Manufacturer | Catalog No. | Lot/Batch No. | Exp. Date |
|---|---|---|---|---|
| Sampling Tubes, 15 mL | | | | |
| Sampling Tubes, 50 mL | | | | |
| Serological Pipets, 5 mL | | | | |
| Serological Pipets, 10 mL | | | | |
| Serological Pipets, 50 mL | | | | |
| Serological Pipets, 100 mL | | | | |
| Sterile Glass Beads | | | | Not applicable |
| Sterile Strainer (100μm) | | | | Not applicable |
| Pipet Tips, 10μL | | | | Not applicable |
| Pipet Tips, 100μL | | | | Not applicable |
| Pipet Tips, 1000μL | | | | Not applicable |
| Combitips Advanced, 10 mL | Eppendorf | 0030089677 | | |
| Matrix Tubes | Thermo Scientific | | | Not applicable |
| 96 well plates | | | | |
| Biohazard Waste Bag | Bel-Art Prod. | 8945C02 | | Not applicable |
| Cryogenic Storage Container | | | | |
| Master Mix Flask | | | | |
| Sterile Towelettes | | | | Not applicable |
| Ziploc Bag | Office Depot | SJN682253 | Not applicable | Not applicable |
| Chocolate Agar Plates | BD Biosciences | 221169 | | |
| Ethanol (70%) | Fisher Scientific | 25-467-01 | | |
| Phosphate Buffered Saline | | | | |
| Water for Injection (WFI) | | | | |
| *Other:* | | | | |

| Performed by (Init/Date) | Comments: |
|---|---|

#### 5. Preparation

| **Labels** | | | **Done (√)** |
|---|---|---|---|
| Print and attach the updated version of the document *DP Mixing* to this batch record and fill out the fields marked with thick dotted lines in this batch record before you start the protocol. | | | |
| Determine the intended number of DP doses in this production run. *Note: Use document DP Mixing or ask supervisor.* | **Dose (CFU)** | **Amount** | |
| **Number of DPs and potency (CFU) for High and Low Dose**<br>*Note: If high and low dose DPs are produced, the volume of 1 high dose DP is spent to produce 10 low dose DPs (e.g. Produce 4 high dose and 10 low dose DPs).* | **High (______)** | | |
| | **Low (______)** | | |
| Generate the Freezerworks entries for the DP cryogenic storage containers.<br>*Note: The barcode on the labels is the samples global unique identifier in Freezerworks.* | | | |
| Print and attach labels for the DP cryogenic storage containers, with the following information:<br>Print one extra label per concentration and put on the back of this page.<br>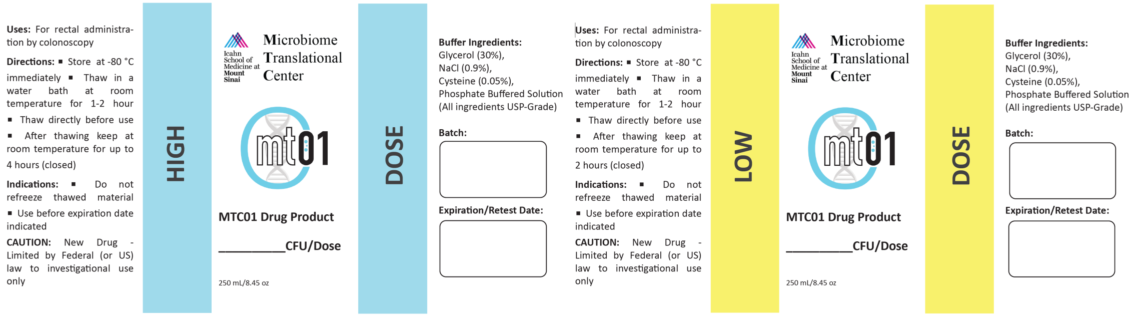 | | | |
| Print and attach 36 labels per batch and concentration for the DP-QC vials, with the following information:<br>Print one extra label and put in box to the left.<br>***Batch Number* DP-QC**<br>**High *or* Low Dose 1-36** | Stick extra label here: | | |
| Print and attach 1 label per DP-QC Box, with the following information:<br>Print one extra label and put in box to the left.<br>**MTC*XX* DP-QC**<br>**Box *Number*** | Stick extra label here: | | |

| Performed by (Init/Date) | Comments: |
|---|---|

| Chamber Preparation | Done (√) |
|---|---|
| Print and attach *MTC-BR-00012_DP Buffer for Live Biotherapeutic Products (LBP)* to this batch record and prepare DP buffer accordingly. | |
| Regenerate catalyst plates:<br>☐ Set the laboratory oven to 200 °C<br>☐ place catalyst plates in the oven<br>☐ Remove catalyst plates after 1 hour and install plates in anaerobic chamber fan<br>*Note: Wear heat resistant gloves when operating the oven and handling hot catalyst plates.* | |
| Aliquot the following supplies under the biosafety cabinet:<br>☐ Sterile glass beads into a sterile sampling tube (15 mL, until the 10 mL mark)<br>☐ 40 mL sterile PBS into a sterile sampling tube (50 mL) | |
| Place equipment and supplies in the anaerobic chamber:<br>☐ Sterile DP buffer<br>☐ Chocolate agar plates (2 stack of 10 plates)<br>☐ Sterile PBS (10 mL)<br>☐ 1 box of each micropipette tips (10 µL, 100 µL, 1000 µL)<br>☐ Pipette controller (<u>recharged</u>)<br>☐ 10x100 mL, 10x50 mL, 10x25 mL, 10x10 mL and 10x5 mL sterile serological pipettes<br>☐ Electronic repeater pipette (<u>recharged</u>)<br>☐ 5 Biopur combitips (10 mL)<br>☐ Matrix tube decapper (<u>recharged</u>)<br>☐ 6 sterile sample tubes (50 mL)<br>☐ 1 sterile Deepwell plate<br>☐ 1 squeeze bottle ethanol (70%)<br>☐ 1 Biohazard waste bags<br>☐ 1 Ziploc bag<br>☐ 5 sterile Towelettes<br>☐ Labelled cryogenic storage containers for the DP (amount determined on page 4)<br>☐ 36 labelled DS-QC Matrix tubes per batch and concentration (in Matrix Tube Rack)<br>☐ Laboratory tray (water bath)<br>☐ 2 L water for injection (WFI)<br>☐ 1 master-mix flask 0.5 L–2 L (Depending on the number of DPs and the total volume of DS)<br>☐ Rack(s) for DS and DP flasks | |
| Print and attach *MTC-MBR-0015_Sterilization of Anaerobic Chambers for Drug Product (DP) of Live Biotherapeutic Products (LBP)* to this batch record and perform the sterilization accordingly, at least 24 hours before the start of formulation. | |
| Tear open the chocolate agar plate bag, place in Ziploc bag and leave about 1 inch of the Ziploc bag open (to prevent plates collecting water and drying out). | |
| Make sure the catalyst fan in the anaerobic chamber is set to 21 °C. | |

| Performed by (Init/Date) | Comments: |
|---|---|

#### 6. DP Formulation

| | | | |
|---|---|---|---|
| If hydrogen ($H_2$) level <2.0% and/or oxygen ($O_2$) level >50 ppm, exchange chamber air with new gas mix | **$H_2$ level [%]** | **$O_2$ level [ppm]** | **Done (√)** |
| Perform **Labelling** steps, described on page 8 and 9, before you proceed thawing the cells. | | | |
| Transfer the DP buffer into the respective DP flasks for high or low dose, using a sterile serological pipette.<br>*Note: Find the volumes in the attached document DP Mixing.* | **DP Buffer Volume [mL]** | | |
| | **High** | **Low** | |
| Fill water bath with 2 L of WFI and place rack for DSs in the water. | | | |
| Retrieve DS cryogenic flasks from -80°C freezer, wipe all sides thoroughly with ethanol, transfer into the anaerobic chamber via the airlock, place into rack (sitting in water) and thaw DS for 15-30 minutes or until completely thawed.<br>*Note: Immediately mix thawed DS and remove spent DS from Freezerworks.* | | | |

| Storage Location | Freezer | Shelf | Rack | Box | DS-BR No. & Flask No. (*yyyy-mm-dd*-MTC01.*StrainID No.*) |
|---|---|---|---|---|---|

| Performed by (Init/Date) | Comments: |
|---|---|

| DP Master Mix | | | | | Done (√) |
|---|---|---|---|---|---|
| Make sure DS is homogeneously mixed before proceeding to the next steps. If DS is not homogeneously mixed, strain it through a sterile 100µm filter.<br>*Note: Indicate straining procedure in table below.* | | | | | |
| Mixing DP Master Mix (Repeat the following steps for each DS Flask):<br>• Remove DS flask from the water bath and mix with a sterile serological pipette 3-times<br>• Transfer the amount given in the table below into the master mix flask<br>• Place the used DS flask outside the water bath<br>*Note: Use aseptic technique and prevent the transfer of water from the water bath into the DS/DP. Volumes are given in the attached document DP Mixing.* | | | | | |
| **Strain ID** | **Strain Name** | **Vol. [mL]** | **Strained** | **Comment** | |
| MTC01.01 | *B. uniformis* | | ☐ | | |
| MTC01.02 | *B. ovatus* | | ☐ | | |
| MTC01.03 | *B. longum* | | ☐ | | |
| MTC01.04 | *B. thetaiotaomicron* | | ☐ | | |
| MTC01.05 | *B. vulgatus* | | ☐ | | |
| MTC01.07 | *P. distasonis* | | ☐ | | |
| MTC01.08 | *B. adolescentis* | | ☐ | | |
| MTC01.09 | *P. merdae* | | ☐ | | |
| MTC01.10 | *C. comes* | | ☐ | | |
| MTC01.11 | *E. rectale* | | ☐ | | |
| MTC01.12 | *B. caccae* | | ☐ | | |
| MTC01.13 | *D. longicatena* | | ☐ | | |
| MTC01.14 | *O. splanchnicus* | | ☐ | | |
| MTC01.15 | *B. cellulosilyticus* | | ☐ | | |
| MTC01.16 | *B. pseudocatenulatum* | | ☐ | | |

| Performed by (Init/Date) | Comments: |
|---|---|

| DP Formulation | | | Done (√) |
|---|---|---|---|
| Determine the volume of Master Mix per DP flask High or Low.<br>*Note: Find volumes in the attached document DP Mixing.* | **Master Mix Volume [mL]** | | |
| | **High** | **Low** | |
| DP aliquoting:<br>• Invert (closed) master mix 10 times before aliquoting<br>• Use a sterile serological pipette to mix (pipetting up and down once) and transfer the volume of the master mix to the respective DP flasks<br>• Close DP flasks and invert 3 times to mix with buffer<br>• Remove DPs immediately and transfer to –80 °C<br>*Note: Master mix must be homogeneously mixed before aliquoting! Close DP flasks tightly and make sure not to break the lid seal.* | | | |
| Determine the volume of Master Mix per DP QC flask High or Low.<br>*Note: Divide the volumes given above by 20.* | **Master Mix Volume [mL]** | | |
| | **High** | **Low** | |
| **Label** two sample tubes (50 mL) with:<br>DP-QC, High *or* Low | | | |
| Transfer the volume from the DP Master Mix for high or low into the respective DP-QC mix tubes, adjust to 12.5 mL with DP buffer and mix well. | | | |
| Aliquot 36 DP-QC vials with 0.25 mL per batch and concentration, using the repeater pipette and a sterile Combitip. | | | |

| | Freezer | Shelf | Batch Number & Flasks |
|---|---|---|---|
| **DP Storage Location** | | | |

| | Freezer | Shelf | Rack | Batch Number & Vials |
|---|---|---|---|---|
| **DP-QC Storage Location** | | | | |

| Performed by (Init/Date) | Comments: |
|---|---|

| Spread Plating for Biotyping | Done (√) |
|---|---|
| **<u>Label</u>** three chocolate agar plates per DP batch and concertation with:<br>*Reference Batch Number*, High *or* Low and *Dilution* ($10^{-5}$-$10^{-7}$ for high DP concentration and $10^{-4}$-$10^{-6}$ for low DP concentration). | |
| Transfer 900 µL sterile PBS to positions A to G of a sterile deep-well plate (A-F for low concentration). | |
| Mix and transfer 100 µL bacterial suspension from a DP-QC vial to position A. Using a new pipette tip, mix 3-5 times and transfer 100 µL to the next position (B) and repeat for the rest of the dilution series. | |
| Add 5-15 sterile glass beads per chocolate agar plate and transfer 250 µL bacterial suspension to the corresponding agar plates. | |
| Close the plates and shake in horizontal plane for 10-15 seconds. | |
| Discard the glass beads, place the spread plates in a Ziploc bag and incubate in an anaerobic chamber at 37 °C for 2-4 days (The different bacterial strains have different optimal incubation times and the plates should be incubated as long as possible without colonies growing into each other).<br>*Note: Closed Ziploc bags can be transferred into any other anaerobic chamber for incubation, if necessary. Transfer bag as swift as possible to the new chamber to prevent oxygen damage.* | |
| Perform biotyping according to the batch record *MTC-MBR-0014_ Biotyping of the Drug Product (DP) for Live Biotherapeutic Products (LBP)*. | |
| Perform CFU count, using the same dilution series, according to the batch record *MTC-MBR-0013_ CFU Count of the Drug Product (DP) for Live Biotherapeutic Products (LBP)*. | |

| USP<61> Testing | | Done (√) |
|---|---|---|
| Send one DP-QC vial (200 µL total) per batch and concentration for USP<61> testing to FOCUS Laboratories (177 N. Commerce Way, Bethlehem, PA 18017). | **Shipping Date**<br><br> | |
| Attach the results from USP<61> testing to this batch record.<br>*Note: If USP<61> testing fails, repeat with two separate QC vials. Destroy DP if all three QC vials fail USP<61> testing*<br>*Specificity: TAMC ($10^3$ CFU/mL), TYMC ($10^2$ CFU/mL); while up to 200% specificity (TAMC $20^3$ CFU/mL, TYMC $20^2$ CFU/mL) acceptable.* | | |

| Performed by (Init/Date) | Comments: |
|---|---|

#### 9. POST-PRODUCTION REVIEW

| End-of-Production Cleaning | Done (√) |
|---|---|
| When the DP formulation is completely finished, discard all remaining flasks and tubes and any leftover solutions and PBS. | |
| Remove all reagents and supplies used during production. | |
| The only equipment remaining in the anaerobic chamber is listed below:<br>☐ Vortex mixer<br>☐ Micropipettes (10 µL, 100 µL, 1000 µL)<br>☐ Waste pouch holder<br>☐ Tweezers<br>☐ Marker<br>☐ Scissor<br>*Note: Clean the entire surface of the equipment, including below with ethanol (70%).* | |
| Clean all non-single use equipment removed from the chamber with ethanol (70%). | |
| Recharge the electronic equipment. | |

| Name | Signature | Date |
|---|---|---|
| Jeremiah Faith, PhD<br>Faculty Director | | |
| Ilaria Mogno, PhD<br>Quality Assurance Specialist | | |

| Performed by (Init/Date) | Comments: |
|---|---|

| **Title: DP Buffer for Live Biotherapeutic Products (LBP)** | **MBR No: MTC-MBR-0012** |
| --- | --- |
| | **Batch No.** |
| | **Effective Date: 8/11/2022**<br>**Supersedes Date: New** |

| **Reviewed By:** | | **Date:** |
| --- | --- | --- |
| Jeremiah Faith, PhD<br>Faculty Director | | |
| **Approved By:** | | **Date:** |
| Ilaria Mogno, PhD<br>Quality Assurance Specialist | | |
| Control No: | | |

| **STUDY ID** |
| --- |
| **Batch number**<br>***(yyyy-mm-dd-MTCXX-DP)*** |

#### 1. REFERENCE SOP(s)

- Generation of Drug Product (DP) for Live Biotherapeutic Products (LBP), MTC-SOP-0011
- DP Buffer for Live Biotherapeutic Products (LBP), MTC-SOP-0012

| Performed by (Init/Date) | Comments: |
| --- | --- |

#### 2. EQUIPMENT CHECKLIST

| Equipment Type | Serial Number | Post-Production Cleaning | |
|---|---|---|---|
| | | Initials | Date |
| Biological Safety Cabinet | 1309819038 | | |
| Pipette Controller | | | |
| Repeater Pipette | | | |
| Balance (BSC) | C132329532 | | |
| *Other:* | | | |
| *Other:* | | | |

#### 3. REAGENTS AND SUPPLIES

| Item | Manufacturer | Catalog No. | Lot/Batch No. | Exp. Date |
|---|---|---|---|---|
| Media Bottle (1 L) | | | | |
| Media Bottle (2 L) | | | | |
| Filter Unit (1 L) | | | | |
| Filter Unit (0.5 L) | | | | |
| Matrix Tubes | Thermo Scientific | | | Not applicable |
| Combitips Advanced, 10 mL | Eppendorf | 0030089677 | | |
| Sterile Spatula | | | | Not applicable |
| Serological Pipettes | | | | |
| USP Grade PBS | | | | |
| USP Grade Glycerol | | | | |
| USP Grade L-cysteine | | | | |
| USP Grade Sodium Chloride | | | | |
| *Other:* | | | | |

| Performed by (Init/Date) | Comments: |
|---|---|

#### 4. Buffer Formulation

| DP Buffer (Perform under BSC and use aseptic technique) | | | | | | | | | Done (√) |
|---|---|---|---|---|---|---|---|---|---|
| Prepare DP buffer at least 24 hours before the start of DP formulation and transfer immediately into anaerobic chamber.<br>*Note: Buffer must not be stored at ambient air >1 h and must be prepared fresh for every production.* | | | | | | | | | |
| Determine the volume of DP buffer needed to generate the intended amount of DP doses in this production run.<br>*Note: Use file Batch Record MTC-MBR-0011_Generation of Drug Product (DP) for Live Biotherapeutic Products (LBP) and the attached document DP Mixing.* | | | | | | | | | |
| **Vol. Buffer DP High** | | **Amount Doses DP High** | | **Vol. Buffer DP Low** | | **Amount Doses DP Low** | | **Vol. Buffer total [mL]** | |
| | x | | + | | x | | = | | |
| Add at least 100 mL to the Vol. Buffer total and round up in steps of 0.5 L.<br>*Note: e.g. if the Vol. Buffer total is 3150 mL, prepare 3500 mL of buffer.* | | | | | | | | | |
| Label the sterile media bottle(s) with: DP Buffer, *Reference Batch Number* | | | | | | | | | |

| Component | Amount per L |
|---|---|
| L-cysteine (USP Grade) | 0.5 g |
| NaCl (USP Grade) * | 9 g |
| Glycerol (USP Grade) | 300 mL |
| Phosphate buffered saline *or* solution * | Adjust to 1 L |
| * NaCl only needs to be added if phosphate buffered <u>solution</u> is used instead of phosphate buffered <u>saline</u>. | |
| Mix components by inverting the flask until homogeneously mixed | |

| Filter Sterilization (Perform under BSC and use aseptic technique) | Done (√) |
|---|---|
| Fill the DP buffer into a filter unit, filter using the vacuum pump in the BSC and close the flask aseptically with the included sterile cap. | |
| Label the flask with: DP-Media, *Reference Batch Number* | |

| Performed by (Init/Date) | Comments: |
|---|---|

| DP Buffer QC Vials (Perform under BSC and Use aseptic technique) | | Done (√) |
|---|---|---|
| Print and attach 3 labels per DP buffer bottle generated, with the following information:<br>Print one extra label each and put in the box to the left.<br>**MTC*XX*.DP Media-QC**<br>**Reference Batch Number** | Stick extra label here: | |
| Print and attach 1 label for the DP-Buffer-QC Box, with the following information:<br>**DP-Media-QC Box *Number***<br>***Date printed*** | Stick extra label here *or* note BR# of DS-Media-QC Box creation: | |
| Aliquot 3 times 0.5 mL per DP buffer bottle into the respective DP-Media-QC vials using the repeater pipette with a sterile Combitip and store at -80 °C. | | |

| | Freezer | Shelf | Rack | Box |
|---|---|---|---|---|
| **DP-Buffer-QC Storage Location** | | | | |

| Performed by (Init/Date) | Comments: |
|---|---|

#### 6. POST-PRODUCTION REVIEW

| End-of-Production Cleaning | Done (√) |
|---|---|
| All dedicated ingredients are placed in the dedicated laboratory area.<br>*Note: Dedicated equipment and ingredients must not be mixed with regular laboratory supplies.* | |
| Clean the BSC with ethanol (70%). | |

| Name | Signature | Date |
|---|---|---|
| Jeremiah Faith, PhD<br>Faculty Director | | |
| Ilaria Mogno, PhD<br>Quality Assurance Specialist | | |

| Performed by (Init/Date) | Comments: |
|---|---|

| Title: CFU Count of the Drug Product (DP) for Live Biotherapeutic Products (LBP) | MBR No: MTC-MBR-0013 |
|---|---|
| | Batch No. |
| | Effective Date: 8/11/2022<br>Supersedes Date: New |

| **Reviewed By:** | | **Date:** |
|---|---|---|
| Jeremiah Faith, PhD<br>Faculty Director | | |
| **Approved By:** | | **Date:** |
| Ilaria Mogno, PhD<br>Quality Assurance Specialist | | |
| Control No: | | |

| |
|---|
| **STUDY ID** |
| **Batch number**<br>***(yyyy-mm-dd*-MTC*XX*-DP*)*** |

#### 1. REFERENCE SOP(s)

- Generation of Drug Product (DP) for Live Biotherapeutic Products (LBP), MTC-SOP-0011
- CFU Count of the Drug Product (DP) for Live Biotherapeutic Products (LBP), MTC-SOP-0013
- Biotyping of the Drug Product (DP) for Live Biotherapeutic Products (LBP), MTC-SOP-0014

| Performed by (Init/Date) | Comments: |
|---|---|

#### 2. EQUIPMENT CHECKLIST

| Equipment Type | Serial Number | Post-Production Cleaning | |
|---|---|---|---|
| | | Initials | Date |
| Anaerobic Chamber | AC21-092 | Not applicable | Not applicable |
| Micropipette (10µL) | G42248J (Chamber 1), G42132J (Chamber 2) | | |
| Micropipette (100µL) | J48012J (Chamber 1), J48210J (Chamber 2) | | |
| Micropipette (1000µL) | J45407J (Chamber 1), J45421J (Chamber 2) | | |
| Multichannel Micropipette | SH25393 (Chamber 1), SH35385 (Chamber 2) | | |
| Vortex Mixer | Not specified | Not applicable | Not applicable |
| *Other:* | | | |

#### 3. REAGENTS AND SUPPLIES

| Item | Manufacturer | Catalog No. | Lot/Batch No. | Exp. Date |
|---|---|---|---|---|
| Pipet Tips, 10µL | | | | Not applicable |
| Pipet Tips, 100µL | | | | Not applicable |
| Pipet Tips, 1000µL | | | | Not applicable |
| Multichannel Tips | Thermo Scientific | 94420043 | | Not applicable |
| Ziploc Bag | Office Depot | Not applicable | Not applicable | Not applicable |
| Chocolate Agar Plates | BD Biosciences | 221169 | | |
| Ethanol (70%) | Fisher Scientific | 25-467-01 | | |
| Phosphate Buffer Solution | | | | |
| *Other:* | | | | |

| Performed by (Init/Date) | Comments: |
|---|---|

#### 4. CFU Count – Drug Product

| Preparation (In the anaerobic chamber) | Done (√) |
|---|---|
| Label three chocolate agar plates per batch and concentration with: *Reference Batch Number,* High *or* Low and CFU-Count 1-3 | |

| Plating (In the anaerobic chamber) | | Done (√) |
|---|---|---|
| Use the multichannel micropipette (program CFU SPOT PLATE), with 4 multichannel pipette tips, to transfer ten-times 1 µL bacterial solution, from positions C-F (dilutions $10^{-3}$-$10^{-6}$ for high DP concentration) or B-E (dilutions $10^{-2}$-$10^{-5}$ for low DP concentration), to the respective chocolate agar plates. *Note: Agar plates need to be dry. If there is water in the agar plate lids, knock lid on sterile towelette to remove water. Change tips for each agar plate.* | **Date/Time** | |
| Leave the plates sitting, lid side up, for 5-10 minutes. | | |
| Place the plates, lid side down, in a Ziploc bag, close tightly and transfer the bag into the QC-anaerobic chamber for incubation at 37 °C until colonies are visible (2-4 days, the different bacterial strains have different optimal incubation times and the plates should be incubated as long as possible without colonies growing into each other). *Note: Perform transfer as swiftly as possible to prevent cell damage by oxygen. Check the colony growth at least every 24 hours.* | | |

| Performed by (Init/Date) | Comments: |
|---|---|

| **Counting (Outside the anaerobic chamber)** | | | | | | **Done (√)** |
|---|---|---|---|---|---|---|
| The countable range of colonies is between 10-100 CFU for the total of 10 spots of one dilution. Everything above is considered too numerous to count (TNTC). | | | | | | |
| **Results for Harvest** | | | | | | |
| Calculate the total CFU for the lowest dilution in the countable range according to: $CFU = (colonies * 25000) \div dilution$ | | | | | **Date/Time** | |
| | **DP High Concentration** | | | | | |
| Dilution | $10^{-3}$ (D3) | $10^{-4}$ (E3) | $10^{-5}$ (F3) | $10^{-6}$ (G3) | Comment | |
| Plate 1 | | | | | | |
| Plate 2 | | | | | | |
| Plate 3 | | | | | | |
| CFU | | | | | | |
| | **DP Low Concentration** | | | | | |
| Dilution | $10^{-2}$ (D3) | $10^{-3}$ (E3) | $10^{-4}$ (F3) | $10^{-5}$ (G3) | Comment | |
| Plate 1 | | | | | | |
| Plate 2 | | | | | | |
| Plate 3 | | | | | | |
| CFU | | | | | | |
| *Note: CFU count is documented to assess potency of the DP. If CFU count of DP is >2 logs below target CFU, repeat CFU count with two separate DP-QC vials. Destroy DP if all three QC vials fail specificity. Target CFU can be derived from the MBR-0011_Generation of Drug Product (DP) for Live Biotherapeutic Products (LBP).* | | | | | | |

| **Target CFU High** | **Target CFU Low** | DP terminated? ☐ yes ☐ no |
|---|---|---|

| Performed by (Init/Date) | Comments: |
|---|---|

#### 6. POST-PRODUCTION REVIEW

| End-of-Production Cleaning | Done (√) |
| --- | --- |
| Clean all equipment after use with ethanol (70%). | |

| Name | Signature | Date |
| --- | --- | --- |
| Jeremiah Faith, PhD<br>Faculty Director | | |
| Ilaria Mogno, PhD<br>Quality Assurance Specialist | | |

| Performed by (Init/Date) | Comments: |
| --- | --- |

| Title: Biotyping of the Drug Product (DP) for Live Biotherapeutic Products (LBP) | MBR No: MTC-MBR-0014 |
|---|---|
| | Batch No. |
| | Effective Date: 8/11/2022<br>Supersedes Date: New |

| **Reviewed By:** | | **Date:** |
|---|---|---|
| Jeremiah Faith, PhD<br>Faculty Director | | |
| **Approved By:** | | **Date:** |
| Ilaria Mogno, PhD<br>Quality Assurance Specialist | | |
| Control No: | | |

| **STUDY ID** |
|---|
| **Batch number**<br>***(yyyy-mm-dd*-MTC*XX*-DP*)*** |

#### 1. REFERENCE SOP(s)

- Generation of Drug Product (DP) for Live Biotherapeutic Products (LBP), MTC-SOP-0011
- CFU Count of the Drug Product (DP) for Live Biotherapeutic Products (LBP), MTC-SOP-0013
- Biotyping of the Drug Product (DP) for Live Biotherapeutic Products (LBP), MTC-SOP-0014

| Performed by (Init/Date) | Comments: |
|---|---|

#### 2. EQUIPMENT CHECKLIST

| Equipment Type | Serial Number | Post-Production Cleaning | |
|---|---|---|---|
| | | Initials | Date |
| Fume Hood | Not applicable | Not applicable | Not applicable |
| Fine Balance | B324443441 | | |
| Micropipette (10µL) | | | |
| Micropipette (100µL) | | | |
| Micropipette (1000µL) | | | |
| Repeater Pipette | | | |
| MALDI Biotyper | 8269944.00964 | Not applicable | Not applicable |
| Target Plates | | | |
| *Other:* | | | |

#### 3. REAGENTS AND SUPPLIES

| Item | Manufacturer | Catalog No. | Lot/Batch No. | Exp. Date |
|---|---|---|---|---|
| Sampling Tubes, 15 mL | | | | |
| Sampling Tubes, 1.5 mL | | | | Not applicable |
| Pipet Tips, 10µL | | | | |
| Pipet Tips, 100µL | | | | |
| Pipet Tips, 1000µL | | | | |
| Combitips Advanced, 0.1 mL | Eppendorf | 0030089618 | | |
| Aluminum Foil | Office Depot | Not applicable | Not applicable | Not applicable |
| Ethanol (70%) | Fisher Scientific | 25-467-01 | | |
| Formic Acid (>98%) | Honeywell Fluka | 94318 | | |
| Acetonitrile (>99%) | Jade Scientific | HBJ34967 | | |
| Mass-spec Solvent | Honeywell Fluka | 191822 | | |
| Matrix (alpha-Cyano-4-hydroxycinnamic acid) | Bruker Daltonics | 8201344 | | |
| Trifluoracetic Acid (TFA) | Jade Scientific | SIGT6508 | | |
| *Other:* | | | | |

| Performed by (Init/Date) | Comments: |
|---|---|

#### 4. Colony Biotyping – Drug Product (performed <u>outside the anaerobic chambers</u>)

| Preparation | Done (√) |
|---|---|
| If not available, prepare 10 mL formic acid (70%) solution by mixing 3 mL distilled water with 7 mL formic acid (>98%), in a sterile sample tube (15 mL).<br>Label the tube with: Formic Acid (70%), *Date*<br>*Note: You have to work under the <u>fume hood</u> when aliquoting formic acid (>98%). The formic acid solution (70%) can be used for multiple production runs and does not expire.* | |
| If not available, transfer 10 mL acetonitrile (>99%) into a sterile sample tube (15 mL).<br>Label the tube with: Acetonitrile, *Date*<br>*Note: You have to work under the <u>fume hood</u> when aliquoting acetonitrile (>99%). The acetonitrile aliquot can be used for multiple production runs and does not expire.* | |
| Prepare 1 mL matrix solution by mixing 10 mg matrix with 1 mL Mass-spec solvent in a sample tube (1.5-2 mL).<br>Label the tube with: Matrix, *Date*<br>Wrap the sample tube with aluminum foil<br>*Note: Use the <u>fine balance</u> to weigh the matrix. The matrix solution expires after two weeks.* | |

| Performed by (Init/Date) | Comments: |
|---|---|

#### 5. Colony Biotyping – Chocolate Plates

| Colony Biotyping - Harvest | | | | Done (√) |
|---|---|---|---|---|
| Pick 288 unique colonies from the chocolate spread plates prepared in *MTC-BR-0011_Generation of Drug Product (DP) for Live Biotherapeutic Products (LBP)* and transfer to clean spots on a target plate.<br>☐ Add 1 µL formic acid solution (70%) per spot and let dry<br>☐ Add 1 µL acetonitrile solution per spot and let dry<br>☐ Add 1 µL matrix solution per spot and let dry | | | | |
| **Strain ID and Name** | **IDs >1.8 confidence score** | | **Comment** | |
| | **High Dose** | **Low Dose** | | |
| **MTC01.01 *B. uniformis*** | | | | |
| **MTC01.02 *B. ovatus*** | | | | |
| **MTC01.03 *B. longum*** | | | | |
| **MTC01.04 *B. thetaiotaomicron*** | | | | |
| **MTC01.05 *B. vulgatus*** | | | | |
| **MTC01.07 *P. distasonis*** | | | | |
| **MTC01.08 *B. adolescentis*** | | | | |
| **MTC01.09 *P. merdae*** | | | | |
| **MTC01.10 *C. comes*** | | | | |
| **MTC01.11 *E. rectale*** | | | | |
| **MTC01.12 *B. caccae*** | | | | |
| **MTC01.13 *D. longicatena*** | | | | |
| **MTC01.14 *O. splanchnicus*** | | | | |
| **MTC01.15 *B. cellulosilyticus*** | | | | |
| **MTC01.16 *B. pseudocatenulatum*** | | | | |
| **Number of correct strain ID's (>1.8)** | | | | |
| **Number of false strain ID's (>1.8)** | | | | |
| **Total number of spots** | | | | |
| *Note: ≥200 of the spots must be high quality identifications (score >1.8) of the target strain. If <200 of the spots are high quality identifications, prepare new spots until ≥200 are high-quality identifications. If any high-quality identification (score >1.8) of a non-target strain occurs repeat biotyping with two separate QC vials. Destroy DP if all three QC vials are contaminated.* | | | | |
| The printout of the MALDI-TOF identification overview table must be attached to this batch record. | | | | |

| Performed by (Init/Date) | Comments: |
|---|---|

#### 7. POST-PRODUCTION REVIEW

| End-of-Production Cleaning | Done (√) |
|---|---|
| Clean target plates after use (work under the fume hood):<br>☐ Add ethanol (70%) and incubate for 5-10 minutes<br>☐ Wipe ethanol with tissue, dip in distilled water and wipe again<br>☐ Add ethanol (70%), wipe with tissue, dip in water and wipe again<br>☐ Add TFA (80 µL) until plate is covered<br>☐ Wipe TFA with tissue, dip in distilled water and wipe again<br>*Note: Discard everything which was in contact with TFA in the appropriate waste container.* | |
| Clean all equipment with ethanol (70%). | |

| Name | Signature | Date |
|---|---|---|
| Jeremiah Faith, PhD<br>Faculty Director | | |
| Ilaria Mogno, PhD<br>Quality Assurance Specialist | | |

| Performed by (Init/Date) | Comments: |
|---|---|

| **Title: Sterilization of Anaerobic Chambers for the Drug Product (DP) of Live Biotherapeutic Products (LBP)** | **MBR No: MTC-MBR-0015** |
|---|---|
| | **Batch No.** |
| | **Effective Date: 8/11/2022**<br>**Supersedes Date: New** |

| **Reviewed By:** | | **Date:** |
|---|---|---|
| Jeremiah Faith, PhD<br>Faculty Director | | |
| **Approved By:** | | **Date:** |
| Ilaria Mogno, PhD<br>Quality Assurance Specialist | | |
| Control No: | | |

| |
|---|
| **STUDY ID** |
| **Batch number**<br>***(yyyy-mm-dd-*MTC*XX*-DP*)*** |

#### 1. REFERENCE SOP(s)

- Generation of Drug Product (DP) for Live Biotherapeutic Products (LBP), MTC-SOP-0011
- Sterilization of Anaerobic Chambers for the Drug Product (DP) of Live Biotherapeutic Products (LBP), MTC-SOP-00015

| Performed by (Init/Date) | Comments: |
|---|---|

#### 2. EQUIPMENT CHECKLIST

| Anaerobic Chamber for this production run: | | | |
|---|---|---|---|
| **Equipment Type** | **Serial Number** | **Post-Production Cleaning** | |
| | | **Initials** | **Date** |
| Anaerobic Chamber | AC21-092 | | |
| Surface Unit | 102-2-110-SS-0032 | | |
| Dräger X-am 5100 hydrazine gas monitor | | Not applicable | Not applicable |
| *Other:* | | | |

#### 3. REAGENTS AND SUPPLIES

| Item | Manufacturer | Catalog No. | Lot/Batch No. | Exp. Date |
|---|---|---|---|---|
| Ziploc Bag | Office Depot | SJN682253 | Not applicable | Not applicable |
| Medium Bottle (Glass) | PYREX | 13951L | Not applicable | Not applicable |
| Iodine Test Paper | La Motte | 2948-BJ | | |
| Distilled Water (Milli-Q) | Millipore-Sigma | Not applicable | Not applicable | Not applicable |
| Hydrogen Peroxide solution (BIT) | TOMI | BIT 400 | | |
| Ethanol (70%) | Fisher Scientific | 25-467-01 | | |
| *Other:* | | | | |

| Performed by (Init/Date) | Comments: |
|---|---|

#### 4. Preparation

| Chamber Preparation | Done (√) |
|---|---|
| Chamber preparation performed according to *MTC-MBR-0011* needs to be finished before the chamber sterilization process. | |
| Turn off the catalytic fan and dehumidifier in the anaerobic chamber. | |
| Remove the rubber plug from the sterilization port and open both airlock doors for at least 2 minutes. | |
| Open the Zip-Loc bags and put one stack of chocolate agar plates in to hold them open. | |
| Stick one iodine test paper to the upper left corner and upper right corner of the anaerobic chamber. | |
| Close the outer airlock door (only the inner door stays open).<br>*Note: If you sterilize two connected chambers at the same time, open only one inner door.* | |

| Surface Unit Preparation | Done (√) |
|---|---|
| Wear protective lab glasses and lab coat. Turn on Dräger X-am 5100 hydrazine gas monitor and attach to your upper body.<br>*Note: Whenever the gas monitor shows a value above 1 ppm for more than 2 minutes turn-off the mister and leave the room until the value is below 1 ppm. Make sure no one can enter the room in the meantime.* | |
| Prepare the tripod stand in front of the chamber. | |
| Plug-in the surface unit and connect the mister to the hydrogen peroxide (HP) solution.<br>*Note: Make sure there is a small air bubble between the remaining water and the HP solution. This step can be skipped for the second chamber if both chambers are sterilized sequentially.* | |
| Set the surface unit to prime mode, set the liquid flow (digital pad) to 20 and empty the remaining water out of the system and into the sink for 2 minutes.<br>*Note: There should be a short halt in the stream when changing from water to HP solution. This step can be skipped for the second chamber if both chambers are sterilized sequentially.* | |
| Place the mister on the tripod stand and insert the nozzle into the sterilization port in the center of the anaerobic chamber.<br>*Note: Make sure the nozzle sits tight in the sterilization port and points slightly upwards. No equipment should be directly in front of the nozzle.* | |

| Performed by (Init/Date) | Comments: |
|---|---|

#### 5. Sterilization Process

| Sterilization | Done (√) |
|---|---|
| Set the surface unit to <u>spray mode</u>, the liquid flow (digital pad) to 10 and spray the chamber for 3-4 minutes.<br>*Note: The air pressure should be 13-15 and the liquid flow = 10. If necessary, adjust airflow with yellow knob and liquid flow with digital pad on the surface unit.* | |
| Turn on the catalyst fan and let the iHP dwell for 4-6 minutes. | |
| After the dwell time, turn off the catalyst fan and spray the chamber again for 3-4 minutes. | |
| Both iodine test papers must change color to dark purple to indicate successful sterilization.<br>*Note: If sterilization not successful, repeat spraying.* | |
| Remove the mister and plug the rubber stopper back into the sterilization port <u>immediately</u>. | |
| Let the iHP dwell for 5-10 minutes (keep catalyst fan turned off) and try to make sure all the equipment in the chamber comes in contact with the iHP, by moving it manually through the iHP mist. | |
| After the dwell time, turn on the catalyst fan (21 °C), remove the front horizontal bar of the chamber, evacuate the iHP air and fill with nitrogen. | |
| Repeat this process once with nitrogen and three times with gas mixture. After filling with gas mixture, wait 5-10 minutes for catalyst reaction (Hydrogen level <2%).<br>*Note: After third gas mix filling hydrogen levels should be between 2-3% and oxygen levels ≤300 ppm, otherwise repeat last step.* | |
| Turn on the dehumidifier on low setting. | |
| After 12-72 hours the oxygen level must be <50 ppm and the hydrogen level must be >2%.<br>*Note: If oxygen level >50 ppm and/or hydrogen level <2 ppm, evacuate chamber and fill with gas mixture until values are in range.* | |

| Performed by (Init/Date) | Comments: |
|---|---|

#### 6. POST-PRODUCTION REVIEW

| End-of-Process Cleaning | Done (√) |
|---|---|
| Disconnect the HP solution from the surface unit and connect a medium bottle with 0.5 liter of distilled water.<br>*Note: Make sure there is a small air bubble between the remaining HP solution and the distilled water.* | |
| Set the surface unit to prime mode, set the liquid flow (digital pad) to 20 and empty the remaining HP solution out of the system and into the sink for 2 minutes.<br>*Note: The system should always have remaining liquid inside.* | |
| Clean the electrodes of the mister with ethanol (70%). | |
| Disconnect the media bottle, unplug the surface unit and store the mister, the cable and the HP solution in the surface unit case. | |
| Recharge the Dräger X-am 5100 hydrazine gas monitor. | |

| Name | Signature | Date |
|---|---|---|
| Jeremiah Faith, PhD<br>Faculty Director | | |
| Ilaria Mogno, PhD<br>Quality Assurance Specialist | | |

| Performed by (Init/Date) | Comments: |
|---|---|

### FMT

Master Batch Records

Bethlehem et.al., 2025

| Title: Generation of CFU normalized Fecal Slurry (FS) for Fecal Microbiota Transplantation (FMT) | MBR No: MTC-MBR-FS-0001 |
|---|---|
| | Batch No. |
| | Effective Date: 08/15/2023<br>Supersedes Date: NEW |

| Reviewed By: | | Date: |
|---|---|---|
| Jeremiah Faith, PhD<br>Faculty Director | | |
| **Approved By:** | | **Date:** |
| Ilaria Mogno, PhD<br>Quality Assurance Specialist | | |
| Control No: | | |

| STUDY ID |
|---|
| **Batch number *(yyyy-mm-dd-FS.DonorID.StoolID)*** |

#### 1. REFERENCE SOP(s)

- Materials Management for Fecal Slurry (FS) for Fecal Microbiota Transplantation (FMT), MTC-SOP-FS-0001
- Generation of CFU normalized Fecal Slurry (FS) for Fecal Microbiota Transplantation (FMT), MTC-SOP-FS-0002
- Fecal Slurry Cryopreservation Media for Fecal Microbiota Transplantation (FMT), MTC-SOP-FS-0003
- Sterilization of Biosafety Cabinet and Stomacher for Fecal Slurry Production for Fecal Microbiota Transplantation (FMT), MTC-SOP-FS-0004
- CFU Count of the Fecal Slurry (FS) for Fecal Microbiota Transplantation (FMT), MTC-SOP-FS-0005

| Performed by (Init/Date) | Comments: |
|---|---|

#### 3. PROCESS OVERVIEW

The goal of this procedure is to generate a homogeneous solution of donor feces normalized to $1x10^{10}$ CFU (low dose) or $1x10^{11}$ CFU (high dose) per 250 ml. To do so the stool sample is divided into smaller portions to maximize the extraction efficiency. The subsamples are weighed and mixed with a PBS based media with glycerol as a cryoprotectant and cysteine to reduce oxygen. The PBS + stool mixtures are homogenized with a stomacher using a filter bag to remove particulate matter from the fecal slurry. The filtered fecal slurries are then pooled in one container. A small aliquot of the concentrated fecal slurry (cFS) is freeze-thawed at -80C and used to measure the potency by CFU/ml. The remainder of the concentrated fecal slurry is frozen at -80C.

Once the CFU/ml of the concentrated fecal slurry is measured, the volume of concentrated fecal slurry per dose is determined. The FMT dose is normalized to a volume of 250 ml with the PBS based media with glycerol as a cryoprotectant and cysteine to reduce oxygen. After production, the CFU/ml of the resulting FMT product is determined and the doses are stored at -80C until use or expiration.

#### 4. EQUIPMENT CHECKLIST

| Equipment Type | Serial Number | Post-Production Cleaning | |
|---|---|---|---|
| | | Initials | Date |
| Stomacher 400 EVO Paddle Blender | EVO - 1078 | | |
| Biological Safety Cabinet | 130475124 A | | |
| Freezer -80 °C | | Not applicable | Not applicable |
| Pipetting Controller | C03324777 | | |
| Single Bag Rack | Not specified | | |
| Multiple Bag Rack | Not specified | | |
| Vial Rack | Not specified | | |
| 8-Channel Decapper | 022415367 | | |
| Balance | C211713971 | | |

| Performed by (Init/Date) | Comments: |
|---|---|

#### 5. REAGENTS AND SUPPLIES

| Item | Manufacturer | Catalog No. | Lot/Batch No. | Exp. Date |
|---|---|---|---|---|
| Plastic Spoons | Nasco | Not specified | | |
| Serological Pipets, 1 mL | | | | |
| Serological Pipets, 50 mL | | | | |
| Serological Pipets, 100 mL | VistaLab | 4090-0100 | | |
| Stomacher Bag | Seward | BA6141/STR | | |
| QC Vial | Thermo Scientific | 3744-BR | | |
| Collection Container 1 L | | | | |
| Cryogenic Storage Container 500 ml | | | | |
| Cryogenic Storage Container 250 ml | | | | |
| Cryogenic Storage Box | Not specified | Not specified | Not applicable | Not applicable |
| Bag Clips | Seward | BA6099 | Not applicable | Not applicable |
| Kimwipes | Kimberly-Clark | Not specified | Not applicable | Not applicable |
| Trash Bag | Not specified | Not specified | Not applicable | Not applicable |
| Ethanol (70%) | Fisher Scientific | A407P-4 | | |
| *Other:* | | | | |

| Performed by (Init/Date) | Comments: |
|---|---|

#### 6. Partitioning of the stool

| **Partitioning of stool samples into stomacher bags (Procedure performed inside the BSC)** | **Done (√)** |
| --- | --- |
| These steps should be done directly when the sample arrives to avoid unnecessary standing of the sample at room temperature. | |
| Print and attach *MTC-MBR-FS-0003_Sterilization of Biosafety Cabinet and Stomacher for Fecal Slurry Production for Fecal Microbiota Transplantation (FMT)* to this batch record and perform the sterilization of the BSC and the equipment listed below accordingly, before the sample arrival.<br>Equipment and supplies to be sterilized:<br>☐ One package of sterile spoons<br>☐ Container with stool sample<br>☐ 6 Stomacher bags<br>☐ 6 bag clips<br>☐ Single bag rack<br>☐ Multi bag rack<br>☐ 1 spray bottle ethanol (70%)<br>☐ Kimwipes<br>☐ Trash bag | |
| Put a sterile stomacher bag in the single bag rack and open the bag and the stool container aseptically. | |
| Use a sterile spoon to transfer 40 – 80 g of the stool into the stomacher bag, close it with a bag clip and put it into the Multi bag rack.<br>*Note: One heaped spoon of stool is 10-15 g.* | |
| Repeat the transfer of the stool into new stomacher bags until the entire sample is partitioned.<br>*Note: A maximum of 6 subsamples can be prepared and processed with this MBR.* | |
| Record the *donor, stool,* and *subsample ID* on the label of the bags containing the stool.<br>*Note: donor ID is e.g. rCD.D283 and stool ID rCD.D283.03 and subsample ID rCD.D283.03.04 if this is the fourth subsample of the third stool sample of the donor 283.* | |
| Put the stomacher bags out of the BSC, weigh the bags and record their weight on the bag labels. | |
| Store the stool subsamples at -80 °C and record location in table below. | |

| **Storage Location** (stool subsamples) | **Freezer** | **Shelf** | **Additional storage information** |
| --- | --- | --- | --- |

| **End-of-Partitioning Cleaning** | **Done (√)** |
| --- | --- |
| When the partitioning of the stool sample is finished, discard all remaining consumables. | |
| Clean all non-single use equipment removed from the chamber with ethanol (70%). | |

| Performed by (Init/Date) | Comments: |
| --- | --- |

#### 7. Preparation for generating concentrated Fecal Slurry

| Initial Preparation for concentrated Fecal Slurry (cFS)<br>(Can be done one to four weeks before the stools are processed) | Done (√) |
| --- | --- |
| Generate the Freezerworks entry for the two cryogenic storage containers (500 ml).<br>*Note: The barcode on the labels is the sample's global unique identifier in Freezerworks.*<br>*If <u>more than 1 L total cFS is produced,</u> add more cryogenic storage containers (e.g. one subsample generates about 300 ml cFS)* | |
| Print 3 labels and attach one label to each cryogenic storage containers (500 ml) and one in the table below in this BR.<br>**MTC.CFS unnormalized** **BARCODE**<br>***Batch date***<br>***DonorID.StoolID***<br>**<u>UNNORMALIZED</u>**<br>**<u>DON'T ADMINISTER</u>** | |
| Print 3 numerated labels for the QC vials and attach the labels to the vials. | |

| Perform the following steps for each subsample taken from the stool sample to be processed: | Done (√) 1 | 2 | 3 | 4 | 5 | 6 |
| --- | --- | --- | --- | --- | --- | --- |
| • Record the identifier of the donor, stool sample and subsample to be processed in the corresponding row/column of the table below. | | | | | | |
| • Record the weight of the stool subsample in the corresponding row/column of the table below.<br>*Note: stomacher bag weight is 9.7 g and clip weight is 18.8 g.* | | | | | | |
| • Calculate the volume of cryogenic media to homogenize the sample (6 ml media/g of stool). To avoid overfilling the stomacher bag for large subsamples the volume should be at most 350 ml <u>including stool weight (i.e., 50 g stool add 300 ml media, 80 g stool add 270 ml media).</u> Record the homogenization volume in the corresponding row/column of the table below. | | | | | | |

| | DonorID StoolID SubsampleID | Stool weight [g] | Homogenization volume [ml] | Extra Label |
| --- | --- | --- | --- | --- |
| 1. Subsample | | | | |
| 2. Subsample | | | | |
| 3. Subsample | | | | |
| 4. Subsample | | | | |
| 5. Subsample | | | | |
| 6. Subsample | | | | |

| Performed by (Init/Date) | Comments: |
| --- | --- |

#### 8. Generation of Concentrated Fecal Slurry (cFS)

| Production of cFS<br>(Procedure performed inside the BSC) | | Done (√) |
|---|---|---|
| Print and attach *MTC-MBR-FS-0002_Fecal Slurry Cryopreservation Media for Fecal Microbiota Transplantation* to this batch record and prepare cryopreservation media accordingly. | | |
| Print and attach *MTC-MBR-FS-0003_Sterilization of Biosafety Cabinet and Stomacher for Fecal Slurry Production for Fecal Microbiota Transplantation (FMT)* to this batch record and perform the sterilization of the Stomacher, BSC and the equipment listed below accordingly, before the start of CFS generation.<br><br>Equipment and supplies to be sterilized:<br>☐ Sterile cryogenic media<br>☐ Pipette controller (recharged)<br>☐ 2 labeled 500 ml cryogenic storage containers<br>☐ 2x100ml serological pipettes<br>☐ 10x50 ml serological pipettes/stool subsample<br>☐ 2x1ml serological pipettes<br>☐ Vial rack with 3 labelled QC vials<br>☐ Multi bag rack with clip closed stomacher bags containing the stool subsamples<br>☐ Single bag rack<br>☐ 8-Channel decapper<br>☐ 1 spray bottle ethanol (70%)<br>☐ Kimwipes<br>☐ Trash bag | | |
| Weigh one empty 1 L collection container (with the lid on) that will be used for the combination of the fecal slurries, sterilize it with ethanol (70%) and put it under the BSC. Record the weight on the right.<br>*Note: the cFS derived from each subsample will be combined in this collection container. After taking aliquots for CFU determination, it will be split among two cryogenic storage containers to allow a shorter thawing time when producing the normalized FS.* | **Pre-weight [g]** | |

| Performed by (Init/Date) | Comments: |
|---|---|

| Step | Done (√) 1 | 2 | 3 | 4 | 5 | 6 |
|---|---|---|---|---|---|---|
| Perform the following steps for all stool subsamples: | | | | | | |
| • Put the bag with the stool subsample in the single bag rack, open it aseptically, add the cryogenic media homogenization volume calculated above (section 6) to the filtering stomacher bag containing the stool using the pipette controller and a sterile serologic pipette (50 mL), and close the bag with a bag clip.<br>*Note: Push as much <u>air out of the stomacher</u> bag as possible!* | | | | | | |
| • Homogenize the feces and the cryogenic media in the stomacher for 60 seconds at 210 rotations per minute. | | | | | | |
| • Spray the surface of the bag with ethanol (70%), place it back in the single rack under the BSC and open it. Using the pipette controller and a sterile serologic pipette (50 mL), transfer the homogenized concentrated fecal slurry from the filtered side of the stomacher bag to the 1 L collection container.<br>*Note: The fecal slurries are pooled in the same collection container before splitting the cFS among the two storage containers.* | | | | | | |

| Step | Post-weight [g] | Done |
|---|---|---|
| Mix the combined concentrated fecal slurries by gently swirling the collection container. | | |
| Open the QC vials with the decapper, collect three 100 µl aliquots of the concentrated slurry for CFU calculation in the QC vials by using the pipette controller and a sterile serological pipette (1 ml), and close the QC vials again with the decapper. | | |
| Weigh the collection container (with the lid on). Record the value on the right. | | |
| Spray the Collection Container with ethanol (70%) and put it back under the BSC. | | |
| Use the pipette controller and a sterile serological pipette (100 ml) to distribute the combined cFS from the collection container evenly between the two labeled cryogenic storage containers. | | |
| Freeze the cryogenic storage containers and the QC vials at -80°C. | | |
| After ≥24 hours, thaw the QC vials at room temperature for 10 minutes, then return to -80 °C for at least 24 hours before performing the CFU count for the cFS.<br>*Note: This is to simulate the freeze thaw cycle for the formulation of the FS FMT Product generation. Mark the QC vial label with a checkmark after performing the freeze thaw cycle.* | | |

| Performed by (Init/Date) | Comments: |
|---|---|

| **Storage Location** (cFS containers) | **Freezer** | **Shelf** | **Additional storage information** | | |
|---|---|---|---|---|---|
| **Storage Location** (cFS QC vials) | **Freezer** | **Shelf** | **Rack** | **Box** | **Vial Numbers** |

| **End-of-Production Cleaning** | **Done (√)** |
|---|---|
| When the production of CFS is finished, discard all remaining consumables and any leftover medium. | |
| Remove all reagents and supplies used during production. | |
| Clean all non-single use equipment removed from the chamber with ethanol (70%). | |

| Performed by (Init/Date) | Comments: |
|---|---|

#### 9. Generation of CFU normalized Fecal Slurry (FS)

| **Initial Preparation for normalized Fecal Slurry (FS) (Takes 1-3 days depending on growth time of strains)** | **Done (√)** |
|---|---|
| Print and attach *MTC-MBR-FS-0004_CFU Count of the Fecal Slurry (FS) for Fecal Microbiota Transplantation (FMT)* to this batch record and perform the CFU/ml determination of the CFS accordingly. | |
| Calculate the following values for the cFS sample to be processed for both high and low dose and record them in the relevant row/column in the table below:<br>*Note: 2-fold higher doses than $1x10^{10}$ and $1x10^{11}$ will be prepared to account for the potential loss of CFU during this final step of production.* | |
| • Note the CFU/ml of cFS calculated with *CFU Count of the Fecal Slurry (FS) for Fecal Microbiota Transplantation (FMT)* (MTC-MBR-FS-0004). Stop production for a sample if the cFS CFU/ml is <$1x10^7$ for three consecutive QC vials. | |
| • cFS total weight = Collection Container post-weight – pre-weight (from above)<br>*Note: fecal slurry is approximately 1 g/ml so g and ml can be used interchangeably.* | |
| • Volume of cFS per dose = $2x10^{10 \text{ or } 11}$ CFU ÷ CFU/ml of cFS (value from below) | |
| • Volume of cryogenic media per dose = 250 ml – volume of cFS per dose<br><br>Example (low dose):<br>cFS potency (CFU/ml) = $1x10^9$<br>ml cFS per dose = $2x10^{10}$ /$1x10^9$ = 20 ml<br>ml cryo media per dose = 250ml - 20ml = 230 ml | |
| • Total doses from cFS = cFS total volume ÷ Volume of cFS per dose | |
| • Decide how many FMT products to prepare per low and high dose based on the calculations and how many low and high doses are still missing for the clinical trial and record the values in the table below.<br>*Note: To prevent a waste of cFS all high doses possible from this sample should be prepared and the remaining cFS volume should be used for the low dose type. Always round down to the nearest integer when calculating from total doses that could be prepared, e.g. the CFU of the cFS could yield 37.5 low doses or 3.75 high doses. In this case, 3 high doses and 7 low doses should be prepared.* | |

| Performed by (Init/Date) | Comments: |
|---|---|

| |
|---|
| For both dose types print N+1 labels and attach one label to a 250 ml cryogenic storage container for each of the N doses where N is the number of doses to be prepared and attach one of the labels of each dose type to this BR in the table below.<br><br>**MTC.FS FMT product** **BARCODE**<br>**$1x10^{10}$ or $1x10^{11}$ CFU dose**<br>***Batch date***<br>***Donor ID Sample ID*** |
| The above label contains a barcode to allow sample tracking in Freezer Works. The cryogenic bottles for clinical use will have an additional label (two labels in total): |

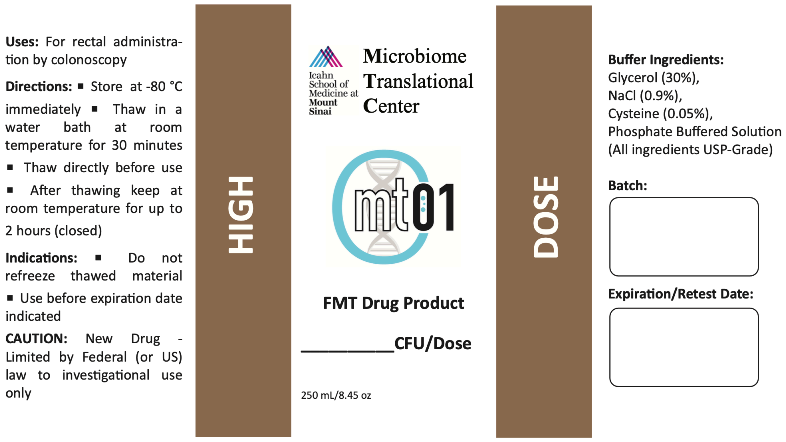

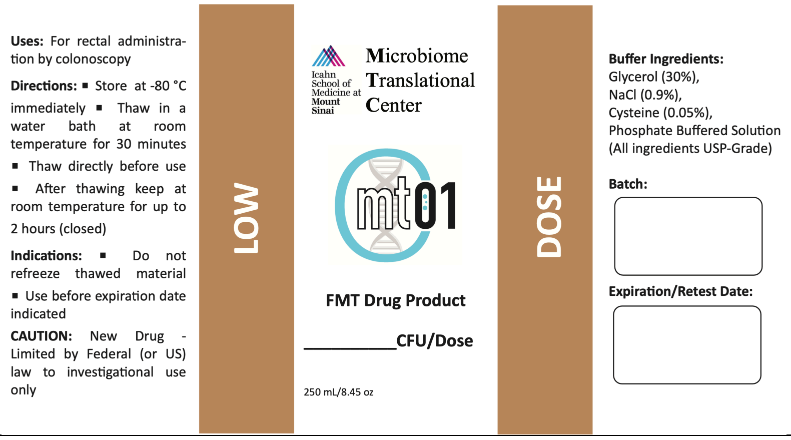

| |
|---|
| Print and attach 10 labels to the 10 QC vials per dose type. |

| Performed by (Init/Date) | Comments: |
|---|---|

| Dose type | CFU/ml of cFS | cFS total weight [g] | Weight of cFS/dose [g] | Volume of media/dose [ml] | Total doses possible per cFS | Doses to be prepared | Extra label |
|---|---|---|---|---|---|---|---|
| **High dose** | | | | | | | |
| **Low dose** | | | | | | | |

| Normalization of cFS to a dose of $1x10^{10/11}$ CFU FS for FMT administration (Procedure performed inside the BSC) | Done (√) |
|---|---|
| Print and attach *MTC-MBR-FS-0002_Fecal Slurry Cryopreservation Media for Fecal Microbiota Transplantation* to this batch record and prepare cryopreservation media accordingly. | |
| Print and attach *MTC-MBR-FS-0003_Sterilization of Biosafety Cabinet and Stomacher for Fecal Slurry Production for Fecal Microbiota Transplantation* to this batch record and perform the sterilization of the BSC and the equipment listed below accordingly, before the start of normalized FS generation.<br><br>Equipment and supplies to be sterilized:<br>☐ Sterile cryogenic media<br>☐ N labeled cryogenic storage containers (250 ml) for each dose type (N = Number of cryogenic storage containers calculated above)<br>☐ Pipette controller (recharged)<br>☐ 2x100ml serological pipettes, 3x50 ml serological pipettes, 2x1ml serological pipettes per dose type<br>☐ Vial rack with 10 labelled QC vials (1-2 mL) per dose type<br>☐ 8-Channel decapper<br>☐ 1 spray bottle ethanol (70 %)<br>☐ Kimwipes<br>☐ Trash bag | |

| Performed by (Init/Date) | Comments: |
|---|---|

| | | |
|---|---|---|
| Remove the Cryogenic storage containers with the cFS to be used for normalized FS FMT production from -80 °C freezer and thaw in a water bath with warm tap water (30–40 °C) with gentle swirling in between.<br>*Note: This takes 40-45 minutes and cFS must not stand any longer than necessary but processed immediately after being thawed.* | | |
| Perform the following steps for both dose types: | **Done (√)** | |
| | High | Low |
| • Ensure the Donor ID and Stool Sample ID for a given sample are identical between the **MTC.CFS unnormalized** container and the **MTC.FS FMT product** label. | | |
| • Transfer the Volume of cryogenic media per dose (calculated above) into each of the **MTC.FS FMT product** containers (250 ml) using the pipette controller and a sterile serologic pipette (50 ml/100 ml). | | |
| • Immediately after thawing the **MTC.CFS unnormalized** containers, spray them with ethanol (70%) and put them under the BSC. Swirl the containers and transfer the volume of cFS per dose into the **MTC.FS FMT product** containers (250 ml) with the pipette controller and a sterile serologic pipette (1 ml/50 ml). | | |
| • Close the lids of all but one of the storage containers tightly so that they snap into place. | | |
| • Homogenize the mixture by gently swirling the containers. | | |
| • Open the QC vials with the decapper. Transfer 10 aliquots of 100 µl normalized FS, from the cryogenic storage container with normalized FS that was not tightly closed, to the QC vials using the pipette controller and a sterile serologic pipette (1 ml) and close the vials again with the decapper. | | |
| Store the **MTC.FS FMT product** containers and the stability QC vials, <u>except one QC vial per dose for immediate CFU determination</u>, at -80 °C until usage or expiration. | | |
| Print and attach *MTC-MBR-FS-0004_CFU Count of the Fecal Slurry (FS) for Fecal Microbiota Transplantation (FMT)* to this batch record and perform the CFU/ml determination of the final FMT product directly after the production is finished for both dose types.<br>*Note: The acceptable limits are $6x10^{9}$ to $5x10^{10}$ CFU per 250 ml low dose and $6x10^{10}$ to $5x10^{11}$ CFU per 250 ml high dose.* | | |

| Performed by (Init/Date) | Comments: |
|---|---|

#### 10. POST-PRODUCTION REVIEW

| End-of-Production Cleaning | Done (√) |
|---|---|
| When the current production run is finished, discard all remaining consumables used and any leftover medium. | |
| Remove all reagents and supplies used during production. | |
| Clean all non-single use equipment removed from the chamber with ethanol (70%). | |

| Name | Signature | Date |
|---|---|---|
| Jeremiah Faith, PhD<br>Faculty Director | | |
| Ilaria Mogno, PhD<br>Quality Assurance Specialist | | |

| Performed by (Init/Date) | Comments: |
|---|---|

| **Reviewed By:** | | **Date:** |
|---|---|---|
| Jeremiah Faith, PhD<br>Faculty Director | | |
| **Approved By:** | | **Date:** |
| Ilaria Mogno, PhD<br>Quality Assurance Specialist | | |
| Control No: | | |

| STUDY ID |
|---|
| **Batch number** *(yyyy-mm-dd-FS.DonorID.StoolID)* |

#### 1. REFERENCE SOP(s)

- Materials Management for Fecal Slurry (FS) for Fecal Microbiota Transplantation (FMT), MTC-SOP-FS-0001
- Generation of CFU normalized Fecal Slurry (FS) for Fecal Microbiota Transplantation (FMT), MTC-SOP-FS-0002
- Fecal Slurry Cryopreservation Media for Fecal Microbiota Transplantation (FMT), MTC-SOP-FS-0003

#### 2. EQUIPMENT CHECKLIST

| Equipment Type | Serial Number | Post-Production Cleaning | |
|---|---|---|---|
| | | Initials | Date |
| Biological Safety Cabinet | 130475124 A | | |
| Balance | B323415058 | | |
| Pipetting Controller | C03324777 | | |
| Anaerobic chamber | AC19-157 | Not applicable | Not applicable |
| *Other:* | | | |
| *Other:* | | | |

| Performed by (Init/Date) | Comments: |
|---|---|

#### 3. REAGENTS AND SUPPLIES

| Item | Manufacturer | Catalog No. | Lot/Batch No. | Exp. Date |
|---|---|---|---|---|
| Serological Pipets, 50 mL | | | | |
| Media Bottle | | | | |
| Bottle Top Filter | | | | |
| Filter Receiver Bottle | | | | |
| Weigh Cups | Fisher | 08-732-113 | Not specified | Not applicable |
| Plastic Spoons | VWR | 89233-128 | Not specified | Not applicable |
| **Item** | **Manufacturer** | **Catalog No.** | **Lot/Batch No.** | **Exp. Date** |
| Ethanol (70%) | Thermo Scientific | A407P-4 | | |
| USP Grade Phosphate Solution | Ricca | R580825020F | | |
| USP Grade Glycerol | J.T. Baker | 2143-01 | | |
| USP Grade L-cysteine | Spectrum | C1473 | | |
| USP Grade Sodium Chloride | Spectrum | SO155 | | |

#### 4. Preparation

| Cleaning | Done (√) |
|---|---|
| Clean balance before use with ethanol (70%).<br>*Note: Before cleaning turn balances off.* | |

| Cryogenic Media (USP Grade)<br>(Use only USP Grade chemicals for this step, perform under BSC and use aseptic technique) | | Done (√) |
|---|---|---|
| The Cryogenic Media is prepared for concentrated Fecal Slurry (cFS) ☐ or normalized FS FMT product ☐.<br>*Note: Cryogenic media must be prepared fresh 2-24 hours before every production.* | | |
| Record the volume of media to be prepared calculated in section 6 (cFS) or section 8 (FS FMT product) of the *MTC-MBR-FS-0001 Generation of CFU normalized Fecal Slurry (FS) for Fecal Microbiota Transplantation.* Add at least 100 mL extra volume and round up in steps of 0.1 L.<br>*Note: e.g. if media volume is 1.35 L prepare 1.5 L.* | Media volume (ml) = ________ | |
| Spray the required equipment with ethanol (70%) and place them under the BSC. | | |

| Performed by (Init/Date) | Comments: |
|---|---|

| | |
|---|---|
| Label a sterile media bottle with: Cryogenic Media, *Reference Batch Number* | |
| Weigh 9 g ***USP Grade*** sodium chloride per 1 L medium into a clean weigh cup using a plastic spoon and add it to the sterile media bottle.<br>*Note: e.g.13.5 g sodium chloride for the preparation of 1.5 L medium. Record the amount added.* | **Amount [g]** |
| Weigh 0.5 g ***USP Grade*** L-cysteine hydrochloride per 1 L medium into a clean weigh cup using a plastic spoon and add it to the sterile media bottle.<br>*Note: e.g. 0.75 g L-cysteine for the preparation of 1.5 L medium. Record the amount added.* | **Amount [g]** |
| Transfer 300 mL of ***USP Grade*** glycerol per 1 L medium with a sterile serological pipette (50 ml) into the media bottle, fill it up to the volume calculated above with sterile ***USP Grade* Phosphate Solution**, and mix by inverting until components are dissolved completely (≥ 10 times).<br>*Note: e.g. 450 ml glycerol for the preparation of 1.5 L medium. Record the amount added.* | **Amount [mL]** |

| **Filter Sterilization (Perform under BSC and use aseptic technique)** | **Done (√)** |
|---|---|
| Fill the cryogenic media into a filter unit, filter using the vacuum pump in the BSC and close the flask aseptically with the included sterile cap. | |
| Label the flask with: Cryogenic Media, *Reference Batch Number* | |
| Store the media in the anerobic chamber. | |

#### 5. POST-PRODUCTION REVIEW

| **End-of-Production Cleaning** | **Done (√)** |
|---|---|
| Clean the balance, the pipetting controller and the BSC with ethanol (70%).<br>*Note: Before cleaning turn balance off.* | |

| **Name** | **Signature** | **Date** |
|---|---|---|
| Jeremiah Faith, PhD<br>Faculty Director | | |
| Ilaria Mogno, PhD<br>Quality Assurance Specialist | | |

| Performed by (Init/Date) | Comments: |
|---|---|

| Microbiome Translational Center (Icahn School of Medicine at Mount Sinai) | **Microbiome Translational Center Master Batch Record** |
|---|---|

| | |
|---|---|
| **Title: Sterilization of Biosafety Cabinet and Stomacher for Fecal Slurry Production for Fecal Microbiota Transplantation** | **MBR No: MTC-MBR-FS-0003** |
| | **Batch No.** |
| | **Effective Date: 08/15/2023**<br>**Supersedes Date: NEW** |

| **Reviewed By:** | | **Date:** |
|---|---|---|
| Jeremiah Faith, PhD<br>Faculty Director | | |
| **Approved By:** | | **Date:** |
| Ilaria Mogno, PhD<br>Quality Assurance Specialist | | |
| Control No: | | |

| **STUDY ID** |
|---|
| **Batch number**<br>***(yyyy-mm-dd- FS.DonorID.StoolID)*** |

#### 1. REFERENCE SOP(s)

- Materials Management for Fecal Slurry (FS) for Fecal Microbiota Transplantation (FMT), MTC-SOP-FS-0001
- Generation of CFU normalized Fecal Slurry (FS) for Fecal Microbiota Transplantation (FMT), MTC-SOP-FS-0002
- Sterilization of Biosafety Cabinet and Stomacher for Fecal Slurry Production for Fecal Microbiota Transplantation (FMT), MTC-SOP-FS-0004

#### 2. EQUIPMENT CHECKLIST

| **Equipment Type** | **Serial Number** | **Post-Production Cleaning** | |
|---|---|---|---|
| | | **Initials** | **Date** |
| Biosafety Cabinet | 130475124 A | Not applicable | Not applicable |
| Stomacher 400 EVO Paddle Blender | EVO - 1078 | Not applicable | Not applicable |
| Graduated Cylinder, 500 ml | Not specified | | |
| Pipette Controller | C03324777 | | |
| Spray Bottle, 500 ml | Not specified | Not applicable | Not applicable |
| *Other:* | | | |

| Performed by (Init/Date) | Comments: |
|---|---|

#### 3. REAGENTS AND SUPPLIES

| Item | Manufacturer | Catalog No. | Lot/Batch No. | Exp. Date |
|---|---|---|---|---|
| Quatricide-PV15 | Pharmacal Research Labs | Fisher, NC9371434 | | |
| Serological Pipette, 1 ml | Corning | 07-200-3 | | |
| Paper Towels | Not specified | Not specified | Not applicable | Not applicable |
| *Other:* | | | | |

#### 4. Preparation

| Quatricide PV-15 Preparation | Done (√) |
|---|---|
| The sterilization is performed for the partitioning of stool sample ☐, the generation of concentrated Fecal Slurry (cFS) ☐ or the generation of normalized FS FMT product ☐. | |
| Wear protective lab glasses, gloves, and lab coat. | |
| Transfer 2 ml Quatricide PV-15 with the pipette controller and a sterile serological pipette (1 ml) into a clean spray bottle, measure 498 ml water in a graduated cylinder and add it to the bottle. | |
| Mix well by inverting 10 times. | |

#### 5. Sterilization Process

| Surface disinfection | Done (√) |
|---|---|
| Spray the biosafety cabinet work surface, the interior walls, and the interior surface of the window with the Quatricide PV-15 solution. | |
| Bring the required equipment and supplies (see *MTC-MBR-FS-0001_Generation of CFU normalized Fecal Slurry (FS) for Fecal Microbiota Transplantation (FMT)* section 6 for stool sample partition, section 8 for cFS or section 9 for normalized FS) into the biological safety cabinet and spray with Quatricide PV-15. | |
| Spray surfaces and the bag compartment of the Stomacher EVO400 with Quatricide PV-15 if cFS is to be generated. | |
| Allow a 10-minute contact time. | |
| Wipe the biosafety cabinet, equipment, supplies, and the Stomacher EVO400 clean to remove residual Quatricide PV-15. | |

| Performed by (Init/Date) | Comments: |
|---|---|

| Name | Signature | Date |
|---|---|---|
| Jeremiah Faith, PhD<br>Faculty Director | | |
| Ilaria Mogno, PhD<br>Quality Assurance Specialist | | |

| Performed by (Init/Date) | Comments: |
|---|---|

| **Reviewed By:** | | **Date:** |
|---|---|---|
| Jeremiah Faith, PhD<br>Faculty Director | | |
| **Approved By:** | | **Date:** |
| Ilaria Mogno, PhD<br>Quality Assurance Specialist | | |
| Control No: | | |

| |
|---|
| **STUDY ID** |
| **Batch number**<br>***(yyyy-mm-dd-FS.DonorID.StoolID)*** |

#### 1. REFERENCE SOP(s)

- Materials Management for Fecal Slurry (FS) for Fecal Microbiota Transplantation (FMT), MTC-SOP-FS-0001
- Generation of CFU normalized Fecal Slurry (FS) for Fecal Microbiota Transplantation (FMT), MTC-SOP-FS-0002
- CFU Count of the Fecal Slurry (FS) for Fecal Microbiota Transplantation (FMT), MTC-SOP-FS-0005

| Performed by (Init/Date) | Comments: |
|---|---|

#### 2. EQUIPMENT CHECKLIST

| Equipment Type | Serial Number | Post-Production Cleaning | |
|---|---|---|---|
| | | Initials | Date |
| Anaerobic Chamber | AC19-157 | Not applicable | Not applicable |
| Micropipette (100 µL) | 18070827 | | |
| Micropipette (10 µL) | H370821 | | |
| Multichannel Micropipette | | | |
| 8-Channel Decapper | 022415367 | | |
| Vial Rack | Not specified | | |
| Vortex Mixer | Not specified | Not applicable | Not applicable |
| *Other:* | | | |

#### 3. REAGENTS AND SUPPLIES

| Item | Manufacturer | Catalog No. | Lot/Batch No. | Exp. Date |
|---|---|---|---|---|
| Pipet Tips, 10µL | | | | Not applicable |
| Pipet Tips, 100µL | | | | Not applicable |
| Multichannel Tips | Thermo Scientific | 94410053 | | Not applicable |
| Ziploc Bag | Office Depot | Not applicable | Not applicable | Not applicable |
| Chocolate Agar Plates | BD Biosciences | 221169 | | |
| Ethanol (70%) | Fisher Scientific | A407P-4 | | |
| Phosphate Buffer Solution | Gibco | 10010-023 | | |
| 96 Well Plate | | | | |
| Absorbent Underpads | VWR | 56616-022 | Not applicable | Not applicable |
| *Other:* | | | | |

| Performed by (Init/Date) | Comments: |
|---|---|

#### 4. CFU Count – homogenized fecal sample after two freeze thaws and final FS FMT product

Donor stool samples are banked at -80C prior to homogenization into concentrated fecal slurry (cFS). While generating cFS, an aliquot is taken and freeze-thawed one more time prior to the step of calculating the CFU/ml and CFU/g to reflect the freeze thaw cycle before combining cFS into normalized fecal slurry (FS). This CFU/ml is then used to determine the volume of cFS to generate the normalized FS to be used as the fecal transplant.

Furthermore, the CFU/ml of the FS product for fecal microbiota transplantation (FMT) is determined. For this purpose, an aliquot is taken at the end of the production that is directly used for CFU count.

| **Preparation<br>(In the anaerobic chamber)** | **Done (√)** |
|---|---|
| CFU count is determined for concentrated Fecal Slurry (cFS) ☐ or normalized FS FMT product (high and/or low dose) ☐. | |
| For the CFU count of cFS make sure the vials have been freeze thawed as described in *MTC-MBR-FS-0001_Generation of CFU normalized Fecal Slurry (FS) for Fecal Microbiota Transplantation (FMT).*<br>*Note: This is indicated by a check mark on the QC vial.* | |
| Spread one underpad on the work surface in the chamber, to protect the chamber from spills, | |
| Label two chocolate agar plates per FS sample to be tested with:<br>*Reference Batch Number* Plate 1 *or* 2 and *sample type* (cFS *or* FS*)*<br>*Note: Agar plates need to be placed in the anaerobic chamber at least 24 h before starting the protocol. Two agar plates as technical replicates for the CFU determination of cFS or normalized FS in MTC-MBR-FS-0001.* | |
| Transfer 90 µL of sterile PBS into positions B1-G1 of a sterile 96 well plate by using a micropipette (100 µL).<br>*Note: Dilutions are identical for cFS and FS. If you have more samples to analyze separate samples by one column; e.g., next sample is prepared in positions B3-G3.* | |

| Performed by (Init/Date) | Comments: |
|---|---|

| Dilution series (In the anaerobic chamber) | Done (√) cFS | high dose | low dose |
|---|---|---|---|
| Perform the following steps for all FS samples to be analyzed:<br>• Homogenize the aliquot of the cFS or FS by vortexing, open the vial with the decapper and transfer 100 µL to position A with the help of a micropipette (100 µL). | | | |
| • Starting in position A, use a micropipette (10 µL) to transfer 10 µL to the next position (B) and mix by pipetting up and down (≥ 3 times). | | | |
| • Dispose the pipette tip, use a new pipette tip to mix the suspension in position B by pipetting up and down (≥ 3 times) and transfer 10 µL to the next well (C). | | | |
| • Repeat the process from this well (C) with a new pipette tip until reaching position G. | | | |

| Plating (In the anaerobic chamber) | Done (√) cFS | high dose | low dose |
|---|---|---|---|
| Perform the following steps for all FS samples to be analyzed: — Date/Time: | | | |
| • Use the multichannel micropipette (program CFU SPOT PLATE), with 4 multichannel pipette tips, to transfer ten-times 1 µL bacterial solution, from positions D-G (dilutions $10^{-3}$-$10^{-6}$) to the respective chocolate agar plate and repeat this step with new tips for the second chocolate agar plate.<br>*Note: Agar plates need to be dry. If there is water in the agar plate lids, knock lid on sterile towelette to remove water. Decrease spacing on multichannel pipette to second stopper position (8) for plating.* | | | |
| • Leave the plates sitting, lid side up, for 5-10 minutes. | | | |
| • Incubate the plates at 37 °C, lid side down, in a Ziploc bag until colonies are visible.<br>*Note: Check the colony growth at least every 24 hours and count colonies as soon as they become clearly visible. <u>Do not let colonies grow too long</u> to prevent inaccurate counts.* | | | |

| Performed by (Init/Date) | Comments: |
|---|---|

| Counting (Outside the anaerobic chamber) | Done (√) |
|---|---|
| The countable range of colonies is between 10-100 CFU for the total of 10 spots of one dilution. Everything above is considered too numerous to count (TNTC). | |
| **Results for Cryogenic Vials** | |
| Calculate the CFU/ml for the lowest dilution in the countable range of all samples to be analyzed using the following table: $CFU/mL = (colonies * 100) * (\frac{1}{dilution})$ — Date/Time: | |

| Sample type | Vial | | | | | |
|---|---|---|---|---|---|---|
| ☐ cFS<br>☐ low dose<br>☐ high dose | Dilution | $10^{-3}$ (D1) | $10^{-4}$ (E1) | $10^{-5}$ (F1) | $10^{-6}$ (G1) | Comment |
| | Agar Plate 1 | | | | | |
| | Agar Plate 2 | | | | | |
| | Ø CFU/ml | | | | | |
| ☐ cFS<br>☐ low dose<br>☐ high dose | Dilution | $10^{-3}$ (D3) | $10^{-4}$ (E3) | $10^{-5}$ (F3) | $10^{-6}$ (G3) | Comment |
| | Agar Plate 1 | | | | | |
| | Agar Plate 2 | | | | | |
| | Ø CFU/mL | | | | | |

#### 5. POST-PRODUCTION REVIEW

| End-of-Production Cleaning | Done (√) |
|---|---|
| Clean all equipment after use with ethanol (70%). | |

| Performed by (Init/Date) | Comments: |
|---|---|

| Name | Signature | Date |
|---|---|---|
| Jeremiah Faith, PhD<br>Faculty Director | | |
| Ilaria Mogno, PhD<br>Quality Assurance Specialist | | |

| Performed by (Init/Date) | Comments: |
|---|---|
