## Supplementary material for "Live biotherapeutic product exhibits similar efficacy and superior engraftment to same donor fecal microbiota transplant for recurrent Clostridioides difficile infection": Bethlehem etal MTC01 preIND

[*] Co-corresponding authors

### Icahn School of Medicine at Mount Sinai

### Pre-IND briefing packet

### MTC01

Jeremiah Faith PhD
Ari Grinspan MD

October 2, 2020

### LIST OF FIGURES

**Figure 1.** The rCDI microbiota has a fitness defect that is therapeutically treatable by FMT. 9
**Figure 2.** Accurate detection of bacterial strains from complex gut communities through low depth metagenomics. 10
**Figure 3.** Quantification of recipient strain persistence and donor strain engraftment. 11
**Figure 4.** Total CFUs transplanted for successful rCDI FMTs using donor material from which MTC01 is derived (n = 8). 18
**Figure 5.** Initial isolation and cell banking system. 20
**Figure 6.** LBP manufacturing facility. 23
**Figure 7.** Predicted antibiotic resistance genes and mobile genetic elements. 46

### LIST OF TABLES

**Table 1.** LBP strains. 12
**Table 2.** Initial isolation. 20
**Table 3.** Major facility equipment. 22
**Table 4.** Treatment strategy 26
**Table 5.** Virulence factors and toxins 43
**Table 6.** Predicted antibiotic resistance genes 44
**Table 7.** Genomic identity. 50
**Table 8.** Bruker MALDI Biotyper identity. 50
**Table 9.** Health records FMT donors. 52

### LIST OF ABBREVIATIONS

| | |
|---|---|
| MALDI-TOF | Matrix-Assisted Laser Desorption/Ionization-Time Of Flight |
| FMT | Fecal Microbiota Transplantation |
| rCDI | recurrent *Clostridioides difficile* infection |
| OTC | Over-the-counter |
| LBP | Live Biotherapeutic Product |
| SAE | Serious Adverse Event |
| MDRO | Multi-Drug Resistant Organisms |
| DSMB | Data and Safety Monitoring Board |
| RCB | Research Cell Bank |
| MCB | Master Cell Bank |
| WCB | Working Cell Bank |
| DS | Drug Substance |
| DP | Drug Product |
| MIC | Minimum Inhibitory Concentration |

### 1 PRE-IND MEETING INFORMATION

#### 1.1 Product name and application number

MTC01

#### 1.2 Chemical name and structure

This is a Live Biotherapeutic Product (LBP) that consists of 16 bacterial strains (Table 1) isolated from healthy Fecal Microbiota Transplantation (FMT) donors that were used to successfully treat recurrent *Clostridioides difficile* infection (rCDI).

#### 1.3 Proposed indication(s)

MTC01 is developed for the treatment of rCDI.

#### 1.4 Dosage form, route of administration and dosing regimen

The dosage form and route of administration will be a mixture of cryopreserved bacterial strains delivered by enema or colonoscopy. The targeted dose will be $10^9$-$10^{11}$ CFU for each microbe per dose.

#### 1.5 Purpose of the meeting

The purpose of the meeting is to gather feedback from the agency to the submitted questions and reach agreement on the proposed pre-clinical, Chemistry, Manufacturing and Controls and the phase I clinical design.

#### 1.6 List of participants (expertise)

The meeting will be Written Responses only. These individuals contributed substantially to this pre-IND submission and will evaluate the written responses from the FDA.

- Jeremiah Faith PhD, Icahn School of Medicine at Mount Sinai (microbiome)
- Ari Grinspan MD, Icahn School of Medicine at Mount Sinai (FMT/gastroenterology)
- Harm van Bakel PhD, Icahn School of Medicine at Mount Sinai (MDRO, *C. difficile*)
- Marcia Meseck, MS, JD, Icahn School of Medicine at Mount Sinai (cGMP expert)

#### 1.7 proposed agenda

The FDA meeting confirmation on Sept 22, 2020 in response to our pre-IND meeting request submitted on Sept 3, 2020 declared we will use a Written Responses meeting format.

#### 1.8 A list of specific questions

##### *1.8.1 Pre-clinical*

1. The sponsor is planning to perform antibiotic resistance profiling on each of the 16 strains individually using a panel of 10 antibiotics approved by an Infectious Disease clinician. MICs will be determined by etest (in-house) and agar plate dilutions (CRO using CSLI methods). Does the Agency agree with this approach to determine antibiotic susceptibility for the strains in the product MTC01?
2. Does the Agency agree that safety data consisting of *in silico* and *in vitro* analysis of antibiotic resistance in combination with the *in human* safety data composed of equivalent dosing of these 16 strains in the form of FMT in one to seven humans

with long-term colonization of all strains without FMT-related SAE (up to 5 years) is sufficient for the proposed phase I study?

##### *1.8.2 Clinical*

1. To select the administration route with the highest success rate in FMT for rCDI, we plan to colonoscopically administer a single dose LBP at an equivalent dose (~$5x10^{11}$ CFU) to that used in FMT for the donors from which the LBP strains were isolated. The selected strains for the LBP are those that engrafted in at least 80% of recipients by FMT with a prevalence in the healthy human population of at least 25%. Does the Agency agree with the plan to conduct a phase I study of a colonoscopically administered LBP with a primary endpoint of safety in rCDI patients?

##### *1.8.3 Chemistry, Manufacturing and Controls*

1. The quantities of bacteria needed for a single dose LBP at equivalent CFU to FMT are sufficiently low to allow anaerobically culturing of relatively small volumes (<1L) of each strain. Towards this, we have worked with Coy Laboratories (a major vendor of anaerobic chambers) to design dedicated anaerobic chambers that we will sterilize between production rounds, similar to the technologies used to maintain germ-free mice for years a time. Each strain will be cultured in a sealed centrifuge bottle that enables the culture to be temporarily removed from the chamber without contamination while remaining anaerobic (e.g., to centrifuge the culture). The sealed container is then surface sterilized when returned to the anaerobic chamber. We are confident these chambers will provide the flexibility to manufacture small batch, pure cultures without the complexity, cost, and contamination risks associated with bioreactor systems where large volumes are transferred with multiple opportunities for contamination. Does the agency agree with this manufacturing strategy?
2. Each strain will be individually cultured in animal product free culture media followed by twice washing in UPS grade sterile PBS-cysteine with final drug substance in USP grade PBS-cysteine-glycerol. The identity of each drug substance strain will be confirmed by plating and MALDI-TOF (Bruker Biotyper), while purity will be assessed by MALDI-TOF of 100 colonies, requiring all good spectra to match the appropriate species. The potency of each strain's drug substance will be measured in CFU. Drug substance will be maintained at -80C. Does the agency agree with this approach for producing and evaluating drug substance?
3. The drug product will be produced by combining each of the 16 strains in roughly equally proportions by CFU. The volume will be adjusted to 125 ml with reduced USP grade PBS+glycerol+cysteine+glucose to have sufficient volume for colonoscopic delivery. Drug product will be maintained at -80C and thawed prior to infusion in a similar manner to FMT. Identity, potency, and purity will be determined in a similar manner to the drug substance, except that we will assay 200 colonies of the drug product by MALDI-TOF to better estimate the proportional representation of the mixed culture. Similar assays will be performed monthly to determine product stability Does the agency agree with this approach for producing and evaluating drug substance?

##### *1.8.4 Regulatory*

1. Does the Agency agree that if safety data are positive from this trial, we can provide LBP material to other investigators for clinical use in recurrent *C. difficile* infection under enforcement discretion, similar to FMT stool banks?

### 2 BACKGROUND

#### 2.1 Clinical Background

*Clostridioides difficile* infection (CDI) is the leading cause of health care associated diarrhea with approximately half a million cases and 29,000 deaths in the US.[1] CDI is associated with antibiotic-induced dysbiosis[2] and treatment consists of discontinuation of the inciting antibiotic followed by antimicrobial therapy.[3] However, the cure rate is only 20 – 30%[4] and a third of the patients develop a recurrence of CDI (rCDI) that is associated with a 33% increased risk of mortality. Fecal Microbiota Transplantation (FMT) is an emerging therapeutic option for patients with rCDI. A recent randomized, active-comparator, open-label clinical trial found that clinical resolution of FMT preceded by vancomycin treatment (FMTv, 92%) was superior to antimicrobial monotherapy (fidaxomicin, 42% or vancomycin, 19%).[5] FMT is generally considered safe with >40,000 FMTs performed and few adverse events. However, FMT is an undefined product and contains hundreds of strains, including beneficial and potentially harmful microbes. For example, two immunocompromised adults who received FMT from the same donor developed invasive infections caused by extended spectrum beta-lactamases (ESBL)-producing *E. coli* that resulted in the death of one individual[6,7]. More recently, there are concerns that even in the context of rCDI performed with FMT product from the most established stool biobanks, that only provides material manufactured from pre-screened donors, can lead to the transmission of enteropathogenic Escherichia coli (EPEC) and Shigatoxin-producing Escherichia coli (STEC) associated with illness and death[8]. Furthermore, the presence of SARS-CoV-2 in the stool of individuals with COVID-19

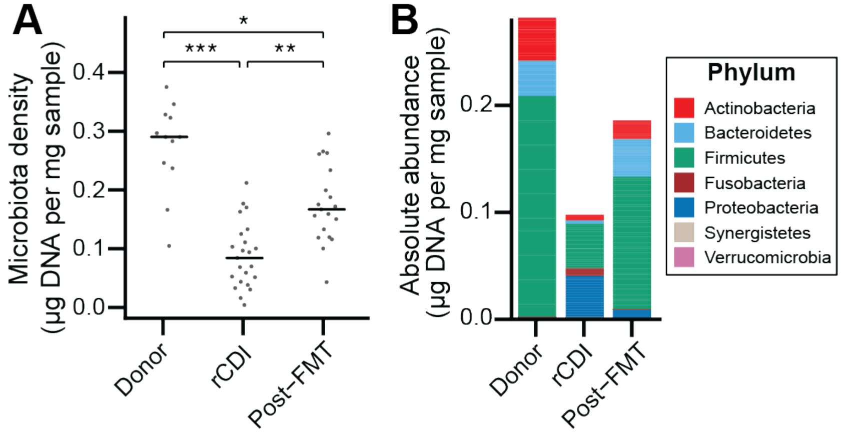

**Figure 1.** The rCDI microbiota has a fitness defect that is therapeutically treatable by FMT.
(A) rCDI subjects have reduced microbiota densities that are significantly increased upon FMT with donor microbiotas. (B) Following FMT, the composition of the microbiota of individuals with rCDI is restored to more closely resemble that of healthy donors in absolute terms. In (A) points represent individual samples, bars indicate median, *p < 0.05, **p < 0.01, and ***p < 0.001 (Kruskal-Wallis with Dunn's post-test corrected for multiple comparisons with the Bonferonni correction).

provides the possibility of viral transmission by FMT[32]. These events highlight the need for stringent screening, specifically for the presence of multidrug resistant organisms and potential pathogens. However, with any undefined product, the risk will be inconclusive and inconsistent. Given the unsurpassed efficacy of FMT for rCDI, a Live Biotherapeutic Product (LBP) composed of a defined cocktail of bacteria with an equivalent efficacy, but improved safety profile and cost-effective manufacturing process is warranted.

Mount Sinai Hospital was an early adopter of FMT for rCDI in the New York region and has a biobank of fecal material from rCDI FMT donor and rCDI recipients from as early as 2014. We have previously used these samples to generally characterize the gut microbiome and the clinical response of individuals with rCDI that do or do not have Inflammatory Bowel Disease (IBD)[9]. We have also used absolute quantification of gut microbiome composition and functional ex-germ-free mouse transfer experiments to demonstrate that the fitness and density of the gut microbiota of rCDI individuals are severely reduced compared to healthy controls and that both gut microbiota fitness[10] and density (Figure 1) are increased after FMT.

The earliest documented use of a defined consortium of bacteria for the treatment of *C. difficile* was that of Tvede and Rask-Madsen in 1989 when five patients were successfully treated with a mixture of 10 bacteria in sterile saline (approximately 2x$10^9$ CFU per strain in a total volume of 200 ml administered rectally)[11]. A key observation in this study was that individuals with *C. difficile* infection were found to not have species from the genus *Bacteroides* in their stools prior to the defined microbiota transplant, whereas such microbes were readily isolatable after the transplant. The transplant contained three *Bacteroides* species, *B. ovatus*, *B. vulgatus*, and *B. thetaiotaomicron* in addition to several *Clostridium* and facultative anaerobes

In the >30 years since this study, there has been an explosion in our ability to characterize the gut microbiome both with culture-independent and culture-dependent methods. Our lab has established a robust high throughput pipeline for the isolation, propagation, storage and recovery of gut microbes from human microbiome samples. To understand the dynamics and stability of FMT engraftment, we developed a novel algorithm that uses reference genomes of bacteria isolated from FMT donors to accurately and sensitively track each microbe in as few as 2.5 million metagenomic reads from each FMT

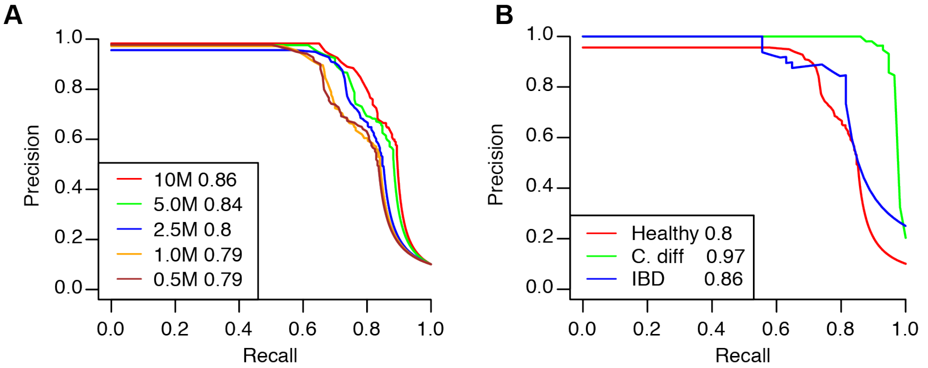

**Figure 2.** Accurate detection of bacterial strains from complex gut communities through low depth metagenomics.
(A) Our algorithm can accurately match 261 strains from different species to the correct donor sample from which they were isolated. 10M, 5M, etc… refers to the number of pair-end metagenomic reads used for strain detection. (B) The algorithm works similarly well on strains isolated from patients with rCDI or IBD with just 2.5M reads with improved performance on the low diversity microbiota from rCDI individuals. AUCs are provided in each legend box to the right of the number of reads.

recipient[31]. Specifically, the algorithm identifies and tracks the strain-unique subset of each strain's genome in fecal metagenome datasets. We validated this algorithm using a dataset of 261 bacterial genomes isolated from 10 individuals whose metagenome was also sequenced. The *Strainer* algorithm detected >75% of the donor strains in the metagenomes with a precision of >0.95 (i.e., less than 5% false positives). The algorithm performed similarly well on test datasets generated from bacterial isolate genomes and metagenomes from individuals with IBD and rCDI (Figure 2). We applied this strain tracking algorithm to quantify the persistence of FMT recipient strains after transplant. Therefore we cultured >3000 bacterial isolates and sequenced bacterial genomes from eight of the recipients in our cohort. As in the study of Tvede and Rask-Madsen, we isolated no species from the genus *Bacteroides* in the recipients before FMT. However, with far more extensive technologies to isolate microbes and more accurate tools to identify the taxonomy of each isolate (using both MALDI-TOF mass spectrometry and whole genome sequencing), we also determined that these recipients lacked bacterial members from the entire phylum of Bacteroidetes. Given that Bacteroidetes typically represent the first or second most abundant phylum in the colon, often encompassing 25-80% of the microbiota, the lack of any representative from the phylum across all tested individuals was remarkable. From our extensive culturing of bacteria over the past 5 years totaling >100 samples from >80 individuals, we culture 8.7±2.3 species from the Bacteroidetes phylum from healthy individuals, and the fewest Bacteroidetes we have ever cultured from a healthy individual was three unique species. These results indicate that, compared to the healthy gut microbiota, the absence of Bacteroidetes in rCDI patients is one of the most defining microbiome characteristics of this disease. Representatives from the other three major phyla in a healthy gut (Actinobacteria, Proteobacteria, and Firmicutes) were all found in subjects with rCDI prior to FMT. Importantly, individuals with rCDI have an excess of uncommon Proteobacteria in their stool that we found were largely eliminated from their stool by 4-8wks post-FMT.

Approximately 25% of the recipient microbiota persists for the duration of our sampling (Figure 3A)[31]. These persistent strains were largely species from the genus

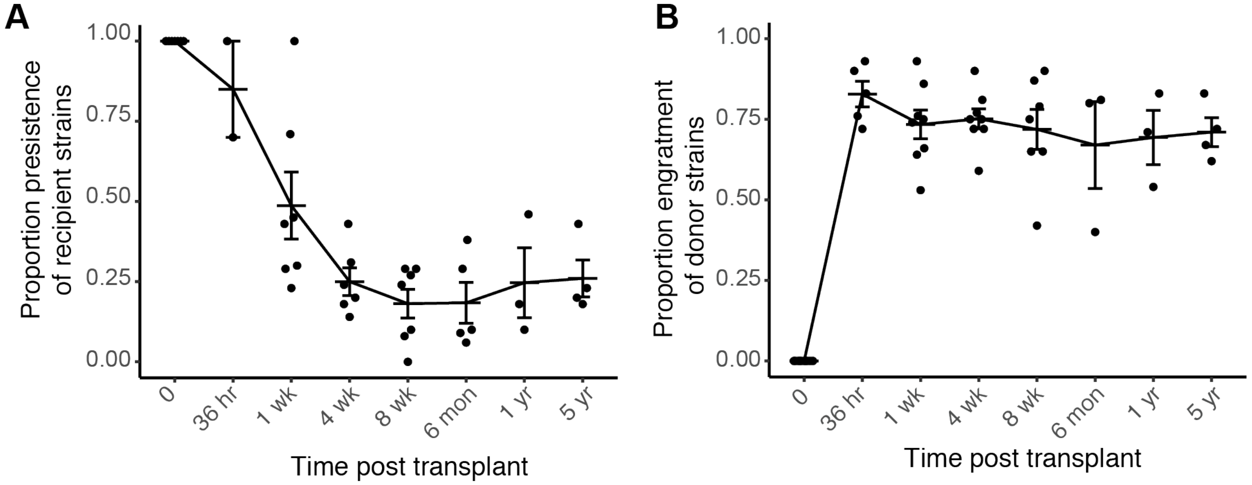

**Figure 3.** Quantification of recipient strain persistence and donor strain engraftment.
A) FMT led to a graduate decline in the proportion of pre-FMT recipient strains that remained stably engrafted for the first 8wks, while those strains (largely from genus Bifidobacterium) that were still present 8 wks were also largely stably colonized for the 5 year duration of the sampling. (B) Unlike the slow decline in recipient persistence, the donor strains engrafted immediately and largely remained stably colonized for the 5 year duration of sampling demonstrating the durability of gut microbiota manipulation as a therapeutic strategy.

Bifidobacterium – the dominant representatives in the healthy gut from the phylum Actinobacteria in the gut.

We found the average proportional engraftment of donor strains into the gut microbiota of recipients that did not relapse in the first 8 wks post-FMT was over >0.7. As in the healthy human gut microbiota[8], we found an incredible stability of these newly engrafted microbes that remained for the >5 year post-FMT sampling in non-relapsing individuals (Figure 3B). Although, we found engraftment of strains from all four major gut microbiota phyla into the recipients, the least successful engrafting strains were those from the genus Bifidobacterium suggesting perhaps their engraftment was prohibited by the persistence of the original Bifidobacterium in the recipient microbiota as discussed above. The most consistent engrafting strains were from phylum Bacteroidetes with an average of 7.8±2.1 different species per recipient post-transplant (not significantly different from healthy individuals) and a proportional stable engraftment of >90% of donor Bacteroidetes strains, compared with no representative across all recipients prior to FMT. Strains that were not stably colonized in the donor microbiota were not stably engrafted in their recipients. Importantly, one donor in this study was used to treat seven independent recipients providing the opportunity to identify strains that stably engraft across numerous individuals.

#### 2.2 Pharmacological Class

##### *2.2.1 Original isolation*

We isolated bacteria from each FMT donor as colonies on agar plates by selective and non-selective conditions in anaerobic, microaerophilic, and aerobic conditions. We have an automated pipeline for bacterial isolation from human stools that has operated for over five years with libraries of bacteria isolated from >100 samples and subsequently used extensively for functional studies of the human gut microbiome[12,13]. For all eight FMT donors successful used at Mount Sinai Hospital and numerous recipients, we isolated bacteria with this pipeline and sequenced the bacterial genomes of the isolates. For the LBP strains in particular, we focused on the donor (1001283) whose stool was transplanted to seven individuals with rCDI. Two of these recipients relapsed but were then retransplanted with the same donor stool and cured of their rCDI. We focused our LBP strain-selection on frequently engrafting strains in the five non-relapsing recipients of donor 1001283. Importantly, there were no SAE in any of the seven recipients of this or any other donor in our studies.

To choose the LBP species and strains (Table 1), we identified 29 strains representing 19 species from donor 1001283 that engrafted in at least 4 of 5 recipients. In an effort to make the LBP cocktail compatible with the most people, we include only species whose frequency in the healthy human population was >25%. We then eliminated *B. fragilis* and *E. coli* due to their associations with clinical pathology. Only one species, *Collinsella aerofaciens*, was found present at >25% in the healthy human gut, was a frequently successful engrafter in our other FMT donors, and was not found in donor 1001283. We therefore included a strain of *C. aerofaciens* from another successful donor in our cohort (1001275). See Table 1 for the strain list, frequency in the healthy human population, and genome sequence. See Appendix 7 for donor health records.

**Table 1.** LBP strains.

| Strain | Species | Healthy Population Frequency | Accession |
|---|---|---|---|
| MTC01.01 | Bacteroides uniformis | 0.96 | SAMN15532497 |
| MTC01.02 | Bacteroides ovatus | 0.83 | SAMN15532699 |
| MTC01.03 | Bifidobacterium longum | 0.79 | SAMN15532405 |

| | | | |
|---|---|---|---|
| MTC01.04 | Bacteroides thetaiotaomicron | 0.75 | SAMN15532862 |
| MTC01.05 | Bacteroides vulgatus | 0.71 | SAMN15533157 |
| MTC01.06 | Collinsella aerofaciens | 0.67 | SAMN15533307 |
| MTC01.07 | Parabacteroides distasonis | 0.63 | SAMN15532962 |
| MTC01.08 | Bifidobacterium adolescentis | 0.54 | SAMN15532697 |
| MTC01.09 | Parabacteroides merdae | 0.50 | SAMN15532955 |
| MTC01.10 | Coprococcus comes | 0.42 | SAMN15532605 |
| MTC01.11 | Eubacterium rectale | 0.33 | SAMN15532976 |
| MTC01.12 | Bacteroides caccae | 0.33 | SAMN15532375 |
| MTC01.13 | Dorea longicatena | 0.33 | SAMN15532943 |
| MTC01.14 | Odoribacter splanchnicus | 0.29 | SAMN15533209 |
| MTC01.15 | Bacteroides cellulosilyticus | 0.29 | SAMN15532683 |
| MTC01.16 | Bifidobacterium pseudocatenulatum | 0.29 | SAMN15533121 |

#### 2.3 Regulatory Background

With the exception of over-the-counter (OTC) probiotics, there are no multispecies LBPs for clinical use. However, with the rise in interest of gut microbiota manipulation as a therapeutic target there are several LBPs in development that have finished phase I with a strong safety record[14,15].

#### 2.4 Mode of Action

The mechanism of action of the defined consortia is presumed to be the same as that of FMT, which is also not completely understood. Both FMT and our defined consortia are presumed to operate through a combination of colonization resistance/competitive exclusion, bile acid metabolism and the restoration of normal gut microbiota[16].

#### 2.5 Safety

##### *2.5.1 Clinical*

FMT for rCDI largely has a positive safety record and an impressive efficacy[5]. However, there is a growing concern that the undefined composition of the gut microbiota in feces can put the recipient at risk for receiving an opportunistic pathogen or pathobiont that was asymptomatically carried in the donor. This risk has been realized now in several infections and deaths[6–8]. To mitigate risks with FMT, donor stools are screened to *exclude* known pathogens and strains previously associated with SAEs. As all of our strains are derived from successful FMT donors, all of our communities have therefore passed these FDA safety criteria.

Importantly the inclusion of individual strains in an LBP vastly reduces the risk compared to the pathogen exclusion approach used for evaluating FMT donor stools. Furthermore, the defined composition of the consortia enables a consistent and well characterized input across all recipients so that risk can be accurately quantified and limited. No species in our consortia has been associated with a serious adverse event (SAE) in any FMT study (i.e., there are no strains of *Escherichia coli* or even representatives from the phylum Proteobacteria). Numerous studies have demonstrated that unrelated individuals do not typically share the same strain of bacteria. Therefore, given the approximately 400 donors used by OpenBiome for ~50,000 combined FMTs[33] and our selection of only species present in >25% of the healthy population (Table 1), we estimate 100-400 different strains of each species in our LBP have been tested from OpenBiome provided FMTs alone with an untarnished safety record in 12500-50000 recipients.

The specific strain of each species in our LBP (MTC01) have each been successfully used to treat rCDI patients and found stably engrafted for up to 5 years in the recipients without any SAE that can be attributed the engraftment of these strains. Although we provide additional safety information about these strains below, we feel these *in human* data are far more valuable than any *in silico* or *in vitro* assay we might perform on each strain.

##### 2.5.2 Non-clinical

To prevent the inclusion of any multi-drug resistant organisms (MDRO) in the LBP all community members have been tested *in silico* and will be tested *in vitro* for antibiotic susceptibly and prominent virulence factors. All microbes are susceptible to multiple antibiotics, providing several options for controlling the entire community in the case of an SAE.

###### 2.5.2.1 In silico evaluation for antibiotic resistance, virulence factors and mobile genetic elements

The results shown in Appendix 3 demonstrate that none of the strains in MTC01 are predicted to have antibiotic resistance genes on mobile genetic elements that associated with MDROs (Table 6, Figure 7) and no prominent virulence or toxin encoded genes were found (Appendix 2, Table 5).

###### 2.5.2.2 In vitro antibiotic testing

Antibiotic testing will be performed by etest, and agar dilution MIC according to CSLI protocols. Agar dilution MIC determinations have been completed on 11 of the 16 strains (APPENDIX 10). As expected, all strains are susceptible to multiple antibiotics. Importantly, the entire community tested thus far is also susceptible to multiple individual antibiotics.

#### 2.6 Clinical Information

##### 2.6.1 Previous Human Experience

As described above, FMT has been safely used in tens of thousands of individuals providing a rich resource of potential SAEs including death[6]. All major SAEs from FMT have involved MDROs or pathogenic strains of *E. coli*. These two concerns can be eliminated in the context of LBPs.

There are to date a limited number of published studies of multi-strain LBPs used for the treatment of *C. difficile*. In the Tvede and Rask-Madsen study described above, five individuals with CDI were treated with a rectal infusion of 10 cultured bacteria ($2x10^9$ CFU per strain; $2x10^{10}$ total dose) with all five patients responding and no SAE [11]. More recently Petrof et.al., treated two subjects with rCDI with a consortium of 33 bacterial isolates from a healthy donor stool [28]. The strains were administered by colonoscopy to the right and mid colon. Both patients responded with no SAE and remained symptom free for 6 months. The microbes were administered in 100 ml of pre-reduced saline at a concentration of $3.5x10^9$ CFU/ml giving a overall total dose of $3.5x10^{11}$. In addition to these two published examples above that did not use cGMP conditions for the manufacture of their consortia, there are press releases describing the results of phase I safety trials for LBP manufactured under cGMP. For example, Vedanta Biosciences gave single doses ranging from $1.6x10^9$ to $8x10^9$ CFU and cumulative doses of $4x10^{10}$ to $1.1x10^{11}$ over five or 14 days[15]. There were no SAEs across 23 healthy volunteers.

Although the defined consortium for this study has never been manufactured and provided to humans, the members have been transplanted to seven individuals in the context of FMT. The mean total CFU provided in these FMT studies was $6x10^{11}$. None of these individuals experienced SAEs, and all were cured of their rCDI (>70% on the first transplant and 100% on the second for those that relapse). We will use the same route of administration (colonoscopy) and dose ($5x10^{11}$ CFU) with the LBP as with the fecal slurry used in the FMT with these same donors.

##### 2.7 Proposed Initial Studies

In light of the widespread use of undefined FMT for the treatment of rCDI, the lack of SAE in all phase I trials of LBPs for healthy and rCDI individuals, the ethical/safety considerations of colonoscopy in healthy volunteers, and the far lower risk profile of a consortium of individually screened bacteria compared with stool, we will perform the original study directly in subjects with rCDI to expedite the development of a safer, more scalable alternative to stool.

#### 3 CLINICAL PLAN

##### 3.1 Summary

Our phase I trial is focused on a safety assessment of using a defined “synthetic stool” solution at a similar concentration to actual stool in human FMT. By choosing the best engrafting strains from a successful FMT donor used to treat multiple individuals with no SAE and eliminating those species that do not engraft well or that are associated with clinical pathology or MDROs, we expect to have a similar efficacy to stool but with a far more controlled and defined safety profile as well as improved scalability. Given the typically high response rate to FMT, we also expect to gain important insights into secondary outcomes of efficacy and microbial engraftment.

Assuming we attain our primary endpoint of safety within the CFU range of human stool-based FMT with a LBP, we would perform a larger efficacy trial and smaller trials to understand mechanism. After establishing a robust safety and efficacy profile for MTC01, we would in parallel move to transition (with other trials) to an oral version of the product to eliminate the need for the colonoscopy.

##### 3.2 Title of study

Defined microbiota transplant for recurrent *Clostridioides difficile* infection with the cultured, engrafting fraction of successful human FMT donors.

##### 3.3 Investigators/study center

###### *3.3.1 Investigators*

Jeremiah Faith, PhD
Associate Professor of Medicine, Clinical Immunology
Expertise: Gut microbiome, microbiology

Ari Grinspan, MD
Associate Professor of Medicine, Gastroenterology
Medicine, Gastroenterology
Expertise: Fecal Microbiota Transplantation clinical studies

Harm van Bakel, PhD
Assistant Professor of Genetics and Genomic Sciences
Expertise: MDROs, C. difficile

###### *3.3.2 Study center*

Mount Sinai Hospital

##### 3.4 Phase of development

Phase I

##### 3.5 Objectives

Safety and efficacy of LBP for rCDI

##### 3.6 Design of study

The study is designed to begin establishing the cultured, engrafting fraction of a successful FMT donor as a direct replacement of our current standard of care for rCDI at Mount Sinai. Specifically individuals with ≥ 2 episodes of confirmed recurrent CDI defined as the presence of diarrhea (Bristol 6 or 7 for 48 hours and a confirmatory test for CDI). Preferred testing will be a two-step method using GDH/EIA toxin, with the most recent episode occurring within the prior 3 months. Subjects will be taken off of all antibiotics for 48 hr prior to the transplant. Our successful human FMTs have transplanted mean viable bacterial loads of approximately $10^{11}$ CFU per transplant of a largely poorly defined mixture of bacteria and metabolites. As described above, we have identified, isolated, and manufactured a subset of strains that consistently engrafted in colonoscopic FMT for rCDI. This engrafting subset will be cultured at scale, washed, and resuspended in sterile GMP-grade PBS with 15% glycerol to retain viability in frozen storage, cysteine to maintain an anaerobic environment for the anaerobic consortium, and 1% glucose to provide an initial food source to the community upon colonization.

For our clinical FMTs at Mount Sinai, we administer on average $3x10^{11}$ microbes to the right colon by colonoscopy in a single infusion of 250ml with PBS as the diluent. Likewise for this LBP largely derived from our most frequent human FMT donor, we will administer between $1x10^{11}$ – $5x10^{11}$ CFU by a single infusion in a volume of 250ml with PBS-glycerol-cysteine-glucose as the diluent.

Each microbe will be evenly pooled in the consortium to reach the target CFU. Strains that do not reach a minimum viability of at least $1x10^{7}$ per dose in stability tests will be eliminated from the cocktail. Strains above the minimum viability that do not have sufficient density to be evenly pooled will be pooled at their maximal possible density. In our preliminary characterization of six strains in this LBP, each of the target strains reached $10^{8}$-$10^{10}$ per ml of culture without concentrating, suggesting that most likely we will be able to include equal quantities of all strains in the LBP drug product.

Each recipient will be monitored in the hospital for 1 hour post-transplant. We will follow-up with each subject at 72hr, at 1, 8, and 12 weeks, and at one year to monitor long term safety. Successful treatment will be determined by clinical cure at 8 weeks. We will collect stool pre-transplant and at 72hr, 1wk, 8wks, 12wks, and 26wks to quantify strain engraftment (see Appendix 6 for study design overview figure).

##### 3.7 Diagnosis and subject selection

###### *3.7.1 C. difficile diagnosis*

Pre*sence of C. difficile* will be determined by PCR or toxin EIA. As well as diarrhea (>=3 loose stools in 24h period, defined as BSS 6-7, x3 days).

##### *3.7.2 Eligibility criteria*

Ages eligible for study: 18 years and older

Sexes eligible for study: All

Accepts healthy volunteers: No

##### *3.7.3 Partial inclusion criteria*

1. Ages eligible for study: 18 years and older
2. Able and willing to provide written informed consent
3. Subjects with a qualifying recurrent CDI episode.
4. Subjects with more than 3 unformed stools per day over 2 consecutive days, a positive *C. difficile* stool test and diarrhea is considered unlikely to have another etiology.
5. CDI symptoms started within 30 days prior to enrollment.

##### *3.7.4 Partial exclusion criteria*

1. Female subjects who are pregnant or are planning to become pregnant during the study.
2. Known or suspected toxic megacolon and/or know small bowel ileus at the time of enrollment.
3. Prior participation in studies of investigational live biotherapeutic products or FMT within the last 6 months.
4. History of active diarrhea associated with inflammatory bowel disease (IBD).
5. Major gastrointestinal surgery within the last 3 months before enrollment.
6. Use of drugs that alter gut motility.
7. History of acute leukemia or hematopoietic stem cell transplantation or myelosuppressive chemotherapy within 2 months prior to enrollment.
8. Unable or unwilling to undergo a colonoscopy
9. Inpatient status, though patients can be screened while inpatients, the must be outpatient for the planned colonoscopy.
10. Anticipated immediate or upcoming surgery within 30 days
11. Need for continued non-anti-CDI antibiotic therapy
12. History of total proctocolectomy
13. Female patients who are pregnant or breastfeeding or plan to become pregnant in the next 6 months.
14. Patients who are unable to give informed consent
15. Participation in a clinical trial in the preceding 30 days or simultaneously during this trial
16. Severe food allergy (anaphylaxis or anaphylactoid-like reaction)
17. Life expectancy < 6 months
18. Unable to adhere to protocol requirements
19. Patient who have received an FMT in the past year
20. Any condition that the physician investigators deems unsafe, including other conditions or medications that the investigator determines that it will put the subject at greater risk from FMT
21. Patient who is diagnosed with class 3 or 4 Heart Failure
22. Lab value of WBC <3.0 x 103/mm3 , Platelets <100 x 103/mm3 , ALT or AST > 1.5 x institutional ULN

23. If a patient is heavily immunosuppressed and is negative for CMV or EBV

#### 3.8 Treatments

Single dose of $1x10^{11}$ to $5x10^{11}$ total CFU ($1x10^{7}$ to $6x10^{10}$ CFU per strain) administered via colonoscopic infusion.

#### 3.9 Dose Justification

The dose is based on the CFU range commonly used in successful FMT, the specific median CFU range used to successfully perform FMT with the donor swhose strains were isolated for the LBP in this study (Figure 4), existing probiotics, and ranges tested in other phase I LBP studies.

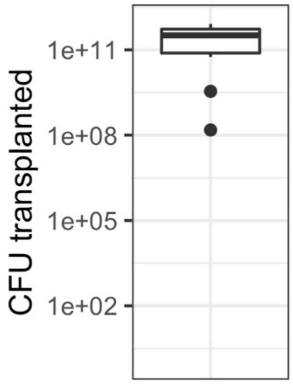

**Figure 4.** Total CFUs transplanted for successful rCDI FMTs using donor material from which MTC01 is derived.

#### 3.10 Main parameters of efficacy

##### *3.10.1 Primary outcome measures*

24. Safety and tolerability of MTC01 determined by incidence of serious adverse events (SAE), adverse events (AE), vital signs, ECG and physical examination up to 24 weeks.

##### *3.10.2 Secondary outcome measures*

1. Recurrence of CDI up to 8 weeks after treatment. Recurrence is defined as positive *C. difficile* toxin EIA as well as diarrhea (>=3 loose stools in 24h period, defined as BSS 6-7, x 2 days) within 8 weeks of receiving FMT.
2. Characterize the engraftment of MTC01 in the fecal microbiome 2 weeks after treatment.
3. Characterize MTC01 related taxonomic changes in the fecal microbiome 2 weeks after treatment.

##### *3.10.3 Other outcome measures*

1. Time to recurrence of CDI (up to 24 weeks).
2. Characterize MTC01 related taxonomic changes and engraftment of MTC01 in the fecal microbiome at 72hr and 1, 8, 12, and 26 weeks after treatment.

#### 3.11 Main parameters of safety

##### *3.11.1 Primary Safety Outcomes Measures*

1. Proportion of participants with an AE through week 8 (± 3 days)
2. Proportion of participants with a SAE through week 8 (± 3 days)

##### *3.11.2 Secondary Safety Outcome Measures:*

3. Proportion of participants with an AE through week 24 (± 5 days)
4. Proportion of participants with a SAE through week 24 (± 5 days)

### 4 CHEMISTRY, MANUFACTURING, AND CONTROL (CMC) INFORMATION

#### 4.1 Description

##### *4.1.1 Identity*

Identity of the Master Cell Bank (MCB) and Working Cell Bank (WCB) is validated by plating an aliquot of the growth media and identification of 10 colonies by MALDI-TOF (Appendix 5 - Table 8). Similarly 100 colonies will be identified by MALDI-TOF for the Drug Substance (DS). Purity, identity and mixing of the microbes in the Drug Product (DP) is validated by plating a proportion of the DP and identification of 200 colonies by MALDI-TOF.

##### *4.1.2 Potency*

To determine the strength of the MCB, WCB, DS and DP, a proportion will be plated on rich media to determine the CFU/mL.

##### *4.1.3 Purity*

Identification of 100 colonies by MALDI-TOF of each DS should the relevant species in MTC01 for high quality spectra. Identification of 200 colonies by MALDI-TOF of the DP should all match only species in MTC01 for high quality spectra. Although the microbes in MTC01 are hypothesized to be strict anaerobes and the cocktail contains no fungi, we are still in the process of determining if any of these MTC01 strains can form colonies in aerobic conditions on Soybean-Casein Digest Agar or Sabouraud Dextrose Agar respectively. Assuming no member of MTC01 forms colonies under these conditions, we will additionally validate purity of the DP and MCB with USP<61>. Alternatively, we will still plate MCB and DP in these conditions and use MALDI-TOF to verify that no colony with high quality spectra matches any bacterium other than those in MTC01.

##### *4.1.4 Stability*

Stability testing will be initiated upon production and will continue for the duration of the study on a monthly basis. Stability of the DS and DP will be determined using CFU counting in combination with a MALDI-TOF biochemical identity test. Preliminary stabilities for select Research Cell Bank (RCB) stocks are shown in Appendix 1 with little to no loss in viability at -80C across approximately 20 wks.

#### 4.2 Summary manufacturing process

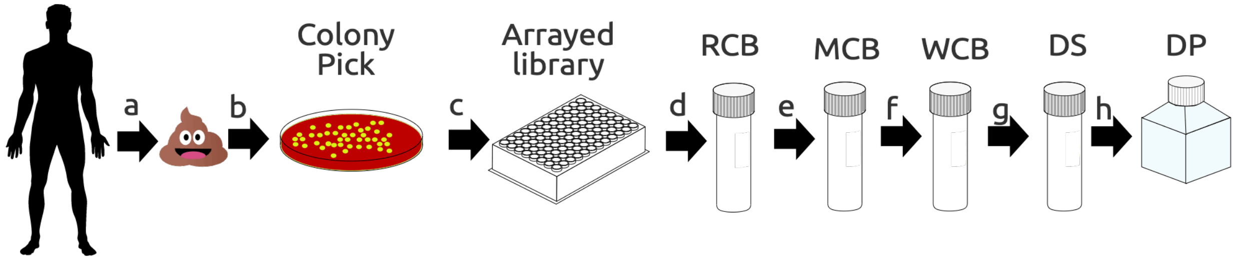

**Figure 5.** Initial isolation and cell banking system.
a) Donor stools were plated under various conditions b) single colonies were picked to generate c) an arrayed culture collection with the unique isolate in each well confirmed by MALDI-TOF, 16S rRNA Sanger sequencing, and genome sequencing d) Research Cell Banks (RCBs) were generated by cryopreservation after colony picking (20 vials per strain) e) Master Cell Banks will be generated from the RCBs by one round of colony picking using agar and broth free of animal products (100 vials per strain) f) Working Cell Banks will be generated from the MCB by inoculation with one cryo vial into 100 mL growth media, harvested after a strain-specific time and resuspended in cryo-preservation media (100 vials per strain) g) Drug Substance (DS) will be generated from the WCBs by inoculation of 250 mL growth media, harvested after a strain specific time and resuspended into 25 mL cryo-preservation media (5x 5 mL); an aliquot will be taken to determine the potency of the DS (CFU/mL) h) The Drug Product (DP) will be generated from the DS by thawing the DS and combining all strains at approximately $3x10^{10-}$CFU per dose for a total dose of approximately $5x10^{11}$ CFU. Manufacturing of the MCB, WCB, DS and DP will all be performed under sterile conditions using a broth free of animal products. DS will be washed and resuspended into USP grade PBS+glycerol+cysteine+glucose.

##### *4.2.1 Initial Strain isolation*

All bacteria were initially isolated from the fecal material of human FMT donors that were used to successfully treat rCDI patients and found to durably engraft for at least 8 weeks as described in section 2.1. The stool samples from the FMT donors were stored at -80$^o$C before processing. Subsequently, the stool sample was pulverized under liquid nitrogen and under strict anaerobic conditions ~500mg of pulverized stool from each donor was blended into a slurry (40 – 50 mg/mL) in pre-reduced LYHBHIv4 media (Appendix 4). The fecal slurries were passed through sterile 100 um strainers to remove large debris. Arrayed culture collections were generated for selected donors as previously described[12,29]. Briefly, clarified and diluted donor stool was plated onto a variety of solid selective and non-selective media under anaerobic, micro-aerophilic and aerobic conditions. Plates were incubated for 48-72 hours at 37$^o$C. 384 single colonies from each donor microbiota were individually picked and regrown in liquid LYHBHIv4 media for 48 hours under anaerobic conditions. Regrown isolates were identified at the species level using a combination of MALDI-TOF mass spectrometry (Bruker Biotyper) and 16S rDNA amplicon sequencing. All regrown isolates were stored in LYHBHIv4 with 15% glycerol at -80$^o$C (Figure 5a-c). All strains for use in MTC01 were genome sequenced with paired-end 150nt reads on an Illumina platform. The solid agar media used for the initial isolation of each strain is listed in Table 2.

**Table 2.** Initial isolation.

| Strain ID | Genus | Species | Media | Passage |
|---|---|---|---|---|
| MTC01.01 | Bacteroides | uniformis | Chocolate Agar | 3 |
| MTC01.02 | Bacteroides | ovatus | Brucella LKV Agar | 3 |
| MTC01.03 | Bifidobacterium | longum | microaerophillic on Columbia CNA w/5% Sheep Blood | 3 |
| MTC01.04 | Bacteroides | thetaiotaomicron | Brucella LKV Agar | 3 |
| MTC01.05 | Bacteroides | vulgatus | Columbia CNA w/5% Sheep Blood | 3 |
| MTC01.06 | Collinsella | aerofaciens | Chocolate Agar | 3 |
| MTC01.07 | Parabacteroides | distasonis | Brucella LKV Agar | 3 |
| MTC01.08 | Bifidobacterium | adolescentis | Chocolate Agar | 3 |
| MTC01.09 | Parabacteroides | merdae | Brucella LKV Agar | 3 |
| MTC01.10 | Coprococcus | comes | Chocolate Agar | 3 |
| MTC01.11 | Eubacterium | rectale | Chocolate Agar | 3 |

| | | | | |
|---|---|---|---|---|
| MTC01.12 | Bacteroides | caccae | Brucella LKV Agar | 3 |
| MTC01.13 | Dorea | longicatena | Columbia CNA w/5% Sheep Blood | 3 |
| MTC01.14 | Odoribacter | splanchnicus | Chocolate Agar | 3 |
| MTC01.15 | Bacteroides | cellulosilyticus | Brucella LKV Agar | 3 |
| MTC01.16 | Bifidobacterium | pseudocatenulatum | Chocolate Agar | 3 |

###### *4.2.2 Preparation of Research Cell Banks (RCB)*

RCBs were generated from the arrayed culture libraries by plating the original arrayed strain on Chocolate agar. The identity of each strain was assayed by MALDI-TOF and a single colony of similar morphology to the test colony was picked to inoculate 11 mL of LYHBHIv4 growth media. After 24-48 hours, 0.5 ml of culture was combined with 0.5 ml of 30% glycerol, for a final volume of 1 ml 15% glycerol, and aliquoted into 20 x 2.0 mL tubes that were then stored at -80$^{o}$C. The strain identity of each RCB was validated from an aliquot of the liquid broth by MALDI-TOF. RCBs are complete. Results are in Appendix 9.

###### *4.2.3 Preparation of Master Cell Banks (MCB)*

All manipulations will take place in a dedicated, sterilized anaerobic chamber with a filtered air supply and sterilization chamber (Appendix 8). The master cell banks will be generated from the RCBs by plating the RCB and picking one colony to inoculate a 10 mL starter culture in LYH + vegitone infusion broth (LYHV; Appendix 4), after 24 hours 2 mL will be used to inoculate one sealable sterile centrifuge tube containing 100 mL LYHV infusion broth and incubated at 37$^{o}$C under anaerobic conditions until within 8hr of peak CFU or 90% of maximum OD600. After reaching 90% of maximum density verified by $OD_{600}$ measurement and validated by CFU, the culture is combined with 100 mL LYHV infusion containing 30% glycerol (final glycerol concentration is 15%) and stored at -80$^{o}$C as 1 ml aliquots in 100 individual air-tight cryovials.

###### *4.2.4 Preparation of Working Cell Banks (WCB)*

One glycerol stock ($10^{7}$-$10^{9}$ CFU in a volume of 1 mL) of the MCB will be thawed and brought into the sterilized anaerobic chamber and cultured in the same manner as the MCB. After reaching 90% of maximum density verified by $OD_{600}$ measurement and validated by CFU, the culture is combined with 100 mL LYHV infusion containing 30% glycerol (final glycerol concentration is 15%) and stored at -80$^{o}$C as 1 ml aliquots in 100 individual air-tight cryovials.

###### *4.2.5 Drug substance (DS)*

One glycerol stock ($10^{7}$-$10^{9}$ cfu in a volume of 1 mL) of the WCB will be thawed and brought into the sterilized anaerobic chamber. Each strain will be inoculated into 25 mL of LYHV infusion broth and grown 24hr. From this starter culture, 5 mL will be used to inoculate 4 x 250 mL sterile LYHV infusion broth media in sealable, sterile centrifuge bottles and incubated at 37$^{o}$C under anaerobic conditions until within 8hr of peak CFU. After reaching 90% of maximum growth density verified by $OD_{600}$ measurement and validated by CFU, the culture is centrifuged and washed twice with GMP grade PBS-cysteine to remove bacterial bioproducts and the original culture media. The final washed bacteria will be resuspended in USP grade PBS-cysteine-glycerol media at 10X concentration and stored as 9 x 10 mL aliquots at -80$^{o}$C. 20 x 0.5 mL aliquots will be stored at -80$^{o}$C for DS stability assays.

###### *4.2.6 Drug product (DP)*

After verifying each strain grew sufficiently dense to meet our CFU boundary conditions, we will thaw all strains and combine them in proportion to their CFU. The final suspension will be $3x10^{10}$ of each strain in 125ml of reduced PBS+glycerol+cysteine+glucose and stored at -20 or -80C (depending on pending stability assays).

###### *4.2.7 Cell Bank System*

The MCB, WCB, DS and DP will be stored in barcoded cryo-vials or containers in an access restricted -80°C. All aliquots and subaliquots will be tracked using FreezerWorks.

##### 4.3 Description of manufacturing facility

###### *4.3.1 Personnel*

All personnel have the education, experience, and training to prepare the phase 1 investigational drug and will follow detailed SOPs detailing the QC principles and acceptable methods to comply with the statutory requirement of cGMP. One individual will be responsible for manufacturing, a second individual will perform QC functions and a third and independent individual will perform periodic review of the manufacturing and QC functions. To assist with SOP design and general expertise on cGMP, Marcia Meseck, MS, JD will serve as a consultant. Marcia is the director of the Vaccine and Cell Therapy laboratory GMP facility at Mount Sinai. She has over 25 years of cGMP experience and has established multiple cGMP facilities.

###### *4.3.2 Facility and equipment*

The facility for the phase I drug substance (DS) and drug product (DP), as well as the MCB and WCB, will be built at the Icahn School of Medicine at Mount Sinai in New York. The facility will use sterilized anaerobic chambers with an air exchange and disinfection chamber to culture the drug substance and formulate the drug product as a 2x washed bacterial consortium in USP grade PBS with glycerol + glucose and a cysteine reducing agent to make the solution anaerobic (Table 3,

Figure **6**). Written SOPs detail gowning/de-gowning, environmental monitoring, disinfection/cleaning and security/access control protocols.

**Table 3.** Major facility equipment.

| *Equipment* | *Purpose* | *Vendor* |
|---|---|---|
| *Shelves* | *Store supplies* | |
| *Fridge (4°C)* | *Store supplies* | |
| *Freezer (-80°C)* | *Store supplies* | |
| *Lab bench* | *Work area* | |
| *Centrifuge* | *Fluidic exchange* | *Beckman* |
| *Anaerobic chamber* | *Fluidic exchange* | *Coy* |
| *Freezer (-80°C)* | *Freeze bacteria* | |

**Figure 6.** LBP manufacturing facility.

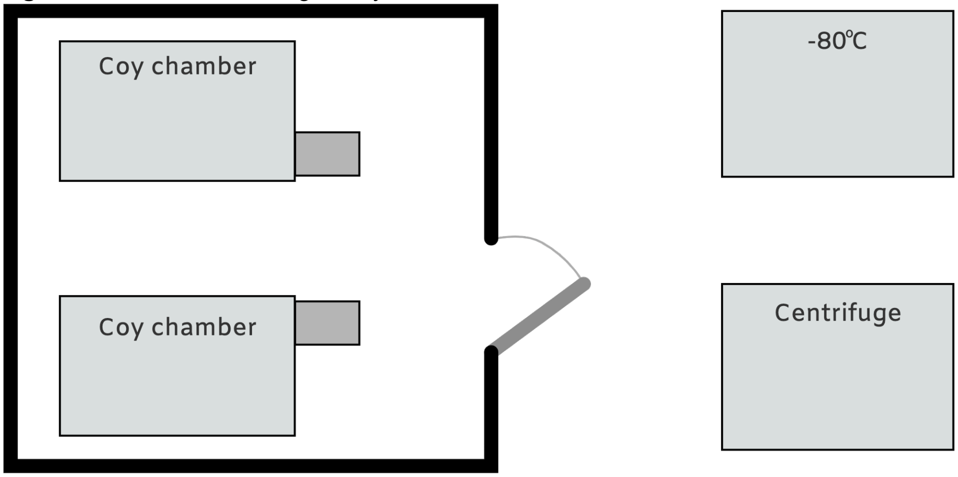

The facility will consist of a 200 square feet lab with two sterile anaerobic chambers and outside the main culture facility the centrifuge and -80°C where sealed, sterilizable containers will be transported for centrifugation and storage respectively.

#### 4.4 Placebo

The proposed phase I study will not have a placebo group.

### 5 STUDY DESIGN

This is an open-label pilot study to measure the safety and microbiological and clinical impacts of a defined LBP in patients with recurrent CDI. We will prospectively enroll 20 patients with recurrent CDI from one tertiary care referral center. A visual overview of the study design is in Appendix 6. The study details are below.

#### 5.1 Visits

##### *5.1.1 Visit 1 - Screening*

Potential subjects will undergo the following screening procedures no more than 4 weeks prior to administration of the LBP to determine if they meet the recipient selection criteria.

1. Medical record review will be done to confirm diagnosis and inclusion criteria. Records will be reviewed for exclusion criteria, however if not available patient self-report will be accepted. The following data will be documented at baseline visit:
    a. Demographic: age, height, gender, weight, race, significant past medical history, and smoking status.
    b. Diarrheal symptom assessment: Average daily Bristol score, number of daily BMs.
    c. CDI history: inciting antibiotic, Number and approximate date of previous CDI episodes, previous positive stool tests—dates, PCR versus EIA, CDI treatment courses (metronidazole, vancomycin, vancomycin taper, or fidaxomicin), CDI related prior hospitalizations and number of CDI related hospitalizations.
2. Baseline symptom assessment using diary and targeted physical exam by study physician or nurse
3. Laboratory assessments:

   a. Blood: CBC, CRP, CMP, HIV, Viral hepatitis, Syphilis, and ONLY subjects who are heavily immunosuppressed will also be screened for CMV and EBV.
   b. Stool: Fecal calprotectin, microbiome analysis. Stool will be collected on chronic anti-CDI therapy.
5. Stool and blood will be banked for metabolomics and future analysis.
6. Urine Pregnancy Test if applicable. If patients are post-menopausal this will be documented and pregnancy test is not needed
7. If not currently on anti-CDI therapy Vancomycin should be started for a minimum of 4 days prior to LBP dosage with instruction to hold for 48 hours prior to LBP dosage.
8. Once patient has passed screening and all labs have been reviewed colonoscopy will be scheduled.

##### *5.1.2 Visit 2 – LBP dosing*

The following baseline assessments will be made in enrolled subjects on day of scheduled LBP treatment.

1. Urine pregnancy test (HCG) for female patients if applicable. If patients of post-menopausal this will be documented, and pregnancy test is not needed.

##### *5.1.3 Visit 3 - Study phone call*

1. AE assessment 72 hours post LBP +/- 1 day

##### *5.1.4 Visit 4, 5, and 6 – Week 1, 8 and 12 post LBP*

Week 1, 8, 12 +/- 3 days post LBP: patients will be evaluated in the clinic for follow-up assessments. The following will be documented:

1. Assessment of efficacy: Assessment of diarrheal symptoms as well as testing for CDI by GDH/EIA and PCR regardless of symptoms at week 1, 8 and 12 post LBP dosage.
2. Assessment for related AEs to LBP will occur at each visit using NIH criteria. Related AEs include but are not limited to:
   a. Diarrhea
   b. Bloating, distention
   c. Constipation
   d. Abdominal pain
   e. bacteremia
   f. infection transition
   g. headache
   h. nausea
   i. pyrexia
3. Laboratory assessments:
   a. Stool: CDI testing (as above), fecal calprotectin, microbiome analysis.
   b. Stool and blood will be banked for metabolomics and future analysis.
4. Vital Signs and targeted physical exam
5. List of current concomitant medications

##### *5.1.5 Visit 7 – Week 26*

Visit 7 (Week 26 +/- 7 days post LBP): Study Phone Call. Patients will be called to assess the following:

1. AE assessment
2. Patients will mail in samples for microbiome analysis only

Patient obligations in the study will end at week 26 however the patient's medical record will be followed prospectively for evaluation of CDI outcomes and procedures done for clinical care for 1-year post-enrollment.

##### *5.1.6 Early Termination Visit:*

In the case of an early termination, study staff will complete an 'Early Termination CRF', if possible, and the following will be assessed:

1. Assessment of efficacy: Assessment of diarrheal symptoms as well as testing for CDI by GDH/EIA and PCR regardless of symptoms
2. Assessment for related AEs to LBP will occur at each visit using NIH criteria. Related AES include but are not limited to:
    a. Diarrhea
    b. Bloating, distention
    c. Constipation
    d. Abdominal pain
    e. bacteremia
    f. infection transition
    g. headache
    h. nausea
    i. pyrexia
3. Laboratory assessments:
    a. Blood: CBC, CRP, CMP
    b. Stool: CDI testing (as above), fecal calprotectin, microbiome analysis.
    c. Stool and blood will be banked for metabolomics and future analysis
4. Vital Signs and targeted physical exam.

##### *5.1.7 Unscheduled Visit*

At any point during the study if patients experience worsening of symptoms they may be brought in by study staff for an unscheduled visit for an assessment. This visit will include:

1. Assessment of efficacy: Assessment of diarrheal symptoms as well as testing for CDI by GDH/EIA and PCR regardless of symptoms at week 1, 8 and 12 post LBP dosage.
2. Assessment for related AEs to LBP dosage will occur at each visit using NIH criteria. Related AEs include but are not limited to:
    a. Diarrhea
    b. Bloating, distention
    c. Constipation
    d. Abdominal pain
    e. bacteremia
    f. infection transition
    g. headache

   h. nausea
   i. pyrexia
3. Laboratory assessments:
   a. Blood: CBC, CRP, CMP (to be done locally)
   b. Stool: CDI testing (as above), fecal calprotectin, microbiome analysis.
4. Stool and blood will be banked for metabolomics and future analysis
5. Vital Signs and targeted physical exam
6. List of current concomitant medications

#### 5.2 Patients Treatment Strategy

The below treatment strategy will be utilized based on results obtained at each visit.

**Table 4.** Treatment strategy

| Symptoms | Step 1: PCR | Step 2: EIA | CDI recurrence | Treatment Course |
|---|---|---|---|---|
| Diarrhea | + | + | Yes | Anti-CDI Therapy including antibiotics or FMT |
| Diarrhea | + | - | No | Likely colonization, treatment at Clinician discretion |
| Diarrhea | - | - | No | No CDI, evaluate for other causes of diarrhea and treat accordingly |
| No Diarrhea | + | + | No | Clinician discretion |
| No Diarrhea | + | - | No | Asymptomatic Carriage, No anti-cdi tx needed |
| No Diarrhea | - | - | No | No treatment needed |

#### 5.3 Recruitment Procedures

Gastroenterologists, including attending physicians and fellows, will be informed of the study's aims and inclusion criteria. These doctors will inform the principal investigator or site lead investigators of patients who meet the study criteria and who may be good candidates for the study. The treating gastroenterologist will introduce the study to the potential patient and request the patient's permission to be approached by study staff. We will not utilize advertisement material or other informational materials for patients beyond the consent form and the posting on clinicaltrials.gov. Patients will be given as much time as they need to decide. They will be given a copy of the consent form to take home, read, and consider, and they will be encouraged to discuss participation with family members and health care providers.

The trial will be posted on clinicaltrials.gov. If patients reach out to us via the provided E-mail or phone number located at the clinicaltrials.gov website, or directly reach out to the study team via any mechanism, we will pre-screen them using questions from the provided E-mail/phone script. If the patient's responses do not indicate any exclusion criteria being met, then we will contact their primary care provider or GI provider via email or phone call (with patient permission) to confirm medical diagnosis. If no records exist for exclusion criteria patient report will be accepted.

#### 5.4 Consent Procedures

The treating gastroenterologist will introduce the study to the potential patient and request the patient's permission to be approached by study staff. With the treating gastroenterologist's permission, either a physician investigator or study coordinator will describe the research study in detail, including participation and risks and alternative courses of treatment, and answer any questions or concerns that the patient may have. Patients will be given as much time as they need to consider participation before signing the consent form.

Subjects will be recruited from the Susan and Leonard Feinstein IBD Clinical Center. In order to avoid coercion, study staff will reinforce that participation is voluntary and that their decision will not affect the medical care that they receive now or in the future. If patients seek more time to consider participation, they will be given a copy of the consent form and encouraged to discuss the study with family, friends, PCP, or others. Study staff will follow-up to see if any questions or concerns have not been addressed. A physician investigator will obtain informed consent signatures.

If patients initiate contact with the study team and the initial phone screen suggests eligibility they will be provided with the consent form via email to review. They will be re-contacted in a few days so that any questions may be answered. If they would like to proceed, a screening visit will be set up.

#### 5.5 Specimen Handling and Shipping

All patients will be assigned a unique participant ID. Clinical samples will be processed at the Microbiome Translation Center at the Icahn School of Medicine at Mount Sinai. Additionally, blood and stool samples will be aliquoted and banked.

#### 5.6 Assessment of safety

Safety will be assessed by the frequency and severity of adverse events (AE)

##### *5.6.1 Definition of an Adverse events (AE)*

Adverse events (AEs) will be recorded at each regular scheduled study visit in the study patient record (source document) as well as on a specific AE CRF.

An AE is any untoward medical occurrence in a study patient or clinical investigation subject administered a pharmaceutical product and which does not necessarily have a causal relationship with this treatment. An AE can therefore be any unfavorable and unintended sign (including an abnormal laboratory finding), symptom, or disease temporally associated with the use of a medicinal product, whether or not related to the medicinal product, e.g.:

- any new clinical diagnosis
- any symptom that requires medical clarification or leads to in-patient admission (surgery or accident)
- any suspected adverse drug reaction (ADR)
- any symptom that appears on the study patient’s medical records
- any event related in time with the application of the study medication and affecting the health of the study patient (including laboratory value changes)

If there is any doubt as to whether a clinical observation is an AE, the event should be reported. AEs must be graded for severity and relationship to study product.

##### *5.6.2 NIH Grading of Severity of the Event*

AEs will be assessed by the clinician using the NIH protocol defined grading system (see Appendix). Briefly, the criteria for estimating adverse event severity grade:

- Grade 1 (Mild): events require minimal or no treatment and do not interfere with the patient’s daily activities.

- Grade 2 (Moderate): events result in a low level of inconvenience or concern with the therapeutic measures. Moderate events may cause some interference with functioning.
- Grade 3 (Severe): events interrupt a patient's usual daily activity.
- Grade 4 (Potentially life threatening): Events result in inability to perform basic selfcare functions or the need for medical or surgical intervention to prevent permanent disability or death.
- Grade 5 (Death)

##### *5.6.3 NIH Adverse Event Relatedness*

The clinician's assessment of an AE's relationship to test LBP is part of the documentation process, but it is not a factor in determining what is or is not reported in the study. If there is any doubt as to whether a clinical observation is an AE, the event should be reported.

The following NIH guidelines of relatedness are used:

- Related: The adverse event is related to the LBP material – i.e. an event that follows a reasonable temporal sequence from administration of the LBP material, follows a known or expected response pattern to the LBP material, that is confirmed by improvement on stopping and reappearance of the event on repeated exposure and that could not be reasonably explained by the known characteristics of the patient's clinical state.
- Possibly Related: The adverse event follows a reasonable temporal relation to LBP administration, however, symptom may be related to other factors.
- Not Related – The adverse event is not related to the LBP material. - i.e. another cause of the event is most plausible; and/or a clinically plausible temporal sequence is inconsistent with the onset of the event and the study intervention and/or a causal relationship is considered biologically implausible

##### *5.6.4 Solicited mild to moderate adverse events:*

In addition to open-ended questions on adverse events meeting the above definitions, specific potential adverse events will be inquired about during the follow up period:

| Symptom that is clinically more severe than participant's baseline | Severity | | | | |
|---|---|---|---|---|---|
| | Grade 1 | Grade 2 | Grade 3 | Grade 4 | Grade 5 |
| Fever* | 38.0 - 39.0 degrees C (100.4 - 102.2 degrees F) | >39.0 - 40.0 degrees C (102.3 - 104.0 degrees F) | >40.0 degrees C (>104.0 degrees F) for <=24 hrs | >40.0 degrees C (>104.0 degrees F) for >24 hrs | Death |
| Diarrhea | Increase of <4 stools per day over baseline pre-LBP; mild increase in ostomy output compared to baseline | Increase of 4 - 6 stools per day over baseline; moderate increase in ostomy output compared to baseline; limiting instrumental ADL | Increase of >=7 stools per day over baseline; incontinence; hospitalization indicated; severe increase in ostomy output compared to baseline; limiting self-care ADL | Life-threatening consequences; urgent intervention indicated | Death |
| Vomiting | 1 - 2 episodes (separated by 5 minutes) in 24 hrs | 3 - 5 episodes (separated by 5 minutes) in 24 hrs | >=6 episodes (separated by 5 minutes) in 24 hrs; tube feeding, TPN | Life-threatening consequences; urgent | Death |

| | | | | | |
|---|---|---|---|---|---|
| | | | or hospitalization indicated | intervention indicated | |
| Abdominal Pain | Mild pain | Moderate pain; limiting instrumental activities of daily life | Severe pain; limiting self care activities of daily life | n/a | n/a |
| Bloating | No change in bowel function or oral intake | Systemic, decreased oral intake; change in bowel function | n/a | n/a | n/a |
| Constipation | Occasional or intermittent symptoms; occasional use of stool softeners, laxatives, dietary modification, or enema | Persistent symptoms with regular use of laxatives or enemas indicated; limiting instrumental activities of daily life | Symptoms interfering with self-care activities of daily life; obstipation with manual evacuation indicated | Life-threatening consequences (e.g. obstruction, toxic megacolon); urgent intervention indicated | Death |

##### 5.6.5 Serious Adverse Events

- An adverse event or suspected adverse reaction is considered "serious" if, in the view of either the investigator or sponsor, it results in any of the following outcomes:
- Death;
- Life-threatening adverse event*;
- Inpatient hospitalization or prolongation of existing hospitalization;
- A congenital anomaly/birth defect;
- Persistent or significant disability or incapacity or substantial disruption of the ability to conduct normal life function.

**Life-threatening adverse event. An adverse event is considered "life-threatening" if, in the view of either the investigator or sponsor, its occurrence places the patient or subject at immediate risk of death. It does not include an adverse event which, had it occurred in a more severe form, might have caused death.*

Important medical events that may not result in death, be life-threatening, or require hospitalization may be considered serious when, based upon appropriate medical judgment, they may jeopardize the patient or subject and may require medical or surgical intervention to prevent one of the outcomes listed in this definition

Any adverse event or suspected adverse reaction that meets the criteria for serious adverse event will be:

- recorded on the appropriate SAE CRF
- followed through resolution by a study clinician
- reviewed and evaluated by a study clinician

##### 5.6.6 Unsolicited Adverse Events

On enrollment in the study, the study participants will be instructed to contact the site PI if an AE occurs. All unsolicited non-serious adverse events will be collected from the time of LBP dosage until 6 months following LBP dosage and will be assessed for relatedness as outlined in section 5.6.3. Patients will be given a patient diary with date, time, details and action taken to help with data collection. Patients will bring this diary to the site PI for evaluation at each follow-up visit and will be instructed to seek immediate medical attention if indicated.

###### *5.6.7 New-Onset Related Chronic Medical Condition*

Study LBP-related chronic medical conditions occurring from the time of the LBP dosage until 6 months following LBP dosage. Specifically, new-onset chronic medical conditions potentially related to LBP dosage such as weight gain, glucose intolerance, autoimmune conditions, and metabolic syndrome will be monitored for. This is be done via patient report.

##### 5.7 Reporting of Adverse Events

Study participants will be instructed to contact the study nurse or doctor if any serious or unexpected adverse event occurs. Study staff will enquire, about using a generally worded question, about AEs at each study visit. Reported AE's will be recorded in detail in an AE CRF.

AE information to be collected in the AE CRF:

- Nature of the event
- Time of onset: date, time
- Concomitant treatment: product (generic name), indication, dosage, dosage interval, presentation, mode of administration, administration regimen
- Duration of the AE
- Severity
- Seriousness
- Causality
- Outcome

The course and outcome of the adverse event will be commented on as follows:

- Recovered without sequelae
- Not yet recovered
- Recovered with sequelae
- Fatal

Any SAE (including death, irrespective of the cause) occurring during the study will be immediately reviewed by the PI, i.e. within 24 hours and referred to the DSMB. In case of a SAE, the information will be reviewed by the PI and reported to the DSMB chair. If the SAE is judged by the DSMB to be related to the treatment, a report will be sent to the IRB of the site. The report must contain a detailed description of the symptoms observed and the concomitant treatment administered. Furthermore, the investigator must comment on a possible causative relationship between the AE and the trial medication. Each SAE must be followed until it is resolved or can be explained satisfactorily.

For non-serious adverse reactions the site PI will complete and submit a report to the lead PI. All non-serious adverse reactions will be reviewed by the DSMB at their regular meeting and or ad/hoc depending on the clinical case at the discretion of the site PI and lead PI.

In accordance with safety requirements, the study PI will inform the local IRB and will make sure that the involved persons will obtain adequate information. The following instructions must be heeded:

- In the case of an intolerable SAE, the study patient must, at the decision of the investigator, be withdrawn from the clinical trial, and symptomatic treatment must be administered.
- The measures taken must be recorded on the CRF.
- In accordance with local legislation, the investigators will submit copies of the final SAE-report to the Regulatory Authorities concerned, if necessary.

#### 5.8 Follow-up of Subjects after Adverse Events

AEs will be followed until resolution or stability even if this extends beyond the study-reporting period. Resolution of an AE is defined as the return to pretreatment status or stabilization of the condition with the expectation that it will remain chronic.

Follow-up procedures, evaluations, and outcomes will be recorded on the subject's case report forms.

#### 5.9 Safety Oversight

##### *5.9.1 Data and Safety Monitoring Board (DSMB)*

Safety oversight will be under the direction of a DSMB. The DSMB is an independent group of experts who will advise the study investigators. The primary responsibilities of the DSMB are to 1) periodically review and evaluate the accumulated study data for subject safety, study conduct and progress, and, when appropriate, efficacy, and 2) make recommendations concerning the continuation, modification, or termination of the trial. The DSMB will be composed of at least 3 voting members. The membership will include a chairperson with prior DSMB experience. There will also be members with clinical expertise in the medical area and subject population being studied. All DSMB members will be separate and independent of study personnel participating in this trial and should not have scientific, financial or other conflict of interest related to the trial. Procedures for DSMB data reviews will be defined in the DSMB Charter that will include DSMB membership, responsibilities, and the scope and frequency of data reviews. The study should be reviewed by the DSMB at least annually otherwise will plan to meet after 10 patients are enrolled.

#### 5.10 Halting Rules

##### *5.10.1 Study enrollment halting rules*

Enrollment in the study will be suspended for conduct of a safety review by the DSMB in the case of:

- A Grade 3 AE of the same organ system deemed related to the study intervention in three or more of the participants.
- Any serious adverse event of an enrolled participant related to the study intervention, including transmission of a pathogen from LBP to recipient.
- An overall pattern of symptomatic, clinical, or laboratory events that the lead PI considers related to study product and that may appear minor in terms of individual events, but that may collectively represent a serious potential concern for safety.

###### *5.10.2 Individual's halting rules*

Subjects who meet any of the following criteria must be assessed by the PI to determine if it is in the subject's best interest to stop the study product(s):

- Subject choice (Withdrawal of consent)
- Participant's non-compliance.
- Development of a significant medical condition and/or participation in the study is no longer in the best interest of the subject.

##### 5.11 Statistical considerations

This is a pilot study, open-label study to determine the safety of LBP in patients with recurrent CDI. Preliminary efficacy/outcome data will also be collected. This study is exploratory in nature so a formal sample size calculation has not been performed. Our goal is to recruit 20 subjects. We believe this will allow us to assess safety, acceptability and engraftment.

##### 5.12 Source documents and access to data/documents

The PI will maintain appropriate medical and research records for this trial, in compliance with institutional requirements for the protection of confidentiality of subjects. Forms for use as source documents will be derived from the electronic CRFs. Original documents and data records include, but are not limited to, hospital records, clinical and office charts, laboratory notes, memoranda, pharmacy dispensing records, recorded data from automated instruments, copies or transcriptions certified after verification as being accurate and complete, microfiches, photographic negatives, microfilm or magnetic media, x-rays, and subject files and records kept at the pharmacy, laboratories, and medico-technical departments involved in the clinical trial. Original source documents will be maintained by each site.

##### 5.13 Quality control and quality assurance

The PI and study coordinators are responsible for conducting routine quality assurance (QA) and quality control (QC) activities to internally monitor study progress and protocol compliance. The PI will have direct access to source data/documents and reports for the purpose of monitoring. Additionally, auditing by local and regulatory authorities will occur at their discretion. The PI will ensure all study personnel are appropriately trained and applicable documentations are maintained on site.

The Principal Investigator, Jeremiah Faith, will assure the validity and integrity of the data and adherence to the IRB-approved protocols.

Study staff will review completed CRF's before each visit to ensure completeness of previous entries. Entries that need clarification will be reviewed by the PI, and the subject and/or treating gastroenterologist will be consulted if needed.

##### 5.14 Ethics/Protection of human subjects

###### *5.14.1 Ethical Standard*

The PI will ensure that this study is conducted in full conformity with principles of the Belmont Report: Ethical Principles and Guidelines for the Protection of Human Subjects of Research of the National Commission for the Protection of Human Subjects of Biomedical and Behavioral Research (April 18, 1979) and codified in 45 CFR 46, 21 CFR

50 and 56, and ICH E6; 62 Federal Regulations 25691 (1997), if applicable. The investigator's Institution will hold a current Federal Wide Assurance (FWA) issued by the Office of Human Research Protection (OHRP) for federally funded research if applicable.

###### *5.14.2 Institutional Review Board (IRB)*

The study investigators will provide for the review and approval of this protocol and the associated informed consent documents, by an appropriate ethics review committee or IRB. Any amendments to the protocol or consent materials must also be approved before they are placed into use unless change is for the safety of the subject. Only those IRB members who are independent of the investigators should provide an opinion on study related matters. Verification of IRB approval of the protocol and the written informed consent will be transmitted by the investigator or designee prior to the shipment of clinical trial material. No deviations from or changes to the protocol will be initiated without prior approval of an appropriate amendment unless change is for the safety of the subject. The study investigators are responsible for ensuring Continuing Review at least once a year and for keeping the IRB apprised of the progress of the study and any changes to the protocol.

###### *5.14.3 Informed Consent Process*

The written consent document will embody the elements of informed consent as described in the Declaration of Helsinki and will adhere to the ICH Harmonized Tripartite Guideline for Good Clinical Practice. Informed consent should be implemented before any protocol-specified procedures or interventions are carried out. Informed consent will be obtained in accordance with 21 CFR 50.25 and 45 CFR 46. Information should be presented both orally and in written form.

An investigator or designee will describe the protocol to potential subjects face-to-face. The Subject Information and Consent Form may be read to the subjects, but, in any event, the investigator shall give the subjects ample opportunity to inquire about details of the study and ask any questions before the signing and dating the consent form.

Study staff must inform subjects and/or legal guardian that the trial involves research, and explain the purpose of the trial, those aspects of the trial that are experimental, any expected benefits, all possible risks (including a statement that the particular treatment or procedure may involve risks to the subject or to the embryo or fetus, if the subject is or may become pregnant or fathers a child, that are currently unforeseeable), the expected duration of the subject's participation in the trial, the procedures of the research study, including all invasive procedures, and the probability for random assignment to treatment groups. Subjects and/or legal guardian will be informed that they will be notified in a timely manner if information becomes available that may be relevant to their willingness to continue participation in the trial. They must also be informed of alternative procedures that may be available, and the important potential benefits and risks of these available alternative procedures. Subjects and/or legal guardian must receive an explanation as to whether any compensation and any medical treatments are available if injury occurs, and, if so, what they consist of, or where further information may be obtained. Subjects and/or legal guardian must be informed of the anticipated financial expenses, if any, to the subject for participating in the trial, as well as any anticipated prorated payments, if any, to the subject for participating in the trial. They must be informed of whom to contact (e.g., the investigator) for answers to any questions relating to the research project. Information will also include the foreseeable circumstances and/or reasons under which the subject's participation in the trial may be terminated. The subjects and/or legal guardian must be informed that participation is voluntary and that they are free to withdraw from the study for any reason at any time without penalty or loss of benefits to which the subject is otherwise entitled.

Neither the investigator, nor the trial staff, should coerce or unduly influence a subject to participate or continue to participate in the trial. The extent of the confidentiality of the subjects' records must be defined, and subjects must be informed that applicable data protection legislation will be followed. Subjects and/or legal guardian must be informed that the monitor(s), auditors(s), IRB, and regulatory authority(ies) will be granted direct access to the subject's medical records for verification of trial procedures and/or data without violating the confidentiality of the subject, to the extent permitted by the applicable laws and regulations, and that, by signing a written informed consent form, the subject is authorizing such access. Subjects and/or legal guardian must be informed that records identifying the subject will be kept confidential, and, to the extent permitted by the applicable laws and/or regulations, will not be made publicly available and, if the results of the trial are published, the subject's identity will remain confidential.

Consent forms must be in a language fully comprehensible to the prospective subjects. Informed consent shall be documented by the use of a written consent form approved by the IRB and signed and dated by the subject and the person who conducted the informed consent discussion. The signature confirms that the consent is based on information that has been provided and all questions have been answered to the prospective subject's satisfaction. Each subject's signed informed consent form must be kept on file by the investigator for possible inspection by Regulatory Authorities and/or the sponsor and Regulatory Compliance persons. The subject should receive a copy of the signed and dated written informed consent form and any other written information provided to the subjects, and should receive copies of any signed and dated consent form updates and any amendments to the written information provided to subjects.

###### *5.14.4 Exclusion of Women, Minorities, and Children (Special Populations)*

Children are excluded for safety reasons.

###### *5.14.5 Subject Confidentiality*

Subject confidentiality is held strictly in trust by the participating investigators, their staff, and the sponsor and their agents. This confidentiality is extended to cover testing of biological samples in addition to the clinical information relating to participating subjects.

The study protocol, documentation, data, and all other information generated will be held in strict confidence. No information concerning the study or the data will be released to any unauthorized third party without prior written approval from the sponsor.

The study monitor or other authorized representatives of the sponsor and FDA may inspect all documents and records required to be maintained by the Investigator, including, but not limited to, medical records (office, clinic, or hospital) and pharmacy records for the subjects in this study. The clinical study site will permit access to such records.

##### 5.14.6 Study Discontinuation

The PI has the right to terminate this study or an individual site's participation at any time. Reasons for terminating the study may include, but are not limited to, the following:

- Incidence or severity of adverse events indicates a potential health hazard;
- Data recording is inaccurate or incomplete;
- Investigator does not adhere to the protocol or applicable regulatory guidelines in conducting the study.

##### 5.14.7 Future Use of Stored Specimens

Any leftover blood and stool specimens will be stored and may be used for future research, under a future protocol, to learn more about fecal transplant in patients with rCDI. These specimens will be stored indefinitely at Mount Sinai Hospital after the study is completed for future testing. In the informed consent document, subjects will be given an opportunity to choose whether or not their de-identified barcoded specimens are stored for future use. For subjects who choose not to allow storage of their samples for future use, these samples will be destroyed at the end of the study. All proposed research projects will be subject to approval by an IRB prior to release of any specimens. No human genetic tests will be performed on specimens.

There are no benefits to subjects in the collection, storage and subsequent research use of specimens. Reports about future research done with subject's samples will NOT be kept in their health records, but subject's samples may be kept with the study records or in other secure areas. Subjects can decide if they want their samples to be used for future research or have their samples destroyed at the end of the study. A subject's decision can be changed at any time before the end of the study by notifying the study doctors or nurses in writing. However, if a subject consents to future use and some of their blood and stool has already been used for research purposes, the information from that research may still be used.

De-identified samples and associated meta-data may be shared with other investigators at other institutions for academic purposes or industry collaborators pending an approved protocol. Each sample will be encoded (labeled) only with a barcode and a unique tracking number to protect subject's confidentiality.

Research using stored specimens may be conducted by other institutions. Any specimens and data provided to the receiving-institution will be coded. Unequivocally, neither individual personal identifiers nor the key linking coded data to individuals will be released to the receiving-institution.

##### 5.14.8 Data handling and record keeping

The investigator is responsible to ensure the accuracy, completeness, legibility, and timeliness of the data reported. All data collection forms should be completed in a neat, legible manner to ensure accurate interpretation of data. Black ink is required to ensure clarity of reproduced copies. When making changes or corrections, cross out the original entry with a single line, and initial and date the change. Do not erase, overwrite, or use correction fluid or tape on the original.

Copies of the electronic CRF (eCRF) will be provided for use as source documents and maintained for recording data for each subject enrolled in the study. Data reported in the eCRF derived from source documents should be consistent with the source documents or the discrepancies should be explained.

##### *5.14.9 Data Management Responsibilities*

All source documents and laboratory reports must be reviewed by the clinical team and data entry staff, who will ensure that they are accurate and complete. Adverse Events must be graded, assessed for severity and causality, and reviewed by the site Principal Investigator or designee.

Data collection is the responsibility of the trial staff at the site under the supervision of the site Principal Investigator. During the study, the investigator must maintain complete and accurate documentation for the study.

##### *5.14.10 Data Capture Methods*

Clinical data (including AEs, concomitant medications, and solicited events data) and clinical laboratory data will be entered into a compliant Internet Data Entry System. The data system includes password protection and internal quality checks, such as automatic range checks, to identify data that appear inconsistent, incomplete, or inaccurate. Clinical data will be entered directly from the source documents.

##### *5.14.11 Types of Data*

Data for this study will include clinical, safety and microbiological outcome measures.

##### *5.14.12 Timing/Reports*

Interim reports for the DSMB will be prepared when approximately 40% and 70% of subjects complete enrollment. Interim statistical reports may be generated as deemed necessary and appropriate by the study PI. Other safety summary reports may be generated for the DSMB. A final report will be prepared following the availability of all the clinical, safety and efficacy data.

##### *5.14.13 Study Records Retention*

Study files (except for future use consent forms) must be maintained for a minimum of two years after the last approval. These documents should be retained for a longer period, however, if required by local regulations. No records will be destroyed without the written consent of the sponsor, if applicable. It is the responsibility of the sponsor to inform the investigator when these documents no longer need to be retained. Consent forms for future use will be maintained as long as the sample exists.

##### *5.14.14 Protocol Deviations*

A protocol deviation is any noncompliance with the clinical trial protocol, GCP, or protocol-specific MOP requirements. The noncompliance may be either on the part of the subject, the investigator, or the study site staff. As a result of deviations, corrective actions are to be developed by the site and implemented promptly.

It is the responsibility of the site PI/study staff to use continuous vigilance to identify and report deviations within five working days of identification of the protocol deviation, or within five working days of the scheduled protocol-required activity. All deviations must be promptly reported to the study PI or designated personnel at MSH.

All protocol deviations, as defined above, must be addressed in study subject source documents. A completed copy of the Protocol Deviation Form must be maintained in the Regulatory File, as well as in the subject's source document. Protocol deviations must be sent to the local IRB per their guidelines. The site PI/study staff is responsible for knowing and adhering to their IRB requirements.

#### 5.15 Potential Risks and Benefits

##### *5.15.1 Risks*

###### *5.15.1.1 LBP*

Some of the known risk associated with FMT are still possible with this LBP such as:

- Altered bowel pattern (diarrhea, constipation)
- Cramping
- Belching

Potential risks of LBP dosage:

- Inclusion of strains with previously undescribed harmful virulence factors
- Alteration in intestinal metabolism and potential drug-drug interactions
- Transmission of allergens

###### *5.15.1.2 Privacy and Confidentiality*

This study involves the collection of personal health information. Accidental release of personal health information is a risk of participation in this study. Measures will be taken to protect the confidentiality of all subjects' information. These measures include keeping all information collected about the subjects' confidential, keeping information in locked rooms, and having physicians who are directly involved with a subject's clinical care involved in the study.

###### *5.15.1.3 Colonoscopy*

Standard potential risks of the endoscopy procedure include discomfort, gastrointestinal bleeding either related or unrelated to biopsies, intestinal perforation, altered bowel habit. Complications of IV conscious sedation during the procedure include respiratory arrest, medication reactions, and aspiration.

###### *5.15.1.4 Venipuncture:*

Risks of having blood drawn include pain, bruising, or infection.

###### *5.15.1.5 Pregnancy*

The risks to fetuses and women who are pregnant are unknown. We will not be enrolling any pregnant of lactating women.

##### *5.15.2 Potential Benefits*

The potential benefits include:

- Treatment of recurrent CDI
- Improvement in clinical symptoms
- Restoration of fecal diversity

As this is a pilot study, it is difficult to quantify the expected benefits.

### REFERENCES

1. Lessa, F. C. *et al.* Burden of *Clostridium difficile* Infection in the United States. *N. Engl. J. Med.* **372,** 825–834 (2015).
2. Ross, C. L., Spinler, J. K. & Savidge, T. C. Structural and functional changes within the gut microbiota and susceptibility to Clostridium difficile infection. *Anaerobe* **41,** 37–43 (2016).
3. McDonald, L. C. *et al.* Clinical Practice Guidelines for Clostridium difficile Infection in Adults and Children: 2017 Update by the Infectious Diseases Society of America (IDSA) and Society for Healthcare Epidemiology of America (SHEA). *Clinical Infectious Diseases* **66,** e1–e48 (2018).
4. Wortelboer, K., Nieuwdorp, M. & Herrema, H. Fecal microbiota transplantation beyond Clostridioides difficile infections. *EBioMedicine* **44,** 716–729 (2019).
5. Hvas, C. L. *et al.* Fecal Microbiota Transplantation Is Superior to Fidaxomicin for Treatment of Recurrent Clostridium difficile Infection. *Gastroenterology* **156,** 1324-1332.e3 (2019).
6. DeFilipp, Z. *et al.* Drug-resistant e. coli bacteremia transmitted by fecal microbiota transplant. *N. Engl. J. Med.* **381,** 2043–2050 (2019).
7. Fecal Microbiota for Transplantation: Safety Communication- Risk of Serious Adverse Reactions Due to Transmission of Multi-Drug Resistant Organisms | FDA. at <https://www.fda.gov/safety/medical-product-safety-information/fecal-microbiota-transplantation-safety-communication-risk-serious-adverse-reactions-due>
8. Fecal Microbiota for Transplantation: Safety Alert - Risk of Serious Adverse Events Likely Due to Transmission of Pathogenic Organisms | FDA. at <https://www.fda.gov/safety/medical-product-safety-information/fecal-microbiota-transplantation-safety-alert-risk-serious-adverse-events-likely-due-transmission>
9. Hirten, R. P. *et al.* Microbial Engraftment and Efficacy of Fecal Microbiota Transplant for Clostridium Difficile in Patients With and Without Inflammatory Bowel Disease. *Inflamm. Bowel Dis.* **25,** 969–979 (2019).
10. Contijoch, E. J. *et al.* Gut microbiota density influences host physiology and is shaped by host and microbial factors. *Elife* **8,** 277095 (2019).
11. Tvede, M. & Rask-Madsen, J. Bacteriotherapy for chronic relapsing Clostridium difficile diarrhoea in six patients. *Lancet (London, England)* **1,** 1156–60 (1989).
12. Britton, G. J. *et al.* Microbiotas from Humans with Inflammatory Bowel Disease Alter the Balance of Gut Th17 and RORγt+ Regulatory T Cells and Exacerbate Colitis in Mice. *Immunity* **50,** 212-224.e4 (2019).
13. Yang, C. *et al.* Fecal IgA Levels Are Determined by Strain-Level Differences in Bacteroides ovatus and Are Modifiable by Gut Microbiota Manipulation. *Cell Host Microbe* **27,** 467-475.e6 (2020).
14. Vedanta Biosciences Announces Positive Topline Data from Two Phase 1 Studies of VE202, a Rationally Defined Bacterial Consortium Being Advanced for Inflammatory Bowel Diseases (IBD) :: Vedanta Biosciences, Inc. at <https://www.vedantabio.com/news-media/press-releases/detail/2682/vedanta-biosciences-announces-positive-topline-data-from>
15. Vedanta Biosciences Announces Successful Phase 1a/1b Data Demonstrating Safety, Tolerability, and Proof of Mechanism for Lead, Rationally Defined Bacterial Consortium Product Candidate, VE303 :: Vedanta Biosciences, Inc. at <https://www.vedantabio.com/news-media/press-releases/detail/2488/vedanta-biosciences-announces-successful-phase-1a1b-data>
16. Britton, R. A. & Young, V. B. Role of the Intestinal Microbiota in Resistance to Colonization by Clostridium difficile. *Gastroenterology* **146,** 1547–1553 (2014).
17. Bankevich, A. *et al.* SPAdes: A new genome assembly algorithm and its applications to single-cell sequencing. *J. Comput. Biol.* **19,** 455–477 (2012).
18. Darling, A. C. E., Mau, B., Blattner, F. R. & Perna, N. T. Mauve: Multiple alignment of conserved genomic sequence with rearrangements. *Genome Res.* **14,** 1394–1403 (2004).
19. Jain, C., Rodriguez-R, L. M., Phillippy, A. M., Konstantinidis, K. T. & Aluru, S. High throughput ANI analysis of 90K prokaryotic genomes reveals clear species boundaries. *Nat. Commun.* **9,** 1–8 (2018).
20. Seemann, T. Prokka: rapid prokaryotic genome annotation. *Bioinformatics* **30,** 2068–2069 (2014).
21. Jia, B. *et al.* CARD 2017: expansion and model-centric curation of the comprehensive antibiotic resistance database. *Nucleic Acids Res.* **45,** D566–D573 (2017).
22. Altschul, S. F., Gish, W., Miller, W., Myers, E. W. & Lipman, D. J. Basic local alignment search tool. *J. Mol. Biol.* **215,** 403–410 (1990).
23. Sayers, S. *et al.* Victors: a web-based knowledge base of virulence factors in human and animal pathogens. *Nucleic Acids Res.* **47,** D693–D700 (2019).

24. Chen, L. VFDB: a reference database for bacterial virulence factors. *Nucleic Acids Res.* **33,** D325–D328 (2004).
25. Wattam, A. R. *et al.* PATRIC, the bacterial bioinformatics database and analysis resource. *Nucleic Acids Res.* **42,** D581-91 (2014).
26. Waack, S. *et al.* Score-based prediction of genomic islands in prokaryotic genomes using hidden Markov models. *BMC Bioinformatics* **7,** 142 (2006).
27. Hsiao, W., Wan, I., Jones, S. J. & Brinkman, F. S. L. IslandPath: aiding detection of genomic islands in prokaryotes. *Bioinformatics* **19,** 418–420 (2003).
28. Petrof, E. O. *et al.* Stool substitute transplant therapy for the eradication of Clostridium difficile infection: 'RePOOPulating' the gut. *Microbiome* **1,** 3 (2013).
29. Faith, J. J., Ahern, P. P., Ridaura, V. K., Cheng, J. & Gordon, J. I. Identifying Gut Microbe-Host Phenotype Relationships Using Combinatorial Communities in Gnotobiotic Mice. *Sci. Transl. Med.* **6,** 220ra11-220ra11 (2014).
30. Sokol, H. *et al.* Faecalibacterium prausnitzii is an anti-inflammatory commensal bacterium identified by gut microbiota analysis of Crohn disease patients. *Proc. Natl. Acad. Sci. U. S. A.* **105,** 16731–16736 (2008).
31. Aggarwala *et.al.,* Quantification of discrete gut bacterial strains following fecal transplantation for recurrent *Clostridioides difficile* infection demonstrates long-term stable engraftment in non-relapsing recipients. bioRxiv. doi: 10.1101/2020.09.10.292136 (2020).
32. Britton et.al., SARS-CoV-2-specific IgA and limited inflammatory cytokines are present in the stool of select patients with acute COVID-19. bioRxiv. doi: 10.1101/2020.09.03.20183947 (2020).
33. Kassam Z, et.al., Donor screening for Fecal Microbiota Transplantation. NEJM. 381(21) 2017-2072 (2019).

### APPENDICES

#### APPENDIX 1 – GROWTH AND STABILITY

**Stability (viability in CFU) of select MTC01 strains over time**

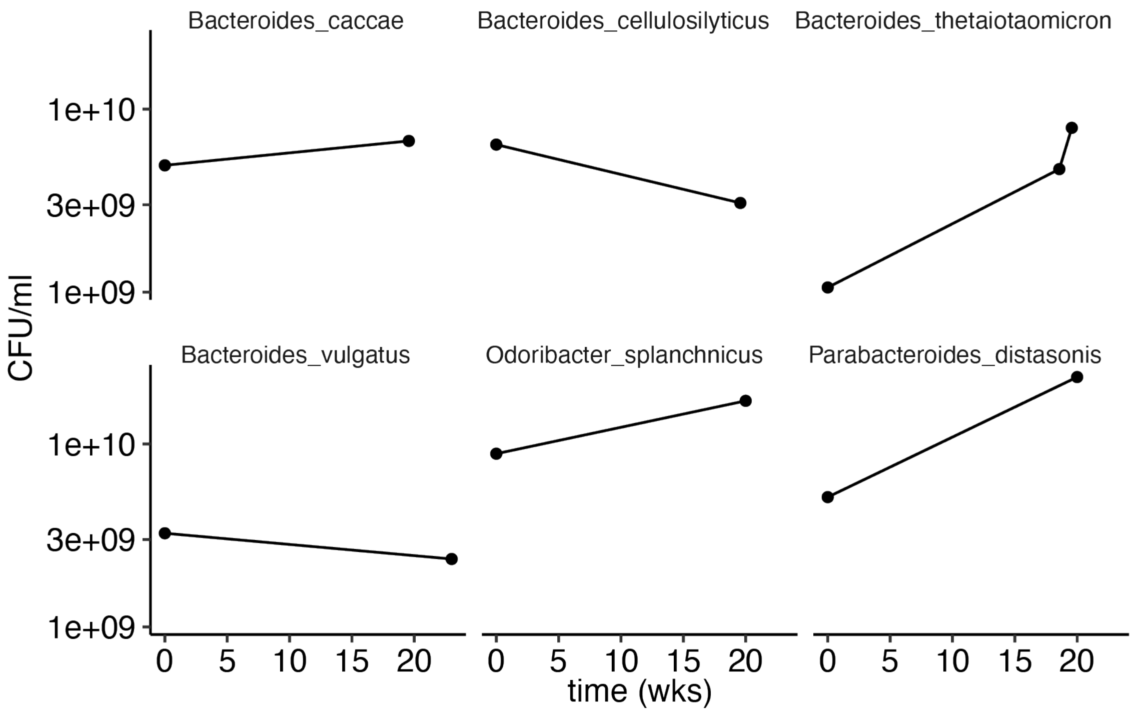

**Viability of select MTC01 strains across growth media at 90% of peak OD**
The composition of LYHBHI, Vegitone, and LYHVegitone (LYHV) are in Appendix 4.

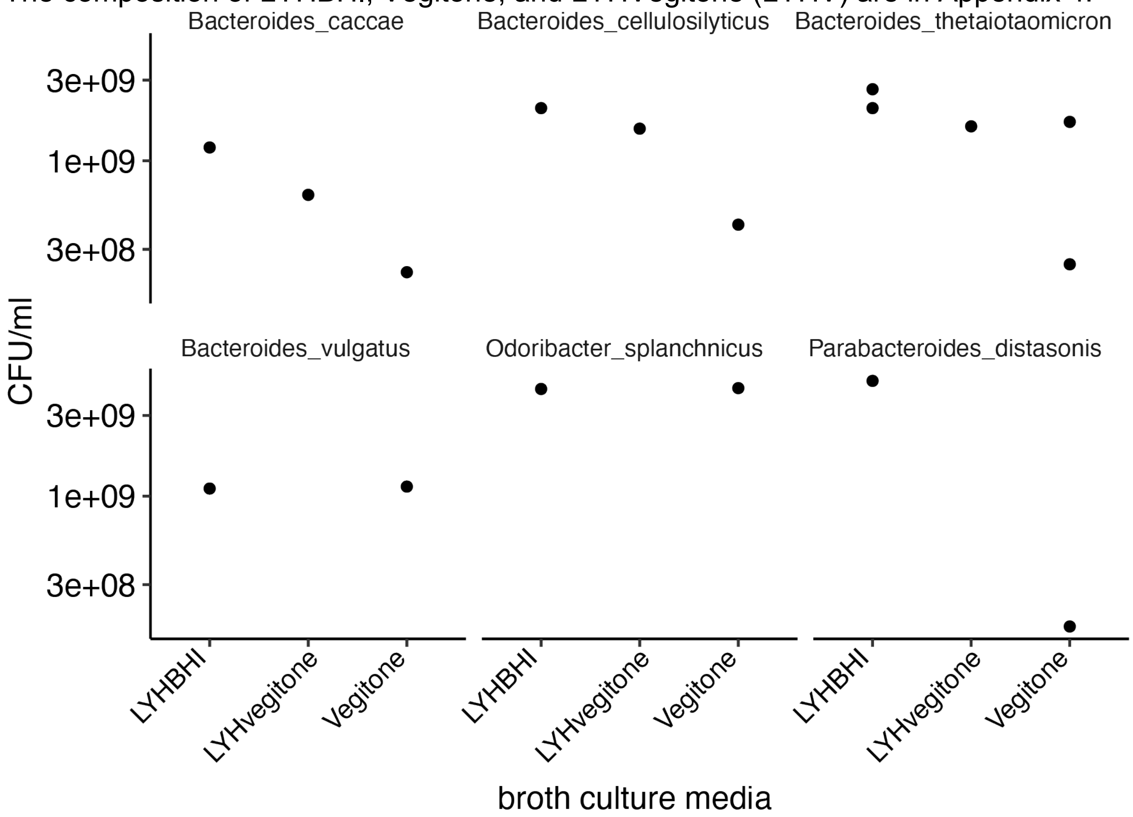

### APPENDIX 2 – VIRULENCE FACTORS AND TOXINS

**Table 5.** Virulence factors and toxins
Contigs from the draft SPAdes assemblies[17] were reordered into scaffolds using Mauve[18] based the closest complete NCBI reference sequence genome as determined by fastANI[19]. Genomes were annotated using Prokka[20] and the resulting translated coding sequences were used for determination of the presence of virulence and toxins encoding genes using BLAST[22] with Victors[23], VFDB[24] and PATRIC_VF[25] as reference databases.

| Strain ID | Species | Property | DB Source | Gene | Identity (%) | E-Value | Comment | doi |
|---|---|---|---|---|---|---|---|---|
| MTC01.01 | Bacteroides uniformis | Virulence Factor | Victors | SP_0320 | 82.759 | 8.14E-162 | Oxioreductase | 10.1046/j.1365-2958.2002.03106.x |
| MTC01.02 | Bacteroides ovatus | Virulence Factor | Victors | SP_0320 | 80.077 | 4.43E-156 | Oxioreductase | 10.1046/j.1365-2958.2002.03106.x |
| MTC01.03 | Bifidobacterium longum | NA | NA | NA | NA | NA | #N/A | #N/A |
| MTC01.04 | Bacteroides thetaiotaomicron | Virulence Factor | Victors | SP_0320 | 82.375 | 2.09E-161 | Oxioreductase | 10.1046/j.1365-2958.2002.03106.x |
| MTC01.05 | Bacteroides vulgatus | Virulence Factor | VFDB | cps4L | 80.916 | 0 | Single gene, not full cps4 locus | 10.1099/jmm.0.000573 |
| MTC01.05 | Bacteroides vulgatus | Virulence Factor | Victors | SP_0320 | 81.609 | 5.92E-161 | Oxioreductase | 10.1046/j.1365-2958.2002.03106.x |
| MTC01.06 | Collinsella aerofaciens | NA | NA | NA | NA | NA | #N/A | #N/A |
| MTC01.07 | Parabacteroides distasonis | NA | NA | NA | NA | NA | #N/A | #N/A |
| MTC01.08 | Bifidobacterium adolescentis | NA | NA | NA | NA | NA | #N/A | #N/A |
| MTC01.09 | Parabacteroides merdae | Virulence Factor | VFDB | cps4L | 80.153 | 0 | Single gene, not full cps4 locus | 10.1099/jmm.0.000573 |
| MTC01.09 | Parabacteroides merdae | Virulence Factor | Victors | SP_0320 | 84.231 | 3.53E-167 | Oxioreductase | 10.1046/j.1365-2958.2002.03106.x |
| MTC01.10 | Coprococcus comes | NA | NA | NA | NA | NA | #N/A | #N/A |
| MTC01.11 | Eubacterium rectale | NA | NA | NA | NA | NA | #N/A | #N/A |
| MTC01.12 | Bacteroides caccae | Virulence Factor | VFDB | cps4L | 81.795 | 0 | Single gene, not full cps4 locus | 10.1099/jmm.0.000573 |
| MTC01.12 | Bacteroides caccae | Virulence Factor | Victors | SP_0320 | 81.609 | 1.32E-159 | Oxioreductase | 10.1046/j.1365-2958.2002.03106.x |
| MTC01.13 | Dorea longicatena | NA | NA | NA | NA | NA | #N/A | #N/A |
| MTC01.14 | Odoribacter splanchnicus | NA | NA | NA | NA | NA | #N/A | #N/A |
| MTC01.15 | Bacteroides cellulosilyticus | Virulence Factor | VFDB | cps4L | 80.662 | 0 | Single gene, not full cps4 locus | 10.1099/jmm.0.000573 |
| MTC01.15 | Bacteroides cellulosilyticus | Virulence Factor | Victors | SP_0320 | 83.908 | 1.02E-166 | Oxioreductase | 10.1046/j.1365-2958.2002.03106.x |
| MTC01.16 | Bifidobacterium pseudocatenulatum | NA | NA | NA | NA | NA | #N/A | #N/A |

### APPENDIX 3 – PREDICTED ANTIBIOTIC RESISTANCE GENES AND MOBILE GENETIC ELEMENTS

**Table 6.** Predicted antibiotic resistance genes
Contigs from the draft SPAdes assemblies[17] were reordered into scaffolds using Mauve[18] based the closest complete NCBI reference sequence genome as determined by fastANI[19]. Genomes were annotated using Prokka[20] and the resulting translated coding sequences were used for prediction antibiotic resistance (AR) genetic elements using CARD[21]. The presence of AR genes on genomic islands and the potential for horizontal gene transfer was determined by SIGI-HMM[26] and IslandPath[27].

| Strain ID | Species | Gene | Drug Class | Resistance Mechanism | Comment |
|---|---|---|---|---|---|
| MTC01.01 | Bacteroides uniformis | adeF | multiple antibiotics | efflux | |
| MTC01.01 | Bacteroides uniformis | tetQ | tetracycline antibiotic | target protection | |
| MTC01.01 | Bacteroides uniformis | CblA-1 | cephalosporin | inactivation | |
| MTC01.02 | Bacteroides ovatus | adeF | multiple antibioticsacterial free fatty acids | efflux | |
| MTC01.02 | Bacteroides ovatus | tetQ | tetracycline antibiotic | target protection | |
| MTC01.03 | Bifidobacterium longum | rpoB mutant | rifamycin antibiotic | target alteration; target replacement | |
| MTC01.04 | Bacteroides thetaiotaomicron | adeF | multiple antibioticsacterial free fatty acids | efflux | |
| MTC01.04 | Bacteroides thetaiotaomicron | tetQ | tetracycline antibiotic | target protection | |
| MTC01.05 | Bacteroides vulgatus | adeF | multiple antibioticsacterial free fatty acids | efflux | |
| MTC01.05 | Bacteroides vulgatus | tetQ | tetracycline antibiotic | target protection | |
| MTC01.05 | Bacteroides vulgatus | ErmF | macrolide antibiotic; lincosamide antibiotic; streptogramin antibiotic | target alteration | |
| MTC01.05 | Bacteroides vulgatus | vanYF | glycopeptide antibiotic | target alteration | Not a full operon, not on a MGE |
| MTC01.06 | Collinsella aerofaciens | IMP-13 | carbapenem; cephalosporin; cephamycin; penam; penem | inactivation | Not on a MGE |
| MTC01.07 | Parabacteroides distasonis | tetB(P) | tetracycline antibiotic | target protection | |
| MTC01.07 | Parabacteroides distasonis | adeF | multiple antibioticsacterial free fatty acids | efflux | |
| MTC01.08 | Bifidobacterium adolescentis | MuxC | multiple antibioticsacterial free fatty acids | efflux | |
| MTC01.08 | Bifidobacterium adolescentis | tetO | tetracycline antibiotic | target protection | |
| MTC01.08 | Bifidobacterium adolescentis | rpoB mutant | rifamycin antibiotic | target alteration; target replacement | |
| MTC01.09 | Parabacteroides merdae | adeF | multiple antibioticsacterial free fatty acids | efflux | |
| MTC01.09 | Parabacteroides merdae | Mef(En2) | multiple antibiotics | efflux | |
| MTC01.09 | Parabacteroides merdae | tetQ | tetracycline antibiotic | target protection | |
| MTC01.11 | Eubacterium rectale | AAC(3)-IId | aminoglycoside antibiotic | inactivation | |
| MTC01.12 | Bacteroides caccae | tetQ | tetracycline antibiotic | target protection | |
| MTC01.12 | Bacteroides caccae | adeF | multiple antibioticsacterial free fatty acids | efflux | |
| MTC01.13 | Dorea longicatena | ErmB | macrolide antibiotic; lincosamide antibiotic; streptogramin antibiotic | target alteration | |
| MTC01.13 | Dorea longicatena | tetM | tetracycline antibiotic | target protection | |
| MTC01.14 | Odoribacter splanchnicus | adeF | multiple antibioticsacterial free fatty acids | efflux | |
| MTC01.14 | Odoribacter splanchnicus | tetQ | tetracycline antibiotic | target protection | |
| MTC01.14 | Odoribacter splanchnicus | ErmF | macrolide antibiotic; lincosamide antibiotic; streptogramin antibiotic | target alteration | |
| MTC01.14 | Odoribacter splanchnicus | tetX | glycylcycline; tetracycline antibiotic | inactivation | |
| MTC01.14 | Odoribacter splanchnicus | aadS | aminoglycoside antibiotic | inactivation | |
| MTC01.15 | Bacteroides cellulosilyticus | tetQ | tetracycline antibiotic | target protection | |
| MTC01.15 | Bacteroides cellulosilyticus | ANT(4')-Ib | aminoglycoside antibiotic | inactivation | |
| MTC01.15 | Bacteroides cellulosilyticus | adeF | multiple antibioticsacterial free fatty acids | efflux | |
| MTC01.16 | Bifidobacterium pseudocatenulatum | MuxC | multiple antibioticsacterial free fatty acids | efflux | |
| MTC01.16 | Bifidobacterium pseudocatenulatum | vanC | glycopeptide antibiotic | target alteration | Not a full operon, not on a MGE |

| Strain ID | Species | Gene | Drug Class | Resistance Mechanism | Comment |
|---|---|---|---|---|---|
| MTC01.16 | Bifidobacterium pseudocatenulatum | dfrF | diaminopyrimidine antibiotic | target replacement | |
| MTC01.16 | Bifidobacterium pseudocatenulatum | rpoB mutant | rifamycin antibiotic | target alteration; target replacement | |
| MTC01.16 | Bifidobacterium pseudocatenulatum | tet(W/N/W) | tetracycline antibiotic | target protection | |

**Figure 7.** Predicted antibiotic resistance genes and mobile genetic elements.
Contigs from the draft SPAdes assemblies[17] were reordered into scaffolds using Mauve[18] based the closest complete NCBI reference sequence genome as determined by fastANI[19]. Genomes were annotated using Prokka[20] and the resulting translated coding sequences were used for prediction antibiotic resistance (AR) genetic elements using CARD[21] (antibiotic resistance gene indicated as text). The presence of AR genes on the full genome (grey) on genomic islands and the potential for horizontal gene transfer was determined by SIGI-HMM[26] (red) and IslandPath[27] (green).

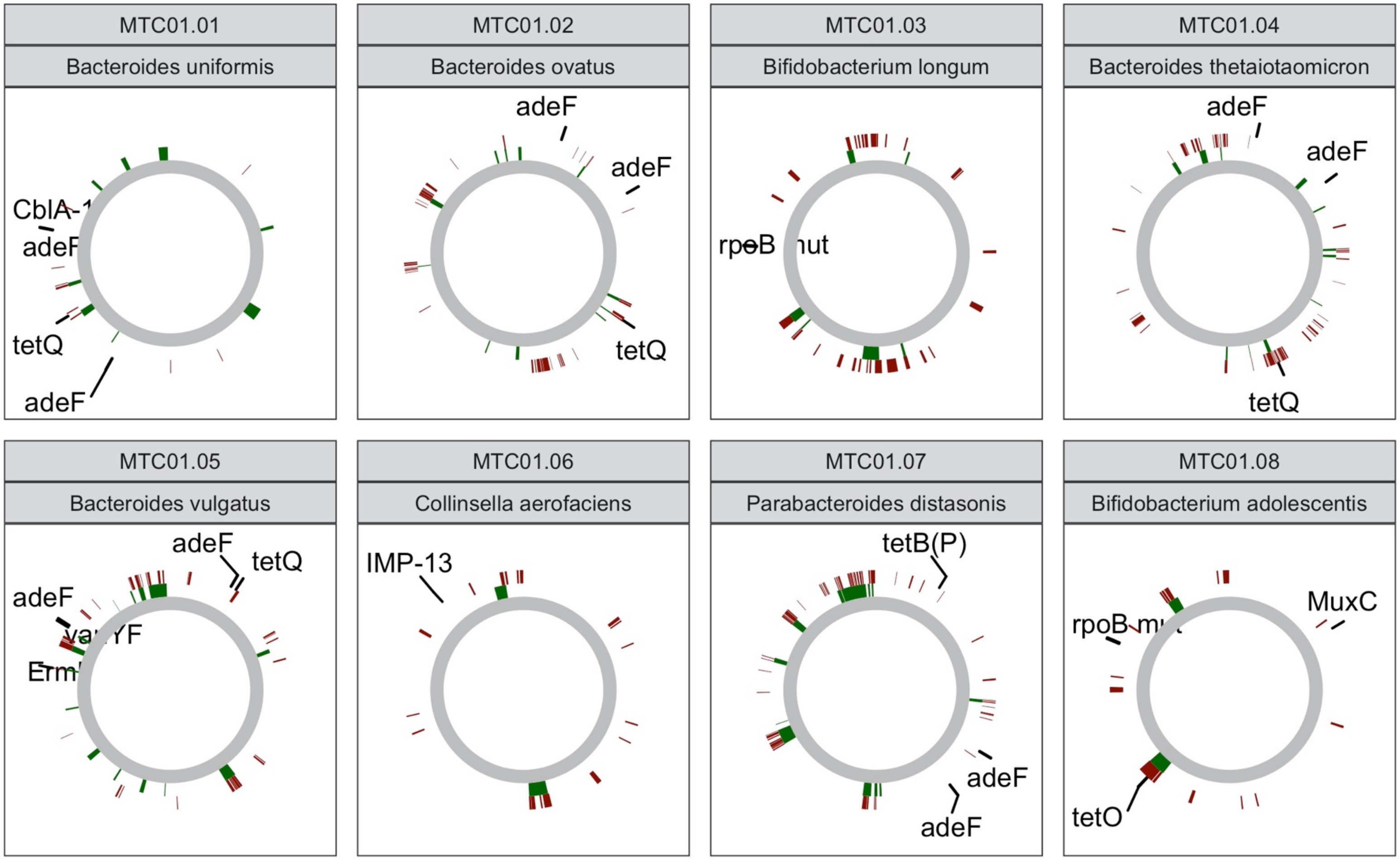

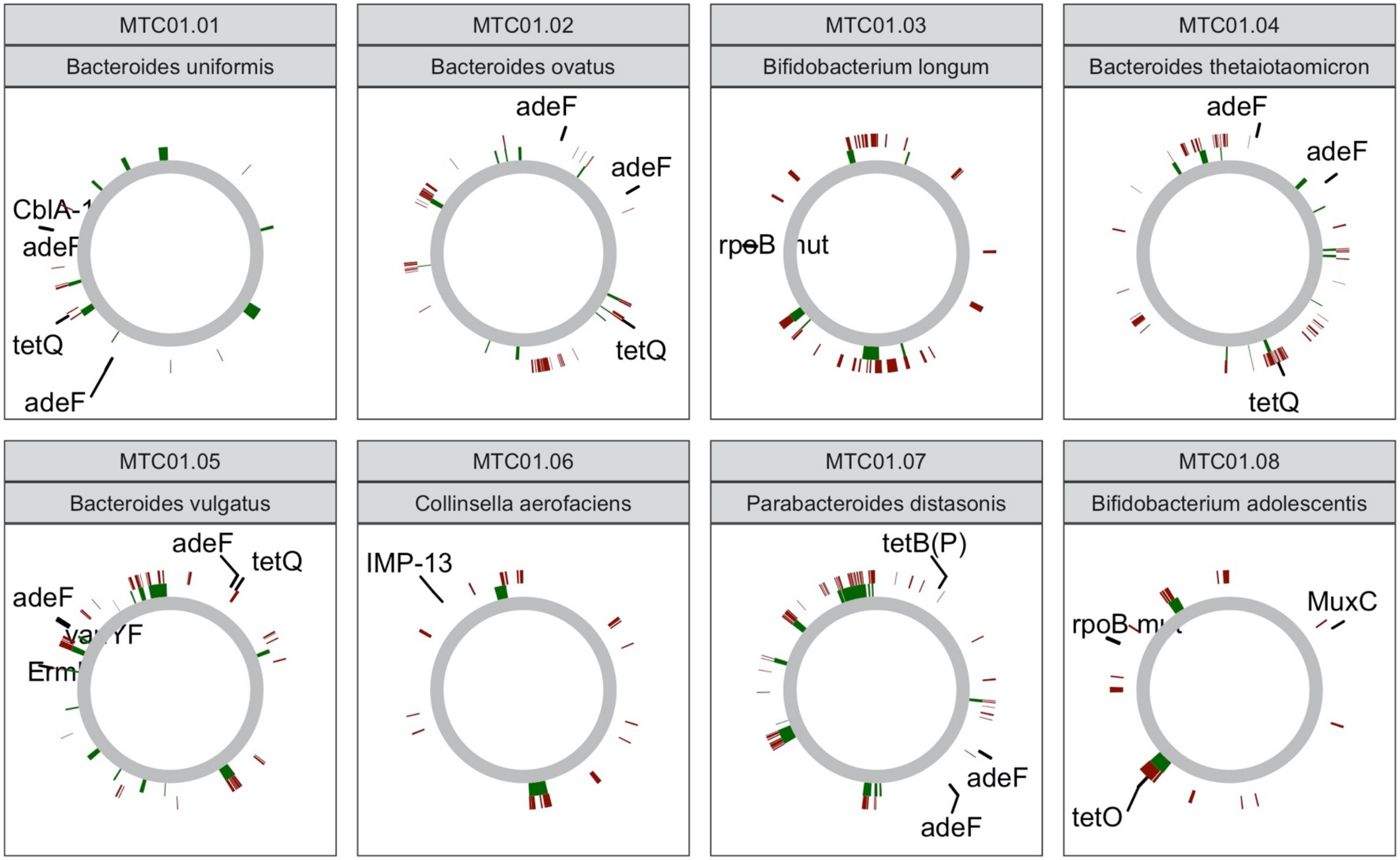
MTC01.01
Bacteroides uniformis
CblA-1
adeF
tetQ
adeF
MTC01.02
Bacteroides ovatus
adeF
adeF
tetQ
MTC01.03
Bifidobacterium longum
rpoB
MTC01.04
Bacteroides thetaiotaomicron
adeF
adeF
tetQ
MTC01.05
Bacteroides vulgatus
adeF
tetQ
adeF
MTC01.06
Collinsella aerofaciens
IMP-13
MTC01.07
Parabacteroides distasonis
tetB(P)
adeF
adeF
MTC01.08
Bifidobacterium adolescentis
MuxC
tetO

### APPENDIX 4 – LIST OF MATERIALS

#### LYHBHIv4

Media is a supplemented BHI media modified from Sokol et al, 2018[30]

##### BHI

| Reagent | | | Amount | |
|---|---|---|---|---|
| 12.5 | g/L | Calf brain, infusion from | 12.5 | g |
| 5 | g/L | Beef heart, infusion from | 5 | g |
| 10 | g/L | Pancreatic digest of casein | 10 | g |
| 2 | g/L | Glucose | 2 | g |
| 5 | g/L | Sodium chloride | 5 | g |
| 2.5 | g/L | Disodium phosphate | 2.5 | g |

| Reagent | | | Amount | |
|---|---|---|---|---|
| 1 | % | Menadione | 0.2 | g |

| Reagent | | | Amount | |
|---|---|---|---|---|
| 37 | g/L | BHI | 37 | g |
| 0.5 | % | Yeast extract | 5 | g |
| 1 | g/L | D-Xylose | 1 | g |
| 1 | g/L | D-Fructose | 1 | g |
| 1 | g/L | D-Glucose | 1 | g |
| 1 | g/L | D-Galactose | 1 | g |
| 0.5 | g/L | N-Acetylglucosamine | 0.5 | g |
| 0.5 | g/L | L-Arabinose | 0.5 | g |
| 1 | g/L | Cellobiose | 1 | g |
| 1 | g/L | Maltose | 1 | g |
| 1 | g/L | Sucrose | 1 | g |
| 0.5 | g/L | Cysteine | 0.5 | g |
| 1 | g/L | Malic acid | 1 | g |
| 1 | g/L | Sodium Sulfate | 1 | g |
| 0.005 | g/L | hemin | 0.005 | g |
| 0.05 | % | Tween 80 | 500 | ul |
| 0.1 | % | Vitamin K solution | 1 | mL |
| 0.5 | % | Resazurin | 2 | mL |
| 0.1 | M | MOPS | 20.92633 | g |
| | | pH 7.2 - KOH, H20 fill to | 1 | L |

Filter sterilize

#### LYH Vegitone infusion broth (LYHV)

Media is based on LYHBHI but with Vegitone Infusion Broth instead of BHI and no hemin

##### Vegitone Infusion Broth [Sigma Aldrich]

| Reagent | | | Amount | |
|---|---|---|---|---|
| 12.5 | g/L | Vegetable special infusion powder | 12.5 | g |
| 5 | g/L | Vegetable infusion powder | 5 | g |
| 10 | g/L | Peptone (vegetable) | 10 | g |
| 2 | g/L | Glucose | 2 | g |
| 5 | g/L | Sodium chloride | 5 | g |
| 2.5 | g/L | Disodium phosphate | 2.5 | g |

| Reagent | | | Amount | |
|---|---|---|---|---|
| 1 | % | Menadione | 0.2 | g |

| Reagent | | | Amount | |
|---|---|---|---|---|
| 37 | g/L | BHI | 37 | g |
| 0.5 | % | Yeast extract | 5 | g |
| 1 | g/L | D-Xylose | 1 | g |
| 1 | g/L | D-Fructose | 1 | g |
| 1 | g/L | D-Glucose | 1 | g |
| 1 | g/L | D-Galactose | 1 | g |
| 0.5 | g/L | N-Acetylglucosamine | 0.5 | g |
| 0.5 | g/L | L-Arabinose | 0.5 | g |
| 1 | g/L | Cellobiose | 1 | g |
| 1 | g/L | Maltose | 1 | g |
| 1 | g/L | Sucrose | 1 | g |
| 0.5 | g/L | Cysteine | 0.5 | g |
| 1 | g/L | Malic acid | 1 | g |
| 1 | g/L | Sodium Sulfate | 1 | g |
| 0.05 | % | Tween 80 | 500 | ul |
| 0.1 | % | Vitamine K solution | 1 | mL |
| 0.2 | % | Resazurin | 2 | mL |
| 0.1 | M | MOPS | 20.92633 | g |
| | | pH 7.2 - KOH, H20 fill to | 1 | L |

Filter sterilize

### APPENDIX 5 – IDENTIFICATION

**Table 7.** Genomic identity.
Genome length determined from the draft SPAdes assemblies[17]. Species named determined by kmer-based genome alignment to >150K RefSeq bacterial genomes.

| Strain ID | Species | Genome Length (bp) |
|---|---|---|
| MTC01.01 | Bacteroides uniformis | 4518790 |
| MTC01.02 | Bacteroides ovatus | 6857847 |
| MTC01.03 | Bifidobacterium longum | 2567636 |
| MTC01.04 | Bacteroides thetaiotaomicron | 7058185 |
| MTC01.05 | Bacteroides vulgatus | 5398426 |
| MTC01.06 | Collinsella aerofaciens | 2185677 |
| MTC01.07 | Parabacteroides distasonis | 5440480 |
| MTC01.08 | Bifidobacterium adolescentis | 2098181 |
| MTC01.09 | Parabacteroides merdae | 4494726 |
| MTC01.10 | Coprococcus comes | 3350557 |
| MTC01.11 | Eubacterium rectale | 3308314 |
| MTC01.12 | Bacteroides caccae | 5732602 |
| MTC01.13 | Dorea longicatena | 3002367 |
| MTC01.14 | Odoribacter splanchnicus | 4744485 |
| MTC01.15 | Bacteroides cellulosilyticus | 6921208 |
| MTC01.16 | Bifidobacterium pseudocatenulatum | 2296089 |

**Table 8.** Bruker MALDI Biotyper identity.

| Strain ID | Genus | Species | Bruker Species | Bruker Score |
|---|---|---|---|---|
| MTC01.01 | Bacteroides | uniformis | Bacteroides uniformis | >1.8 |
| MTC01.02 | Bacteroides | ovatus | Bacteroides ovatus | >1.8 |
| MTC01.03 | Bifidobacterium | longum | Bifidobacterium longum | >1.8 |
| MTC01.04 | Bacteroides | thetaiotaomicron | Bacteroides thetaiotaomicron | >1.8 |
| MTC01.05 | Bacteroides | vulgatus | Bacteroides vulgatus | >1.8 |
| MTC01.06 | Collinsella | aerofaciens | Collinsella aerofaciens | >1.8 |
| MTC01.07 | Parabacteroides | distasonis | Parabacteroides distasonis | >1.8 |
| MTC01.08 | Bifidobacterium | adolescentis | Bifidobacterium adolescentis | >1.8 |
| MTC01.09 | Parabacteroides | merdae | Parabacteroides johnsonii | >1.8 |
| MTC01.10 | Coprococcus | comes | Coprococcus comes | >1.8 |
| MTC01.11 | Eubacterium | rectale | Eubacterium rectale | >1.8 |
| MTC01.12 | Bacteroides | caccae | Bacteroides caccae | >1.8 |
| MTC01.13 | Dorea | longicatena | Dorea longicatena | >1.8 |
| MTC01.14 | Odoribacter | splanchnicus | Odoribacter splanchnicus | >1.8 |
| MTC01.15 | Bacteroides | cellulosilyticus | Bacteroides cellulosilyticus | >1.8 |
| MTC01.16 | Bifidobacterium | pseudocatenulatum | Bifidobacterium pseudocatenulatum | >1.8 |

For species with very small colonies (e.g., *E. rectale*), identification will alternatively be determined by transferring a colony into broth where it may be grown to an OD600 > 0.1 and identification will be determined from MALDI-TOF of the broth with a minimum Bruker score >1.6. For both broth and colonies, the correct species must be the top hit.

#### APPENDIX 6 – STUDY SCHEMA

20 individuals will follow the following study schema

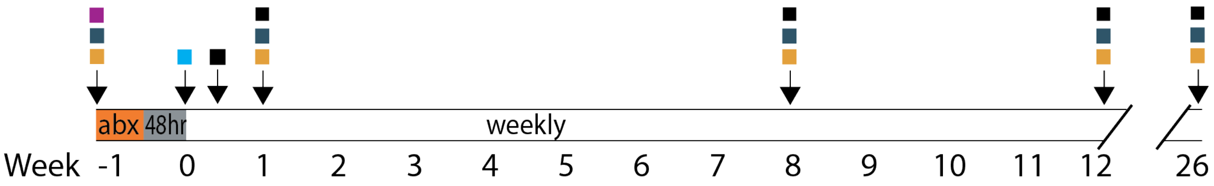

**Assay Legend**

- consent
- blood
- stool for C.diff PCR/ELISA, microbiome, calprotectin
- colonoscopy to deliver FMT
- Adverse event assessment

**Therapeutics Legend**

| | |
|---|---|
| abx | vancomycin (minimum 4 days) |
| 48hr | antibiotic washout (halt vancomycin) |
| weekly | capsules taken once per week |

Note *C. difficile* will assayed with both methods for all stool samples except for the pre-intervention sample when the individual is taking antibiotics.

### APPENDIX 7 – DONOR HEALTH RECORDS

**Table 9.** Health records FMT donors.

| | Donor | |
|---|---|---|
| | **1001275** | **1001283** |
| History | | |
| Medical history | No | No |
| Surgical history | No | No |
| Family history of any significant medical problems | No | No |
| Medications | No | No |
| Known allergies | No | No |
| 10 point Review of Systems negative | No | No |
| Stool tests | | |
| Shiga toxin EIA | Not detected | Not detected |
| Salmonella/Shigella culture | Negative | Negative |
| Campylobacter culture | Negative | Negative |
| Ova and Parasites | Negative | Negative |
| C diff GDH antigen | Negative | N/A |
| C diff Toxin EIA | Negative | N/A |
| C diff PCR | N/A | Negative |
| Blood tests | | |
| RPR | Negative | Negative |
| Hepatitis A antibody | Nonreactive | Nonreactive |
| Hepatitis B surface Antigen | Nonreactive | Nonreactive |
| Hepatitis B surface Antibody | Reactive | Nonreactive |
| Hepatitis B core antibody | Nonreactive | Nonreactive |
| Hepatitis C antibody | Nonreactive | Nonreactive |
| HIV 1/2 | Nonreactive | Nonreactive |
| CMV IgG Antibody | Negative | Negative |

#### APPENDIX 8 – CUSTOM ANAEROBIC CHAMBER

To manufacture the strains in this LBP, a custom anaerobic Coy chamber has been developed in collaboration with Coy Laboratories, a leading source of anaerobic chambers. Although all strain growth for MTC01 will be performed with individually contained and sterilized culture vessels, the risk of contamination of these vessels can be minimized tremendously by operating in a controlled environment (e.g., a hood or clean room). Given the relatively small volumes required for this study, we will take advantage of technologies and SOPs from working with germ-free mice to reduce microbial loads in our anaerobic chamber to near zero. The chamber is designed to be easily chemically sterilizable with ports for atomizing and draining sterilant. Equipment that is not autoclavable or filter sterilizable will be sterilized by irradiation prior to import. This system will be far cleaner than a clean room at substantially reduced cost and more than enough capacity for this study.

Design schema of the new chamber provided by the engineers at Coy Labs.

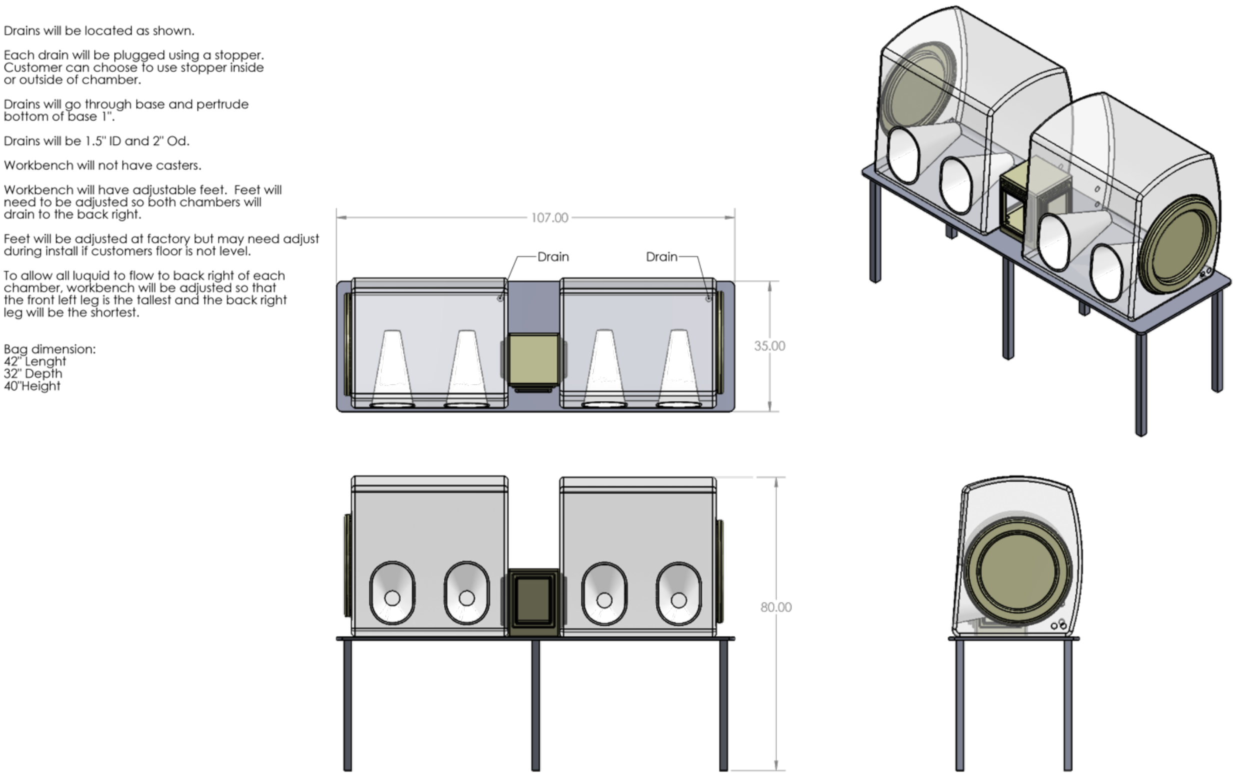

### APPENDIX 9 – RESEARCH CELL BANK RESULTS

| StrainID | Species | Agar Plating Date | Plate Incubation Time (hrs) | Colony Pick Date | Colony Bruker Score | Broth Incubation Time (hrs) | Harvest Date | Harvest OD600 | Broth Bruker Score |
|---|---|---|---|---|---|---|---|---|---|
| MTC01.01 | Bacteroides uniformis | 8/18/20 | 24 | 8/19/20 | 2.35 | 24 | 8/20/20 | 1.10 | 2.30 |
| MTC01.02 | Bacteroides ovatus | 8/24/20 | 48 | 8/26/20 | 1.87 | 24 | 8/27/20 | 1.49 | 2.00 |
| MTC01.03 | Bifidobacterium longum | 8/18/20 | 24 | 8/19/20 | 2.01 | 24 | 8/20/20 | 1.41 | 2.02 |
| MTC01.04 | Bacteroides thetaiotaomicron | 8/31/20 | 48 | 9/2/20 | 2.30 | 24 | 9/3/20 | 1.44 | 2.16 |
| MTC01.05 | Bacteroides vulgatus | 8/31/20 | 48 | 9/2/20 | 2.53 | 24 | 9/3/20 | 1.15 | 2.40 |
| MTC01.06 | Collinsella aerofaciens | 8/28/20 | 72 | 8/31/20 | 2.17 | 24 | 9/1/20 | 1.36 | 2.17 |
| MTC01.07 | Parabacteroides distasonis | 8/31/20 | 48 | 9/2/20 | 2.21 | 24 | 9/3/20 | 1.27 | 2.15 |
| MTC01.08 | Bifidobacterium adolescentis | 8/18/20 | 24 | 8/19/20 | 2.22 | 24 | 8/20/20 | 1.52 | 1.79 |
| MTC01.09 | Parabacteroides merdae | 8/21/20 | 72 | 8/24/20 | 2.00 | 24 | 8/25/20 | 0.72 | 2.45 |
| MTC01.10 | Coprococcus comes | 8/21/20 | 72 | 8/24/20 | 2.14 | 24 | 8/25/20 | 1.46 | 2.18 |
| MTC01.11 | Eubacterium rectale | 8/21/20 | 72 | 8/24/20 | colonies too small for detection | 48 | 8/26/20 | 0.26 | 2.36 |
| MTC01.12 | Bacteroides caccae | 8/31/20 | 48 | 9/2/20 | 2.23 | 24 | 9/3/20 | 1.31 | 2.27 |
| MTC01.13 | Dorea longicatena | 8/24/20 | 48 | 8/26/20 | 1.82 | 24 | 8/27/20 | 1.13 | 1.62 |
| MTC01.14 | Odoribacter splanchnicus | 8/31/20 | 48 | 9/2/20 | 2.37 | 24 | 9/3/20 | 1.20 | 2.45 |
| MTC01.15 | Bacteroides cellulosilyticus | 8/31/20 | 48 | 9/2/20 | 2.14 | 24 | 9/3/20 | 1.26 | 2.30 |
| MTC01.16 | Bifidobacterium pseudocatenulatum | 8/24/20 | 48 | 8/26/20 | 2.14 | 24 | 8/27/20 | 1.20 | 2.23 |

Note that for every Bruker MALDI-TOF species identification reading above, the correct species was the top hit.

### APPENDIX 10 – IN VITRO ANTIBIOTIC SUSCEPTIBILITY

| Organism | | MIC (µg/mL) | | | | | | | | | |
|---|---|---|---|---|---|---|---|---|---|---|---|
| | ATCC/ID | VAN | MTZ | TGC | SAM | AMC | MEM | TZP | CLI | CRO | MOX |
| *Bacteroides fragilis* | 25285 | >32 | 0.5 (0.25-1) | 0.25 (0.12-1) | 1/0.5 (0.5/0.25-2/1) | 1/0.5 (0.25/0.12-1/0.5) | 0.12 (0.03-0.25) | 0.25/4 (0.12/4-0.5/4) | 1 (0.5-2) | 64 (32-128) | 0.25 (0.12-0.5) |
| *Bacteroides thetaiotaomicron* | 29741 | >32 | 1 (0.5-2) | 0.25 (0.5-2) | 1/0.5 (0.5/0.25-2/1) | 0.5/0.25 (0.5/0.25-2/1) | 0.12 (0.12-0.5) | 4/4 (4/4-16/4) | 4 (2-8) | 64 (64-256) | 2 (1-4) |
| *Clostridiodes difficile* | 700057 | 2 (0.5-4) | 0.5 (0.12-0.5) | 1 (0.06-0.5) | 2/1 (0.5/0.25-4/2) | 1/0.5 (0.25/0.12/1/0.5) | 2 (0.5-4) | 16/4 (4/4-16/4) | 4 (2-8) | 32 | 2 (1-4) |
| *Eggerthella lenta* | 43055 | 2 | 0.5 | 0.12 | 1/0.5 (0.25/0.12-2/1) | 1/0.5 | 0.5 (0.12-1) | 32/4 (4/4-16/4) | 0.12 (0.06-0.25) | 256 | 0.25 (0.12-0.5) |
| *Bacteroides uniformis* | MTCO1.01 | >32 | 1 | 0.25 | 1/0.5 | 1/0.5 | 0.12 | 0.25/4 | 2 | 64 | 2 |
| *Bacteroides ovatus* | MTCO1.02 | >32 | 1 | 0.25 | 1/0.5 | 0.5/0.25 | 0.25 | 4/4 | 2 | 64 | 2 |
| *Bacteroides thetaiotaomicron* | MTCO1.04 | >32 | 0.5 | 2 | 1/0.5 | 1/0.5 | 0.25 | 4/4 | 4 | 64 | 1 |
| *Bacteroides vulgatus* | MTCO1.05 | 32 | 0.5 | 2 | 4/2 | 0.5/0.25 | 0.25 | 4/4 | >8 | 16 | >16 |
| *Collinsella aerofaciens* | MTCO1.06 | 2 | 2 | 0.25 | 1/0.5 | 0.5/0.25 | 0.25 | 4/4 | <0.03 | 16 | 0.5 |
| *Parabacteroides distasonis* | MTCO1.07 | >32 | 1 | 1 | 8/4 | 7/2 | 0.5 | >32/4 | 0.25 | 32 | 4 |
| *Parabacteroides merdae* | MTCO1.09 | >32 | 1 | 1 | 2/1 | 2/1 | 0.25 | 16/4 | 2 | 16 | 0.25 |
| *Bacteroides caccae* | MTCO1.12 | >32 | 1 | >8 | 2/1 | 0.5/0.25 | 0.12 | 4/4 | 1 | 16 | 2 |
| *Odoribacter splanchnicus* | MTCO1.14 | >32 | 1 | 0.12 | 4/2 | 2/1 | 0.12 | <0.06/4 | >8 | 16 | 2 |
| *Bacteroides cellulosilyticus* | MTCO1.15 | >32 | 1 | 2 | 4/2 | 2/1 | 0.25 | 8/4 | 2 | 64 | 4 |
| *Bifdobacterium pseudocatenulatum* | MTCO1.16 | 0.5 | 2 | 1 | 0.12/0.06 | 0.06/0.03 | 0.12 | 0.5/5 | <0.03 | <0.5 | 4 |

**Color Legend**
Susceptible
Intermediate
Resistant

The above table provides interim results of Minimum Inhibitory Concentrations (MIC) determined for 11 of the 16 strains in MTC01. The remaining five strains are in process. The top four strains are control strains from ATCC. MIC is being determined for 10 antibiotics by agar dilution in accordance with guidelines from the Clinical and Laboratory Standards Institute (CLSI; 1,2). These experiments are being performed by Micromyx, LLC.

Antibiotic abbreviations: vancomycin (VAN), metronidazole (MTZ), tigecycline (TGC), ampicillin-sulbactam (SAM), amoxicillin-clavulante (AMC), meropenem (MEM), piperacillin-tazobactam (TZP), clindamycin (CLI), ceftriaxone (CRO), amoxicillin (MOX).
