## Supplementary material for "Live biotherapeutic product exhibits similar efficacy and superior engraftment to same donor fecal microbiota transplant for recurrent Clostridioides difficile infection": Bethlehem etal MTC01 IND

[*] Co-corresponding authors

Icahn School of Medicine at Mount Sinai
*Conduits*
The Institutes for Translational Sciences

Nov 1, 2022

Food and Drug Administration
Center for Biologics Evaluation and Research
Central Document Room
5901-B Ammendale Rd.
Beltsville, MD 20705-1266

RE: **Initial Investigator New Drug Application**
**Serial Number 0000**

Dear Sir/Madam;

We are submitting this initial application for a Sponsor-Investigator IND. We propose to evaluate a defined bacterial consortia MTC01 in comparison with fecal microbiota transplant under this IND for safety and efficacy for the treatment of recurrent C. difficile infection (rCDI).

This submission contains the initial study protocol entitled, "Phase 1b comparison of defined bacterial consortium MTC01 and Fecal Microbiota Transplantation for the treatment of recurrent Clostridioides difficile infection." MTC01 is a defined consortia of the best engrafting strains from donor D283, whose stool was used for 25 FMTs at Mount Sinai. We will compare MTC01 with donor stool from D283 in individuals with rCDI infection. Approximately 60 patients with rCDI will be enrolled in this study and will undergo a single administration of a microbial therapeutic by colonoscopy to allow a comparison of safety, efficacy, and engraftment when the same strains are transmitted by stool or defined consortia. The primary outcome is safety at eight and twelve weeks. MTC01 was described in Pre-IND PS005985, CRMTS # 12870.

If you have any questions regarding this submission, please contact myself at 212-241-7757 or.

Sincerely,

Ari Grinspan, MD
Associate Professor
Division of Gastroenterology
Icahn School of Medicine at Mount Sinai

Icahn School of Medicine at Mount Sinai

*Conduits*
The Institutes for Translational Sciences

### Investigational New Drug Application

**Regulatory Sponsor:** Ari Grinspan MD,
Department of Medicine/Division of Gastroenterology
Icahn School of Medicine at Mount Sinai, New York, NY
One Gustave L. Levy Place Box 1069
New York NY 10029
212-241-7757

**Funding Sponsor:** National Institute of Diabetes and Digestive and Kidney Diseases
6707 Democracy Boulevard
Bethesda, MD 20892

**Study Product:** MTC01 (defined LBP) and FMT

**Protocol Number:** 1.0

**Date: Nov 1, 2022**

### 1 FDA Form 1571 [21 CFR 312.23(a)(1)]

Next Page | Export Data | Import Data | Reset Form

**DEPARTMENT OF HEALTH AND HUMAN SERVICES**
Food and Drug Administration

### INVESTIGATIONAL NEW DRUG APPLICATION (IND)

*(Title 21, Code of Federal Regulations (CFR) Part 312)*

Form Approved: OMB No. 0910-0014
Expiration Date: March 31, 2022
*See PRA Statement on page 3.*

NOTE: No drug/biologic may be shipped or clinical investigation begun until an IND for that investigation is in effect (21 CFR 312.40)

1. Name of Sponsor
Ari Grinspan

2. Date of Submission *(mm/dd/yyyy)*
11/01/2022

3. Sponsor Address

Address 1 *(Street address, P.O. box, company name c/o)*
One Gustave L. Levy Place

Address 2 *(Apartment, suite, unit, building, floor, etc.)*
Box 1069

City: New York
State/Province/Region: NY
Country: USA
ZIP or Postal Code: 10029

4. Telephone Number *(Include country code if applicable and area code)*
212-241-7757

5. Name of Drug *(Include all available names: Trade, Generic, Chemical, or Code)*
MTC01 (defined live biotherapeutic product); fecal microbiota transplantation comparator

Continuation Page for #5

6A. IND Number *(If previously assigned)*

6B. Select One: ☐ Commercial ☑ Research

7A. (Proposed) Indication for Use
recurrent C. difficile infection

Is this indication for a rare disease (prevalence <200,000 in U.S.)? ☑ Yes ☐ No

Does this product have an FDA Orphan Designation for this indication? ☐ Yes ☑ No

If yes, provide the Orphan Designation number for this indication:

Continuation Page for #7

7B. SNOMED CT Indication Disease Term *(Use continuation page for each additional indication and respective coded disease term)*
789685005

8. Phase of Clinical Investigation to be conducted ☑ Phase 1 ☐ Phase 2 ☐ Phase 3 ☐ Other *(Specify)*:

9. List numbers of all Investigational New Drug Applications (21 CFR Part 312), New Drug Applications (21 CFR Part 314) , Drug Master Files (21 CFR Part 314.420) , and Biologics License Applications (21 CFR Part 601) referred to in this application.

10. IND submission should be consecutively numbered. The initial IND should be numbered "Serial number: 0000." The next submission (e.g., amendment, report, or correspondence) should be numbered "Serial Number: 0001." Subsequent submissions should be numbered consecutively in the order in which they are submitted..

Serial Number: 0 0 0 0

11. This submission contains the following *(Select all that apply)*

- ☑ Initial Investigational New Drug Application (IND)
- ☐ Request For Reactivation Or Reinstatement
- ☐ Development Safety Update Report (DSUR)
- ☐ Response to Clinical Hold
- ☐ Annual Report
- ☐ Other *(Specify)*:
- ☐ Response To FDA Request For Information
- ☐ General Correspondence

**Protocol Amendment**
- ☐ New Protocol
- ☐ Change in Protocol
- ☐ New Investigator
- ☐ PMR/PMC Protocol
- ☐ Human Factors Protocol

**Information Amendment**
- ☐ Chemistry/Microbiology
- ☐ Pharmacology/Toxicology
- ☐ Clinical/Safety
- ☐ Statistics
- ☐ Clinical Pharmacology

**Request for**
- ☐ Meeting
- ☐ Proprietary Name Review
- ☐ Special Protocol Assessment
- ☐ Formal Dispute Resolution

**IND Safety Report**
- ☐ Initial Written Report
- ☐ Follow-up to a Written Report

12. For Originals, is the product a combination product (21 CFR 3.2(e))? ☐ Yes ☑ No

Combination Product Type *(See instructions)*

Request for Designation (RFD) Number

13. Select the following only if applicable. *(Justification statement must be submitted with application for any items selected below. Refer to the cited CFR section for further information.)*

- ☐ Emergency Research Exception From Informed Consent Requirements, 21 CFR 312.23 (f)
- ☐ Charge Request, 21 CFR 312.8

*Expanded Access Use, 21 CFR 312.300*
- ☐ Individual Patient, Non-Emergency 21 CFR 312.310
- ☐ Individual Patient, Emergency 21 CFR 312.310(d)
- ☐ Intermediate Size Patient Population, 21 CFR 312.315
- ☐ Treatment IND or Protocol, 21 CFR 312.320

#### For FDA Use Only

| CBER/DCC Receipt Stamp | DDR Receipt Stamp | Division Assignment |
|---|---|---|
| | | IND Number Assigned |

PSC Publishing Services (301) 443-6740 EF

Previous Page | Next Page

14. Contents of Application – This application contains the following items *(Select all that apply)*

- [x] 1. Form FDA 1571 *(21 CFR 312.23(a)(1))*
- [x] 2. Table of Contents *(21 CFR 312.23(a)(2))*
- [x] 3. Introductory statement *(21 CFR 312.23(a)(3))*
- [x] 4. General Investigational plan *(21 CFR 312.23(a)(3))*
- [ ] 5. Investigator's brochure *(21 CFR 312.23(a)(5))*
- [ ] 6. Protocol *(21 CFR 312.23(a)(6))*
  - [ ] a. Study protocol *(21 CFR 312.23(a)(6))*
  - [ ] b. Investigator data *(21 CFR 312.23(a)(6)(iii)(b))* or completed Form FDA 1572
  - [ ] c. Facilities data *(21 CFR 312.23(a)(6)(iii)(b))* or completed Form FDA 1572

6. Protocol *(Continued)*
  - [ ] d. Institutional Review Board data (21 CFR 312.23(a)(6)(iii)(b)) or completed Form FDA 1572
- [ ] 7. Chemistry, manufacturing, and control data *(21 CFR 312.23(a)(7))*
  - [ ] Environmental assessment or claim for exclusion *(21 CFR 312.23(a)(7)(iv)(e))*
- [ ] 8. Pharmacology and toxicology data (21 CFR 312.23(a)(8))
- [ ] 9. Previous human experience *(21 CFR 312.23(a)(9))*
- [ ] 10. Additional information *(21 CFR 312.23(a)(10))*
- [ ] 11. Biosimilar User Fee Cover Sheet *(Form FDA 3792)*
- [ ] 12. Clinical Trials Certification of Compliance *(Form FDA 3674)*

15. Is any part of the clinical study to be conducted by a contract research organization? [ ] Yes [x] No

If Yes, will any sponsor obligations be transferred to the contract research organization? [ ] Yes [ ] No

If Yes, provide a statement containing the name and address of the contract research organization, identification of the clinical study, and a listing of the obligations transferred *(use continuation page)*.

Continuation Page for #15

16. Name and Title of the person responsible for monitoring the conduct and progress of the clinical investigations

Ari Grinspan MD, Associate Professor, Icahn School of Medicine at Mount Sinai

17. Name and Title of the person responsible for review and evaluation of information relevant to the safety of the drug

Ari Grinspan MD, Associate Professor, Icahn School of Medicine at Mount Sinai

**I agree not to begin clinical investigations until 30 days after FDA's receipt of the IND unless I receive earlier notification by FDA that the studies may begin. I also agree not to begin or continue clinical investigations covered by the IND if those studies are placed on clinical hold or financial hold. I agree that an Institutional Review Board (IRB) that complies with the requirements set forth in 21 CFR Part 56 will be responsible for initial and continuing review and approval of each of the studies in the proposed clinical investigation. I agree to conduct the investigation in accordance with all other applicable regulatory requirements.**

18. Name of Sponsor or Sponsor's Authorized Representative

Ari Grinspan

19. Telephone Number *(Include country code if applicable and area code)*

212-241-7757

20. Facsimile (FAX) Number *(Include country code if applicable and area code)*

646-537-8921

21. Address

Address 1 *(Street address, P.O. box, company name c/o)*: One Gustave L. Levy Place

Address 2 *(Apartment, suite, unit, building, floor, etc.)*: Box 1069

City: New York

State/Province/Region: New York

Country: USA

ZIP or Postal Code: 10029

22. Email Address

23. Date of Sponsor's Signature *(mm/dd/yyyy)*

11/02/2022

24. Name of Countersigner

25. Address of Countersigner

Address 1 *(Street address, P.O. box, company name c/o)*:

Address 2 *(Apartment, suite, unit, building, floor, etc.)*:

City:

State/Province/Region:

Country: United States of America

ZIP or Postal Code:

26. Email Address

**WARNING : A willfully false statement is a criminal offense (U.S.C. Title 18, Sec. 1001).**

27. Signature of Sponsor or Sponsor's Authorized Representative

Ari Grinspan Digitally signed by Ari Grinspan Date: 2022.11.02 14:58:55 -04'00'

Sign

28. Signature of Countersigner

Sign

**The information below applies only to requirements of the Paperwork Reduction Act of 1995.**

The burden time for this collection of information is estimated to average 100 hours per response, including the time to review instructions, search existing data sources, gather and maintain the data needed and complete and review the collection of information. Send comments regarding this burden estimate or any other aspect of this information collection, including suggestions for reducing this burden to the address to the right:

*"An agency may not conduct or sponsor, and a person is not required to respond to, a collection of information unless it displays a currently valid OMB number."*

Department of Health and Human Services
Food and Drug Administration
Office of Operations
Paperwork Reduction Act (PRA) Staff
**

***Please do NOT send your completed form to this PRA Staff email address.***

### 2 Table of Contents [21 CFR 312.23(a)(2)]

**1 FDA Form 1571 [21 CFR 312.23(a)(1)] ii**

**2 Table of Contents [21 CFR 312.23(a)(2)] iii**

**3 Introduction [21 CFR 312.23(a)(3)] 1**

3.1 Investigational Agent 2
3.2 Dose and Exposure 2
3.3 Overview of Previous Human Experience 2
3.4 Overview of Preclinical Data 2

**4 General Investigational Plan [21 CFR 312.23(a)(4)] 2**

4.1 Research Rationale and Objectives 2
- The change in the mucosal immune profile before and after FMT 3
4.2 Proposed clinical research 3
4.3 Anticipated risks from study drug 3

**5 Investigator's Brochure [21 CFR 312.23(a)(5)] 3**

**6 Protocol [21 CFR 312.23(a)(6)] 3**

6.1 Investigator and Facilities Data 3
*6.1.1 Form FDA 1572 3*
*6.1.2 Sponsor-Investigator Credentials 3*
*6.1.3 Subinvestigator(s) Credentials (CVs) 3*
*6.1.4 Disclosure of Financial Interests 3*
6.2 Clinical Trial Registration 3

**7 Chemistry and Manufacturing [21 CFR 312.23(a)(7)] 3**

7.1 General Method of Preparation and packaging 3
7.2 Drug Components and Drug Product 4
7.3 Placebo Product 4
7.4 Labeling 4
7.5 Environmental Analysis Requirements 4

**8 Pharmacology and Toxicology [21 CFR 312.23(a)(8)] 4**

8.1 Pharmacodynamics 4
*8.1.1 Primary pharmacodynamics 4*
8.2 Safety pharmacology 4
8.3 Pharmacokinetics 5
*8.3.1 Absorption 5*
*8.3.2 Distribution 5*
*8.3.3 Metabolism 5*
*8.3.4 Excretion 5*
8.4 Pharmacology Summary 5
8.5 Pharmacology Conclusions 5
8.6 Toxicology 5
8.7 Genetic Toxicology 5
8.8 Carcinogenicity 5
8.9 Reproductive and Developmental Toxicology 6
8.10 Special Toxicology Studies 6
8.11 Toxicology Summary 6

8.12 Toxicology Conclusions 6

**9 Previous Human Experience with the Investigational Agent [21 CFR 312.23(a)(9)] 6**

9.1 Prior Clinical Research Experience 6
9.2 Clinical Care Experience 6

**10 Additional Information [21 CFR 312.23(a)(10)] 6**

**11 Bibliography 6**

**12 Attachments 6**

### 3 Introduction [21 CFR 312.23(a)(3)]

We are applying for IND approval for a phase 1b study in recurrent *Clostridioides difficile* infection (rCDI) comparing a defined live biotherapeutic product (LBP), MTC01, derived from a frequently successful fecal microbiota transplantation (FMT) donor (D283) compared with FMT product from the donor D283. Specifically, we will examine the efficacy and safety of MTC01 and the FMT for rCDI as well as investigate the engraftment of each bacterial strain when administered via a defined bacterial consortia or fecal slurry (FS).

The remarkable efficacy of Fecal Microbiota Transplantation (FMT) for the treatment of *Clostridioides difficile* infection (CDI) with cure rates often >90% represents the greatest clinical demonstration to date of the potential for gut microbiota manipulation to impact health. Although this success was evident even in an early randomized clinical trial comparing FMT with antibiotic standard of care [1], the subsequent years have been highly productive in determining key aspects of such therapies including that colonic administration appears to outperform enema [2] and that the gut microbiota 24-72 hr post-transplant typically is restored to contain members of the bacterial groups in healthy individuals [3,4]. FMT studies have also shown that single-dose oral administration in capsules may be less effective than colonoscopic administration, while multiple oral doses can achieve non-inferiority with FMT [5,6].

With more widespread use of FMT for rCDI, some of the challenges of this therapeutic modality are emerging. First it is difficult to scale as it requires a constant supply of human stool donations, safety screening of every donor microbiota, and labor-intensive processing. These challenges are exacerbated in indications beyond rCDI, such as ulcerative colitis, where it appears multiple FMT doses may be more effective than single doses [7–9]. A second challenge has been the emergence of serious adverse events (SAEs) associated with the transmission of opportunistic pathogens from the donor to the recipient, including extended spectrum beta-lactamases (ESBL)-producing *E. coli* [10,11], enteropathogenic *E. coli* (EPEC), and Shigatoxin-producing *E. coli* (STEC) [12], as well as the potential risk of transmitting SARS-CoV-2. Although rare, these SAEs can be life threatening to the affected individuals, and each potentially new opportunistic pathogen identified increases the cost to screen FMT donors and decreases the number of donors that pass these screens, thereby reducing the amount of available “drug” in the human population. Finally, the identification of safety concerns is a challenge with an undefined FMT product, as few FMT recipients receive the same gut microbiota from the same donor, and therefore only the most penetrant and severe adverse events can be associated with a given donor microbiota.

These challenges with FMT have spurred interest in developing defined consortia of fecal bacteria that could be used as a synthetic alternative to FMT termed a defined live biotherapeutic product (LBP). Defined LBPs would be more scalable, could avoid including multi-drug resistant organisms (MDROs) or strains associated with SAEs, and provide far greater ability to control community composition and gain mechanistic insights. To date, there are a limited number of published studies of multi-strain bacterial LBPs used for the treatment of rCDI, including a 10-member [13], 12-member [14,15], 33 member [16], and 40 member [17] community. Although these studies were important demonstrations of feasibility and safety, our current rate of four published LBP clinical studies for rCDI in the past 31 years is not a promising road to rapid discovery.

This difference in progress between FMT and LBPs is in part due to the far higher manufacturing challenges associated with LBPs. For a given LBP, each component strain must be manufactured at scale and to a clinical grade, which includes safety, stability, purity, and potency checks. Although our knowledge of microbiome culturing has increased tremendously in the past decade [18–22], there is little published know-how for manufacturing clinical grade bacteria beyond common probiotic strains. Industry trials often remain unpublished, and if they are published, key aspects of manufacturing know-how are unlikely to be provided. Therefore, there is a large and unexplored niche of small, innovative LBP human clinical trials for understanding the mechanisms of gut microbiota manipulation as a therapeutic modality. If these studies are performed in academia with sharing of know-how similar to FMT, it would accelerate the translation of the large investment in human microbiome research into clinical care for both academia and industry.

A key open question in the field is if LBP bacterial strains engraft as well as those in FMT. In an LBP, the manufacturing process allows control over the growth parameters of each strain. Particularly if the strains are grown individually, they can be combined evenly by potency or any other desired metric to improve engraftment. However, when the strains are administered in the context of a fecal slurry, they have the advantage of being present in their native nutrient environment with the complete set of bacterial community members that they have stably co-evolved with. In this trial, we used our success with prior FMT donor D283 to design a defined consortium of 15 bacterial strains isolated from this donor that are frequent in the healthy human gut microbiota (i.e., present in >25% of individuals) and that engrafted in the majority of recipients in prior fecal transplants. We will compare this 15-member cocktail, MTC01, with fecal slurry from the same donor in a randomized open-label trial in 60 individuals with rCDI. For each drug modality (defined LBP vs FMT), we will test either a high dose based on the common dose of $1x10^{11}$ that we successfully use for FMT at Mount Sinai and at a low dose $1x10^{10}$ that would allow significantly lower cost of goods and manufacturability in future trials. We will compare the safety, efficacy, and strain engraftment across the two doses and two drug types in 60 total subjects with 15 subjects for each dose (high/low) and drug type (defined LBP / FMT).

##### *3.1 Investigational Agent*

The investigational agent is MTC01, a defined live biotherapeutic product (LBP) composed of 15 gut bacterial strains isolated from a human fecal transplant donor. A comparator agent will be FMT performed with donor fecal slurry from the same individual from which MTC01 was isolated.

##### *3.2 Dose and Exposure*

- MTC01 and human donor stool slurry; each agent will be provided by a single administration of either a high dose ($1x10^{11}$) or a low dose ($1x10^{10}$) with a specification of $6x10^{10}$ – $5x10^{11}$ and $6x10^{9}$ – $5x10^{10}$ respectively for high dose and low dose
- Formulation: the formulation buffer for both agents will be Phosphate Buffered Saline (9 g NaCl/L) + 30% glycerol + 0.5% cysteine (all USP grade)
- Route of administration: Colonoscopy to the right colon
- Planned exposure: Single dose via colonoscopy

The dose is based on the CFU range commonly used in successful FMT, the mean dose used by this donor in the past, and ranges tested in other phase I LBP studies. The high dose is designed to be near the median CFU of our previous successful FMT ($6x10^{11}$), while remaining within the practical limits of our current manufacturing facility. While the low dose is still almost an order of magnitude higher than the lowest successful dose in an FMT at Mount Sinai ($3x10^{9}$). Given that successful FMT has been performed with far larger amounts of FS (and thereby higher CFU) than this study, we do not expect dose to be a factor in the safety profile for either the high or low dose.

##### *3.3 Overview of Previous Human Experience*

**Other defined bacterial consortium tested in human rCDI**

FMT has been safely used in tens of thousands of individuals providing a rich resource of potential SAEs including death [10]. All major SAEs from FMT have involved MDROs or pathogenic strains of *E. coli*. These two concerns can be largely eliminated in the context of LBPs.

FMT has been used extensively in the treatment of rCDI. There are a limited number of published studies of multi-strain LBPs used for the treatment of *C. difficile*. In the Tvede and Rask-Madsen study described above, five individuals with CDI were treated with a rectal infusion of 10 cultured bacteria ($2x10^{9}$ CFU per strain; $2x10^{10}$ total dose) with all five patients responding and no SAE [13]. Lactobacillus rhamnosus and Lactobacillus acidophilus were added to the mixture in the late 1990s and it was subsequently reported this 12-strain mixture was given to 55 rCDI patients by enema at a dose of approximately $4x10^{11}$ CFU/dose and at least $4x10^{9}$ of each strain with a success rate of 63.6% [15]. This consortium was subsequently tested in an open-label 3-arm randomized control trial with 31 subjects receiving vancomycin alone, 31 subjects receiving vancomycin followed by 3 enema infusions of the 12-strain consortium, and 34 subjects receiving vancomycin followed by 1-3 enema infusions of FMT (50 grams stool/dose). FMT was provided as a rescue therapy to the defined consortia and failures did not receive additional defined consortia. FMT following vancomycin was superior to vancomycin alone but the defined consortium combined administered after vancomycin was not superior to vancomycin alone. 10% of subjects had serious adverse events. None of the hospital admissions were related to either FMT or defined consortium except a potentially related case of suspected urosepsis in the defined consortium group [14]. In a long-term follow-up of results for up to 5 years with this defined consortium compared with FMT, the result in FMT was not associated with increased mortality, risk of hospital admission or onset of disease following treatment compared to the 12-member consortium [23].

Petrof et.al., treated two subjects with rCDI with a consortium of 33 bacterial isolates from a healthy donor stool [16]. The strains were administered by colonoscopy to the right and mid colon. Both patients responded with no SAE and remained symptom free for 6 months. The microbes were administered in 100 ml of pre-reduced saline at a concentration of $3.5x10^{9}$ CFU/ml giving an overall total dose of $3.5x10^{11}$. A follow-up study with some of the same authors and the biotech company Nubiyota explored a 40 member consortium (MET-2) administered orally to 20 individuals at a dose of $1x10^{10}$ for 2 days followed by a dose of $3x10^{9}$ for 3 days and an optional follow-up dosing upon relapse. They found 15 of 19 individuals (79%) receiving MET-2 achieved the primary outcome of no CDI recurrence by day 50 with no short-term safety concerns. In healthy volunteers, Vedanta Biosciences recently published results demonstrating engraftment of a multi-strain defined 8-strain LBP, VE303, at single doses ranging from $1.6x10^{9}$ to $8x10^{9}$ CFU and cumulative doses of $4x10^{10}$ to $1.1x10^{11}$ over five or 14 days with no drug related SAEs across 23 healthy volunteers [24]. According to non-peer reviewed press releases, they have also described VE303 as providing benefit in the context of rCDI. Although the numbers are too small to compare with the far more utilized FMT approach in rCDI, we would expect drug-related SAEs to be exceptionally low for a defined LBP given the safety record of FMT in generally and the mitigation of the major risks in FMT by using a defined LBP. To date, this expectation appears to be the case. Importantly, no study has of yet compared FMT to a defined LBP using the same donor for the FMT and the LBP to determine if the strain engraftment, which we have demonstrated is directly related to efficacy [21], differs between the two drug types.

**MTC01 for rCDI**

Although MTC01 has never been manufactured and provided to humans, the 15 members of MTC01 have been transplanted to seven individuals in the context of FMT. And although stool was only collected to enable confirmation of strain engraftment in seven recipients, donor D283 was used in a total of 25 recipients with a 84% success rate. The comparator FMT group will receive a fecal slurry from the same donor at the same concentrations and in the same formulation buffer as MTC01. The sponsor has extensive experience with FMT studies for rCDI [21,25–30] including the studies with donor D283 that led to the current phase 1b trial [21,25].

##### 3.4 *Overview of Preclinical Data*

Given the large literature demonstrating the efficacy of FMT for human rCDI, the growing support for defined communities for human rCDI, the demonstration that even small communities can be successful for CDI in mouse models [24,31], and the prior successful usage of stool from the FMT from the donor to be used in this study (D283), we have not tested the FMT or MTC01 in mice models of CDI. Rather we designed MTC01 by identifying the strains that transferred most frequently from FMT donor D283 to successful rCDI FMT recipients [21].

To determine the potential immunomodulation potential of MTC01 as well as to confirm that it did not drive tissue inflammation in the intestine, we colonized germ-free mice with 1.5 x $10^8$ CFU of MTC01 by oral gavage (Figure 1). The strains colonized at relative abundances that were expected based on previous gnotobiotic colonizations with strains from larger donor D283 cultured communities and other cultured communities of bacteria isolated from humans in our published work (Figure 1C; [32,33]). MTC01 robustly induced regulatory T cells in the colonic lamina propria and did not drive histological inflammation (Figures 1F and 1G). Tissue sections were scored using the method of Koelink et.al., 2018 [34]

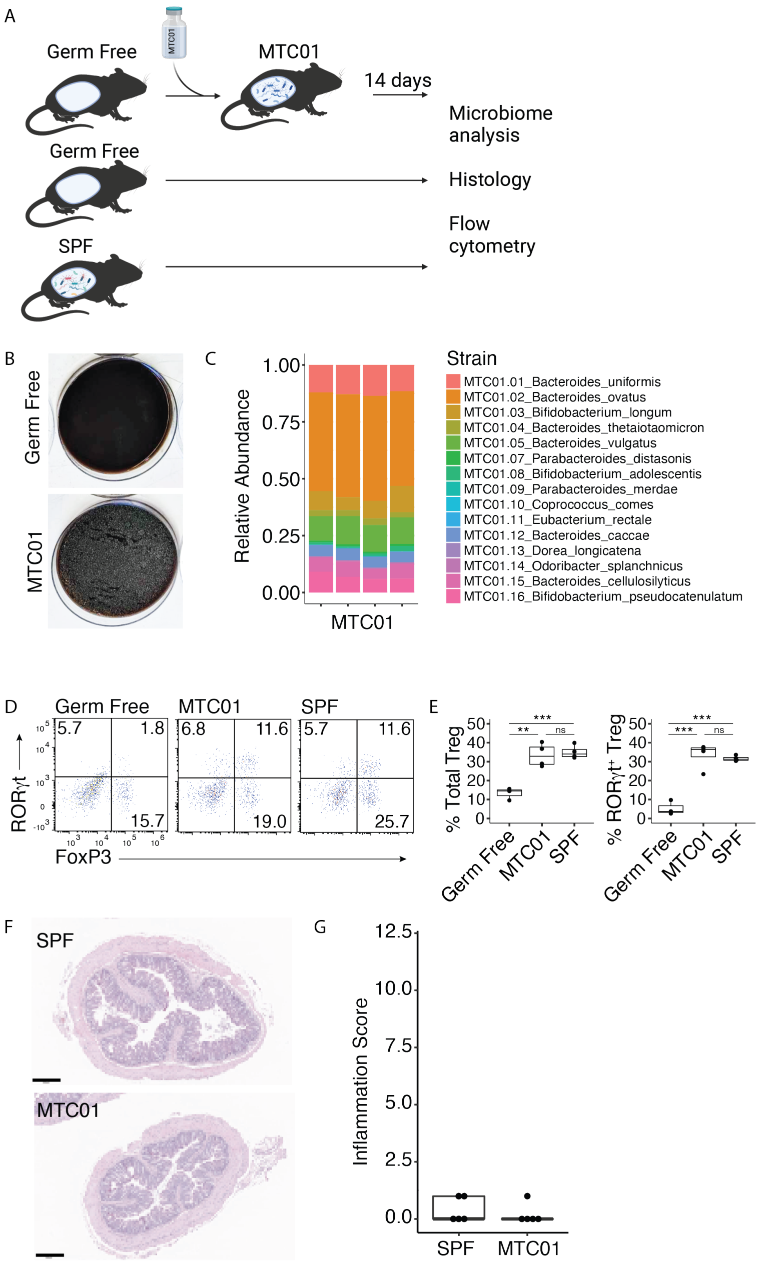

**Figure 1.** (A) We colonized germ-free mice with MTC01. Fourteen days post colonization we (B) verified the colonization status of the germ-free control and MTC01 gnotobiotic mice respectively. (C) The relative abundances of MTC01 strains were as expected from prior studies [32,33] with colonization dominated by Bacteroides. (D-E) MTC01 robustly induced regulatory T cells to comparable levels of specific pathogen free (SPF) mice. (F-G) No histological inflammation was observed in the intestine of MTC01 colonized mice.

### 4 General Investigational Plan [21 CFR 312.23(a)(4)]

#### 4.1 Research Rationale and Objectives

This phase 1b trial is focused on a safety assessment of using a defined LBP, MTC01, compared to FMT sourced from the donor from which MTC01 was isolated. By choosing the best engrafting strains from a successful FMT donor, D283, used to treat multiple individuals with no SAE and eliminating those species that do not engraft well, we expect to have a similar efficacy to stool but with a far more controlled and defined safety profile as well as improved scalability. If a defined consortium performs similarly to FMT it will also provide insights into potential mechanisms of action and insights into characteristics of what is the minimal community of strains needed for successful FMT.

The primary endpoint measure and assessment is: the number of patients with MTC01 and FMT related adverse events through week 8 classified according to the Medical Dictionary for Regulatory Activities (MedDRA) and categorized according to the Common Terminology Criteria for Adverse Events (CTCAE).

Given the typically high response rate to FMT, we also expect to gain important insights into secondary outcomes of efficacy and microbial engraftment.

The secondary endpoint measures and assessments are:
- The number of patients experiencing a CDI recurrence through week 8
- The number of patients with a negative C. difficile test result 8 weeks post treatment
- The number of patients with absence of diarrhea 8 weeks post treatment
- The proportion of MTC01 strains engrafting when transmitted via LBP
- The proportion of MTC01 strains engrafting when transmitted via D283 donor stool
- The proportion of non-MTC01 strains engrafting when transmitted via D283 donor stool

#### 4.2 Proposed clinical research

We propose a single center, open-label pilot study to evaluate the efficacy and safety of MTC01 compared to healthy donor FMT from the donor whose stool was used to isolate the strains in MTC01. We will also evaluate the differences in engraftment between the strains transmitted via defined LBP and FMT. The products will be administered to individuals $\geq$18 years old with recurrent *C. difficile* infection. FMT will be derived from previously successful donor D283 and delivered via colonoscopy to the right colon. If D283 stool does not pass the donor screening criteria at the time of the study, we will alternatively use FMT material from an approved, screened donor. We plan to enroll 60 patients with rCDI (25% low dose MTC01, 25% low dose FMT, 25% high dose MTC01, 25% high dose FMT).

#### 4.3 Anticipated risks from study drug

Related or possibly related Adverse Events (AEs) of significance include: bloating, distention, flatulence, diarrhea, constipation, abdominal pain, fever, vomiting, bacteremia, infection transmission.

### 5 Investigator's Brochure [21 CFR 312.23(a)(5)]

**See Section 12, Attachments.**

### 6 Protocol [21 CFR 312.23(a)(6)]

**See Section 12, Attachments.**

#### *6.1 Investigator and Facilities Data*

##### 6.1.1 Form FDA 1572

##### 6.1.2 Sponsor-Investigator Credentials

##### 6.1.3 Subinvestigator(s) Credentials (CVs)

##### 6.1.4 Disclosure of Financial Interests

#### *6.2 Clinical Trial Registration*

### 7 Chemistry and Manufacturing [21 CFR 312.23(a)(7)]

**MTC01**

The Drug Product (DP) MTC01 is generated by combining the drug substance for each of 15 individually cultured strains of bacteria that were previously isolated from FMT donor D283. MTC01 is generated in the Microbiome Translational Center (MTC) in the Icahn School of Medicine at Mount Sinai using established SOPs and MBRs. The MTC is located at:

Microbiome Translational Center
Icahn School of Medicine at Mount Sinai
1470 Madison Ave 8-201
New York, NY 10029

The MTC01 DP is at a potency of $1x10^{11}$ CFU/dose in a volume of 250ml (high dose) or $1x10^{10}$ CFU/dose in a volume of 250ml (low dose). The high dose specification is $6x10^{10}$-$5x10^{11}$, and the low dose specification is $6x10^{9}$-$5x10^{10}$

**FMT**

The fecal slurry (FS) used for FMT in this study will be generated by the homogenization of feces from pre-screened donor individual D283, whose microbiome was previously used for numerous successful rCDI FMTs. The filtered FMT drug product will be at the same volume (250 ml) and potency as MTC01 (high dose = $1x10^{11}$ CFU with a specification of $6x10^{10}$ - $5x10^{11}$; low dose = $1x10^{10}$ CFU with a specification of $6x10^{9}$-$5x10^{10}$).

The FMT drug will be generated in the Microbiome Translational Center (MTC) in the Icahn School of Medicine at Mount Sinai using established SOPs and MBRs. The MTC is located at:

Microbiome Translational Center
Icahn School of Medicine at Mount Sinai
1470 Madison Ave 8-201
New York, NY 10029

Prior to release for processing of stool samples into FS for use in FMT, donor samples will be banked and must pass safety screens. The details of these screens are in the clinical protocol.

Briefly, blood tests will be performed on the donor 14 days before collection and 30 days after collection for:

- CBC w/ differential
- LFTs, complete
- Bun/Cr
- Fasting glucose
- High Sensitivity CRP
- Serum triglycerides, LDL, HDL cholesterol
- HIV 1/2 (IgG) and E1A
- HAV (IgG and IgM)
- HBV (surface antigen/antibody; core antibody)
- HCV (IgG)
- Syphilis serology (Trep-sure ELISA; RPR done if treponemal test is positive)
- HTLV 1&2

Stool tests will be performed on days 1 and 30 of collection for:
- Clostridium difficile toxin by PCR
- Fecal Giardia antigen
- Fecal Cryptosporidium antigen
- Ova and parasites (light microscopy)
- Helicobacter pylori fecal antigen
- NAAT/Biofire
- VRE
- MSRA
- ESBL

Finally, FMT donors will be tested for SARS-CoV-2 at the following time points: 14 days before collection, day 1 of collection, day 15 of collection, day 30 of collection, and then 30 days after final day of collection. Priority will be given to donor D283 given their direct relevance to MTC01. However, if D283 stool and blood fail to pass these required assays we will use a different donor that does pass these criteria. In this case, the secondary aim of comparing engraftment of MTC01 strains will be performed on legacy FMT data from this donor [21].

#### *7.1 General Method of Preparation and packaging*

##### MTC01

##### *Overview of MTC01 bacterial strain selection and manufacturing*

All strains in MTC01 were isolated from stool samples of a frequently successful rCDI FMT donor (D283) at Mount Sinai Hospital (Appendix 1 and 2). Various selective media were used to enrich different bacterial groups (Table 1). Each strain was subsequently tracked via metagenomics using a recently published algorithm that precisely and sensitively detects individual strains in a metagenome using the informative (rare) regions from the genome sequence of the tracked strain [21]. Strains were tracked in multiple D283 FMT recipients for up to five years. The D283 FMT donor was also sampled for 5 years to look at the stability of the strains in the human donor. As expected from our prior research demonstrating that the human gut microbiome is largely stable over time [19], we observed that the large majority of strains isolated and sequenced from donor D283 were present five years after the first FMT sample was obtained from the donor. Importantly, we also discovered that the majority of donor strains engraft in the recipient and that the majority of strains engrafted at 8 weeks are stably maintained in the recipient for years post-FMT without an additional transplant [21] (Investigators Brochure Figure 1).

To avoid the known translation challenges of *in vivo* or *in vitro* models, we used *in human* discovery to design an LBP consisting of all prevalent human gut microbes (i.e., present in >25% of healthy subjects) that stably engraft in a majority of recipients while eliminating multi-drug resistant organisms and species associated with SAE in FMT (e.g., *E. coli*), resulting in a community of 15 strains from donor D283 (Figure 2).

**Table 1.** Initial isolation media for each strain.

| Strain ID | Genus | Species | Media | Passage |
|---|---|---|---|---|
| MTC01.01 | *Bacteroides* | *uniformis* | Chocolate Agar | 3 |
| MTC01.02 | *Bacteroides* | *ovatus* | Brucella LKV Agar | 3 |
| MTC01.03 | *Bifidobacterium* | *longum* | microaerophillic on Columbia CNA w/5% Sheep Blood | 3 |
| MTC01.04 | *Bacteroides* | *thetaiotaomicron* | Brucella LKV Agar | 3 |
| MTC01.05 | *Bacteroides* | *vulgatus* | Columbia CNA w/5% Sheep Blood | 3 |
| MTC01.07 | *Parabacteroides* | *distasonis* | Brucella LKV Agar | 3 |
| MTC01.08 | *Bifidobacterium* | *adolescentis* | Chocolate Agar | 3 |
| MTC01.09 | *Parabacteroides* | *merdae* | Brucella LKV Agar | 3 |
| MTC01.10 | *Coprococcus* | *comes* | Chocolate Agar | 3 |
| MTC01.11 | *Eubacterium* | *rectale* | Chocolate Agar | 3 |
| MTC01.12 | *Bacteroides* | *caccae* | Brucella LKV Agar | 3 |
| MTC01.13 | *Dorea* | *longicatena* | Columbia CNA w/5% Sheep Blood | 3 |
| MTC01.14 | *Odoribacter* | *splanchnicus* | Chocolate Agar | 3 |
| MTC01.15 | *Bacteroides* | *cellulosilyticus* | Brucella LKV Agar | 3 |
| MTC01.16 | *Bifidobacterium* | *pseudocatenulatum* | Chocolate Agar | 3 |

Each 15 of these strains selected for MTC01 was individually manufactured as a DS in a sterilized anaerobic chamber and subsequently combined in a sterilized anaerobic chamber to make the DP to be used in this clinical trial. First, research cell banks (RCB) were made for each of the 15 strains. These RCB stocks were tested for antibiotic resistance internally by Etest and externally by CRO using MIC plating according to CLSI standards [Micromyx] with a high correlation between the two approaches (Figure 2C; Appendix 3). MTC01 strains were also tested for suitability for USP<61> and found suitable (i.e., no growth under the USP<61> conditions and no inhibition of positive control strains) by an external CRO [FOCUS Labs] (Appendix 4**)**. After RCB generation, we moved the manufacturing into individually sterilized anaerobic chambers for each subsequent step (Appendix 5; Figure 3). Under these conditions, we generated master cell banks (MCB) of each strain – all passed USP<61>. We generated DS for each strain – all passed USP<61> (Appendix 4 and 6). We generated the DP by combining the DS in equal potency for 9 of the 15 strains and at maximal possible potency for the 6 strains that could not be practically manufactured at sufficient concentration to reach equal potency (Appendix 6**)**.

All manufacturing of MCB, DS, and DP was performed with sterile single-use growth flasks, filters, and tubes.

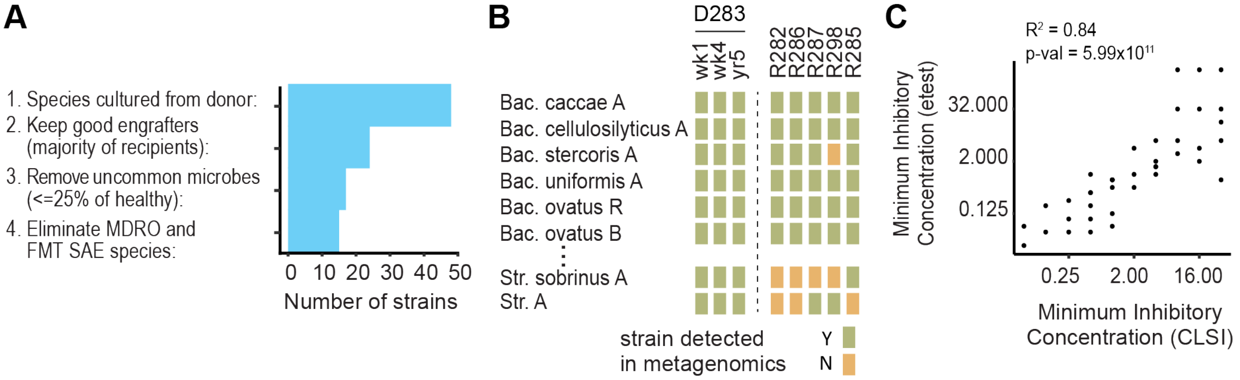

**Figure 2.** (A) We used a series of criteria to choose the subset of strains for inclusion in the LBP for this clinical trial. (B) A key dataset in this selection was the frequency of engraftment of strains isolated from donor D283 whose gut microbiota successfully led to remission of 5 individuals with a single dose. Shown here is a subset of the D283 strains. Notice the donor strains are largely stable over 5 years (i.e., green squares across the first three columns). Strains that did not engraft in the majority of recipients (e.g., <3 green squares out of the five recipients in this figure) were eliminated. (C) An advantage of LBPs is the potential to screen them for any antibiotic resistance concerns. We found similar results between an internal test (Etest) and antibiotic plating dilutions by CLSI standards performed by a CRO.

##### *MTC01 Strain Isolation*

All bacteria were initially isolated from the fecal samples of healthy human FMT donor (D283) that was used to successfully treat multiple rCDI patients [21,25]. The stool samples from the FMT donor were stored at -80$^{o}$C before processing. Subsequently, the stool sample was pulverized under liquid nitrogen and under strict anaerobic conditions ~500mg of pulverized stool from each donor was blended into a fecal slurry (40 – 50 mg/mL) in pre-reduced LYHBHIv4 media (Appendix 9). The fecal slurries were passed through sterile 100 um strainers to remove large debris. Arrayed culture collections were generated for selected donors as previously described[35,36]. Briefly, clarified and diluted donor stool was plated onto a variety of solid selective and non-selective media under anaerobic, micro-aerophilic and aerobic conditions. Plates were incubated for 48-72 hours at 37$^{o}$C. 384 single colonies from each donor microbiota were individually picked and regrown in liquid LYHBHIv4 media for 48 hours under anaerobic conditions. Regrown isolates were identified at the species level using a combination of MALDI-TOF mass spectrometry (Bruker Biotyper) and 16S rDNA amplicon sequencing. All regrown isolates were stored in LYHBHIv4 with 15% glycerol at -80$^{o}$C. All strains for use in MTC01 were genome sequenced with paired-end 150nt reads on an Illumina platform. The solid agar media used for the initial isolation of each strain is listed in Table 1.

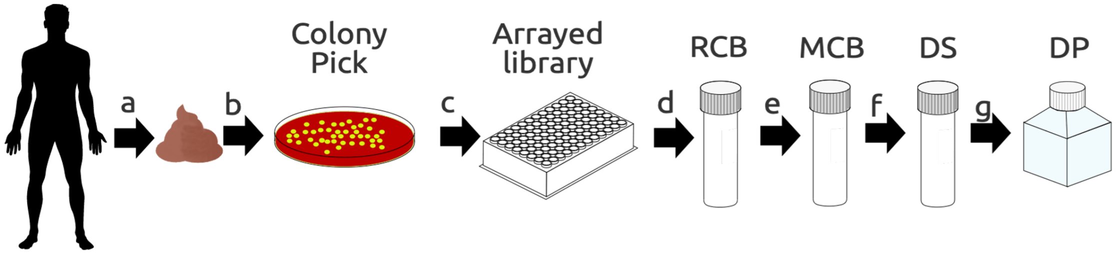

**Figure 3. Initial isolation and cell banking system.**
a) Donor stools were plated under various conditions b) single colonies were picked to generate c) an arrayed culture collection with the unique isolate in each well confirmed by MALDI-TOF, 16S rRNA Sanger sequencing, and genome sequencing d) Research Cell Banks (RCBs) were generated by cryopreservation after colony picking (~20 vials per strain) e) Master Cell Banks (MCBs) were generated from the RCBs (~192 vials per strain) f) Drug Substance (DS) was generated from the MWCBs by inoculation of 1.4 L growth media, harvested after a strain specific time and resuspended into ~140 mL cryo-preservation media (4x 35 mL); an aliquot was taken to determine the potency of the DS (CFU/mL) g) The Drug Product (DP) was generated from the DS by thawing the DS and combining all strains at approximately $3x10^{10}$ CFU per dose for a total dose of approximately $1x10^{11}$ CFU. Manufacturing of the MCB, DS and DP were all performed under sterile conditions using a culture medium free of animal products. DS was washed with USP grade PBS+cysteine and resuspended into USP grade PBS+glycerol+cysteine. The DP was formulated in USP grade PBS+glycerol+cysteine.

##### *Preparation of Research Cell Banks (RCB)*

RCBs were generated from the arrayed culture libraries by plating the original arrayed strain on Chocolate agar. The identity of each strain was assayed by MALDI-TOF and a single colony of similar morphology to the test colony was picked to inoculate 11 mL of LYHBHIv4 growth media (Appendix 9**)**. After 24-48 hours, 0.5 ml of dense liquid bacterial culture was combined with 0.5 ml of 30% glycerol, for a final volume of 1 ml 15% glycerol, and aliquoted into 20 x 2.0 mL tubes that were then stored at -80$^{o}$C. The strain identity of each RCB was confirmed from an aliquot of the liquid broth by MALDI-TOF. Antibiotic susceptibility testing was performed on all 15 strains internally by Etest and by MIC plating according to CLSI standards by Micromyx (Appendix 3; Figure 2C). All RCBs are complete and results are in Appendix 6. All strain genomes were analyzed in silico for antibiotic resistance, virulence factors, and mobile genetic elements

Appendix 7 and 8). As detailed below in the safety subsection of the pharmacology and toxicology section, there were no *in vitro* or *in silico* concerns with antibiotic susceptibility.

***Design of the manufacturing suite and cell bank system***

All manipulations of master cell bank strains, drug substance, and drug product took place in a dedicated, sterilized anaerobic chamber with a filtered air supply and sterilization chamber (Figure 4). This manufacturing system was designed by incorporating concepts from anaerobic culturing and germ-free mouse technology. The system uses a vinyl bag under positive pressure with a HEPA filtered air supply to maintain a controlled anaerobic environment. The anaerobic chamber has a sterilization port that enables rapid fogging with ionized hydrogen peroxide [TOMI Steramist] that rapidly kills microbes while sparing electronics. The combination of rapid sterilization between manufacturing runs and a contained anaerobic environment under positive pressure creates an ideal system for manufacturing and has had outstanding containment, as almost all equipment and resources are sterilized in place while the few items that must be imported (e.g., the strains to be cultured) can be surface sterilized and imported through the anaerobic exchange port that also uses HEPA filtered air. Strict aseptic technique was used throughout the manufacturing process to further limit any potential contamination.

In addition to being a completely closed system, the manufacturing chambers are housed in a room that is not used to culture any microbial pathogens. The room is in a secure area that requires card access and is monitored by video camera. The -80°C freezers used are access restricted and dedicated to microbial therapeutics and dedicated shelves for LBPs. The MCB, DS and DP are stored in barcoded cryo-vials or storage containers with all aliquots and subaliquots tracked using FreezerWorks.

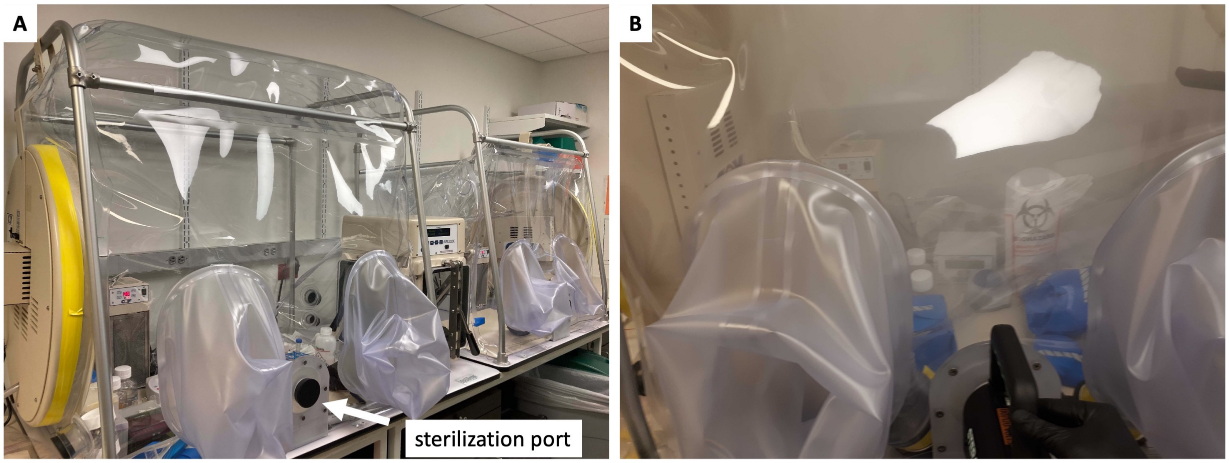

**Figure 4. Anaerobic manufacturing chamber.** (A) In collaboration with Coy Laboratories, we designed a small flexible film anaerobic chamber that has an integrated sterilization port to easily fog with sterilant; the system has two independent chambers. All materials are preloaded into the chamber and sterilized in place. All consumables are purchased sterile with no autoclaved components throughout the manufacturing. For steps, such as centrifugation, that require temporarily removing and returning items from the chamber, the items can be sterilized similarly with a smaller sterilant in the HEPA filtered air exchange port positioned between the two coy chambers. (B) 2x3 minutes of sterilant contact time with a fog of ionized hydrogen peroxide is sufficient for passing all chemical and biological indicator tests, including a *Geobacillus sterothermophilus* spore test.

***Procedures for cleanup and changeover of the anaerobic manufacturing chambers***

At the beginning of the manufacturing process for a new strain (either MCB, DS, or DP), all necessary equipment and consumables are imported into the relevant anaerobic chamber. The chamber has two sides and each is operated independently with a shared, sterilizable air exchange port. All consumables for manufacturing are single-use and no autoclaved components were used in the manufacturing. After all consumables and equipment are brought into the anaerobic chamber, the catalytic fan and dehumidifier are turned off. Iodine test papers are placed into the upper left and upper right corners of the anaerobic chamber to test for effective sterilization.

To sterilize the chamber the rubber stopper in the sterilization port is removed (Figure 4) and ionized hydrogen peroxide (iHP) is sprayed into the chamber for 3-4 minutes. The catalyst fan is then turned on for an iHP dwell time of 5 minutes. After the dwell time is complete, the catalyst fan is again turned off and the chamber fogged for an additional 3-4 minutes. The rubber stopper is then placed into the sterilization port for an iHP dwell time of 10 minutes. During this final 10 minute dwell time the equipment is flipped to ensure contact of iHP to all sides. After the dwell time is complete, the catalytic fan is turned on. The air in the chamber is then exchanged with HEPA filtered nitrogen and then a gas mix with 5% hydrogen to bring the chamber into anaerobic conditions. Once the chamber is anaerobic, a bacterial strain (for MCB or DS) or all bacterial strains (for DP) can be imported into the chamber through the air exchange port to initiate the bacterial growth and mixing steps of the manufacturing.

After a manufacturing production run is complete all reagents and supplies used in the production are removed, and the equipment remaining in the chamber is cleaned with 70% ethanol.

***Preparation of Master Cell Banks (MCB)***

The master cell banks were generated from the RCBs by inoculating a 10 ml starter culture in LYH_VIB media (Appendix 9), after 24-96 hours the starter culture was used to inoculate one flask containing 100 ml LYH_VIB to an optical density ($OD_{600}$) of 0.1-0.2 and incubated at 37$^{o}$C under anaerobic conditions until within 90% of maximum $OD_{600}$. After reaching 90% of maximum $OD_{600}$ measurement, the culture is combined with 100 ml Cryogenic medium (PBS with 0.05% cysteine, 30% glycerol; final glycerol concentration is 15%; Appendix 9;) and stored at -80$^{o}$C as 1 ml aliquots in up to 192 individual air-tight cryovials. All MCBs passed USP<61> [FOCUS labs] (Appendix 4), are in the range of $10^7$-$10^9$ and tested for identity by MALDI-TOF of at least 48 unique colonies with no single contamination. All MCBs are complete and results are (Appendix 6).

***Drug substance (DS)***

To produce DS for each strain, three glycerol stock MCB ($10^7$-$10^9$ CFU in a volume of 1 ml) of a given strain were used to inoculate a starter culture flask with 100 ml of LYH_VIB media and grown 24-96 hr. From this starter culture, a flask containing 1.4 L of LYH_VIB media was inoculated to an $OD_{600}$ of 0.1-0.2 and incubated at 37$^{o}$C under anaerobic conditions until within 90% of maximum $OD_{600}$. After reaching 90% of maximum growth density, the culture was centrifuged and washed with wash media (PBS with 0.05% cysteine; Appendix 9; all USP grade) to remove bacterial bioproducts and the original culture medium. The washed bacteria were centrifuged and resuspended in formulation medium (PBS with 0.05% cysteine, 30% glycerol medium; Appendix 9; all USP grade) at 10X concentration and stored as 30-50 ml aliquots at -80$^{o}$C. 10 x 0.1 mL aliquots were stored at -80$^{o}$C for DS stability assays. Total DS volume for each strain is 130-160 ml and the identity of all strains was tested by MALDI-TOF of at least 48 unique colonies with no single contamination. DS has been generated for all 15 strains and the results are in Appendix 6. All DS passed USP<61> [FOCUS labs] (Appendix 4).

Master batch records and standard operating procedures were used throughout the MTC01 manufacturing process.

| Name | Type | Title |
|---|---|---|

| | | |
|---|---|---|
| MTC-MBR--0001 | MBR | Generation of Master or Working Cell Banks (MCB, WCB) for Live Biotherapeutic Products (LBP) |
| MTC-MBR--0002 | MBR | Culture Media for Live Biotherapeutic Products (LBP) |
| MTC-MBR--0003 | MBR | CFU Count for Live Biotherapeutic Products (LBP) |
| MTC-MBR--0004 | MBR | Biotyping for Live Biotherapeutic Products (LBP) |
| MTC-MBR--0005 | MBR | Sterilization of Anaerobic Chambers for Live Biotherapeutic Products (LBP) |
| MTC-MBR--0006 | MBR | Generation of Drug Substance (DS) for Live Biotherapeutic Products (LBP) |
| MTC-MBR--0007 | MBR | Drug Substance (DS) Culture Media for Live Biotherapeutic Products (LBP) |
| MTC-MBR--0008 | MBR | CFU Count of the Drug Substance (DS) for Live Biotherapeutic Products (LBP) |
| MTC-MBR--0009 | MBR | Biotyping of the Drug Substance (DS) for Live Biotherapeutic Products (LBP) |
| MTC-MBR--0010 | MBR | Sterilization of Anaerobic Chambers for the Drug Substance (DS) of Live Biotherapeutic Products (LBP) |
| MTC-MBR--0011 | MBR | Generation of Drug Product (DP) for Live Biotherapeutic Products (LBP) |
| MTC-MBR--0012 | MBR | DP Buffer for Live Biotherapeutic Products (LBP) |
| MTC-MBR--0013 | MBR | CFU Count of the Drug Product (DP) for Live Biotherapeutic Products (LBP) |
| MTC-MBR--0014 | MBR | Biotyping of the Drug Product (DP) for Live Biotherapeutic Products (LBP) |
| MTC-MBR--0015 | MBR | Sterilization of Anaerobic Chambers for the Drug Product (DP) of Live Biotherapeutic Products (LBP) |
| MTC-SOP--0002 | SOP | Culture Media for Live Biotherapeutic Products (LBP) |
| MTC-SOP--0003 | SOP | CFU Count for Live Biotherapeutic Products (LBP) |
| MTC-SOP--0004 | SOP | Biotyping for Live Biotherapeutic Products (LBP) |
| MTC-SOP--0005 | SOP | Sterilization of Anaerobic Chambers for Live Biotherapeutic Products (LBP) |
| MTC-SOP--0006 | SOP | Generation of Drug Substance (DS) for Live Biotherapeutic Products (LBP) |
| MTC-SOP--0007 | SOP | Drug Substance (DS) Culture Media for Live Biotherapeutic Products (LBP) |
| MTC-SOP--0008 | SOP | CFU Count of the Drug Substance (DS) for Live Biotherapeutic Products (LBP) |
| MTC-SOP--0009 | SOP | Biotyping of the Drug Substance (DS) for Live Biotherapeutic Products (LBP) |
| MTC-SOP--0010 | SOP | Sterilization of Anaerobic Chambers for the Drug Substance (DS) of Live Biotherapeutic Products (LBP) |

| | | |
|---|---|---|
| MTC-SOP--0011 | SOP | Generation of Drug Product (DP) for Live Biotherapeutic Products (LBP) |
| MTC-SOP--0012 | SOP | DP Buffer for Live Biotherapeutic Products (LBP) |
| MTC-SOP--0013 | SOP | CFU Count of the Drug Product (DP) for Live Biotherapeutic Products (LBP) |
| MTC-SOP--0014 | SOP | Biotyping of the Drug Product (DP) for Live Biotherapeutic Products (LBP) |
| MTC-SOP--0015 | SOP | Sterilization of Anaerobic Chambers for the Drug Product (DP) of Live Biotherapeutic Products (LBP) |

**FMT**

The FS FMT product for this study will be generated by homogenization of human donor stool in in formulation buffer composed of Phosphate Buffered Saline (9 g NaCl/L) + 30% glycerol + 0.5% cysteine with glycerol as a cryoprotectant and cysteine as an oxygen scavenger to facilitate the preservation of strict anaerobes (this is the same Formulation medium as the MTC01 DP and DS; Appendix 9). The stool will initially be homogenized in cryopreservation medium with a Stomacher using a filter bag to remove particulate matter. This crude extract will be stored at -80°C and a small aliquot will be used to quantify the concentration of viable bacterial cells per gram in the filtered homogenate. The crude extract will then be diluted to a target dose of $1x10^{11}$ CFU in 250 ml (high dose) or $1x10^{10}$ CFU in 250 ml (low dose) and stored at -80°C until use or expiration.

The stool will initially be homogenized in this formulation buffer with a stomacher using a filter bag to remove particulate matter. To generate concentrated fecal slurry (cFS), the stool sample is homogenized with a stomacher (60 seconds at 210 rotations per minute) after mixing 1:1 volume:grams of health donor stool and formulation buffer. To avoid overfilling the stomacher bag, less volume is used when media (ml) + stool (g) is >400. In this case, we use sufficient cryogenic media to bring the final media + stool to 400. For example, for 300 g of stool use 100 ml of media and for 260g stool we add 140 ml media. This crude cFS extract will be stored at -80C.

A small aliquot of cFS is collected at the time of generation. It is also frozen at -80C and used to quantify the concentration of viable bacterial cells per gram in the filtered homogenate in terms of colony forming units by serial dilution plating on solid Chocolate agar media (Appendix 10). The cFS extract will then be diluted in formulation buffer to a target dose of $1x10^{10}$ CFU/dose (low dose) fecal slurry (FS) or $1x10^{10}$ CFU/dose (high dose) FS in total volume of 250ml and stored at -80C until use or expiration. The SOP for generating cFS and FS are provided in Appendix 11. Given the estimated potency (in CFU) of human stools each dose will contain approximately 7 grams of fecal slurry in a volume of 250 ml formulation buffer. We will not use fecal samples that require using substantial enough amounts of feces that the glycerol concentration drops below 15%.

The normalized filtered FS FMT product will be stored at -80C. The first sample of a series of collections from a given donor will be placed under a stability program and monitored at 1 month, 3 months, 6 months, 12 months, 18 months, and 24 months. Drug from a donor series will be considered expired if the stability reading drops below our minimum spec of $6x10^{9}$ and $6x10^{10}$ for low dose and high dose across three consecutive readings. Based on the extensive literature for FMT in rCDI using healthy donor stool samples, we anticipate the product will be stable at -80C for the entire 24-month testing period.

Master batch records and standard operating procedures are used throughout the manufacturing process.

| Name | Type | Title |
|---|---|---|
| MTC-MBR-FS-0001 | MBR | Generation of CFU normalized Fecal Slurry (FS) for Fecal Microbiota Transplantation (FMT) |
| MTC-MBR-FS-0002 | MBR | Fecal Slurry Cryopreservation Media for Fecal Microbiota Transplantation (FMT) |
| MTC-MBR-FS-0003 | MBR | Sterilization of Biosafety Cabinet and Stomacher for Fecal Slurry Production for Fecal Microbiota Transplantation (FMT) |
| MTC-MBR-FS-0004 | MBR | CFU Count of the Fecal Slurry (FS) for Fecal Microbiota Transplantation (FMT) |
| MTC-SOP-FS-0001 | SOP | [SOP] Materials Management for Fecal Slurry (FS) for Fecal Microbiota Transplantation (FMT) |
| MTC-SOP-FS-0002 | SOP | [SOP] Generation of CFU normalized Fecal Slurry (FS) for Fecal Microbiota Transplantation (FMT) |
| MTC-SOP-FS-0003 | SOP | [SOP] Fecal Slurry Cryopreservation Media for Fecal Microbiota Transplantation (FMT) |
| MTC-SOP-FS-0004 | SOP | [SOP] Sterilization of Biosafety Cabinet and Stomacher for Fecal Slurry Production for Fecal Microbiota Transplantation (FMT) |
| MTC-SOP-FS-0005 | SOP | [SOP] CFU Count of the Fecal Slurry (FS) for Fecal Microbiota Transplantation (FMT) |

**Description of the FMT manufacturing facilities**

Although the equipment for FS generation is used to handle human feces that could contain spore-forming pathogens, we do not culture or isolate spore-forming pathogens in the biological safety cabinet or stomacher used for generating FS. In addition, we store the FS doses in a freezer dedicated to clinical grade microbial therapeutics (fecal slurry and defined consortia). The facilities description is in Appendix 5. The facility for generating MTC01 is physically separate from the facility used to generate FS for FMT. They only brought in close proximity in the -80C freezer for long-term storage. However, even in the -80C freezer they are stored on separate shelves and have visually distinct labels (section 7.4 below).

#### *7.2 Drug Components and Drug Product*

**MTC01**

##### *Drug Product*

To make a batch of drug product, we thawed DS from all 15 strains and combined them in equal proportion to their CFU where possible and maximum volume where not possible (Appendix 6). The final suspension of the high dose MTC01 is $1x10^{11}$ per dose with $3x10^{10}$ of 9 strains, $7.8x10^{8}$ of strain MTC01.12, $1.6x10^{10}$ of strain MTC01.03 and MTC01.13, $2x10^{10}$ of strain MTC01.09 and MTC01.10 and $2.8x10^{10}$ of strain MTC01.01 in a total volume of 250 ml of formulation buffer (PBS with 0.05% cysteine, 30% glycerol; Appendix 9; all USP grade) and stored at -80°C. Low dose DP was made in an identical manner but with $1:10^{th}$ the amount of drug substance per strain for a final dose of $1x10^{10}$.

##### *Identity*

Each of the 15 strains in MTC01 (Table 1) were manufactured separately in anaerobic chambers sterilized with ionized hydrogen peroxide [TOMI Steramist] (Figure 4) for both the Master Cell Banks (MCB) and the Drug Substances (DS). Identity of the MCB and DS was determined by plating an aliquot on rich media (chocolate agar) followed by reading 96 colonies (or at least 48 reads with high quality spectra) by MALDI-TOF (Appendix 6). All high quality MALDI-TOF spectra matched the correct MTC01 strain during MCB and DS manufacturing as expected.

Purity, identity and mixing of the microbes in the Drug Product (DP) was determined by plating a proportion of the DP and identification reading of 288 colonies (or at least 200 colonies with high quality spectra) by MALDI-TOF. All high quality MALDI-TOF spectra matched MTC01 strains as expected.

##### *Potency*

The strength of the DS and DP were determined by plating on chocolate agar to determine the CFU/mL (Appendix 6).

##### *Purity*

Each MCB and DS was plated on rich chocolate agar and 96 colonies (or at least 48 high-quality spectra) were read by MALDI-TOF. For each MCB and DS, only the correct strain was detected (Appendix 6). In addition, at least 200 high-quality MALDI-TOF spectra were detected from individual colonies from the DP. All colonies matched only species in MTC01 (Appendix 6). The microbes in MTC01 are strict anaerobes and the cocktail contains no fungi. The contract research organization (FOCUS Labs) confirmed that no MTC01 strain grows under the aerobic conditions necessary for USP<61> testing. In addition, FOCUS labs determined for each strain and the mixture of all strains in MTC01 that the positive control organisms of the USP<61> assay are not inhibited by any or all of the MTC01 strains. All MCB, DS and the DP passed the USP<61> conditions required for a rectally administered product of $10^3$ CFU/ml for Total Aerobic Microbial Count and $10^2$ CFU/ml for Total Combined Yeasts/Molds Count with no colony detected, as expected [FOCUS labs] (Appendix 4).

##### *Stability*

The stability of each of the 15 strains individually stored at -80°C in the same formulation buffer as the DS and DP has been measured for 48 weeks with little to no loss in viability (Appendix 12). We will continue to monitor the stability of these strains at 96 weeks and 144 weeks to inform the shelf-life of MTC01. Since each strain undergoes two total freeze-thaws prior to administration (the same as the FMT product), we also determined the impact of multiple freeze-thaw cycles on each strain (Appendix 12). Most strains were unaffected, while strains MTC01.03, MTC01.09, MTC01.12 and MTC01.16 appear to have some loss due to freeze thaw. However, this loss is less than one log (Appendix 6). In addition, each strain is left at room temperature during the mixing of DS into DP. Therefore, we also tested the stability of each strain under anaerobic conditions in formulation buffer and found no or minor losses in viability. Finally, the DP must be thawed prior to use in the clinic and must be then poured into a clinical tray where it remains briefly during aspiration into the syringes used to administer MTC01 by colonoscopy. Given the need for flexible time windows when performing clinical procedures, we quantified a large variation in times for these two clinical steps and found the DP is viable in the cryogenic storage bottle post freeze-thaw for 4 hours and in the clinical tray for at least 60 minutes.

Stability testing on the DP was initiated upon the generation of the first batch of DP on Aug 12, 2022. We have measured stability at 0 wks and 4wks with no detected difference in viability, as expected. We will continue to measure the stability of MTC01 DP at 12, 24, 48, 72 and 96 wks.

**FMT**

FMT drug products are complex in nature and vary depending on the human donor used for the transplant. However, as we have previously demonstrated in the context of healthy individuals in general [19] and FMT specifically [21], fecal samples coming from the same individual over time are quite stable, and a majority of the strains remaining the same across periods of multiple years.

Specifically, in this trial we will focus on donor D283 stool, which was previously used to treat several individuals with rCDI. In addition, we have recently obtained a sample from this individual to verify that all of the strains in MTC01 have been stably maintained in donor D283 over the 5 years since the first subject was administered their donor stool. Therefore, it is highly likely that MTC01 strains will remain in this donor throughout the rest of the study, although this will be tested to verify. It is exceedingly rare to find unrelated individuals that share the same strains of bacteria [19,21,37] and likely impossible that any other individual contains all the strains in MTC01 (except the previous recipients of D283 FMT). Therefore, we strongly prefer to use D283 as the donor in this study. If this is not possible due to logistic challenges or safety screen failures, we will try to limit the number of donors to as few as possible to have an opportunity to observe the same donor strains engrafting into multiple recipients.

The defined components of the drug product will be the same formulation medium used for MTC01 (PBS with 0.05% cysteine and 30% glycerol; Appendix 9; all USP grade). The complex components of the drug product will be live microorganisms from durably healthy stool samples. The product will be generated in a sterile, disposable Stomacher filter bag with all manipulations of the open sample performed in a sterilized biological safety cabinet with to produce a final formulation of $1x10^{10}$ low dose and $1x10^{11}$ high dose. The facilities used for the FMT are separate from those used to produce MTC01. FMT will be processed aerobically as is typical in the field and was the case for our previous work with D283, while MTC01 will be generated anaerobically in a sterilized anaerobic chamber dedicated to defined LBP production. The non-disposable objects in the process of generating pFS, consisting of a biological safety cabinet, stomacher, and pipettes, will be sterilized with Quatricide-PV15 [Pharmacal] with a contact time of 10 minutes (SOP and Data sheet in Appendix 13). Quatricide-PV15 is a disinfectant that is a broad range bactericidal, virucidal, and fungicidal containing benzalkonium chloride and dodecyl dimethyl ammonium chloride. The remaining supplies used to generate the fecal slurry are single-use disposable items.

The stability of the FMT product will be monitored by CFU plating of drug product aliquots from the first sample in each series at the time intervals listed above in section 7.1.

**Instructions for use of MTC01 and FMT Drug Product.**
Drug product are stored at -80C until the day of use or expiration (i.e., stability reading drops below our minimum specification of $6x10^{9}$ for low dose and $6x10^{10}$ for high dose across three separate readings). On the day of use, a dose of MTC01 or FMT is thawed in a room temperature water bath and used ideally within 1 hour and not more than 4 hours. To aspirate the solution into syringes for administration by colonoscopy, the DP may be placed in a clinical tray for up as short as possible and not more than 60 minutes.

**Additional comments regarding FDA comments in preIND.**

Many alterations and advancements in MTC01 manufacturing since our preIND filing in fall 2020 (Pre-IND PS005985, CRMTS # 12870) have been in response to feedback in the Written Responses to the preIND. There are two challenges proposed by the FDA for later stage development of MTC01 that we have pursued but not adequately solved yet. We provide general feedback on our experience with these unique challenges to LBP development below.

*Strain specific potency*

Although the FDA found our proposed method of MALDI-TOF identification of individual colonies combined with a total CFU potency metric suitable for this phase of development, it was stated that “as product development continues it will be necessary to quantify the number of each organism included in the final product to ensure manufacturing consistency.” In response to this suggestion, we have begun testing methods towards this goal.

Strain specific potency determination can be viewed as a two-step process, firstly discriminating between live and dead cells and secondly discriminating the strains. A DNA based live/dead discrimination can potentially be performed using propidium monoazide (PMA), a photoreactive DNA-binding dye, that can permeate membranes of damaged cells but not live cells. Since PMA modified DNA is not accessible for PCR based quantification methods. we developed strain specific qPCR primers using RUCS [38] to discriminate between the strains of MTC01.

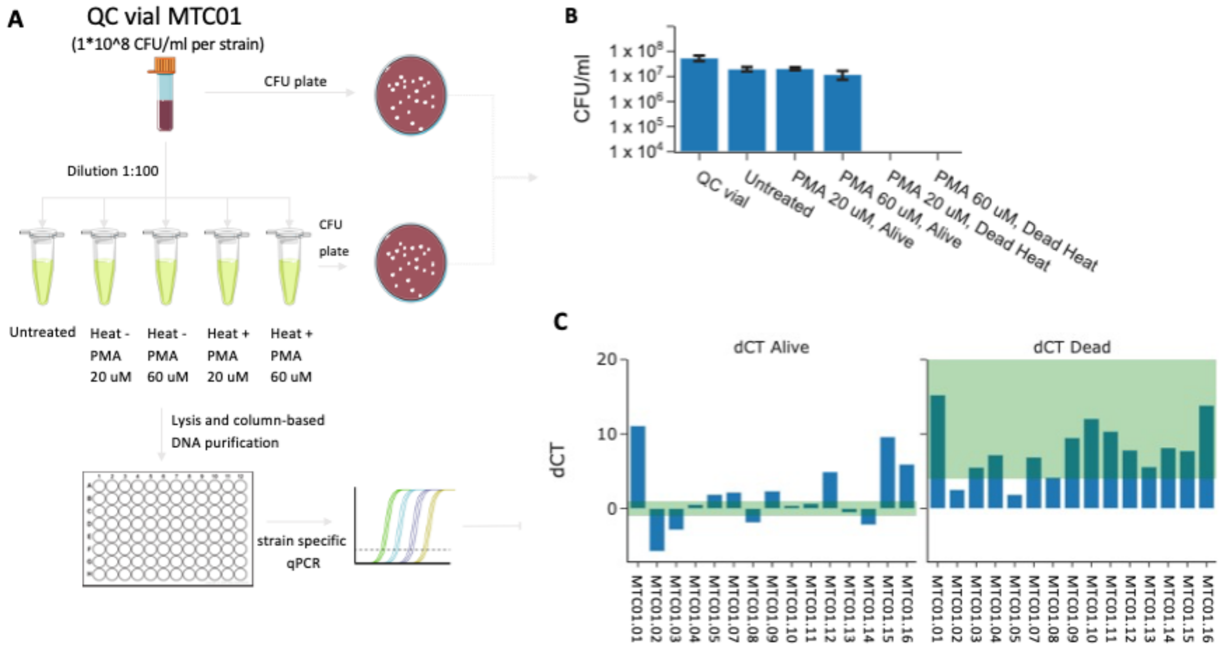

**Figure 5. Challenges with PMA-based quantification of live bacterial mixtures.** (A) A quality control vial of MTC01 was thawed and diluted 100-fold using PBS buffer to have sufficiently few bacterial cells to saturate them with PMA. The resultant cells were split into 5 aliquots. Two simulate (extreme) cell death, two aliquots were heated to 90C for 10 minutes. PMA treatment (Biotium) was performed at either 20 or 60 uM with a 10 minute incubation in the dark. PMA-Lite photolysis device (Biotium) was used to photoactivate the cells for 15 minutes. (B) CFU plating of each aliquot verified that PMA had a minimal impact on cell viability and that 90C lead to a total loss of viability. (C) delta CT values between the untreated sample and the 20 uM PMA treated sample suggest that for a few strains the PMA limits the amplification of live cells (e.g., strains 15 and 16). In addition, the method has some ability to differentiate live cells from dead cells, but it has a small dynamic range for many strains including MTC01.02, MTC01.05, and MTC01.08. Given the poor performance in this extreme example of 90C cell death over multiple orders of magnitude and high variability across strains, we are doubtful that further optimizations of this PMA-based approach will yield a usable assay in the much more subtle type of death MTC01 likely incurs while stored at -80C with losses expected to be <2 orders of magnitude across >1 year (based on our CFU stability data to date).

We attempted PMA-based approaches as these were previously used for FMT studies and highlighted by an FDA scientist as a potential approach [39–41]. However, in our hands with defined consortium, we found these methods unsuitable as a method for identifying strain-specific potency (Figure 5), and we are doubtful of their utility for FMT beyond very broad conclusions. As previously published for FMT [42], we found the utility of this method was highly dependent on getting the optimal ratio of PMA to microbial DNA. However, in testing this method across all multiple strains of MTC01 we found the dynamic range was not suitable and more importantly it was highly variable across strains with a few strains demonstrating partial reliability with this method and many strains not functioning at all (Figure 5). Given these poor results, we do not plan to continue advancing PMA-based methods for FMT or defined consortia.

A second approach to live/dead cell discrimination is growth based. Growth based identification methods are challenged by the variable growth conditions and times that individual strains require, but they have the major advantage that growth, by definition, indicates a living cell. We have had success in identifying strain-specific potency of low abundance and outcompeted strains by using antibiotic enrichment to selectively quantify the strain-specific potency of drug product on agar plates. We are able to utilize cheap and scalable *in silico* predictions of antibiotic resistance (CARD) [43] and confirm the predicted selective conditions quickly. We have developed tools to quantify 14 of the 15 MTC01 strains by this method. As would be expected, the CFU estimate of a strain on antibiotic enrichment media is often slightly lower than the CFU on non-antibiotic media. In addition to developing methods to enrich and quantify the 15$^{th}$ strain, we are using dilution series of each strain on antibiotic and non-antibiotic media to develop scaling factors to adjust for this viability loss on antibiotic media.

Thus far the most promising approach (also still in development) has been the use of limiting dilution to the point of single cells using Poisson statistics to allow each strain to grow individually without the competition that occurs on agar plates. We have previously used this method for high throughput strain isolation with great success [20]. In small pilots on our MTC01 drug product, it has been promising as the single-cell per well approach limits the competition that occurs when all 16 strains are on a single agar plate. We are currently scaling up the method to identify the potency of strains across a range of $10^1$-$10^4$. With the current mixing strategy for MTC01 this dynamic range should give us 1-2 logs more dynamic range than necessary to quantify the potency of each strain. An additional benefit of this approach is that by using redundant 16S rRNA amplicon sequencing of each well, we can generate high confidence consensus identifications of the strain(s) in each well [20]. The limiting dilution has the benefit having a clearer definition of viability than PMA based assays without the necessity to optimize the assay for every individual MTC01 strain.

We will continue to work on these assays in the coming year and will publish any advancements for the broader scientific community if we develop a reliable method.

*Extraneous anaerobic microbial counts*

In our preIND document, the following FDA comment was provided in response to our plans to evaluate the drug product, “We acknowledge you are determining if any of the product organisms grow under aerobic conditions and intend to perform USP <61>. However, because the organisms in your product are grown anaerobically during production, we encourage you to consider how to perform total extraneous anaerobic microbial counts as part of your bioburden testing. If you determine that any of your product strains are able to grow in the presence of oxygen, please describe your approach to determining microbial purity of those drug substances and of your final product, as an alternative to total aerobic microbial count (TAMC) and total combined yeasts and molds count (TYMC) as per USP<61>.”

As described above and in the USP<61> reports, none of the MTC01 strains grew under aerobic conditions, and none of our strains inhibited strains used as controls in the USP<61> assay. Therefore, we are confident in the ability of the USP<61> assay, performed by an external CRO, to provide an important quality control against extraneous aerobes. Aerobes are the most likely contaminants to result in adverse events. Therefore, this result significantly derisks the MTC01 characterization process.

In response to your comment that for future studies we develop methods to perform total extraneous anaerobic microbial counts as part of bioburden testing, we have thought extensively about potential solutions to what is an immensely difficult challenge. Our method of MALDI-TOF of several hundred colonies of DP provides a simple means to detect gross contamination of anerobic strains that are not inhibited by the 15 strains in MTC01. We have not detected any extraneous strains in our DP in this manner across all DP tests to date. In addition, the single cell limiting dilution culturing method we are developing as a potential strain specific potency assay provides a second approach with much more scalability than a MALDI-TOF approach. However, even this approach could realistically only be applied to $10^4$-$10^5$ cells. With a DP potency of $1x10^{11}$ per dose, that still leaves any anaerobes at $<10^6$ potency unaccounted for. Without clear criteria of which specific anaerobes to test for (e.g., by enrichment), we are therefore still left with a significant challenge. FACs sorting with antibodies towards 15 strains could allow you to perform negative selection to sort all non-labeled strains and check a smaller group for contamination with a bandwidth several orders of magnitude higher than our single cell approach. However, maintaining viability of strict anaerobes in FACs is a challenge. Furthermore, we have recently published work on the specificity of antibodies towards gut microbes [44]. Although we found antibodies can indeed be strain-specific, there are still substantial false-positives and false-negatives that would plague such a method [44,45]. The only realistic solution we have considered to date is to have strains in defined LBPs that are auxotrophs or have some way to effectively and specifically eliminate them from the DP. After all strains in MTC01 were killed, the remaining DP could be plated to determine if any extraneous anaerobes remained. Towards this we are now exploring approaches to develop genetically engineered versions of MTC01 strains with these features. However, this is a long and unpredictable scientific challenge, so we do not have an estimated timeline for substantial progress. We have been inspired by this question of extraneous anaerobes in an anaerobic LBP and have presented the idea widely at scientific conferences around the world to hopefully inspire others to also pursue this unique challenge.

#### *7.3 Placebo Product*

We will not have a placebo product in this phase 1b study.

#### *7.4 Labeling*

Below is an example of the label that will be placed on the cryogenic bottle containing the high dose and low dose MTC01 as well as the high dose and low dose FS for FMT.

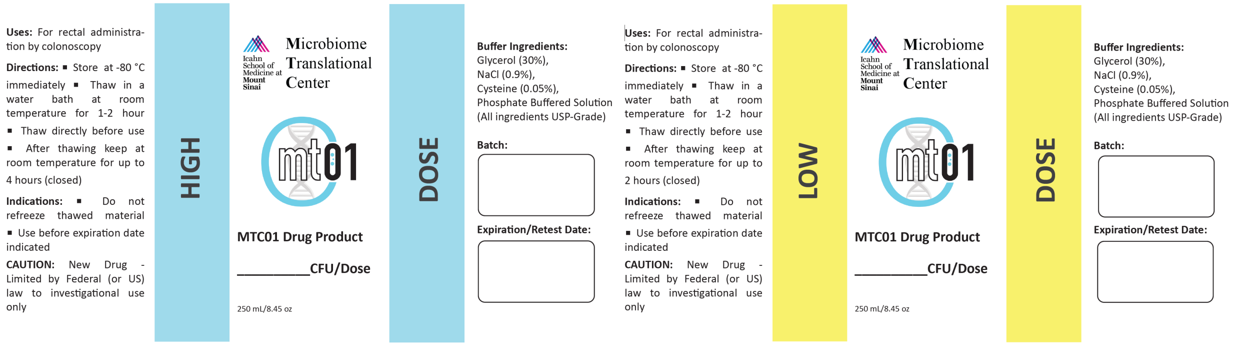

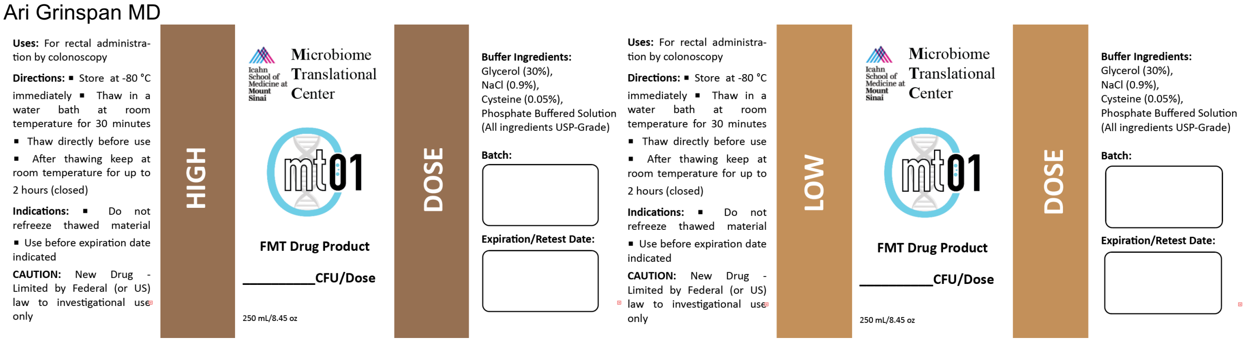

#### *7.5 Environmental Analysis Requirements*

Not applicable.

### 8 Pharmacology and Toxicology [21 CFR 312.23(a)(8)]

#### *8.1 Pharmacodynamics*

##### 8.1.1 Primary pharmacodynamics

Due to the unique nature of fecal material compared to most therapeutics, as well as the historical development of FMT within medical practice, there is a paucity of relevant pharmacokinetic or metabolic data available. Moreover, the applicability of pharmacokinetic or metabolic data is questionable given the nature of the active agent. Briefly, the active agents associated with FMT are thought to be viable gastrointestinal microbial strains that will colonize the patient's gut and proliferate to form new commensal populations. The inactive components of formulation buffer and fecal matter will pass through the patient via normal excretion pathways, either by being absorbed in the intestine or eliminated as stool. MTC01 will have active components of the 15 bacterial strains from donor D283 and inactive components of the formulation buffer.

#### *8.2 Safety pharmacology*

**MTC01**

To prevent the inclusion of any multi-drug resistant organisms (MDRO) in the LBP all community members have been tested in silico and in vitro for antibiotic susceptibly and prominent virulence factors. All microbes are susceptible to multiple antibiotics, providing several options for controlling the entire community in the case of an SAE. None of the strains in MTC01 are predicted to have antibiotic resistance genes on mobile genetic elements that are associated with MDROs (Appendix 8) and no prominent virulence or toxin encoded genes were found (Appendix 7). Antibiotic testing was performed by E-test, and agar dilution MIC according to CSLI protocols with a high correlation between the two methods (Figure 2C; Appendix 3 and 8). As expected, all strains are susceptible to multiple antibiotics. Importantly, the entire community tested thus far is also susceptible to multiple individual antibiotics.

**FMT**

The active agents associated with FMT are intended to colonize the patient's gut and proliferate to form new commensal microbiota. To that end, related or possibly related safety events of significance are gastrointestinal and include: bloating, distention, flatulence, diarrhea, constipation, abdominal pain, fever, vomiting, bacteremia, infection transmission.

#### 8.3 Pharmacokinetics

##### 8.3.1 Absorption

The inactive components of fecal matter will pass through the patient via normal excretion pathways, either by being absorbed in the intestine or eliminated as stool. Both MTC01 and FMT area formulated with USP grade PBS + cysteine + glycerol.

##### 8.3.2 Distribution

MTC01 and FMT will be administered directly into the right colon for distribution to the remainder of the colon.

##### 8.3.3 Metabolism

MTC01 and FMT involves administration of minimally manipulated microbial communities from an *in vitro* manufactured community (MTC01) or the stool of a healthy donor (FMT) into the gastrointestinal tract of an unhealthy patient. There is no metabolism of the FMT product. Instead, there is engraftment with the goal of normalization of the recipient's gut microbiota composition and functionality to treat rCDI.

##### 8.3.4 Excretion

The inactive components of MTC01 and fecal matter will be eliminated as stool.

#### 8.4 Pharmacology Summary

MTC01 and FMT are used to normalize the recipient's fecal microbial community structure. FMT product is not absorbed or metabolized. FMT product is engrafted and established into the recipient's microbiome.

#### 8.5 Pharmacology Conclusions

Due to the unique nature of MTC01 and FMT compared to most therapeutics, there is a paucity of relevant pharmacokinetic or metabolic data available. Moreover, the applicability of pharmacokinetic or metabolic metrics is questionable given the nature of the active agent. The active agents associated with MTC01 and FMT are thought to be viable gastrointestinal microbiota, and microbes that are not metabolized, in the same manner as a conventional drug. Instead, some fraction of the microbes will colonize the patient's gut and proliferate to form new commensal populations.

#### 8.6 Toxicology

Drug related SAEs for FMT have been MDROs and opportunistic pathogenic strains that were transferred from the FMT donor to the recipient. We use an extensive stool screening program to eliminate donors harboring such organisms. Far fewer individuals have been treated with defined LBPs than FMTs and fewer still individuals have received a specific LBP, little information is therefore available about adverse events related to defined LBPs. Given the absence of Proteobacteria in our cocktail which has been most associated with SAEs, we expect MTC01 to have a better safety profile than FMT.

#### 8.7 Genetic Toxicology

Not applicable

#### 8.8 Carcinogenicity

Not applicable

#### 8.9 Reproductive and Developmental Toxicology

Not applicable

#### 8.10 Special Toxicology Studies

Not applicable

##### 8.11 Toxicology Summary

Studies have found that FMT is largely safe and well tolerated. Similar rates of mild to moderate adverse events have been observed in participants who receive FMT compared to those who receive placebo. The most common reported adverse events with FMT are gastrointestinal and include nausea, constipation, diarrhea, low-grade fever, and vomiting. MTC01 has not been tested in humans although it is composed of a subset of bacterial strains from a human rCDI FMT donor.

##### 8.12 Toxicology Conclusions

Previous case studies involving the administration of FMT have on rare occasion shown a risk of transmission of infectious agents from donor to patients. This risk is thoroughly addressed through the donor screening process.

#### 9 Previous Human Experience with the Investigational Agent [21 CFR 312.23(a)(9)]

##### 9.1 Prior Clinical Research Experience

Numerous studies have demonstrated the outstanding efficacy of FMT for rCDI including the impact of storage time, processing method, and route of administration. This efficacy is highly correlated with the engraftment of donor strains. To a far more limited extent, defined LBPs have been used as an alternative to FMT in rCDI. One published phase 2 trial demonstrated that enema administered FMT was successful at treating rCDI while an enema administered 10-member consortium was not successful [14]. Notably in this study, subjects that failed FMT received a rescue FMT that counted towards success while those that received the LBP did not get a rescue LBP therapy. Smaller case studies have suggested LBPs can be successful in the treatment of rCDI including a 40 member community from Nubiyota with promising results [17].

MTC01 has not been tested previously in humans. However, the strains from MTC01 have been transferred to 25 rCDI recipients via FMT from donor D283.

##### 9.2 Clinical Care Experience

Not applicable.

#### 10 Additional Information [21 CFR 312.23(a)(10)]

Not applicable

#### 11 Bibliography

1. van Nood, E. *et al.* Duodenal infusion of donor feces for recurrent Clostridium difficile. *N Engl J Med* **368**, 407–415 (2013).
2. Ramai, D. *et al.* Fecal Microbiota Transplantation (FMT) with Colonoscopy Is Superior to Enema and Nasogastric Tube While Comparable to Capsule for the Treatment of Recurrent Clostridioides difficile Infection: A Systematic Review and Meta-Analysis. *Dig Dis Sci* (2020) doi:10.1007/s10620-020-06185-7.
3. Seekatz, A. M. *et al.* Recovery of the gut microbiome following fecal microbiota transplantation. *mBio* **5**, e00893-00814 (2014).
4. Hamilton, M. J., Weingarden, A. R., Unno, T., Khoruts, A. & Sadowsky, M. J. High-throughput DNA sequence analysis reveals stable engraftment of gut microbiota following transplantation of previously frozen fecal bacteria. *Gut Microbes* **4**, 125–135 (2013).
5. Youngster, I. *et al.* Oral, capsulized, frozen fecal microbiota transplantation for relapsing Clostridium difficile infection. *JAMA* **312**, 1772–1778 (2014).
6. Kao, D. *et al.* Effect of Oral Capsule- vs Colonoscopy-Delivered Fecal Microbiota Transplantation on Recurrent Clostridium difficile Infection: A Randomized Clinical Trial. *JAMA* **318**, 1985–1993 (2017).
7. Paramsothy, S. *et al.* Multidonor intensive faecal microbiota transplantation for active ulcerative colitis: a randomised placebo-controlled trial. *Lancet* **389**, 1218–1228 (2017).

8. Moayyedi, P. *et al.* Fecal Microbiota Transplantation Induces Remission in Patients With Active Ulcerative Colitis in a Randomized Controlled Trial. *Gastroenterology* **149**, 102-109.e6 (2015).
9. Costello, S. P. *et al.* Effect of Fecal Microbiota Transplantation on 8-Week Remission in Patients With Ulcerative Colitis: A Randomized Clinical Trial. *JAMA* **321**, 156–164 (2019).
10. DeFilipp, Z. *et al.* Drug-resistant e. coli bacteremia transmitted by fecal microbiota transplant. *New England Journal of Medicine* **381**, 2043–2050 (2019).
11. Fecal Microbiota for Transplantation: Safety Communication- Risk of Serious Adverse Reactions Due to Transmission of Multi-Drug Resistant Organisms | FDA. https://www.fda.gov/safety/medical-product-safety-information/fecal-microbiota-transplantation-safety-communication-risk-serious-adverse-reactions-due.
12. Fecal Microbiota for Transplantation: Safety Alert - Risk of Serious Adverse Events Likely Due to Transmission of Pathogenic Organisms | FDA. https://www.fda.gov/safety/medical-product-safety-information/fecal-microbiota-transplantation-safety-alert-risk-serious-adverse-events-likely-due-transmission.
13. Tvede, M. & Rask-Madsen, J. Bacteriotherapy for chronic relapsing Clostridium difficile diarrhoea in six patients. *Lancet* **1**, 1156–1160 (1989).
14. Rode, A. A. *et al.* Randomised clinical trial: a 12-strain bacterial mixture versus faecal microbiota transplantation versus vancomycin for recurrent Clostridioides difficile infections. *Aliment Pharmacol Ther* **53**, 999–1009 (2021).
15. Tvede, M., Tinggaard, M. & Helms, M. Rectal bacteriotherapy for recurrent Clostridium difficile-associated diarrhoea: results from a case series of 55 patients in Denmark 2000-2012. *Clin Microbiol Infect* **21**, 48–53 (2015).
16. Petrof, E. O. *et al.* Stool substitute transplant therapy for the eradication of Clostridium difficile infection: 'RePOOPulating' the gut. *Microbiome* **1**, 3 (2013).
17. Kao, D. *et al.* The effect of a microbial ecosystem therapeutic (MET-2) on recurrent Clostridioides difficile infection: a phase 1, open-label, single-group trial. *Lancet Gastroenterol Hepatol* **6**, 282–291 (2021).
18. Browne, H. P. *et al.* Culturing of 'unculturable' human microbiota reveals novel taxa and extensive sporulation. *Nature* **533**, 543–546 (2016).
19. Faith, J. J. *et al.* The long-term stability of the human gut microbiota. *Science* **341**, 1237439 (2013).
20. Goodman, A. L. *et al.* Extensive personal human gut microbiota culture collections characterized and manipulated in gnotobiotic mice. *Proc. Natl. Acad. Sci. U.S.A.* **108**, 6252–6257 (2011).
21. Aggarwala, V. *et al.* Precise quantification of bacterial strains after fecal microbiota transplantation delineates long-term engraftment and explains outcomes. *Nat Microbiol* **6**, 1309–1318 (2021).
22. Poyet, M. *et al.* A library of human gut bacterial isolates paired with longitudinal multiomics data enables mechanistic microbiome research. *Nat. Med.* **25**, 1442–1452 (2019).
23. Cold, F., Svensson, C. K., Petersen, A. M., Hansen, L. H. & Helms, M. Long-Term Safety Following Faecal Microbiota Transplantation as a Treatment for Recurrent Clostridioides difficile Infection Compared with Patients Treated with a Fixed Bacterial Mixture: Results from a Retrospective Cohort Study. *Cells* **11**, 435 (2022).
24. Dsouza, M. *et al.* Colonization of the live biotherapeutic product VE303 and modulation of the microbiota and metabolites in healthy volunteers. *Cell Host Microbe* **30**, 583-598.e8 (2022).
25. Hirten, R. P. *et al.* Microbial Engraftment and Efficacy of Fecal Microbiota Transplant for Clostridium Difficile in Patients With and Without Inflammatory Bowel Disease. *Inflamm. Bowel Dis.* **25**, 969–979 (2019).
26. Cho, S., Spencer, E., Hirten, R., Grinspan, A. & Dubinsky, M. C. Fecal Microbiota Transplant for Recurrent Clostridium difficile Infection in Pediatric Inflammatory Bowel Disease. *J Pediatr Gastroenterol Nutr* **68**, 343–347 (2019).
27. Kelly, C. R. *et al.* Fecal Microbiota Transplantation Is Highly Effective in Real-World Practice: Initial Results From the FMT National Registry. *Gastroenterology* (2020) doi:10.1053/j.gastro.2020.09.038.
28. Kelly, C. R., Fischer, M., Grinspan, A. & Allegretti, J. R. Patients Eligible for Trials of Microbe-Based Therapeutics Do Not Represent the Population With Recurrent Clostridioides difficile Infection. *Clin Gastroenterol Hepatol* **18**, 1099–1101 (2020).
29. Luo, Y., Tixier, E. N. & Grinspan, A. M. Fecal Microbiota Transplantation for Clostridioides difficile in High-Risk Older Adults Is Associated with Early Recurrence. *Dig Dis Sci* **65**, 3647–3651 (2020).
30. Contijoch, E. J. *et al.* Gut microbiota density influences host physiology and is shaped by host and microbial factors. *Elife* **8**, (2019).
31. Lawley, T. D. *et al.* Targeted restoration of the intestinal microbiota with a simple, defined bacteriotherapy resolves relapsing Clostridium difficile disease in mice. *PLoS Pathog* **8**, e1002995 (2012).

32. Yang, C. *et al.* Fecal IgA Levels Are Determined by Strain-Level Differences in Bacteroides ovatus and Are Modifiable by Gut Microbiota Manipulation. *Cell Host Microbe* **27**, 467-475.e6 (2020).
33. Britton, G. J. *et al.* Defined microbiota transplant restores Th17/RORγt+ regulatory T cell balance in mice colonized with inflammatory bowel disease microbiotas. *Proc. Natl. Acad. Sci. U.S.A.* **117**, 21536–21545 (2020).
34. Koelink, P. J. *et al.* Development of Reliable, Valid and Responsive Scoring Systems for Endoscopy and Histology in Animal Models for Inflammatory Bowel Disease. *J Crohns Colitis* **12**, 794–803 (2018).
35. Britton, G. J. *et al.* Microbiotas from Humans with Inflammatory Bowel Disease Alter the Balance of Gut Th17 and RORγt+ Regulatory T Cells and Exacerbate Colitis in Mice. *Immunity* **50**, 212-224.e4 (2019).
36. Faith, J. J., Ahern, P. P., Ridaura, V. K., Cheng, J. & Gordon, J. I. Identifying Gut Microbe-Host Phenotype Relationships Using Combinatorial Communities in Gnotobiotic Mice. *Science Translational Medicine* **6**, 220ra11-220ra11 (2014).
37. Faith, J. J. *et al. Strain population structure varies widely across bacterial species and predicts strain colonization in unrelated individuals*. http://biorxiv.org/lookup/doi/10.1101/2020.10.17.343640 (2020) doi:10.1101/2020.10.17.343640.
38. Thomsen, M. C. F., Hasman, H., Westh, H., Kaya, H. & Lund, O. RUCS: rapid identification of PCR primers for unique core sequences. *Bioinformatics* **33**, 3917–3921 (2017).
39. Carlson, P. E. Regulatory Considerations for Fecal Microbiota Transplantation Products. *Cell Host Microbe* **27**, 173–175 (2020).
40. Chu, N. D., Smith, M. B., Perrotta, A. R., Kassam, Z. & Alm, E. J. Profiling Living Bacteria Informs Preparation of Fecal Microbiota Transplantations. *PLoS One* **12**, e0170922 (2017).
41. Papanicolas, L. E. *et al.* Bacterial viability in faecal transplants: Which bacteria survive? *EBioMedicine* **41**, 509–516 (2019).
42. Papanicolas, L. E. *et al.* Optimisation of a propidium monoazide based method to determine the viability of microbes in faecal slurries for transplantation. *J Microbiol Methods* **156**, 40–45 (2019).
43. Alcock, B. P. *et al.* CARD 2020: antibiotic resistome surveillance with the comprehensive antibiotic resistance database. *Nucleic Acids Research* gkz935 (2019) doi:10.1093/nar/gkz935.
44. Yang, C. *et al.* Immunoglobulin A antibody composition is sculpted to bind the self gut microbiome. *Sci Immunol* **7**, eabg3208 (2022).
45. Rollenske, T. *et al.* Parallelism of intestinal secretory IgA shapes functional microbial fitness. *Nature* **598**, 657–661 (2021).
46. Bankevich, A. *et al.* SPAdes: A new genome assembly algorithm and its applications to single-cell sequencing. *Journal of Computational Biology* **19**, 455–477 (2012).
47. Darling, A. C. E., Mau, B., Blattner, F. R. & Perna, N. T. Mauve: Multiple alignment of conserved genomic sequence with rearrangements. *Genome Research* **14**, 1394–1403 (2004).
48. Jain, C., Rodriguez-R, L. M., Phillippy, A. M., Konstantinidis, K. T. & Aluru, S. High throughput ANI analysis of 90K prokaryotic genomes reveals clear species boundaries. *Nature Communications* **9**, 1–8 (2018).
49. Seemann, T. Prokka: rapid prokaryotic genome annotation. *Bioinformatics* **30**, 2068–2069 (2014).
50. Altschul, S. F., Gish, W., Miller, W., Myers, E. W. & Lipman, D. J. Basic local alignment search tool. *Journal of Molecular Biology* **215**, 403–410 (1990).
51. Sayers, S. *et al.* Victors: a web-based knowledge base of virulence factors in human and animal pathogens. *Nucleic Acids Research* **47**, D693–D700 (2019).
52. Chen, L. VFDB: a reference database for bacterial virulence factors. *Nucleic Acids Research* **33**, D325–D328 (2004).
53. Wattam, A. R. *et al.* PATRIC, the bacterial bioinformatics database and analysis resource. *Nucleic acids research* **42**, D581-91 (2014).
54. Jia, B. *et al.* CARD 2017: expansion and model-centric curation of the comprehensive antibiotic resistance database. *Nucleic Acids Research* **45**, D566–D573 (2017).
55. Waack, S. *et al.* Score-based prediction of genomic islands in prokaryotic genomes using hidden Markov models. *BMC Bioinformatics* **7**, 142 (2006).
56. Hsiao, W., Wan, I., Jones, S. J. & Brinkman, F. S. L. IslandPath: aiding detection of genomic islands in prokaryotes. *Bioinformatics* **19**, 418–420 (2003).
57. Sokol, H. *et al.* Faecalibacterium prausnitzii is an anti-inflammatory commensal bacterium identified by gut microbiota analysis of Crohn disease patients. *Proceedings of the National Academy of Sciences of the United States of America* **105**, 16731–16736 (2008).

#### 12 Attachments

The following lists the attachments to this IND application:

- Cover letter
- Investigator's Brochure [21 CFR 312.23(a)(5)]
- FDA Form 1572 [21 CFR 312.23(a)(6)]
- FDA Form 3674 [21 CFR 312.23(a)(6)]
- Protocol(s*)* [21 CFR 312.23(a)(6)]
- Donor Informed Consent Form [21 CFR 312.23(a)(6)]
- Recipient Informed Consent Form [21 CFR 312.23(a)(6)]
- CVs of all investigators [21 CFR 312.23(a)(6)]
- Data safety and monitoring board
- Appendices:
    1. MTC01 Donor Health Record
    2. MTC01 Strain identification
    3. MTC01 in vitro antibiotic susceptibility
    4. MTC01 USP<61> Reports
    5. MTC01 and FMT manufacturing facilities report
    6. MTC01 potency, identity, and purity results
    7. MTC01 virulence factors and toxins
    8. MTC01 predicted antibiotic resistance genes and mobile genetic elements
    9. MTC01 and FMT growth media and formulation buffers
    10. Calculation of MTC01 DS, MTC01 DP, and FS potency by colony forming units
    11. Generation of fecal slurry from the stool samples of human donors that pass safety screening
    12. MTC01 stability results
    13. Sterilization of non-disposable items in the generation of FS and MTC01 DP
    14. Donor Health Screening Questionnaire and Physical Exam
    15. CLIA Certificates
    16. NIH Adverse Event and Serious Adverse Event Guidelines

### INVESTIGATOR'S BROCHURE

[21 CFR 312.23(a)(5)]

Icahn School of Medicine at Mount Sinai

*Conduits*
The Institutes for Translational Sciences

### INVESTIGATOR'S BROCHURE (IB)

**PRODUCT NAME:** MTC01 (defined LBP) and FMT

**SPONSOR:**
Ari Grinspan MD,

Department of Medicine/Division of Gastroenterology
Icahn School of Medicine at Mount Sinai Hospital, New York, NY
One Gustave L. Levy Place Box 1069
New York NY 10029
212-241-7757


**For Principal Investigators:**

| |
|---|
| I hereby certify that I have read this document. |
| **Name:** |
| **Date and signature:** |

### CONFIDENTIALITY STATEMENT

**Important:** This document contains proprietary and confidential information belonging to Dr. Ari Grinspan. Its contents may not be disclosed or used without the express permission of Dr. Grinspan. This confidential document is for the sole information and use of the investigator's team and regulatory representatives.

### LIST OF CONTRIBUTORS

The Investigator's Brochure was compiled by:

| | |
|---|---|
| **Role: Sponsor** | **Ari Grinspan MD**, Associate Professor of Medicine, Gastroenterology<br>Address: One Gustave Levy Place Box 1069<br>Tel : 212-241-7757<br> |
| **Role: co-director Microbiome Translational Center** | **Jeremiah Faith PhD**, Associate Professor of Medicine, Clinical Immunology<br>Address: One Gustave Levy Pl Box 1498<br>Tel : 212-824-8953<br> |

### LIST OF ABBREVIATIONS AND DEFINITIONS OF TERMS

| Abbreviation or special term | Explanation |
|---|---|
| AE | Adverse Event |
| FMT | Fecal Microbiota Transplantation |
| FS | Fecal Slurry |
| LBP | Live Biotherapeutic Product |
| MCB | Master Cell Bank |
| RCB | Research Cell Bank |
| SAE | Serious Adverse Event |
| rCDI | recurrent *Clostridiodes. difficile* infection |

### 1 SUMMARY

MTC01 is a defined live biotherapeutic product (LBP) composed of 15 bacterial strains for the treatment of recurrent *Clostridioides difficile* infection (rCDI). The strains were isolated from stools of a frequently successful fecal microbiota transplantation donor (D283). In this phase 1b trial, we will compare the safety, engraftment, and efficacy of MTC01 with stool from the same FMT donor providing the opportunity to study the transmissibility of the same strains when *in vitro* manufactured compared to those generated *in vivo* in a human host.

Clinical experience:
In human studies, stool samples from donor D283 were used for 25 fecal transplants in rCDI. For 7 of these transplants, stool samples were collected to enable strain tracking of reference genomes from MTC01 isolates in the FMT recipient metagenome. The MTC01 strains were selected based on their engraftment in more than half of the recipients and their representing species of bacteria that are common in the human gut microbiota (>25% of individuals) [cite].

Nonclinical experience:
The genomes of MTC01 strains were analyzed in silico for toxin genes and mobile antibiotic resistance genes. No known toxin genes were found in the MTC01 genomes. No antibiotic resistance genes were detected on transferable regions of their genomes. *In vitro* studies identified multiple antibiotics that all MTC01 strains are susceptible to sulbactam, amoxicillin-clavulante, meropenem, and imipenem/relebactam.

Trial summary:
Subjects in this study will receive either FMT or MTC01 (30 per group). The FMT will consist of fecal slurry for donor D283, assuming the stool passes the latest safety screens. Otherwise, we will use fecal slurry from another healthy donor. Within both the FMT and MTC01 groups, we will test a high dose ($1x10^{11}$) and a low dose ($1x10^{10}$). The complete groupings and number of subjects per group will therefore be high dose D283 FMT (N=15), low dose D283 FMT (N=15), high dose MTC01 (N=15), and low dose MTC01 (N=15).

### 2 INTRODUCTION

The remarkable efficacy of Fecal Microbiota Transplantation (FMT) for the treatment of *Clostridioides difficile* infection (CDI) with cure rates often >90% represents the greatest clinical demonstration to date of the potential for gut microbiota manipulation to impact health. Although this success was evident even in an early randomized clinical trial comparing FMT with antibiotic standard of care [1], the subsequent years have been highly productive in determining key aspects of such therapies including that colonic administration appears to outperform enema [2] and that the gut microbiota 24-72hr post-transplant typically is restored to contain members of the bacterial groups in healthy individuals [3,4]. FMT studies have identified bacterial species that may be key to the mechanism of FMT success [5]. And FMT studies have shown that single-dose oral administration in capsules may be less effective than colonoscopic administration, while multiple oral doses can achieve non-inferiority with FMT [6,7]. The major contributors to these FMT innovations have been small investigator initiated academic clinical trials fostered by the relative ease of producing the “drug”, which unlike the complex manufacturing requirements for generating patient-grade biologics or small molecule drugs requires just blending the stool sample, coarse filtering the fecal slurry for administration, and screening assays for safety. The sharing of know-how by the scientists leading such studies, ranging from how to produce and bank the fecal slurry [8,9] to how to file an Investigational New Drug (IND) application to the FDA [10], has also been key to lowering the barriers to entry for innovative trials which now span numerous indications.

With more widespread use of FMT for rCDI, some of the challenges of this therapeutic modality are emerging. First it is difficult to scale as it requires a constant supply of human stool donations, safety screening of every donor microbiota, and labor-intensive processing. These challenges are exacerbated in indications beyond rCDI, such as ulcerative colitis, where it appears multiple FMT doses may be more effective than single doses [11–13]. A second challenge has been the emergence of serious adverse events (SAEs) associated with the transmission of opportunistic pathogens from the donor to the recipient, including extended spectrum beta-lactamases (ESBL)-producing *E. coli* [14,15], enteropathogenic *E. coli* (EPEC), and Shigatoxin-producing *E. coli* (STEC) [16], as well as the potential risk of transmitting SARS-CoV-2 and monkey pox. Although rare, these SAEs can be life threatening to the affected individuals, and each potentially new opportunistic pathogen identified increases the cost to screen FMT donors and decreases the number of donors that pass these screens, thereby reducing the amount of available "drug" in the human population. Finally, the identification of safety concerns is a significant challenge with an undefined FMT product, as few FMT recipients receive the same gut microbiota from the same donor, and therefore only the most penetrant and severe adverse events can be associated with a given donor microbiota.

These challenges with FMT have spurred interest in developing defined consortia of fecal bacteria that could be used as a synthetic alternative to FMT termed a defined live biotherapeutic product (LBP). Defined LBPs would be more scalable, could avoid including multi-drug resistant organisms (MDROs) or strains associated with SAEs, and provide far greater ability to control community composition and gain mechanistic insights. To date, there are a limited number of published studies of multi-strain LBPs used for the treatment of CDI. Tvede and Rask-Madsen treated five individuals with CDI with a rectal infusion of 10 cultured bacteria ($2x10^9$ colony forming units (CFU) per strain; $2x10^{10}$ total dose) with all five patients responding and no SAE [17]. More recently Petrof et.al., treated two subjects with rCDI with a consortium of 33 bacterial isolates from a healthy donor stool [18]. The strains were administered by colonoscopy to the right and mid colon. Both patients responded with no SAE and remained symptom free for 6 months. The microbes were administered in 100 ml of pre-reduced saline at a concentration of $3.5x10^9$ CFU/ml giving an overall total dose of $3.5x10^{11}$ CFU. And most recently the company NuBiyota published an open label study with a 40-member oral lyophilized consortia given to 19 individuals with rCDI that attained cure rates similar to oral FMT (79% on first treatment, 95% on retreatment of relapsed individuals) [19]. Although these three studies were important demonstrations of feasibility and safety, we need more trials to understand minimal consortia necessary for a safe therapeutic for rCDI, and we are far from a similar consortium for addition promising indications for FMT like ulcerative colitis. There is a large and unexplored niche of small, innovative LBP human clinical trials for understanding the mechanisms of gut microbiota manipulation as a therapeutic modality. If these studies are performed in academia with sharing of know-how similar to FMT, it would accelerate the translation of the large investment in human microbiome research into clinical care for both academia and industry. This study represents one such example of an academically driven trial with the goal of translating microbiome discoveries from successful human FMT in rCDI to a scalable defined consortium derived of strains from the same donor. By comparing safety, engraftment, and efficacy between the FMT and the defined consortia we will gain important insights to bring microbial therapeutics more broadly to the clinic in rCDI and beyond.

***Tracking the stable engraftment and persistence of strains in rCDI FMT recipients*** — For more than a decade, we have refined high throughput pipelines for the isolation, propagation, storage, and recovery of gut microbes from human microbiome samples [20–24]. We recently published a novel algorithm, termed *Strainer*, that uses reference genomes of bacteria isolated from FMT donors to accurately and sensitively track each microbe in as few as 2.5 million metagenomic reads from each FMT recipient [25]. Specifically, the algorithm identifies and tracks the strain-unique subset of each strain's genome in fecal metagenome datasets. We validated this algorithm using a dataset of >350 bacterial genomes isolated from 19 individuals whose metagenome was also sequenced. The *Strainer* algorithm detected >75% of the donor strains in the metagenomes with a precision of >0.95 (i.e., less than 5% false positives). The algorithm performed similarly well on test datasets generated from bacterial isolate genomes and metagenomes from individuals with IBD and rCDI (**Fig. 1A**) [25].

To apply this new algorithm to understand strain engraftment and persistence in rCDI FMT recipients, we isolated and sequenced 1,008 unique bacterial strains (207 species) from 8 FMT healthy donors and 14 rCDI FMT recipients [25]. As in prior studies [26,27], these cultured strains represented the majority of the metagenome with 70% (sd = 16%) of bacterial metagenomic reads mapping to the cultured strain genomes. The proportion of donor strains engrafting in the recipient at 8 weeks was 71% across all rCDI individuals with no early relapse within 2 months post-FMT (**Fig. 1B**). To determine the stability of this engraftment, we tracked recipients for up to five years after FMT and found consistently high engraftment of donor strains at all time points (**Fig. 1C**). In these individuals, we report an average engraftment of 83% (sd = 9%) at 36 hours, which stabilizes at 71% (sd = 16%) at 8 weeks and remains consistently high at 71% (sd = 9%) even 5 years later. In contrast, the majority of the recipient microbiota is eliminated in the first 8 weeks post-FMT with the small subset that persists for 8 weeks stably remaining for the duration of the 5-year sampling (**Fig. 1D**). Overall these results demonstrate that gut microbiota manipulation by FMT can lead to a near permanent engraftment of a new stable set of bacterial strains in subjects with rCDI.

***in human discovery based design of an LBP for rCDI*** — Although most drug discovery pipelines involve some combination of *in vitro*, *in silico*, and *in vivo* experimentation with model systems, the translation from model to human is an established challenge. An advantage of the empirical use of FMT to treat human disease is the potential to use *in human* discovery to identify putative therapeutic microbiotas and strains [5,12]. We used the human FMT transplants for rCDI described above to design the synthetic mixture of strains for an LBP to treat rCDI. To perform the strain tracking detailed above, we isolated hundreds of strains from all Mount Sinai FMT donors to provide the initial raw materials for the cocktail. In our FMT program at Mount Sinai Hospital, we used one particular human FMT donor (D283) seven times with relapse-free remission in 5 of 7 individuals with one transplant and 7 of 7 achieving relapse-free remission with two transplants. We focused our LBP design on this donor D283 for which we had isolated 49 strains from 29 species. Manufacturing this complete set of strains would be a large undertaking and given the substantial variation in gut microbiota composition between successful FMT donors, it is likely unnecessary. We first eliminated the numerous donor strains that did not engraft in the majority of the five D283 recipients with no relapse. To further limit the number of

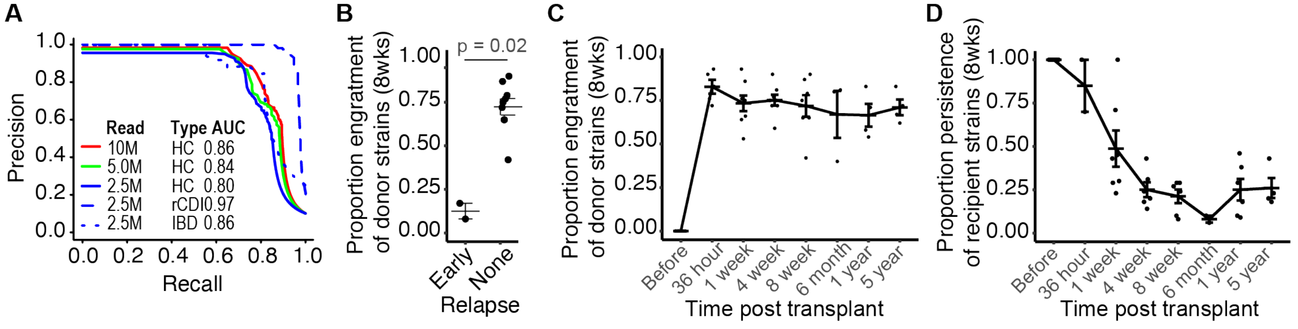

**Figure 1.** Accurate strain tracking of FMT donor strain engraftment and recipient strain persistence. (A) Our algorithm can accurately match >350 strains from different species to the correct donor sample from which they were isolated and performs well on samples from individuals with rCDI, IBD, and healthy controls. (B) Proportional engraftment of donor strains is lower at 8 wks in individuals that relapse early (prior to 8 wks). (C) Strains from the donor remain stably engrafted in non-relapsing post-FMT subjects for at least 5 years after transplant, while (D) strains isolated from a recipient prior to FMT are rapidly lost with a small proportion persisting at longer timescales.

strains, we preferred to maximize species diversity over strain diversity; therefore, we selected only the most abundant strain of each species. To focus on species that will be effective in the broadest population of recipients, we eliminated species that are found in <25% of healthy individuals. After applying these selection criteria on the culturable fraction of D283, we were left with 15 unique strains from 15 unique species for the defined LBP MTC01 (**Table 1**).

### 3 PHYSICAL, CHEMICAL AND PHARMACEUTICAL PROPERTIES AND FORMULATION

Both MTC01 and D283 FMT products are liquid formulations of live microbes in a volume of 250 ml, thawed at room temperature and used within 4 hours of thawing. Administration of MTC01 and D283 FMT is by colonoscopy with product delivery to the right colon.

D283 FMT fecal slurry consists of either a high dose ($1x10^{11}$ CFU) or low dose ($1x10^{10}$ CFU) of bacteria and other fecal products and metabolites coarse filtered and homogenized in a formulation of USP grade PBS, 0.5% cysteine (oxygen scavenger), and 30% glycerol (cryoprotectant).

MTC01 was grown in vitro in animal free rich media, washed 2x in USP grade PBS with 0.5% cysteine (oxygen scavenger). The drug substance and drug product were formulated in same media (USP grade PBS, 0.5% cysteine, 30% glycerol) as the fecal slurry.

Both MTC01 and D283 FMT are stored at -80C until release for the study. FMT has been previously demonstrated to be stable at -80C for >12 months [28]. We found little change in stability of MTC01 strains stored in -80C for up to 48 weeks.

**Table 1. The 15 strains in MTC01.**

| Strain ID | Genus | Species | Phylum |
|---|---|---|---|
| MTC01.01 | Bacteroides | uniformis | Bacteroidetes |
| MTC01.02 | Bacteroides | ovatus | Bacteroidetes |
| MTC01.03 | Bifidobacterium | longum | Actinobacterium |
| MTC01.04 | Bacteroides | thetaiotaomicron | Bacteroidetes |
| MTC01.05 | Bacteroides | vulgatus | Bacteroidetes |
| MTC01.07 | Parabacteroides | distasonis | Bacteroidetes |
| MTC01.08 | Bifidobacterium | adolescentis | Actinobacterium |
| MTC01.09 | Parabacteroides | merdae | Bacteroidetes |
| MTC01.11 | Eubacterium | rectale | Firmicutes |
| MTC01.12 | Bacteroides | caccae | Bacteroidetes |
| MTC01.13 | Dorea | longicatena | Firmicutes |
| MTC01.14 | Odoribacter | splanchnicus | Bacteroidetes |
| MTC01.15 | Bacteroides | cellulosilyticus | Bacteroidetes |
| MTC01.16 | Bifidobacterium | pseudocatenulatum | Actinobacterium |

### 4 NON/PRE CLINICAL STUDIES

#### 4.1 Non Clinical Pharmacology

The genomes of MTC01 strains have been analyzed using comparative genomes tools to identify that MTC01 strain do not contain known toxins. In addition, all strains are susceptible multiple antibiotics in MIC testing using CLSI methods including ampicillin/sulbactam, amoxicillin/clavulanic acid, and meropenem.

#### 4.2 Pharmacokinetics and Product Metabolism in Animals

We colonized germ-free mice with MTC01 and found it induced no tissue damage and was able to induce tolerogenic regulatory T cells.

#### 4.3 Toxicology

No known toxins were found in the genomes of MTC01 strains.

### 5 EFFECTS IN HUMANS

MTC01 and FMT are predicted to operate by a one or more mechanisms in rCDI. Likely most importantly, the native gut microbiota of subjects with rCDI is decimated and therefore does not have colonization resistance to pathogens. Restoring a normal gut ecosystem likely boosts colonization resistance by competing for similar niches to C. difficile. In addition, the gut microbiota may influence toxin generation by C. difficile, improve barrier function, promote immune tolerance, and alter bile salt conditions to provide a less hospitable environment for *C. difficile*.

#### 5.1 Pharmacokinetics and Product Metabolism in Humans

MTC01 and FMT are both live biotherapeutic products. For FMT we have previously demonstrated that durable engraftment (for at least 5 years) occurs from a single FMT dose in rCDI. Determining the engraftment durability of a defined consortia compared to FMT in rCDI is one of the goals of this study.

#### 5.2 Safety and Efficacy

MTC01 has not been tested in humans. However, it is derived from FMT donor stools from a successful FMT donor (D283) that did not previously lead to an SAE across 25 recipients with a success rate of 84%.

#### 5.3 Marketing Experience

MTC01 has not been tested in humans and is not currently a marketed drug.

### 6 SUMMARY OF DATA AND GUIDANCE FOR THE INVESTIGATOR

MTC01 has not been tested in humans with rCDI. FMT has however and provides some safety risk guidance. SAE resulting from FMT across all indications have included the transfer of multiple infectious agents [15,16], pathobionts [29], and even death (in an immunocompromised subject)[14]. To date, all drug related SAE from FMT has resulted from the transfer of gram negative Proteobacteria from the donor to the recipient. For our FMT product in this study, we follow all the latest screening guidance including testing for ESBL. For MTC01, Proteobacteria have not been included in the product. Furthermore, all MTC01 strains are strict anaerobes and do not inhibit the growth of USP<61> positive control organisms. All MTC01 MCB, DS, and DP have passed USP<61> testing by an external CRO.

### 7 REFERENCES

1. van Nood, E. *et al.* Duodenal infusion of donor feces for recurrent Clostridium difficile. *N Engl J Med* **368**, 407–415 (2013).
2. Ramai, D. *et al.* Fecal Microbiota Transplantation (FMT) with Colonoscopy Is Superior to Enema and Nasogastric Tube While Comparable to Capsule for the Treatment of Recurrent Clostridioides difficile Infection: A Systematic Review and Meta-Analysis. *Dig Dis Sci* (2020) doi:10.1007/s10620-020-06185-7.
3. Seekatz, A. M. *et al.* Recovery of the gut microbiome following fecal microbiota transplantation. *mBio* **5**, e00893-00814 (2014).
4. Hamilton, M. J., Weingarden, A. R., Unno, T., Khoruts, A. & Sadowsky, M. J. High-throughput DNA sequence analysis reveals stable engraftment of gut microbiota following transplantation of previously frozen fecal bacteria. *Gut Microbes* **4**, 125–135 (2013).
5. Buffie, C. G. *et al.* Precision microbiome reconstitution restores bile acid mediated resistance to Clostridium difficile. *Nature* **517**, 205–208 (2015).
6. Youngster, I. *et al.* Oral, capsulized, frozen fecal microbiota transplantation for relapsing Clostridium difficile infection. *JAMA* **312**, 1772–1778 (2014).
7. Kao, D. *et al.* Effect of Oral Capsule- vs Colonoscopy-Delivered Fecal Microbiota Transplantation on Recurrent Clostridium difficile Infection: A Randomized Clinical Trial. *JAMA* **318**, 1985–1993 (2017).
8. Hamilton, M. J., Weingarden, A. R., Sadowsky, M. J. & Khoruts, A. Standardized frozen preparation for transplantation of fecal microbiota for recurrent Clostridium difficile infection. *Am J Gastroenterol* **107**, 761–767 (2012).
9. Satokari, R., Mattila, E., Kainulainen, V. & Arkkila, P. E. T. Simple faecal preparation and efficacy of frozen inoculum in faecal microbiota transplantation for recurrent Clostridium difficile infection--an observational cohort study. *Aliment Pharmacol Ther* **41**, 46–53 (2015).
10. Kelly, C. R., Kunde, S. S. & Khoruts, A. Guidance on preparing an investigational new drug application for fecal microbiota transplantation studies. *Clin Gastroenterol Hepatol* **12**, 283–288 (2014).
11. Paramsothy, S. *et al.* Multidonor intensive faecal microbiota transplantation for active ulcerative colitis: a randomised placebo-controlled trial. *Lancet* **389**, 1218–1228 (2017).
12. Moayyedi, P. *et al.* Fecal Microbiota Transplantation Induces Remission in Patients With Active Ulcerative Colitis in a Randomized Controlled Trial. *Gastroenterology* **149**, 102-109.e6 (2015).
13. Costello, S. P. *et al.* Effect of Fecal Microbiota Transplantation on 8-Week Remission in Patients With Ulcerative Colitis: A Randomized Clinical Trial. *JAMA* **321**, 156–164 (2019).
14. DeFilipp, Z. *et al.* Drug-resistant e. coli bacteremia transmitted by fecal microbiota transplant. *New England Journal of Medicine* **381**, 2043–2050 (2019).
15. Fecal Microbiota for Transplantation: Safety Communication- Risk of Serious Adverse Reactions Due to Transmission of Multi-Drug Resistant Organisms | FDA. https://www.fda.gov/safety/medical-product-safety-information/fecal-microbiota-transplantation-safety-communication-risk-serious-adverse-reactions-due.
16. Fecal Microbiota for Transplantation: Safety Alert - Risk of Serious Adverse Events Likely Due to Transmission of Pathogenic Organisms | FDA. https://www.fda.gov/safety/medical-product-safety-information/fecal-microbiota-transplantation-safety-alert-risk-serious-adverse-events-likely-due-transmission.
17. Tvede, M. & Rask-Madsen, J. Bacteriotherapy for chronic relapsing Clostridium difficile diarrhoea in six patients. *Lancet (London, England)* **1**, 1156–60 (1989).
18. Petrof, E. O. *et al.* Stool substitute transplant therapy for the eradication of Clostridium difficile infection: 'RePOOPulating' the gut. *Microbiome* **1**, 3 (2013).
19. Kao, D. *et al.* The effect of a microbial ecosystem therapeutic (MET-2) on recurrent Clostridioides difficile infection: a phase 1, open-label, single-group trial. *Lancet Gastroenterol Hepatol* **6**, 282–291 (2021).

20. Britton, G. J. *et al.* Defined microbiota transplant restores Th17/RORγt+ regulatory T cell balance in mice colonized with inflammatory bowel disease microbiotas. *Proc. Natl. Acad. Sci. U.S.A.* **117**, 21536–21545 (2020).
21. Yang, C. *et al.* Fecal IgA Levels Are Determined by Strain-Level Differences in Bacteroides ovatus and Are Modifiable by Gut Microbiota Manipulation. *Cell Host Microbe* **27**, 467-475.e6 (2020).
22. Goodman, A. L. *et al.* Extensive personal human gut microbiota culture collections characterized and manipulated in gnotobiotic mice. *Proc. Natl. Acad. Sci. U.S.A.* **108**, 6252–6257 (2011).
23. Faith, J. J., Ahern, P. P., Ridaura, V. K., Cheng, J. & Gordon, J. I. Identifying gut microbe-host phenotype relationships using combinatorial communities in gnotobiotic mice. *Sci Transl Med* **6**, 220ra11 (2014).
24. Britton, G. J. *et al.* Microbiotas from Humans with Inflammatory Bowel Disease Alter the Balance of Gut Th17 and RORγt+ Regulatory T Cells and Exacerbate Colitis in Mice. *Immunity* **50**, 212-224.e4 (2019).
25. Aggarwala, V. *et al. Quantification of discrete gut bacterial strains following fecal transplantation for recurrent* Clostridioides difficile *infection demonstrates long-term stable engraftment in non-relapsing recipients*. http://biorxiv.org/lookup/doi/10.1101/2020.09.10.292136 (2020) doi:10.1101/2020.09.10.292136.
26. Lau, J. T. *et al.* Capturing the diversity of the human gut microbiota through culture-enriched molecular profiling. *Genome medicine* **8**, 72 (2016).
27. Browne, H. P. *et al.* Culturing of 'unculturable' human microbiota reveals novel taxa and extensive sporulation. *Nature* **533**, 543–546 (2016).
28. Allegretti, J. R. *et al.* Stool processing speed and storage duration do not impact the clinical effectiveness of fecal microbiota transplantation. *Gut Microbes* **11**, 1806–1808 (2020).
29. Drewes, J. L. *et al.* Transmission and clearance of potential procarcinogenic bacteria during fecal microbiota transplantation for recurrent Clostridioides difficile. *JCI Insight* **4**, 130848 (2019).

### FDA Form 1572

[21 CFR 312.23(a)(6)]

| DEPARTMENT OF HEALTH AND HUMAN SERVICES<br>FOOD AND DRUG ADMINISTRATION<br>**STATEMENT OF INVESTIGATOR**<br>***(TITLE 21, CODE OF FEDERAL REGULATIONS (CFR) PART 312)***<br>(See instructions on reverse side.) | Form Approved: OMB No. 0910-0014<br>Expiration Date: March 31, 2025<br>*See OMB Statement on Reverse.*<br>**NOTE:** No investigator may participate in an investigation until he/she provides the sponsor with a completed, signed Statement of Investigator, Form FDA 1572 (21 CFR 312.53(c)). |
|---|---|

**1. NAME AND ADDRESS OF INVESTIGATOR**

Name of Clinical Investigator: Ari Grinspan

| Address 1 | Address 2 | | |
|---|---|---|---|
| One Gustave L. Levy Place Box 1069 | | | |
| **City** | **State/Province/Region** | **Country** | **ZIP or Postal Code** |
| New York | New York | USA | 10029 |

**2. EDUCATION, TRAINING, AND EXPERIENCE THAT QUALIFY THE INVESTIGATOR AS AN EXPERT IN THE CLINICAL INVESTIGATION OF THE DRUG FOR THE USE UNDER INVESTIGATION. ONE OF THE FOLLOWING IS PROVIDED** *(Select **one** of the following.)*

☒ Curriculum Vitae ☐ Other Statement of Qualifications

**3. NAME AND ADDRESS OF ANY MEDICAL SCHOOL, HOSPITAL, OR OTHER RESEARCH FACILITY WHERE THE CLINICAL INVESTIGATION(S) WILL BE CONDUCTED**

CONTINUATION PAGE for Item 3

Name of Medical School, Hospital, or Other Research Facility: Icahn School of Medicine at Mount Sinai Hospital

| Address 1 | Address 2 | | |
|---|---|---|---|
| 1468 Madison Ave | | | |
| **City** | **State/Province/Region** | **Country** | **ZIP or Postal Code** |
| New York | New York | USA | 10029 |

**4. NAME AND ADDRESS OF ANY CLINICAL LABORATORY FACILITIES TO BE USED IN THE STUDY**

CONTINUATION PAGE for Item 4

Name of Clinical Laboratory Facility: Microbiome Translational Center

| Address 1 | Address 2 | | |
|---|---|---|---|
| 1470 Madison Ave | 8-201 | | |
| **City** | **State/Province/Region** | **Country** | **ZIP or Postal Code** |
| New York | New York | USA | 10029 |

**5. NAME AND ADDRESS OF THE INSTITUTIONAL REVIEW BOARD (IRB) THAT IS RESPONSIBLE FOR REVIEW AND APPROVAL OF THE STUDY(IES)**

CONTINUATION PAGE for Item 5

Name of IRB: Program for the Protection of Human Subjects

| Address 1 | Address 2 | | |
|---|---|---|---|
| One Gustave L. Levy Place Box 1081 | | | |
| **City** | **State/Province/Region** | **Country** | **ZIP or Postal Code** |
| New York | New York | USA | 10029 |

**6. NAMES OF SUBINVESTIGATORS** *(If not applicable, enter "None")*

None

CONTINUATION PAGE – for Item 6

**7. NAME AND CODE NUMBER, IF ANY, OF THE PROTOCOL(S) IN THE IND FOR THE STUDY(IES) TO BE CONDUCTED BY THE INVESTIGATOR**

See attached study protocol (MTC01_Study_Protocol)

PSC Publishing Services (301) 443-6740 EF

8. PROVIDE THE FOLLOWING CLINICAL PROTOCOL INFORMATION. *(Select **one** of the following.)*

☒ For Phase 1 investigations, a general outline of the planned investigation including the estimated duration of the study and the maximum number of subjects that will be involved.

☐ For Phase 2 or 3 investigations, an outline of the study protocol including an approximation of the number of subjects to be treated with the drug and the number to be employed as controls, if any; the clinical uses to be investigated; characteristics of subjects by age, sex, and condition; the kind of clinical observations and laboratory tests to be conducted; the estimated duration of the study; and copies or a description of case report forms to be used.

9. COMMITMENTS

I agree to conduct the study(ies) in accordance with the relevant, current protocol(s) and will only make changes in a protocol after notifying the sponsor, except when necessary to protect the safety, rights, or welfare of subjects.

I agree to personally conduct or supervise the described investigation(s).

I agree to inform any patients, or any persons used as controls, that the drugs are being used for investigational purposes and I will ensure that the requirements relating to obtaining informed consent in 21 CFR Part 50 and institutional review board (IRB) review and approval in 21 CFR Part 56 are met.

I agree to report to the sponsor adverse experiences that occur in the course of the investigation(s) in accordance with 21 CFR 312.64. I have read and understand the information in the investigator's brochure, including the potential risks and side effects of the drug.

I agree to ensure that all associates, colleagues, and employees assisting in the conduct of the study(ies) are informed about their obligations in meeting the above commitments.

I agree to maintain adequate and accurate records in accordance with 21 CFR 312.62 and to make those records available for inspection in accordance with 21 CFR 312.68.

I will ensure that an IRB that complies with the requirements of 21 CFR Part 56 will be responsible for the initial and continuing review and approval of the clinical investigation. I also agree to promptly report to the IRB all changes in the research activity and all unanticipated problems involving risks to human subjects or others. Additionally, I will not make any changes in the research without IRB approval, except where necessary to eliminate apparent immediate hazards to human subjects.

I agree to comply with all other requirements regarding the obligations of clinical investigators and all other pertinent requirements in 21 CFR Part 312.

#### INSTRUCTIONS FOR COMPLETING FORM FDA 1572 STATEMENT OF INVESTIGATOR

1. Complete all sections. Provide a separate page if additional space is needed.
2. Provide curriculum vitae or other statement of qualifications as described in Section 2.
3. Provide protocol outline as described in Section 8.
4. Sign and date below.
5. FORWARD THE COMPLETED FORM AND OTHER DOCUMENTS BEING PROVIDED TO THE SPONSOR. The sponsor will incorporate this information along with other technical data into an Investigational New Drug Application (IND). INVESTIGATORS SHOULD NOT SEND THIS FORM DIRECTLY TO THE FOOD AND DRUG ADMINISTRATION.

| 10. DATE *(mm/dd/yyyy)* | 11. SIGNATURE OF INVESTIGATOR Sign |
|---|---|
| 11/02/2022 | Ari Grinspan Digitally signed by Ari Grinspan Date: 2022.11.02 14:56:00 -04'00' |

(**WARNING**: A willfully false statement is a criminal offense. U.S.C. Title 18, Sec. 1001.)

**The information below applies only to requirements of the Paperwork Reduction Act of 1995.**

The burden time for this collection of information is estimated to average 100 hours per response, including the time to review instructions, search existing data sources, gather and maintain the data needed and complete and review the collection of information. Send comments regarding this burden estimate or any other aspect of this information collection, including suggestions for reducing this burden to the address to the right:

Department of Health and Human Services
Food and Drug Administration
Office of Operations
Paperwork Reduction Act (PRA) Staff
**

*"An agency may not conduct or sponsor, and a person is not required to respond to, a collection of information unless it displays a currently valid OMB number."*

**DO NOT SEND YOUR COMPLETED FORM TO THIS PRA STAFF EMAIL ADDRESS.**

### FDA Form 3674

[21 CFR 312.23(a)(6)]

**DEPARTMENT OF HEALTH AND HUMAN SERVICES**
Food and Drug Administration

### Certification of Compliance

#### Under 42 U.S.C. § 282(j)(5)(B), with Requirements of ClinicalTrials.gov Data Bank

(For submission with an application/submission, including amendments, supplements, and resubmissions, under §§ 505, 515, 520(m), or 510(k) of the Federal Food, Drug, and Cosmetic Act or § 351 of the Public Health Service Act.)

##### SPONSOR / APPLICANT / SUBMITTER INFORMATION

1. Name of Sponsor/Applicant/Submitter: Ari Grinspan

2. Date of the Application/Submission: 11/01/2022

3. Address

Address 1 *(Street address, P.O. box, company name c/o)*: One Gustave L. Levy Place Box 1069

Address 2 *(Apartment, suite, unit, building, floor, etc.)*:

| City | State/Province/Region |
|---|---|
| New York | New York |

| Country | ZIP or Postal Code |
|---|---|
| USA | 10029 |

4. Telephone and Fax Numbers *(Include country code if applicable and area code)*

(Tel): 212-241-7757

(Fax): 646-537-8921

##### PRODUCT INFORMATION

5. **For Drugs/Biologics:** Include Any/All Available Established, Proprietary and/or Chemical/Biochemical/Blood/Cellular/Gene Therapy Product Name(s).
**For Devices:** Include Any/All Common or Usual Name(s), Classification, Trade or Proprietary or Model Name(s) and/or Model Number(s)

MTC01, defined live biotherapeutic product consisting of 15 bacterial strains, defined microbial therapeutic

Fecal microbiota transplant

Continuation Page for #5

##### APPLICATION / SUBMISSION INFORMATION

6. Type of Application/Submission Which This Certification Accompanies

☒ IND ☐ NDA ☐ ANDA ☐ BLA ☐ PMA ☐ HDE ☐ 510(k) ☐ PDP ☐ Other

7. Include IND/NDA/ANDA/BLA/PMA/HDE/510(k)/PDP/Other Number *(If number previously assigned)*:

If BLA was selected in item 6, provide Supplement Number:

8. Serial Number Assigned to Application/Submission Which This Certification Accompanies: 00000

##### CERTIFICATION STATEMENT / INFORMATION

9. Check only one of the following boxes *(See instructions for additional information and explanation)*

☐ A. I certify that the requirements of 42 U.S.C. § 282(j), Section 402(j) of the Public Health Service Act, including 42 CFR part 11, do not apply because the application/submission which this certification accompanies does not reference any clinical trial.

☒ B. I certify that the requirements of 42 U.S.C. § 282(j), Section 402(j) of the Public Health Service Act, including 42 CFR part 11, do not apply to any clinical trial referenced in the application/submission which this certification accompanies.

☐ C. I certify that the requirements of 42 U.S.C. § 282(j), section 402(j) of the Public Health Service Act, apply to one or more of the clinical trials referenced in the application/submission which this certification accompanies and that the requirements of 42 U.S.C. 282(j), including any applicable provisions of 42 CFR part 11, have been met.

*Certification Statement / Information section continued on page 2*

#### CERTIFICATION STATEMENT / INFORMATION *(Continued)*

10. If you checked box C, in number 9, provide the National Clinical Trial (NCT) Number(s) for any "applicable clinical trial(s)," for which you (the sponsor/applicant/submitter) are the "responsible party" under 42 U.S.C. § 282(j)(1)(a)(i), section 402(j)(1)(a)(i) of the Public Health Service Act referenced in the application/ submission which this Certification accompanies. (Add continuation page as necessary.)

NCT Number(s): ________ ________ ________ ________ ________

Continuation Page for #10

The undersigned declares, to the best of her/his knowledge, that this is an accurate, true, and complete submission of information. I understand that the failure to submit the certification required by 42 U.S.C. § 282(j)(5)(B), section 402(j)(5)(B) of the Public Health Service Act, and the knowing submission of a false certification under such section are prohibited acts under 21 U.S.C. § 331, section 301 of the Federal Food, Drug, and Cosmetic Act.

**Warning:** A willfully and knowingly false statement is a criminal offense, U.S. Code, title 18, section 1001.

11. Name and Title of the Person who Signs Number 15

| Name | Title |
|---|---|
| Ari Grinspan | Associate Professor |

12. Address

Address 1 *(Street address, P.O. box, company name c/o)*: One Gustave L. Levy Place Box 1069

Address 2 *(Apartment, suite, unit, building, floor, etc.)*: New York

City: New York

State/Province/Region: New York

Country: USA

ZIP or Postal Code: 10029

13. Telephone and Fax Numbers *(Include country code if applicable and area code)*

(Tel): 212-241-7757

(Fax): 646-537-8921

14. Date of Certification

11/02/2022

15. Signature of Sponsor/Applicant/Submitter or an Authorized Representative *(Sign)*

Sign

Ari Grinspan Digitally signed by Ari Grinspan Date: 2022.11.02 14:54:51 -04'00'

This section applies only to requirements of the Paperwork Reduction Act of 1995.

*****DO NOT SEND YOUR COMPLETED FORM TO THE PRA STAFF EMAIL ADDRESS BELOW.*****

The burden time for this collection of information is estimated to average 15 minutes and 45 minutes (depending on the type of application/ submission) per response, including the time to review instructions, search existing data sources, gather and maintain the data needed and complete and review the collection of information. Send comments regarding this burden estimate or any other aspect of this information collection, including suggestions for reducing this burden to:

Department of Health and Human Services
Food and Drug Administration
Office of Chief Information Officer
Paperwork Reduction Act (PRA) Staff
**

*"An agency may not conduct or sponsor, and a person is not required to respond to, a collection of information unless it displays a currently valid OMB number."*

#### INSTRUCTIONS FOR COMPLETION OF FORM FDA 3674 – CERTIFICATION OF COMPLIANCE
##### Under 42 U.S.C. § 282(j)(5)(B), with Requirements for ClinicalTrials.gov Data Bank

Form FDA 3674 must accompany an application/submission, including certain amendments, supplements, and resubmissions, submitted under §§ 505, 515, 520(m), or 510(k) of the Federal Food, Drug, and Cosmetic Act or § 351 of the Public Health Service Act.

**1. Name of Sponsor/Applicant/Submitter –** This is the name of the sponsor/applicant/submitter of the drug/ biologic/device application/submission which the certification accompanies. The name must be identical to that listed on the application/submission.

**2. Date –** This is the date of the application/submission which the certification accompanies or should have accompanied, if submitted at a subsequent date.

**3. & 4.** Provide complete address, telephone number, and fax number of the sponsor/applicant/submitter.

**5. Product Information –** ***For Drugs/Biologics:*** Provide the established, proprietary name, and/or chemical/ biochemical/blood product/cellular/ gene therapy name(s) for the product covered by the application/ submission. Include all available names by which the product is known. ***For Devices:*** Provide the common or usual name, classification, trade or proprietary or model name(s), and/or model number(s). Include all available names/model numbers by which the product is known.

Use a continuation page only if you have filled in all available spaces.

**6. Type of Application/Submission –** Identify the type of application/submission which the certification accompanies by checking the appropriate box. If the name of the type of application/submission is not identified (i.e. a new IND clinical protocol), check the box labeled "Other."

**7. IND/NDA/ANDA/BLA/PMA/HDE/510(k)/PDP/Other Number –** If FDA has previously assigned a number associated with the application/ submission which this certification accompanies, list that number in this field. For example, if the application/submission accompanied by this certification is an IND protocol amendment and the IND number has already been issued by FDA, that number should be provided in this field.

**8. Serial Number –** In some instances a sequential serial number is assigned to the submission/application. If there is such a serial number, provide it in this field. If there is no such number, leave this field blank.

**9. Certification –** This section contains three different check-off boxes.

**Box A** should be checked if the sponsor/applicant/submitter has concluded that the requirements of 42 U.S.C. § 282(j), section 402(j) of the Public Health Service Act, including 42 CFR part 11, do not apply because no data from clinical trials are included, relied upon, or otherwise referred to, in the application/submission which the certification accompanies.

**Box B** should be checked if the sponsor/applicant/submitter has concluded that the requirements of 42 U.S.C. § 282(j), section 402(j) of the Public Health Service Act, including 42 CFR part 11, do not apply at the time of submission of the certification to any clinical trials for which data are included, relied upon, or otherwise referred to, in the application/submission which the certification accompanies.

**Box C** should be checked if the sponsor/applicant/submitter has concluded that the requirements of 42 U.S.C. § 282(j), section 402(j) of the Public Health Service Act, do apply, on the date the certification is signed, to some or all of the clinical trials for which data are included, relied upon, or otherwise referred to, in the application/submission which the certification accompanies and for which the sponsor/ applicant/submitter is the "responsible party" of the applicable clinical trial included in the application/ submission. This means that, as of the date the certification is signed, the requirements of 42 U.S.C. § 282(j), section 402(j) of the Public Health Service Act, and any provisions of 42 CFR part 11 that apply to one or more of the applicable clinical trials included, relied upon, or otherwise referred to, in the application/submission which this certification accompanies have been met by the sponsor/applicant/ submitter as the "responsible party."

**10. National Clinical Trial (NCT) Numbers –** If you have checked Box C in number 9 (Certification), provide the NCT Number obtained from *www.ClinicalTrials.gov* for each applicable clinical trial for which the sponsor/applicant/submitter is the "responsible party" and for which data is included, relied upon, or otherwise referred to, in the application/submission which the certification accompanies. Type only the number, as the term "NCT" will be added automatically before number. Include any and all NCT numbers that, as of the date the certification is signed, have been assigned to the clinical trials included, relied upon, or otherwise referred to, in the application/submission which this certification accompanies. Multiple NCT numbers may be required for a particular certification, depending on the number of "applicable clinical trials" for which data is included, relied upon, or otherwise referred to, in the application/submission which the certification accompanies and for which the sponsor/applicant/submitter is the "responsible party." Leave this field blank if you have checked Box 9.C but, at the time the certification is completed, you have not yet received any NCT number(s) for the "applicable clinical trial(s)" for which data is included, relied upon, or otherwise referred to in the application/ submission. Use a continuation page only if you have filled in all available spaces.

**11. Name and Title of Person Who Signed in number 11 –** Include the name and title of the person who is signing the certification. If the person signing the certification is not the sponsor/applicant/submitter of the application/submission, he or she must be an authorized representative of the sponsor/applicant/submitter.

**12 & 13.** Provide the full address, telephone and fax numbers of the person who is identified in number 11 and signs the certification in number 15.

**14.** Provide the date the certification is signed. This date may be different from the date provided in number 2.

**15. Signature of Sponsor/Applicant/Submitter or an Authorized Representative –** The person signing the certification must sign in this field.

### PROTOCOL

[21 CFR 312.23(a)(6)]

### CLINICAL STUDY PROTOCOL

Phase 1b comparison of defined bacterial consortium MTC01 and Fecal Microbiota Transplantation for the treatment of recurrent *Clostridioides difficile* infection.

**PRODUCT NAME:** MTC01 (defined LBP) and FMT

**SPONSOR:** Ari Grinspan MD,
Department of Medicine/Division of Gastroenterology
Icahn School of Medicine at Mount Sinai Hospital, New York, NY
One Gustave L. Levy Place Box 1069
New York NY 10029
212-241-7757


#### 1. SYNOPSIS

##### Title of study
Phase 1b comparison of defined bacterial consortium MTC01 and Fecal Microbiota Transplantation for the treatment of recurrent *Clostridioides difficile* infection.

##### Investigator
Ari Grinspan, MD
Associate Professor of Medicine, Gastroenterology
Medicine, Gastroenterology
Expertise: Fecal Microbiota Transplantation (FMT) clinical studies

##### Study center
Mount Sinai Hospital

##### Phase of development
Phase 1b

##### Objectives
Safety, efficacy, and engraftment of defined 15 strain aaaaaaa, MTC01, for rCDI compared with FMT

##### Study synopsis
In this clinical trial for rCDI, individuals will be block randomized to receive either a high dose ($1x10^{11}$) or a low dose ($1x10^{10}$) of a complex FMT or a 15-strain defined LBP bacterial communities administered colonoscopically to the right colon (N = 15 individuals per group; randomization block size = 4; Figure 1). The groups will consist of an FMT and a defined LBP derived from a single FMT donor previously used to treat multiple rCDI recipients. Group 1 (D283.FMT) will be donor stool alone, while group 2 (MTC01) will consist of a defined community of 15 frequently engrafting bacterial strains isolated from donor D283.

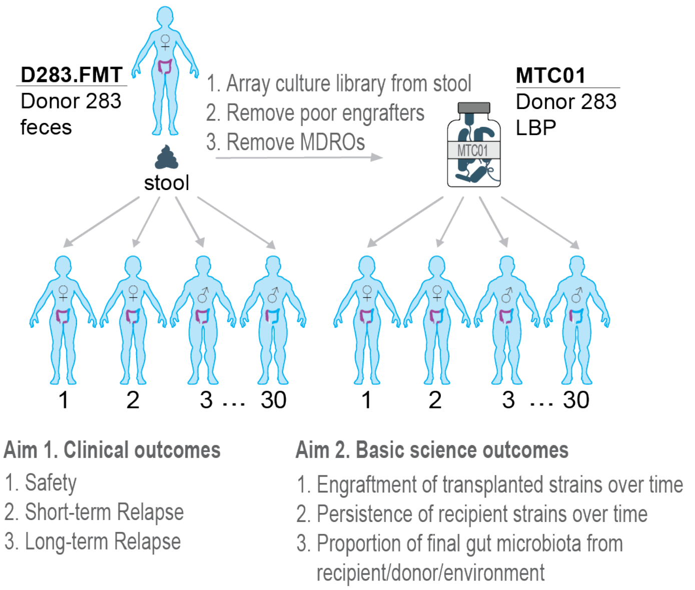

**Figure 1. Study synopsis.** Individuals will be block randomized (N=15/group; block size=4) to receive colonoscopic administration of either FMT from donor D283 [D283.FMT] or an LBP composed of the 15 frequent engrafting strains of D283 [MTC01] at either a high dose ($1x10^{11}$) or a low dose ($1x10^{10}$). We will compare safety, clinical response, donor microbiota engraftment and recipient microbiota persistence across both groups to understand differences between these therapeutic approaches.

We will compare clinical outcomes of safety, short-term recurrence (first 8 weeks), and long-term relapse (up to 6 months) between individuals receiving a stool transplant compared to those receiving an LBP transplant. We will also compare the engraftment of each strain in both the natural complex donor community and the synthetic *in vitro* manufactured community derived from the same donor. In addition, we will determine if the proportional engraftment of donor strains or the persistence of recipient strains is predictive of any clinical outcome while also quantifying the proportion of donor, recipient, and environmental strains making up the final gut microbiota of the recipient.

#### 2. INTRODUCTION

This study will evaluate the cultured, engrafting strain fraction of a successful FMT donor as a direct replacement of our current standard of care of FMT for rCDI at Mount Sinai. As per the IDSA definition of rCDI [1], "A recurrent case is defined as an episode of symptom onset and positive assay result following an episode with positive assay result in the previous 2–8 weeks". We will recruit individuals with ≥ 2 episodes of confirmed recurrent CDI defined as the presence of diarrhea (Bristol 6 or 7 for 48 hours) and a confirmatory test for CDI. Preferred testing will be a two-step method using GDH/EIA toxin, with the most recent episode occurring within the prior 3 months. Subjects will be taken off all antibiotics for 48 hr prior to the transplant and undergo a standard colonoscopy bowel preparation within 12 hours of their procedure. Our successful human FMTs have transplanted mean viable bacterial loads of approximately $10^{11}$ CFU per transplant of a largely poorly defined mixture of bacteria and metabolites. We have identified, isolated, and manufactured a subset of 15 strains from a single donor that consistently engrafted in colonoscopic FMT for rCDI [2]. This engrafting subset was cultured at scale in a sterile anaerobic environment, washed in USP grade PBS + cysteine (0.5%), and resuspended in USP grade PBS with 0.5% cysteine (oxygen scavenger) and 30% glycerol (cryoprotectant). We will compare the safety, efficacy, and engraftment of this defined 15-strain live biotherapeutic product, MTC01, with fecal microbiota transplantation, ideally from the same successful donor D283 that the MTC01 strains were isolated from. All 15 strains were detected in a recent stool sample from D283 collected approximately 5 years after the first use of this donor's stool for FMT. Therefore, it is likely that the transmission of the same strains will be tested in both the FMT groups and the MTC01 groups in addition to the larger set of strains from D283 that are not in MTC01.

For our clinical FMTs at Mount Sinai, we administer approximately $10^{11}$ microbes to the right colon by colonoscopy in a single infusion of 250ml with PBS as the diluent. Likewise for this trial, we will administer either FMT or MTC01 by a single infusion of $1 \times 10^{11}$ microbes [high dose] or $1 \times 10^{10}$ [low dose] in a volume of 250ml with USP PBS-glycerol-cysteine as the formulation media. For the FMT product, after safety screens are completed on the initial and final stool sample of a given batch, each stool sample will be homogenized and coarse filtered with a stomacher. The potency (CFU/ml) will be determined anaerobically and used to generate doses of $10^{11}$ in a volume of 250ml. For MTC01, each microbe will be evenly pooled in the consortium

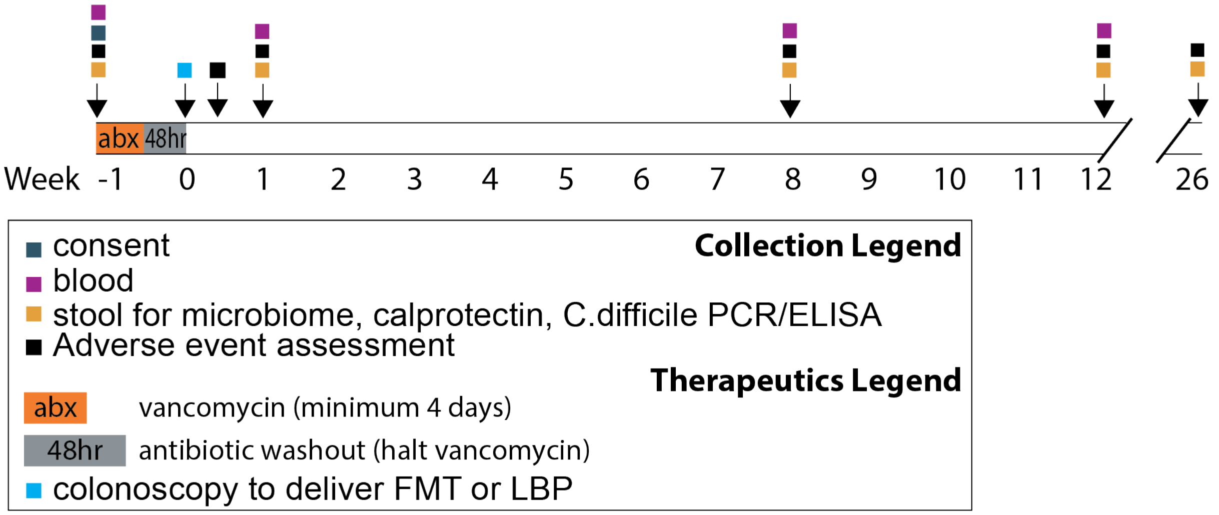

**Figure 2. Study design.**

where possible to reach the target CFU. The six strains that did not have sufficient density to be evenly pooled at $3x10^{10}$ were added at their maximal possible density at a range of $7.8x10^{8}$ (MTC01.12) to $2.8x10^{10}$ MTC01.01. Both MTC01 and FMT will also be tested at a lower dose of approximately $1x10^{10}$ to determine if less material could be used with similar engraftment and efficacy. Given that our high dose is on the low range of prior FMT studies, we do not expect these doses to impact safety. Therefore, we will block randomize (block size of 4) into all four groups rather than perform a dose escalation.

Each recipient will be monitored in the hospital for 1-hour post-transplant. We will follow-up with each subject at 72hr, at 1, 8, and 12 weeks, and at 26 weeks to monitor long term safety. Successful treatment will be determined by clinical cure at 8 weeks. We will collect stool pre-transplant and at 72hr, 1wk, 8wks, 12wks, and 26wks to quantify strain engraftment (Figure 2).

#### 3. SUBJECT SELECTION

***C. difficile diagnosis***

Pre*sence of C. difficile* will be determined by PCR or toxin EIA. As well as diarrhea (>=3 unformed stools in 24h period, defined as BSS 6-7, x2 days).

***Eligibility criteria***

Ages eligible for study: 18 years and older
Sexes eligible for study: All
Accepts healthy volunteers: No

***Inclusion criteria***

1. Ages eligible for study: 18 years and older
2. Able and willing to provide written informed consent
3. History of recurrent CDI defined as 2 episodes of CDI occurring within the previous 6 months (inclusive of the current episode)
4. Subjects with a qualifying recurrent CDI episode, defined as:
    a. History of diarrhea (>=3 unformed stools per day for 2 or more consecutive days that is clinically consistent with CDI
    b. Documented positive stool test by local laboratory for toxigenic *C. difficile* (toxin EIA or PCR-based testing) for the current CDI episode within 60 days prior to randomization.
    c. Received a course of standard-of-care (SOC) CDI antibiotics for the most recent CDI episode (for 10 to 42 days, with exact duration, antibiotic type and dose at the discretion of the Investigator)
    d. Demonstrated adequate clinical response, defined as <= 3 unformed stools per day for 2 or more consecutive days during SOC CDI antibiotics prior to randomization.
5. CDI symptoms started within 60 days prior to randomization.

***Exclusion criteria***

1. Female subjects who are pregnant or breastfeeding or are planning to become pregnant during the study. Women with reproductive potential should use a reliable method of birth control:
    a. Consistent use of an approved hormonal contraception (birth control pill/patches, rings)
    b. An intrauterine device (IUD)
    c. Contraceptive injection (Depo-Provera)

   d. Double barrier methods (Diaphragm with spermicidal gel or condoms with contraceptive foam)
   e. Sexual abstinence (no sexual intercourse) or
   f. Sterilization
2. Known or suspected toxic megacolon, ileus or bowel obstruction at the time of enrollment.
3. Subjects with active gastroenteritis due to infectious causes other than CDI
4. Subjects with allergies to ingredients present in the investigational product
5. Prior participation in studies of investigational live biotherapeutic products or FMT within the last 6 months.
6. History of active diarrhea associated with inflammatory bowel disease (IBD).
7. Major gastrointestinal surgery within the last 3 months before enrollment.
8. Use of drugs that alter gut motility.
9. History of acute leukemia or hematopoietic stem cell transplantation or myelosuppressive chemotherapy within 2 months prior to enrollment.
10. Unable or unwilling to undergo a colonoscopy
11. Inpatient status, though patients can be screened while inpatients, the must be outpatient for the planned colonoscopy.
12. Anticipated immediate or upcoming surgery within 30 days
13. Need for continued non-anti-CDI antibiotic therapy
14. History of total proctocolectomy
15. Patients who are unable to give informed consent
16. Participation in a clinical trial in the preceding 30 days or simultaneously during this trial
17. Severe food allergy (anaphylaxis or anaphylactoid-like reaction)
18. Life expectancy < 6 months
19. Unable to adhere to protocol requirements
20. Patient who have received an FMT in the past year
21. Any condition that the physician investigators deems unsafe, including other conditions or medications that the investigator determines that it will put the subject at greater risk from FMT
22. Clinically significant abnormal lab values including but not limited to WBC >15 x $10^3/mm^3$, ANC <0.5 x $10^3/mm^3$, or laboratory evidence of acute kidney injury at Investigator's discretion, at screening
23. If a patient is heavily immunosuppressed and is negative for CMV or EBV

#### 4. TREATMENTS

Single dose of either: 1) $1x10^{11}$ CFU FMT, 2) $1x10^{10}$ CFU FMT, 3) $1x10^{11}$ CFU MTC01, or 4) $1x10^{10}$ CFU MTC01 ($8x10^8$ to $3x10^{10}$ CFU per strain in high dose; 1:10 lower in low dose) administered via colonoscopic infusion to the right colon.

#### 5. OUTCOMES

***Primary outcome measures***
Safety and tolerability of MTC01 and FMT determined by incidence of serious adverse events (SAE), adverse events (AE), vital signs, ECG and physical examination up to 24 weeks.

***Secondary outcome measures***
1. Recurrence of CDI up to 8 weeks after treatment. Recurrence is defined as positive *C. difficile* toxin EIA as well as diarrhea (>=3 loose stools in 24h period, defined as BSS 6-7, x 2 days) within 8 weeks of receiving FMT.

2. Characterize the engraftment of MTC01 and FMT in the fecal microbiome 2 weeks after treatment.
3. Characterize MTC01 and FMT related taxonomic changes in the fecal microbiome 2 weeks after treatment.

***Other outcome measures***
1. Time to recurrence of CDI (up to 24 weeks).
2. Characterize MTC01 and FMT related taxonomic changes and engraftment of MTC01 in the fecal microbiome at 72hr and 1, 8, 12, and 26 weeks after treatment.

**Main parameters of safety**

***Primary Safety Outcomes Measures***
1. Proportion of participants with an AE through week 8 (± 3 days)
2. Proportion of participants with a SAE through week 8 (± 3 days)

***Secondary Safety Outcome Measures:***
1. Proportion of participants with an AE through week 24 (± 5 days)
2. Proportion of participants with a SAE through week 24 (± 5 days)

#### 6. FMT DONOR STOOL SELECTION AND SCREENING

**6.1 FMT Donor selection:** Donor stool will be collected from the frequently used FMT donor D283. If the donor stools from this individual fail donor stool screening, we will use another healthy volunteer stool.

**6.2 Donor screening:** Health screening procedures for the donor will include questionnaires that will assess for the following (Appendix 14): (1) Infectious risk factors: risk for factors for multi-drug resistant organisms; high risk sexual behaviors and social history including illicit drug use, (2) Potential microbiome-mediated conditions and general health status: gastrointestinal comorbidities, metabolic comorbidities, neurological comorbidities, psychiatric comorbidities, chronic pain syndromes, infectious diseases, autoimmune diseases, atopy, asthma and allergies (food and other), malignancy, surgeries/other medical history, current symptoms medications including antimicrobial therapy, diet, and family history, (3) Pregnancy and breastfeeding status for female donors. Donors will be tested for SARS-CoV-2 via nasal swab PCR at the following time points: 14 days before collection, day 1 of collection, day 15 of collection, day 30 of collection, and then 30 days after final day of collection. Each stool donation will be cleared by confirmation that the donor remained asymptomatic for 14 days post donation. Additional screening will include the tests detailed in Sections 3.1.1. and 3.1.2. Stool screening test will be within 30 days of donation. HIV serological testing will be performed at least 2 weeks after the donation and a negative result will be confirmed prior to administration of product manufactured from this donation. The donor will complete a brief questionnaire to monitor general health during each donation visit (Appendix 14).

6.2.1 Blood tests 14-21 days before collection and 30 days after collection will be performed by the CLIA certified Icahn School of Medicine at Mount Sinai Pathology Laboratory (CLIA certificate Appendix 15):
- CBC w/ differential
- LFTs, complete

- Bun/Cr
- Fasting glucose
- High Sensitivity CRP
- Serum triglycerides, LDL, HDL cholesterol
- HIV 1/2 (IgG) and E1A
- HAV (IgG and IgM)
- HBV (surface antigen/antibody; core antibody)
- HCV (IgG)
- Syphilis serology (RPR done if treponemal test is positive)
- HTLV 1&2 antibody

6.2.2 Stool tests on days 1 and 30 of collection will be performed by the CLIA certified Icahn School of Medicine at Mount Sinai Pathology Laboratory (CLIA certificate Appendix 15):

- Clostridium difficile toxin by PCR
- Fecal Giardia antigen
- Fecal Cryptosporidium antigen
- Ova and parasites by light microscopy
- Helicobacter pylori fecal antigen
- Mount Sinai Hospital NAAT/Biofire: campylobacter, plesiomonas, salmonella, yersinia, vibrio spp, vibrio cholerae, enteroaggregative e coli, enteropathogenic e coli, enterotoxigenic e coli, shiga-like toxin producing e coli, e coli 0157, shigella/enteroinvasive e coli, cryptosporidium spp, Cyclospora cayetanensis, entamoeba histolytic, giardia lamblia, adenovirus F 40/41, astrovirus, norovirus gi/gii, rotavirus A, sapovirus (I, II, IV, V)

6.2.3 Stool tests on days 1 and 30 of collection will be performed by the Icahn School of Medicine Microbiome Translational Center, not CLIA certified:

- VRE via agar plates
- MSRA via agar plates
- ESBL fecal surveillance culture via Mayo Clinic Laboratories

6.2.4 Donor exclusion criteria:

- Any past or current malignancy
- History of irritable bowel syndrome, excessive gas, bloating, lymphocytic colitis, idiopathic chronic constipation, chronic use of laxatives or chronic diarrhea
- Any chronic medications with the exception of hormonal birth control
- Use of probiotics or any OTC aids for regulating digestion
- Antibiotics within the preceding 6 months
- Major immunosuppressive medications, e.g., calcineurin inhibitors, exogenous glucocorticoids, biologic agents, etc.
- Systemic anti-neoplastic agents
- History of gastrointestinal surgery (e.g., gastric bypass) or endoscopy
- Metabolic syndrome
- Neurological, neurodevelopmental disorder e.g. Parkinson's disease, autism, etc.
- Systemic autoimmunity, e.g., multiple sclerosis, psoriasis, vasculitis, connective tissue disease, any rheumatological or inflammatory condition
- Atopic diseases, e.g. asthma and eczema, food allergies, eosinophilic disorders of the gastrointestinal tract
- Chronic pain syndromes, e.g., chronic fatigue syndrome, fibromyalgia

- Travel to underdeveloped countries in the past year. Short travel to major developed cities in Europe, Asia, and Australia will be subject to PI discretion.
- Positive screening test result
- Suspected or confirmed SARs-CoV-2 infection or exposure within the previous 8 weeks
  - Detection of SARs-CoV-2 will result in exclusion of the donor and exclusion from clinical use of any FMT manufactured from stool donated by the affected donor beginning 4 weeks prior to the first positive test.
  - We will reinstate donors who test positive or have a known or suspected exposure after a minimum of 8 weeks following the positive test or exposure.
- Suspected or confirmed monkeypox infection or exposure
  - Suspected or confirmed monkeypox infection or exposure will result in exclusion of the donor and exclusion from clinical use of any FMT manufactured from stool donated by the affected donor beginning 8 weeks prior to the first positive test.

6.2.5 Stool collection: Stool samples will be collected at the Inflammatory Bowel Disease Center at the Icahn School of Medicine on 17E 102nd Street, New York, New York 10029. The sample will be collected by the patient. The patient will deposit the bowel movement into the collection hat and then use a wooden stick to move stool to the different sample containers. The samples will be transported by the research staff to the Microbiome Translational Center. The period of time over which donations and screening will occur will not exceed 60 days per screening window. All FMT products will be held in quarantine until after post-donation testing and screening procedures are complete.

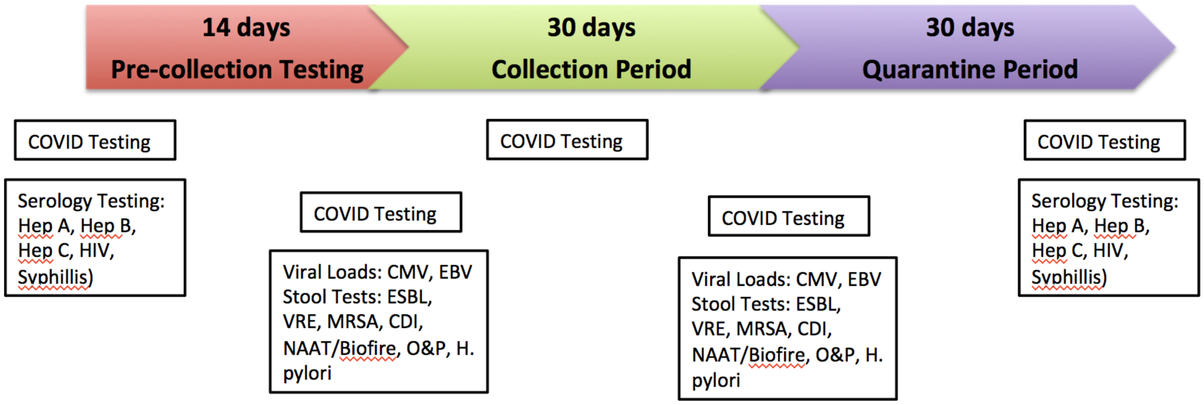

**Figure 3. FMT donor screening**

#### 7. STUDY DESIGN

This is an open-label pilot study to assess the safety, microbiological and clinical impacts of a defined LBP MTC01 compared with FMT in patients with recurrent CDI. We will prospectively enroll 60 patients with recurrent CDI from one tertiary care referral center over the course of 3 years. Patients will receive either a high dose $10^{11}$ or low dose $10^{11}$ of MTC01 or FMT (4 groups; 15 patients per group). A visual overview of the study design is in Figure 2. The study details are below.

##### 7.1 Visits

***Visit 1 – Screening (Day -45 to Day 0)***
Potential subjects will undergo the following screening procedures no more than 4 weeks prior to administration of MTC01 or FMT to determine if they meet the recipient selection criteria.

1. Medical record review will be done to confirm diagnosis and inclusion criteria. Records will be reviewed for exclusion criteria, however if not available patient self-report will be accepted. The following data will be documented at baseline visit:
    a. Demographic: age, height, gender, weight, race, significant past medical history, and smoking status.
    b. Diarrheal symptom assessment: Average daily Bristol score, number of daily BMs.
    c. CDI history: inciting antibiotic, Number and approximate date of previous CDI episodes, previous positive stool tests—dates, PCR versus EIA, CDI treatment courses (metronidazole, vancomycin, vancomycin taper, or fidaxomicin), CDI related prior hospitalizations and number of CDI related hospitalizations.
2. Baseline symptom assessment using diary and targeted physical exam by study physician or nurse
3. Laboratory assessments:
    a. Blood: CBC, CRP, CMP, HIV, Viral hepatitis, Syphilis, and ONLY subjects who are heavily immunosuppressed will also be screened for CMV and EBV.
    b. Stool: Fecal calprotectin, microbiome analysis. Stool will be collected on chronic anti-CDI therapy.
4. Stool and blood will be banked for metabolomics and future analysis.
5. Urine Pregnancy Test if applicable. If patients are post-menopausal this will be documented and pregnancy test is not needed
6. If not currently on anti-CDI therapy Vancomycin should be started for a minimum of 4 days prior to MTC01 or FMT dosage with instruction to hold for 48 hours prior to MTC01 or FMT dosage.
7. Once patient has passed screening and all labs have been reviewed colonoscopy will be scheduled.

***Visit 2 – Day 1 (Randomization to MTC01 or FMT)***
The following baseline assessments will be made in enrolled subjects on day of scheduled microbial therapeutic treatment with either MTC01 or FMT. Subjects will be randomized (block size 4) to one of the four treatment groups of low/high dose with FMT/MTC01. The microbial therapeutic will be administered in the right colon by colonoscopy. Standard colonoscopy procedures will be followed per Mount Sinai Hospital policy. Patients will be monitored for at least 30 minutes after the procedure to ensure FMT tolerance and record/address any treatment related adverse event.

1. Urine pregnancy test (HCG) for female patients if applicable. If patients of post-menopausal this will be documented, and pregnancy test is not needed.

#### ***Visit 3, 4, 5 – Week 1, 8 and 12 post MTC01 or FMT***
Week 1, 8, 12 +/- 3 days post MTC01 or FMT: patients will be evaluated in the clinic for follow-up assessments. The following will be documented:

1. Assessment of efficacy: Assessment of diarrheal symptoms as well as testing for CDI by GDH/EIA and PCR regardless of symptoms at week 1, 8 and 12 post MTC01 or FMT dosage.
2. Assessment for related AEs to MTC01 or FMT will occur at each visit using NIH criteria. Related AEs include but are not limited to:

    a. Diarrhea
    b. Bloating, distention, and flatulence
    c. Constipation
    d. Abdominal pain
    e. bacteremia
    f. infection transition
    g. headache
    h. nausea and vomiting
    i. pyrexia
3. Laboratory assessments:
    a. Stool: CDI testing (as above), fecal calprotectin, microbiome analysis.
    b. Stool and blood will be banked for metabolomics and future analysis.
4. Vital Signs and targeted physical exam
5. List of current concomitant medications

***Visit 6 – Week 26***

Visit 6 (Week 26 +/- 7 days post MTC01 or FMT): Study Phone Call. Patients will be called to assess the following:

1. AE assessment
2. Patients will mail in samples for microbiome analysis only

Patient obligations in the study will end at week 26 however the patient's medical record will be followed prospectively for evaluation of CDI outcomes and procedures done for clinical care for 1-year post-enrollment.

***Early Termination Visit:***

In the case of an early termination, study staff will complete an 'Early Termination CRF', if possible, and the following will be assessed:

1. Assessment of efficacy: Assessment of diarrheal symptoms as well as testing for CDI by GDH/EIA and PCR regardless of symptoms
2. Assessment for related AEs to MTC01 or FMT will occur at each visit using NIH criteria. Related AES include but are not limited to:
    a. Diarrhea
    b. Bloating, distention
    c. Constipation
    d. Abdominal pain
    e. bacteremia
    f. infection transition
    g. headache
    h. nausea
    i. pyrexia
3. Laboratory assessments:
    a. Blood: CBC, CRP, CMP
    b. Stool: CDI testing (as above), fecal calprotectin, microbiome analysis.
    c. Stool and blood will be banked for metabolomics and future analysis
4. Vital Signs and targeted physical exam.

***Unscheduled Visit***

At any point during the study if patients experience worsening of symptoms they may be brought in by study staff for an unscheduled visit for an assessment. This visit will include:

1. Assessment of efficacy: Assessment of diarrheal symptoms as well as testing for CDI by GDH/EIA and PCR regardless of symptoms at week 1, 8 and 12 post MTC01 or FMT dosage.
2. Assessment for related AEs to MTC01 or MTC dosage will occur at each visit using NIH criteria. Related AEs include but are not limited to:
    a. Diarrhea
    b. Bloating, distention
    c. Constipation
    d. Abdominal pain
    e. Bacteremia
    f. infection transition
    g. headache
    h. nausea
    i. pyrexia
3. Laboratory assessments:
    a. Blood: CBC, CRP, CMP (to be done locally)
    b. Stool: CDI testing (as above), fecal calprotectin, microbiome analysis.
4. Stool and blood will be banked for metabolomics and future analysis.
5. Vital Signs and targeted physical exam
6. List of current concomitant medications

##### 7.1.1 Schedule of assessments

| Assessments | Screening V1 | Open-label treatment period | | | | Follow-up V6 |
|---|---|---|---|---|---|---|
| | | V2 | V3 | V4 | V5 | |
| | Day -45 to Day 0 | Day 1 | Day 7 (± 3 days) | Day 56 (± 3 days) | Day 84 (± 3 days) | Week 26 (± 7 days) |
| Medical History | x | | | | | |
| Baseline Symptoms | x | | | | | |
| Physical exam | x | | x | x | x | |
| Demography | x | | | | | |
| Inclusion/Exclusion Criteria | x | | | | | |
| List of concomitant medications | x | x | x | x | x | x |
| safety assessment | | | x | x | x | x |
| Blood tests: | | | | | | |
| *-CBC, CRP, CMP, HIV, Viral hepatitis, syphilis* | x | | | | | |
| *-CMV and EBV (immunosuppressed subjects only)* | x | | | | | |
| Stool tests: | | | | | | |
| *-microbiome analysis* | x | | x | x | x | x |
| Urine test: | | | | | | |
| -pregnancy test (if applicable) | x | x | | | | |
| Randomization | | x | | | | |
| Colonoscopic infustion of microbial therapeutic | | x | | | | |
| CDI tests: | | | | | | |
| *-GDH/EIA* | | | x | x | x | |
| *-PCR* | | | x | x | x | |
| -diarrheal symptoms | | | x | x | x | |

#### 7.2 Patients Treatment Strategy

The below treatment strategy will be utilized based on results obtained at each visit.

**Table 1.** Treatment strategy

| Symptoms | Step 1: PCR | Step 2: EIA | CDI recurrence | Treatment Course |
|---|---|---|---|---|
| Diarrhea | + | + | Yes | Anti-CDI Therapy including antibiotics or FMT |
| Diarrhea | + | - | No | Likely colonization, treatment at Clinician discretion |
| Diarrhea | - | - | No | No CDI, evaluate for other causes of diarrhea and treat accordingly |
| No Diarrhea | + | + | No | Asymptomatic Carriage, No anti-CDI treatment needed |
| No Diarrhea | + | - | No | Asymptomatic Carriage, No anti-CDI treatment needed |
| No Diarrhea | - | - | No | No treatment needed |

#### 7.3 Recruitment Procedures

Gastroenterologists, including attending physicians and fellows, will be informed of the study's aims and inclusion criteria. These doctors will inform the principal investigator or site lead investigators of patients who meet the study criteria and who may be good candidates for the study. The treating gastroenterologist will introduce the study to the potential patient and request the patient's permission to be approached by study staff. We will not utilize advertisement material or other informational materials for patients beyond the consent form and the posting on clinicaltrials.gov. Patients will be given as much time as they need to decide. They will be given a copy of the consent form to take home, read, and consider, and they will be encouraged to discuss participation with family members and health care providers.

The trial will be posted on clinicaltrials.gov. If patients reach out to us via the provided E-mail or phone number located at the clinicaltrials.gov website, or directly reach out to the study team via any mechanism, we will pre-screen them using questions from the provided E-mail/phone script. If the patient's responses do not indicate any exclusion criteria being met, then we will contact their primary care provider or GI provider via email or phone call (with patient permission) to confirm medical diagnosis. If no records exist for exclusion criteria patient report will be accepted.

We plan to recruit approximately 20 subjects per year (5 per group).

#### 7.4 Consent Procedures

The treating gastroenterologist will introduce the study to the potential patient and request the patient's permission to be approached by study staff. With the treating gastroenterologist's permission, either a physician investigator or study coordinator will describe the research study in detail, including participation and risks and alternative courses of treatment, and answer any questions or concerns that the patient may have. Patients will be given as much time as they need to consider participation before signing the consent form.

Subjects will be recruited from the Susan and Leonard Feinstein IBD Clinical Center. In order to avoid coercion, study staff will reinforce that participation is voluntary and that their decision will not affect the medical care that they receive now or in the future. If patients seek more time to consider participation, they will be given a copy of the consent form and encouraged to discuss the study with family, friends, PCP, or others. Study staff will follow-up to see if any questions or concerns have not been addressed. A physician investigator will obtain informed consent signatures.

If patients initiate contact with the study team and the initial phone screen suggests eligibility they will be provided with the consent form via email to review. They will be re-contacted in a few days so that any questions may be answered. If they would like to proceed, a screening visit will be set up.

**7.5 Specimen Handling and Shipping**

All patients will be assigned a unique participant ID. Clinical samples will be processed at the Microbiome Translation Center at the Icahn School of Medicine at Mount Sinai. Additionally, blood and stool samples will be aliquoted and banked.

**7.6 Assessment of safety**

Safety will be assessed by the frequency and severity of adverse events (AE)

***Definition of an Adverse events (AE)***

Adverse events (AEs) will be recorded at each regular scheduled study visit in the study patient record (source document) as well as on a specific AE CRF.

An AE is any untoward medical occurrence in a study patient or clinical investigation subject administered a pharmaceutical product and which does not necessarily have a causal relationship with this treatment. An AE can therefore be any unfavorable and unintended sign (including an abnormal laboratory finding), symptom, or disease temporally associated with the use of a medicinal product, whether or not related to the medicinal product, e.g.:

- any new clinical diagnosis
- any symptom that requires medical clarification or leads to in-patient admission (surgery or accident)
- any suspected adverse drug reaction (ADR)
- any symptom that appears on the study patient's medical records
- any event related in time with the application of the study medication and affecting the health of the study patient (including laboratory value changes)

If there is any doubt as to whether a clinical observation is an AE, the event should be reported. AEs must be graded for severity and relationship to study product.

***NIH Grading of Severity of the Event***

AEs will be assessed by the clinician using the NIH protocol defined grading system (see Appendix 16). Briefly, the criteria for estimating adverse event severity grade:

- Grade 1 (Mild): events require minimal or no treatment and do not interfere with the patient's daily activities.
- Grade 2 (Moderate): events result in a low level of inconvenience or concern with the therapeutic measures. Moderate events may cause some interference with functioning.
- Grade 3 (Severe): events interrupt a patient's usual daily activity.
- Grade 4 (Potentially life threatening): Events result in inability to perform basic selfcare functions or the need for medical or surgical intervention to prevent permanent disability or death.
- Grade 5 (Death)

***NIH Adverse Event Relatedness***

The clinician's assessment of an AE's relationship to MTC01 or FMT is part of the documentation process, but it is not a factor in determining what is or is not reported in the study. If there is any doubt as to whether a clinical observation is an AE, the event should be reported.
The following NIH guidelines of relatedness are used:

- Related: The adverse event is related to the MTC01 or FMT material – i.e. an event that follows a reasonable temporal sequence from administration of the MTC01 or FMT material, follows a known or expected response pattern to the MTC01 or FMT material, that is confirmed by improvement on stopping and reappearance of the event on repeated exposure and that could not be reasonably explained by the known characteristics of the patient's clinical state.
- Possibly Related: The adverse event follows a reasonable temporal relation to MTC01 or FMT administration, however, symptom may be related to other factors.
- Not Related – The adverse event is not related to the MTC01 or FMT material. - i.e. another cause of the event is most plausible; and/or a clinically plausible temporal sequence is inconsistent with the onset of the event and the study intervention and/or a causal relationship is considered biologically implausible

***Solicited mild to moderate adverse events:***
In addition to open-ended questions on adverse events meeting the above definitions, specific potential adverse events will be inquired about during the follow up period:

| Symptom that is clinically more severe than participant's baseline | Severity | | | | |
|---|---|---|---|---|---|
| | Grade 1 | Grade 2 | Grade 3 | Grade 4 | Grade 5 |
| Fever* | 38.0 - 39.0 degrees C (100.4 - 102.2 degrees F) | >39.0 - 40.0 degrees C (102.3 - 104.0 degrees F) | >40.0 degrees C (>104.0 degrees F) for <=24 hrs | >40.0 degrees C (>104.0 degrees F) for >24 hrs | Death |
| Diarrhea | Increase of <4 stools per day over baseline pre-MTC01 or pre-FMT; mild increase in ostomy output compared to baseline | Increase of 4 - 6 stools per day over baseline; moderate increase in ostomy output compared to baseline; limiting instrumental ADL | Increase of >=7 stools per day over baseline; incontinence; hospitalization indicated; severe increase in ostomy output compared to baseline; limiting self-care ADL | Life-threatening consequences; urgent intervention indicated | Death |
| Vomiting | 1 - 2 episodes (separated by 5 minutes) in 24 hrs | 3 - 5 episodes (separated by 5 minutes) in 24 hrs | >=6 episodes (separated by 5 minutes) in 24 hrs; tube feeding, TPN or hospitalization indicated | Life-threatening consequences; urgent intervention indicated | Death |
| Abdominal Pain | Mild pain | Moderate pain; limiting instrumental activities of daily life | Severe pain; limiting self care activities of daily life | n/a | n/a |
| Bloating | No change in bowel function or oral intake | Systemic, decreased oral intake; change in bowel function | n/a | n/a | n/a |
| Constipation | Occasional or intermittent symptoms; occasional use of stool softeners, laxatives, dietary | Persistent symptoms with regular use of laxatives or enemas indicated; limiting instrumental | Symptoms interfering with self-care activities of daily life; obstipation with manual evacuation | Life-threatening consequences (e.g. obstruction, toxic megacolon); urgent | Death |

| | | | | |
|---|---|---|---|---|
| | modification, or enema | activities of daily life | indicated | intervention indicated |

***Serious Adverse Events***

- An adverse event or suspected adverse reaction is considered “serious” if, in the view of either the investigator or sponsor, it results in any of the following outcomes:
- Death;
- Life-threatening adverse event*;
- Inpatient hospitalization or prolongation of existing hospitalization;
- A congenital anomaly/birth defect;
- Persistent or significant disability or incapacity or substantial disruption of the ability to conduct normal life function.

**Life-threatening adverse event. An adverse event is considered “life-threatening” if, in the view of either the investigator or sponsor, its occurrence places the patient or subject at immediate risk of death. It does not include an adverse event which, had it occurred in a more severe form, might have caused death.*

Important medical events that may not result in death, be life-threatening, or require hospitalization may be considered serious when, based upon appropriate medical judgment, they may jeopardize the patient or subject and may require medical or surgical intervention to prevent one of the outcomes listed in this definition
Any adverse event or suspected adverse reaction that meets the criteria for serious adverse event will be:

- recorded on the appropriate SAE CRF
- followed through resolution by a study clinician
- reviewed and evaluated by a study clinician

***Unsolicited Adverse Events***

On enrollment in the study, the study participants will be instructed to contact the study PI if an AE occurs. All unsolicited non-serious adverse events will be collected from the time of MTC01 or FMT dosage until 6 months following MTC01 or FMT dosage and will be assessed for relatedness as outlined in section 5.6.3. Patients will be given a patient diary with date, time, details and action taken to help with data collection. Patients will bring this diary to the study PI for evaluation at each follow-up visit and will be instructed to seek immediate medical attention if indicated.

***Medically Attended Adverse Events (MAAEs)***

A MAAE is defined as an AE that leads to an unscheduled visit to a healthcare provider. MAAEs will be recorded for six months after FMT administration.

***New-Onset Related Chronic Medical Condition***

Study MTC01 or FMT-related chronic medical conditions occurring from the time of the MTC01 or FMT dosage until 6 months following MTC01 or FMT dosage. Specifically, new-onset chronic medical conditions potentially related to MTC01 or FMT dosage such as weight gain, glucose intolerance, autoimmune conditions, and metabolic syndrome will be monitored for. This is be done via patient report.

#### 7.7. Reporting of Adverse Events

Study participants will be instructed to contact the study nurse or doctor if any serious or unexpected adverse event occurs. Study staff will enquire about using a generally worded question, about AEs at each study visit. Reported AE's will be recorded in detail in an AE CRF.
AE information to be collected in the AE CRF:

- Nature of the event
- Time of onset: date, time
- Concomitant treatment: product (generic name), indication, dosage, dosage interval, presentation, mode of administration, administration regimen
- Duration of the AE
- Severity
- Seriousness
- Causality
- Outcome

The course and outcome of the adverse event will be commented on as follows:

- Recovered without sequelae
- Not yet recovered
- Recovered with sequelae
- Fatal

Any SAE (including death, irrespective of the cause) occurring during the study will be immediately reviewed by the PI, i.e. within 24 hours and referred to the DSMB. In case of a SAE, the information will be reviewed by the PI and reported to the DSMB chair. If the SAE is judged by the DSMB to be related to the treatment, a report will be sent to the IRB of the site. The report must contain a detailed description of the symptoms observed and the concomitant treatment administered. Furthermore, the investigator must comment on a possible causative relationship between the AE and the trial medication. Each SAE must be followed until it is resolved or can be explained satisfactorily.

All non-serious adverse reactions will be reviewed by the DSMB at their regular meeting and or ad/hoc depending on the clinical case at the discretion of the study PI.
In accordance with safety requirements, the study PI will inform the local IRB and will make sure that the involved persons will obtain adequate information. The following instructions must be heeded:

- In the case of an intolerable SAE, the study patient must, at the decision of the investigator, be withdrawn from the clinical trial, and symptomatic treatment must be administered.
- The measures taken must be recorded on the CRF.
- In accordance with local legislation, the investigators will submit copies of the final SAE-report to the Regulatory Authorities concerned, if necessary.

##### 7.8 Follow-up of Subjects after Adverse Events

AEs will be followed until resolution or stability even if this extends beyond the study-reporting period. Resolution of an AE is defined as the return to pretreatment status or stabilization of the condition with the expectation that it will remain chronic.
Follow-up procedures, evaluations, and outcomes will be recorded on the subject's case report forms.

##### 7.9 Safety Oversight

Dr. Ryan Ungaro will chair the data safety and monitoring board for this clinical trial. Dr. Ungaro is well versed in reporting procedures as they are currently required for all research conducted at Mount Sinai. All research staff will complete training in compliance with Mount Sinai's IRB for conducting human subjects research.

**Data and Safety Monitoring Board (DSMB)**

Safety oversight for the trial will be under the direction of a DSMB. The DSMB is an independent group of experts, led by chairperson Dr. Ungaro, who will advise the study investigators. The primary responsibilities of the DSMB are to 1) periodically review and evaluate the data for subject safety, study conduct, study progress, and efficacy, and 2) make recommendations concerning the continuation, modification, or termination of the trial. The DSMB will be composed of at least 3 voting members. Members will have clinical expertise in the medical area and subject population being studied. All DSMB members will be separate and independent of study personnel participating in this trial and will not have scientific, financial or other conflict of interest related to the trial. Procedures for DSMB data reviews will be defined in the DSMB Charter that will include DSMB membership, responsibilities, and the scope and frequency of data reviews. The study will be reviewed by the DSMB at least annually and after 10 patients are enrolled.

##### 7.10 Halting Rules

***Study enrollment halting rules***

Enrollment in the study will be suspended for conduct of a safety review by the DSMB in the case of:

- A grade 3 or higher AE or SAE that is other than 'unrelated' to the study intervention.
- One case of allergic reaction or anaphylaxis, or one infection assessed as anything other than 'unrelated' to study intervention
- An overall pattern of symptomatic, clinical or laboratory events that the PI considers related to study product and that may appear minor in terms of individual events, but that may collectively represent a serious potential concern for safety.

***Individual's halting rules***

Subjects who meet any of the following criteria must be assessed by the PI to determine if it is in the subject's best interest to stop the study product(s):

- Subject choice (Withdrawal of consent)
- Participant's non-compliance.
- Development of a significant medical condition and/or participation in the study is no longer in the best interest of the subject.

##### 7.11 Statistical considerations

This is a pilot study, open-label study to determine the safety of MTC01 compared to FMT in patients with recurrent CDI. Preliminary efficacy/outcome data will also be collected. This study is exploratory in nature so a formal sample size calculation has not been performed. Our goal is to recruit 60 subjects. We believe this will allow us to assess safety, acceptability and engraftment.

##### 7.12 Source documents and access to data/documents

The PI will maintain appropriate medical and research records for this trial, in compliance with institutional requirements for the protection of confidentiality of subjects. Forms for use as source documents will be derived from the electronic CRFs. Original documents and data records include, but are not limited to, hospital records, clinical and office charts, laboratory notes, memoranda, pharmacy dispensing records, recorded data from automated instruments, copies or transcriptions certified after verification as being accurate and complete, microfiches, photographic negatives, microfilm or magnetic media, x-rays, and subject files and records kept at the pharmacy, laboratories, and medico-technical departments involved in the clinical trial. Original source documents will be maintained by each site.

**7.13 Quality control and quality assurance**
The PI and study coordinators are responsible for conducting routine quality assurance (QA) and quality control (QC) activities to internally monitor study progress and protocol compliance. The PI will have direct access to source data/documents and reports for the purpose of monitoring. Additionally, auditing by local and regulatory authorities will occur at their discretion. The PI will ensure all study personnel are appropriately trained and applicable documentations are maintained on site.

The Principal Investigator, Ari Grinspan, will assure the validity and integrity of the data and adherence to the IRB-approved protocols.

Study staff will review completed CRF's before each visit to ensure completeness of previous entries. Entries that need clarification will be reviewed by the PI, and the subject and/or treating gastroenterologist will be consulted if needed.

**5.14 Ethics/Protection of human subjects**

***Ethical Standard***
The PI will ensure that this study is conducted in full conformity with principles of the Belmont Report: Ethical Principles and Guidelines for the Protection of Human Subjects of Research of the National Commission for the Protection of Human Subjects of Biomedical and Behavioral Research (April 18, 1979) and codified in 45 CFR 46, 21 CFR 50 and 56, and ICH E6; 62 Federal Regulations 25691 (1997), if applicable. The investigator's Institution will hold a current Federal Wide Assurance (FWA) issued by the Office of Human Research Protection (OHRP) for federally funded research if applicable.

***Institutional Review Board (IRB)***
The study investigators will provide for the review and approval of this protocol and the associated informed consent documents, by an appropriate ethics review committee or IRB. Any amendments to the protocol or consent materials must also be approved before they are placed into use unless change is for the safety of the subject. Only those IRB members who are independent of the investigators should provide an opinion on study related matters. Verification of IRB approval of the protocol and the written informed consent will be transmitted by the investigator or designee prior to the shipment of clinical trial material. No deviations from or changes to the protocol will be initiated without prior approval of an appropriate amendment unless change is for the safety of the subject. The study investigators are responsible for ensuring Continuing Review at least once a year and for keeping the IRB apprised of the progress of the study and any changes to the protocol.

***Informed Consent Process***
The written consent document will embody the elements of informed consent as described in the Declaration of Helsinki and will adhere to the ICH Harmonized Tripartite Guideline for Good Clinical Practice. Informed consent should be implemented before any protocol-specified procedures or interventions are carried out. Informed consent will be obtained in accordance with 21 CFR 50.25 and 45 CFR 46. Information should be presented both orally and in written form.
An investigator or designee will describe the protocol to potential subjects face-to-face. The Subject Information and Consent Form may be read to the subjects, but, in any event, the

investigator shall give the subjects ample opportunity to inquire about details of the study and ask any questions before the signing and dating the consent form.

Study staff must inform subjects and/or legal guardian that the trial involves research, and explain the purpose of the trial, those aspects of the trial that are experimental, any expected benefits, all possible risks (including a statement that the particular treatment or procedure may involve risks to the subject or to the embryo or fetus, if the subject is or may become pregnant or fathers a child, that are currently unforeseeable), the expected duration of the subject's participation in the trial, the procedures of the research study, including all invasive procedures, and the probability for random assignment to treatment groups. Subjects and/or legal guardian will be informed that they will be notified in a timely manner if information becomes available that may be relevant to their willingness to continue participation in the trial. They must also be informed of alternative procedures that may be available, and the important potential benefits and risks of these available alternative procedures. Subjects and/or legal guardian must receive an explanation as to whether any compensation and any medical treatments are available if injury occurs, and, if so, what they consist of, or where further information may be obtained. Subjects and/or legal guardian must be informed of the anticipated financial expenses, if any, to the subject for participating in the trial, as well as any anticipated prorated payments, if any, to the subject for participating in the trial. They must be informed of whom to contact (e.g., the investigator) for answers to any questions relating to the research project. Information will also include the foreseeable circumstances and/or reasons under which the subject's participation in the trial may be terminated. The subjects and/or legal guardian must be informed that participation is voluntary and that they are free to withdraw from the study for any reason at any time without penalty or loss of benefits to which the subject is otherwise entitled.

Neither the investigator, nor the trial staff, should coerce or unduly influence a subject to participate or continue to participate in the trial. The extent of the confidentiality of the subjects' records must be defined, and subjects must be informed that applicable data protection legislation will be followed. Subjects and/or legal guardian must be informed that the monitor(s), auditors(s), IRB, and regulatory authority(ies) will be granted direct access to the subject's medical records for verification of trial procedures and/or data without violating the confidentiality of the subject, to the extent permitted by the applicable laws and regulations, and that, by signing a written informed consent form, the subject is authorizing such access. Subjects and/or legal guardian must be informed that records identifying the subject will be kept confidential, and, to the extent permitted by the applicable laws and/or regulations, will not be made publicly available and, if the results of the trial are published, the subject's identity will remain confidential.

Consent forms must be in a language fully comprehensible to the prospective subjects. Informed consent shall be documented by the use of a written consent form approved by the IRB and signed and dated by the subject and the person who conducted the informed consent discussion. The signature confirms that the consent is based on information that has been provided and all questions have been answered to the prospective subject's satisfaction. Each subject's signed informed consent form must be kept on file by the investigator for possible inspection by Regulatory Authorities and/or the sponsor and Regulatory Compliance persons. The subject should receive a copy of the signed and dated written informed consent form and any other written information provided to the subjects, and should receive copies of any signed and dated consent form updates and any amendments to the written information provided to subjects.

***Exclusion of Women, Minorities, and Children (Special Populations)***
Children are excluded for safety reasons.

***Subject Confidentiality***

Subject confidentiality is held strictly in trust by the participating investigators, their staff, and the sponsor and their agents. This confidentiality is extended to cover testing of biological samples in addition to the clinical information relating to participating subjects.
The study protocol, documentation, data, and all other information generated will be held in strict confidence. No information concerning the study or the data will be released to any unauthorized third party without prior written approval from the sponsor.
The study monitor or other authorized representatives of the sponsor and FDA may inspect all documents and records required to be maintained by the Investigator, including, but not limited to, medical records (office, clinic, or hospital) and pharmacy records for the subjects in this study. The clinical study site will permit access to such records.

***Study Discontinuation***
The PI has the right to terminate this study or an individual site's participation at any time. Reasons for terminating the study may include, but are not limited to, the following:

- Incidence or severity of adverse events indicates a potential health hazard;
- Data recording is inaccurate or incomplete;
- Investigator does not adhere to the protocol or applicable regulatory guidelines in conducting the study.

***Future Use of Stored Specimens***
Any leftover blood and stool specimens will be stored and may be used for future research, under a future protocol, to learn more about fecal transplant in patients with rCDI. These specimens will be stored indefinitely at Mount Sinai Hospital after the study is completed for future testing. In the informed consent document, subjects will be given an opportunity to choose whether or not their de-identified barcoded specimens are stored for future use. For subjects who choose not to allow storage of their samples for future use, these samples will be destroyed at the end of the study. All proposed research projects will be subject to approval by an IRB prior to release of any specimens. No human genetic tests will be performed on specimens.

There are no benefits to subjects in the collection, storage and subsequent research use of specimens. Reports about future research done with subject's samples will NOT be kept in their health records, but subject's samples may be kept with the study records or in other secure areas. Subjects can decide if they want their samples to be used for future research or have their samples destroyed at the end of the study. A subject's decision can be changed at any time before the end of the study by notifying the study doctors or nurses in writing. However, if a subject consents to future use and some of their blood and stool has already been used for research purposes, the information from that research may still be used.

De-identified samples and associated meta-data may be shared with other investigators at other institutions for academic purposes or industry collaborators pending an approved protocol. Each sample will be encoded (labeled) only with a barcode and a unique tracking number to protect subject's confidentiality.

Research using stored specimens may be conducted by other institutions. Any specimens and data provided to the receiving-institution will be coded. Unequivocally, neither individual personal identifiers nor the key linking coded data to individuals will be released to the receiving-institution.

***Data handling and record keeping***
The investigator is responsible to ensure the accuracy, completeness, legibility, and timeliness of the data reported. All data collection forms should be completed in a neat, legible manner to ensure accurate interpretation of data. Black ink is required to ensure clarity of reproduced copies. When making changes or corrections, cross out the original entry with a single line, and initial and date the change. Do not erase, overwrite, or use correction fluid or tape on the original.

Copies of the electronic CRF (eCRF) will be provided for use as source documents and maintained for recording data for each subject enrolled in the study. Data reported in the eCRF derived from source documents should be consistent with the source documents or the discrepancies should be explained.

***Data Management Responsibilities***
All source documents and laboratory reports must be reviewed by the clinical team and data entry staff, who will ensure that they are accurate and complete. Adverse Events must be graded, assessed for severity and causality, and reviewed by the site Principal Investigator or designee. Data collection is the responsibility of the trial staff at the site under the supervision of the site Principal Investigator. During the study, the investigator must maintain complete and accurate documentation for the study.

***Data Capture Methods***
Clinical data (including AEs, concomitant medications, and solicited events data) and clinical laboratory data will be entered into a compliant Internet Data Entry System. The data system includes password protection and internal quality checks, such as automatic range checks, to identify data that appear inconsistent, incomplete, or inaccurate. Clinical data will be entered directly from the source documents.

***Types of Data***
Data for this study will include clinical, safety and microbiological outcome measures.

***Timing/Reports***
Interim reports for the DSMB will be prepared when approximately 40% and 70% of subjects complete enrollment. Interim statistical reports may be generated as deemed necessary and appropriate by the study PI. Other safety summary reports may be generated for the DSMB. A final report will be prepared following the availability of all the clinical, safety and efficacy data.

***Study Records Retention***
Study files (except for future use consent forms) must be maintained for a minimum of two years after the last approval. These documents should be retained for a longer period, however, if required by local regulations. No records will be destroyed without the written consent of the sponsor, if applicable. It is the responsibility of the sponsor to inform the investigator when these documents no longer need to be retained. Consent forms for future use will be maintained as long as the sample exists.

***Protocol Deviations***
A protocol deviation is any noncompliance with the clinical trial protocol, GCP, or protocol-specific MOP requirements. The noncompliance may be either on the part of the subject, the investigator, or the study site staff. As a result of deviations, corrective actions are to be developed by the site and implemented promptly.

It is the responsibility of the study PI/study staff to use continuous vigilance to identify and report deviations within five working days of identification of the protocol deviation, or within five working days of the scheduled protocol-required activity. All deviations must be promptly reported to the study PI or designated personnel at MSH.

All protocol deviations, as defined above, must be addressed in study subject source documents. A completed copy of the Protocol Deviation Form must be maintained in the Regulatory File, as well as in the subject's source document. Protocol deviations must be sent to the local IRB per their guidelines. The study PI/study staff is responsible for knowing and adhering to their IRB requirements.

**7.15 Potential Risks and Benefits**

***7.15.1 Risks***

***7.15.1.1 MTC01 and FMT***
Some of the known risk associated with FMT are still possible with MTC01 such as:
- Altered bowel pattern (diarrhea, constipation)
- Cramping
- Belching
- Alteration in intestinal metabolism and potential drug-drug interactions.
- Transmission of allergens

Potential risks of MTC01 dosage:
- Inclusion of strains with previously undescribed harmful virulence factors

Potential risk of FMT dosage:
- Transmission of pathogenic bacteria, viruses, fungi, transmission of allergens

***7.15.1.2 Privacy and Confidentiality***
This study involves the collection of personal health information. Accidental release of personal health information is a risk of participation in this study. Measures will be taken to protect the confidentiality of all subjects' information. These measures include keeping all information collected about the subjects' confidential, keeping information in locked rooms, and having physicians who are directly involved with a subject's clinical care involved in the study.

***7.15.1.3 Colonoscopy***
Standard potential risks of the endoscopy procedure include discomfort, gastrointestinal bleeding either related or unrelated to biopsies, intestinal perforation, altered bowel habit. Complications of IV conscious sedation during the procedure include respiratory arrest, medication reactions, and aspiration.

***7.15.1.4 Venipuncture:***
Risks of having blood drawn include pain, bruising, or infection.

***7.15.1.5 Pregnancy***
The risks to fetuses and women who are pregnant are unknown. We will not be enrolling any pregnant of lactating women.

***7.15.2 Potential Benefits***
The potential benefits include:
- Treatment of recurrent CDI
- Improvement in clinical symptoms
- Restoration of fecal diversity

As this is a pilot study, it is difficult to quantify the expected benefits.

#### References

1. McDonald, L. C. *et al.* Clinical Practice Guidelines for Clostridium difficile Infection in Adults and Children: 2017 Update by the Infectious Diseases Society of America (IDSA) and Society for Healthcare Epidemiology of America (SHEA). *Clinical Infectious Diseases* **66**, e1–e48 (2018).
2. Aggarwala, V. *et al.* Precise quantification of bacterial strains after fecal microbiota transplantation delineates long-term engraftment and explains outcomes. *Nat Microbiol* **6**, 1309–1318 (2021).

### INFORMED CONSENT FORM(S)

[21 CFR 312.23(a)(6)]

#### TITLE OF RESEARCH STUDY:

Title: Phase 1b comparison of defined bacterial consortium MTC01 and Fecal Microbiota Transplantation for the treatment of recurrent *Clostridioides difficile* infection.

#### PRINCIPAL INVESTIGATOR (HEAD RESEARCHER) NAME AND CONTACT INFORMATION:

Name: Ari Grinspan

Physical Address:

The Susan and Leonard Feinstein Inflammatory Bowel Disease Clinical Center at Mount Sinai (IBD Center)

17 East 102nd Street, NY, NY 10029

5th Floor

Mailing Address: 1 Gustave L Levy Place, Box 1069, NY, NY 10029

#### WHAT IS A RESEARCH STUDY?

A research study is when scientists try to answer a question about something that we don't know enough about. Participating may not help you or others.

People volunteer to be in a research study. The decision about whether or not to take part is totally up to you. You can also agree to take part now and later change your mind. Whatever you decide is okay. It will not affect your ability to get medical care within the Mount Sinai Health System.

Someone will explain this research study to you. Feel free to ask all the questions you want before you decide. Any new information that develops during this research study that might make you change your mind about participating will be given to you promptly.

Basic information about this study will appear on the website http://www.ClinicalTrials.gov. There are a few reasons for this: the National Institutes of Health (NIH) encourages all researchers to post their research; some medical journals only accept articles if the research was posted on the website; and, for research studies the U.S. Food and Drug Administration (FDA) calls "applicable clinical trials" a description of this clinical trial will be available on http://www.ClinicalTrials.gov, as required by U.S. Law. This Web site will not include information that can identify you. At most, the Web site will include a summary of the results. You can search this Web site at any time.

#### PURPOSE OF THIS RESEARCH STUDY:

The purpose of this study is to compare two different treatments for patients with recurrent *Clostridiodies difficile* infections: MTC01 vs fecal microbiota transplantation (FMT). FMT is the transfer of bacteria from a healthy donor's colon to a recipient's colon. To do this, stool from a healthy donor is blended with salt water and made into a liquid solution rich in bacteria. This solution is sprayed into your colon during a colonoscopy. This treatment is now considered standard medical care for recurrent *Clostridioides difficile* (i.e., *C diff*) infections.

One FMT dose contains the entire collection of microbes in a healthy donor and is made up of billions of microbes. Each dose of FMT is different from the next and it is unknown exactly what microbes are present in each dose. A live biotherapeutic product (LBP) is a defined collection of microbes. LBPs could be potentially safer and easier to administer to patients than using an entire collection of microbes. We previously looked at patients who had a successful FMT treatment for their *C diff* infection and found that there are a small number of microbes from the FMT donor that remain in the recipient's colon for years after their treatment. We believe that this collection of donor microbes that colonize with FMT are key to the therapy's success. We have created a new LBP called MTC01 using these small number of microbes.

We will be randomizing patients who have recurrent *C diff* infection and are recommended to undergo FMT to receive either a standard FMT or MTC01. Both treatments will be administered during a colonoscopy, which is the standard way to give this treatment.

This is a pilot study. Pilot studies are done on a small group of subjects to learn if a larger study would be useful.

You may qualify to take part in this research study because you have already agreed to have a fecal transplant for your CDI.

Funds for conducting this research are provided by NIH (National Institute of Health)

#### LENGTH OF TIME AND NUMBER OF PEOPLE EXPECTED TO PARTICIPATE

Your participation in this research study is expected to last 26 weeks

The number of people expected to take part in this research study is 60. Subjects will be enrolled at the Mount Sinai Hospital.

#### DESCRIPTION OF WHAT'S INVOLVED:

If you agree to participate in this research study, the following information describes what may be involved.

**Screening (Visit 1)**

The Screening Visit will take about 45 minutes. During this visit, we will do some tests and procedures to see if you qualify to take part in this research study. The study doctor will review the results of these tests and procedures. If you do not qualify, the study doctor will explain why you do not fit the requirements for the study. This visit will take place at the IBD center.

At this visit, we will:

- Do a physical exam
- Review your medical history. We may review your medical record to do this.
- We will draw a total of about 5 tablespoons of blood during the entire course of the research study. Your blood will be tested for HIV, Hepatitis B and C as well as Syphilis. Any positive test results are required to be reported to the public health authorities.
- Ask you for a stool sample to look specifically at what bacteria are in your stool as well as their byproducts.
- Test your urine for pregnancy, if you are a female able to become pregnant. Pregnant women can't take part in this research study.
- Do a Symptoms Assessment by asking questions about current GI symptoms.

**Study Visit 2: Receive FMT vs LBP during colonoscopy**

Visit 2 will take about 4 hours (including procedure and recovery time). This will visit will take place the Mount Sinai Hospital Endoscopy Suite, located at 1184 $5^{th}$ Avenue, $7^{th}$ floor, NY, NY. At this visit, we will:

- Perform a urine pregnancy test if you are a female
- Give you questionnaires for you to complete about your disease activity and quality of life
- Give you a sedative medication and do a colonoscopy. During the colonoscopy, you will receive the microbiota transplant. Because you will be receiving a sedative, you will have to be escorted home. This is being done as part of your standard medical care
- Give you a symptom diary. You will use this diary to record any unpleasant things you experience during the study. Please bring this diary with you to each study visit so we can track your progress.

**Study Visit 3 (Week 1), Study Visit 4 (Week 8), Study Visit 5 (Week 12)**

These visits will each take about 15 minutes. These visits will each take place at the IBD center. At this visit, we will:

- Do a physical exam
- Review your medical history. We may review your medical record to do this.
- We will draw a total of about 1 tablespoons of blood at each visit.

- Ask you for a stool sample to look specifically at what bacteria are in your stool as well as their byproducts.
- Do a Symptoms Assessment by asking questions about current GI symptoms.
- Do a Safety Assessment by reviewing any side effects you have experienced, illnesses or hospitalizations.

**Telephone Call: Week 26**

This telephone call will take about 5 minutes. During this phone call we will:

- Do a Safety Assessment by reviewing any side effects you have experienced, illnesses or hospitalizations.
- Ask you to mail in a stool sample to look specifically at what bacteria are in your stool as well as their byproducts.

**After You Complete the Study**

After you complete the study, we will refer you to your doctor for your ongoing medical care

**For Women**

Since you are participating in a study that involves drugs or investigational treatment with potential risks to a developing fetus, it is recommended, for your protection, that you not become pregnant for the duration of the study. Also, you should not participate if you are breastfeeding. Therefore, practicing effective contraception is important. No individual contraceptive is 100% effective.

Recommended methods of birth control are:

- The consistent use of an approved hormonal contraception (birth control pill/patches, rings),
- An intrauterine device (IUD),
- Contraceptive injection (Depo-Provera),
- Double barrier methods (Diaphragm with spermicidal gel or condoms with contraceptive foam),
- Sexual abstinence (no sexual intercourse) or
- Sterilization.

Hormonal contraceptives, implants, and injections are only considered effective if used properly and started at least one month before you begin the study, continuing throughout the study and for one month after the end of the study. You should ask your study doctor if you should continue birth control for longer than 30 days after the end of the study. If you are unsure whether the method of birth control you use is acceptable to use while participating in this study, you should ask your study doctor before you begin the study. If you are less than one year post-menopausal, there is the potential that you could become pregnant. If you or your partner becomes pregnant or thinks either of you may be pregnant at any time during the trial, it is important that you tell your study doctor immediately. The trial drug may be stopped and a referral will be made to an obstetrician/gynecologist for follow-up. If you plan to become pregnant in the year following a clinical trial, speak with your study doctor.

If you have any questions about birth control, your study coordinator or study doctor will be able to answer your questions and give you advice.

Should you become pregnant, regardless of the outcome, the sponsor may ask for information on your pregnancy, even if you are withdrawn from the study. Your written consent will be obtained separately in the case that this happens.

The researchers would like to ask your permission to keep specimens (like blood, tissue, hair, or any other body matter) collected from you during this study to use them in future research studies. They would also like to know your wishes about how they might use your specimens in future research studies. You should also know that it is possible that products may someday be developed with the help of your specimens, and there are no plans to share any profits from such products with you.

(1) Will you allow the researchers to store your specimens to use in future research studies?

Yes________ No_________ If no, please stop here. If yes, please continue to the next question.

(2) The researchers can keep your specimens stored in one of two different ways: one way will store your specimens in a way that it is linked to your identity (through the use of a code that can indicate the information came from you personally) and the other way will store your specimens anonymously (no one will know who the information is from). It will not be stored both ways, so you must choose one of these two options. Please note that if you choose to have your specimens stored anonymously, you will not be able to change your mind to ask for your specimens to be destroyed at a future date.

How would you like your specimens stored? Please initial ONE choice:

I would like my specimens stored with a link to my identity___________

I would like my specimens stored anonymously _____________

(3) Do you give the researchers permission to contact you in the future to collect additional information about you, discuss how your specimens might be used, or to discuss possible participation in another research project? Please initial your choice:

Yes_________ No__________

(4) Do you give the researchers permission to keep the specimens indefinitely and use them for future studies that are directly related to the purpose of the current study? Please initial your choice:

Yes_________ No__________

(5) Do you give the researchers permission to keep the specimens indefinitely and use them for future studies that are not related to the purpose of the current study (for example, a different area of research)? Please initial your choice:

Yes_________ No__________

(a) If the future research in a different area can be done without having to know that the specimens came from you personally, that will be done.

(b) If the future research in a different area requires that it is known specifically who the specimens came from, then one of the following will be done:

(i) If you allowed the researchers to contact you in the future, they will be able to contact you to explain why your specimen is needed and what will be done with it. Your permission will be asked to use your specimens in that research project.

(ii) If you do not give permission to be contacted in the future, or if it is found that contacting you is not practical, for example, because you have moved, your specimens may still be used. Either all links to your identity will be removed from the specimens, or an Institutional Review Board will be asked for permission to use the specimens linked to your identity. The Institutional Review Board (IRB) is a committee of doctors and scientists and non-scientists and people not associated with this hospital or medical school whose job it is to protect people who participate in research. The IRB can give permission for researchers to use and share health information connected to specimens that are linked to people's identities, but only if it determines that doing this will not be more than a minimal risk to people or their privacy.

(6) Do you give permission to have portions of the specimens given to other researchers at Mount Sinai or other institutions for use in research that is either related or not related to the purpose of this study? Please initial your choice:

Yes_________ No__________

<u>If you change your mind later</u> and want your samples destroyed, contact the study doctor.

#### YOUR RESPONSIBILITIES IF YOU TAKE PART IN THIS RESEARCH:

If you decide to take part in this research study you will be responsible for the following things:

It is your responsibility to complete all the study visits and procedures and follow instructions given to you by the study doctor or the study team.

It is your responsibility to update the study team with any new medications or treatments prescribed by your physicians and any new signs or symptoms that develop during the study protocol.

#### COSTS OR PAYMENTS THAT MAY RESULT FROM PARTICIPATION:

If you agree to take part in this research study, we will pay you *** for each visit you complete. Payment will occur at the end of your participation in the study. Payment will be in the form of a check for a maximum of *** if you complete each visit. Checks require some time to be prepared and will be given to you as available.

Tax law may require the Mount Sinai Finance Department to report the amount of payment you receive from Mount Sinai to the Internal Revenue Service (IRS) or other agencies, as applicable. Generally this reporting would take place if you receive payments that equal $600 or more from Mount Sinai in a calendar year. You would be responsible for the payment of any tax that may be due.

#### POSSIBLE BENEFITS:

It is important to know that you may not get any benefit from taking part in this research. Others with CDI may benefit in the future from what we learn in this study.

#### REASONABLY FORESEEABLE RISKS AND DISCOMFORTS:

**Risks of Fecal Microbiota Transplant and MTC01**

You will be getting stool bacteria from a healthy person who has been evaluated carefully by physicians at Mount Sinai Hospital for good health and absence of any intestinal infections. There is a risk of transmission of infection by FMT or transmission of antibiotic-resistant bacteria. We screen stool donors by asking about health and COVID exposures. Donors are tested for the virus during their donation period. No screening tests are perfect, and it is possible that some problems may not be detected by the screening tests. We also screen stool donors for monkeypox by asking about health and monkeypox exposures. Monkeypox may be transmitted via FMT, although the risk of transmission is unknown. There are possible risks of transferring one person’s stool bacteria to another that are not related to infection. These include the possibility that inflammatory, allergic or metabolic conditions or weight/obesity could be changed by a specific combination of stool bacteria.

**Risks of Blood Draws**

You may have a bruise (a black and blue mark) or pain where we take the blood samples. The risks of a blood draw include pain, bruising, and the slight possibility of infection at the place where the needle goes in. Some people feel dizzy or may faint during or after a blood draw. As part of this research study, we will test your blood for HIV, Hepatitis B and C as well as Syphilis. For most studies, this means that results will become part of your hospital medical record. If these tests show that you have HIV, Hepatitis B and C, or Syphilis, you cannot be in the study. We will refer you for medical care if we find you have these infections. By law, healthcare providers must report positive test results for infectious diseases to public health authorities. These reports are required to identify you by name..

**Risks of Colonoscopy**

The current standard of care way to deliver an FMT is through a colonoscopy. We will be performing a colonoscopy to give you the study drugs: MTC01 or FMT. There is a risk of infection, bleeding, and/or perforation of the colon with colonoscopy. The overall risk of infection, bleeding and/or perforation is less than 1%.

**Privacy Risks**

This study involves the collection of personal health information. Accidental release of your personal health information is a risk of taking part in this study. To keep your study information private, every effort will be made to keep your information confidential. We will store your information in locked rooms and in secure electronic files. As explained in this consent form in the section, “Sending study information to Research Collaborators outside Partners,” your personal information will be removed from study information and samples, and study codes will be used. Only persons involved in this study will be able to see your study information.

#### OTHER POSSIBLE OPTIONS TO CONSIDER:

You may decide not to take part in this research study without any penalty. The choice is totally up to you. Instead of being in this research study, your choices may include:

1) You can still receive an FMT for your recurrent CDI even if you don't want to participate in this study.

#### IN CASE OF INJURY DURING THIS RESEARCH STUDY:

If you are injured or made sick from taking part in this research study, medical care will be provided. Generally, this care will be billed to you or your insurance in the ordinary manner and you will be responsible for all treatment costs not covered by your insurance, including deductibles, co-payments and coinsurance. This does not prevent you from seeking payment for injury related to malpractice or negligence. Contact the investigator for more information.

#### ENDING PARTICIPATION IN THE RESEARCH STUDY:

You may stop taking part in this research study at any time without any penalty. This will not affect your ability to receive medical care at any of the Mount Sinai Health System hospitals or to receive any benefits to which you are otherwise entitled.

If you decide to stop being in the research study, please contact the Principal Investigator or the research staff.

If you stop being in the research study, already collected information may not be removed from the research study database and will continue to be used to complete the research analysis. You may be asked whether the investigator can collect information from your routine medical care. If you agree, this data will be handled the same as research data

You may also withdraw your permission for the use and disclosure of any of your protected information for research, but you must do so in writing to the Principal Investigator at the address on the first page. Even if you withdraw your permission, the Principal Investigator for the research study may still use the information that was already collected if that information is necessary to complete the research study. Your health information may still be used or shared after you withdraw your authorization if you have an adverse event (a bad effect) from participating in the research study.

Withdrawal without your consent: The study doctor, the sponsor or the institution may stop your involvement in this research study at any time without your consent. This may be because the research study is being stopped, the instructions of the study team have not been followed, the investigator believes it is in your best interest, or for any other reason. If specimens or data have been stored as part of the research study, they too can be destroyed without your consent. More possible reasons for removal from the study include:

- The colonoscopy cannot be completed to deliver the fecal transplant
- The study doctor thinks it is best for you to not receive the fecal transplant
- You can't complete study requirements
- The Sponsor decides to stop the study
- We stop doing the study for other reasons

If you have a colonoscopy but do not receive the fecal transplant, we will follow-up with you for 2 weeks to make sure you do not develop any health problems from the procedure.

#### CONTACT PERSON(S):

If you have any questions, concerns, or complaints at any time about this research, or you think the research has hurt you, please contact the office of the research team and/or the Principal Investigator at phone number 212-241-7757

If you experience an emergency during your participation in this research, contact Dr. Grinspan and call 911 or go to the emergency room.

This research has been reviewed and approved by an Institutional Review Board. You may reach a representative of the Program for Protection of Human Subjects at the Icahn School of Medicine at Mount Sinai at telephone number (212) 824-8200 during standard work hours for any of the reasons listed below. This office will direct your call to the right person within the Mount Sinai Health System:

- Your questions, concerns, or complaints are not being answered by the research team.
- You cannot reach the research team.
- You are not comfortable talking to the research team.

- You have questions about your rights as a research subject.
- You want to get information or provide input about this research.

#### DISCLOSURE OF FINANCIAL INTERESTS:

Sometimes, physicians/researchers receive payments for consulting or similar work performed for industry. Effective September 2014 Mount Sinai reviews only payments to an individual totaling more than $5,000 a year per entity when determining potential conflicts of interest. If you have questions regarding industry relationships, we encourage you to talk your physician/researcher or visit our website at http://icahn.mssm.edu/ where Mount Sinai publicly discloses the industry relationships of our faculty.

#### MAINTAINING CONFIDENTIALITY – HIPAA AUTHORIZATION:

As you take part in this research project it will be necessary for the research team and others to use and share some of your private protected health information. Consistent with the federal Health Insurance Portability and Accountability Act (HIPAA), we are asking your permission to receive, use and share that information.

<u>What protected health information is collected and used in this study, and might also be disclosed (shared) with others?</u>

As part of this research project, the research team at the hospital(s) involved in the research will collect your name, address, telephone/fax numbers, date of birth, medical records number.

The researchers will also get information from your medical record from Mount Sinai Hospital or your personal doctor.

During the study the researchers will gather information by:

- completing the tests, procedures, questionnaires and interviews explained in the description section of this consent.
- reviewing HIV-related information, which includes any information indicating that you have had an HIV related test, or have HIV infection, HIV related illness or AIDS, or any information which could indicate that you have been potentially exposed to HIV

<u>Why is your protected health information being used?</u>

Your personal contact information is important to be able to contact you during the study. Your health information and the results of any tests and procedures being collected as part of this research study will be used for the purpose of this study as explained earlier in this consent form. The results of this study could be published or presented at scientific meetings, lectures, or other events, but would not include any information that would let others know who you are, unless you give separate permission to do so.

The Principal Investigator may also use and share the results of these tests and procedures to treat you in collaboration with others in the Mount Sinai Health System.

The research team and other authorized members of The Mount Sinai Health System ("Mount Sinai") workforce may use and share your information to ensure that the research meets legal, institutional or accreditation requirements. For example, the School's Program for the Protection of Human Subjects is responsible for overseeing research on human subjects, and may need to see your information. If you receive any payments for taking part in this study, the Mount Sinai Finance Department may need your name, address, social security number, payment amount, and related information for tax reporting purposes. If the research team uncovers abuse, neglect, or reportable diseases, this information may be disclosed to appropriate authorities.

Who, outside Mount Sinai, might receive your protected health information?

As part of the study, the Principal Investigator, study team and others in the Mount Sinai workforce may disclose your protected health information, including the results of the research study tests and procedures, to the following people or organizations: (It is possible that there may be changes to the list during this research study; you may request an up-to-date list at any time by contacting the Principal Investigator.)

- Other collaborating research center(s) and their associated research/clinical staff who are working with the investigators on this project: Brigham and Women's Hospital
- Outside laboratory who will be performing laboratory analysis for all the research centers involved in this project: Brigham and Women's Hospital
- The commercial sponsor and/or their representative (who will use the results for submissions to the Food and Drug Administration): Brigham and Women's Hospital
- A Data Safety Monitoring Board or other committee that will monitor the study on an ongoing basis for safety.
- The United States Food and Drug Administration
- The United States Department of Health and Human Services and the Office of Human Research Protection.

In almost all disclosures outside of Mount Sinai, you will not be identified by name, social security number, address, telephone number, or any other direct personal identifier. Some records and information disclosed may be identified with a unique code number. The Principal Investigator will ensure that the key to the code will be kept in a locked file, or will be securely stored electronically. The code will not be used to link the information back to you without your permission, unless the Institutional Review Board allows it after determining that there would be minimal risk to your privacy. The Certificate of Confidentiality obtained from the Department of Health and Human Services will not be used to prevent disclosure to local authorities of child abuse and neglect, or harm to self or others. It is possible that a sponsor or their representatives, a data coordinating office, or a contract research organization, will come to inspect your records. Even if those records are identifiable when inspected, the information leaving the institution will be stripped of direct identifiers. Additionally, the Office of Human Subjects Protection (OHRP) of the Department of Health and Human Services as well as the Food and Drug

Administration (FDA) will be granted direct access to your medical records for verification of the research procedures and data. They are authorized to remove information with identifiers if necessary to complete their task. By signing this document you are authorizing this access. We may publish the results of this research. However, we will keep your name and other identifying information confidential.

For how long will Mount Sinai be able to use or disclose your protected health information?

Your authorization for use of your protected health information for this specific study does not expire.

Will you be able to access your records?

During your participation in this study, you will have access to your medical record and any study information that is part of that record. The investigator is not required to release to you research information that is not part of your medical record.

Do you need to give us permission to obtain, use or share your health information?

NO! If you decide not to let us obtain, use or share your health information you should not sign this form, and you will not be allowed to volunteer in the research study. If you do not sign, it will not affect your treatment, payment or enrollment in any health plans or affect your eligibility for benefits.

Can you change your mind?

You may withdraw your permission for the use and disclosure of any of your protected information for research, but you must do so in writing to the Principal Investigator at the address on the first page. Even if you withdraw your permission, the Principal Investigator for the research study may still use your protected information that was already collected if that information is necessary to complete the study. Your health information may still be used or shared after you withdraw your authorization if you should have an adverse event (a bad effect) from being in the study. If you withdraw your permission to use your protected health information for research that means you will also be withdrawn from the research study, but standard medical care and any other benefits to which you are entitled will not be affected. You can also tell us you want to withdraw from the research study at any time without canceling the Authorization to use your data.

It is important for you to understand that once information is disclosed to others outside Mount Sinai, the information may be re-disclosed and will no longer be covered by the federal privacy protection regulations. However, even if your information will no longer be protected by federal regulations, where possible, Mount Sinai has entered into agreements with those who will receive your information to continue to protect your confidentiality.

If as part of this research project your medical records are being reviewed, or a medical history is being taken, it is possible that HIV-related information may be revealed to the researchers. If that is the case, the following information concerns you. If this research does not involve any review of medical records or questions about your medical history or conditions, then the following section may be ignored.

---

**Notice Concerning HIV-Related Information**

If you are authorizing the release of HIV-related information, you should be aware that the recipient(s) is (are) prohibited from re-disclosing any HIV-related information without your authorization unless permitted to do so under federal or state law. You also have a right to request a list of people who may receive or use your HIV-related information without authorization. If you experience discrimination because of the release or disclosure of HIV-related information, you may contact the New York State Division of Human Rights at (888) 392-3644 or the New York City Commission on Human Rights at (212) 306-5070. These agencies are responsible for protecting your rights.

---

**Signature Block for Capable Adult**

Your signature below documents your permission to take part in this research and to the use and disclosure of your protected health information. A signed and dated copy will be given to you.

| | |
|---|---|
| Signature of subject | Date |
| Printed name of subject | Time |

**Person Explaining Study and Obtaining Consent**

| | |
|---|---|
| Signature of person obtaining consent | Date |
| Printed name of person obtaining consent | Time |

***Witness Section: For use when a witness is required to observe the consent process,, document below (for example, subject is illiterate or visually impaired, or this accompanies a short form consent):***

*My signature below documents that the information in the consent document and any other written information was accurately explained to, and apparently understood by, the subject, and that consent was freely given by the subject.*

| | |
|---|---|
| *Signature of witness to consent process* | *Date* |
| *Printed name of person witnessing consent process* | Time |

#### TITLE OF RESEARCH STUDY:

Title: Phase 1b comparison of defined bacterial consortium MTC01 and Fecal Microbiota Transplantation for the treatment of recurrent *Clostridioides difficile* infection.

#### PRINCIPAL INVESTIGATOR (HEAD RESEARCHER) NAME AND CONTACT INFORMATION:

Name: Ari Grinspan

Physical Address:

The Susan and Leonard Feinstein Inflammatory Bowel Disease Clinical Center at Mount Sinai (IBD Center)

17 East 102nd Street, NY, NY 10029

5th Floor

Mailing Address: 1 Gustave L Levy Place, Box 1069, NY, NY 10029

#### WHAT IS A RESEARCH STUDY?

A research study is when scientists try to answer a question about something that we don't know enough about. Participating may not help you or others.

People volunteer to be in a research study. The decision about whether or not to take part is totally up to you. You can also agree to take part now and later change your mind. Whatever you decide is okay. It will not affect your ability to get medical care within the Mount Sinai Health System.

Someone will explain this research study to you. Feel free to ask all the questions you want before you decide. Any new information that develops during this research study that might make you change your mind about participating will be given to you promptly.

Basic information about this study will appear on the website http://www.ClinicalTrials.gov. There are a few reasons for this: the National Institutes of Health (NIH) encourages all researchers to post their research; some medical journals only accept articles if the research was posted on the website; and, for research studies the U.S. Food and Drug Administration (FDA) calls "applicable clinical trials" a description of this clinical trial will be available on http://www.ClinicalTrials.gov, as required by U.S. Law. This Web site will not include information that can identify you. At most, the Web site will include a summary of the results. You can search this Web site at any time.

#### PURPOSE OF THIS RESEARCH STUDY:

The purpose of this study is to compare two different treatments for patients with recurrent *Clostridiodies difficile* infections: MTC01 vs fecal microbiota transplantation (FMT). FMT is the transfer of bacteria from a healthy donor's colon to a recipient's colon. To do this, stool from a healthy donor is blended with salt water and made into a liquid solution rich in bacteria. This solution is sprayed into your colon during a colonoscopy. This treatment is now considered standard medical care for recurrent *Clostridioides difficile* (i.e., *C diff*) infections.

One FMT dose contains the entire collection of microbes in a healthy donor and is made up of billions of microbes. Each dose of FMT is different from the next and it is unknown exactly what microbes are present in each dose. A live biotherapeutic product (LBP) is a defined collection of microbes. LBPs could be potentially safer and easier to administer to patients than using an entire collection of microbes. We previously looked at patients who had a successful FMT treatment for their *C diff* infection and found that there are a small number of microbes from the FMT donor that remain in the recipient's colon for years after their treatment. We believe that this collection of donor microbes that colonize with FMT are key to the therapy's success. We have created a new LBP called MTC01 using these small number of microbes.

We will be randomizing patients who have recurrent *C diff* infection and are recommended to undergo FMT to receive either a standard FMT or MTC01. Both treatments will be administered during a colonoscopy, which is the standard way to give this treatment.

This is a pilot study. Pilot studies are done on a small group of subjects to learn if a larger study would be useful.

You are being consented as a stool donor for this pilot.

Funds for conducting this research are provided by NIH (National Institute of Health)

#### LENGTH OF TIME AND NUMBER OF PEOPLE EXPECTED TO PARTICIPATE

Your participation in this research study is expected to last 1 week.

The number of people expected to take part in this research study is 60. Subjects will be enrolled at the Mount Sinai Hospital.

#### DESCRIPTION OF WHAT'S INVOLVED:

If you agree to participate in this research study, the following information describes what may be involved.

- History and physical exam by study doctor
- Completion of screening questionnaires
- Blood tests at two time points
- Stool tests at two time points
- Stool donation (minimum of 5 donations) within a 30-day donation period

- COVID-19 nasopharyngeal swabs at two time points

The researchers would like to ask your permission to keep specimens (like blood, tissue, hair, or any other body matter) collected from you during this study to use them in future research studies. They would also like to know your wishes about how they might use your specimens in future research studies. You should also know that it is possible that products may someday be developed with the help of your specimens, and there are no plans to share any profits from such products with you.

(1) Will you allow the researchers to store your specimens to use in future research studies?

Yes________ No_________ If no, please stop here. If yes, please continue to the next question.

(2) The researchers can keep your specimens stored in one of two different ways: one way will store your specimens in a way that it is linked to your identity (through the use of a code that can indicate the information came from you personally) and the other way will store your specimens anonymously (no one will know who the information is from). It will not be stored both ways, so you must choose one of these two options. Please note that if you choose to have your specimens stored anonymously, you will not be able to change your mind to ask for your specimens to be destroyed at a future date.

How would you like your specimens stored? Please initial ONE choice:

I would like my specimens stored with a link to my identity___________

I would like my specimens stored anonymously _____________

(3) Do you give the researchers permission to contact you in the future to collect additional information about you, discuss how your specimens might be used, or to discuss possible participation in another research project? Please initial your choice:

Yes_________ No__________

(4) Do you give the researchers permission to keep the specimens indefinitely and use them for future studies that are directly related to the purpose of the current study? Please initial your choice:

Yes_________ No__________

(5) Do you give the researchers permission to keep the specimens indefinitely and use them for future studies that are not related to the purpose of the current study (for example, a different area of research)? Please initial your choice:

Yes_________ No__________

(a) If the future research in a different area can be done without having to know that the specimens came from you personally, that will be done.

(b) If the future research in a different area requires that it is known specifically who the specimens came from, then one of the following will be done:

(i) If you allowed the researchers to contact you in the future, they will be able to contact you to explain why your specimen is needed and what will be done with it. Your permission will be asked to use your specimens in that research project.

(ii) If you do not give permission to be contacted in the future, or if it is found that contacting you is not practical, for example, because you have moved, your specimens may still be used. Either all links to your identity will be removed from the specimens, or an Institutional Review Board will be asked for permission to use the specimens linked to your identity. The Institutional Review Board (IRB) is a committee of doctors and scientists and non-scientists and people not associated with this hospital or medical school whose job it is to protect people who participate in research. The IRB can give permission for researchers to use and share health information connected to specimens that are linked to people's identities, but only if it determines that doing this will not be more than a minimal risk to people or their privacy.

(6) Do you give permission to have portions of the specimens given to other researchers at Mount Sinai or other institutions for use in research that is either related or not related to the purpose of this study? Please initial your choice:

Yes_________ No__________

<u>If you change your mind later</u> and want your samples destroyed, contact the study doctor.

#### YOUR RESPONSIBILITIES IF YOU TAKE PART IN THIS RESEARCH:

If you decide to take part in this research study you will be responsible for the following things:

It is your responsibility to complete all the study visits and procedures and follow instructions given to you by the study doctor or the study team.

It is your responsibility to update the study team with any new medications or treatments prescribed by your physicians and any new signs or symptoms that develop during the study protocol.

#### COSTS OR PAYMENTS THAT MAY RESULT FROM PARTICIPATION:

You will be paid for participating in this research study. Being in this research study will not lead to extra costs to you. You will be reimbursed for your travel that may be required for study visits.

The samples collected from you as part of this research may be used for commercial profit. Regardless of whether your identifiable information is removed, there are no plans to share any profits with you.

#### POSSIBLE BENEFITS:

It is important to know that you will not get any benefit from taking part in this research. Others with recurrent C. difficile infection may benefit in the future from what we learn in this study.

#### REASONABLY FORESEEABLE RISKS AND DISCOMFORTS:

##### Risks of Blood Draws

You may have a bruise (a black and blue mark) or pain where we take the blood samples. The risks of a blood draw include pain, bruising, and the slight possibility of infection at the place where the needle goes in. Some people feel dizzy or may faint during or after a blood draw.

As part of this research study, we will test your blood for HIV, Hepatitis B and C as well as Syphilis. For most studies, this means that results will become part of your hospital medical record. If these tests show that you have HIV, Hepatitis B and C, or Syphilis, you cannot be in the study. We will refer you for medical care if we find you have these infections. By law, healthcare providers must report positive test results for infectious diseases to public health authorities. These reports are required to identify you by name.

##### Privacy Risks

This study involves the collection of personal health information. Accidental release of your personal health information is a risk of taking part in this study. To keep your study information private, every effort will be made to keep your information confidential. We will store your information in locked rooms and in secure electronic files. Only persons involved in this study will be able to see your study information.

#### OTHER POSSIBLE OPTIONS TO CONSIDER:

You may decide not to take part in this research study without any penalty. The choice is totally up to you.

#### IN CASE OF INJURY DURING THIS RESEARCH STUDY:

If you are injured or made sick from taking part in this research study, medical care will be provided. Generally, this care will be billed to you or your insurance in the ordinary manner and you will be responsible for all treatment costs not covered by your insurance, including deductibles, co-payments and coinsurance. This does not prevent you from seeking payment for injury related to malpractice or negligence. Contact the investigator for more information.

#### ENDING PARTICIPATION IN THE RESEARCH STUDY:

You may stop taking part in this research study at any time without any penalty. This will not affect your ability to receive medical care at any of the Mount Sinai Health System hospitals or to receive any benefits to which you are otherwise entitled.

If you decide to stop being in the research study, please contact the Principal Investigator or the research staff.

If you stop being in the research study, already collected information may not be removed from the research study database and will continue to be used to complete the research analysis. You may be asked whether the investigator can collect information from your routine medical care. If you agree, this data will be handled the same as research data

You may also withdraw your permission for the use and disclosure of any of your protected information for research, but you must do so in writing to the Principal Investigator at the address on the first page. Even if you withdraw your permission, the Principal Investigator for the research study may still use the information that was already collected if that information is necessary to complete the research study. Your health information may still be used or shared after you withdraw your authorization if you have an adverse event (a bad effect) from participating in the research study.

Withdrawal without your consent: The study doctor, the sponsor or the institution may stop your involvement in this research study at any time without your consent. This may be because the research study is being stopped, the instructions of the study team have not been followed, the investigator believes it is in your best interest, or for any other reason. If specimens or data have been stored as part of the research study, they too can be destroyed without your consent. More possible reasons for removal from the study include:

- You can’t complete study requirements
- The Sponsor decides to stop the study
- We stop doing the study for other reasons

#### CONTACT PERSON(S):

If you have any questions, concerns, or complaints at any time about this research, or you think the research has hurt you, please contact the office of the research team and/or the Principal Investigator at phone number 212-241-7757

If you experience an emergency during your participation in this research, contact Dr. Kayal and call 911 or go to the emergency room.

This research has been reviewed and approved by an Institutional Review Board. You may reach a representative of the Program for Protection of Human Subjects at the Icahn School of Medicine at Mount Sinai at telephone number (212) 824-8200 during standard work hours for any of the reasons listed below. This office will direct your call to the right person within the Mount Sinai Health System:

- Your questions, concerns, or complaints are not being answered by the research team.
- You cannot reach the research team.
- You are not comfortable talking to the research team.
- You have questions about your rights as a research subject.
- You want to get information or provide input about this research.

#### DISCLOSURE OF FINANCIAL INTERESTS:

Sometimes, physicians/researchers receive payments for consulting or similar work performed for industry. Effective September 2014 Mount Sinai reviews only payments to an individual totaling more than $5,000 a year per entity when determining potential conflicts of interest. If you have questions regarding industry relationships, we encourage you to talk your physician/researcher or visit our website at http://icahn.mssm.edu/ where Mount Sinai publicly discloses the industry relationships of our faculty.

#### MAINTAINING CONFIDENTIALITY – HIPAA AUTHORIZATION:

As you take part in this research project it will be necessary for the research team and others to use and share some of your private protected health information. Consistent with the federal Health Insurance Portability and Accountability Act (HIPAA), we are asking your permission to receive, use and share that information.

<u>What protected health information is collected and used in this study, and might also be disclosed (shared) with others?</u>

As part of this research project, the research team at the hospital(s) involved in the research will collect your name, address, telephone/fax numbers, date of birth, medical records number.

The researchers will also get information from your medical record from Mount Sinai Hospital or your personal doctor.

During the study the researchers will gather information by:

- completing the tests, procedures, questionnaires and interviews explained in the description section of this consent.

– reviewing HIV-related information, which includes any information indicating that you have had an HIV related test, or have HIV infection, HIV related illness or AIDS, or any information which could indicate that you have been potentially exposed to HIV

Why is your protected health information being used?

Your personal contact information is important to be able to contact you during the study. Your health information and the results of any tests and procedures being collected as part of this research study will be used for the purpose of this study as explained earlier in this consent form. The results of this study could be published or presented at scientific meetings, lectures, or other events, but would not include any information that would let others know who you are, unless you give separate permission to do so.

The Principal Investigator may also use and share the results of these tests and procedures to treat you in collaboration with others in the Mount Sinai Health System.

The research team and other authorized members of The Mount Sinai Health System ("Mount Sinai") workforce may use and share your information to ensure that the research meets legal, institutional or accreditation requirements. For example, the School's Program for the Protection of Human Subjects is responsible for overseeing research on human subjects, and may need to see your information. If you receive any payments for taking part in this study, the Mount Sinai Finance Department may need your name, address, social security number, payment amount, and related information for tax reporting purposes. If the research team uncovers abuse, neglect, or reportable diseases, this information may be disclosed to appropriate authorities.

Who, outside Mount Sinai, might receive your protected health information?

As part of the study, the Principal Investigator, study team and others in the Mount Sinai workforce may disclose your protected health information, including the results of the research study tests and procedures, to the following people or organizations: (It is possible that there may be changes to the list during this research study; you may request an up-to-date list at any time by contacting the Principal Investigator.)

- A Data Safety Monitoring Board or other committee that will monitor the study on an ongoing basis for safety.
- The United States Food and Drug Administration
- The United States Department of Health and Human Services and the Office of Human Research Protection.

In almost all disclosures outside of Mount Sinai, you will not be identified by name, social security number, address, telephone number, or any other direct personal identifier. Some records and information disclosed may be identified with a unique code number. The Principal Investigator will ensure that the key to the code will be kept in a locked file, or will be securely stored electronically. The code will not be used to link the information back to you without your permission, unless the Institutional Review Board allows it after determining that there would be minimal risk to your privacy. The Certificate of Confidentiality obtained from the Department of Health and Human Services will not be used to prevent disclosure to local authorities of child abuse and neglect, or harm to self or others.

It is possible that a sponsor or their representatives, a data coordinating office, or a contract research organization, will come to inspect your records. Even if those records are identifiable when inspected, the information leaving the institution will be stripped of direct identifiers. Additionally, the Office of Human Subjects Protection (OHRP) of the Department of Health and Human Services as well as the Food and Drug Administration (FDA) will be granted direct access to your medical records for verification of the research procedures and data. They are authorized to remove information with identifiers if necessary to complete their task. By signing this document you are authorizing this access. We may publish the results of this research. However, we will keep your name and other identifying information confidential.

For how long will Mount Sinai be able to use or disclose your protected health information?

Your authorization for use of your protected health information for this specific study does not expire.

Will you be able to access your records?

During your participation in this study, you will have access to your medical record and any study information that is part of that record. The investigator is not required to release to you research information that is not part of your medical record.

Do you need to give us permission to obtain, use or share your health information?

NO! If you decide not to let us obtain, use or share your health information you should not sign this form, and you will not be allowed to volunteer in the research study. If you do not sign, it will not affect your treatment, payment or enrollment in any health plans or affect your eligibility for benefits.

Can you change your mind?

You may withdraw your permission for the use and disclosure of any of your protected information for research, but you must do so in writing to the Principal Investigator at the address on the first page. Even if you withdraw your permission, the Principal Investigator for the research study may still use your protected information that was already collected if that information is necessary to complete the study. Your health information may still be used or shared after you withdraw your authorization if you should have an adverse event (a bad effect) from being in the study. If you withdraw your permission to use your protected health information for research that means you will also be withdrawn from the research study, but standard medical care and any other benefits to which you are entitled will not be affected. You can also tell us you want to withdraw from the research study at any time without canceling the Authorization to use your data.

It is important for you to understand that once information is disclosed to others outside Mount Sinai, the information may be re-disclosed and will no longer be covered by the federal privacy protection regulations. However, even if your information will no longer be protected by federal regulations, where possible, Mount Sinai has entered into agreements with those who will receive your information to continue to protect your confidentiality.

If as part of this research project your medical records are being reviewed, or a medical history is being taken, it is possible that HIV-related information may be revealed to the researchers. If that is the case, the following information concerns you. If this research does not involve any review of medical records or questions about your medical history or conditions, then the following section may be ignored.

---

**Notice Concerning HIV-Related Information**

If you are authorizing the release of HIV-related information, you should be aware that the recipient(s) is (are) prohibited from re-disclosing any HIV-related information without your authorization unless permitted to do so under federal or state law. You also have a right to request a list of people who may receive or use your HIV-related information without authorization. If you experience discrimination because of the release or disclosure of HIV-related information, you may contact the New York State Division of Human Rights at (888) 392-3644 or the New York City Commission on Human Rights at (212) 306-5070. These agencies are responsible for protecting your rights.

---

#### Signature Block for Capable Adult

Your signature below documents your permission to take part in this research and to the use and disclosure of your protected health information. A signed and dated copy will be given to you.

| | |
|---|---|
| Signature of subject | Date |
| Printed name of subject | Time |

#### Person Explaining Study and Obtaining Consent

| | |
|---|---|
| Signature of person obtaining consent | Date |
| Printed name of person obtaining consent | Time |

***Witness Section: For use when a witness is required to observe the consent process,, document below (for example, subject is illiterate or visually impaired, or this accompanies a short form consent):***

*My signature below documents that the information in the consent document and any other written information was accurately explained to, and apparently understood by, the subject, and that consent was freely given by the subject.*

| | |
|---|---|
| *Signature of witness to consent process* | *Date* |
| *Printed name of person witnessing consent process* | Time |

### CURRICULA VITAE OF ALL INVESTIGATORS

[21 CFR 312.23(a)(10)]

Ari Grinspan
1623 3rd Avenue Apt 35A
NY, NY 10128


**APPOINTMENTS**
2014-Present Associate Professor of Medicine, Icahn School of Medicine at Mount Sinai, NY, NY

**GAP IN EMPLOYMENT** N/A

**EDUCATION**
2000-2004 BA, University of Pennsylvania, Philadelphia, PA
2004-2008 MD, Albert Einstein College of Medicine, Bronx, NY
2008-2011 Residency in Internal Medicine, New York-Presbyterian Hospital, NY, NY
2011-2014 Fellowship in Gastroenterology, Mount Sinai Hospital, NY, NY

**CERTIFICATION**
2011-2021 Internal Medicine
2014-2024 Gastroenterology

**LICENSURE**
2011-Present New York State License #261448

**HONORS/AWARDS**
2004 Magna Cum Laude, University of Pennsylvania
2004 Phi Beta Kappa, University of Pennsylvania
2008 Distinction in Gastroenterology, Albert Einstein College of Medicine
2008 Alpha Omega Alpha, Albert Einstein College of Medicine
2011 Assistant Chief Resident, New York-Presbyterian Hospital
2011 Daniel M. Libby Program Director's Award, New York-Presbyterian Hospital
2013 Chief Fellow, Division of Gastroenterology, Mount Sinai Hospital
2016 Outstanding Outpatient Preceptor of the Year Award, Mount Sinai Hospital
2017 World Trade Center Appreciation Award
2018 Outstanding Teacher of the Year Award, Icahn School of Medicine at Mount Sinai GI Fellowship
2019 Solomon Silver Award in Clinical Medicine, Mount Sinai Hospital
2020 Outstanding Teacher of the Year Award, Icahn School of Medicine at Mount Sinai GI Fellowship
2021 Fellow, American College of Gastroenterology
2022 Fellow, American Gastroenterological Association

**PATENTS** N/A

**OTHER PROFESSIONAL ROLES**
Internal
2014-Present Member, Admissions Committee, Mount Sinai Hospital GI Fellowship program
2014-Present Course Director, Gastroenterology Grand Rounds
2017-Present Member, Antibiotic Stewardship Committee
2019-Present Physician Communication Ambassador
2019-Present Well-being Champion, Division of Gastroenterology

2019-Present Well-being Champion, Gastroenterology Fellowship

External

Professional Memberships

| | |
|---|---|
| 2011-Present | American Gastroenterological Association, Fellow |
| 2011-Present | American College of Gastroenterology, Fellow |

Professional Journals

| | |
|---|---|
| 2014 | Reviewer, Gastrointestinal Endoscopy |
| 2014 | Reviewer, ACG Case Reports Journal |
| 2014-Present | Reviewer, Journal of Gastroenterology and Hepatology |
| 2016-Present | Reviewer, American Journal of Gastroenterology |
| 2016-Present | Reviewer, Surgical Endoscopy |
| 2016-Present | Reviewer, Journal of Clinical Gastroenterology |
| 2016-Present | Reviewer, Inflammatory Bowel Disease |
| 2017-Present | Reviewer, Liver Transplantation |
| 2018-Present | Reviewer, BMJ Open |
| 2018-Present | Reviewer, BMC Gastroenterology |
| 2018-Present | Reviewer, Gastroenterology |
| 2019-Present | Reviewer, Digestive Disease Sciences |
| 2020-Present | Reviewer, Gut |

Professional Committees

2019-Present Patient Care Committee, American College of Gastroenterology (ACG)

**RESEARCH PROFILE**

My research focuses on the manipulation of the gut microbiome to treat human disease, namely *Clostridioides difficile* infection (CDI) and Inflammatory Bowel Disease (IBD). I have developed collaborations with colleagues across the US (Harvard Medical School, Warren Alpert Medical School of Brown University and Indiana University School of Medicine) and we have engaged in multiple collaborations exploring the use of FMT in CDI and IBD. Together, we received a CCF grant to prospectively study FMT in patients with CDI and IBD (ICON) which showed in a prospective study that FMT is an effective treatment for recurrent CDI in patients with IBD. We are now approved for additional round of funding to further study FMT in CDI and IBD (ICON 2). Partnering with Dr. Jean-Fred Colombel, Dr. Sergio Lira, Dr. Judy Cho, Dr. Jeremiah Faith, and Dr. Jose Clemente, we obtained a private donation to fund 2 successful research collaborations exploring Ulcerative Colitis (SUCCESS 1.0 and SUCCESS 2.0). We performed deep microbial analysis of patients with UC undergoing FMT to treat CDI. While multiple publications have come from this research, I first-authored a manuscript accepted to a top IBD journal where we describe novel findings of long-term efficacy of FMT for CDI in patients with IBD. Due to the tremendous success of these 2 grants, we recently obtained additional funding from a private donor to prospectively study the safety and efficacy of FMT in active UC (SUCCESS 3.0). I have obtained additional funding from a private donor to help support my research efforts in FMT (George Waechter Family Foundation). I am also active in clinical trials, serving as site PI for 2 national trials exploring the use of FMT in recurrent *Clostridioides difficile* infection as well as one of the top recruiters for the American Gastroenterology Association's Fecal Microbiota Transplantation National Registry. I am currently the principal investigator for a trial exploring the use of encapsulated FMT to treat patients with active UC. In collaboration with Dr. Jeremiah Faith and Dr. Maia Kayal, we have been awarded funding to explore the utility of FMT to treat chronic pouchitis.

**CLINICAL PROFILE**

As a clinician, I pioneered the Fecal Microbiota Transplantation (FMT) program at Mount Sinai Hospital for the treatment of recurrent and refractory *Clostridioides difficile* infection (CDI). There was a clear need in both the inpatient and outpatient setting for a more definitive treatment for CDI as rates of CDI and its complications have become an epidemic. FMT has proven to be an incredibly effective treatment with ~90% cure rate. As an active member of the Antibiotic Stewardship Committee, we have instituted a revised guideline for the treatment of *Clostridioides difficile* for the Mount Sinai Health System. I have built the FMT program into a national and international referral center. I have been referred patients from across the nation and internationally from Canada, Mexico, Panama, Switzerland, Israel, China and Japan. I have been recognized by the AGA as having a robust FMT practice and have been chosen as one of the first sites for its National FMT registry to track the long-term safety outcomes of patients undergoing FMT.

**IMPACT**

From a clinical perspective, I have built a very busy practice in IBD, general GI and recalcitrant *Clostridioides difficile* infection. Since establishing the FMT program at MSH in 2013, I have grown the FMT program into one of the largest in the country, performing over 50 FMTs per year. I am not only performing FMT on outpatients with recurrent CDI, but also in hospitalized patients with severe or fulminant CDI where we have shown a 77% mortality benefit over standard of care treatment. We recently presented these findings in a top tier gastroenterology journal.

As an educator, I have been actively involved in teaching medical students, residents and fellows. I currently lecture in the 2$^{nd}$ year Gastroenterology course at Icahn School of Medicine at Mount Sinai as well as the 1$^{st}$ year Infectious Disease course at New York Medical College. I have a passion for teaching and have been awarded outstanding clinic preceptor of the year as well as outstanding teacher of the year by the GI fellows. I have also lectured middle school students, high school students and adults on the role of the microbiome in our health in nationally recognized forums including the American Museum of Natural History in New York City and the Aspen Ideas Festival in Aspen, Colorado. I have been invited to speak at the Aspen Institute, Society of Fellows conference in Washington DC as well as Canada Digestive Disease Week in Calgary, Alberta. Through our clinical efforts and renown for performing FMT, Mount Sinai has been designated as a referral center for FMT.

**GRANTS, CONTRACTS, FOUNDATION SUPPORT**

**PAST GRANTS**

| Funding Source | Role | Dates | Direct Costs | Supplemental |
|---|---|---|---|---|
| Burrill B. Crohn Research Foundation<br>Fecal Microbiota Transplantation to Treat Ulcerative Colitis<br>GCO 14-1819 | PI | 2014-2016 | $50,000 | - |
| Anonymous Donor<br>SUCCESS (Sinai Ulcerative Colitis: Clinical, Experimental and Systems Studies) | Co-I | 2014-2017 | $3,000,000 | - |
| Anonymous Donor<br>SUCCESS 2.0 (Sinai Ulcerative Colitis: Clinical, Experimental and Systems Studies) | Co-I | 2017-2018 | $1,000,000 | - |
| CCF | Co-I | 2017-2019 | | - |

| | | | | |
|---|---|---|---|---|
| Inflammatory Bowel Disease and Recurrent Clostridium difficile Infection: Outcomes after Fecal Microbiota Transplantation (ICON) | | | | |
| George Waechter Family Foundation<br>Fecal Microbiota Transplantation: Clinical and mechanistic studies | PI | 2016-2019 | $124,000 | - |

**CURRENT GRANTS**

| | | | | |
|---|---|---|---|---|
| Anonymous Donor<br>SUCCESS 3.0 (Sinai Ulcerative Colitis: Clinical, Experimental and Systems Studies) | Co-I | 2018-2022 | $500,000 | Renewal will be sought |
| NIH<br>AGA Fecal Microbiota Transplantation National Registry | Co-I | 2018-2028 | $1000 | |
| Icahn School of Medicine at Mount Sinai<br>Genetics and Genomic Sciences Pilot Grant | Co-I | 2019-2020 | $25,000 | No-cost extension to 2022 |
| CCF<br>Inflammatory Bowel Disease and Recurrent Clostridium difficile Infection: Outcomes after Fecal Microbiota Transplantation + Bezlotuxumab (ICON 2.0) | Co-I | 2019-2022 | | |
| CCF – Litwin IBD Pioneers Grant<br>Defining the interactions between the immune system, strain engraftment, and patient outcomes in the REFOCUS trial of FMT capsules for UC | Co-I | 2020-2022 | | |
| NIDDK – R01 DK124133<br>Uncovering the rules of gut microbiome strain transmission | Co-I | 2020-2023 | | |
| CCF<br>Uncovering the rule of gut microbiome strain transmission | Co-I | 2020-2022 | | |
| CCF<br>Safety and Efficacy of healthy to inflamed pouch FMT | Co-I | 2021-2023 | $150,000 | |

**PENDING GRANTS**

| | | |
|---|---|---|
| NIH – U01<br>Computational Modeling of Immunometabolism (CMI) | Co-I | 2021-2026 |
| Kenneth Rainin Foundation<br>Microbial and immunological assessment in patients with chronic pouchitis undergoing FMT | Co-I | 2021-2022 |

**CLINICAL TRIALS PARTICIPATION**

| Sponsor | Role | Dates | Award | Other |
|---|---|---|---|---|
| Finch Therapeutics<br>Ranomized Double-blind, placebo-controlled, phase 3 trial of efficacy, safety and tolerability of a single oral administration of CP101 for the prevention of Recurrent clostridioides difficile infection (PRISM4) | Site PI | 2021- | N/A | Multi-center trial Phase III |
| Merck<br>A Randomized controlled trial to compare the effectiveness of Fecal Microbiota Transplantation in combination with bezlotoxumab to FMT alone for the prevention of CDI recurrence in patients with inflammatory bowel disease | Site PI | 2020- | N/A | Multi-center Trial |

**TRAINEES**

| Name | Level of Trainee | Role in Training & Inclusive Dates of Training | Publications/Awards | Trainees' Current Status/Employment |
|---|---|---|---|---|
| Yuying Luo | Resident | Mentor (2016 - present) | 6 publications | GI Fellow, MSH, NY, NY |
| Rebecca Roediger | Resident | Mentor (2015-2016) | 1 publication | Assistant Professor of Medicine, Mount Sinai Hospital |
| Nancy Yang | Med Student | Mentor (2015-2017) | 2 publications | Chief GI Fellow, Duke, Durham, NC |
| Joanna Lopez | Fellow | Mentor (2014-2016) | 1 publication<br>Chief Fellow | Director of IBD, GastroHealth, Miami, FL |
| Eugenia Schmidt | Fellow | Mentor (2016) | | Assistant Professor of Medicine, University of Minnesota, Minneapolis, MN |
| Emily Tixier | Med Student | Mentor (2018 – present) | 3 publications<br>Digestive Disease Research Foundation Awardee | Medicine Resident, Massachussetts General Hospital, Boston, MA |

| | | | | |
|---|---|---|---|---|
| Maia Kayal | Fellow | Mentor (2018 - present) | 1 publication<br><br>Chief Fellow | Assistant Professor of Medicine, MSH, NY, NY |
| Elijah Verheyen | Resident | Mentor (2016 - present) | 2 publications | GI Fellow, MSSL, NY, NY |
| Shivani Gupta | Fellow | Mentor (2017 - present) | | Assistant Professor, Henry Ford Health System, Detroit, MI |
| David Faleck | Fellow | Mentor (2016-2018) | | Assistant Professor, Memorial Sloan Kettering, NY, NY |
| Marion Protano | Fellow | Mentor 2014-2015) | | Director of FMT, Capital Health, Pennington, NJ |
| Victoria Mroz | Medical Student | Mentor (2015-) | | Medical Student, Icahn School of Medicine, NY, NY |
| Katie Dunleavy | Resident | Mentor (2019 - present) | | GI Fellow, Mayo Clinic, Rochester, MN |
| Joseph Rinaldi | Resident | Mentor (2019 - present) | | GI Fellow, Montefiore, NY, NY |
| Elizabeth Spencer | Fellow | Mentor (2018 - present) | 1 publication | Assistant Professor of Medicine, Mount Sinai Hospital |
| Stanley Cho | Resident | Mentor (2018 – 2019) | 1 publication | Pediatric GI Fellow, Baylor College of Medicine, Houston, TX |
| Kelly Suchman | Medical Student | Mentor (2019 – present) | 1 publication | Medical Resident, Northwell Health, NY, NY |

| | | | | |
|---|---|---|---|---|
| Zach Borman | Fellow | Mentor (2018-2020) | | Attending, Capital Health, Bethesda, MD |
| Akash Kumar | Fellow | Mentor (2018-2020) | Chief Fellow | Assistant Professor of Medicine, Montefiore Medical Center |
| Sheila Rustgi | Fellow | Mentor (2018-2020) | Chief Fellow | Assistant Professor of Medicine, Columbia University Medical Center |
| Yichun Fu | Resident | Mentor (2019- | 2 publication | Medicine Resident, Mount Sinai Hospital |
| Charles Du | Resident | Mentor (2019- | 2 publications | Medicine Resident, Mount Sinai Hospital |
| Agnieszka Nagorna | Resident | Mentor (2020- | | Medicine Resident, Mount Sinai Hospital |
| Joyce Chang | Resident | Mentor (2021- | | Medicine Resident, Mount Sinai Hospital |

**TEACHING ACTIVITIES**

| Activity | Level | Role | Learners | Time | Years |
|---|---|---|---|---|---|
| 2nd year GI course, Icahn School of Medicine | Medical School | Small Group Leader | Students (20) | 12 hours per year | 2014-present |
| Inpatient Rounds | Division Level | Attending | Fellows (8) | 1 month per year | 2014-present |
| Endoscopy Teaching | Division Level | Attending | Fellows | 32 hours per year | 2014-present |
| 2nd year GI Course, Icahn School of Medicine | Medical School | Lecturer | Students (180) | 1 hour per year | 2018-present |
| 2nd year ID course, New York Medical College | Medical School | Lecturer | Students (180) | 1 hour per year | 2019-present |

| | | | | | |
|---|---|---|---|---|---|
| Clinical Preparation Session leader, Icahn School of Medicine | Medical School | Small Group Leader | Students (5) | 3 hours per year | 2020-present |

**ADMINISTRATIVE LEADERSHIP APPOINTMENTS**

***Internal***

**Clinical**:

2015 – Present Director of GI Microbial Therapeutics

**Teaching**:

2015 – Present Course Director, Mount Sinai Intensive Review in GI & Hepatology

**PUBLICATIONS**

1. **Grinspan** AM, Kelly CR. Fecal Microbiota Transplantation for Ulcerative Colitis: Not Just Yet. Gastroenterology. 2015 Jul;149(1):15-8
2. **Grinspan** A, Kornbluth A. Positioning Therapy for UC. Curr Gastroenterol Rep. 2015 Aug;17(8):29
3. Lopez J, **Grinspan A**. Fecal Microbiota Transplantation for Inflammatory Bowel Disease. Gastroenterol Hepatol (NY) 2016;12:374-9.
4. Llewellyn SR, Britton GJ, Contijoch EJ, Vennaro OH, Mortha A, Colombel JF, **Grinspan A**, Clemente JC, Merad M, Faith JJ. Interactions between diet and the intestinal microbiota alter intestinal permeability and colitis severity in mice. Gastroenterology. 2018 Mar;154(4):1037-1046
5. Cho S, Spencer E, Hirten R, **Grinspan A**, Dubinsky MC. Fecal Microbiota Transplant for Recurrent Clostridium difficile infection in Pediatric Inflammatory Bowel Disease. J Pediatr Gastroenterol Nutr. 2019 Mar;68(3):343-347
6. Mankal PK, Abed J, Latte-Naor S, **Grinspan A**, Kotler DP. Fidaxomicin and Fecal Microbiota Transplants for Severe Clostridium difficile Colitis. Am J Ther. 2019 Jan/Feb;26(1):e115-117
7. Britton GJ, Contijoch EJ, Mogno I, Vennaro OH, Llewellyn SR, Ng R, Li Z, Mortha A, Merad M, Das A, Gevers D, McGovern DPB, Singh N, Braun J, Jacobs JP, Clemente JC, **Grinspan** A, Sands BE, Colombel JF, Dubinsky MC, Faith JJ. Microbiotas from Humans with Inflammatory Bowel Disease Alter the Balance of Gut Th17 and RORγt$^{+}$ Regulatory T Cells and Exacerbate Colitis in Mice Immunity. 2019 Jan 15;50(1):212-224
8. Contijoch EJ, Britton GJ, Yang C, Mogno I, Li Z, Ng R, Llewellyn SR, Hira S, Johnson C, Rabinowitz KM, Barkan R, Dotan I, Hirten RP, Fu SC, Luo Y, Yang N, Luong T, Labrias PR, Lira S, Peter I, **Grinspan A**, Clemente JC, Kosoy R, Kim-Schulze S, Qin X, Castillo A, Hurley A, Atreja A, Rogers J, Fasihuddin F, Saliaj M, Nolan A, Reyes-Mercedes P, Rodriguez C, Aly S, Santa-Cruz K, Peters L, Suárez-Fariñas M, Huang R, Hao K, Zhu J, Zhang B, Losic B, Irizar H, Song WM, Di Narzo A, Wang W, Cohen BL, DiMaio C, Greenwald D, Itzkowitz S, Lucas A, Marion J, Maser E, Ungaro R, Naymagon S, Novak J, Shah B, Ullman T, Rubin P, George J, Legnani P, Telesco SE, Friedman JR, Brodmerkel C, Plevy S, Cho JH, Colombel JF, Schadt EE, Argmann C, Dubinsky M, Kasarskis A, Sands B, Faith JJ. Gut microbiota density influences host physiology and is shaped by host and microbial factors. Elife 2019;8:e40553
9. Verheyen E, Dalapathi V, Arora S, Patel K, Mankal PK, Kumar V, Lung E, Kotler DP, **Grinspan A**. High 30-day readmission rates associated with Clostridium difficile infection. Am J Infect Cont 2019;47(8):922-927

10. Hirten R*, **Grinspan A***, Fu S, Luo Y, Suarez-Farinas M, Rowland J, Contijoch EJ, Mogno I, Yang N, Luong T, Labrias PR, Peter I, Cho JH, Sands BE, Colombel JF, Faith JJ, Clemente JC. Microbial Engraftment and Efficacy of Fecal Microbiota Transplant for Clostridium difficile in Patients with and without Inflammatory Bowel Disease. Inflamm Bowel Dis 2019;25(6):969-979
11. Santiago, M, Eysenbach L, Allegretti J, Aroniadis O, Brandt LJ, Fischer M, **Grinspan A**, Kelly C, Morrow C, Rodriguez M, Osman M, Kassam Z, Smith MB, Timberlake S. Microbiome predictors of dysbiosis and VRE decolonization in patients with recurrent Clostridium difficile infections in a multi-center retrospective study. AIMS Microbiology 2019;5(1):1-18
12. Kelly CR, Fischer M, **Grinspan A**, Allegretti JR. Patients Eligible for Trials of Microbe-based Therapeutics do not Represent the Population With Recurrent Clostridioides difficile Infection. Clin Gastroenterol Hepatol 2020;18(5):1099-1101
13. Luo Y, Lucas AL, **Grinspan AM**. Fecal Transplants by Colonoscopy and Capsules are Cost-Effective Strategies for Treating Recurrent Clostridioides difficile infection. Dig Dis Sci 2020;65(4):1125-1133
14. Tixier EN, Verheyen E, Ungaro RC, **Grinspan AM**. Faecal microbiota transplant decreases mortality in severe and fulminant Clostridioides difficile infection in critically ill patients. Aliment Pharmacol Ther 2019;50(10):1094-1099
15. **Grinspan A**. Fecal Microbiota Transplantation in Inflammatory Bowel Disease Patients with Clostridium difficile infection. Gastroenterol Hepatol (NY) 2019;15:481-3
16. Luo Y, Tixier EN, **Grinspan AM**. Fecal Microbiota Transplantation for Clostridioides difficile in high-risk older adults is associated with early recurrence. Dig Dis Sci 2020 Feb 20 [epub ahead of print]
17. **Grinspan A**. FMT for Severe C. difficile Infection: If at First You Do No Harm, the Second Time's a Charm. Dig Dis Sci 2020 May 14 [epub ahead of print]
18. Kayal M, Tixier E, Plietz M, Radcliffe M, Rizvi A, Riggs A, Trivedi P, Khaitov S, Sylla P, Greenstein A, Dubinsky MC, **Grinspan A**. Clostridioides difficile infection is a rare cause of infectious pouchitis. Inflamm Intest Dis. 2020;5(2):59-64
19. Hirten RP, Stanley S, Danieletto M, Borman Z, **Grinspan A**, Rao P, Sauk J, Chang L, Arnrich B, Bottinger E, Keefer L, Sands BE. Wearable Devices Are Well Accepted by Patients in the Study and Management of Inflammatory Bowel Disease: A Survey Study. Dig Dis Sci 2020 [epub ahead of print]
20. Allegretti JR, Kelly CR, **Grinspan A**, Mullish BH, Kassam Z, Fischer M. Outcomes of Fecal Microbiota Transplantation in Patients With Inflammatory Bowel Diseases and Recurrent Clostridioides difficile Infection. Gastroenterology 2020 [epub ahead of print]
21. Britton GJ, Contijoch EJ, Spindler MP, Aggarwala V, Dogan B, Bongers G, San Mateo L, Baltus A, Das A, Gevers D, Borody TJ, Kaakoush NO, Kamm MA, Mitchell H, Paramsothy S, Clemente JC, Colombel JF, Simpson KW, Dubinsky MC, **Grinspan A**, Faith JJ. Defined microbiota transplant restores Th17/RORγt + regulatory T cell balance in mice colonized with inflammatory bowel disease microbiotas. Proc Natl Acad Sci U S A. 2020;117(35):21536-21545
22. Luo Y, Grinspan LT, Fu Y, Adams-Sommer V, Willey DK, Patel G, **Grinspan AM**. Hospital-Onset Clostridioides difficile Infections during the COVID-19 Pandemic. Infect Control Hosp Epidemiol 2021 Sep;42(9):1165-1166
23. Livanos AE, Jha D, Cossarini F, Gonzalez-Reiche AS, Tokuyama M, Aydillo T, Parigi TL, Ramos I, Dunleavy K, Lee B, Dixon R, Chen ST, Martinez-Delgado G, Nagula S, Ko HM, Reidy J, Naymagon S, **Grinspan A**, Ahmad J, Tankelevich M, Gordon R, Sharma K, Britton GJ, Chen-Liaw A, Spindler MP, Plitt T, Wang P, Cerutti A, Faith JJ, Colombel JF, Kenigsberg E, Argmann C, Merad M, Gnjatic S, Harpaz N, Danese S, Rahman A, Kumta NA, Aghemo A, Petralia F, van Bakel H, Garcia-Sastre A,

Mehandru S. Intestinal Host Response to SARS-CoV-2 Infection and COVID-19 Outcomes in Patients With Gastrointestinal Symptoms. 2021 Jun;160(7):2435-2450

24. Kelly CR, Yen EF, **Grinspan AM**, Kahn SA, Atreja A, Lewis JD, Moore TA, Rubin DT, Kim AM, Serra S, Nersesova Y, Fredell L, Hunsicker D, McDonald D, Knight R, Allegretti JR, Pekow J, Absah I, Hsu R, Vincent J, Khanna S, Tangen L, Crawford CV, Mattar MC, Chen LA, Fischer M, Arsenescu RI, Feuerstadt P, Goldstein J, Kerman D, Ehrlich AC, Wu GD, Laine L. Fecal Microbiota Transplantation is hightly effective in real-world practice: Initial results from the FMT National Registry. Gastroenterology 2021;160(1):183-192.
25. Allegretti JR, Kelly CR, **Grinspan A**, Mullish BH, Hurtado J, Carrellas M, Marcus J, Marchesi JR, McDonald JAK, Gerardin Y, Silverstein M, Pechlivanis A, Barker GF, Miguens Blanco J, Alexander JL, Gallagher KI, Pettee W, Phelps E, Nemes S, Sagi SV, Bohm M, Kassam Z, Fischer M. Inflammatory Bowel Disease outcomes following Fecal Microbiota Transplantation for recurrent C. difficile infection. Inflamm Bowel Dis. 2021 Aug 19;27(9):1371-1378
26. Livanos AE, Jha D, Cossarini F, Gonzalez-Reiche AS, Tokuyama M, Aydillo T, Parigi TL, Ladinsky MS, Ramos I, Dunleavy K, Lee B, Dixon R, Chen ST, Martinez-Delgado G, Nagula S, Bruce EA, Ko HM, Glicksberg BS, Nadkarni G, Pujadas E, Reidy J, Naymagon S, **Grinspan A**, Ahmad J, Tankelevich M, Bram Y, Gordon R, Sharma K, Houldsworth J, Britton GJ, Chen-Liaw A, Spindler MP, Plitt T, Wang P, Cerutti A, Faith JJ, Colombel JF, Kenigsberg E, Argmann C, Merad M, Gnjatic S, Harpaz N, Danese S, Cordon-Cardo C, Rahman A, Schwartz RE, Kumta NA, Aghemo A, Bjorkman PJ, Petralia F, van Bakel H, Garcia-Sastre A, Mehandru S. Intestinal Host Response to SARS-CoV-2 Infection and COVID-19 Outcomes in Patients With Gastrointestinal Symptoms. Gastroenterology. 2021 Mar 4:S0016-5085(21)00461-3 [online ahead of print]
27. Du C, Luo Y, Walsh S, **Grinspan A.** Oral Fecal Microbiota Transplant Capsules are safe and effective for recurrent Clostridioides difficile infection: A systematic review and meta-analysis. J Clin Gastroenterol. 2021 Apr 1;55(4):300-308
28. Tixier EN, Verheyen E, Luo Y, Grinspan LT, Du CH, Ungaro RC, Walsh S, **Grinspan** AM. Systematic Review with Meta-Analysis: Fecal Microbiota Transplantation for Severe or Fulminant Clostridioides difficile. Dig Dis Sci. 2021 Mar 22. [online ahead of print]
29. Fu Y, Luo Y, **Grinspan AM**. Epidemiology of community-acquired and recurrent Clostridioides difficile infection. Therap Adv Gastroenterol. 2021 May 22
30. Aggarwala V, Mogno I, Li Z, Yang C, Britton GJ, Chen-Liaw A, Mitcham J, Bongers G, Gevers D, Clemente JC, Colombel JF, **Grinspan A**, Faith J. Precise quantification of bacterial strains after fecal microbiota transplantation delineates long-term engraftment and explains outcomes. Nat Microbiol. 2021 Oct;6(10):1309-1318
31. Suchman K, Luo Y, **Grinspan A**. Fecal Microbiota Transplant for Clostridioides difficile infection is safe and efficacious in an immunocompromised cohort. Dig Dis Sci. 2022 Jan 9 [online ahead of print]

**<u>BOOK CHAPTERS</u>**

**Grinspan** A, Kornbluth A. Ischemic Disease of the Small and Large Bowel. In: Mount Sinai Expert Guides: Gastroenterology. Sands BE (Ed.), Wiley-Blackwell, 2015; 422-434.

**Grinspan A,** Boutros M. Organ preserving strategies in the management of fulminant Clostridium difficile colitis, In: The SAGES Manual of Colorectal Surgery. Sylla P, Kaiser AM, Popowich DA (Ed.), Springer, 2019

Grinspan LT, **Grinspan A**. Discharge: How Should You Follow and Care For Patients After FMT?, In: The 6 D’s of Fecal Microbiota Transplantation: A Practical Primer for Clinicians from Decision to Discharge and Beyond. Allegretti JA, Kassam A (Ed.), SLACK, 2019

**INVITED LECTURES/PRESENTATIONS**

International:

2019 The Role of FMT in IBD. Canada DDW Annual Meeting, Banff, Canada. March 2019

National:

2014 Career Mentoring Program. American College of Gastroenterology Annual Meeting. Philadelphia, PA. October 2014

2018 The Power of Poop. Teen SciCafe, American Museum of Natural History, NY, NY

2018 The Power of Poop. SciCafe, American Museum of Natural History, NY, NY

2018 The Power of Poop. Aspen Ideas Festival. Aspen, CO

2019 The Power of Poop. SciChat, American Museum of Natural History, NY, NY

2019 The Role of the Microbiome in Human Health and Disease. Aspen Institute, Society of Fellows, Washington DC

2020 Optimal Management of Clostridioides difficile infection (CDI) and CDI recurrent in Patients with IBD. AIBD 2020

2021 The Changing Landscape of Fecal Microbial Transplantation (FMT). Postgraduate course, DDW 2021

Local/Regional:

2013 Fecal Microbial Transplantation for IBD. Dr. Stephen Matarazzo IBD Conference, CCFA Philadelphia/Delaware Valley Chapter, Galloway, NJ.

2013 Have You Heard About Fecal Transplant? Dr. Stephen Matarazzo IBD Conference, CCFA Philadelphia/Delaware Valley Chapter, Galloway, NJ

2014 Fecal Microbiota Transplantation: C diff and beyond. Grand Rounds, Department of Medicine, Elmhurst Hospital, Queens, NY

2014 Fecal Microbiota Transplantation: C diff and IBD. Grand Rounds, Division of Gastroenterology, St. Luke's/Roosevelt Hospital, NY, NY.

2014 Fecal Microbiota Transplantation in IBD. 11th Annual Mount Sinai Inflammatory Bowel Disease Consultant's Course. NY, NY.

2014 Fecal Microbiota Transplantation: Lessons from Clostridium difficile. Grand Rounds, Department of Medicine, Elmhurst Hospital, Queens, NY

2015 Fecal Microbiota Transplantation for Clostridium difficile and Beyond. Grand Rounds, Department of Medicine, Mount Sinai Hospital, NY, NY

2015 Fecal Microbiota Transplantation: Role in IBD. 12th Annual Mount Sinai Inflammatory Bowel Disease Consultant's Course. NY, NY

2015 Fecal Microbiota Transplantation: C diff and IBD. Grand Rounds, Division of Gastroenterology, Mount Sinai Beth Israel, NY, NY

2016 Clostridium difficile infection: Treatment Strategies. Grand Rounds, Division of Gastroenterology. New York-Presbyterian Hospital, Weill-Cornell Medical Center, NY, NY

2016 Clostridium difficile Infection: Concepts and Management. Grand Rounds, Department of Medicine, Mount Sinai West, NY, NY

2016 Clostridium difficile infection: Current Management Strategies. Grand Rounds, Department of Medicine, Lincoln Medical Center, Bronx, NY

2016 Management of Severe Clostridium difficile infection. Grand Rounds, Department of Surgery, Mount Sinai Hospital, NY, NY

2016 Fecal Microbiota Transplantation for Clostridium difficile and Beyond. Grand Rounds, Department of Pediatrics, Mount Sinai Hospital, NY, NY

2016 The Current Status of Clostridium difficile infection: Diagnosis, Management and Challenges. New York Metropolitan Society of Gastroenterology Nurses and Associates, Region 40. Spring Course 2016, NY, NY

2016 Fecal Microbiota Transplantation for Clostridium difficile and Beyond. Grand Rounds, Department of Medicine, Mount Sinai Hospital Queens, Queens, NY

2017 The Current Status of Clostridium difficile infection: Diagnosis, Management and Challenges. New York Metropolitan Society of Gastroenterology Nurses and Associates, Region 40. Fall Course 2017, NY, NY

2017 Manipulating the Microbiome: The Role of FMT. Grand Rounds, Division of Gastroenterology, Mount Sinai Hospital, NY, NY

2017 Novel Therapeutics to Altering the Microbiome. Medical Scientist Grand Rounds, Mount Sinai Hospital, NY, NY

2017 Fecal Microbiota Transplantation for Severe Clostridium difficile. Grand Rounds, Colorectal Surgery, Mount Sinai Hospital, NY, NY

2017 Fecal Microbiota Transplantation for Clostridium difficile and Beyond. Grand Rounds, Department of Medicine, Richmond University Medical Center, NY, NY

2017 Fecal Microbiota Transplantation for Clostridium difficile and Beyond. Grand Rounds, Department of Pediatrics, Staten Island University Hospital, NY, NY

2017 Clostridium difficile infection: Current Management Strategies. Grand Rounds, Department of Medicine, Mount Sinai Beth Israel, NY NY

2018 Update on Fecal Microbiota Transplantation in IBD. IBD Conference, Division of Gastroenterology, Mount Sinai Hospital. NY, NY

2018 Update in management of Clostridium difficile. Grand Rounds, Department of Medicine, Metropolitan Hospital, NY, NY.

2018 Fecal Microbiota Transplantation in IBD. 15th Annual Mount Sinai Inflammatory Bowel Disease Consultant's Course. NY, NY

2018 The Power of Poop: FMT for Clostridium difficile and IBD. Grand Rounds, Division of Pediatric Gastroenterology, Mount Sinai Hospital. NY, NY.

2018 Clostridium difficile and Fecal Microbiota Transplant. Spring Review in Internal Medicine. Department of Medicine, Richmond University Medical Center. Staten Island, NY.

2018 Fecal Microbiota Transplantation: Clostridium difficile and Beyond. 2nd Annual Mount Sinai Live Endoscopy Course, Mount Sinai Hospital, NY, NY.

2018 The Power of Poop: Role of FMT in IBD. Crohn's & Colitis Foundation: Fairfield/Westchester Patient Education Symposium, Greenwich Hospital, Greenwich, CT

2018 The Role of the Microbiome in Human Health and Disease. HIV Grand Rounds, Mount Sinai West, NY, NY

2018 *Clostridioides difficile* infection. Bone Marrow Transplant Grand Rounds, Mount Sinai Hospital, NY, NY

2019 Fecal Microbiota Transplantation: Expanding role in the clinic. 3rd Annual Mount Sinai Live Endoscopy Course, Mount Sinai Hospital, NY, NY.

2019 *Clostridioides difficile* infection: Management of severe and fulminant disease. Colorectal Surgery Grand Rounds, Mount Sinai Hospital, NY, NY

2019 The Power of Poop: How FMT is transforming medicine. 2nd Annual Microbiome Symposium, New York Medical College, Valhalla, NY

2019 Fecal Microbiota Transplantation: What do Gastroenterologists need to know. Grand Rounds, Division of Gastroenterology, Memorial Sloan Kettering Cancer Center, NY, NY

2019 Fecal Microbiota Transplantation for IBD. New York Society of Gastrointestinal Endoscopy (NYSGE) IBD Seminar, NY, NY

2019 Fecal Microbiota Transplantation: What do health care providers need to know. New York Metropolitan Society of Gastroenterology Nurses and Associates, Region 40. Fall Course 2019, NY, NY

2019 Bacteriotherapy in IBD: What can we do now and what may we do in the future? 16th Annual Mount Sinai Inflammatory Bowel Disease Consultant's Course. NY, NY

2019 Update on *Clostridioides difficile* infection and FMT. Grand Rounds, Division of Hospital Medicine, Mount Sinai Hospital, NY, NY

2019 Hitchhiker's Guide to Fecal Microbiota Transplant. Grand Rounds, Department of Medicine, Mount Sinai Hospital, NY, NY

2019 Hitchhiker's Guide to the Gut Microbiome. Medical School lecture, Albert Einstein College of Medicine, Bronx, NY

2020 Hitchhiker's Guide to the Microbiome. Grand Rounds, Department of Medicine, Metropolitan Hospital, NY, NY

2020 Hitchhiker's Guide to the Microbiome. Keynote Speaker, Medical Student Research Forum, New York Medical College, Valhalla, NY

2020 Hitchhiker's Guide to the Microbiome. Grand Rounds, Division of Gastroenterology, Northwell Health, Manhasset, NY

2020 FMT for Fulminant C diff infections. Grand Rounds, Division of Colorectal Surgery, Mount Sinai Hospital, NY, NY

2021 Fecal Microbiota Transplantation: *Clostridiodies difficile* and Beyond. Grand Rounds, Department of Medicine, Richmond University Medical Center, Staten Island, NY

2021 Fulminant *Clostrioides dificile* infection: FMT is a game-changer. Grand rounds, Division of Colorectal Surgery, Mount Sinai Hospital, NY, NY

2021 Clostridioides difficile: What to order and When to treat. 18th Annual Mount Sinai Consultant's Course, Mount Sinai Hospital, NY, NY

2021 Fecal Microbiota Transplant: Hitchhikers' Guide to the Microbiome. Grand Rounds, Department of Medicine, Mount Sinai Morningside

2021 Fecal Microbiota Transplant: Hitchhikers' Guide to the Microbiome. Research Seminar, Department of Medicine, Renaissance School of Medicine at Stony Brook University, Stony Brook, NY

2021 Fecal Microbiota Transplant for IBD. IBD conference, Mount Sinai Hospital, NY, NY

2021 Current treatments for recurrent and refractory *Clostridioides* infection. Grand Rounds, Division of Infectious Diseases, Donald and Barbara Zucker School of Medicine at Hofstra Northwell, Manhassat, NY

2022 Current treatments for severe and fulminant *Clostridioides* infection. Grand rounds, Division of colorectal surgery, Mount Sinai Hospital, NY, NY

2022 Current treatments for recurrent and refractory *Clostridioides* infection. Intern report, Department of Medicine, Mount Sinai Hospital, NY, NY

2022 Current treatments for recurrent and refractory *Clostridioides* infection. Resident report, Department of Medicine, Mount Sinai Hospital, NY, NY

2022 Current treatments for recurrent and refractory *Clostridioides* infection. Grand Rounds, Department of Medicine, Mount Sinai South Nassau, Oceanside, NY.

#### SELECTED VOLUNTARY PRESENTATIONS

2008 **Grinspan A**, Bernheim O, Yousefazadeh E, Rahmani R, Southern W, Brandt L. *Clostridium difficile* as a causative agent of post-infection Irritable Bowel Syndrome (PI-IBS). Oral Presentation at Digestive Disease Week, San Diego, CA

2016 Eysenbach L, Allegretti JR, Aroniadis O, Brandt L, Donovan D, Fischer M, **Grinspan A**, Kassam Z, Kelly CR, Kim C, Morrow C, Osman M, Quan J, Rodriguez M, Walker T, Smith M. Clearance of Vancomycin-Resistant Enterococcus Colonization with Fecal Microbiota Transplantation Among Patients with Recurrent Clostridium difficile Infection. ID Week 2016, New Orleans, LA.

2017 Cho S, Spencer E, Hirten R, **Grinspan A**, Dubinsky M. High Recurrence Rate after Fecal Microbiota Transplant for Recurrent Clostridium difficile Infection in Pediatric Inflammatory Bowel Disease. Poster Presentation at NASPHGAN in Las Vegas, NV.

2018 Luo Y, Lucas A, **Grinspan A**. Cost-Effectiveness Analysis of Multiple Strategies for Recurrent Clostridium difficile Infection. Oral Presentation at DDW, Washington DC

2019 Allegretti, JR, Hurtado J, Carrellas M, Marcus J, Phelps E, Wong WF, Marchesi J, Mullish BH, McDonald JAK, Pechlivanis A, Barker GF, Blanco JM, Sagi S, Bohm M, Kelly CR, Kassam Z, **Grinspan A**, Fischer M. The ICON Study: Inflammatory Bowel Disease and recurrent Clostridium difficile infection: outcomes after fecal microbiota transplantation. Plenary presentation at DDW, San Diego, CA.

2019 Tixier E, Verheyen E, Ungaro R, **Grinspan A**. Fecal Microbiota Transplant Decreases Mortality in Patients with Severe and Fulminant *Clostridium difficile* Infection. Poster of Distinction at DDW, San Diego, CA

2020 Kelly, CR, Yen EF, **Grinspan A**, et al. Fecal Microbiota Transplantation is highly effective in real-world practice: initial results from the American Gastroenterological Association Fecal Microbiota Transplantation National Registry. Oral presentation at DDW, Chicago, IL

**MEDIA RESOURCE EDUCATIONAL MATERIALS**

2015 Guest commentary: The exciting future of fecal microbiota transplant. Healio Gastroenterology
2017 Fecal Transplant Promising In Colitis. MedPage Today
2017 Everything you need to know about FMT. Women's Health
2018 Fecal Transplant in Human Health and Disease. Greek Radio Podcast
2018 The Poop Cure. American Museum of Natural History
2020 The Smartest Doctor in the Room. Dean Mitchell Podcast
2022 Current state of *Clostridioides difficile* infection: Role of FMT. Physician's Weekly.

#### DATA SAFETY AND MONITORING PLAN

Dr. Ryan Ungaro will chair the data safety and monitoring board for this clinical trial. Dr. Ungaro is well versed in reporting procedures as they are currently required for all research conducted at Mount Sinai. All research staff will complete training in compliance with Mount Sinai's IRB for conducting human subjects research.

##### Data and Safety Monitoring Board (DSMB)

Safety oversight for the trial will be under the direction of a DSMB. The DSMB is an independent group of experts, led by chairperson Dr. Ungaro, who will advise the study investigators. The primary responsibilities of the DSMB are to 1) periodically review and evaluate the data for subject safety, study conduct, study progress, and efficacy, and 2) make recommendations concerning the continuation, modification, or termination of the trial. The DSMB will be composed of at least 3 voting members. Members will have clinical expertise in the medical area and subject population being studied. All DSMB members will be separate and independent of study personnel participating in this trial and will not have scientific, financial or other conflict of interest related to the trial. Procedures for DSMB data reviews will be defined in the DSMB Charter that will include DSMB membership, responsibilities, and the scope and frequency of data reviews. The study will be reviewed by the DSMB at least annually and after 15 patients are enrolled.

##### Halting Rules

###### *Study enrollment halting rules*

Enrollment in the study will be suspended for conduct of a safety review by the DSMB in the case of:

- A grade 3 or higher AE or SAE that is other than 'unrelated' to the study intervention.
- One case of allergic reaction or anaphylaxis, or one infection assessed as anything other than 'unrelated' to study intervention
- An overall pattern of symptomatic, clinical or laboratory events that the PI considers related to study product and that may appear minor in terms of individual events, but that may collectively represent a serious potential concern for safety.

###### *Individual's halting rules*

Subjects who meet any of the following criteria must be assessed by the PI to determine if it is in the subject's best interest to stop the study product(s):

- Subject choice (Withdrawal of consent)
- Participant's non-compliance.
- Development of a significant medical condition and/or participation in the study is no longer in the best interest of the subject.

### APPENDICIES

### Appendix 1 – MTC01 Donor Health Record

Health record FMT donor D283.

| | Donor D283 |
|---|---|
| History | |
| Medical history | No |
| Surgical history | No |
| Family history of any significant medical problems | No |
| Medications | No |
| Known allergies | No |
| 10 point Review of Systems negative | No |
| Stool tests | |
| Shiga toxin EIA | Not detected |
| Salmonella/Shigella culture | Negative |
| Campylobacter culture | Negative |
| Ova and Parasites | Negative |
| C diff GDH antigen | N/A |
| C diff Toxin EIA | N/A |
| C diff PCR | Negative |
| Blood tests | |
| RPR | Negative |
| Hepatitis A antibody | Nonreactive |
| Hepatitis B surface Antigen | Nonreactive |
| Hepatitis B surface Antibody | Nonreactive |
| Hepatitis B core antibody | Nonreactive |
| Hepatitis C antibody | Nonreactive |
| HIV 1/2 | Nonreactive |
| CMV IgG Antibody | Negative |

### Appendix 2 – MTC01 Strain identification

**Genomic identity**
Genome length determined from the draft SPAdes assemblies[46]. Species named determined by kmer-based genome alignment to >150K RefSeq bacterial genomes.

| Strain ID | Species | Genome Length (Mbp) |
|---|---|---|
| MTC01.01 | *Bacteroides uniformis* | 4.52 |
| MTC01.02 | *Bacteroides ovatus* | 6.86 |
| MTC01.03 | *Bifidobacterium longum* | 2.57 |
| MTC01.04 | *Bacteroides thetaiotaomicron* | 7.06 |
| MTC01.05 | *Bacteroides vulgatus* | 5.40 |
| MTC01.07 | *Parabacteroides distasonis* | 5.44 |
| MTC01.08 | *Bifidobacterium adolescentis* | 2.10 |
| MTC01.09 | *Parabacteroides merdae* | 4.49 |
| MTC01.10 | *Coprococcus comes* | 3.35 |
| MTC01.11 | *Eubacterium rectale* | 3.31 |
| MTC01.12 | *Bacteroides caccae* | 5.73 |
| MTC01.13 | *Dorea longicatena* | 3.00 |
| MTC01.14 | *Odoribacter splanchnicus* | 4.74 |
| MTC01.15 | *Bacteroides cellulosilyticus* | 6.92 |
| MTC01.16 | *Bifidobacterium pseudocatenulatum* | 2.30 |

**Bruker MALDI Biotyper identity**

| Strain ID | Genus | Species | Bruker Species | Bruker Score |
|---|---|---|---|---|
| MTC01.01 | *Bacteroides* | *uniformis* | Bacteroides uniformis | >1.8 |
| MTC01.02 | *Bacteroides* | *ovatus* | Bacteroides ovatus | >1.8 |
| MTC01.03 | *Bifidobacterium* | *longum* | Bifidobacterium longum | >1.8 |
| MTC01.04 | *Bacteroides* | *thetaiotaomicron* | Bacteroides thetaiotaomicron | >1.8 |
| MTC01.05 | *Bacteroides* | *vulgatus* | Bacteroides vulgatus | >1.8 |
| MTC01.06 | *Collinsella* | *aerofaciens* | Collinsella aerofaciens | >1.8 |
| MTC01.07 | *Parabacteroides* | *distasonis* | Parabacteroides distasonis | >1.8 |
| MTC01.08 | *Bifidobacterium* | *adolescentis* | Bifidobacterium adolescentis | >1.8 |
| MTC01.09 | *Parabacteroides* | *merdae* | Parabacteroides johnsonii | >1.8 |
| MTC01.10 | *Coprococcus* | *comes* | Coprococcus comes | >1.8 |
| MTC01.11 | *Eubacterium* | *rectale* | Eubacterium rectale | >1.8 |
| MTC01.12 | *Bacteroides* | *caccae* | Bacteroides caccae | >1.8 |
| MTC01.13 | *Dorea* | *longicatena* | Dorea longicatena | >1.8 |
| MTC01.14 | *Odoribacter* | *splanchnicus* | Odoribacter splanchnicus | >1.8 |
| MTC01.15 | *Bacteroides* | *cellulosilyticus* | Bacteroides cellulosilyticus | >1.8 |
| MTC01.16 | *Bifidobacterium* | *pseudocatenulatum* | Bifidobacterium pseudocatenulatum | >1.8 |

For species with very small colonies (e.g., E. rectale), identification will alternatively be determined by transferring a colony into broth where it may be grown to an OD600 > 0.1 and identification will be determined from MALDI-TOF of the broth with a minimum Bruker score >1.8. For broth and colonies, the correct species must be the top hit. Strains MTC01.08 *B. adolescentis*, MTC01.10 *C. comes*, MTC01.11 *E. rectale* and MTC01.13 *D. longicatena* are identified by custom MALDI-TOF MSP produced internally and verified by genome sequencing.

### Appendix 3 – MTC01 in vitro antibiotic susceptibility

| Organism | ID | MIC (µg/mL) | | | | | | | | | | | | | | | | | | | | | |
|---|---|---|---|---|---|---|---|---|---|---|---|---|---|---|---|---|---|---|---|---|---|---|---|
| | | VAN | | MTZ | | TGC | | SAM | | AMC | | MEM | | TZP | | CLI | | CRO | | MOX | | etest only | |
| | | CLSI | etest | CLSI | etest | CLSI | etest | CLSI | etest | CLSI | etest | CLSI | etest | CLSI | etest | CLSI | etest | CLSI | etest | CLSI | etest | CAZ | IMI |
| *Bacteroides uniformis* | MTCO1 .01 | >32 | 32 | 1 | 1 | 0.25 | 0.047 | 1/0.5 | 1 | 1/0.5 | 0.38 | 0.12 | 0.094 | 0.25/4 | 0.25 | 2 | 1.5 | 64 | 38 | 2 | 1.5 | >256 | 0.064 |
| *Bacteroides ovatus* | MTCO1 .02 | >32 | >256 | 1 | 0.75 | 0.25 | 0.094 | 1/0.5 | 0.75 | 0.5/0.25 | 0.25 | 0.25 | 0.094 | 4/4 | 1.5 | 2 | 0.064 | 64 | 32 | 2 | 1 | >256 | 0.094 |
| *Bifidobacterium longum* | MTCO1 .03 | 1 | 0.25 | 8 | 8 | 0.5 | 0.125 | 0.5/0.25 | 0.032 | 0.5/0.25 | 0.064 | 0.06 | 4 | 0.5/4 | 0.19 | 0.06 | 0.047 | 4 | 2 | 0.5 | 8 | 0.38 | 0.016 |
| *Bacteroides thetaiotaomicron* | MTCO1 .04 | >32 | 12 | 1 | 1 | 2 | 0.5 | 1/0.5 | 0.75 | 1/0.5 | 0.25 | 0.25 | 0.094 | 4/4 | 1 | 4 | 1.5 | 64 | 48 | 1 | 0.75 | >256 | 0.064 |
| *Bacteroides vulgatus* | MTCO1 .05 | 32 | 12 | 0.5 | 1 | 2 | 0.5 | 4/2 | 1 | 0.5/0.25 | 0.125 | 0.25 | 0.19 | 4/4 | 1.5 | >8 | >256 | 16 | 4 | >16 | >32 | 32 | 0.023 |
| *Parabacteroides distasonis* | MTCO1 .07 | >32 | >256 | 1 | 0.75 | 1 | 0.125 | 8/4 | 3 | 4/2 | 1.5 | 0.5 | 0.19 | >32/4 | 6 | 0.25 | 0.094 | 32 | 0.75 | 4 | 6 | 24 | 0.19 |
| *Bifidobacterium adolescentis* | MTCO1 .08 | 1 | 0.5 | >16 | >256 | .5 | 0.094 | 1/0.5 | 0.064 | 0.25/0.12 | 0.047 | 0.12 | 0.047 | <0.06/4 | 0.023 | <=0.06 | 0.064 | <=0.5 | 0.047 | 8 | >256 | 0.5 | 0.016 |
| *Parabacteroides merdae* | MTCO1 .09 | >32 | 24 | 1 | 0.25 | 1 | 0.094 | 2/1 | 3 | 2/1 | 0.75 | 0.25 | 0.25 | 16/4 | 4 | 2 | 1 | 16 | 32 | 0.25 | 0.19 | 64 | 0.38 |
| *Coprococcus comes* | MTCO1 .10 | 0.5 | 0.25 | 0.5 | 0.094 | 0.12 | 0.016 | 0.03/0.015 | 0.016 | <=0.015/0.0 08 | 0.016 | 0.06 | 0.094 | 0.12/4 | 0.016 | <=0.03 | 0.016 | <=0.5 | 0.094 | 8 | 6 | 0.75 | 0.016 |
| *Eubacterium rectale* | MTCO1 .11 | 0.5 | 0.25 | 0.06 | 0.094 | 0.06 | 0.016 | ND | 0.023 | ND | 0.016 | ND | 0.016 | ND | 0.032 | <=0.06 | 0.016 | ND | 0.026 | 4 | >32 | 0.38 | 0.016 |
| *Bacteroides caccae* | MTCO1 .12 | >32 | 32 | 1 | 0.5 | >8 | 4 | 2/1 | 0.75 | 0.5/0.25 | 0.19 | 0.12 | 0.125 | 4/4 | 1 | 1 | 0.38 | 16 | 4 | 2 | 2 | 32 | 0.125 |
| *Dorea longicatena* | MTCO1 .13 | 0.5 | 0.5 | 0.06 | 0.5 | 0.25 | 0.094 | 0.5/0.25 | 0.38 | 0.25/0.12 | 0.19 | 0.5 | 0.38 | 1/4 | 0.25 | 4 | 1.5 | 2 | 0.38 | 8 | >32 | 4 | 0.125 |
| *Odoribacter splanchnicus* | MTCO1 .14 | >32 | >256 | 1 | 0.19 | 0.12 | 0.016 | 4/2 | 0.75 | 2/1 | 0.19 | 0.12 | 0.023 | <0.06/4 | 0016 | >8 | >256 | 16 | 4 | 2 | 1 | 6 | 0.032 |
| *Bacteroides cellulosilyticus* | MTCO1 .15 | >32 | 24 | 1 | 0.75 | 2 | 0.25 | 4/2 | 3 | 2/1 | 1 | 0.25 | n.a. | 8/4 | 1.5 | 2 | 1 | 64 | 128 | 4 | 3 | >256 | 0.25 |
| *Bifdobacterium pseudocatenulatum* | MTCO1 .16 | 0.5 | 0.38 | 2 | 48 | 1 | 0.032 | 0.12/0.06 | 0.125 | 0.06/0.03 | 0.094 | 0.12 | 0.064 | 0.5/5 | 0.38 | <0.03 | 0.047 | <0.5 | 0.38 | 4 | 4 | 1 | 0.047 |

| Color Legend |
|---|
| Susceptible |
| Intermediate |
| Resistant |
| Intermediate or Resistant |

The above table provides results of Minimum Inhibitory Concentrations (MIC) determined for 15 strains in MTC01. MIC was determined for by agar dilution in accordance with guidelines from the Clinical and Laboratory Standards Institute (CLSI; 1,2). These CLSI experiments were performed by Micromyx, LLC (their report is on the following pages). Similarly we performed an etest internally that was highly correlated with the CLSI results (Figure 2C). An additional two antibiotics were tested by etest only.

Antibiotic abbreviations: vancomycin (VAN), metronidazole (MTZ), tigecycline (TGC), ampicillin-sulbactam (SAM), amoxicillin-clavulante (AMC), meropenem (MEM), piperacillin-tazobactam (TZP), clindamycin (CLI), ceftriaxone (CRO), amoxicillin (MOX), ceftazidime/avibactam (CAZ), imipenem/relebactam (IMI).

### Micromyx, LLC in vitro antibiotic susceptibility report

### FINAL REPORT

**TITLE:** Antibiograms of 16 strains from a Live Bacterial Therapeutic by Anaerobic Agar Dilution

**REPORT NUMBER:** 10-08-2020-Mt. Sinai 1

**QUOTE NUMBER:** 08-24-2020-Mt. Sinai 1

**DATE:** October 12, 2020

**AUTHORS:**

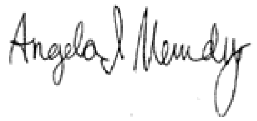

Angela Mendez
R&D Technician

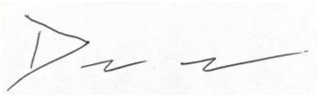

David Hufnagel, Ph.D.
Senior Scientist

**STUDY DIRECTOR:**

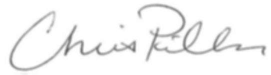

Chris Pillar, Ph. D.
Director of Science and Operations

**SITE:** Micromyx, LLC
4717 Campus Drive
Kalamazoo, MI, USA 49008

#### INTRODUCTION

Mt. Sinai is currently evaluating a live biotherapeutic product (LBP), which consists of 16 isolates of anaerobic bacteria. Understanding the antibiotic-resistance profile of these organism is integral to LBP development. In this study, we determined the antibiograms of the 16 component isolates of the LBP and 2 supplemental isolates by anaerobic agar dilution. Agar dilution susceptibility testing was conducted following guidelines recommended by the Clinical and Laboratory Standards Institute (CLSI; 1, 2).

#### MATERIALS AND METHODS

##### Test Compounds

Antibiotics were provided by Micromyx. All stock solutions were allowed to stand for at least 1 hr prior to use to auto-sterilize. Leftover stock solutions of drugs were aliquoted and stored at -80°C. Suppliers, catalog/lot numbers, solvents, stock concentrations and testing ranges were as follows.

| Test Agents | Supplier | Catalog Number | Lot Number | Solvent/Diluent | Testing Range (µg/mL) |
|---|---|---|---|---|---|
| Vancomycin | Sigma | V2002 | 080M1341V | $H_2O$ / $H_2O$ | 0.12-32 |
| Metronidazole | Sigma | M3761 | 09560693 | DMSO / $H_2O$ | 0.015-16 |
| Tigecycline | USP | 1667643 | R09410 | $H_2O$ / $H_2O$ | 0.015-8 |
| Ampicillin | Sigma | A9518 | 127M4894V | PBS pH-8 / PBS pH-6 | 0.015-64 |
| Sulbactam | USP | 1623670 | R03980 | $H_2O$ / $H_2O$ | 0.008-32 |
| Amoxicillin | Sigma | A8523 | 100M0828V | PBS pH-6 / PBS pH-6 | 0.015-8 |
| Clavulanate | Sigma | 33454 | STBH5214 | PBS pH-6 / PBS pH-6 | 0.008-4 |
| Meropenem | USP | 1392454 | JOK434 | $H_2O$ / $H_2O$ | 0.015-16 |
| Piperacillin | Sigma | P8396 | SLBC6856V | $H_2O$ / $H_2O$ | 0.06-32 |
| Tazobactam | USP | 1643383 | R080X0 | $H_2O$ / $H_2O$ | 4 |
| Clindamycin | Selleck Chem | 52457 | 01 | $H_2O$ / $H_2O$ | 0.03-8 |
| Ceftriaxone | Sigma | C5793 | 077K0598 | $H_2O$ / $H_2O$ | 0.05-256 |
| Moxifloxacin | Chem Pacific | 35185 | CHPCO62915MHV | $H_2O$ / $H_2O$ | 0.03-16 |

##### Test Isolates

Test organisms consisted of reference quality control (QC) strains from the American Type Culture Collection (ATCC; Manassas, VA) and the test isolates sent from Mt. Sinai. Prior to testing, the isolates were sub-cultured from frozen glycerol stocks onto supplemented Brucella agar (SBA; Becton Dickenson [BD]/BBL; Lot No.0134828; Sparks, MD) in the Bactron II anaerobe chamber (Shel Lab, Cornelius, WA). All organisms were grown in the anaerobe chamber at 35ºC for 48 hr.

On each day of testing, *Bacteroides fragilis* ATCC 25285, *Bacteroides thetaiotaomicron* ATCC 29741, *Clostridioides difficile* ATCC 700057, and *Eggerthella lenta* ATCC 43055 were included for purposes of QC.

**Test Media**

The test medium for this study was supplemented Brucella agar (SBA) as recommended by guidelines established by the CLSI (1, 2) for the testing of anaerobes.

**Agar Dilution MIC Methodology**

Each test agent was mixed with molten (50-55ºC) Brucella agar (BD; Lot No. 9128845) supplemented with hemin (Sigma; Lot No. SLBP5720V; St. Louis, MO), Vitamin K (Sigma; Lot No. MKCB9432), and 5% sheep blood (Hemostat; Lot No. 529190; Dixon, CA) in accordance with CLSI guidelines (1, 2). Vancomycin, metronidazole, tigecycline, meropenem, clindamycin, ceftriaxone, and moxifloxacin were added to the media in a ratio of 2 mL 10X test agent to 18 mL agar. Ampicillin and sulbactam were added to the media in a ratio of 1 mL 20X test agent to 18 mL agar in order to create plates with ampicillin/sulbactam (2:1 fixed ratio w/w). Amoxicillin and clavulanate were added to the media in a ratio of 1 mL 20X test agent to 18 mL agar in order to create plates with amoxicillin/clavulanate (2:1 fixed ratio w/w). Piperacillin was added to the media in a ratio of 2 mL 10X test agent to 18 mL agar containing 4 µg/mL tazobactam. The growth control plates used in between drug plates contained 2 mL of water or phosphate buffer. An additional growth control with 4 µg/mL tazobactam was also evaluated. The vancomycin, metronidazole, tigecycline, ampicillin/sulbactam, clindamycin, ceftriaxone, and moxifloxacin plates were made the day before and used within 24 hrs. The other plates were made on the day of testing.

The agar dilution plates were allowed to pre-reduce in the anaerobe chamber for 1-2 hr prior to inoculation. In the anaerobe chamber, each isolate was suspended to the equivalent of a 0.5 McFarland standard in saline (NERL Blood Bank Saline 0.9%; Thermo Scientific; Lot No. 910360; Middletown, VA). Each bacterial cell suspension was then transferred to wells in a stainless-steel replicator block. The prongs on the replicator deliver approximately 1-2 µl of inoculum to an agar surface. The resulting inoculum spots contained approximately $10^5$ CFU/spot.

Each agar plate containing either test compound or no drug (control) was stamped with the replicator in the anaerobe chamber. All plates were placed on the anaerobe chamber surface with the agar surface up to allow for the inoculum to soak into the agar. The plates were inverted and incubated anaerobically at 35°C for 48 hr, after which they were removed from the chamber and inspected for growth. The MIC was defined as the lowest test agent concentration that substantially inhibited bacterial growth on the agar surface.

For enumeration of the inoculum suspensions, a 0.3 mL aliquot was removed from each 0.5 standard McFarland, and pipetted into column 1 of a 96-well plate that had 270 µL of chilled saline in columns 2-12. Serial dilutions of the plate were performed on the Biomek 2000. During track dilution plating, a 10 µl aliquot of each dilution was spotted in duplicate across the top of a

TSA plate. The plate was then tilted at a 45 – 90° angle to allow the 10 µL aliquot to track across the agar surface to the opposite side of the plate. The plates were laid flat, allowed to dry at room temperature, then inverted and incubated at 35°C for approximately 48 hr (in some cases prolonged incubation was required). Colonies were then counted to determine the viable count in CFU/mL.

#### RESULTS AND DISCUSSION

The susceptibility of a total of 16 isolates that make up an LBP from Mt. Sinai along with a couple of extra isolates were evaluated by anaerobic agar dilution for 10 antibiotics relevant to treatment of anaerobic infections. All organisms were enumerated and identification was attempted by MALDI. All 4 QC organisms tested on two separate days were within established CLSI QC ranges (1,2), except for tigecycline when testing against *C. difficile* ATCC 700057 and tazobactam/piperacillin when testing against *Eggerthella lenta* ATCC 43055 (**Appendix 1**). In both instances, a minimum of two other QC organisms were within QC, thus validating the assay (**Appendix 1**).

**Table 1** shows the MIC values of the 10 antibiotics generated against the LBP test strains by anaerobic agar dilution. In select instances (*Bacteroides cellulosilyticus* MTCO1.15, *Odoribacter splanchnicus* MTCO1.14, *Coprococcus comes* MTCO1.10), more than one morphology was noted during primary plating of the provided frozen stocks. Both morphologies were tested and the MIC results were identical or within 2-fold for each morphology. MALDI ID and spectra when available were also consistent across morphologies; MALDI spectra has been provided to the Sponsor for all test isolates.

Overall, among the LBP isolates, all were susceptible to ampicillin/sulbactam, amoxicillin/clavulanate, and meropenem. All but a single isolate were susceptible to metronidazole (*Bifidobacterium adolescentis* MTCO1.08 was resistant), tigecycline (*Bacteroides caccae* MTCO1.12 was resistant), and piperacillin/tazobactam (*Parabacteroides distasonis* MTCO1.07 was either intermediate or resistant). In contrast, intermediate-susceptibility or resistance among the LBP isolates to clindamycin, ceftriaxone, and moxifloxacin was more common. As expected, vancomycin activity was restricted to Gram-positive anaerobes. Of note, MIC results for the two supplemental isolates can be found in **Appendix 2**.

No spectrum was returned by MALDI for either morphology of *Coprococcus comes* MTCO1.10, and no ID was able to be determined from the specta of both *Eubacterium rectale* MTCO1.11 and *Dorea longicatena* MTCO1.13. These results were as expected as the Bruker Biotyper MALDI ID database does not have spectra for these species (personal communication from the Sponsor). The IDs of all the remaining LBP isolates were confirmed by MALDI. Of note, MALDI ID also failed to confirm QC isolate *Eggerthella lenta* ATCC 43055 and supplemental isolate *Collinsella aerofaciens* MTCO1.EXTRAB;

Enumeration data (**Table 1**) demonstrated that an inoculum of approximately 1-5 x $10^5$ CFU/spot was achieved for nearly all isolates excluding *Bifidobacterium longum* MTCO1.03 and *Dorea longicatena* MTCO1.13 where the inoculum density was low and select strains were counts were not able to be determined (*Coprococcus comes* MTCO1.10 and *Eubacterium rectale*

MTCO1.11). However, growth control spots in these instances were sufficient to interpret the MIC in most cases; in select cases, no growth was seen on the no drug control plate for the indicated test agent and as a result MIC values were not determined (ND) (**Table 1**). It is possible that the viability of these strains is compromised over time when suspended in normal saline, as control plates for antibiotics at the end of the run had a higher frequency of being ND for these three strains. This could also explain the inability to enumerate, since enumerations were performed after the last agar dilution plate was stamped.

In summary, all LBP isolates were susceptible to meropenem, ampicillin/sulbactam, amoxicillin/clavulanate and all but one isolate was susceptible to tigecycline, metronidazole, and piperacillin/tazobactam. The evaluated organisms had much higher rates of intermediate and resistance phenotypes when tested against clindamycin, ceftriaxone, and moxifloxacin.

#### REFERENCES

1.) Clinical and Laboratory Standards Institute (CLSI). *Methods for Antimicrobial Susceptibility Testing of Anaerobic Bacteria; 9th ed*. CLSI standard M11. CLSI, 940 West Valley Road, Suite 1400, Wayne, Pennsylvania 19087-1898 USA, 2018.

2.) CLSI. *Performance Standards for Antimicrobial Susceptibility Testing; 30th ed*. CLSI supplement M100. CLSI, 940 West Valley Road, Suite 1400, Wayne, Pennsylvania 19087-1898 USA, 2020.

**Table 1.** Agar dilution MIC values against isolates from the live bacterial therapeutic

| Organism | | CFU/spot inoculated | MIC (µg/mL) | | | | | | | | | |
|---|---|---|---|---|---|---|---|---|---|---|---|---|
| | | | VAN | MTZ | TGC | SAM | AMC | MEM | TZP | CLI | CRO | MOX |
| Gram-negative | *Bacteroides uniformis* MTCO1.01 | 7.30E+05 | >32 | 1 | 0.25 | 1/0.5 | 1/0.5 | 0.12 | 0.25/4 | 2 | 64 | 2 |
| | *Bacteroides ovatus* MTCO1.02 | 7.90E+05 | >32 | 1 | 0.25 | 1/0.5 | 0.5/0.25 | 0.25 | 4/4 | 2 | 64 | 2 |
| | *Bacteroides thetaiotaomicron* MTCO1.04 | 9.60E+05 | >32 | 1 | 2 | 1/0.5 | 1/0.5 | 0.25 | 4/4 | 4 | 64 | 1 |
| | *Bacteroides vulgatus* MTCO1.05 | 6.50E+05 | 32 | 0.5 | 2 | 4/2 | 0.5/0.25 | 0.25 | 4/4 | >8 | 16 | >16 |
| | *Bacteroides caccae* MTCO1.12 | 7.60E+05 | >32 | 1 | >8 | 2/1 | 0.5/0.25 | 0.12 | 4/4 | 1 | 16 | 2 |
| | *Bacteroides cellulosilyticus* (Round) MTCO1.15 | 7.00E+05 | >32 | 1 | 2 | 4/2 | 2/1 | 0.25 | 8/4 | 2 | 64 | 4 |
| | *Bacteroides cellulosilyticus* (Flat) MTCO1.15 | 5.80E+05 | >32 | 1 | 2 | 4/2 | 2/1 | 0.25 | 8/4 | 2 | 64 | 4 |
| | *Parabacteroides distasonis* MTCO1.07 | 1.00E+06 | >32 | 1 | 1 | 8/4 | 4/2 | 0.5 | >32/4 | 0.25 | 32 | 4 |

| Organism | | CFU/spot inoculated | MIC (µg/mL) | | | | | | | | | |
|---|---|---|---|---|---|---|---|---|---|---|---|---|
| | | | VAN | MTZ | TGC | SAM | AMC | MEM | TZP | CLI | CRO | MOX |
| Gram-negative | *Parabacteroides merdae* MTCO1.09 | 2.30E+06 | >32 | 1 | 1 | 2/1 | 2/1 | 0.25 | 16/4 | 2 | 16 | 0.25 |
| | *Odoribacter splanchnicus* (Large) MTCO1.14 | 8.40E+05 | >32 | 0.5 | 0.12 | 4/2 | 2/1 | 0.12 | ≤0.06/4 | >8 | 16 | 2 |
| | *Odoribacter splanchnicus* (Small) MTCO1.14 | 9.30E+05 | >32 | 0.5 | 0.12 | 4/2 | 2/1 | 0.12 | ≤0.06/4 | >8 | 16 | 1 |
| Gram-positive | *Bifidobacterium longum* MTCO1.03 | 6.60E+02 | 1 | 8 | 0.5 | 0.5/.25 | 0.5/0.25 | 0.06 | 0.5/4 | 0.06 | 4 | 0.5 |
| | *Bifidobacterium adolescentis* MTCO1.08 | 2.10E+05 | 1 | >16 | 0.5 | 1/0.5 | 0.25/0.12 | 0.12 | ≤0.06/4 | ≤0.06 | ≤0.5 | 8 |
| | *Bifidobacterium pseudocatenulatum* MTCO1.16 | 4.10E+05 | 0.5 | 2 | 1 | 0.12/0.06 | 0.06/0.03 | 0.12 | 0.5/4 | ≤0.03 | ≤0.5 | 4 |
| | *Collinsella aerofaciens* MTCO1.06 | 4.40E+05 | 2 | 2 | 0.25 | 1/0.5 | 0.5/0.25 | 0.25 | 4/4 | ≤0.03 | 16 | 0.5 |
| | *Coprococcus comes* (Small) MTCO1.10 | ND | 0.5 | 0.5 | 0.06 | 0.03/0.015 | ≤0.015/0.008 | 0.03 | 0.12/4 | ≤0.03 | ≤0.5 | 8 |
| | *Coprococcus comes* (Large) MTCO1.10 | ND | 0.5 | 0.5 | 0.12 | 0.03/0.015 | ≤0.015/0.008 | 0.06 | 0.12/4 | ≤0.03 | ≤0.5 | 8 |

| Organism | | CFU/spot inoculated | MIC (µg/mL) | | | | | | | | | |
|---|---|---|---|---|---|---|---|---|---|---|---|---|
| | | | VAN | MTZ | TGC | SAM | AMC | MEM | TZP | CLI | CRO | MOX |
| Gram-positive | *Eubacterium rectale* MTCO1.11 | ND | 0.5 | 0.06 | 0.06 | ND | ND | ND | ND | ≤0.06 | ND | 4 |
| | *Dorea longicatena* MTCO1.13 | 7.30E+02 | 0.5 | 0.06 | 0.25 | 0.5/0.25 | 0.25/0.12 | 0.5 | 1/4 | 4 | 2 | 8 |

Susceptible isolates are shaded green, intermediate isolates are shaded orange, and resistant isolates are shaded red (yellow means this isolate is either intermediate or resistant); Susceptible, intermediate, and resistant breakpoint categorizations are from CLSI guidelines (tigecycline breakpoints were obtained from the FDA labeling); VAN, vancomycin; MTZ, metronidazole; TGC, tigecycline; SAM, ampicillin/sulbactam; AMC, amoxicillin/clavulanate; MEM, meropenem; TZP, tazobactam/piperacillin; CLI, clindamycin; CRO, ceftriaxone; MOX, moxifloxacin; ND, not determined due to no growth on the enumeration plate or corresponding growth control plate

**Appendix 1.** QC organisms MIC values on multiple run days

| Organism | Run | CFU/spot inoculated | MIC (µg/mL) | | | | | | | | | |
|---|---|---|---|---|---|---|---|---|---|---|---|---|
| | | | VAN | MTZ | TGC | SAM | AMC | MEM | TZP | CLI | CRO | MOX |
| *Bacteroides fragilis* ATCC 25285 | 1 | 6.50E+05 | >32 | 0.5 (0.25-1) | 0.25 (0.12-1) | 1/0.5 (0.5/0.25-2/1) | 1/0.5 (0.25/0.12-1/0.5) | 0.12 (0.03-0.25) | 0.25/4 (0.12/4-0.5/4) | 1 (0.5-2) | 64 (32-128) | 0.25 (0.12-0.5) |
| | 2 | 2.35E+05 | >32 | 0.5 (0.25-1) | 0.5 (0.12-1) | 2/1 (0.5/0.25-2/1) | 1/0.5 (0.25/0.12-1/0.5) | 0.12 (0.03-0.25) | 0.25/4 (0.12/4-0.5/4) | 1 (0.5-2) | 64 (32-128) | 0.25 (0.12-0.5) |
| *Bacteroides thetaiotaomicron* ATCC 29741 | 1 | 3.30E+05 | >32 | 1 (0.5-2) | 1 (0.5-2) | 1/0.5 (0.5/0.25-2/1) | 0.5/0.25 (0.5/0.25-2/1) | 0.12 (0.12-0.5) | 4/4 (4/4-16/4) | 4 (2-8) | 64 (64-256) | 2 (1-4) |
| | 2 | 5.80E+04 | >32 | 1 (0.5-2) | 2 (0.5-2) | 1/0.5 (0.5/0.25-2/1) | 1/0.5 (0.5/0.25-2/1) | 0.12 (0.12-0.5) | 2/4 (4/4-16/4) | 4 (2-8) | 64 (64-256) | 2 (1-4) |
| *Clostridiodes difficile* ATCC 700057 | 1 | 1.40E+04 | 2 (0.5-4) | 0.5 (0.12-0.5) | 1 (0.06-0.5) | 2/1 (0.5/0.25-4/2) | 1/0.5 (0.25/0.12/1/0.5) | 2 (0.5-4) | 16/4 (4/4-16/4) | 4 (2-8) | 32 | 2 (1-4) |
| | 2 | 9.00E+04 | 2 (0.5-4) | 0.25 (0.12-0.5) | 1 (0.06-0.5) | 2/1 (0.5/0.25-4/2) | 1/0.5 (0.25/0.12/1/0.5) | 2 (0.5-4) | 16/4 (4/4-16/4) | 8 (2-8) | 64 | 2 (1-4) |
| *Eggerthella lenta* ATCC 43055 | 1 | 8.60E+05 | 2 | 0.5 | 0.12 (0.06-0.5) | 1/0.5 (0.25/0.12-2/1) | 1/0.5 | 0.5 (0.12-1) | 32/4 (4/4-16/4) | 0.12 (0.06-0.25) | 256 | 0.25 (0.12-0.5) |
| | 2 | 2.00E+05 | 2 | 0.5 | 0.25 (0.06-0.5) | 2/1 (0.25/0.12-2/1) | 2/1 | 0.5 (0.12-1) | 32/4 (4/4-16/4) | 0.12 (0.06-0.25) | 256 | 0.25 (0.12-0.5) |

Accepted CLSI QC ranges shown in parentheses; VAN, vancomycin; MTZ, metronidazole; TGC, tigecycline; SAM, ampicillin/sulbactam; AMC, amoxicillin/clavulanate; MEM, meropenem; TZP, tazobactam/piperacillin; CLI, clindamycin; CRO, ceftriaxone; MOX, moxifloxacin

**Appendix 2.** Agar dilution MIC values against isolates from the live bacterial therapeutic

| Organism | Enumeration | MIC (µg/mL) | | | | | | | | | |
|---|---|---|---|---|---|---|---|---|---|---|---|
| | | VAN | MTZ | TGC | SAM | AMC | MEM | TZP | CLI | CRO | MOX |
| *Bacteroides ovatus* MTCO1.EXTRAA | 4.00E+08 | >32 | 1 | 0.25 | 16/8 | >8/4 | 0.5 | 8/4 | 0.5 | >256 | 4 |
| *Collinsella aerofaciens* MTCO1.EXTRAB | 5.80E+07 | 0.5 | 2 | 0.25 | 1/0.5 | 0.5/0.25 | 0.25 | 4/4 | ≤0.03 | 16 | 0.5 |

Susceptible isolates are shaded green, intermediate isolates are shaded orange, and resistant isolates are shaded red; Susceptible, intermediate, and resistant breakpoint categorizations are from CLSI guidelines (tigecycline breakpoints were obtained from the FDA labeling); VAN, vancomycin; MTZ, metronidazole; TGC, tigecycline; SAM, ampicillin/sulbactam; AMC, amoxicillin/clavulanate; MEM, meropenem; TZP, tazobactam/piperacillin; CLI, clindamycin; CRO, ceftriaxone; MOX, moxifloxacin

### Appendix 4 – MTC01 USP<61> Reports

### FOCUS Labs single strain USP<61> Method suitability reports

FOCUS LABORATORIES

177 North Commerce Way
Bethlehem, PA 18017
(610) 866-7272
www.FOCUS-lab.com

### Test Report

**Client:** Icahn School of Medicine at Mount Sinai
Hess Center for Science and Medicine
1470 Madison Avenue, 8th FL.
New York, NY 10029

**Contact:** Lukas Bethlehem, PhD

212-824-9088

**Study Number:** 211635-1-R1

**Test Start Date:** 25-May-2021

**Test End Date:** 01-Jun-2021

**Testing Guideline:** USP <61>

| Sample Name |
|---|
| MTC01.01_*B. uniformis* |

**Test Results:**

Table 1: USP <61>

| Test | Result | Specification | Pass/Fail |
|---|---|---|---|
| Aerobic Plate Count | ≤ 100 cfu/mL | ≤ 1000 cfu/mL | Pass |
| Yeast and Mold Count | ≤ 100 cfu/mL | ≤ 100 cfu/mL | Pass |

Table 2: USP <61> Method Suitability Results

| Organism | Control count | Test Count | % Recovery | Pass/Fail |
|---|---|---|---|---|
| *S. aureus* ATCC 6538 | 94 cfu | 92 cfu | 98% | Pass |
| *P. aeruginosa* ATCC 9027 | 57 cfu | 82 cfu | 144% | Pass |
| *B. subtilis* ATCC 6633 | 48 cfu | 65 cfu | 135% | Pass |
| *C. albicans* ATCC 10231 30°C – 35°C | 31 cfu | 28 cfu | 90% | Pass |
| *A. brasiliensis* ATCC 16404 30°C – 35°C | 15 cfu | 27 cfu | 180% | Pass |
| *C. albicans* ATCC 10231 20°C – 25°C | 25 cfu | 28 cfu | 112% | Pass |
| *A. brasiliensis* ATCC 16404 20°C – 25°C | 20 cfu | 28 cfu | 140% | Pass |

**Conclusion:** A 1:1 dilution in TSB+ is a suitable method for MTC01.01_*B. uniformis.*

QA Reviewed / Approved by/date: [signature] 16 Jul 2021

FOCUS Laboratories is ISO/IEC 17025:2017 accredited by PJLA in the field of testing, Accreditation #77499

†indicated specific test is outside the scope of ISO accreditation, however FOCUS is an FDA registered cGMP laboratory. This document is issued under the company's terms and conditions which were agreed upon during sample submission. FOCUS Laboratories' liability regarding this analysis is limited to the cost of the tests associated with this Test Report. The results are the property of the client listed above and FOCUS Laboratories. These results only relate to items tested above and apply to the samples as received. This report shall not be reproduced except in full without approval of FOCUS Laboratories.

FOCUS LABORATORIES

177 North Commerce Way
Bethlehem, PA 18017
(610) 866-7272
www.FOCUS-lab.com

### Test Report

**Client:** Icahn School of Medicine at Mount Sinai
Hess Center for Science and Medicine
1470 Madison Avenue, 8th FL.
New York, NY 10029

**Contact:** Lukas Bethlehem, PhD

212-824-9088

**Study Number:** 211635-2-R1

**Test Start Date:** 25-May-2021
**Test End Date:** 01-Jun-2021

**Testing Guideline:** USP <61>

| Sample Name |
|---|
| MTC01.02_ *B. ovatus* |

**Test Results:**

Table 1: USP <61>

| Test | Result | Specification | Pass/Fail |
|---|---|---|---|
| Aerobic Plate Count | ≤ 100 cfu/mL | ≤ 1000 cfu/mL | Pass |
| Yeast and Mold Count | ≤ 100 cfu/mL | ≤ 100 cfu/mL | Pass |

Table 2: USP <61> Method Suitability Results

| Organism | Control count | Test Count | % Recovery | Pass/Fail |
|---|---|---|---|---|
| *S. aureus* ATCC 6538 | 94 cfu | 99 cfu | 105% | Pass |
| *P. aeruginosa* ATCC 9027 | 57 cfu | 62 cfu | 109% | Pass |
| *B. subtilis* ATCC 6633 | 48 cfu | 61 cfu | 127% | Pass |
| *C. albicans* ATCC 10231 30°C – 35°C | 31 cfu | 29 cfu | 94% | Pass |
| *A. brasiliensis* ATCC 16404 30°C – 35°C | 15 cfu | 13 cfu | 87% | Pass |
| *C. albicans* ATCC 10231 20°C – 25°C | 25 cfu | 35 cfu | 140% | Pass |
| *A. brasiliensis* ATCC 16404 20°C – 25°C | 20 cfu | 24 cfu | 120% | Pass |

**Conclusion:** A 1:1 dilution in TSB+ is a suitable method for MTC01.02_ *B. ovatus*.

QA Reviewed / Approved by/date: [signature] 16 Jul 2021

FOCUS Laboratories is ISO/IEC 17025:2017 accredited by PJLA in the field of testing, Accreditation #77499

†Indicated specific test is outside the scope of ISO accreditation, however FOCUS is an FDA registered cGMP laboratory. This document is issued under the company's terms and conditions which were agreed upon during sample submission. FOCUS Laboratories' liability regarding this analysis is limited to the cost of the tests associated with this Test Report. The results are the property of the client listed above and FOCUS Laboratories. These results only relate to items tested above and apply to the samples as received. This report shall not be reproduced except in full without approval of FOCUS Laboratories.

FOCUS LABORATORIES

177 North Commerce Way
Bethlehem, PA 18017
(610) 866-7272
www.FOCUS-lab.com

### Test Report

**Client:** Icahn School of Medicine at Mount Sinai
Hess Center for Science and Medicine
1470 Madison Avenue, 8th FL.
New York, NY 10029

**Contact:** Lukas Bethlehem, PhD

212-824-9088

**Study Number:** 211635-3-R1

**Test Start Date:** 25-May-2021
**Test End Date:** 01-Jun-2021

**Testing Guideline:** USP <61>

| Sample Name |
|---|
| MTC01.03_*B. longum* |

**Test Results:**

Table 1: USP <61>

| Test | Result | Specification | Pass/Fail |
|---|---|---|---|
| Aerobic Plate Count | ≤ 100 cfu/mL | ≤ 1000 cfu/mL | Pass |
| Yeast and Mold Count | ≤ 100 cfu/mL | ≤ 100 cfu/mL | Pass |

Table 2: USP <61> Method Suitability Results

| Organism | Control count | Test Count | % Recovery | Pass/Fail |
|---|---|---|---|---|
| *S. aureus* ATCC 6538 | 94 cfu | 87 cfu | 93% | Pass |
| *P. aeruginosa* ATCC 9027 | 57 cfu | 58 cfu | 102% | Pass |
| *B. subtilis* ATCC 6633 | 48 cfu | 46 cfu | 96% | Pass |
| *C. albicans* ATCC 10231 30°C – 35°C | 31 cfu | 30 cfu | 97% | Pass |
| *A. brasiliensis* ATCC 16404 30°C – 35°C | 15 cfu | 17 cfu | 113% | Pass |
| *C. albicans* ATCC 10231 20°C – 25°C | 25 cfu | 29 cfu | 116% | Pass |
| *A. brasiliensis* ATCC 16404 20°C – 25°C | 20 cfu | 20 cfu | 100% | Pass |

**Conclusion:** A 1:1 dilution in TSB+ is a suitable method for MTC01.03_*B. longum*.

QA Reviewed / Approved by/date: [signature] 16 Jul 2021

FOCUS Laboratories is ISO/IEC 17025:2017 accredited by PJLA in the field of testing, Accreditation #77499
†Indicated specific test is outside the scope of ISO accreditation, however FOCUS is an FDA registered cGMP laboratory. This document is issued under the company's terms and conditions which were agreed upon during sample submission. FOCUS Laboratories' liability regarding this analysis is limited to the cost of the tests associated with this Test Report. The results are the property of the client listed above and FOCUS Laboratories. These results only relate to items tested above and apply to the samples as received. This report shall not be reproduced except in full without approval of FOCUS Laboratories.

FOCUS LABORATORIES

177 North Commerce Way
Bethlehem, PA 18017
(610) 866-7272
www.FOCUS-lab.com

### Test Report

**Client:** Icahn School of Medicine at Mount Sinai
Hess Center for Science and Medicine
1470 Madison Avenue, 8th FL.
New York, NY 10029

**Contact:** Lukas Bethlehem, PhD

212-824-9088

**Study Number:** 211635-4-R1

**Test Start Date:** 25-May-2021

**Test End Date:** 01-Jun-2021

**Testing Guideline:** USP <61>

| Sample Name |
|---|
| MTC01.04_*B. thetaiotaomicron* |

**Test Results:**

Table 1: USP <61>

| Test | Result | Specification | Pass/Fail |
|---|---|---|---|
| Aerobic Plate Count | ≤ 100 cfu/mL | ≤ 1000 cfu/mL | Pass |
| Yeast and Mold Count | ≤ 100 cfu/mL | ≤ 100 cfu/mL | Pass |

Table 2: USP <61> Method Suitability Results

| Organism | Control count | Test Count | % Recovery | Pass/Fail |
|---|---|---|---|---|
| *S. aureus* ATCC 6538 | 94 cfu | 76 cfu | 81% | Pass |
| *P. aeruginosa* ATCC 9027 | 57 cfu | 65 cfu | 114% | Pass |
| *B. subtilis* ATCC 6633 | 48 cfu | 65 cfu | 135% | Pass |
| *C. albicans* ATCC 10231 30°C – 35°C | 31 cfu | 30 cfu | 97% | Pass |
| *A. brasiliensis* ATCC 16404 30°C – 35°C | 15 cfu | 23 cfu | 153% | Pass |
| *C. albicans* ATCC 10231 20°C – 25°C | 25 cfu | 33 cfu | 132% | Pass |
| *A. brasiliensis* ATCC 16404 20°C – 25°C | 20 cfu | 14 cfu | 70% | Pass |

**Conclusion:** A 1:1 dilution in TSB+ is a suitable method for MTC01.04_*B. thetaiotaomicron*

QA Reviewed / Approved by/date: [signature] 16 Jul 2021

FOCUS Laboratories is ISO/IEC 17025:2017 accredited by PJLA in the field of testing, Accreditation #77499
†indicated specific test is outside the scope of ISO accreditation, however FOCUS is an FDA registered cGMP laboratory. This document is issued under the company's terms and conditions which were agreed upon during sample submission. FOCUS Laboratories' liability regarding this analysis is limited to the cost of the tests associated with this Test Report. The results are the property of the client listed above and FOCUS Laboratories. These results only relate to items tested above and apply to the samples as received. This report shall not be reproduced except in full without approval of FOCUS Laboratories.

FOCUS LABORATORIES

177 North Commerce Way
Bethlehem, PA 18017
(610) 866-7272
www.FOCUS-lab.com

### Test Report

**Client:** Icahn School of Medicine at Mount Sinai
Hess Center for Science and Medicine
1470 Madison Avenue, 8th FL.
New York, NY 10029

**Contact:** Lukas Bethlehem, PhD

212-824-9088

**Study Number:** 211635-5-R1

**Test Start Date:** 25-May-2021
**Test End Date:** 01-Jun-2021

**Testing Guideline:** USP <61>

| Sample Name |
|---|
| MTC01.05_*B. vulgatus* |

**Test Results:**

Table 1: USP <61>

| Test | Result | Specification | Pass/Fail |
|---|---|---|---|
| Aerobic Plate Count | ≤ 100 cfu/mL | ≤ 1000 cfu/mL | Pass |
| Yeast and Mold Count | ≤ 100 cfu/mL | ≤ 100 cfu/mL | Pass |

Table 2: USP <61> Method Suitability Results

| Organism | Control count | Test Count | % Recovery | Pass/Fail |
|---|---|---|---|---|
| *S. aureus* ATCC 6538 | 94 cfu | 90 cfu | 96% | Pass |
| *P. aeruginosa* ATCC 9027 | 57 cfu | 73 cfu | 128% | Pass |
| *B. subtilis* ATCC 6633 | 48 cfu | 56 cfu | 117% | Pass |
| *C. albicans* ATCC 10231 30°C – 35°C | 31 cfu | 30 cfu | 97% | Pass |
| *A. brasiliensis* ATCC 16404 30°C – 35°C | 15 cfu | 10 cfu | 67% | Pass |
| *C. albicans* ATCC 10231 20°C – 25°C | 25 cfu | 23 cfu | 92% | Pass |
| *A. brasiliensis* ATCC 16404 20°C – 25°C | 20 cfu | 25 cfu | 125% | Pass |

**Conclusion:** A 1:1 dilution in TSB+ is a suitable method for MTC01.05_*B. vulgatus*.

QA Reviewed / Approved by/date: Jim Bepstein 16Jul2021

FOCUS Laboratories is ISO/IEC 17025:2017 accredited by PJLA in the field of testing, Accreditation #77499
†Indicated specific test is outside the scope of ISO accreditation, however FOCUS is an FDA registered cGMP laboratory. This document is issued under the company's terms and conditions which were agreed upon during sample submission. FOCUS Laboratories' liability regarding this analysis is limited to the cost of the tests associated with this Test Report. The results are the property of the client listed above and FOCUS Laboratories. These results only relate to items tested above and apply to the samples as received. This report shall not be reproduced except in full without approval of FOCUS Laboratories.

FOCUS LABORATORIES

177 North Commerce Way
Bethlehem, PA 18017
(610) 866-7272
www.FOCUS-lab.com

### Test Report

**Client:** Icahn School of Medicine at Mount Sinai
Hess Center for Science and Medicine
1470 Madison Avenue, 8th FL.
New York, NY 10029

**Contact:** Lukas Bethlehem, PhD

212-824-9088

**Study Number:** 211635-6-R1

**Test Start Date:** 25-May-2021
**Test End Date:** 01-Jun-2021

**Testing Guideline:** USP <61>

| Sample Name |
|---|
| MTC01.06_*C. aerofaciens* |

**Test Results:**

Table 1: USP <61>

| Test | Result | Specification | Pass/Fail |
|---|---|---|---|
| Aerobic Plate Count | ≤ 100 cfu/mL | ≤ 1000 cfu/mL | Pass |
| Yeast and Mold Count | ≤ 100 cfu/mL | ≤ 100 cfu/mL | Pass |

Table 2: USP <61> Method Suitability Results

| Organism | Control count | Test Count | % Recovery | Pass/Fail |
|---|---|---|---|---|
| *S. aureus* ATCC 6538 | 94 cfu | 88 cfu | 94% | Pass |
| *P. aeruginosa* ATCC 9027 | 57 cfu | 78 cfu | 137% | Pass |
| *B. subtilis* ATCC 6633 | 48 cfu | 50 cfu | 104% | Pass |
| *C. albicans* ATCC 10231 30°C – 35°C | 31 cfu | 31 cfu | 100% | Pass |
| *A. brasiliensis* ATCC 16404 30°C – 35°C | 15 cfu | 13 cfu | 87% | Pass |
| *C. albicans* ATCC 10231 20°C – 25°C | 25 cfu | 34 cfu | 136% | Pass |
| *A. brasiliensis* ATCC 16404 20°C – 25°C | 20 cfu | 27 cfu | 135% | Pass |

**Conclusion:** A 1:1 dilution in TSB+ is a suitable method for MTC01.06_*C. aerofaciens*.

QA Reviewed / Approved by/date: [signature] 16 Jul 2021

FOCUS Laboratories is ISO/IEC 17025:2017 accredited by PJLA in the field of testing, Accreditation #77499
†indicated specific test is outside the scope of ISO accreditation, however FOCUS is an FDA registered cGMP laboratory. This document is issued under the company's terms and conditions which were agreed upon during sample submission. FOCUS Laboratories' liability regarding this analysis is limited to the cost of the tests associated with this Test Report. The results are the property of the client listed above and FOCUS Laboratories. These results only relate to items tested above and apply to the samples as received. This report shall not be reproduced except in full without approval of FOCUS Laboratories.

FOCUS LABORATORIES

177 North Commerce Way
Bethlehem, PA 18017
(610) 866-7272
www.FOCUS-lab.com

### Test Report

**Client:** Icahn School of Medicine at Mount Sinai
Hess Center for Science and Medicine
1470 Madison Avenue, 8th FL.
New York, NY 10029

**Contact:** Lukas Bethlehem, PhD

212-824-9088

**Study Number:** 211635-7-R1

**Test Start Date:** 25-May-2021

**Test End Date:** 01-Jun-2021

**Testing Guideline:** USP <61>

| Sample Name |
|---|
| MTC01.07_*P. distasonis* |

**Test Results:**

Table 1: USP <61>

| Test | Result | Specification | Pass/Fail |
|---|---|---|---|
| Aerobic Plate Count | ≤ 100 cfu/mL | ≤ 1000 cfu/mL | Pass |
| Yeast and Mold Count | ≤ 100 cfu/mL | ≤ 100 cfu/mL | Pass |

Table 2: USP <61> Method Suitability Results

| Organism | Control count | Test Count | % Recovery | Pass/Fail |
|---|---|---|---|---|
| *S. aureus* ATCC 6538 | 94 cfu | 103 cfu | 110% | Pass |
| *P. aeruginosa* ATCC 9027 | 57 cfu | 64 cfu | 112% | Pass |
| *B. subtilis* ATCC 6633 | 48 cfu | 61 cfu | 127% | Pass |
| *C. albicans* ATCC 10231 30°C – 35°C | 31 cfu | 30 cfu | 97% | Pass |
| *A. brasiliensis* ATCC 16404 30°C – 35°C | 15 cfu | 19 cfu | 127% | Pass |
| *C. albicans* ATCC 10231 20°C – 25°C | 25 cfu | 33 cfu | 132% | Pass |
| *A. brasiliensis* ATCC 16404 20°C – 25°C | 20 cfu | 36 cfu | 180% | Pass |

**Conclusion:** A 1:1 dilution in TSB+ is a suitable method for MTC01.07_*P. distasonis.*

QA Reviewed / Approved by/date: [signature] 16 Jul 2021

FOCUS Laboratories is ISO/IEC 17025:2017 accredited by PJLA in the field of testing, Accreditation #77499

†Indicated specific test is outside the scope of ISO accreditation, however FOCUS is an FDA registered cGMP laboratory. This document is issued under the company's terms and conditions which were agreed upon during sample submission. FOCUS Laboratories' liability regarding this analysis is limited to the cost of the tests associated with this Test Report. The results are the property of the client listed above and FOCUS Laboratories. These results only relate to items tested above and apply to the samples as received. This report shall not be reproduced except in full without approval of FOCUS Laboratories.

FOCUS LABORATORIES

177 North Commerce Way
Bethlehem, PA 18017
(610) 866-7272
www.FOCUS-lab.com

### Test Report

**Client:** Icahn School of Medicine at Mount Sinai
Hess Center for Science and Medicine
1470 Madison Avenue, 8th FL.
New York, NY 10029

**Contact:** Lukas Bethlehem, PhD

212-824-9088

**Study Number:** 211635-8-R1

**Test Start Date:** 25-May-2021
**Test End Date:** 15-Jul-2021

**Testing Guideline:** USP <61>

| Sample Name | Lot |
|---|---|
| B. adolescentis | 1001283B150304_161114-H5 |

**Test Results:**

Table 1: USP <61>

| Test | Result | Specification | Pass/Fail |
|---|---|---|---|
| Aerobic Plate Count | 1800 cfu/mL | ≤ 1000 cfu/mL | Fail* |
| Yeast and Mold Count | ≤ 100 cfu/mL | ≤ 100 cfu/mL | Pass |

*Out of Specification was performed. (OOS #2142PA)

Table 2: USP <61> Retest

| Test | Result | Specification | Pass/Fail |
|---|---|---|---|
| Aerobic Plate Count | < 100 cfu/mL | ≤ 1000 cfu/mL | Pass |
| Yeast and Mold Count | < 100 cfu/mL | ≤ 100 cfu/mL | Pass |

*Out of Specification was performed. (OOS #2142PA)

Table 3: USP <61> Method Suitability Results

| Organism | Control count | Test Count | % Recovery | Pass/Fail |
|---|---|---|---|---|
| *S. aureus* ATCC 6538 | 94 cfu | 99 cfu | 105% | Pass |
| *P. aeruginosa* ATCC 9027 | 57 cfu | 76 cfu | 133% | Pass |
| *B. subtilis* ATCC 6633 | 48 cfu | 49 cfu | 102% | Pass |
| *C. albicans* ATCC 10231 30°C – 35°C | 31 cfu | 33 cfu | 106% | Pass |
| *A. brasiliensis* ATCC 16404 30°C – 35°C | 15 cfu | 10 cfu | 67% | Pass |
| *C. albicans* ATCC 10231 20°C – 25°C | 25 cfu | 36 cfu | 144% | Pass |
| *A. brasiliensis* ATCC 16404 20°C – 25°C | 20 cfu | 37 cfu | 185% | Pass |

**Conclusion:** A 1:1 dilution in TSB+ is a suitable method for MTC01.08_*B. adolescentis.*

QA Reviewed / Approved by/date: [signature] 16 Jul 2021

FOCUS Laboratories is ISO/IEC 17025:2017 accredited by PJLA in the field of testing, Accreditation #77499
†indicated specific test is outside the scope of ISO accreditation, however FOCUS is an FDA registered cGMP laboratory. This document is issued under the company's terms and conditions which were agreed upon during sample submission. FOCUS Laboratories' liability regarding this analysis is limited to the cost of the tests associated with this Test Report. The results are the property of the client listed above and FOCUS Laboratories. These results only relate to items tested above and apply to the samples as received. This report shall not be reproduced except in full without approval of FOCUS Laboratories.

FOCUS
LABORATORIES

177 North Commerce Way
Bethlehem, PA 18017
(610) 866-7272
www.FOCUS-lab.com

### Test Report

**Client:** Icahn School of Medicine at Mount Sinai
Hess Center for Science and Medicine
1470 Madison Avenue, 8th FL.
New York, NY 10029

**Contact:** Lukas Bethlehem, PhD

212-824-9088

**Study Number:** 211635-9-R1

**Test Start Date:** 25-May-2021
**Test End Date:** 01-Jun-2021

**Testing Guideline:** USP <61>

| Sample Name |
|---|
| MTC01.09_*P. merdae* |

**Test Results:**

Table 1: USP <61>

| Test | Result | Specification | Pass/Fail |
|---|---|---|---|
| Aerobic Plate Count | ≤ 100 cfu/mL | ≤ 1000 cfu/mL | Pass |
| Yeast and Mold Count | ≤ 100 cfu/mL | ≤ 100 cfu/mL | Pass |

Table 2: USP <61> Method Suitability Results

| Organism | Control count | Test Count | % Recovery | Pass/Fail |
|---|---|---|---|---|
| *S. aureus* ATCC 6538 | 94 cfu | 108 cfu | 115% | Pass |
| *P. aeruginosa* ATCC 9027 | 57 cfu | 70 cfu | 123% | Pass |
| *B. subtilis* ATCC 6633 | 48 cfu | 58 cfu | 121% | Pass |
| *C. albicans* ATCC 10231 30°C – 35°C | 31 cfu | 30 cfu | 97% | Pass |
| *A. brasiliensis* ATCC 16404 30°C – 35°C | 15 cfu | 8 cfu | 53% | Pass |
| *C. albicans* ATCC 10231 20°C – 25°C | 25 cfu | 34 cfu | 136% | Pass |
| *A. brasiliensis* ATCC 16404 20°C – 25°C | 20 cfu | 23 cfu | 115% | Pass |

**Conclusion:** A 1:1 dilution in TSB+ is a suitable method for MTC01.09_*P. merdae*.

QA Reviewed / Approved by/date: [signature] 16 Jul 2021

FOCUS Laboratories is ISO/IEC 17025:2017 accredited by PJLA in the field of testing, Accreditation #77499

†indicated specific test is outside the scope of ISO accreditation, however FOCUS is an FDA registered cGMP laboratory. This document is issued under the company's terms and conditions which were agreed upon during sample submission. FOCUS Laboratories' liability regarding this analysis is limited to the cost of the tests associated with this Test Report. The results are the property of the client listed above and FOCUS Laboratories. These results only relate to items tested above and apply to the samples as received. This report shall not be reproduced except in full without approval of FOCUS Laboratories.

FOCUS LABORATORIES

177 North Commerce Way
Bethlehem, PA 18017
(610) 866-7272
www.FOCUS-lab.com

### Test Report

**Client:** Icahn School of Medicine at Mount Sinai
Hess Center for Science and Medicine
1470 Madison Avenue, 8th FL.
New York, NY 10029

**Contact:** Lukas Bethlehem, PhD

212-824-9088

**Study Number:** 211635-10-R1

**Test Start Date:** 25-May-2021
**Test End Date:** 01-Jun-2021

**Testing Guideline:** USP <61>

| Sample Name |
|---|
| MTC01.10_*C. comes* |

**Test Results:**

Table 1: USP <61>

| Test | Result | Specification | Pass/Fail |
|---|---|---|---|
| Aerobic Plate Count | ≤ 100 cfu/mL | ≤ 1000 cfu/mL | Pass |
| Yeast and Mold Count | ≤ 100 cfu/mL | ≤ 100 cfu/mL | Pass |

Table 2: USP <61> Method Suitability Results

| Organism | Control count | Test Count | % Recovery | Pass/Fail |
|---|---|---|---|---|
| *S. aureus* ATCC 6538 | 94 cfu | 89 cfu | 95% | Pass |
| *P. aeruginosa* ATCC 9027 | 57 cfu | 64 cfu | 112% | Pass |
| *B. subtilis* ATCC 6633 | 48 cfu | 55 cfu | 115% | Pass |
| *C. albicans* ATCC 10231 30°C – 35°C | 31 cfu | 29 cfu | 94% | Pass |
| *A. brasiliensis* ATCC 16404 30°C – 35°C | 15 cfu | 13 cfu | 87% | Pass |
| *C. albicans* ATCC 10231 20°C – 25°C | 25 cfu | 33 cfu | 132% | Pass |
| *A. brasiliensis* ATCC 16404 20°C – 25°C | 20 cfu | 22 cfu | 110% | Pass |

**Conclusion:** A 1:1 dilution in TSB+ is a suitable method for MTC01.10_*C. comes*.

QA Reviewed / Approved by/date: [signature] 16 Jul 2021

FOCUS Laboratories is ISO/IEC 17025:2017 accredited by PJLA in the field of testing, Accreditation #77499
†indicated specific test is outside the scope of ISO accreditation, however FOCUS is an FDA registered cGMP laboratory. This document is issued under the company's terms and conditions which were agreed upon during sample submission. FOCUS Laboratories' liability regarding this analysis is limited to the cost of the tests associated with this Test Report. The results are the property of the client listed above and FOCUS Laboratories. These results only relate to items tested above and apply to the samples as received. This report shall not be reproduced except in full without approval of FOCUS Laboratories.

FOCUS
LABORATORIES

177 North Commerce Way
Bethlehem, PA 18017
(610) 866-7272
www.FOCUS-lab.com

### Test Report

**Client:** Icahn School of Medicine at Mount Sinai
Hess Center for Science and Medicine
1470 Madison Avenue, 8th FL.
New York, NY 10029

**Contact:** Lukas Bethlehem, PhD

212-824-9088

**Study Number:** 211635-11-R1

**Test Start Date:** 25-May-2021
**Test End Date:** 01-Jun-2021

**Testing Guideline:** USP <61>

| Sample Name |
|---|
| MTC01.11_*E. rectale* |

**Test Results:**

Table 1: USP <61>

| Test | Result | Specification | Pass/Fail |
|---|---|---|---|
| Aerobic Plate Count | ≤ 100 cfu/mL | ≤ 1000 cfu/mL | Pass |
| Yeast and Mold Count | ≤ 100 cfu/mL | ≤ 100 cfu/mL | Pass |

Table 2: USP <61> Method Suitability Results

| Organism | Control count | Test Count | % Recovery | Pass/Fail |
|---|---|---|---|---|
| *S. aureus* ATCC 6538 | 94 cfu | 100 cfu | 106% | Pass |
| *P. aeruginosa* ATCC 9027 | 57 cfu | 59 cfu | 104% | Pass |
| *B. subtilis* ATCC 6633 | 48 cfu | 58 cfu | 121% | Pass |
| *C. albicans* ATCC 10231 30°C – 35°C | 31 cfu | 28 cfu | 90% | Pass |
| *A. brasiliensis* ATCC 16404 30°C – 35°C | 15 cfu | 22 cfu | 147% | Pass |
| *C. albicans* ATCC 10231 20°C – 25°C | 25 cfu | 32 cfu | 128% | Pass |
| *A. brasiliensis* ATCC 16404 20°C – 25°C | 20 cfu | 17 cfu | 85% | Pass |

**Conclusion:** A 1:1 dilution in TSB+ is a suitable method for MTC01.11_*E. rectale*.

QA Reviewed / Approved by/date: [signature] 16 Jul 2021

FOCUS Laboratories is ISO/IEC 17025:2017 accredited by PJLA in the field of testing, Accreditation #77499
†indicated specific test is outside the scope of ISO accreditation, however FOCUS is an FDA registered cGMP laboratory. This document is issued under the company's terms and conditions which were agreed upon during sample submission. FOCUS Laboratories' liability regarding this analysis is limited to the cost of the tests associated with this Test Report. The results are the property of the client listed above and FOCUS Laboratories. These results only relate to items tested above and apply to the samples as received. This report shall not be reproduced except in full without approval of FOCUS Laboratories.

FOCUS LABORATORIES

177 North Commerce Way
Bethlehem, PA 18017
(610) 866-7272
www.FOCUS-lab.com

### Test Report

**Client:** Icahn School of Medicine at Mount Sinai
Hess Center for Science and Medicine
1470 Madison Avenue, 8th FL.
New York, NY 10029

**Contact:** Lukas Bethlehem, PhD

212-824-9088

**Study Number:** 211635-12-R1

**Test Start Date:** 25-May-2021
**Test End Date:** 01-Jun-2021

**Testing Guideline:** USP <61>

| Sample Name |
|---|
| MTC01.12_*B. caccae* |

**Test Results:**

Table 1: USP <61>

| Test | Result | Specification | Pass/Fail |
|---|---|---|---|
| Aerobic Plate Count | ≤ 100 cfu/mL | ≤ 1000 cfu/mL | Pass |
| Yeast and Mold Count | ≤ 100 cfu/mL | ≤ 100 cfu/mL | Pass |

Table 2: USP <61> Method Suitability Results

| Organism | Control count | Test Count | % Recovery | Pass/Fail |
|---|---|---|---|---|
| *S. aureus* ATCC 6538 | 94 cfu | 102 cfu | 109% | Pass |
| *P. aeruginosa* ATCC 9027 | 57 cfu | 74 cfu | 130% | Pass |
| *B. subtilis* ATCC 6633 | 48 cfu | 63 cfu | 131% | Pass |
| *C. albicans* ATCC 10231 30°C – 35°C | 31 cfu | 30 cfu | 97% | Pass |
| *A. brasiliensis* ATCC 16404 30°C – 35°C | 15 cfu | 11 cfu | 73% | Pass |
| *C. albicans* ATCC 10231 20°C – 25°C | 25 cfu | 28 cfu | 112% | Pass |
| *A. brasiliensis* ATCC 16404 20°C – 25°C | 20 cfu | 28 cfu | 140% | Pass |

**Conclusion:** A 1:1 dilution in TSB+ is a suitable method for MTC01.12_*B. caccae.*

QA Reviewed / Approved by/date: [signature] 16 Jul 2021

FOCUS Laboratories is ISO/IEC 17025:2017 accredited by PJLA in the field of testing, Accreditation #77499

†indicated specific test is outside the scope of ISO accreditation, however FOCUS is an FDA registered cGMP laboratory. This document is issued under the company's terms and conditions which were agreed upon during sample submission. FOCUS Laboratories' liability regarding this analysis is limited to the cost of the tests associated with this Test Report. The results are the property of the client listed above and FOCUS Laboratories. These results only relate to items tested above and apply to the samples as received. This report shall not be reproduced except in full without approval of FOCUS Laboratories.

FOCUS LABORATORIES

177 North Commerce Way
Bethlehem, PA 18017
(610) 866-7272
www.FOCUS-lab.com

### Test Report

**Client:** Icahn School of Medicine at Mount Sinai
Hess Center for Science and Medicine
1470 Madison Avenue, 8th FL.
New York, NY 10029

**Contact:** Lukas Bethlehem, PhD

212-824-9088

**Study Number:** 211635-13-R1

**Test Start Date:** 25-May-2021

**Test End Date:** 01-Jun-2021

**Testing Guideline:** USP <61>

| Sample Name |
|---|
| MTC01.13_*D. longicatena* |

**Test Results:**

Table 1: USP <61>

| Test | Result | Specification | Pass/Fail |
|---|---|---|---|
| Aerobic Plate Count | ≤ 100 cfu/mL | ≤ 1000 cfu/mL | Pass |
| Yeast and Mold Count | ≤ 100 cfu/mL | ≤ 100 cfu/mL | Pass |

Table 2: USP <61> Method Suitability Results

| Organism | Control count | Test Count | % Recovery | Pass/Fail |
|---|---|---|---|---|
| *S. aureus* ATCC 6538 | 94 cfu | 89 cfu | 95% | Pass |
| *P. aeruginosa* ATCC 9027 | 57 cfu | 62 cfu | 109% | Pass |
| *B. subtilis* ATCC 6633 | 48 cfu | 47 cfu | 98% | Pass |
| *C. albicans* ATCC 10231 30°C – 35°C | 31 cfu | 34 cfu | 110% | Pass |
| *A. brasiliensis* ATCC 16404 30°C – 35°C | 15 cfu | 13 cfu | 87% | Pass |
| *C. albicans* ATCC 10231 20°C – 25°C | 25 cfu | 32 cfu | 128% | Pass |
| *A. brasiliensis* ATCC 16404 20°C – 25°C | 20 cfu | 27 cfu | 135% | Pass |

**Conclusion:** A 1:1 dilution in TSB+ is a suitable method for MTC01.13_*D. longicatena*

QA Reviewed / Approved by/date: [signature] 16 Jul 2021

FOCUS Laboratories is ISO/IEC 17025:2017 accredited by PJLA in the field of testing, Accreditation #77499

†indicated specific test is outside the scope of ISO accreditation, however FOCUS is an FDA registered cGMP laboratory. This document is issued under the company's terms and conditions which were agreed upon during sample submission. FOCUS Laboratories' liability regarding this analysis is limited to the cost of the tests associated with this Test Report. The results are the property of the client listed above and FOCUS Laboratories. These results only relate to items tested above and apply to the samples as received. This report shall not be reproduced except in full without approval of FOCUS Laboratories.

FOCUS LABORATORIES

177 North Commerce Way
Bethlehem, PA 18017
(610) 866-7272
www.FOCUS-lab.com

### Test Report

**Client:** Icahn School of Medicine at Mount Sinai
Hess Center for Science and Medicine
1470 Madison Avenue, 8th FL.
New York, NY 10029

**Contact:** Lukas Bethlehem, PhD

212-824-9088

**Study Number:** 211635-14-R1

**Test Start Date:** 25-May-2021
**Test End Date:** 01-Jun-2021

**Testing Guideline:** USP <61>

| Sample Name |
|---|
| MTC01.14_*O. splachnicus* |

**Test Results:**

Table 1: USP <61>

| Test | Result | Specification | Pass/Fail |
|---|---|---|---|
| Aerobic Plate Count | ≤ 100 cfu/mL | ≤ 1000 cfu/mL | Pass |
| Yeast and Mold Count | ≤ 100 cfu/mL | ≤ 100 cfu/mL | Pass |

Table 2: USP <61> Method Suitability Results

| Organism | Control count | Test Count | % Recovery | Pass/Fail |
|---|---|---|---|---|
| *S. aureus* ATCC 6538 | 94 cfu | 85 cfu | 90% | Pass |
| *P. aeruginosa* ATCC 9027 | 57 cfu | 61 cfu | 107% | Pass |
| *B. subtilis* ATCC 6633 | 48 cfu | 60 cfu | 125% | Pass |
| *C. albicans* ATCC 10231 30°C – 35°C | 31 cfu | 26 cfu | 84% | Pass |
| *A. brasiliensis* ATCC 16404 30°C – 35°C | 15 cfu | 15 cfu | 100% | Pass |
| *C. albicans* ATCC 10231 20°C – 25°C | 25 cfu | 30 cfu | 120% | Pass |
| *A. brasiliensis* ATCC 16404 20°C – 25°C | 20 cfu | 31 cfu | 155% | Pass |

**Conclusion:** A 1:1 dilution in TSB+ is a suitable method for MTC01.14_*O. splachnicus*

QA Reviewed / Approved by/date: [signature] 16 Jul 2021

FOCUS Laboratories is ISO/IEC 17025:2017 accredited by PJLA in the field of testing, Accreditation #77499
†indicated specific test is outside the scope of ISO accreditation, however FOCUS is an FDA registered cGMP laboratory. This document is issued under the company's terms and conditions which were agreed upon during sample submission. FOCUS Laboratories' liability regarding this analysis is limited to the cost of the tests associated with this Test Report. The results are the property of the client listed above and FOCUS Laboratories. These results only relate to items tested above and apply to the samples as received. This report shall not be reproduced except in full without approval of FOCUS Laboratories.

FOCUS LABORATORIES

177 North Commerce Way
Bethlehem, PA 18017
(610) 866-7272
www.FOCUS-lab.com

### Test Report

**Client:** Icahn School of Medicine at Mount Sinai
Hess Center for Science and Medicine
1470 Madison Avenue, 8th FL.
New York, NY 10029

**Contact:** Lukas Bethlehem, PhD

212-824-9088

**Study Number:** 211635-15-R1

**Test Start Date:** 25-May-2021

**Test End Date:** 01-Jun-2021

**Testing Guideline:** USP <61>

| Sample Name |
|---|
| MTC01.15_*B. cellulosilyticus* |

**Test Results:**

Table 1: USP <61>

| Test | Result | Specification | Pass/Fail |
|---|---|---|---|
| Aerobic Plate Count | ≤ 100 cfu/mL | ≤ 1000 cfu/mL | Pass |
| Yeast and Mold Count | ≤ 100 cfu/mL | ≤ 100 cfu/mL | Pass |

Table 2: USP <61> Method Suitability Results

| Organism | Control count | Test Count | % Recovery | Pass/Fail |
|---|---|---|---|---|
| *S. aureus* ATCC 6538 | 94 cfu | 98 cfu | 104% | Pass |
| *P. aeruginosa* ATCC 9027 | 57 cfu | 60 cfu | 105% | Pass |
| *B. subtilis* ATCC 6633 | 48 cfu | 58 cfu | 121% | Pass |
| *C. albicans* ATCC 10231 30°C – 35°C | 31 cfu | 31 cfu | 100% | Pass |
| *A. brasiliensis* ATCC 16404 30°C – 35°C | 15 cfu | 18 cfu | 120% | Pass |
| *C. albicans* ATCC 10231 20°C – 25°C | 25 cfu | 34 cfu | 136% | Pass |
| *A. brasiliensis* ATCC 16404 20°C – 25°C | 20 cfu | 39 cfu | 195% | Pass |

**Conclusion:** A 1:1 dilution in TSB+ is a suitable method for MTC01.15_*B. cellulosilyticus*

QA Reviewed / Approved by/date:

FOCUS Laboratories is ISO/IEC 17025:2017 accredited by PJLA in the field of testing, Accreditation #77499

†indicated specific test is outside the scope of ISO accreditation, however FOCUS is an FDA registered cGMP laboratory. This document is issued under the company's terms and conditions which were agreed upon during sample submission. FOCUS Laboratories' liability regarding this analysis is limited to the cost of the tests associated with this Test Report. The results are the property of the client listed above and FOCUS Laboratories. These results only relate to items tested above and apply to the samples as received. This report shall not be reproduced except in full without approval of FOCUS Laboratories.

FOCUS LABORATORIES

177 North Commerce Way
Bethlehem, PA 18017
(610) 866-7272
www.FOCUS-lab.com

### Test Report

**Client:** Icahn School of Medicine at Mount Sinai
Hess Center for Science and Medicine
1470 Madison Avenue, 8th FL.
New York, NY 10029

**Contact:** Lukas Bethlehem, PhD

212-824-9088

**Study Number:** 211635-16-R1

**Test Start Date:** 25-May-2021

**Test End Date:** 01-Jun-2021

**Testing Guideline:** USP <61>

| Sample Name |
|---|
| MTC01.16_*B. pseudocatenulatum* |

**Test Results:**

Table 1: USP <61>

| Test | Result | Specification | Pass/Fail |
|---|---|---|---|
| Aerobic Plate Count | ≤ 100 cfu/mL | ≤ 1000 cfu/mL | Pass |
| Yeast and Mold Count | ≤ 100 cfu/mL | ≤ 100 cfu/mL | Pass |

Table 2: USP <61> Method Suitability Results

| Organism | Control count | Test Count | % Recovery | Pass/Fail |
|---|---|---|---|---|
| *S. aureus* ATCC 6538 | 94 cfu | 92 cfu | 98% | Pass |
| *P. aeruginosa* ATCC 9027 | 57 cfu | 69 cfu | 121% | Pass |
| *B. subtilis* ATCC 6633 | 48 cfu | 52 cfu | 108% | Pass |
| *C. albicans* ATCC 10231 30°C – 35°C | 31 cfu | 29 cfu | 94% | Pass |
| *A. brasiliensis* ATCC 16404 30°C – 35°C | 15 cfu | 18 cfu | 120% | Pass |
| *C. albicans* ATCC 10231 20°C – 25°C | 25 cfu | 31 cfu | 124% | Pass |
| *A. brasiliensis* ATCC 16404 20°C – 25°C | 20 cfu | 32 cfu | 160% | Pass |

**Conclusion:** A 1:1 dilution in TSB+ is a suitable method for MTC01.16_*B. pseudocatenulatum.*

Report 211635 was revised on 16-Jul-2021 to add repeat testing for sample -08.

REVISED
JB 16 Jul 2021

QA Reviewed / Approved by/date: [signature] 16 Jul 2021

FOCUS Laboratories is ISO/IEC 17025:2017 accredited by PJLA in the field of testing, Accreditation #77499

†Indicated specific test is outside the scope of ISO accreditation, however FOCUS is an FDA registered cGMP laboratory. This document is issued under the company's terms and conditions which were agreed upon during sample submission. FOCUS Laboratories' liability regarding this analysis is limited to the cost of the tests associated with this Test Report. The results are the property of the client listed above and FOCUS Laboratories. These results only relate to items tested above and apply to the samples as received. This report shall not be reproduced except in full without approval of FOCUS Laboratories.

### FOCUS Labs MTC01 mixed strain USP<61> Method suitability report

FOCUS LABORATORIES

#### Certificate of Analysis No. 22-00237

**Customer:** Icahn School of Medicine
**Address:** 1470 Madison Ave, Fl. 8 RM# 201
New York City, NY 10029
**Date Authorized:** February 1, 2022

**Contact:** Lukas Bethlehem
**Total Number of Pages:** 1

#### Test Results

**Sample No.** 22-00237-01
**Product Name:** DP-Mix_Test
**Lot Number:** 2021-01-19

**Sample Received Date:** January 21, 2022
**Sample Description:** NA

| Test | Method | Results | Specification | Pass/Fail | Completed | Lab |
|---|---|---|---|---|---|---|
| 20-25C A. brasiliensis % Recovery | USP <61> | 78 % | 50 200 % | PASS | 02/01/2022 | FOCUSPA |
| 20-25C C. albicans % Recovery | USP <61> | 100 % | 50 200 % | PASS | 02/01/2022 | FOCUSPA |
| 30-35C A. brasiliensis % Recovery | USP <61> | 100 % | 50 200 % | PASS | 02/01/2022 | FOCUSPA |
| 30-35C B. subtilis % Recovery | USP <61> | 98 % | 50 200 % | PASS | 02/01/2022 | FOCUSPA |
| 30-35C C. albicans % Recovery | USP <61> | 90 % | 50 200 % | PASS | 02/01/2022 | FOCUSPA |
| 30-35C P. aeruginosa % Recovery | USP <61> | 70 % | 50 200 % | PASS | 02/01/2022 | FOCUSPA |
| 30-35C S. aureus % Recovery | USP <61> | 89 % | 50 200 % | PASS | 02/01/2022 | FOCUSPA |

##### Reference Information:

**Laboratories:**

FOCUSPA : Focus Laboratories Pennsylvania, 177 N Commerce Way, Bethlehem, PA, 18017 United States

Reviewed / Approved by:

Nathan Evans, QA Data Reviewer for PA

Signed Date: February 1, 2022

FOCUS Laboratories Pennsylvania is ISO/IEC 17025:2017 accredited by PJLA in the field of testing, Accreditation #77499

†indicated specific test is outside the scope of ISO accreditation, however FOCUS is an FDA registered cGMP laboratory.
This document is issued under the company's terms and conditions which were agreed upon during sample submission. FOCUS Laboratories' liability regarding this analysis is limited to the cost of the tests associated with this Test Report. The results are the property of the client listed above and FOCUS Laboratories. These results only relate to items tested above and apply to the samples as received. This report shall not be reproduced except in full without approval of FOCUS Laboratories.

### FOCUS Labs USP<61> MCB reports

FOCUS LABORATORIES

177 North Commerce Way
Bethlehem, PA 18017
(610) 866-7272
www.FOCUS-lab.com

### Test Report

**Client:** Icahn School of Medicine at Mount Sinai
Hess Center for Science and Medicine
1470 Madison Avenue, 8th FL.
New York, NY 10029

**Contact:** Lukas Bethlehem, PhD

212-824-9087

**Study Number:** 212978-1

**Test Start Date:** 27-Aug-2021
**Test End Date:** 01-Sep-2021

**Testing Guideline:** USP <61>

| Sample Name | Lot Number |
|---|---|
| B. uniformis | 2021-07-30-MTC01.01-MCB |

**Test Results:**

| Test | Result | Specification | Pass/Fail |
|---|---|---|---|
| Aerobic Plate Count | < 10 cfu/mL | ≤ 1000 cfu/mL | Pass |
| Yeast and Mold Count | < 10 cfu/mL | ≤ 100 cfu/mL | Pass |

QA Reviewed / Approved by/date: Jessica Bergstein 02Sep2021

FOCUS Laboratories is ISO/IEC 17025:2017 accredited by PJLA in the field of testing, Accreditation #77499
†Indicated specific test is outside the scope of ISO accreditation, however FOCUS is an FDA registered cGMP laboratory. This document is issued under the company's terms and conditions which were agreed upon during sample submission. FOCUS Laboratories' liability regarding this analysis is limited to the cost of the tests associated with this Test Report. The results are the property of the client listed above and FOCUS Laboratories. These results only relate to items tested above and apply to the samples as received. This report shall not be reproduced except in full without approval of FOCUS Laboratories.

FOCUS LABORATORIES

177 North Commerce Way
Bethlehem, PA 18017
(610) 866-7272
www.FOCUS-lab.com

### Test Report

**Client:** Icahn School of Medicine at Mount Sinai
Hess Center for Science and Medicine
1470 Madison Avenue, 8th FL.
New York, NY 10029

**Contact:** Lukas Bethlehem, PhD

212-824-9087

**Study Number:** 212978-2

**Test Start Date:** 27-Aug-2021
**Test End Date:** 01-Sep-2021

**Testing Guideline:** USP <61>

| Sample Name | Lot Number |
|---|---|
| B. ovatus | 2021-08-06-MTC01.02-MCB |

**Test Results:**

| Test | Result | Specification | Pass/Fail |
|---|---|---|---|
| Aerobic Plate Count | < 10 cfu/mL | ≤ 1000 cfu/mL | Pass |
| Yeast and Mold Count | < 10 cfu/mL | ≤ 100 cfu/mL | Pass |

QA Reviewed / Approved by/date: Jessica Bergsheim 02Sep2021

FOCUS Laboratories is ISO/IEC 17025:2017 accredited by PJLA in the field of testing, Accreditation #77499

†Indicated specific test is outside the scope of ISO accreditation, however FOCUS is an FDA registered cGMP laboratory. This document is issued under the company's terms and conditions which were agreed upon during sample submission. FOCUS Laboratories' liability regarding this analysis is limited to the cost of the tests associated with this Test Report. The results are the property of the client listed above and FOCUS Laboratories. These results only relate to items tested above and apply to the samples as received. This report shall not be reproduced except in full without approval of FOCUS Laboratories.

FOCUS LABORATORIES

177 North Commerce Way
Bethlehem, PA 18017
(610) 866-7272
www.FOCUS-lab.com

### Test Report

**Client:** Icahn School of Medicine at Mount Sinai
Hess Center for Science and Medicine
1470 Madison Avenue, 8th FL.
New York, NY 10029

**Contact:** Lukas Bethlehem, PhD

212-824-9087

**Study Number:** 212978-3

**Test Start Date:** 27-Aug-2021
**Test End Date:** 01-Sep-2021

**Testing Guideline:** USP <61>

| Sample Name | Lot Number |
|---|---|
| B. longum | 2021-08-13-MTC01.03-MCB |

**Test Results:**

| Test | Result | Specification | Pass/Fail |
|---|---|---|---|
| Aerobic Plate Count | < 10 cfu/mL | ≤ 1000 cfu/mL | Pass |
| Yeast and Mold Count | < 10 cfu/mL | ≤ 100 cfu/mL | Pass |

QA Reviewed / Approved by/date: Jessica Bergstein 02Sep2021

FOCUS Laboratories is ISO/IEC 17025:2017 accredited by PJLA in the field of testing, Accreditation #77499

†Indicated specific test is outside the scope of ISO accreditation, however FOCUS is an FDA registered cGMP laboratory. This document is issued under the company's terms and conditions which were agreed upon during sample submission. FOCUS Laboratories' liability regarding this analysis is limited to the cost of the tests associated with this Test Report. The results are the property of the client listed above and FOCUS Laboratories. These results only relate to items tested above and apply to the samples as received. This report shall not be reproduced except in full without approval of FOCUS Laboratories.

FOCUS LABORATORIES

177 North Commerce Way
Bethlehem, PA 18017
(610) 866-7272
www.FOCUS-lab.com

### Test Report

**Client:** Icahn School of Medicine at Mount Sinai
Hess Center for Science and Medicine
1470 Madison Avenue, 8th FL.
New York, NY 10029

**Contact:** Lukas Bethlehem, PhD

212-824-9088

**Study Number:** 213161-1

**Test Start Date:** 09-Sep-2021
**Test End Date:** 14-Sep-2021

**Testing Guideline:** USP <61>

| Sample Name | Lot |
|---|---|
| *B. vulgatus* | 2021-08-19-MTC01.05-MCB |

**Test Results:**

| Test | Result | Specification | Pass/Fail |
|---|---|---|---|
| Aerobic Plate Count | < 100 cfu/mL | ≤ 1000 cfu/mL | Pass |
| Yeast and Mold Count | < 100 cfu/mL | ≤ 100 cfu/mL | Pass |

QA Reviewed / Approved by/date: Jessica Bergstein 16Sep2021

FOCUS Laboratories is ISO/IEC 17025:2017 accredited by PJLA in the field of testing, Accreditation #77499

†Indicated specific test is outside the scope of ISO accreditation, however FOCUS is an FDA registered cGMP laboratory. This document is issued under the company's terms and conditions which were agreed upon during sample submission. FOCUS Laboratories' liability regarding this analysis is limited to the cost of the tests associated with this Test Report. The results are the property of the client listed above and FOCUS Laboratories. These results only relate to items tested above and apply to the samples as received. This report shall not be reproduced except in full without approval of FOCUS Laboratories.

177 North Commerce Way
Bethlehem, PA 18017
(610) 866-7272
www.FOCUS-lab.com

### Test Report

**Client:** Icahn School of Medicine at Mount Sinai
Hess Center for Science and Medicine
1470 Madison Avenue, 8th FL.
New York, NY 10029

**Contact:** Lukas Bethlehem, PhD

212-824-9088

**Study Number:** 213161-2

**Test Start Date:** 09-Sep-2021
**Test End Date:** 14-Sep-2021

**Testing Guideline:** USP <61>

| Sample Name | Lot |
|---|---|
| *C. aerofaciens* | 2021-08-27-MTC01.06-MCB |

**Test Results:**

| Test | Result | Specification | Pass/Fail |
|---|---|---|---|
| Aerobic Plate Count | < 100 cfu/mL | ≤ 1000 cfu/mL | Pass |
| Yeast and Mold Count | < 100 cfu/mL | ≤ 100 cfu/mL | Pass |

QA Reviewed / Approved by/date: 16Sep2021

FOCUS Laboratories is ISO/IEC 17025:2017 accredited by PJLA in the field of testing, Accreditation #77499

†Indicated specific test is outside the scope of ISO accreditation, however FOCUS is an FDA registered cGMP laboratory. This document is issued under the company's terms and conditions which were agreed upon during sample submission. FOCUS Laboratories' liability regarding this analysis is limited to the cost of the tests associated with this Test Report. The results are the property of the client listed above and FOCUS Laboratories. These results only relate to items tested above and apply to the samples as received. This report shall not be reproduced except in full without approval of FOCUS Laboratories.

FOCUS LABORATORIES

177 North Commerce Way
Bethlehem, PA 18017
(610) 866-7272
www.FOCUS-lab.com

### Test Report

**Client:** Icahn School of Medicine at Mount Sinai
Hess Center for Science and Medicine
1470 Madison Avenue, 8th FL.
New York, NY 10029

**Contact:** Lukas Bethlehem, PhD

212-824-9088

**Study Number:** 213161-3

**Test Start Date:** 09-Sep-2021
**Test End Date:** 14-Sep-2021

**Testing Guideline:** USP <61>

| Sample Name | Lot |
|---|---|
| *P. distasonis* | 2021-08-19-MTC01.07-MCB |

**Test Results:**

| Test | Result | Specification | Pass/Fail |
|---|---|---|---|
| Aerobic Plate Count | < 100 cfu/mL | ≤ 1000 cfu/mL | Pass |
| Yeast and Mold Count | < 100 cfu/mL | ≤ 100 cfu/mL | Pass |

QA Reviewed / Approved by/date: [illegible] 16Sep2021

FOCUS Laboratories is ISO/IEC 17025:2017 accredited by PJLA in the field of testing, Accreditation #77499

†Indicated specific test is outside the scope of ISO accreditation, however FOCUS is an FDA registered cGMP laboratory. This document is issued under the company's terms and conditions which were agreed upon during sample submission. FOCUS Laboratories' liability regarding this analysis is limited to the cost of the tests associated with this Test Report. The results are the property of the client listed above and FOCUS Laboratories. These results only relate to items tested above and apply to the samples as received. This report shall not be reproduced except in full without approval of FOCUS Laboratories.

FOCUS LABORATORIES

177 North Commerce Way
Bethlehem, PA 18017
(610) 866-7272
www.FOCUS-lab.com

### Test Report

**Client:** Icahn School of Medicine at Mount Sinai
Hess Center for Science and Medicine
1470 Madison Avenue, 8th FL.
New York, NY 10029
**Contact:** Lukas Bethlehem, PhD

212-824-9088

**Study Number:** 213161-4

**Test Start Date:** 09-Sep-2021
**Test End Date:** 14-Sep-2021

**Testing Guideline:** USP <61>

| Sample Name | Lot |
|---|---|
| *B. adolescentis* | 2021-08-27-MTC01.08-MCB |

**Test Results:**

| Test | Result | Specification | Pass/Fail |
|---|---|---|---|
| Aerobic Plate Count | < 100 cfu/mL | ≤ 1000 cfu/mL | Pass |
| Yeast and Mold Count | < 100 cfu/mL | ≤ 100 cfu/mL | Pass |

QA Reviewed / Approved by/date: Jessica Bergheim 16Sep2021

FOCUS Laboratories is ISO/IEC 17025:2017 accredited by PJLA in the field of testing, Accreditation #77499
†Indicated specific test is outside the scope of ISO accreditation, however FOCUS is an FDA registered cGMP laboratory. This document is issued under the company's terms and conditions which were agreed upon during sample submission. FOCUS Laboratories' liability regarding this analysis is limited to the cost of the tests associated with this Test Report. The results are the property of the client listed above and FOCUS Laboratories. These results only relate to items tested above and apply to the samples as received. This report shall not be reproduced except in full without approval of FOCUS Laboratories.

FOCUS
LABORATORIES

177 North Commerce Way
Bethlehem, PA 18017
(610) 866-7272
www.FOCUS-lab.com

### Test Report

**Client:** Icahn School of Medicine at Mount Sinai
Hess Center for Science and Medicine
1470 Madison Avenue, 8th FL.
New York, NY 10029

**Contact:** Lukas Bethlehem, PhD

212-824-9087

**Study Number:** 212978-4

**Test Start Date:** 27-Aug-2021
**Test End Date:** 01-Sep-2021

**Testing Guideline:** USP <61>

| Sample Name | Lot Number |
|---|---|
| B. thetaiotaomicron | 2021-08-13-MTC01.04-MCB |

**Test Results:**

| Test | Result | Specification | Pass/Fail |
|---|---|---|---|
| Aerobic Plate Count | < 10 cfu/mL | ≤ 1000 cfu/mL | Pass |
| Yeast and Mold Count | < 10 cfu/mL | ≤ 100 cfu/mL | Pass |

QA Reviewed / Approved by/date: Jessica Bergstein 02Sep2021

FOCUS Laboratories is ISO/IEC 17025:2017 accredited by PJLA in the field of testing, Accreditation #77499

†Indicated specific test is outside the scope of ISO accreditation, however FOCUS is an FDA registered cGMP laboratory. This document is issued under the company's terms and conditions which were agreed upon during sample submission. FOCUS Laboratories' liability regarding this analysis is limited to the cost of the tests associated with this Test Report. The results are the property of the client listed above and FOCUS Laboratories. These results only relate to items tested above and apply to the samples as received. This report shall not be reproduced except in full without approval of FOCUS Laboratories.

FOCUS LABORATORIES

### Certificate of Analysis No. 21-05312

**Customer:** Icahn School of Medicine
**Address:** 1470 Madison Ave, Fl. 8 RM# 201
New York City, NY 10029
**Date Authorized:** December 9, 2021

**Contact:** Lukas Bethlehem
**Total Number of Pages:** 2

#### Test Results

**Sample No.** 21-05312-01
**Product Name:** P. merdae
**Lot Number:** 2021-10-21-MTC01.09-MCB
**Comments:** PO:SC524970J
**Sample Received Date:** November 23, 2021
**Sample Description:** NA

| Test | Method | Results | Specification | Pass/Fail | Completed | Lab |
|---|---|---|---|---|---|---|
| Aerobic Plate Count (g) | USP <61> | <10 cfu/g | <1000 cfu/g | PASS | 12/06/2021 | FOCUSPA |
| Yeast and Mold Count (g) | USP <61> | <10 cfu/g | <100 cfu/g | PASS | 12/06/2021 | FOCUSPA |

**Sample No.** 21-05312-02
**Product Name:** C. comes
**Lot Number:** 2021-10-01-MTC01.10-MCB
**Comments:** PO:SC524970J
**Sample Received Date:** November 23, 2021
**Sample Description:** NA

| Test | Method | Results | Specification | Pass/Fail | Completed | Lab |
|---|---|---|---|---|---|---|
| Aerobic Plate Count (g) | USP <61> | <10 cfu/g | <1000 cfu/g | PASS | 12/06/2021 | FOCUSPA |
| Yeast and Mold Count (g) | USP <61> | <10 cfu/g | <100 cfu/g | PASS | 12/06/2021 | FOCUSPA |

**Sample No.** 21-05312-03
**Product Name:** E. rectale
**Lot Number:** 2021-10-08-MTC01.11-MCB
**Comments:** PO:SC524970J
**Sample Received Date:** November 23, 2021
**Sample Description:** NA

| Test | Method | Results | Specification | Pass/Fail | Completed | Lab |
|---|---|---|---|---|---|---|
| Aerobic Plate Count (g) | USP <61> | <10 cfu/g | <1000 cfu/g | PASS | 12/06/2021 | FOCUSPA |
| Yeast and Mold Count (g) | USP <61> | <10 cfu/g | <100 cfu/g | PASS | 12/06/2021 | FOCUSPA |

**Sample No.** 21-05312-04
**Product Name:** B. caccae
**Lot Number:** 2021-09-17-MTC01.12-MCB
**Comments:** PO:SC524970J
**Sample Received Date:** November 23, 2021
**Sample Description:** NA

| Test | Method | Results | Specification | Pass/Fail | Completed | Lab |
|---|---|---|---|---|---|---|
| Aerobic Plate Count (g) | USP <61> | <10 cfu/g | <1000 cfu/g | PASS | 12/06/2021 | FOCUSPA |
| Yeast and Mold Count (g) | USP <61> | <10 cfu/g | <100 cfu/g | PASS | 12/06/2021 | FOCUSPA |

FOCUS Laboratories Pennsylvania is ISO/IEC 17025:2017 accredited by PJLA in the field of testing, Accreditation #77499
†Indicated specific test is outside the scope of ISO accreditation, however FOCUS is an FDA registered cGMP laboratory.
This document is issued under the company's terms and conditions which were agreed upon during sample submission. FOCUS Laboratories' liability regarding this analysis is limited to the cost of the tests associated with this Test Report. The results are the property of the client listed above and FOCUS Laboratories. These results only relate to items tested above and apply to the samples as received. This report shall not be reproduced except in full without approval of FOCUS Laboratories.

| | | | |
|---|---|---|---|
| **Sample No.** | 21-05312-05 | **Sample Received Date:** | November 23, 2021 |
| **Product Name:** | D. longicatena | **Sample Description:** | NA |
| **Lot Number:** | 2021-10-21-MTC01.13-MCB | | |
| **Comments:** | PO:SC524970J | | |

| Test | Method | Results | Specification | Pass/Fail | Completed | Lab |
|---|---|---|---|---|---|---|
| Aerobic Plate Count (g) | USP <61> | <10 cfu/g | <1000 cfu/g | PASS | 12/06/2021 | FOCUSPA |
| Yeast and Mold Count (g) | USP <61> | <10 cfu/g | <100 cfu/g | PASS | 12/06/2021 | FOCUSPA |

| | | | |
|---|---|---|---|
| **Sample No.** | 21-05312-06 | **Sample Received Date:** | November 23, 2021 |
| **Product Name:** | O. splanchnicus | **Sample Description:** | NA |
| **Lot Number:** | 2021-09-17-MTC01.14-MCB | | |
| **Comments:** | PO:SC524970J | | |

| Test | Method | Results | Specification | Pass/Fail | Completed | Lab |
|---|---|---|---|---|---|---|
| Aerobic Plate Count (g) | USP <61> | <10 cfu/g | <1000 cfu/g | PASS | 12/06/2021 | FOCUSPA |
| Yeast and Mold Count (g) | USP <61> | <10 cfu/g | <100 cfu/g | PASS | 12/06/2021 | FOCUSPA |

| | | | |
|---|---|---|---|
| **Sample No.** | 21-05312-07 | **Sample Received Date:** | November 23, 2021 |
| **Product Name:** | B. cellulosilyticus | **Sample Description:** | NA |
| **Lot Number:** | 2021-11-05-MTC01.15-MCB | | |
| **Comments:** | PO:SC524970J | | |

| Test | Method | Results | Specification | Pass/Fail | Completed | Lab |
|---|---|---|---|---|---|---|
| Aerobic Plate Count (g) | USP <61> | <10 cfu/g | <1000 cfu/g | PASS | 12/06/2021 | FOCUSPA |
| Yeast and Mold Count (g) | USP <61> | <10 cfu/g | <100 cfu/g | PASS | 12/06/2021 | FOCUSPA |

| | | | |
|---|---|---|---|
| **Sample No.** | 21-05312-08 | **Sample Received Date:** | November 23, 2021 |
| **Product Name:** | B. pseudocatenulatum | **Sample Description:** | NA |
| **Lot Number:** | 2021-09-24-MTC01.16-MCB | | |
| **Comments:** | PO:SC524970J | | |

| Test | Method | Results | Specification | Pass/Fail | Completed | Lab |
|---|---|---|---|---|---|---|
| Aerobic Plate Count (g) | USP <61> | <10 cfu/g | <1000 cfu/g | PASS | 12/06/2021 | FOCUSPA |
| Yeast and Mold Count (g) | USP <61> | <10 cfu/g | <100 cfu/g | PASS | 12/06/2021 | FOCUSPA |

#### Reference Information:

**Laboratories:**

FOCUSPA : Focus Laboratories Pennsylvania, 177 N Commerce Way, Bethlehem, PA, 18017 United States

Reviewed / Approved by: *Nath Evans*

Nathan Evans, QA Data Reviewer for PA

Signed Date: December 9, 2021

### FOCUS Labs USP<61> DS reports

FOCUS LABORATORIES

#### Certificate of Analysis No. 22-02526

**Customer:** Icahn School of Medicine
**Address:** 1470 Madison Ave, Fl. 8 RM# 201
New York City, NY 10029
**Date Authorized:** June 29, 2022

**Contact:** Lukas Bethlehem
**Total Number of Pages:** 4

##### Test Results

**Sample No.** 22-02526-01
**Product Name:** B. uniformis
**Lot Number:** 2022-04-01-MTC01.01
**Comments:** NA
**Sample Received Date:** June 24, 2022
**Sample Description:** NA

| Test | Method | Results | Specification | Pass/Fail | Completed | Lab |
|---|---|---|---|---|---|---|
| Aerobic Plate Count (mL) | USP <61> | <100 cfu/mL | ≤1000 cfu/mL | PASS | 06/29/2022 | FOCUSPA |
| Yeast and Mold Count (mL) | USP <61> | <100 cfu/mL | ≤100 cfu/mL | PASS | 06/29/2022 | FOCUSPA |

**Sample No.** 22-02526-02
**Product Name:** B. ovatus
**Lot Number:** 2022-03-18-MTC01.02
**Comments:** NA
**Sample Received Date:** June 24, 2022
**Sample Description:** NA

| Test | Method | Results | Specification | Pass/Fail | Completed | Lab |
|---|---|---|---|---|---|---|
| Aerobic Plate Count (mL) | USP <61> | <100 cfu/mL | ≤1000 cfu/mL | PASS | 06/29/2022 | FOCUSPA |
| Yeast and Mold Count (mL) | USP <61> | <100 cfu/mL | ≤100 cfu/mL | PASS | 06/29/2022 | FOCUSPA |

**Sample No.** 22-02526-03
**Product Name:** B. longum
**Lot Number:** 2022-03-25-MTC01.03
**Comments:** NA
**Sample Received Date:** June 24, 2022
**Sample Description:** NA

| Test | Method | Results | Specification | Pass/Fail | Completed | Lab |
|---|---|---|---|---|---|---|
| Aerobic Plate Count (mL) | USP <61> | <100 cfu/mL | ≤1000 cfu/mL | PASS | 06/29/2022 | FOCUSPA |
| Yeast and Mold Count (mL) | USP <61> | <100 cfu/mL | ≤100 cfu/mL | PASS | 06/29/2022 | FOCUSPA |

**Sample No.** 22-02526-04
**Product Name:** B. thetaiotaomicron
**Lot Number:** 2022-03-25-MTC01.04
**Comments:** NA
**Sample Received Date:** June 24, 2022
**Sample Description:** NA

| Test | Method | Results | Specification | Pass/Fail | Completed | Lab |
|---|---|---|---|---|---|---|
| Aerobic Plate Count (mL) | USP <61> | <100 cfu/mL | ≤1000 cfu/mL | PASS | 06/29/2022 | FOCUSPA |
| Yeast and Mold Count (mL) | USP <61> | <100 cfu/mL | ≤100 cfu/mL | PASS | 06/29/2022 | FOCUSPA |

FOCUS Laboratories Pennsylvania is ISO/IEC 17025:2017 accredited by PJLA in the field of testing, Accreditation #77499
†indicated specific test is outside the scope of FOCUS Pennsylvania ISO accreditation, however FOCUS is an FDA registered cGMP laboratory.
This document is issued under the company's terms and conditions which were agreed upon during sample submission. FOCUS Laboratories' liability regarding this analysis is limited to the cost of the tests associated with this Test Report. The results are the property of the client listed above and FOCUS Laboratories. These results only relate to items tested above and apply to the samples as received. This report shall not be reproduced except in full without approval of FOCUS Laboratories.

**Sample No.** 22-02526-05
**Product Name:** B. vulgatus
**Lot Number:** 2022-04-01-MTC01.05
**Comments:** NA
**Sample Received Date:** June 24, 2022
**Sample Description:** NA

| Test | Method | Results | Specification | Pass/Fail | Completed | Lab |
|---|---|---|---|---|---|---|
| Aerobic Plate Count (mL) | USP <61> | <100 cfu/mL | ≤1000 cfu/mL | PASS | 06/29/2022 | FOCUSPA |
| Yeast and Mold Count (mL) | USP <61> | <100 cfu/mL | ≤100 cfu/mL | PASS | 06/29/2022 | FOCUSPA |

**Sample No.** 22-02526-06
**Product Name:** C. aerofaciens
**Lot Number:** 2022-04-08-MTC01.06
**Comments:** NA
**Sample Received Date:** June 24, 2022
**Sample Description:** NA

| Test | Method | Results | Specification | Pass/Fail | Completed | Lab |
|---|---|---|---|---|---|---|
| Aerobic Plate Count (mL) | USP <61> | <100 cfu/mL | ≤1000 cfu/mL | PASS | 06/29/2022 | FOCUSPA |
| Yeast and Mold Count (mL) | USP <61> | <100 cfu/mL | ≤100 cfu/mL | PASS | 06/29/2022 | FOCUSPA |

**Sample No.** 22-02526-07
**Product Name:** P. distasonis
**Lot Number:** 2022-04-08-MTC01.08
**Comments:** NA
**Sample Received Date:** June 24, 2022
**Sample Description:** NA

| Test | Method | Results | Specification | Pass/Fail | Completed | Lab |
|---|---|---|---|---|---|---|
| Aerobic Plate Count (mL) | USP <61> | <100 cfu/mL | ≤1000 cfu/mL | PASS | 06/29/2022 | FOCUSPA |
| Yeast and Mold Count (mL) | USP <61> | <100 cfu/mL | ≤100 cfu/mL | PASS | 06/29/2022 | FOCUSPA |

**Sample No.** 22-02526-08
**Product Name:** B. adolescentis
**Lot Number:** 2022-04-15-MTC01.08
**Comments:** NA
**Sample Received Date:** June 24, 2022
**Sample Description:** NA

| Test | Method | Results | Specification | Pass/Fail | Completed | Lab |
|---|---|---|---|---|---|---|
| Aerobic Plate Count (mL) | USP <61> | <100 cfu/mL | ≤1000 cfu/mL | PASS | 06/29/2022 | FOCUSPA |
| Yeast and Mold Count (mL) | USP <61> | <100 cfu/mL | ≤100 cfu/mL | PASS | 06/29/2022 | FOCUSPA |

**Sample No.** 22-02526-09
**Product Name:** P. merdae
**Lot Number:** 2022-05-20-MTC01.09
**Comments:** NA
**Sample Received Date:** June 24, 2022
**Sample Description:** NA

| Test | Method | Results | Specification | Pass/Fail | Completed | Lab |
|---|---|---|---|---|---|---|
| Aerobic Plate Count (mL) | USP <61> | <100 cfu/mL | ≤1000 cfu/mL | PASS | 06/29/2022 | FOCUSPA |
| Yeast and Mold Count (mL) | USP <61> | <100 cfu/mL | ≤100 cfu/mL | PASS | 06/29/2022 | FOCUSPA |

**Sample No.** 22-02526-10
**Product Name:** C. comes
**Lot Number:** 2022-04-15-MTC01.10
**Comments:** NA
**Sample Received Date:** June 24, 2022
**Sample Description:** NA

| Test | Method | Results | Specification | Pass/Fail | Completed | Lab |
|---|---|---|---|---|---|---|
| Aerobic Plate Count (mL) | USP <61> | <100 cfu/mL | ≤1000 cfu/mL | PASS | 06/29/2022 | FOCUSPA |
| Yeast and Mold Count (mL) | USP <61> | <100 cfu/mL | ≤100 cfu/mL | PASS | 06/29/2022 | FOCUSPA |

**Sample No.** 22-02526-11
**Product Name:** E. rectale
**Lot Number:** 2022-05-20-MTC01.11
**Comments:** NA
**Sample Received Date:** June 24, 2022
**Sample Description:** NA

| Test | Method | Results | Specification | Pass/Fail | Completed | Lab |
|---|---|---|---|---|---|---|
| Aerobic Plate Count (mL) | USP <61> | <100 cfu/mL | ≤1000 cfu/mL | PASS | 06/29/2022 | FOCUSPA |
| Yeast and Mold Count (mL) | USP <61> | <100 cfu/mL | ≤100 cfu/mL | PASS | 06/29/2022 | FOCUSPA |

**Sample No.** 22-02526-12
**Product Name:** B. caccae
**Lot Number:** 2022-05-27-MTC01.12
**Comments:** NA
**Sample Received Date:** June 24, 2022
**Sample Description:** NA

| Test | Method | Results | Specification | Pass/Fail | Completed | Lab |
|---|---|---|---|---|---|---|
| Aerobic Plate Count (mL) | USP <61> | <100 cfu/mL | ≤1000 cfu/mL | PASS | 06/29/2022 | FOCUSPA |
| Yeast and Mold Count (mL) | USP <61> | <100 cfu/mL | ≤100 cfu/mL | PASS | 06/29/2022 | FOCUSPA |

**Sample No.** 22-02526-13
**Product Name:** D. longicatena
**Lot Number:** 2022-05-06-MTC01.13
**Comments:** NA
**Sample Received Date:** June 24, 2022
**Sample Description:** NA

| Test | Method | Results | Specification | Pass/Fail | Completed | Lab |
|---|---|---|---|---|---|---|
| Aerobic Plate Count (mL) | USP <61> | <100 cfu/mL | ≤1000 cfu/mL | PASS | 06/29/2022 | FOCUSPA |
| Yeast and Mold Count (mL) | USP <61> | <100 cfu/mL | ≤100 cfu/mL | PASS | 06/29/2022 | FOCUSPA |

**Sample No.** 22-02526-14
**Product Name:** O. splanchnicus
**Lot Number:** 2022-04-22-MTC01.14
**Comments:** NA
**Sample Received Date:** June 24, 2022
**Sample Description:** NA

| Test | Method | Results | Specification | Pass/Fail | Completed | Lab |
|---|---|---|---|---|---|---|
| Aerobic Plate Count (mL) | USP <61> | <100 cfu/mL | ≤1000 cfu/mL | PASS | 06/29/2022 | FOCUSPA |
| Yeast and Mold Count (mL) | USP <61> | <100 cfu/mL | ≤100 cfu/mL | PASS | 06/29/2022 | FOCUSPA |

**Sample No.** 22-02526-15
**Product Name:** B. cellulosilyticus
**Lot Number:** 2022-04-29-MTC01.15
**Comments:** NA
**Sample Received Date:** June 24, 2022
**Sample Description:** NA

| Test | Method | Results | Specification | Pass/Fail | Completed | Lab |
|---|---|---|---|---|---|---|
| Aerobic Plate Count (mL) | USP <61> | <100 cfu/mL | ≤1000 cfu/mL | PASS | 06/29/2022 | FOCUSPA |
| Yeast and Mold Count (mL) | USP <61> | <100 cfu/mL | ≤100 cfu/mL | PASS | 06/29/2022 | FOCUSPA |

**Sample No.** 22-02526-16
**Product Name:** B. pseudocatenulatum
**Lot Number:** 2022-04-29-MTC01.16
**Comments:** NA
**Sample Received Date:** June 24, 2022
**Sample Description:** NA

| Test | Method | Results | Specification | Pass/Fail | Completed | Lab |
|---|---|---|---|---|---|---|
| Aerobic Plate Count (mL) | USP <61> | <100 cfu/mL | ≤1000 cfu/mL | PASS | 06/29/2022 | FOCUSPA |
| Yeast and Mold Count (mL) | USP <61> | <100 cfu/mL | ≤100 cfu/mL | PASS | 06/29/2022 | FOCUSPA |

**Reference Information:**

**Laboratories:**

FOCUSPA : Focus Laboratories Pennsylvania, 177 N Commerce Way, Bethlehem, PA, 18017 United States

Reviewed / Approved by:

Nathan Evans, Quality Specialist

Signed Date: June 29, 2022

### FOCUS Labs USP<61> DP report

FOCUS LABORATORIES

### Certificate of Analysis No. 22-03285 v1

**Customer:** Icahn School of Medicine
**Address:** 1470 Madison Ave, Fl. 8 RM# 201
New York City, NY 10029
**Date Authorized:** August 29, 2022

**Contact:** Lukas Bethlehem
**Total Number of Pages:** 1

#### Test Results

**Sample No.** 22-03285-01
**Product Name:** DP-High
**Lot Number:** 2022-08-12-MTC01-DP-QC High Dose 2
**Comments:** NA
**Sample Received Date:** August 18, 2022
**Sample Description:** NA

| Test | Method | Results | Specification | Pass/Fail | Completed | Lab |
|---|---|---|---|---|---|---|
| Aerobic Plate Count (cfu/mL) | USP <61> | <100 cfu/mL | ≤1000 cfu/mL | PASS | 08/29/2022 | FOCUSPA |
| Yeast & Mold (cfu/mL) | USP <61> | <100 cfu/mL | ≤100 cfu/mL | PASS | 08/29/2022 | FOCUSPA |

**Sample No.** 22-03285-02
**Product Name:** DP-Low
**Lot Number:** 2022-08-12-MTC01-DP-QC Low Dose 2
**Comments:** NA
**Sample Received Date:** August 18, 2022
**Sample Description:** NA

| Test | Method | Results | Specification | Pass/Fail | Completed | Lab |
|---|---|---|---|---|---|---|
| Aerobic Plate Count (cfu/mL) | USP <61> | <100 cfu/mL | ≤1000 cfu/mL | PASS | 08/29/2022 | FOCUSPA |
| Yeast & Mold (cfu/mL) | USP <61> | <100 cfu/mL | ≤100 cfu/mL | PASS | 08/29/2022 | FOCUSPA |

##### Reference Information:

**Laboratories:**

FOCUSPA - FOCUS Laboratories PA
177 N Commerce Way
Bethlehem, PA 18017

Reviewed / Approved by:

Nathan Evans, Quality Specialist

Signed Date: August 29, 2022

FOCUS Laboratories Pennsylvania is ISO/IEC 17025:2017 accredited by PJLA in the field of testing, Accreditation #77499

†indicated specific test is outside the scope of FOCUS Pennsylvania ISO accreditation, however FOCUS is an FDA registered cGMP laboratory.
This document is issued under the company's terms and conditions which were agreed upon during sample submission. FOCUS Laboratories' liability regarding this analysis is limited to the cost of the tests associated with this Test Report. The results are the property of the client listed above and FOCUS Laboratories. These results only relate to items tested above and apply to the samples as received. This report shall not be reproduced except in full without approval of FOCUS Laboratories.

### Appendix 5 – MTC01 and FMT manufacturing facilities report

#### Facility Layout

The manufacturing facility is housed in a room where no known bacterial pathogens are grown. Storage of MCB, DS, DP is in a freezer dedicated to microbial therapeutics with a dedicated shelf for defined LBP MCB, DS, and DP. Both the centrifuge and -80 °C freezer are stored outside the manufacturing room. However, all transfers from the -80 °C freezer or centrifuge are in sealed containers that are sprayed with EtOH inside the anaerobic air exchange (which is also sprayed) prior to reentry into the anaerobic environment to minimize contamination risk. The anaerobic coy chamber, -80 °C freezer, and centrifuge are all in an access-controlled building with dedicated security staff on the 1st floor, card access elevators, and card access to the laboratory space. Extensive video recording systems monitor all the entrances and hallways.

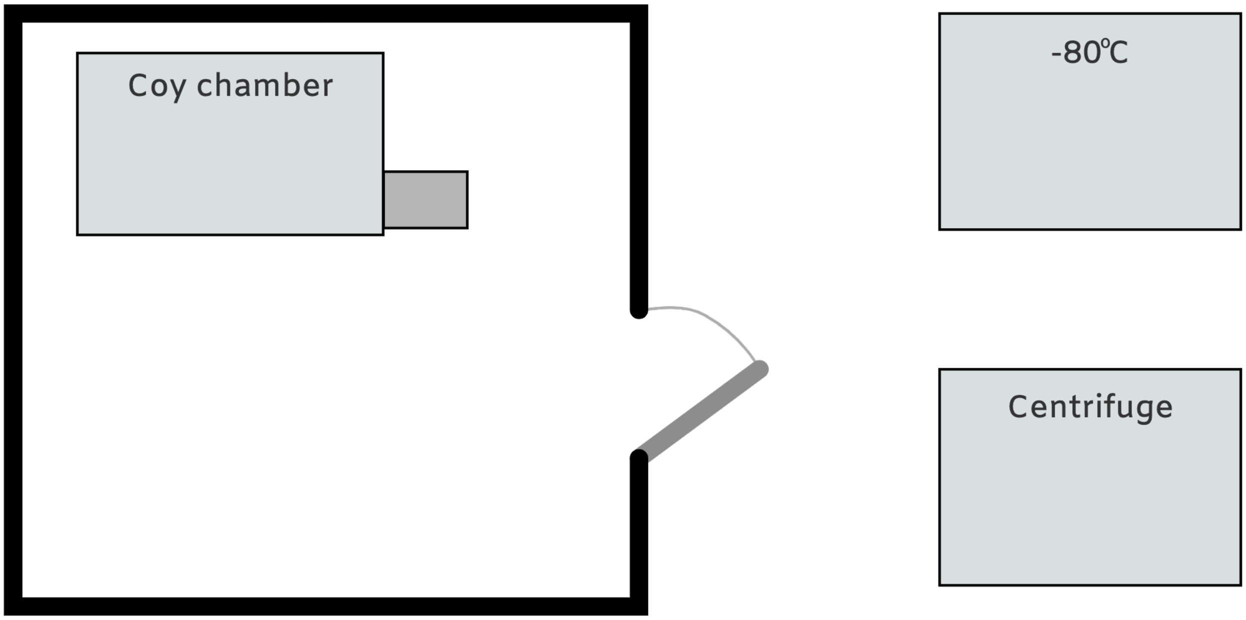

Below is an additional facilities layout with more details of the room housing the manufacturing coy chamber.

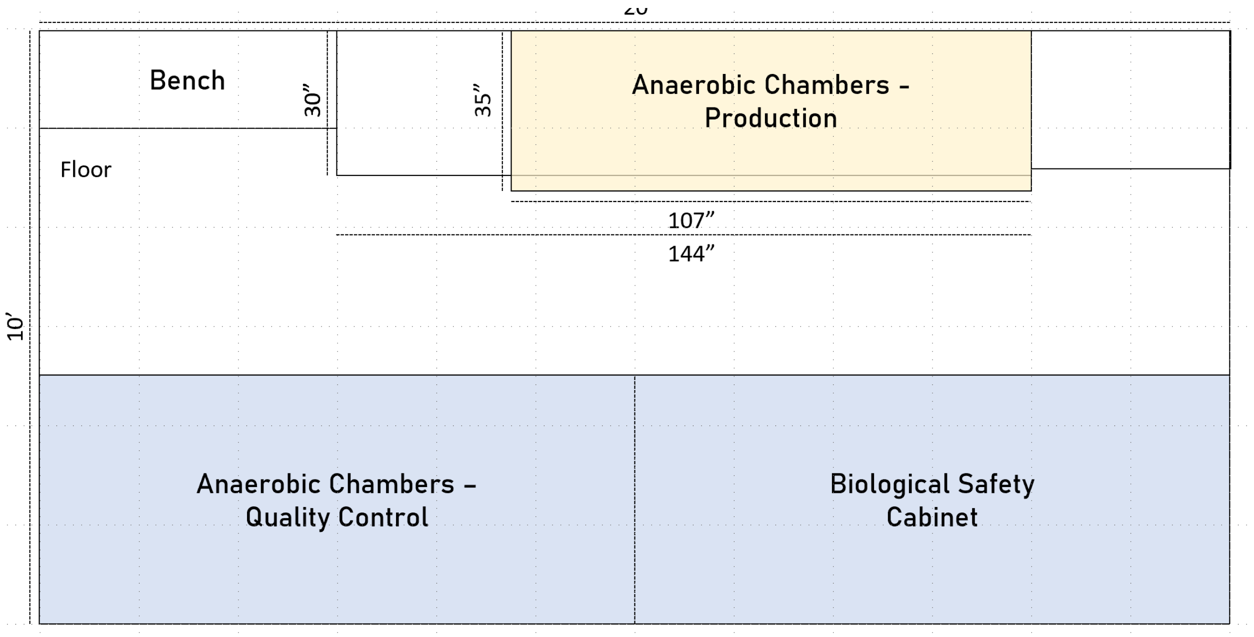

### Anaerobic chambers for defined LBP manufacturing

The anaerobic chamber for manufacturing the MCB, DS, and DP is a custom anaerobic Coy chamber that has been developed in collaboration with Coy Laboratories, a leading source of anaerobic chambers. Although all strain growth for MTC01 will be performed with individually contained and sterilized culture vessels, the risk of contamination of these vessels can be minimized tremendously by operating in a controlled environment (e.g., a hood or clean room). Given the relatively small volumes required for this study, we took advantage of technologies and SOPs from working with germ-free mice to reduce microbial loads in our anaerobic chamber to near zero. The chamber is designed to be easily chemically sterilizable with ports for ionizing and draining sterilant. This system is far cleaner and simpler than a clean room at substantially reduced cost. It had more than enough capacity for this study. Ionized hydrogen peroxide [Sterimist TOMI] is used to sterilize each anaerobic chamber. Since it is sparing of electronic equipment, all materials used for the manufacturing, except the bacterial strain itself, are sterilized in place.

Design schema of the new chamber provided by the engineers at Coy Labs. While photographs of the anaerobic chamber can be seen in Figure 4.

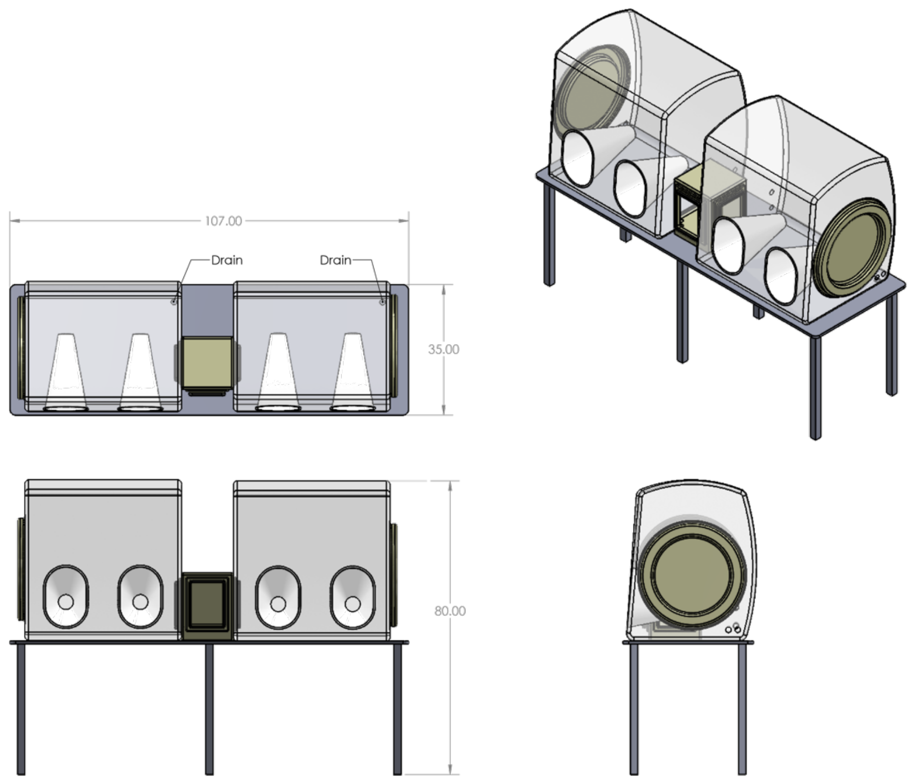

### Facilities for FMT FS processing

The healthy donor fecal slurry used for FMT is produced in a biological safety cabinet sterilized with Quatricide PV-15. A dedicated stomacher paddle blender [Seward Stomacher 400 Evo] is used to homogenize each stool sample and is in the same room. The non-disposable materials for pFS generation are also used for human sample including the generation of fecal slurry for human FMT with healthy human donor stool. We do not culture or isolate spore-forming pathogens with this equipment.

Schema of room for fecal slurry generation for human FMT.

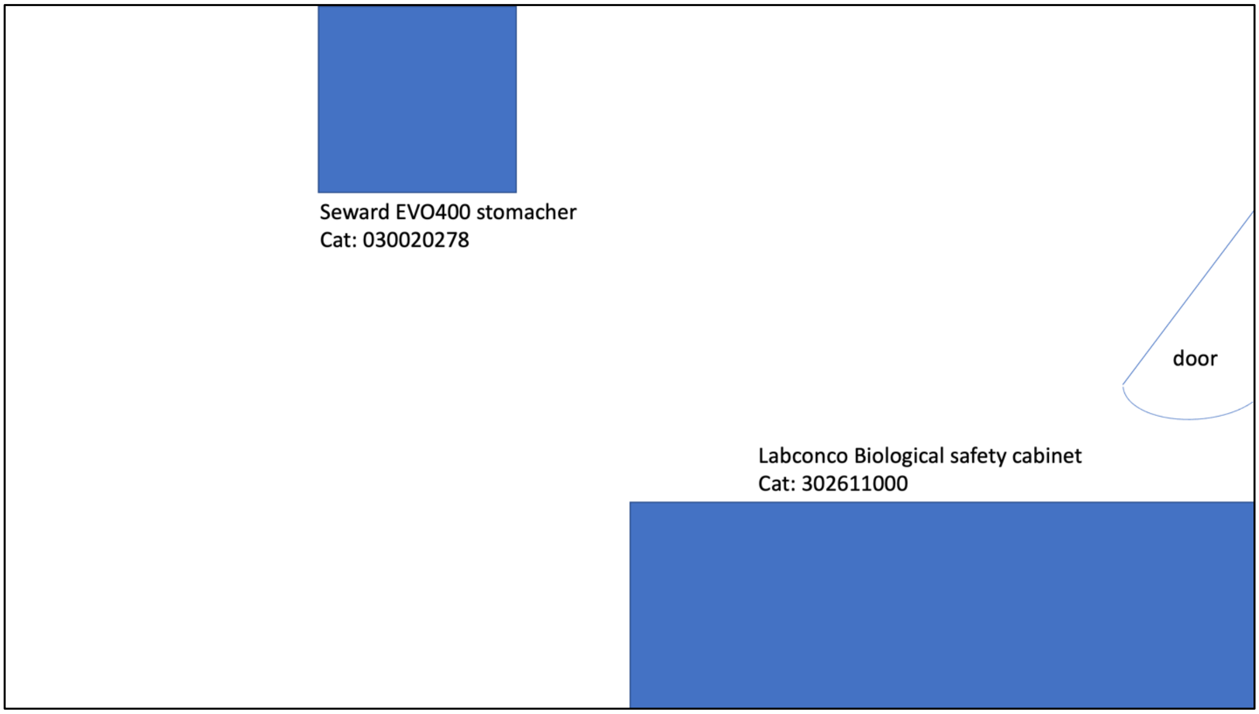

### Appendix 6 – MTC01 potency, identity, and purity results

#### Research Cell Banks (identify, purity)

| StrainID | Species | Plate Incubation Time (hrs) | Colony Bruker Score | Broth Incubation Time (hrs) | Harvest OD600 | Broth Bruker Score |
|---|---|---|---|---|---|---|
| MTC01.01 | Bacteroides uniformis | 24 | 2.35 | 24 | 1.10 | 2.30 |
| MTC01.02 | Bacteroides ovatus | 48 | 1.87 | 24 | 1.49 | 2.00 |
| MTC01.03 | Bifidobacterium longum | 24 | 2.01 | 24 | 1.41 | 2.02 |
| MTC01.04 | Bacteroides thetaiotaomicron | 48 | 2.30 | 24 | 1.44 | 2.16 |
| MTC01.05 | Bacteroides vulgatus | 48 | 2.53 | 24 | 1.15 | 2.40 |
| MTC01.07 | Parabacteroides distasonis | 48 | 2.21 | 24 | 1.27 | 2.15 |
| MTC01.08 | Bifidobacterium adolescentis | 24 | 2.22 | 24 | 1.52 | 1.79 |
| MTC01.09 | Parabacteroides merdae | 72 | 2.00 | 24 | 0.72 | 2.45 |
| MTC01.10 | Coprococcus comes | 72 | 2.14 | 24 | 1.46 | 2.18 |
| MTC01.11 | Eubacterium rectale | 72 | colonies too small for detection | 48 | 0.26 | 2.36 |
| MTC01.12 | Bacteroides caccae | 48 | 2.23 | 24 | 1.31 | 2.27 |
| MTC01.13 | Dorea longicatena | 48 | 1.82 | 24 | 1.13 | 1.62 |
| MTC01.14 | Odoribacter splanchnicus | 48 | 2.37 | 24 | 1.20 | 2.45 |
| MTC01.15 | Bacteroides cellulosilyticus | 48 | 2.14 | 24 | 1.26 | 2.30 |
| MTC01.16 | Bifidobacterium pseudocatenulatum | 48 | 2.14 | 24 | 1.20 | 2.23 |

#### Master Cell Banks (potency, identify, purity)

All MALDI readings identified the correct species.
USP<61> specification (rectally administered product):
$<10^3$ CFU/ml for Total Aerobic Microbial Count and $<10^2$ CFU/ml for Total Combined Yeasts/Molds Count

No colony was detected for all assays

| Strain ID | Species | Culture | Volume [mL] | Incubation Time [h] | $OD_{600}$ | CFU/mL | MALDI IDs (total) | MALDI IDs (>1.8) | USP<61> |
|---|---|---|---|---|---|---|---|---|---|
| MTC01.01 | *B. uniformis* | Starter | 10 | 32 | 1.45 | | 48 | 45 | |
| | | Production | 100 | 21 | 1.24 | 1.20E+09 | 96 | 95 | passed |
| MTC01.02 | *B. ovatus* | Starter | 10 | 33 | 2.68 | | 48 | 48 | |
| | | Production | 100 | 21 | 1.08 | 7.97E+08 | 96 | 91 | passed |
| MTC01.03 | *B. longum* | Starter | 10 | 31 | 2.52 | | 48 | 39 | |
| | | Production | 100 | 21 | 2 | 2.40E+09 | 96 | 76 | passed |
| MTC01.04 | *B. thetaiotaomicron* | Starter | 10 | 31 | 1.8 | | 48 | 46 | |
| | | Production | 100 | 26 | 2.56 | 5.77E+08 | 96 | 96 | passed |
| MTC01.05 | *B. vulgatus* | Starter | 10 | 30 | 1.04 | | 48 | 48 | |
| | | Production | 100 | 25 | 2 | 4.20E+08 | 96 | 93 | passed |
| MTC01.07 | *P. distasonis* | Starter | 10 | 30 | 3.04 | | 48 | 47 | |
| | | Production | 100 | 25 | 0.8 | 6.40E+08 | 96 | 94 | passed |
| MTC01.08 | *B. adolescentis* | Starter | 10 | 28 | 3.72 | | 48 | 40 | |
| | | Production | 100 | 19 | 3.6 | 2.30E+09 | 287 | 57 | passed |
| MTC01.09 | *P. merdae* | Starter | 10 | 32 | 0.48 | | 72 | 72 | |
| | | Production | 100 | 23 | 0.48 | 4.60E+07 | 96 | 96 | passed |
| MTC01.10 | *C. comes* | Starter | 10 | 22 | 1.08 | | 48 | 37 | |
| | | Production | 100 | 10 | 2.36 | 4.50E+07 | 96 | 77 | passed |
| MTC01.11 | *E. rectale* | Starter | 10 | 54 | 1.12 | | 72 | 72 | |
| | | Production | 100 | 18 | 1.04 | 1.00E+08 | 96 | 74 | passed |
| MTC01.12 | *B. caccae* | Starter | 10 | 27 | 2.12 | | 48 | 47 | |
| | | Production | 100 | 27 | 1.76 | 3.53E+07 | 96 | 94 | passed |
| MTC01.13 | *D. longicatena* | Starter | 10 | 31 | | | 168 | 22 | |
| | | Production | 100 | 43 | 6.24 | 1.00E+08 | 96 | 68 | passed |
| MTC01.14 | *O. splanchnicus* | Starter | 10 | 27 | 2.96 | | 48 | 48 | |
| | | Production | 100 | 26 | 2.8 | 1.73E+09 | 96 | 96 | passed |
| MTC01.15 | *B. cellulosilyticus* | Starter | 10 | 44 | 0.84 | | 48 | 47 | |
| | | Production | 100 | 53 | 0.8 | 1.10E+08 | 96 | 82 | passed |
| MTC01.16 | *B. pseudocatenulatum* | Starter | 10 | 31 | 3.08 | | 48 | 40 | |
| | | Production | 100 | 17 | 3.88 | 7.40E+08 | 96 | 92 | passed |

#### Drug Substance (potency, identify, purity)

All MALDI readings identified the correct species.
USP<61> specification (rectally administered product):
$<10^3$ CFU/ml for Total Aerobic Microbial Count and $<10^2$ CFU/ml for Total Combined Yeasts/Molds Count

No colony was detected for all assays

| Strain ID | Species | Culture | Volume [mL] | Incubation Time [h] | $OD_{600}$ | CFU/mL | MALDI IDs (total) | MALDI IDs (>1.8) | USP<61> |
|---|---|---|---|---|---|---|---|---|---|
| MTC01.01 | *B. uniformis* | Starter 1 | | | 0.72 | | | | |
| | | Starter 2 | 100 | 27 | 0.68 | | 24 | 24 | |
| | | Starter 3 | | | 0.88 | | | | |
| | | Production | 1400 | 21 | 2.4 | 6.20E+09 | 96 | 96 | passed |
| MTC01.02 | *B. ovatus* | Starter 1 | | | 1.12 | | | | |
| | | Starter 2 | 100 | 52 | 1.24 | | 24 | 24 | |
| | | Starter 3 | | | 1.28 | | | | |
| | | Production | 1400 | 31 | 1.32 | 9.67E+08 | 96 | 85 | passed |
| MTC01.03 | *B. longum* | Starter 1 | | | 2 | | | | |
| | | Starter 2 | 100 | 17 | 5.2 | | 24 | 24 | |
| | | Starter 3 | | | 8 | | | | |
| | | Production | 1400 | 10 | 1.6 | 1.47E+08 | 96 | 96 | passed |
| MTC01.04 | *B. thetaiotaomicron* | Starter 1 | | | 0.76 | | | | |
| | | Starter 2 | 100 | 26 | 0.8 | | 60 | 43 | |
| | | Starter 3 | | | 0.84 | | | | |
| | | Production | 1400 | 20 | 2.16 | 3.07E+09 | 96 | 79 | passed |
| MTC01.05 | *B. vulgatus* | Starter 1 | | | 2.44 | | | | |
| | | Starter 2 | 100 | 19 | 2.84 | | 24 | 24 | |
| | | Starter 3 | | | 2.16 | | | | |
| | | Production | 1400 | 9 | 4.32 | 3.20E+09 | 96 | 96 | passed |
| MTC01.07 | *P. distasonis* | Starter 1 | | | 0.72 | | | | |
| | | Starter 2 | 100 | 32 | 0.76 | | 24 | 24 | |
| | | Starter 3 | | | 0.92 | | | | |
| | | Production | 1400 | 32 | 1.64 | 2.00E+09 | 96 | 95 | passed |
| MTC01.08 | *B. adolescentis* | Starter 1 | | | 1.2 | | | | |
| | | Starter 2 | 100 | 17 | 1.96 | | 24 | 4 | |
| | | Starter 3 | | | 3.72 | | | | |
| | | Production | 1400 | 7 | 3.32 | 1.27E+09 | 192 | 83 | passed |
| MTC01.09 | *P. merdae* | Starter 1 | | | 0.48 | | | | |
| | | Starter 2 | 100 | 77 | 0.4 | | 24 | 23 | |
| | | Starter 3 | | | 0.2 | | | | |
| | | Production | 1400 | 24 | 0.72 | 2.03E+09 | 96 | 96 | passed |
| MTC01.10 | *C. comes* | Starter 1 | | | 1.4 | | | | |
| | | Starter 2 | 100 | 34 | 3 | | NA | NA | |
| | | Starter 3 | | | 2.16 | | | | |
| | | Production | 1400 | 14 | 4.8 | 3.37E+08 | 96 | 68 | passed |
| MTC01.11 | *E. rectale* | Starter 1 | | | 0.2 | | | | |
| | | Starter 2 | 100 | 119 | 0.68 | | 24 | 11 | |
| | | Starter 3 | | | 0.28 | | | | |
| | | Production | 1400 | 24 | 0.48 | 4.13E+08 | 96 | 77 | passed |
| MTC01.12 | *B. caccae* | Starter 1 | | | 0.68 | | | | |
| | | Starter 2 | 100 | 52 | 0.56 | | 24 | 21 | |
| | | Starter 3 | | | 0.8 | | | | |
| | | Production | 1400 | 24 | 1.36 | 1.60E+09 | 96 | 48 | passed |
| MTC01.13 | *D. longicatena* | Starter 1 | | | NA | | | | |
| | | Starter 2 | 100 | 211 | NA | | 12 | 2 | |
| | | Starter 3 | | | NA | | | | |
| | | Production | 1400 | 48 | 2.84 | 2.90E+08 | 96 | 48 | passed |
| MTC01.14 | *O. splanchnicus* | Starter 1 | | | 4.08 | | | | |
| | | Starter 2 | 100 | 54 | 3.92 | | 24 | 24 | |
| | | Starter 3 | | | 3.92 | | | | |
| | | Production | 1400 | 24 | 3.32 | 7.33E+09 | 96 | 96 | passed |
| MTC01.15 | *B. cellulosilyticus* | Starter 1 | | | 1.56 | | | | |
| | | Starter 2 | 100 | 51 | 1.28 | | 24 | 22 | |
| | | Starter 3 | | | 1.48 | | | | |
| | | Production | 1400 | 20 | 2.88 | 2.90E+09 | 96 | 96 | passed |
| MTC01.16 | *B. pseudocatenulatum* | Starter 1 | | | 1.7 | | | | |
| | | Starter 2 | 100 | 32 | 2.9 | | 24 | 20 | |
| | | Starter 3 | | | 2.6 | | | | |
| | | Production | 1400 | 14 | 4.08 | 3.17E+09 | 96 | 52 | passed |

#### Drug Product (potency, identify, purity)

USP<61> specification (rectally administered product):
<$10^3$ CFU/ml for Total Aerobic Microbial Count and <$10^2$ CFU/ml for Total Combined Yeasts/Molds Count

No colony was detected for all assays

All MALDI readings identified only MTC01 species.

| DP Dose | MALDI IDs (total) | MALDI IDs (>1.8) | USP<61> |
|---|---|---|---|
| High | 288 | 269 | passed |
| Low | 288 | 283 | passed |

Estimated potency of each strain per dose in DP
(based on DS CFU/ml and the number of ml of each strain added to the final DP)

| Strain ID | Species | CFU DP High | CFU DP Low |
|---|---|---|---|
| MTC01.01 | *B. uniformis* | 3.13E+10 | 3.13E+09 |
| MTC01.02 | *B. ovatus* | 3.13E+10 | 3.13E+09 |
| MTC01.03 | *B. longum* | 1.41E+10 | 1.41E+09 |
| MTC01.04 | *B. thetaiotaomicron* | 3.13E+10 | 3.13E+09 |
| MTC01.05 | *B. vulgatus* | 3.13E+10 | 3.13E+09 |
| MTC01.07 | *P. distasonis* | 3.13E+10 | 3.13E+09 |
| MTC01.08 | *B. adolescentis* | 2.28E+10 | 2.28E+09 |
| MTC01.09 | *P. merdae* | 2.59E+10 | 2.59E+09 |
| MTC01.10 | *C. comes* | 2.11E+10 | 2.11E+09 |
| MTC01.11 | *E. rectale* | 3.13E+10 | 3.13E+09 |
| MTC01.12 | *B. caccae* | 1.26E+09 | 1.26E+08 |
| MTC01.13 | *D. longicatena* | 1.60E+10 | 1.60E+10 |
| MTC01.14 | *O. splanchnicus* | 3.13E+10 | 3.13E+09 |
| MTC01.15 | *B. cellulosilyticus* | 3.13E+10 | 3.13E+09 |
| MTC01.16 | *B. pseudocatenulatum* | 3.13E+10 | 3.13E+09 |

### Appendix 7 – MTC01 virulence factors and toxins

Contigs from the draft SPAdes assemblies[46] were reordered into scaffolds using Mauve[47] based the closest complete NCBI reference sequence genome as determined by fastANI[48]. Genomes were annotated using Prokka[49] and the resulting translated coding sequences were used for determination of the presence of virulence and toxins encoding genes using BLAST[50] with Victors[51], VFDB[52] and PATRIC_VF[53] as reference databases.

| Strain ID | Species | Property | DB | Gene | Identity (%) | E-Value | Comment | doi |
|---|---|---|---|---|---|---|---|---|
| MTC01.01 | Bacteroides uniformis | Virulence Factor | Victors | SP_0320 | 82.759 | 8.14E-162 | Oxioreductase | 10.1046/j.1365-2958.2002.03106.x |
| MTC01.02 | Bacteroides ovatus | Virulence Factor | Victors | SP_0320 | 80.077 | 4.43E-156 | Oxioreductase | 10.1046/j.1365-2958.2002.03106.x |
| MTC01.03 | Bifidobacterium longum | NA | NA | NA | NA | NA | #N/A | #N/A |
| MTC01.04 | Bacteroides thetaiotaomicron | Virulence Factor | Victors | SP_0320 | 82.375 | 2.09E-161 | Oxioreductase | 10.1046/j.1365-2958.2002.03106.x |
| MTC01.05 | Bacteroides vulgatus | Virulence Factor | VFDB | cps4L | 80.916 | 0 | Single gene, not full cps4 locus | 10.1099/jmm.0.000573 |
| MTC01.05 | Bacteroides vulgatus | Virulence Factor | Victors | SP_0320 | 81.609 | 5.92E-161 | Oxioreductase | 10.1046/j.1365-2958.2002.03106.x |
| MTC01.07 | Parabacteroides distasonis | NA | NA | NA | NA | NA | #N/A | #N/A |
| MTC01.08 | Bifidobacterium adolescentis | NA | NA | NA | NA | NA | #N/A | #N/A |
| MTC01.09 | Parabacteroides merdae | Virulence Factor | VFDB | cps4L | 80.153 | 0 | Single gene, not full cps4 locus | 10.1099/jmm.0.000573 |
| MTC01.09 | Parabacteroides merdae | Virulence Factor | Victors | SP_0320 | 84.231 | 3.53E-167 | Oxioreductase | 10.1046/j.1365-2958.2002.03106.x |
| MTC01.10 | Coprococcus comes | NA | NA | NA | NA | NA | #N/A | #N/A |
| MTC01.11 | Eubacterium rectale | NA | NA | NA | NA | NA | #N/A | #N/A |
| MTC01.12 | Bacteroides caccae | Virulence Factor | VFDB | cps4L | 81.795 | 0 | Single gene, not full cps4 locus | 10.1099/jmm.0.000573 |
| MTC01.12 | Bacteroides caccae | Virulence Factor | Victors | SP_0320 | 81.609 | 1.32E-159 | Oxioreductase | 10.1046/j.1365-2958.2002.03106.x |
| MTC01.13 | Dorea longicatena | NA | NA | NA | NA | NA | #N/A | #N/A |
| MTC01.14 | Odoribacter splanchnicus | NA | NA | NA | NA | NA | #N/A | #N/A |
| MTC01.15 | Bacteroides cellulosilyticus | Virulence Factor | VFDB | cps4L | 80.662 | 0 | Single gene, not full cps4 locus | 10.1099/jmm.0.000573 |
| MTC01.15 | Bacteroides cellulosilyticus | Virulence Factor | Victors | SP_0320 | 83.908 | 1.02E-166 | Oxioreductase | 10.1046/j.1365-2958.2002.03106.x |
| MTC01.16 | Bifidobacterium pseudocatenulatum | NA | NA | NA | NA | NA | #N/A | #N/A |

### Appendix 8 – MTC01 predicted antibiotic resistance genes and mobile genetic elements

Contigs from the draft SPAdes assemblies[46] were reordered into scaffolds using Mauve[47] based the closest complete NCBI reference sequence genome as determined by fastANI[48]. Genomes were annotated using Prokka[49] and the resulting translated coding sequences were used for prediction antibiotic resistance (AR) genetic elements using CARD[54]. The presence of AR genes on genomic islands and the potential for horizontal gene transfer was determined by SIGI-HMM[55] and IslandPath[56].

| Strain ID | Species | Gene | Drug Class | Resistance Mechanism | Comment |
|---|---|---|---|---|---|
| MTC01.01 | Bacteroides uniformis | adeF | multiple antibiotics | efflux | |
| MTC01.01 | Bacteroides uniformis | tetQ | tetracycline antibiotic | target protection | |
| MTC01.01 | Bacteroides uniformis | CblA-1 | cephalosporin | inactivation | |
| MTC01.02 | Bacteroides ovatus | adeF | multiple antibioticsacterial free fatty acids | efflux | |
| MTC01.02 | Bacteroides ovatus | tetQ | tetracycline antibiotic | target protection | |
| MTC01.03 | Bifidobacterium longum | rpoB mutant | rifamycin antibiotic | target alteration; target replacement | |
| MTC01.04 | Bacteroides thetaiotaomicron | adeF | multiple antibioticsacterial free fatty acids | efflux | |
| MTC01.04 | Bacteroides thetaiotaomicron | tetQ | tetracycline antibiotic | target protection | |
| MTC01.05 | Bacteroides vulgatus | adeF | multiple antibioticsacterial free fatty acids | efflux | |
| MTC01.05 | Bacteroides vulgatus | tetQ | tetracycline antibiotic | target protection | |
| MTC01.05 | Bacteroides vulgatus | ErmF | macrolide antibiotic; lincosamide antibiotic; streptogramin antibiotic | target alteration | |
| MTC01.05 | Bacteroides vulgatus | vanYF | glycopeptide antibiotic | target alteration | Not a full operon, not on a MGE |
| MTC01.07 | Parabacteroides distasonis | tetB(P) | tetracycline antibiotic | target protection | |
| MTC01.07 | Parabacteroides distasonis | adeF | multiple antibioticsacterial free fatty acids | efflux | |
| MTC01.08 | Bifidobacterium adolescentis | MuxC | multiple antibioticsacterial free fatty acids | efflux | |
| MTC01.08 | Bifidobacterium adolescentis | tetO | tetracycline antibiotic | target protection | |
| MTC01.08 | Bifidobacterium adolescentis | rpoB mutant | rifamycin antibiotic | target alteration; target replacement | |
| MTC01.09 | Parabacteroides merdae | adeF | multiple antibioticsacterial free fatty acids | efflux | |
| MTC01.09 | Parabacteroides merdae | Mef(En2) | multiple antibiotics | efflux | |
| MTC01.09 | Parabacteroides merdae | tetQ | tetracycline antibiotic | target protection | |
| MTC01.11 | Eubacterium rectale | AAC(3)-IId | aminoglycoside antibiotic | inactivation | |
| MTC01.12 | Bacteroides caccae | tetQ | tetracycline antibiotic | target protection | |
| MTC01.12 | Bacteroides caccae | adeF | multiple antibioticsacterial free fatty acids | efflux | |
| MTC01.13 | Dorea longicatena | ErmB | macrolide antibiotic; lincosamide antibiotic; streptogramin antibiotic | target alteration | |
| MTC01.13 | Dorea longicatena | tetM | tetracycline antibiotic | target protection | |

| Strain ID | Species | Gene | Drug Class | Resistance Mechanism | Comment |
|---|---|---|---|---|---|
| MTC01.14 | Odoribacter splanchnicus | adeF | multiple antibioticsacterial free fatty acids | efflux | |
| MTC01.14 | Odoribacter splanchnicus | tetQ | tetracycline antibiotic | target protection | |
| MTC01.14 | Odoribacter splanchnicus | ErmF | macrolide antibiotic; lincosamide antibiotic; streptogramin antibiotic | target alteration | |
| MTC01.14 | Odoribacter splanchnicus | tetX | glycylcycline; tetracycline antibiotic | inactivation | |
| MTC01.14 | Odoribacter splanchnicus | aadS | aminoglycoside antibiotic | inactivation | |
| MTC01.15 | Bacteroides cellulosilyticus | tetQ | tetracycline antibiotic | target protection | |
| MTC01.15 | Bacteroides cellulosilyticus | ANT(4')-Ib | aminoglycoside antibiotic | inactivation | |
| MTC01.15 | Bacteroides cellulosilyticus | adeF | multiple antibioticsacterial free fatty acids | efflux | |
| MTC01.16 | Bifidobacterium pseudocatenulatum | MuxC | multiple antibioticsacterial free fatty acids | efflux | |
| MTC01.16 | Bifidobacterium pseudocatenulatum | vanC | glycopeptide antibiotic | target alteration | Not a full operon, not on a MGE |
| MTC01.16 | Bifidobacterium pseudocatenulatum | dfrF | diaminopyrimidine antibiotic | target replacement | |
| MTC01.16 | Bifidobacterium pseudocatenulatum | rpoB mutant | rifamycin antibiotic | target alteration; target replacement | |
| MTC01.16 | Bifidobacterium pseudocatenulatum | tet(W/N/W) | tetracycline antibiotic | target protection | |

Contigs from the draft SPAdes assemblies[46] were reordered into scaffolds using Mauve[47] based the closest complete NCBI reference sequence genome as determined by fastANI[48]. Genomes were annotated using Prokka[49] and the resulting translated coding sequences were used for prediction antibiotic resistance (AR) genetic elements using CARD[54] (antibiotic resistance gene indicated as text). The presence of AR genes on the full genome (grey) on genomic islands and the potential for horizontal gene transfer was determined by SIGI-HMM[55] (red) and IslandPath[56] (green).

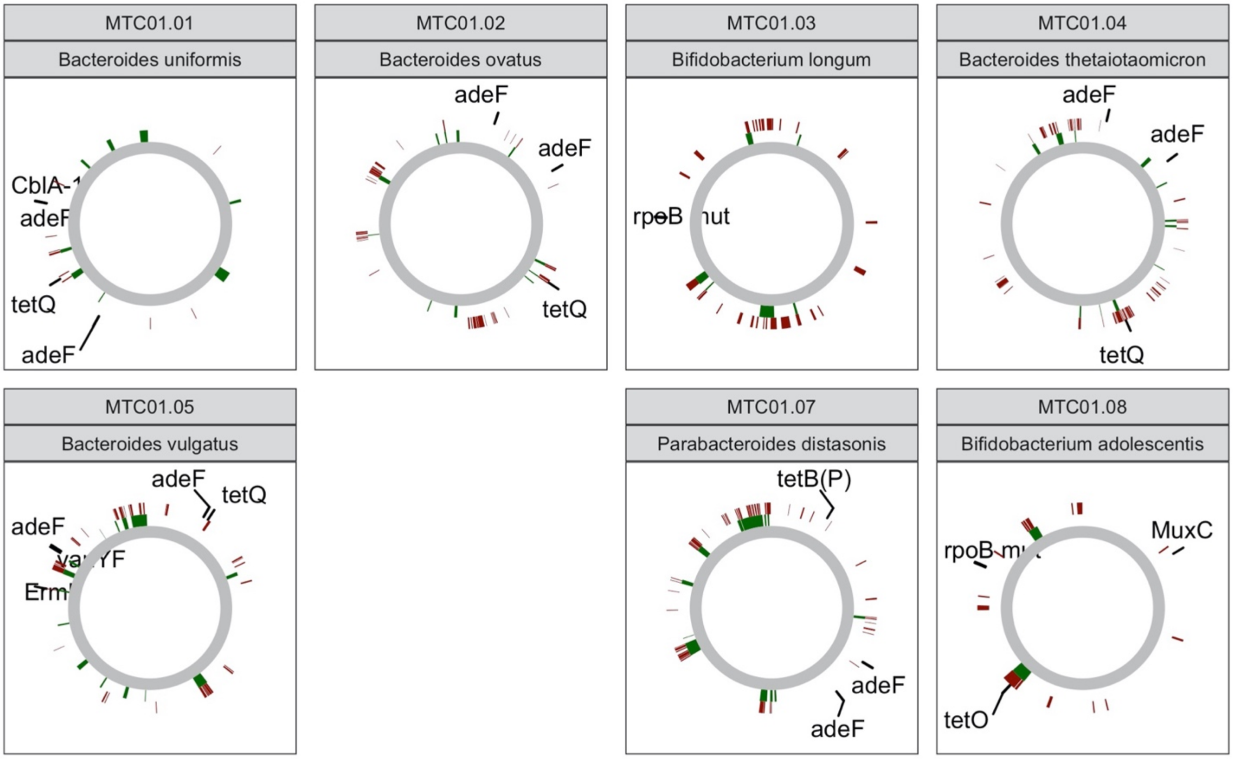

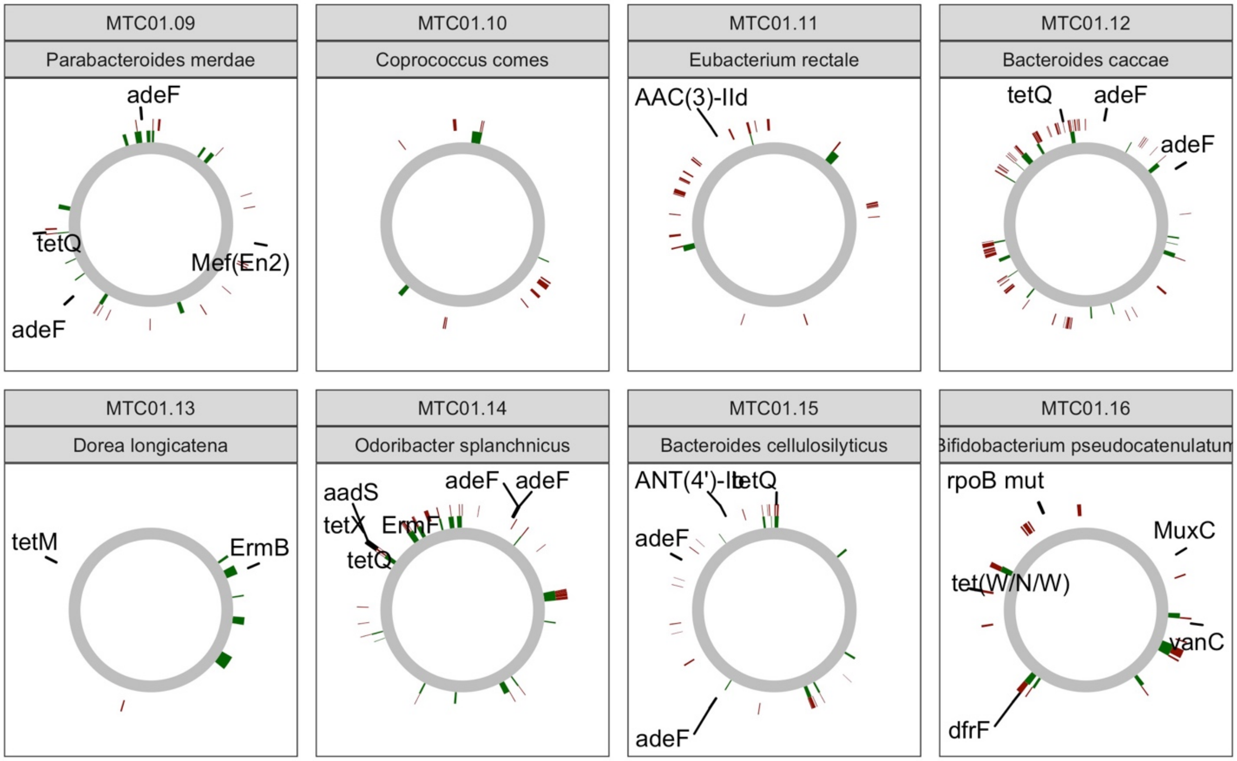
MTC01.09
Parabacteroides merdae
adeF
tetQ
Mef(En2)
adeF
MTC01.10
Coprococcus comes
MTC01.11
Eubacterium rectale
AAC(3)-IId
MTC01.12
Bacteroides caccae
tetQ
adeF
adeF
MTC01.13
Dorea longicatena
tetM
ErmB
MTC01.14
Odoribacter splanchnicus
aadS
adeF
adeF
tetX
ErmF
tetQ
MTC01.15
Bacteroides cellulosilyticus
ANT(4')-Ib
tetQ
adeF
adeF
MTC01.16
ifidobacterium pseudocatenulatu
rpoB mut
MuxC
tet(W/N/W)
vanC
dfrF

### Appendix 9 – MTC01 and FMT growth media and formulation buffers

#### Growth Media 1: LYHBHIv4 – used for RCB generation

Media is a supplemented BHI media modified from Sokol et al, 2018[57]

**BHI**

| Concentration | | Reagent | Amount | |
|---|---|---|---|---|
| 12.5 | g/L | Calf brain, infusion from | 12.5 | g |
| 5 | g/L | Beef heart, infusion from | 5 | g |
| 10 | g/L | Pancreatic digest of casein | 10 | g |
| 2 | g/L | Glucose | 2 | g |
| 5 | g/L | Sodium chloride | 5 | g |
| 2.5 | g/L | Disodium phosphate | 2.5 | g |

| Concentration | | Reagent | Amount | |
|---|---|---|---|---|
| 1 | % | Menadione | 0.2 | g |

| Concentration | | Reagent | Amount | |
|---|---|---|---|---|
| 37 | g/L | BHI | 37 | g |
| 0.5 | % | Yeast extract | 5 | g |
| 1 | g/L | D-Xylose | 1 | g |
| 1 | g/L | D-Fructose | 1 | g |
| 1 | g/L | D-Glucose | 1 | g |
| 1 | g/L | D-Galactose | 1 | g |
| 0.5 | g/L | N-Acetylglucosamine | 0.5 | g |
| 0.5 | g/L | L-Arabinose | 0.5 | g |
| 1 | g/L | Cellobiose | 1 | g |
| 1 | g/L | Maltose | 1 | g |
| 1 | g/L | Sucrose | 1 | g |
| 0.5 | g/L | Cysteine | 0.5 | g |
| 1 | g/L | Malic acid | 1 | g |
| 1 | g/L | Sodium Sulfate | 1 | g |
| 0.005 | g/L | hemin | 0.005 | g |
| 0.05 | % | Tween 80 | 500 | ul |
| 0.1 | % | Vitamin K solution | 1 | mL |
| 0.5 | % | Resazurin | 2 | mL |
| 0.1 | M | MOPS | 20.92633 | g |
| | | pH 7.2 - KOH, H20 fill to | 1 | L |

Filter sterilize

#### Growth Media 2: LYH Vegitone Infusion Broth (LYH_VIB) – used for growth of MCB and DS

This animal-free media is based on LYHBHI but with Vegitone Infusion Broth instead of BHI and no hemin.

LYH_VIB Medium

| Reagent | Amount [g/L] |
|---|---|
| Vegitone infusion broth (VIB) | 37 |
| Yeast extract | 5 |
| Monosaccharide mix | 4 |
| - D-xylose | 1 |
| - D-fructose | 1 |
| - D-glucose | 1 |
| - D-galactose | 1 |
| - N-acetylglucosamine | 0.5 |
| - L-arabinose | 0.5 |
| Disaccharide mix | 3 |
| - D-Cellobiose | 1 |
| - D-Maltose | 1 |
| - Sucrose | 1 |
| L-cysteine hydrochloride | 0.5 |
| L-Malic acid | 1 |
| Sodium sulfate | 2 |
| MOPS | 20.9 |
| | **Volume [mL/L]** |
| Vitamin-K solution | 1 |
| Tween 80 | 0.5 |
| $H_2O$ (WIFI) | Adjust to 1 L |
| | pH 7.2 (NaOH) |

Vitamin-K solution

| Component | Volume [mg/mL] |
|---|---|
| Menadione | 1 |
| Ethanol (190 Proof) | Adjust to 1 mL |

#### Formulation Buffer / Cryogenic medium – used as the freezing media for the concentrated DS and as the formulation buffer for the DP

This media contains PBS buffered solution (9 g NaCl/L) with 30% glycerol for cryogenic preservation of the bacterial strains as well as cysteine to reduce $O_2$ in the media. All components are USP grade.

Cryogenic Buffer

| Reagent | Amount [g/L] |
|---|---|
| Sodium chloride [USP grade]* | 9 |
| L-cysteine hydrochloride [USP grade] | 0.5 |
| Glycerol [USP grade] | 300 |
| | **Volume [mL/L]** |
| Phosphate buffered saline *or* solution (pH 7.5) [USP grade] | Adjust to 1 L |

* NaCl only needs to be added if phosphate buffered solution is used instead of phosphate buffered saline.

#### Wash Media – used to wash the DS grown in LYH_VIB

This media contains PBS buffered solution (9 g NaCl/L) with cysteine to reduce $O_2$ in the media. All components are USP grade. It is used to wash the DS culture grown in LYH_VIB media.

Wash Media

| Reagent | Amount [g/L] |
|---|---|
| Sodium chloride [USP grade]* | 9 |
| L-cysteine hydrochloride [USP grade] | 0.5 |
| | **Volume [mL/L]** |
| Phosphate buffered saline *or* solution (pH 7.5) [USP grade] | Adjust to 1 L |

* NaCl only needs to be added if phosphate buffered solution is used instead of phosphate buffered saline.

### SOP for Formulation buffer

| Icahn School of Medicine at Mount Sinai — Microbiome Translational Center | **Microbiome Translational Center Standard Operating Procedures** |
|---|---|
| **Title: Fecal Slurry Cryopreservation Media for Fecal Microbiota Transplantation** | **SOP No: MTC-SOP-FS-0003**<br>**Revision: 0** |
| | **Effective Date: 10/16/2022**<br>**Supersedes Date: NEW** |

| **Written by:** | | **Date:** |
|---|---|---|
| Lukas Bethlehem, PhD | | |
| **Reviewed By:** | | **Date:** |
| Jeremiah Faith, PhD<br>Faculty Director | | |
| **Approved By:** | | **Date:** |
| Ilaria Mogno, PhD<br>Quality Assurance Specialist | | |

#### 1.0 INTRODUCTION

The purpose of this SOP is to provide instructions for the preparation of cryogenic media for the generation of fecal slurry (FS) for use in Fecal Microbiota Transplantation (FMT).

#### 2.0 RESPONSIBLE PERSONNEL

2.1 The Facility Director is responsible for identifying new SOPs to be written, drafting new SOPs, suggesting revisions to existing SOPs, reviewing and approving all SOPs, and ensuring that all laboratory personnel is properly trained and following all SOPs.

2.2 The Quality Assurance (QA) Lead is responsible for coordinating the preparation, approval, distribution, and maintenance of this SOP, for suggesting revisions to existing SOPs, and for ensuring that all SOPs are being followed by the required personnel.

2.3 The Approver can and should name representatives, who are qualified to take over the duties in case of an emergency or during absence of the Approver/reviewer.

2.4 Technicians and other Lab personal are responsible for performing the procedures described in this SOP.

#### 3.0 DEFINITIONS

3.1 SOP = Standard Operating Procedure
3.2 QA = Quality Assurance
3.3 MTC = Microbiome Translational Center

3.4 FMT = Fecal Microbiota Transplantation
3.5 FS = Fecal Slurry
3.6 BSC = biological safety cabinet

#### 4.0 SAFETY

4.1 Wear protective gloves and lab coat while performing this procedure.
4.2 Perform all steps with the dedicated equipment for production of FS and follow instructions carefully to avoid contamination of the product or exposure to the operator.

#### 5.0 MATERIALS

5.1 Equipment

| Item | Manufacturer | Catalog No. |
|---|---|---|
| Biological Safety Cabinet (BSC) | Labconco | 302619100 |
| Pipetting Controller | | |
| Balance | Mettler Toledo | XS6002S |
| Fine Balance | Mettler Toledo | XP205 |

5.2 Supplies

| Item | Manufacturer | Catalog No. |
|---|---|---|
| Serological Pipets, 50 mL | | |
| Media Bottle, 1 L | Thermo Scientific | 3420201000 |
| Filtration Flask, 1 L | Corning | 431098 |

5.3 Reagents

| Item | Manufacturer | Catalog No. |
|---|---|---|
| USP Grade Phosphate solution | Ricca | R5808250 |
| USP Grade Sodium chloride | Spectrum | S0155 |
| USP Grade Glycerol (>99.5%) | JT-Baker | 2143-01 |
| USP Grade L-cysteine | Spectrum | C1473 |
| Ethanol (190 Proof) | Fisher Scientific | 04-355-225 |

#### 6.0 STORAGE

6.1 Ethanol is stored at room temperature separate from all other reagents.

6.2 All other reagents are stored at room temperature.

#### 7.0 PROCEDURE

##### Media Formulation

7.1 Make fresh media for each new production run.

7.2 Prepare media in a new disposable media bottle.

7.3 For cryogenic medium (1 L):

7.3.1 In the disposable media bottle add 200 mL sterile phosphate solution (H2KO4P pH 7.5). Add 9g sodium chloride, and 0.5 g L-cysteine hydrochloride and mix into solution.

7.3.2 Add 300 ml of glycerol

7.3.3 Bring total volume to 1000 mL with sterile phosphate solution (approximately 491 ml)

7.4 Filter sterilize solution, working under a BSC.

#### 8.0 ASSOCIATED DOCUMENTS

8.1 MTC-SOP-FS-0002_Generation of CFU normalized Fecal Slurry (FS) for Fecal Microbiota Transplantation (FMT)

#### 9.0 REVISION HISTORY

| DATE | DESCRIPTION OF CHANGE | INITIATOR |
|---|---|---|

| Icahn School of Medicine at Mount Sinai — Microbiome Translational Center | **Microbiome Translational Center Standard Operating Procedures** |
|---|---|
| **Title: Drug Substance (DS) Culture Media for Live Biotherapeutic Products (LBP)** | **SOP No: 0007**<br>**Revision: 0**<br>**Effective Date: 02/02/2022**<br>**Supersedes Date: NEW** |

| **Written by:** | | **Date:** |
|---|---|---|
| Lukas Bethlehem, PhD | | |
| **Reviewed By:** | | **Date:** |
| Jeremiah Faith, PhD<br>Faculty Director | | |
| **Approved By:** | | **Date:** |
| Ilaria Mogno, PhD<br>Quality Assurance Specialist | | |

#### 1.0 INTRODUCTION

The purpose of this SOP is to provide instructions for the preparation of culture media, wash media and cryogenic media for the growth and preparation of anaerobic bacteria included in the Drug Substance (DS) of a Live Biotherapeutic Product (LBP).

#### 2.0 RESPONSIBLE PERSONNEL

2.1 The Faculty Director is responsible for identifying new SOPs to be written, drafting new SOPs, suggesting revisions to existing SOPs, reviewing and approving all SOPs, and ensuring that all laboratory personnel is properly trained and following all SOPs.

2.2 The Quality Assurance (QA) Specialist is responsible for coordinating the preparation, approval, distribution, and maintenance of this SOP, for suggesting revisions to existing SOPs, and for ensuring that all SOPs are being followed by the required personnel.

2.3 The Approver can and should name representatives, who are qualified to take over the duties in case of an emergency or during absence of the Approver/Reviewer.

2.4 Technicians and other Lab personal are responsible for performing the procedures described in this SOP.

#### 3.0 DEFINITIONS

3.1 SOP = Standard Operating Procedure
3.2 QA = Quality Assurance
3.3 MTC = Microbiome Translational Center
3.4 LBP = Live Biotherapeutic Product
3.5 MCB = Master Cell Bank
3.6 RCB = Research Cell Bank
3.7 WCB = Working Cell Bank
3.8 DS = Drug Substance
3.9 DP = Drug Product
3.10 PBS = Phosphate Buffer Solution
3.11 WFI = Water for Injection
3.12 PBS = Phosphate Buffered Solution
3.13 BSC = Biological Safety Cabinet

#### 4.0 SAFETY

4.1 Wear protective gloves and lab coat while performing this procedure.
4.2 Perform all steps with the dedicated equipment for production of LBPs and follow instructions carefully to avoid contamination of the product or exposure to the operator.
4.3 Dispose all sharp waste into the appropriate waste containers.

#### 5.0 MATERIALS

##### 5.1 Equipment

| Item | Manufacturer | Catalog No. |
|---|---|---|
| Biological Safety Cabinet (BSC) | Labconco | 302619100 |
| Anaerobic Chamber | Coy Laboratories | 7100000 |
| Aerobic Incubator | Scientific Industries | SI-G1700 |
| Refrigerator | Thermo Scientific | FBG12RPGA |
| Micropipettes | Eppendorf | 3123000900 |
| Repeater Pipette | Eppendorf | 4987000134 |
| Magnetic Stirrer | Thermo Scientific | 14-513-61SIX |
| Balance | Mettler Toledo | XS6002S |
| Fine Balance | Mettler Toledo | XP205 |
| Balance (BSC) | Ohaus | 30253019 |
| pH Meter | Mettler Toledo | 30019028 |

##### 5.2 Supplies

| Item | Manufacturer | Catalog No. |
|---|---|---|
| Pipet Tips | USA Scientific | Not specified |
| Media Bottle | Thermo Scientific | 3420200250 |
| Filtration Unit | Corning | 431153 |
| Sampling Tubes | Thermo Scientific | Not specified |
| Matrix Tubes | Thermo Scientific | 3744 |
| Combitips Advanced | Eppendorf | 0030089677 / 0030089618 |
| Ziploc Bag | Office Depot | Not specified |
| Spatulas | LevGo | 17251 |
| Glass Beads | Zymo Research | S1001 |
| Chocolate Agar Plates | BD Biosciences | 221169 |
| Tryptic Soy Agar Plates (TSA) | Edge Biologicals | P1405 |
| Aluminum Foil | Office Depot | Not specified |

5.3 Reagents

| Item | Manufacturer | Catalog No. |
|---|---|---|
| WFI Water | Cytiva | SH30221 |
| USP Grade PBS | Ricca Chemical Co. | R580825020F |
| USP Grade Glycerol | JT Baker | 214301 |
| USP Grade Sodium Chloride | Spectrum | SO155 |
| USP Grade L-cysteine | Spectrum | C1473 |
| Ethanol (190 Proof) | Fisher Scientific | 04-355-225 |
| Glycerol (>99.5%) | Fisher Scientific | BP229 |
| L-cysteine | Fisher Scientific | C1276 |
| Vegitone Infusion Broth | Sigma Aldrich | 41960 |
| Yeast Extract | Thermo Scientific | DF0127179 |
| D-xylose | Sigma Aldrich | X1500 |
| D-fructose | Sigma Aldrich | F0127 |
| D-glucose | Sigma Aldrich | G8270 |
| D-galactose | Sigma Aldrich | G5388 |
| N-acetyl-D-glucosamine | Sigma Aldrich | A3286 |
| L-arabinose | Sigma Aldrich | W325501 |
| D-cellobiose | Sigma Aldrich | C7252 |
| D-maltose | Sigma Aldrich | M5885 |
| Sucrose | Sigma Aldrich | S0389 |
| L-malic Acid | Sigma Aldrich | M6413 |
| Sodium Sulfate | Sigma Aldrich | S5640 |
| Tween 80 | Sigma Aldrich | P1754 |
| Menadione | Sigma Aldrich | M5625 |
| MOPS | Sigma Aldrich | M1254 |
| Sodium Hydroxide | Thermo Scientific | S318 |

#### 6.0 STORAGE

6.1 Vitamin-K solution is stored at 4 °C (protected from light), for up to two weeks.

6.2 Sodium hydroxide (NaOH) is stored at room temperature, separately from other components in a safety cabinet.

6.3 N-acetyl-D-glucosamine is stored at -20 °C.

6.4 Ethanol is stored at room temperature separate from all other reagents.

6.5 All other reagents are stored at room temperature.

6.6 Chocolate agar and TSA agar plates are stored at 4°C.

#### 7.0 PROCEDURE

##### Media Formulation

7.1 Prepare fresh media for each new production run.

7.2 Utilize WFI water to formulate the medium in dedicated, sterile media bottles.

7.3 For culture media:

7.3.1 Formulate 2 L culture medium according to the recipe listed below.

7.4 Only use USP grade reagents for the formulation of wash media and cryogenic media.

7.4.1 Prepare wash media and cryogenic media under the BSC.

7.5 For cryogenic media:

7.5.1 Prepare 60 g glycerol, add 140 mL sterile Phosphate Buffered Solution (PBS), 0.1 g L-cysteine hydrochloride and 1.8 g sodium chloride to formulate 200 mL cryogenic media.

7.6 For wash media:

7.6.1 Prepare 500 mL sterile PBS, 0.26 g L-cysteine hydrochloride and 4.5 g sodium chloride to formulate 500 mL wash media.

7.7 Filter sterilize all media and buffer working under a BSC.

**Media Sterility Test**

7.8 Print four labels each for culture, wash and cryogenic QC vials and for the corresponding storage box and generate the respective entries for the databank (Freezerworks).

7.9 For the media sterility test, work under the BSC.

7.9.1 Transfer 250 µL of the culture, wash and cryogenic media each to a chocolate plate and a TSA plate, add 5-15 sterile glass beads and shake in horizontal plane 10-15 times.

7.9.2 Discard the glass beads and incubate the TSA plates in an aerobic incubator at 37 °C and the chocolate plates in an anaerobic chamber at 37 °C for 14 days.

7.9.3 If any colonies form, identify via biotyping and repeat sterility test with a respective QC vial. If contamination can be re-isolated, eliminate production run.

7.10 Transfer 500 µL of the culture, wash and cryogenic media each into the respective media QC vials and store at -80 °C.

**Media Recipes**

LYH_VIB Medium

| Component | Amount [g/L] |
|---|---|
| Vegitone infusion broth (VIB) | 37 |
| Yeast extract | 5 |
| Monosaccharide mix | 4 |
| - D-xylose | 1 |
| - D-fructose | 1 |
| - D-glucose | 1 |
| - D-galactose | 1 |
| - N-acetylglucosamine | 0.5 |
| - L-arabinose | 0.5 |
| Disaccharide mix | 3 |
| - D-Cellobiose | 1 |
| - D-Maltose | 1 |
| - Sucrose | 1 |
| L-cysteine hydrochloride | 0.5 |
| L-Malic acid | 1 |
| Sodium sulfate | 2 |
| MOPS | 20.9 |
| | **Volume [mL/L]** |
| Vitamin-K solution | 1 |
| Tween 80 | 0.5 |
| $H_2O$ (WIFI) | Adjust to 1 L |
| | pH 7.2 (NaOH) |

Vitamin-K solution

| Component | Volume [mg/mL] |
|---|---|
| Menadione | 1 |
| Ethanol (190 Proof) | Adjust to 1 mL |

#### 8.0 ASSOCIATED DOCUMENTS

8.1 MTC-SOP-0006_Generation of Drug Substance (DS) for Live Biotherapeutic Products (LBP)

#### 9.0 REVISION HISTORY

| DATE | DESCRIPTION OF CHANGE | INITIATOR |
|---|---|---|

| Icahn School of Medicine at Mount Sinai — Microbiome Translational Center | **Microbiome Translational Center**<br>**Standard Operating Procedures** |
|---|---|
| **Title: Drug Product (DP) Buffer for Live Biotherapeutic Products (LBP)** | **SOP No: 0012**<br>**Revision: 0**<br>**Effective Date: 08/11/2022**<br>**Supersedes Date: NEW** |

| **Written by:** | | **Date:** |
|---|---|---|
| Lukas Bethlehem, PhD | | |
| **Reviewed By:** | | **Date:** |
| Jeremiah Faith, PhD<br>Faculty Director | | |
| **Approved By:** | | **Date:** |
| Ilaria Mogno, PhD<br>Quality Assurance Specialist | | |

#### 1.0 INTRODUCTION

The purpose of this SOP is to provide instructions for the preparation of formulation buffer for the preparation of the Drug Product (DP) to be used as a Live Biotherapeutic Product (LBP).

#### 2.0 RESPONSIBLE PERSONNEL

2.1 The Faculty Director is responsible for identifying new SOPs to be written, drafting new SOPs, suggesting revisions to existing SOPs, reviewing and approving all SOPs, and ensuring that all laboratory personnel is properly trained and following all SOPs.

2.2 The Quality Assurance (QA) Specialist is responsible for coordinating the preparation, approval, distribution, and maintenance of this SOP, for suggesting revisions to existing SOPs, and for ensuring that all SOPs are being followed by the required personnel.

2.3 The Approver can and should name representatives, who are qualified to take over the duties in case of an emergency or during absence of the Approver/Reviewer.

2.4 Technicians and other Lab personal are responsible for performing the procedures described in this SOP.

#### 3.0 DEFINITIONS

3.1 SOP = Standard Operating Procedure
3.2 QA = Quality Assurance
3.3 MTC = Microbiome Translational Center
3.4 LBP = Live Biotherapeutic Product
3.5 MCB = Master Cell Bank
3.6 RCB = Research Cell Bank
3.7 WCB = Working Cell Bank
3.8 DS = Drug Substance
3.9 DP = Drug Product
3.10 PBS = Phosphate Buffer Solution
3.11 WFI = Water for Injection
3.12 PBS = Phosphate Buffered Solution
3.13 BSC = Biological Safety Cabinet

#### 4.0 SAFETY

4.1 Wear protective gloves and lab coat while performing this procedure.
4.2 Perform all steps with the dedicated equipment for production of LBPs and follow instructions carefully to avoid contamination of the product or exposure to the operator.
4.3 Dispose all sharp waste into the appropriate waste containers.

#### 5.0 MATERIALS

##### 5.1 Equipment

| Item | Manufacturer | Catalog No. |
|---|---|---|
| Biological Safety Cabinet (BSC) | Labconco | 302619100 |
| Pipette Controller | Corning | 4099 |
| Repeater Pipette | Eppendorf | 4987000134 |
| Balance (BSC) | Ohaus | 30253019 |

##### 5.2 Supplies

| Item | Manufacturer | Catalog No. |
|---|---|---|
| Serological Pipettes | Corning | Not specified |
| Media Bottle | Thermo Scientific | 3420200250 |
| Filtration Unit | Corning | 431153 |
| Matrix Tubes | Thermo Scientific | 3744 |
| Combitips Advanced | Eppendorf | 0030089677 / 0030089618 |
| Sterile Spatulas | LevGo | 17251 |

##### 5.3 Reagents

| Item | Manufacturer | Catalog No. |
|---|---|---|
| Ethanol (70%) | Fisher Scientific | 2546701 |
| USP Grade PBS | Ricca Chemical Co. | R580825020F |
| USP Grade Glycerol | JT Baker | 214301 |
| USP Grade Sodium Chloride | Spectrum | SO155 |
| USP Grade L-cysteine | Spectrum | C1473 |

#### 6.0 STORAGE

6.1 Ethanol is stored at room temperature separate from all other reagents.

6.2 All other reagents are stored at room temperature.

#### 7.0 PROCEDURE

##### Buffer Formulation

7.1 Prepare fresh DP formulation buffer for each new production run.

7.2 Utilize USP grade chemicals and sterile media bottles to prepare sufficient volume of DP formulation (depending on amount of DP doses produced and volumes of DS added to DP; see *MTC-SOP-0011_Generation of Drug Product (DP) for Live Biotherapeutic Products (LBP)*.

7.2.1 Prepare DP formulation buffer under the BSC, according to recipe below.

7.3 Filter sterilize the DP formulation buffer working under a BSC.

##### DP Buffer QC vials

7.4 Print three labels for each (sterile) DP formulation buffer bottle generated and for the corresponding storage box.

7.5 Transfer 500 µL of each bottle into the respective media QC vials, using sterile equipment, and store at -80 °C.

##### Buffer Recipe

DP Formulation Buffer

| Component | Amount per L | |
|---|---|---|
| L-cysteine (USP Grade) | 0.5 | g |
| Glycerol (USP Grade) | 300 | mL |
| NaCl (USP Grade) | 9 | g |
| Phosphate buffered solution | Adjust to 1 | L |

#### 8.0 ASSOCIATED DOCUMENTS

8.1 MTC-SOP-0011_Generation of Drug Product (DP) for Live Biotherapeutic Products (LBP)

#### 9.0 REVISION HISTORY

| DATE | DESCRIPTION OF CHANGE | INITIATOR |
|---|---|---|

### Sample Certificates of Analysis for all Formulation Buffer Components

### Certificate Of Analysis

| Item Number | C1473 | Lot Number | 1LA0153 |
|---|---|---|---|
| Item | Cysteine Hydrochloride, Monohydrate, USP | CAS Number | 7048-04-6 |
| Molecular Formula | $C_3H_7NO_2S•HCl•H_2O$ | Molecular Weight | 175.64 |

| TEST | SPECIFICATION | | RESULT |
|---|---|---|---|
| | MIN | MAX | |
| ASSAY (DRIED BASIS) | 98.5 | 101.5 % | 99.84 % |
| SPECIFIC ROTATION $[a]_D$ | +5.7° to +6.8° | | +5.98° |
| LOSS ON DRYING | 8.0 | 12.0 % | 10.26 % |
| RELATED COMPOUNDS: | | | . |
| INDIVIDUAL IMPURITY | | 0.5 % | <0.5 % |
| TOTAL IMPURITIES | | 2.0 % | <2.0 % |
| RESIDUE ON IGNITION | | 0.4 % | 0.01 % |
| SULFATE | | 0.03 % | <0.03 % |
| IRON | | 30 ppm | <30 ppm |
| ELEMENTAL IMPURITIES | AS REPORTED | | NO ELEMENTAL IMPURITIES PRESENT |
| IDENTIFICATION (FTIR) | SPECTRUM MATCHES REFERENCE | | SPECTRUM MATCHES REFERENCE |
| CERTIFIED HALAL | | | CERTIFIED HALAL |
| APPEARANCE | | | WHITE CRYSTALS |
| EXPIRATION DATE | | | 29-MAR-2023 |
| DATE OF MANUFACTURE | | | 30-MAR-2021 |
| RESIDUAL SOLVENTS | AS REPORTED | | NO RESIDUAL SOLVENTS USED |
| MONOGRAPH EDITION | | | (USP) 2021 |

Certificate of Analysis Results Entered By:

HBERMUDEZ
Hernan Bermudez
17-JAN-22 02:18:37

Certificate of Analysis Results Approved By:

HBERMUDEZ
Hernan Bermudez
17-JAN-22 02:19:00

Spectrum Chemical Mfg Corp
14422 South San Pedro Street
Gardena 90248 CA

**All pharmaceutical ingredients are tested using current edition of applicable pharmacopeia.**

**Read and understand label and SDS before handling any chemicals. All Spectrum's chemicals are for manufacturing, processing, repacking or research purposes by experienced personnel only. It is the customer's responsibility to provide adequate hazardous material training and ensure that appropriate Personal Protective Equipment (PPE) is used before handling any chemical.**

The Elemental Impurities standards implemented by USP and other Pharmaceutical Compendia reflect a growing understanding of the toxicology of trace levels of elemental impurities that can remain in drug substances originating from either raw materials or manufacturing processes. Identifying and quantifying impurities can be critical to predicting the best possible patient outcomes. Elemental Impurities has been a requirement of all products meeting USP/NF, EP and BP monographs since January 1, 2018. More information can be found in USP sections <232> Elemental Impurities – Limits and <233> Elemental Impurities – Procedures. Data for drug substances furnished by Spectrum Chemical Mfg. Corp can be used to ensure that patient daily exposures by oral administration to the selected elements are not exceeded in the formulation of pharmaceutical products.

Glycerin, U.S.P.
Multi-Compendial
100% Vegetable Based
(glycerol)

Material No.: 2143-01
Batch No.: 0000289984
Manufactured Date: 2021-02-07
Retest Date: 2026-02-06
Release Date: 2021-09-10
Revision No.: 0

### Certificate of Analysis

Meets B.P. Chemical Specifications, Meets J.P. Chemical Specifications, Meets E.P. Chemical Specifications, Meets U.S.P Requirements, GMP Manufactured Product

| Test | Specification | Result |
|---|---|---|
| USP - Identification A | Passes Test | Passes Test |
| USP - Identification B - Diethylene Glycol | ≤ 0.10 % | < 0.10 % |
| USP - Identification B - Ethylene Glycol | ≤ 0.10 % | < 0.10 % |
| USP - Identification C | Passes Test | Passes Test |
| USP - Assay (as $HOCH_2CHOHCH_2OH$) (calculated on anhydrous basis) | 99.0 - 101.0 % | 101.0 % |
| USP - Specific Gravity at 25°/25°C | ≥ 1.249 | 1.261 |
| USP - Color | Passes Test | Passes Test |
| USP - Residue on Ignition | ≤ 0.01 % | < 0.01 % |
| USP - Chloride (Cl) | ≤ 0.001 % | < 0.001 % |
| USP - Sulfate ($SO_4$) | ≤ 0.002 % | < 0.002 % |
| USP - Chlorinated Compounds (as Cl) | ≤ 0.003 % | < 0.003 % |
| USP - Fatty Acids and Esters | Passes Test | Passes Test |
| USP - Water ($H_2O$) | ≤ 5.0 % | 0.6 % |
| USP - Related Compounds: Individual Impurity | ≤ 0.1 % | < 0.1 % |
| USP - Related Compo unds: Total Impurities | ≤ 1.0 % | < 1.0 % |
| EP/BP - Identification A | Passes Test | Passes Test |
| EP/BP - Identification B | Passes Test | Passes Test |
| EP/BP - Appearance of Solution | Passes Test | Passes Test |
| EP/BP - Acidity or Alkalinity | Passes Test | Passes Test |
| EP/BP - Refractive Index, $n^{20}D$ | 1.470 - 1.475 | 1.474 |
| EP/BP - Aldehydes | ≤ 10 ppm | < 10 ppm |
| EP/BP - Ester | Passes Test | Passes Test |
| EP/BP - Halogenated Compounds | ≤ 35 ppm | < 35 ppm |
| EP/BP - Sugars | Passes Test | Passes Test |
| EP/BP - Chloride (Cl) | ≤ 10 ppm | < 10 ppm |
| EP/BP - Water ($H_2O$) | ≤ 2.0 % | 0.6 % |
| EP/BP - Ash (sulfated) | ≤ 0.01 % | < 0.01 % |
| EP/BP - Impurity A and Related Substances - Impurity A | ≤ 0.1 % | < 0.1 % |

>>> Continued on page 2 >>>

For questions on this Certificate of Analysis please contact Technical Services at 855.282.6867 or +1.610.386.1700
Avantor Performance Materials, LLC

Glycerin, U.S.P.
Multi-Compendial
100% Vegetable Based
(glycerol)

Material No.: 2143-01
Batch No.: 0000289984

| Test | Specification | Result |
|---|---|---|
| EP/BP - Impurity A and Related Substances - Other Impurities with R^T<Glycerol | ≤ 0.1 % | < 0.1 % |
| EP/BP - Impurity A and Related Substances - Total Impurities R^T>Glycerol | ≤ 0.5 % | < 0.5 % |
| EP - Assay (anhydrous basis) | 98.0 - 101.0 % | 100.3 % |
| JP - Assay (anhydrous basis) | 98.0 - 101.0 % | 100.9 % |
| JP - Identification | Passes Test | Passes Test |
| JP - Specific Gravity at 20°/20°C | ≥ 1.258 | 1.264 |
| JP - Refractive Index, $n^{20}D$ | ≥ 1.470 | 1.474 |
| JP - Color | Passes Test | Passes Test |
| JP - Residue on Ignition | ≤ 0.01 % | < 0.01 % |
| JP - Chloride (Cl) | ≤ 0.001 % | < 0.001 % |
| JP - Sulfate ($SO_4$) | ≤ 0.002 % | < 0.002 % |
| JP - Heavy Metals (as Pb) | ≤ 5 ppm | < 5 ppm |
| JP - Fatty Acids and Esters(as butyric acid) | Passes Test | Passes Test |
| JP - Readily Carbonizable Substances | Passes Test | Passes Test |
| JP - Acidity or Alkalinity | Passes Test | Passes Test |
| JP - Acrolein, Glucose, and Other Reducing Substances | Passes Test | Passes Test |
| JP - Ammonium ($NH_4$) | Passes Test | Passes Test |
| JP - Calcium (Ca) | Passes Test | Passes Test |
| JP - Arsenic (As) | ≤ 2 ppm | < 2 ppm |
| JP - Water ($H_2O$) | ≤ 2.0 % | 0.6 % |
| JP - Ethylene Glycol | ≤ 0.1 % | < 0.1 % |
| JP - Diethylene Glycol | ≤ 0.1 % | < 0.1 % |
| JP - Any Other Impurity | ≤ 0.1 % | < 0.1 % |
| JP - Total Impurities | ≤ 1.0 % | < 0.1 % |
| Endotoxin Concentration (EU/g) | ≤ 5 | < 1 |

>>> Continued on page 3 >>>

Glycerin, U.S.P.
Multi-Compendial
100% Vegetable Based
(glycerol)

Material No.: 2143-01
Batch No.: 0000289984

| Test | Specification | Result |
| --- | --- | --- |

GMP Manufactured Product
Bulk Pharmaceutical Chemical
CAUTION: For Manufacturing, processing or repackaging
No Class 1,2,3 or other solvents are used or produced in the manufacturing or purification of the product.
Elemental Impurities (USP 232, EP 5.20) - Information on elemental impurities for this product is available on the associated Product Regulatory Data Sheet and elemental impurity profile report.
Country of Origin: USA
Packaging Site: Paris Mfg Ctr & DC
Manufacturer: 00005030
Manufacturer source batch: 0002375284

James T Ethier
Jamie Ethier
Vice President Global Quality

spectrum CHEMICAL MFG CORP Trusted Since 1971

### Certificate Of Analysis

| | | | |
|---|---|---|---|
| Item Number | SO155 | Lot Number | 1LH0541 |
| Item | Sodium Chloride, Granular, USP | Manufacturer Lot | RI22189003 |
| CAS Number | 7647-14-5 | Manufacturer Code | 12349 |
| Molecular Formula | NaCl | Molecular Weight | 58.44 |

| TEST | SPECIFICATION | | RESULT |
|---|---|---|---|
| | MIN | MAX | |
| APPEARANCE | | | WHITE GRANULES |
| IDENTIFICATION (A) | POSITIVE FOR SODIUM | | POSITIVE FOR SODIUM |
| IDENTIFICATION (B) | PRECIPITATE DISSOLVES | | PRECIPITATE DISSOLVES |
| ASSAY (DRIED BASIS) | 99.0 | 100.5 % | 100.5 % |
| APPEARANCE OF SOLUTION | CLEAR COLORLESS | | CLEAR COLORLESS |
| ACIDITY OR ALKALINITY | | 0.5 ml | 0.05 % |
| LOSS ON DRYING | | 0.5% | 0.0 % |
| BROMIDES | | 100 ppm | <100 ppm |
| PHOSPHATES | | 25 ppm | <25 ppm |
| IODIDES | NO BLUE COLOR | | NO BLUE COLOR |
| MAGNESIUM AND ALKALINE-EARTH METALS (as Ca) | | 100 ppm | <100 ppm |
| ARSENIC (As) | | 1 ppm | <0.05 ppm |
| IRON (Fe) | | 2 ppm | <2 ppm |
| BARIUM (Ba) | OPALESCENCE LESS THAN REFERENCE | | OPALESCENCE LESS THAN REFERENCE |
| FERROCYANIDES | NO BLUE COLOR | | NO BLUE COLOR |
| SULFATE ($SO_4$) | | 200 ppm | <200 ppm |
| NITRITES | | 0.01 | 0.00 |
| BACTERIAL ENDOTOXINS | | 5 IU/g | <5 IU/g |
| ELEMENTAL IMPURITIES | AS REPORTED | | NO ELEMENTAL IMPURITIES PRESENT (M) |

| TEST | SPECIFICATION | | RESULT |
|---|---|---|---|
| | MIN | MAX | |
| CERTIFIED KOSHER | | | CERTIFIED KOSHER |
| CERTIFIED HALAL | | | CERTIFIED HALAL |
| RETEST DATE | | | 08-JUL-2025 |
| DATE OF MANUFACTURE | | | 08-JUL-2022 |
| RESIDUAL SOLVENTS | -AS REPORTED | | NO RESIDUAL SOLVENTS USED (M) |
| MONOGRAPH EDITION | | | (USP) 2022 |
| NOTE:All results reported with (M) are from manufacturer certificate of analysis | | | . |

Certificate of Analysis Results Entered By:

HBERMUDEZ
Hernan Bermudez
15-SEP-22 09:10:16

Certificate of Analysis Results Approved By:

JRUIZ
Jesus Ruiz
15-SEP-22 04:52:10

Spectrum Chemical Mfg Corp
14422 South San Pedro Street
Gardena 90248 CA

**All pharmaceutical ingredients are tested using current edition of applicable pharmacopeia.**

**Read and understand label and SDS before handling any chemicals. All Spectrum's chemicals are for manufacturing, processing, repacking or research purposes by experienced personnel only. It is the customer's responsibility to provide adequate hazardous material training and ensure that appropriate Personal Protective Equipment (PPE) is used before handling any chemical.**

The Elemental Impurities standards implemented by USP and other Pharmaceutical Compendia reflect a growing understanding of the toxicology of trace levels of elemental impurities that can remain in drug substances originating from either raw materials or manufacturing processes. Identifying and quantifying impurities can be critical to predicting the best possible patient outcomes. Elemental Impurities has been a requirement of all products meeting USP/NF, EP and BP monographs since January 1, 2018. More information can be found in USP sections <232> Elemental Impurities – Limits and <233> Elemental Impurities – Procedures. Data for drug substances furnished by Spectrum Chemical Mfg. Corp can be used to ensure that patient daily exposures by oral administration to the selected elements are not exceeded in the formulation of pharmaceutical products.

Spectrum Chemical Mfg. Corp. repackages Active Pharmaceutical Ingredient chemicals provided by other manufacturers and suppliers. For traceability purposes, Spectrum's Certificate of Analysis contains an identifier code unique to each manufacturer or supplier, as well as the chemical's lot number provided on the original manufacturer Certificate of Analysis. This material is manufactured by Morton Salt, Inc. at 151 South Industrial Avenue, Rittman, Ohio (OH) 44270, United States. Copies of the original Certificate of Analysis are available upon request from your Spectrum Sales Representative or by calling Spectrum's Quality Department at 1-800-772-8786.

RICCA CHEMICAL COMPANY®

1841 Broad Street
Pocomoke City, MD 21851
http://www.riccachemical.com
1-888-GO-RICCA


### Certificate of Analysis

#### Phosphate Buffer Solution, pH 7.5, USP 27 Reagent

**Lot Number:** 2207968 **Product Number:** R5808250

**Manufacture Date:** JUL 08, 2022
**Expiration Date:** JUN 2024

| Name | CAS# | Grade |
|---|---|---|
| Water | 7732-18-5 | ACS/ASTM/USP/EP |
| Potassium Dihydrogen Phosphate | 7778-77-0 | ACS |
| Sodium Hydroxide | 1310-73-2 | Reagent |

| Test | Specification | Result | NIST SRM# |
|---|---|---|---|
| Appearance | Colorless liquid | Passed | |
| pH at 25°C (Method: SQCP027, SQCP033) | 7.48-7.52 | 7.51 | 186-I-g, 186-II-g, 191d |

Volumetric glassware complies with Class A tolerance requirements of ASTM E 288 and NIST Circular 434; it is calibrated before first use and recalibrated regularly in accordance with ASTM E 542 and NIST Procedure NBSIR 74-461. Balances are calibrated regularly with weights certified traceable to the NIST national mass standard. Thermometers and temperature probes are calibrated before first use and recalibrated regularly with a thermometer traceable to NIST standards. All products are prepared according to master documents that assure manufacture according to validated methods. Batch records document raw material traceability and production and testing history for each lot manufactured.

| Part Number | Size / Package Type | Shelf Life (Unopened Container) |
|---|---|---|
| R5808250-20F | 20 L Cubitainer® | 24 months |

**Recommended Storage:** 15°C - 30°C (59°F - 86°F)

Myrlande Gilles (07/08/2022)
Quality Control

This Certificate of Analysis is designed to comply with ISO Guide 31 "Reference Materials -- Contents of Certificates and Labels."

This test report shall not be reproduced, except in full, without the written approval of Ricca Chemical Company.

### Appendix 10 – calculation of MTC01 and FS potency by colony forming units.

### SOP for MTC01 CFU

| **Written by:** | | **Date:** |
|---|---|---|
| Lukas Bethlehem, PhD | | |
| **Reviewed By:** | | **Date:** |
| Jeremiah Faith, PhD<br>Faculty Director | | |
| **Approved By:** | | **Date:** |
| Ilaria Mogno, PhD<br>Quality Assurance Specialist | | |

#### 1.0 INTRODUCTION

The purpose of this SOP is to provide instructions for the enumeration of colony forming units (CFU) as a measure of potency of the Drug Product (DS) to be used as a Live Biotherapeutic Product (LBP).

#### 2.0 RESPONSIBLE PERSONNEL

2.1 The Faculty Director is responsible for identifying new SOPs to be written, drafting new SOPs, suggesting revisions to existing SOPs, reviewing and approving all SOPs, and ensuring that all laboratory personnel is properly trained and following all SOPs.

2.2 The Quality Assurance (QA) Specialist is responsible for coordinating the preparation, approval, distribution, and maintenance of this SOP, for suggesting revisions to existing SOPs, and for ensuring that all SOPs are being followed by the required personnel.

2.3 The Approver can and should name representatives, who are qualified to take over the duties in case of an emergency or during absence of the Approver/Reviewer.

2.4 Technicians and other Lab personal are responsible for performing the procedures described in this SOP.

#### 3.0 DEFINITIONS

3.1 SOP = Standard Operating Procedure
3.2 QA = Quality Assurance
3.3 MTC = Microbiome Translational Center
3.4 LBP = Live Biotherapeutic Product
3.5 MCB = Master Cell Bank
3.6 RCB = Research Cell Bank
3.7 WCB = Working Cell Bank
3.8 DS = Drug Substance
3.9 DP = Drug Product
3.10 CFU = Colony Forming Unit
3.11 PBS = Phosphate Buffered Saline
3.12 TNTC = Too Numerous to Count

#### 4.0 SAFETY

4.1 Wear protective gloves and lab coat while performing this procedure.
4.2 Perform all steps with the dedicated equipment for production of LBPs and follow instructions carefully to avoid contamination of the product or exposure to the operator.
4.3 Dispose all biological waste into the appropriate waste containers.
4.4 Dispose all sharp waste into the appropriate waste containers.

#### 5.0 MATERIALS

##### 5.1 Equipment

| Item | Manufacturer | Catalog No. |
|---|---|---|
| Anaerobic Chamber | Coy Laboratories | 7100000 |
| Micropipettes | Eppendorf | 3123000900 |
| Multichannel Pipette | Thermo Scientific | 4672020BT |
| Vortex Mixer | Fisher Scientific | 14-955-151 |

##### 5.2 Supplies

| Item | Manufacturer | Catalog No. |
|---|---|---|
| Pipet Tips | USA Scientific | Not specified |
| Multichannel Tips | Thermo Scientific | Not specified |
| Syringes | Fisher Scientific | Not specified |
| Syringe Needle | BD Biosciences | 305155 |
| Ziploc Bag | Office Depot | Not specified |
| Chocolate Agar Plates | BD Biosciences | 221169 |

##### 5.3 Reagents

| Item | Manufacturer | Catalog No. |
|---|---|---|
| Phosphate Buffered Saline (PBS) | Caisson Labs | PBL06 |
| Ethanol (70%) | Fisher Scientific | Not specified |

#### 6.0 STORAGE

6.1 Ethanol for cleaning is stored at room temperature separate from all the other reagents.

6.2 Chocolate agar plates are stored at 4°C.

#### 7.0 PROCEDURE

##### CFU Plating – DP

7.1 Label three chocolate agar plates with:
*Reference Batch Number*, *DP-Dose*, *Plate Number*

7.2 Use dilutions prepared in *MTC-SOP-0011_Generation of Drug Product (DP) for Live Biotherapeutic Products (LBP)*.

7.2.1 Utilize the multichannel pipette to mix and transfer ten times 1 µL (dilutions $10^{-3}$-$10^{-6}$ for High doses (>1E+11 total CFU) and $10^{-2}$-$10^{-5}$ for Low doses (>1E+10 total CFU)) from wells C-F or D-E to the respective chocolate agar plate.

7.2.2 Leave the plates, facing upright, for 5-10 minutes to prevent spots from running.

7.3 Place the plates in a Ziploc bag, to prevent dehydration, and incubate at 37 °C in the anaerobic chamber until colonies are visible (2-4 days).

7.3.1 The different strains in a DP have different optimal incubation times and the plates should be incubated as long as possible without colonies growing into each other.

7.3.2 The countable range of colonies is between 10-100 CFU per dilution. Everything above is considered too numerous to count (TNTC).

7.3.3 The CFU/mL is determined for the lowest dilution in the countable range.

7.3.4 Counting of the colonies can be performed outside of the chamber.

7.3.5 CFU count is documented to assess potency of the DP. If CFU count of DP is >2 logs below target CFU, repeat CFU count with two separate DP-QC vials. Destroy DP if all three QC vials fail specificity.

#### 8.0 ASSOCIATED DOCUMENTS

8.1 MTC-SOP-0011_Generation of Drug Product (DP) for Live Biotherapeutic Products (LBP)

#### 9.0 REFERENCES

#### 10.0 REVISION HISTORY

| DATE | DESCRIPTION OF CHANGE | INITIATOR |
|---|---|---|

### SOP for FS CFU

| Microbiome Translational Center (Icahn School of Medicine at Mount Sinai) | **Microbiome Translational Center Standard Operating Procedures** |
|---|---|
| **Title: CFU Count of the Fecal Slurry (FS) for Fecal Microbiota Transplantation (FMT)** | **SOP No: MTC-SOP-FS-0005**<br>**Revision: 0** |
| | **Effective Date: 06/30/2022**<br>**Supersedes Date: NEW** |

| **Written by:** | | **Date:** |
|---|---|---|
| Lukas Bethlehem, PhD | | |
| **Reviewed By:** | | **Date:** |
| Jeremiah Faith, PhD<br>Faculty Director | | |
| **Approved By:** | | **Date:** |
| Ilaria Mogno, PhD<br>Quality Assurance Specialist | | |

#### 1.0 INTRODUCTION

The purpose of this SOP is to provide instructions for the enumeration of colony forming units (CFU) as a measure of potency of human fecal slurry to be used for Fecal Microbiota Transplantation (FMT).

#### 2.0 RESPONSIBLE PERSONNEL

2.1 The Facility Director is responsible for identifying new SOPs to be written, drafting new SOPs, suggesting revisions to existing SOPs, reviewing and approving all SOPs, and ensuring that all laboratory personnel is properly trained and following all SOPs.

2.2 The Quality Assurance (QA) Lead is responsible for coordinating the preparation, approval, distribution, and maintenance of this SOP, for suggesting revisions to existing SOPs, and for ensuring that all SOPs are being followed by the required personnel.

2.3 The Approver can and should name representatives, who are qualified to take over the duties in case of an emergency or during absence of the Approver/reviewer.

2.4 Technicians and other Lab personal are responsible for performing the procedures described in this SOP.

#### 3.0 DEFINITIONS

3.1 SOP = Standard Operating Procedure

3.2 QA = Quality Assurance

3.3 MTC = Microbiome Translational Center
3.4 FS = Fecal Slurry
3.5 cFS = concentrated Fecal Slurry
3.6 FMT = Fecal Microbiota Transplantation
3.7 CFU = Colony Forming Unit
3.8 PBS = Phosphate Buffered Saline
3.9 TNTC = Too Numerous to Count

#### 4.0 SAFETY

4.1 Wear protective gloves and lab coat while performing this procedure.
4.2 Perform all steps with the dedicated equipment for production of FS and follow instructions carefully to avoid contamination of the product or exposure to the operator.
4.3 Dispose all biological waste into the appropriate waste containers.

#### 5.0 MATERIALS

5.1 Equipment

| Item | Manufacturer | Catalog No. |
|---|---|---|
| Anaerobic Chamber | Coy Laboratories | AC19-157 |
| Micropipettes | Eppendorf | 3123000900 |
| Multichannel Pipette | Thermo Scientific | 4672020BT |
| Vortex Mixer | Fisher Scientific | 14-955-151 |

5.2 Supplies

| Item | Manufacturer | Catalog No. |
|---|---|---|
| Pipet Tips | USA Scientific | Not specified |
| Multichannel Tips | Thermo Scientific | Not specified |
| Syringes | Fisher Scientific | Not specified |
| Syringe Needle | BD Biosciences | 305155 |
| Ziploc Bag | Office Depot | Not specified |
| 96-well plate | | |

5.3 Reagents

| Item | Manufacturer | Catalog No. |
|---|---|---|

| | | |
|---|---|---|
| Phosphate Buffered Saline (PBS) | Caisson Labs | PBL06 |
| Chocolate Agar Plates | BD Biosciences | 221169 |
| Ethanol (70%) | Fisher Scientific | Not specified |

#### 6.0 STORAGE

6.1 Ethanol for cleaning is stored at room temperature separate from all the other reagents.

6.2 Chocolate agar plates are stored at 4°C.

#### 7.0 PROCEDURE

##### CFU Plating – Cryogenic Vials

7.1 Label two chocolate agar plates with:
*Reference Batch Number*, *Plate Number*

7.2 For a dilution series, fill 90 µL sterile PBS into unused positions B to G of a sterile 96 well plate.

7.2.1 You can use the same plate for several samples of the same donor production run, but not for different production runs.

7.2.2 Leave one column free between samples on the 96 well plate, if using the same plate for several samples of the same production run to prevent contamination.

7.3 To position A1 of the sterile 96 well plate, add a concentrated fecal slurry (cFS) aliquot, as generated in *MTC-SOP-FS-0001_Generation of CFU normalized Fecal Slurry (FS) for Fecal Microbiota Transplantation (FMT).* Additional samples from the same donor batch could be placed in A3, A5, …, A11

7.4 Mix the cFS in position A, using a 10 µL pipette and transfer 10 µL to the next position (B). Dispose the pipette tip and repeat the process with a fresh pipette tip starting at position B, until reaching position G.

7.4.1 Dilution series should generate dilutions from $10^{-1}$-$10^{-6}$ in positions B to G of the 96 well plate.

7.5 Utilize the multichannel pipette to transfer ten times 1 µL (dilutions $10^{-3}$-$10^{-6}$) from wells B to E to the respective chocolate agar plate.

7.5.1 Leave the plates, facing upright, for 5-10 minutes to prevent spots from running.

7.6 Place the plates in a Ziploc bag, to prevent dehydration, and incubate at 37 °C in the anaerobic chamber until colonies are visible.

7.6.1 Count colonies as soon as they become clearly visible to prevent overgrowth and inaccurate counting.

7.6.2 The countable range of colonies is between 10-100 CFU per dilution. Everything above is considered too numerous to count (TNTC).

7.6.3 The CFU/mL is determined for the lowest dilution in the countable range.

7.6.4 Counting of the colonies can be performed outside of the chamber.

#### 8.0 ASSOCIATED DOCUMENTS

8.1 MTC-SOP-FS-0002_Generation of CFU normalized Fecal Slurry (FS) for Fecal Microbiota Transplantation (FMT)

#### 9.0 REFERENCES

#### 10.0 REVISION HISTORY

| DATE | DESCRIPTION OF CHANGE | INITIATOR |
|---|---|---|

### Appendix 11 – Generation of fecal slurry from the stool samples of human donors that pass safety screening

### SOP for generation of cFS and FS

| Microbiome Translational Center (Icahn School of Medicine at Mount Sinai) | **Microbiome Translational Center Standard Operating Procedures** |
|---|---|
| **Title: Generation of CFU normalized Fecal Slurry (FS) for Fecal Microbiota Transplantation (FMT)** | **SOP No: MTC-SOP-FS-0002**<br>**Revision: 0**<br>**Effective Date: 10/16/2022**<br>**Supersedes Date: NEW** |

| **Written by:** | | **Date:** |
|---|---|---|
| Lukas Bethlehem, PhD | | |
| **Reviewed By:** | | **Date:** |
| Jeremiah Faith, PhD<br>Faculty Director | | |
| **Approved By:** | | **Date:** |
| Ilaria Mogno, PhD<br>Quality Assurance Specialist | | |

#### 1.0 INTRODUCTION

The purpose of this SOP is to provide instructions for generating Fecal Slurry (FS) that has been normalized to a dose of $1x10^{11}$ or $1x10^{10}$ CFU/dose. Each dose will be from the stool of healthy fecal donors free that pass extensive screening. The resulting FS will be used in Fecal Microbiota Transplantation of recipients with recurrent *C. difficile* infection.

To generate fecal material for fecal microbiota transplantation, a series of fecal samples are longitudinally collected form a given donor. Prior to processing the series samples must clear the safety screens for known infectious agents. Including the following:

**Blood tests 14 days before collection and 30 days after collection**

- CBC w/ differential
- LFTs, complete
- Bun/Cr
- Fasting glucose
- High Sensitivity CRP
- Serum triglycerides, LDL, HDL cholesterol
- HIV 1/2 (IgG) and E1A
- HAV (IgG and IgM)
- HBV (surface antigen/antibody; core antibody)
- HCV (IgG)
- Syphilis serology (Trep-sure ELISA; RPR done if treponemal test is positive)
- HTLV 1&2

**Stool tests on days 1 and 30 of collection**

- Clostridium difficile toxin by PCR
- Fecal Giardia antigen
- Fecal Cryptosporidium antigen
- Ova and parasites by light microscopy
- Helicobacter pylori fecal antigen
- Mount Sinai Hospital NAAT/Biofire: campylobacter, plesiomonas, salmonella, yersinia, vibrio spp, vibrio cholerae, enteroaggregative e coli, enteropathogenic e coli, enterotoxigenic e coli, shiga-like toxin producing e coli, e coli 0157, shigella/enteroinvasive e coli, cryptosporidium spp, Cyclospora cayetanensis, entamoeba histolytic, giardia lamblia, adenovirus F 40/41, astrovirus, norovirus gi/gii, rotavirus A, sapovirus (I, II, IV, V)
- VRE via agar plates
- MSRA via agar plates
- ESBL via agar plates

**COVID screens**
Donors will be tested for SARS-CoV-2 via nasal swab PCR at the following time points: 14 days before collection, day 1 of collection, day 15 of collection, day 30 of collection, and then 30 days after final day of collection. Each stool donation will be cleared by confirmation that the donor remained asymptomatic for 14 days post donation.

*Stools will not be released for processing unless these tests are complete and have passed.*

In the first processing step, a concentrated form of the fecal slurry (cFS) is generated by homogenizing the original stool in a PBS/glycerol/cysteine-based media using a stomacher with filter bag. The resulting cFS is frozen at -80C. A small aliquot of cFS is freeze-thawed again, and subsequently the potency of cFS is determined by CFU count. A normalized dose of $1x10^{11}$ [high dose] or $1x10^{10}$ [low dose] in 250 ml is generated from the cFS using the potency CFU count data. One small aliquot of FS is taken from all slurries for potential future use. An additional 9 aliquots of FS are taken to quantify the stability over time of the donor series. All samples of cFS and cFS are stored at -80C.

#### 2.0 RESPONSIBLE PERSONNEL

2.1 The Facility Director is responsible for identifying new SOPs to be written, drafting new SOPs, suggesting revisions to existing SOPs, reviewing and approving all SOPs, and ensuring that all laboratory personnel is properly trained and following all SOPs.

2.2 The Quality Assurance (QA) Lead is responsible for coordinating the preparation, approval, distribution, and maintenance of this SOP, for suggesting revisions to existing SOPs, and for ensuring that all SOPs are being followed by the required personnel.

2.3 The Approver can and should name representatives, who are qualified to take over the duties in case of an emergency or during absence of the Approver/reviewer.

2.4 Technicians and other Lab personal are responsible for performing the procedures described in this SOP.

#### 3.0 DEFINITIONS

3.1 SOP = Standard Operating Procedure
3.2 QA = Quality Assurance
3.3 MTC = Microbiome Translational Center
3.4 FMT = Fecal Microbiota Transplantation
3.5 EtOH = 70% ethanol
3.6 FS = fecal slurry
3.7 cFS = concentrated fecal slurry
3.8 BSC = biological safety cabinet

#### 4.0 SAFETY

4.1 Wear protective gloves and lab coat while performing this procedure.

4.2 Perform all steps with the dedicated equipment for production of FS and follow instructions carefully to avoid contamination of the product or exposure to the operator.

#### 5.0 MATERIALS

5.1 Equipment

| Item | Manufacturer | Catalog No. |
|---|---|---|
| Biological Safety Cabinet (BSC) | Labconco | 302611000 |
| Micropipettes | Eppendorf | 3123000900 |
| Pipetting Controller | | |
| Repeater pipette | | |
| Balance (BSC) | Ohaus | 30253019 |
| Stomacher 400 EVO Paddle Blender | Stomacher | 0400/002/00 |

5.2 Supplies

| Item | Manufacturer | Catalog No. |
|---|---|---|
| 400 ml stomacher filter bag | Seward | 030020278 |
| Cryogenic Storage Container 500 ml | Thermo Scientific | 342020-0500 |
| Cryogenic Storage Container 250 ml | Thermo Scientific | 342020-0250 |
| Serological pipettes (50 ml) | | |
| Combitips Advanced, 10 mL | Eppendorf | 0030089677 |

5.3 Reagents

| Item | Manufacturer | Catalog No. |
|---|---|---|
| Ethanol (70%) | | |

#### 6.0 STORAGE

6.1 All reagents are stored at room temperature.

#### 7.0 PROCEDURE

**Generating concentrated Fecal Slurry (cFS)**

7.1 Prepare cryogenic medium according to *MTC-SOP-FS-0003_Fecal Slurry Cryopreservation Media for Fecal Microbiota Transplantation*

7.2 Clean the biosafety cabinet according to *MTC-SOP-FS-0004_Sterilization of Biosafety Cabinet and Stomacher for Fecal Slurry Production for Fecal Microbiota Transplantation (FMT)*

7.3 Weigh the stool sample to be processed.

7.4 Weigh an empty 500 ml cryogenic storage container (with lid on)

7.5 In a stomacher bag, homogenize (60 seconds at 210 rotations per minute) the stool samples with cryogenic media with a 1 to 1 media to stool ratio. To avoid overfilling the stomacher bag, use less volume when media (ml) + stool (g) is >400. In this case use sufficient cryogenic media to bring the final media + stool to 400. For example, for 300 g of stool use 100 ml of media and for 260g stool add 140 ml media.

7.6 From the filtered portion of the stomacher bag, transfer the homogenized concentrated fecal slurry (cFS) to the 500 ml cryogenic storage container using a 50 ml serologic pipette

7.7 Collect one 100 µl aliquot of the cFS to a cryogenic vial.

7.8 Weigh the cFS in the 500 ml cryogenic storage container with the lid on to calculate the total weight of the homogenized sample.

7.9 Freeze at -80C the 500 ml storage container and the vial containing the cFS

7.10 After ≥24 hours at -80C, thaw the cryovial at room temperature for 10 minutes, then return to -80C for at least 24 additional hours to generate a freeze-thaw cycle prior to estimating cFS potency.

7.11 Surface clean all equipment with EtOH.

**Generating CFU normalized Fecal Slurry (cFS)**

7.12 Prepare cryogenic medium according to *MTC-SOP-FS-0003_Fecal Slurry Cryopreservation Media for Fecal Microbiota Transplantation*

7.13 Clean the biosafety cabinet according to *MTC-SOP-FS-0004_Sterilization of Biosafety Cabinet and Stomacher for Fecal Slurry Production for Fecal Microbiota Transplantation (FMT)*

7.14 Calculate the CFU/ml of cFS with *CFU Count of the Fecal Slurry (cFS) for Fecal Microbiota Transplantation (FMT)* (MTC-SOP-cFS-0005). Stop production if the CFU / ml is $<4x10^8$.

7.15 Calculate the volume of cFS needed for a dose of $1x10^8$ CFU, assuming 1 g cFS = 1 ml cFS

7.16 Calculate the volume of cryopreservation media needed for a total volume of 250 ml (i.e., if a $1x10^{11}$ CFU dose requires 50 ml of cFS, 200 ml of cryopreservation media will make for a total volume of 250 ml)

7.17 Transfer the calculated volume of cryopreservation media to a 250 ml cryogenic storage bottle with a 50 ml serologic pipette

7.18 Transfer the calculated volume of cFS for a dose of $1x10^{11}$ CFU [high dose] or $1x10^{10}$ CFU [low dose] with a 50 ml serologic pipette.

7.18.1 Mix the normalized FS with the 50 ml serologic pipette.

7.19 Repeat the generation of 250 ml volume $1x10^{11}$ FS [high] and $1x10^{10}$ FS [low] doses until there is not sufficient cFS for an additional dose.

7.20 If the sample is the first in a donor series (i.e., from day 1 of the FMT donor collection), remove 1.2 ml from the FS in the storage bottle using a repeat pipettor

7.20.1 Save 8 x 100 µl aliquots for future stability testing.

7.21 If the sample is not the first in a donor series, transfer 100 ul of FS to a cryovial for long-term storage.

7.22 Store the FS FMT products and all generated cryovials at -80C until usage or expiration.

7.23 Surface clean all equipment with EtOH.

#### 8.0 ASSOCIATED DOCUMENTS

8.1 MTC-SOP-FS-0003_ Fecal Slurry Cryopreservation Media for Fecal Microbiota Transplantation (FMT)

8.2 MTC-SOP-FS-0004_Sterilization of Biosafety Cabinet and Stomacher for Fecal Slurry Production for Fecal Microbiota Transplantation (FMT)

8.3 MTC-SOP-FS-0005_CFU Count of the Fecal Slurry (FS) for Fecal Microbiota Transplantation (FMT)

#### 9.0 REVISION HISTORY

| DATE | DESCRIPTION OF CHANGE | INITIATOR |
|---|---|---|

### Appendix 12 – MTC01 stability results

#### Stability MTC01 strains in Formulation Buffer over 48 wks

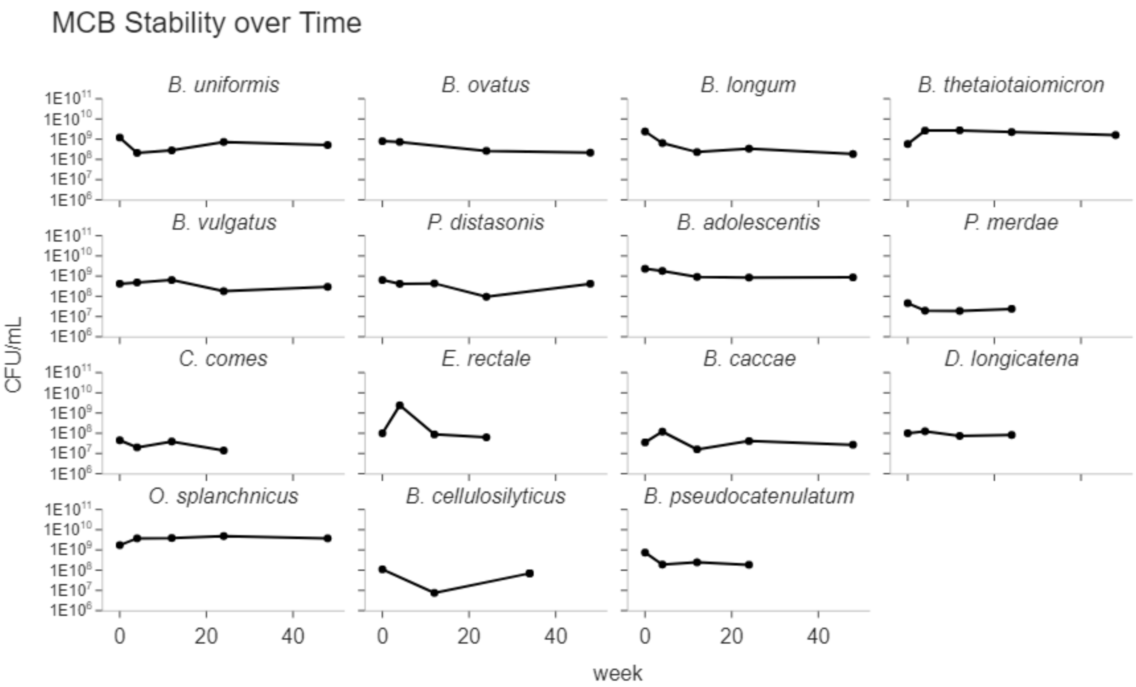

#### Stability MTC01 strain drug substance in Formulation Buffer over 12 wks

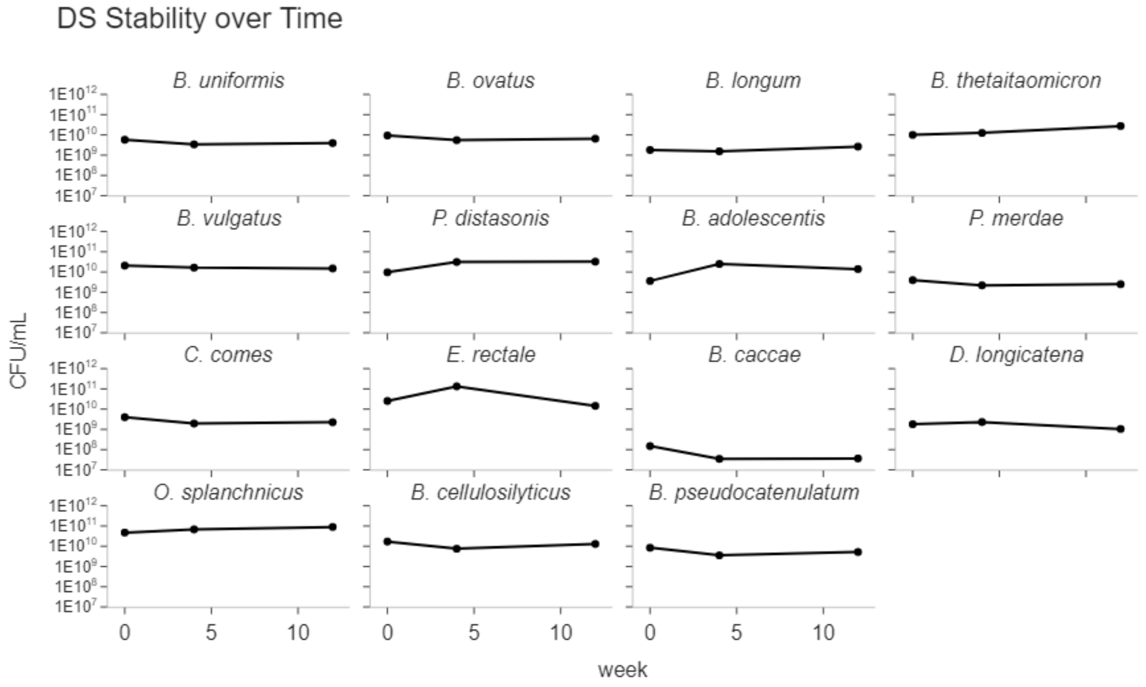

#### Stability of MTC01 strains to multiple freeze-thaws

MCB Freeze-Thaw Stability

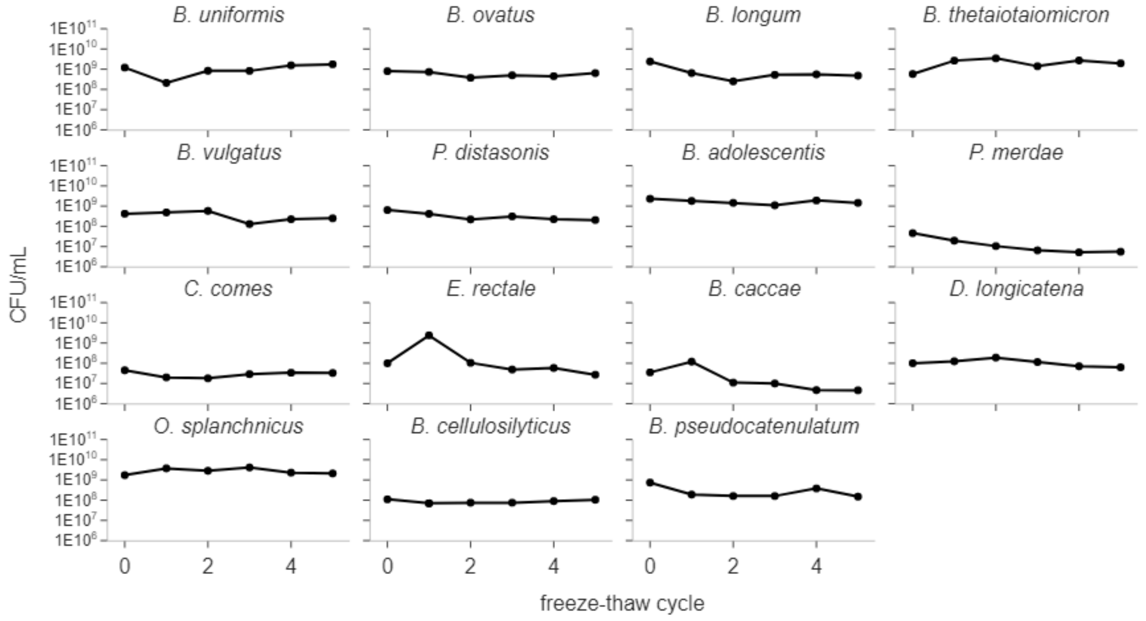

DS Freeze-Thaw Stability

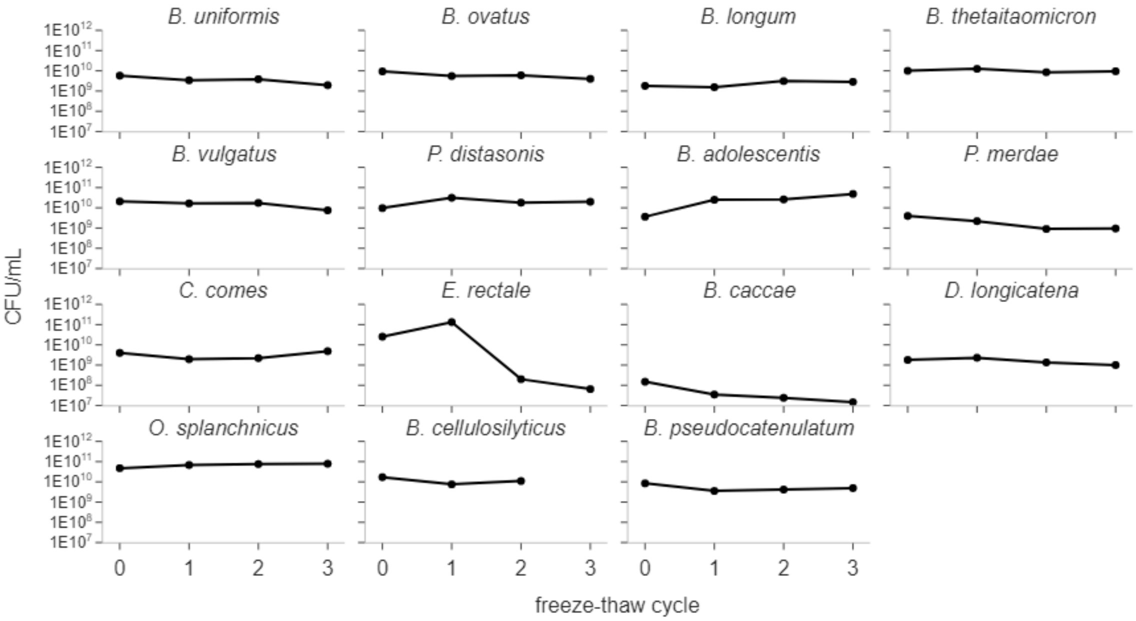

**Stability of MTC01 strains to an extended thaw cycle in glycerol formulation buffer** (to simulate the process of combining strains from DS to make DP)

MCB stability in formulation buffer in anaerobic atmosphere

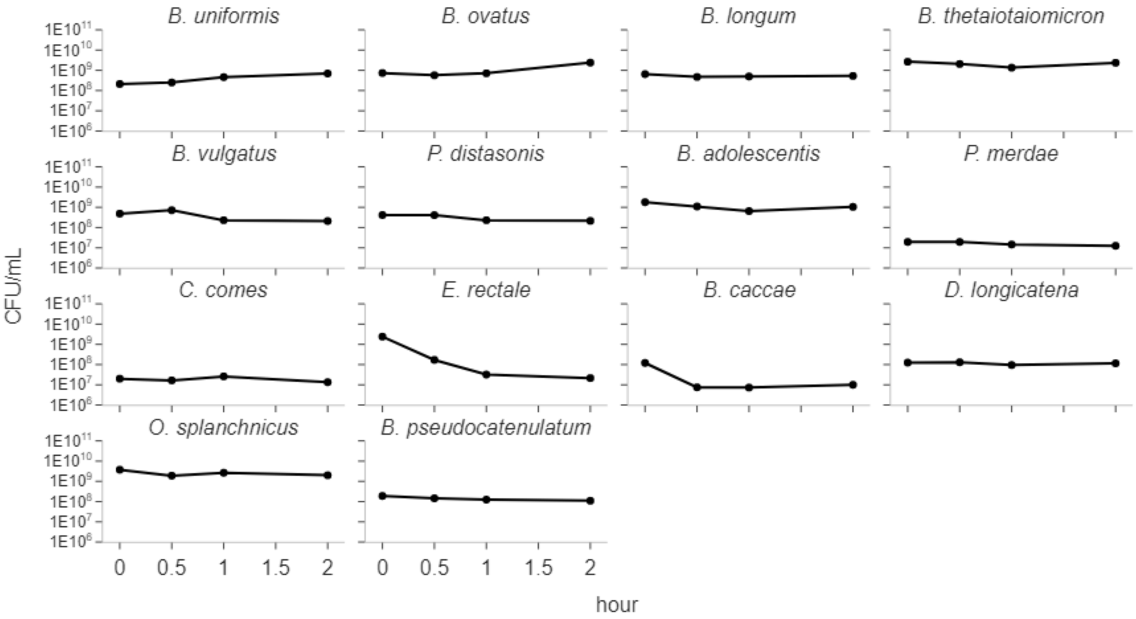

DS stability in formulation buffer in anaerobic atmosphere

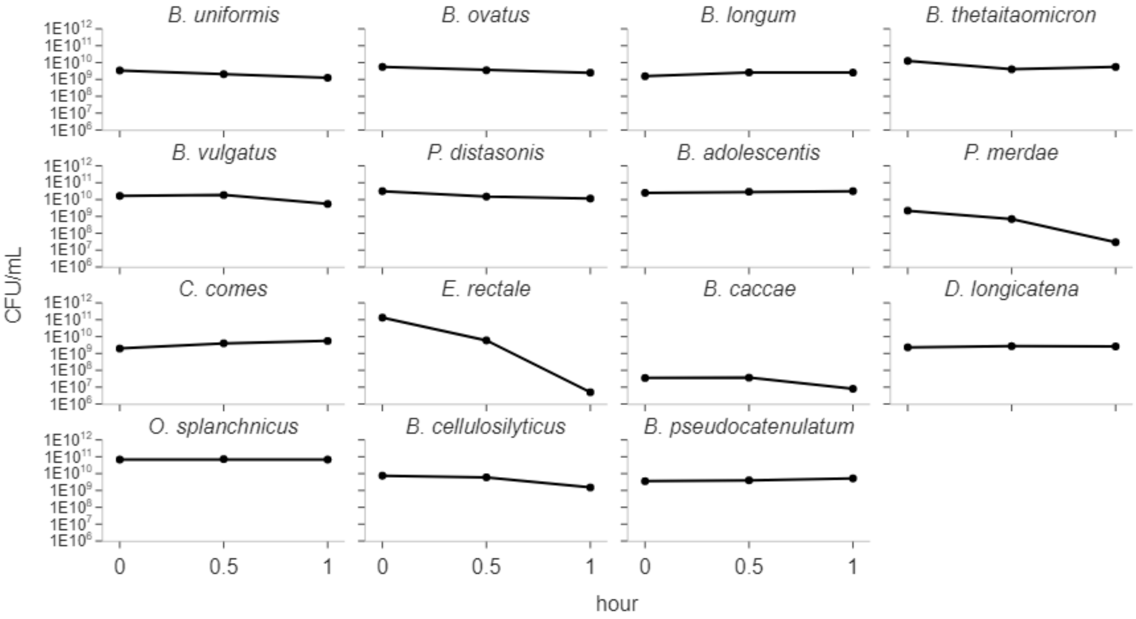
B. uniformis
B. ovatus
B. longum
B. thetaitaomicron
B. vulgatus
P. distasonis
B. adolescentis
P. merdae
C. comes
E. rectale
B. caccae
D. longicatena
O. splanchnicus
B. cellulosilyticus
B. pseudocatenulatum
CFU/mL
1E10^12
1E10^11
1E10^10
1E10^9
1E10^8
1E10^7
1E10^6
0
0.5
1
hour

**DP stability at RT in storage bottle in aerobic environment**
(to simulate the thawing process in the clinic)

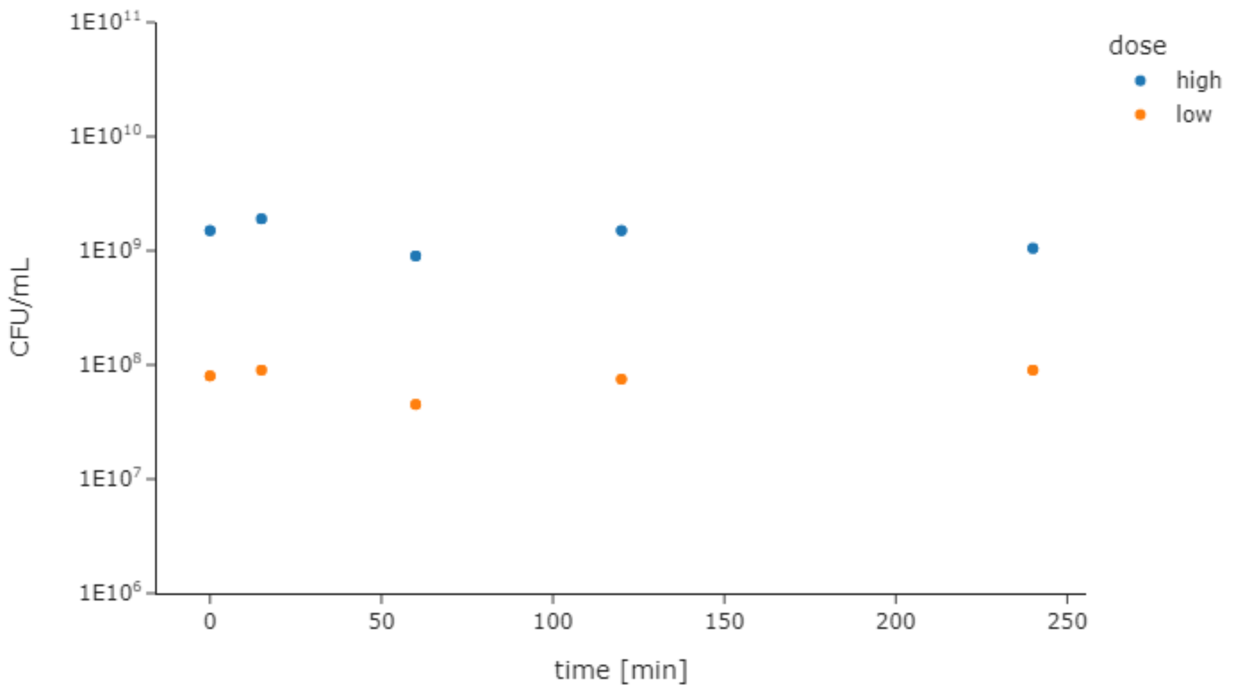

**DP stability at RT in trough in aerobic environment**
(to simulate the process of filling syringes for colonoscopy)

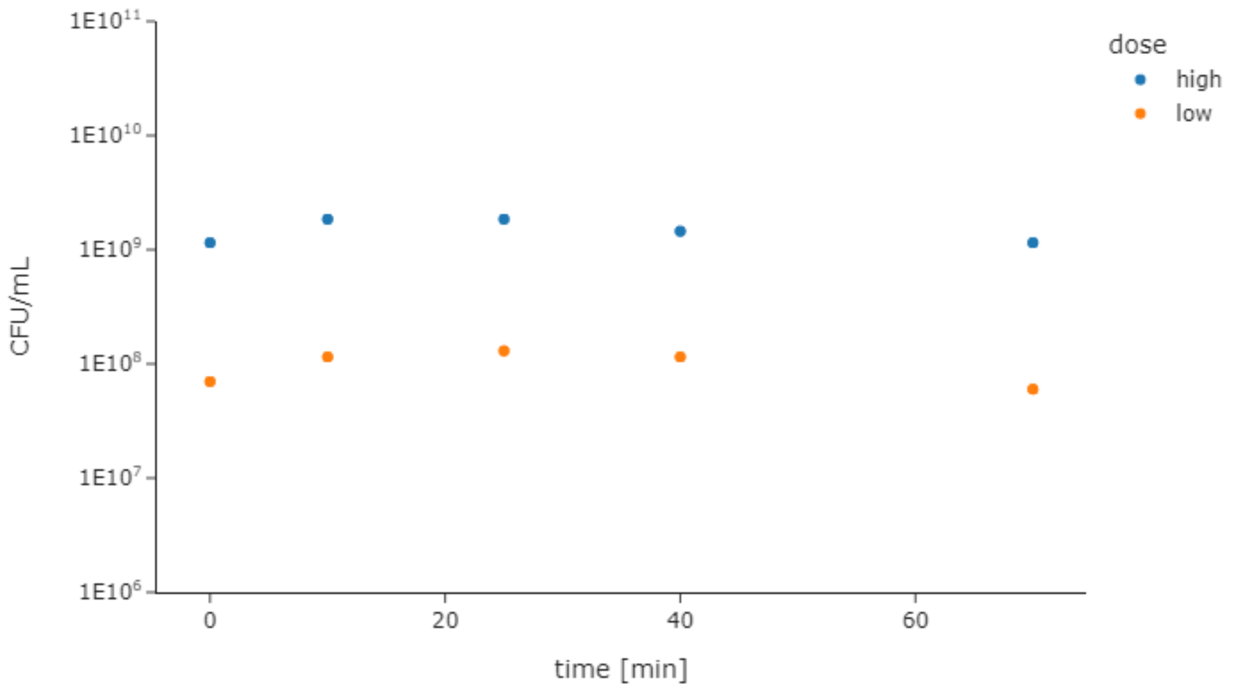

#### DP stability in formulation buffer at -80C (long-term storage conditions)

| | | Total Viable CFU/dose | |
|---|---|---|---|
| **Date** | **Time** | **Low Dose** | **High Dose** |
| ***Specification*** | ***Specification*** | *$6x10^9$-$5x10^{10}$* | *$6x10^{10}$-$5x10^{11}$* |
| **08/12/2022** | **0 weeks** | $6.9x10^9$ | $8.3x10^{10}$ |
| **09/09/2022** | **4 weeks** | $6.6x10^9$ | $8.9x10^{10}$ |
| 11/04/2022 | 12 weeks | | |
| 01/27/2023 | 24 weeks | | |
| 07/14/2023 | 48 weeks | | |
| 12/29/2023 | 72 weeks | | |
| 06/14/2024 | 96 weeks | | |

### Appendix 13 – Sterilization of non-disposable items in the generation of FS

### SOP for non-disposable item sterilization with Quatricide PV-15 prior to the generation of FS

| Microbiome Translational Center (Icahn School of Medicine at Mount Sinai) | Microbiome Translational Center<br>Standard Operating Procedures |
|---|---|
| **Title: Sterilization of Biosafety Cabinet and Stomacher for the Fecal Slurry Production for Fecal Microbiota Transplantation** | **SOP No: MTC-SOP-FS-0004**<br>**Revision: 0**<br>**Effective Date: 10/16/2022**<br>**Supersedes Date: NOW** |

| **Written by:** | | **Date:** |
|---|---|---|
| Lukas Bethlehem, PhD | | |
| **Reviewed By:** | | **Date:** |
| Jeremiah Faith, PhD<br>Faculty Director | | |
| **Approved By:** | | **Date:** |
| Ilaria Mogno, PhD<br>Quality Assurance Specialist | | |

#### 1.0 INTRODUCTION

The purpose of this SOP is to provide instructions for the ionized hydrogen peroxide (iHP) sterilization of the biosafety cabinet used for the generation of human fecal slurries as Drug Substance (DS) for fecal microbiota transplantion (FMT).

#### 2.0 RESPONSIBLE PERSONNEL

2.1 The Facility Director is responsible for identifying new SOPs to be written, drafting new SOPs, suggesting revisions to existing SOPs, reviewing and approving all SOPs, and ensuring that all laboratory personnel is properly trained and following all SOPs.

2.2 The Quality Assurance (QA) Lead is responsible for coordinating the preparation, approval, distribution, and maintenance of this SOP, for suggesting revisions to existing SOPs, and for ensuring that all SOPs are being followed by the required personnel.

2.3 The Approver can and should name representatives, who are qualified to take over the duties in case of an emergency or during absence of the Approver/reviewer.

2.4 Technicians and other Lab personal are responsible for performing the procedures described in this SOP.

#### 3.0 DEFINITIONS

3.1 SOP = Standard Operating Procedure

3.2 QA = Quality Assurance

3.3 MTC = Microbiome Translational Center
3.4 FMT = Fecal Microbiota Transplantation

#### 4.0 SAFETY

4.1 Wear protective gloves, lab coat and goggles while performing this procedure.
4.2 Perform all steps with the dedicated equipment for production of LBPs and follow instructions carefully to avoid contamination of the product or exposure to the operator.

#### 5.0 MATERIALS

5.1 Equipment

| Item | Manufacturer | Catalog No. |
|---|---|---|
| Biosafety Cabinet | Labconco | 130475124 A |
| Stomacher 400 EVO Paddle Blender | Seward | 0400/002/00 |

5.2 Supplies

| Item | Manufacturer | Catalog No. |
|---|---|---|
| Spray Bottle | Not specified | Not specified |

5.3 Reagents

| Item | Manufacturer | Catalog No. |
|---|---|---|
| Distilled Water (Milli-Q) | Millipore-Sigma | Not specified |
| Quatricide-PV15 | Pharmacal Research Labs | Fisher, NC9371434 |

#### 6.0 PROCEDURE

##### Preparation

Prepare a spray bottle of Quatricide PV-15 with 1.95 ml Quatricide PV-15 and water to a final volume of 500 ml.

##### Sterilization

6.1 Surface disinfection.

6.1.1 Spray the biosafety cabinet work surface, the interior walls, and the interior surface of the window with the Quatricide PV-15 solution.

6.1.2 Bring the pipettes inside the biosafety cabinet and spray them with Quatricide PV-15 solution.

6.1.3 Spray the stomacher with Quatricide PV-15 solution.

6.1.4 Allow a 10 minute contact time.

6.1.5 Wipe all surfaces of the biological safety cabinet, pipettes, and stomacher clean with a paper towel.

#### 7.0 ASSOCIATED DOCUMENTS

7.1 MTC-SOP-FS-0002_Generation of CFU normalized Fecal Slurry (FS) for Fecal Microbiota Transplantation (FMT)

#### 8.0 REVISION HISTORY

| DATE | DESCRIPTION OF CHANGE | INITIATOR |
|---|---|---|

### Technical data sheet of activity of Quatricide PV-15

### Pharmacal Research Laboratories, Inc.

technical information

#### Hospital Disinfection

##### Efficacy For QUATRICIDE PV Second Generation (at 2 ounces per gallon)
##### Efficacy For QUATRICIDE PV-15 Second Generation (at 1/2 ounce per gallon)

This product is bactericidal according to the AOAC Use Dilution Test method on hard inanimate surfaces modified in the presence of 5% organic serum and 400 ppm hard water with a 10 minutes contact time against:

(Testing is performed per the AOAC UDT/GST method (DIS/TSS-1). Sixty carriers are required on 3 separate lots, one of which must be > 60 days old against Pseudomonas aeruginosa, Salmonella choleraesuis and Staphylococcus aureus. Killing of 59 out of 60 carriers is required (total carriers = 540).)

| | CARRIER POPULATION | SAMPLE | # CARRIERS | # POSITIVE |
|---|---|---|---|---|
| *Pseudomonas aeruginosa* ATCC #15442 | 3.9 X $10^4$ CFU/Carrier | A (60 Days Old) | 60 | 0/60 |
| | | B | 60 | 0/60 |
| | | C | 60 | 1/60 |
| *Salmonella choleraesuis* ATCC #10708 | 1.03 X $10^6$ CFU/Carrier | A (60 Days Old) | 60 | 1/60 |
| | | B | 60 | 1/60 |
| | | C | 60 | 0/60 |
| *Staphylococcus aureus* ATCC #6538 | 7.0 X $10^4$ CFU/Carrier | A (60 Days Old) | 60 | 0/60 |
| | | B | 60 | 0/60 |
| | | C | 60 | 0/60 |

#### Supplemental Organisms

(Testing is performed per the AOAC UDT/GST method. Ten carriers are required on 2 separate lots against each supplemental organism. Killing of 10 out of 10 carriers is required (total carriers = 20).)

| | CARRIER POPULATION | SAMPLE | # CARRIERS | # POSITIVE |
|---|---|---|---|---|
| *Acinetobacter baumannii* ATCC 19003 | 5.1 X $10^6$ CFU/Carrier | A | 10 | 0/10 |
| | | B | 10 | 0/10 |
| *Acinetobacter Iwoffi* ATCC 15309 | 5.7 X $10^5$ CFU/Carrier | A | 10 | 0/10 |
| | | B | 10 | 0/10 |
| *Acinetobacter Iwoffi* ATCC 9957 | 4.0 X $10^5$ CFU/Carrier | A | 10 | 0/10 |
| | | B | 10 | 0/10 |
| *Bordetella bronchiseptica* ATCC 10580 | 9.4 X $10^6$ CFU/Carrier | A | 10 | 0/10 |
| | | B | 10 | 0/10 |
| *Citrobacter freundii* ATCC 8090 | 3.9 X $10^5$ CFU/Carrier | A | 10 | 0/10 |
| | | B | 10 | 0/10 |
| *Clostridium difficile ATCC 9689* | 9.7 X $10^5$ CFU/Carrier | A | 10 | 0/10 |
| | | B | 10 | 0/10 |
| | | C | 10 | 0/10 |
| *Enterobacter aerogenes* ATCC 13048 | 2.35 X $10^7$ CFU/Carrier | A | 10 | 0/10 |
| | | B | 10 | 0/10 |

| | | | | |
|---|---|---|---|---|
| *Enterobacter agglomerans* ATCC 27155 | 3.9 X $10^{5}$ CFU/Carrier | A | 10 | 0/10 |
| | | B | 10 | 0/10 |
| *Enterobacter cloacae* ATCC 13047 | 3.3 X $10^{7}$ CFU/Carrier | A | 10 | 0/10 |
| | | B | 10 | 0/10 |
| *Enterococcus faecalis* ATCC 19433 | 6.2 X $10^{5}$ CFU/Carrier | A | 10 | 0/10 |
| | | B | 10 | 0/10 |
| *Enterococcus faecalis* Vancomycin Resistant (VRE) ATCC 51299 | 1.3 X $10^{7}$ CFU/Carrier | A | 10 | 0/10 |
| | | B | 10 | 0/10 |
| *Enterococcus hirae* ATCC 10541 | 1.19 X $10^{5}$ CFU/Carrier | A | 10 | 0/10 |
| | | B | 10 | 0/10 |
| *Escherichia coli* ATCC 11229 | 1.3 X $10^{7}$ CFU/Carrier | A | 10 | 0/10 |
| | | B | 10 | 0/10 |
| *Escherichia coli Spectrum B-Lactamase (ESBL) ATCC BAA-196* | 4.6 X $10^{6}$ CFU/Carrier | A | 10 | 0/10 |
| | | B | 10 | 0/10 |
| *Escherichia coli O111:H8 ATCC BAA-184* | 4.3 X $10^{6}$ CFU/Carrier | A | 10 | 0/10 |
| | | B | 10 | 0/10 |
| *Escherichia coli* Tetracycline Resistant ATCC 47041 | 3.1 X $10^{5}$ CFU/Carrier | A | 10 | 0/10 |
| | | B | 10 | 0/10 |
| *Fusobacterium necrophorum* ATCC 27852 | 5.8 X $10^{5}$ CFU/Carrier | A | 10 | 0/10 |
| | | B | 10 | 0/10 |
| *Klebsiella oxytoca* ATCC 13182 | 1.07 X $10^{6}$ CFU/Carrier | A | 10 | 0/10 |
| | | B | 10 | 0/10 |
| *Klebsiella pneumoniae* ATCC 13883 | 1.2 X $10^{6}$ CFU/Carrier | A | 10 | 0/10 |
| | | B | 10 | 0/10 |
| *Listeria monocytogenes* ATCC 19117 | 7.7 X $10^{6}$ CFU/Carrier | A | 10 | 0/10 |
| | | B | 10 | 0/10 |
| *Micrococcus luteus* ATCC 14452 | 1.1 X $10^{5}$ CFU/Carrier | A | 10 | 0/10 |
| | | B | 10 | 0/10 |
| *Micrococcus luteus* ATCC 4698 | 4.8 X $10^{5}$ CFU/Carrier | A | 10 | 0/10 |
| | | B | 10 | 0/10 |
| *Pasturella multocida* ATCC 12947 | 1.32 X $10^{7}$ CFU/Carrier | A | 10 | 0/10 |
| | | B | 10 | 0/10 |
| *Proteus vulgaris* ATCC 13315 | 1.9 X $10^{4}$ CFU/Carrier | A | 10 | 0/10 |
| | | B | 10 | 0/10 |
| *Proteus vulgaris* ATCC 9920 | 1.24 X $10^{5}$ CFU/Carrier | A | 10 | 0/10 |
| | | B | 10 | 0/10 |
| *Pseudomonas aeruginosa* Tetracycline Resistant ATCC 27853 | 3.5 X $10^{6}$ CFU/Carrier | A | 10 | 0/10 |
| | | B | 10 | 0/10 |

| | | | | |
|---|---|---|---|---|
| *Pseudomonas cepacia* ATCC 25416 | 1.63 X $10^6$ CFU/Carrier | A | 10 | 0/10 |
| | | B | 10 | 0/10 |
| *Salmonella enterica* ATCC 23564 | 9.2 X $10^4$ CFU/Carrier | A | 10 | 0/10 |
| | | B | 10 | 0/10 |
| *Salmonella enterica* ATCC 4931 | 1.3 X $10^6$ CFU/Carrier | A | 10 | 0/10 |
| | | B | 10 | 0/10 |
| *Salmonella choleraesuis serotype* pullorum ATCC 19945 | 7.1 X $10^5$ CFU/Carrier | A | 10 | 0/10 |
| | | B | 10 | 0/10 |
| *Salmonella typhi* ATCC 6539 | 8.3 X $10^6$ CFU/Carrier | A | 10 | 0/10 |
| | | B | 10 | 0/10 |
| *Salmonella typhimurium ATCC 23564* | 1.5 X $10^5$ CFU/Carrier | A | 10 | 0/10 |
| | 5.6 X $10^5$ CFU/Carrier | B | 10 | 0/10 |
| *Serratia marcescens* ATCC 14756 | 6.2 X $10^6$ CFU/Carrier | A | 10 | 0/10 |
| | | B | 10 | 0/10 |
| *Serratia marcescens* ATCC 9103 | 6.0 X $10^6$ CFU/Carrier | A | 10 | 0/10 |
| | | B | 10 | 0/10 |
| *Shigella flexneri* ATCC 12022 | 2.6 X $10^4$ CFU/Carrier | A | 10 | 0/10 |
| | | B | 10 | 0/10 |
| *Shigella flexneri* ATCC 9380 | 1.99 X $10^6$ CFU/Carrier | A | 10 | 0/10 |
| | | B | 10 | 0/10 |
| *Shigella sonnei* ATCC 25931 | 1.04 X $10^6$ CFU/Carrier | A | 10 | 0/10 |
| | | B | 10 | 0/10 |
| *Staphylococcus aureus* ATCC 14154 | 9.2 X $10^5$ CFU/Carrier | A | 10 | 0/10 |
| | | B | 10 | 0/10 |
| *Staphylococcus aureus* ATCC 25923 | 6.6 X $10^6$ CFU/Carrier | A | 10 | 0/10 |
| | | B | 10 | 0/10 |
| *Staphylococcus aureus sub species* aureus ATCC 33586 | 7.2 X $10^4$ CFU/Carrier | A | 10 | 0/10 |
| | | B | 10 | 0/10 |
| *Staphylococcus aureus* Methicillin Resistant (MRSA) ATCC 33592 | 5.4 X $10^6$ CFU/Carrier | A | 10 | 0/10 |
| | | B | 10 | 0/10 |
| *Staphylococcus aureus Community Associated Methicillin Resistant (CA-MRSA)* | 6.3 X $10^6$ CFU/Carrier | A | 10 | 0/10 |
| | | B | 10 | 0/10 |
| *Staphylococcus aureus Community Associated Methicillin Resistant (CA-MRSA) (NARSA NRS384) Genotype USA300)* | 1.60 X $10^6$ CFU/Carrier | A | 10 | 0/10 |
| | | B | 10 | 0/10 |
| *Staphylococcus aureus* Vancomycin Intermediate Resistant (VISA) ATCC 5836 | 3.2 X $10^6$ CFU/Carrier | A | 10 | 0/10 |
| | | B | 10 | 0/10 |
| *Staphylococcus epidermidis* ATCC 14990 | 1.56 X $10^6$ CFU/Carrier | A | 10 | 0/10 |
| | | B | 10 | 0/10 |

| | | | | |
|---|---|---|---|---|
| *Staphylococcus epidermidis* Antibiotic resistant ATCC 51625 | 8.6 X $10^5$ CFU/Carrier | A | 10 | 0/10 |
| | | B | 10 | 0/10 |
| *Staphylococcus haemolyticus* ATCC 29970 | 9.5 X $10^5$ CFU/Carrier | A | 10 | 0/10 |
| | | B | 10 | 0/10 |
| *Streptococcus agalactiae* ATCC 13813 | 5.6 X $10^6$ CFU/Carrier | A | 10 | 0/10 |
| | | B | 10 | 0/10 |
| *Streptococcus mutans ATCC 25175* | 1.02 X $10^6$CFU/Carrier | A | 10 | 0/10 |
| | 1.3 X $10^4$CFU/Carrier | B | 10 | 0/10 |
| *Streptococcus pneumonia* Penicillin Resistant ATCC 51915 | 9.6 X $10^4$ CFU/Carrier | A | 10 | 0/10 |
| | | B | 10 | 0/10 |
| *Streptococcus pyogenes* ATCC 19615 | 4.7 X $10^4$ CFU/Carrier | A | 10 | 0/10 |
| | | B | 10 | 0/10 |
| *Vibrio cholera* ATCC 11623 | 1.0 X $10^6$ CFU/Carrier | A | 10 | 0/10 |
| | | B | 10 | 0/10 |
| *Yersinia enterocolitica* ATCC 23715 | 1.2 X $10^7$ CFU/Carrier | A | 10 | 0/10 |
| | | B | 10 | 0/10 |

#### Virucidal against

##### Efficacy For QUATRICIDE PV Second Generation (at 2 ounces per gallon)
##### Efficacy For QUATRICIDE PV-15 Second Generation (at 1/2 ounce per gallon)

This product was evaluated in the presence of 5% serum and 400 ppm hard water with a 10 minute contact time and found to be effective against the following viruses on hard nonporous environmental surfaces.

(Testing is performed per EPA Guidance (DIS/TSS-7). Two separate lots are tested. Inactivation of virus must be demonstrated at all dilutions when no cytotoxicity is observed or at all dilutions above the cytotoxic level when it is observed. The data must demonstrate a 3-log reduction in viral titer for both lots.) (3 lots and 4-Logreduction for Canada).

| | DRIED VIRUS CONTROL | SAMPLE | RESULT | LOG REDUCTION |
|---|---|---|---|---|
| Avian Infectious Bronchitis virus Beaudette IB42 | 6.42 $Log_{10}$ | A | ≤0.5 $Log_{10}$ | ≥5.92 $Log_{10}$ |
| | | B | ≤0.5 $Log_{10}$ | ≥5.92 $Log_{10}$ |
| | 6.5 $Log_{10}$ | C | ≤0.5 $Log_{10}$ | ≥6.0 $Log_{10}$ |
| Avian Influenza A (H3N2) virus (Avian Reassortant) (ATTC VR-2072) | 4.75 $Log_{10}$ | A | ≤0.5 $Log_{10}$ | ≥4.25 $Log_{10}$ |
| | | B | ≤0.5 $Log_{10}$ | ≥4.25 $Log_{10}$ |
| | | C | ≤0.5 $Log_{10}$ | ≥4.25 $Log_{10}$ |
| Avian Influenza A (H5N1) virus | 6.75 $Log_{10}$ | A | ≤0.5 $Log_{10}$ | ≥6.25 $Log_{10}$ |
| | | B | ≤0.5 Log | ≥6.25 $Log_{10}$ |
| Canine Coronavirus ATCC VR-809 | 4.5 $Log_{10}$ | A | ≤0.5 $Log_{10}$ | ≥4.0 $Log_{10}$ |
| | | B | ≤0.5 $Log_{10}$ | ≥4.0 $Log_{10}$ |
| | 4.75 $Log_{10}$ | C | ≤0.5 $Log_{10}$ | ≥4.25 $Log_{10}$ |
| Canine Distemper virus ATCC VR-128 | 6.25 $Log_{10}$ | A | ≤0.5 $Log_{10}$ | ≥5.75 $Log_{10}$ |
| | | B | ≤0.5 $Log_{10}$ | ≥5.75 $Log_{10}$ |
| | 6.75 $Log_{10}$ | C | ≤0.5 $Log_{10}$ | ≥6.25 $Log_{10}$ |

| | | | | |
|---|---|---|---|---|
| Chlamydia psittaci ATCC VR-125 | 7.25 $Log_{10}$ | A | ≤0.5 $Log_{10}$ | ≥6.75 $Log_{10}$ |
| | | B | ≤0.5 $Log_{10}$ | ≥6.75 $Log_{10}$ |
| | 4.75 $Log_{10}$ | C | ≤0.5 $Log_{10}$ | ≥4.25 $Log_{10}$ |
| Cytomegalovirus ATCC VR-538 | 4.5 $Log_{10}$ | A | ≤0.5 $Log_{10}$ | ≥4.0 $Log_{10}$ |
| | | B | ≤0.5 $Log_{10}$ | ≥4.0 $Log_{10}$ |
| | | C | ≤0.5 $Log_{10}$ | ≥4.0 $Log_{10}$ |
| Feline Picornavirus ATCC VR-649 | 4.5 $Log_{10}$ | A | ≤0.5 $Log_{10}$ | ≥4.0 $Log_{10}$ |
| | | B | ≤0.5 $Log_{10}$ | ≥4.0 $Log_{10}$ |
| | 5.75 $Log_{10}$ | C | ≤0.5 $Log_{10}$ | ≥5.25 $Log_{10}$ |
| Hepatitis B Virus | 5.06 $Log_{10}$ | A | 0.27 $Log_{10}$ | 4.79 $Log_{10}$ |
| | 5.20 $Log_{10}$ | B | 0.41 $Log_{10}$ | 4.79 $Log_{10}$ |
| | 5.06 $Log_{10}$ | Confirmatory B | 0.27 $Log_{10}$ | 4.79 $Log_{10}$ |
| Hepatitis C Virus | 6.21 $Log_{10}$ | A | 0.24 $Log_{10}$ | 5.97 $Log_{10}$ |
| | 6.21 $Log_{10}$ | B | 0.42 $Log_{10}$ | 5.79 $Log_{10}$ |
| | 6.06 $Log_{10}$ | Confirmatory B | 0.13 $Log_{10}$ | 5.93 $Log_{10}$ |
| Herpes Simplex Virus Type 1 ATCC VR-773 | 5.5 $Log_{10}$ | A | ≤0.5 $Log_{10}$ | ≥5.0 $Log_{10}$ |
| | | B | ≤0.5 $Log_{10}$ | ≥5.0 $Log_{10}$ |
| | 6.0 $Log_{10}$ | C | ≤0.5 $Log_{10}$ | ≥5.5 $Log_{10}$ |
| Herpes Simplex Virus Type 2 ATCC VR-734 | 6.0 $Log_{10}$ | A | ≤0.5 $Log_{10}$ | ≥5.5 $Log_{10}$ |
| | | B | ≤0.5 $Log_{10}$ | ≥5.5 $Log_{10}$ |
| | 5.75 $Log_{10}$ | C | ≤0.5 $Log_{10}$ | ≥5.25 $Log_{10}$ |
| Human Coronavirus ATCC VR-740 | 4.5 $Log_{10}$ | A | ≤0.5 $Log_{10}$ | ≥4.0 $Log_{10}$ |
| | | B | ≤0.5 $Log_{10}$ | ≥4.0 $Log_{10}$ |
| | 4.5 $Log_{10}$ | C | ≤0.5 $Log_{10}$ | ≥4.0 $Log_{10}$ |
| Human Immunodeficiency Virus type 1 (HIV 1) HTLV-IIIB | 5.75 $Log_{10}$ | A | ≤1.5 $Log_{10}$ | ≥4.25$Log_{10}$ |
| | | B | ≤1.5 $Log_{10}$ | ≥4.25$Log_{10}$ |
| | | C | ≤1.5 $Log_{10}$ | ≥4.25$Log_{10}$ |
| Infectious Bovine Rhinotracheitis virus ATCC VR-188 | 4.5 $Log_{10}$ | A | ≤0.5 $Log_{10}$ | ≥4.0 $Log_{10}$ |
| | | B | ≤0.5 $Log_{10}$ | ≥4.0 $Log_{10}$ |
| | 4.75 $Log_{10}$ | C | ≤0.5 $Log_{10}$ | ≥4.25 $Log_{10}$ |
| Influenza A virus ATCC VR-544 | 6.5 $Log_{10}$ | A | ≤0.5 $Log_{10}$ | ≥6.0 $Log_{10}$ |
| | | B | ≤0.5 $Log_{10}$ | ≥6.0 $Log_{10}$ |
| | 6.0 $Log_{10}$ | C | ≤0.5 $Log_{10}$ | ≥5.5 $Log_{10}$ |

| | | | | |
|---|---|---|---|---|
| Pseudorabies virus ATCC VR-135 | 6.25 $Log_{10}$ | A | ≤0.5 $Log_{10}$ | ≥5.75 $Log_{10}$ |
| | | B | ≤0.5 $Log_{10}$ | ≥5.75 $Log_{10}$ |
| | 5.5 $Log_{10}$ | C | ≤0.5 $Log_{10}$ | ≥5.0 $Log_{10}$ |
| Respiratory syncytial virus ATCC VR-26 | 4.5 $Log_{10}$ | A | ≤0.5 $Log_{10}$ | ≥4.0 $Log_{10}$ |
| | | B | ≤0.5 $Log_{10}$ | ≥4.0 $Log_{10}$ |
| | 5.0 $Log_{10}$ | C | ≤0.5 $Log_{10}$ | ≥4.5 $Log_{10}$ |
| SARS Associated Coronavirus | 6.5 $Log_{10}$ | A | ≤3.5 $Log_{10}$ | ≥3.0 $Log_{10}$ |
| | | B | ≤3.5 $Log_{10}$ | ≥3.0 $Log_{10}$ |
| Transmissible Gastroenteritis virus | 4.75 $Log_{10}$ | A | ≤0.5 $Log_{10}$ | ≥4.25 $Log_{10}$ |
| | | B | ≤0.5 $Log_{10}$ | ≥4.25 $Log_{10}$ |
| | 6.25 $Log_{10}$ | C | ≤0.5 $Log_{10}$ | ≥5.75 $Log_{10}$ |
| Vaccinia virus ATCC VR-119 | 6.75 $Log_{10}$ | A | ≤0.5 $Log_{10}$ | ≥6.25 $Log_{10}$ |
| | | B | ≤0.5 $Log_{10}$ | ≥6.25 $Log_{10}$ |
| | 6.5 $Log_{10}$ | C | ≤0.5 $Log_{10}$ | ≥6.0 $Log_{10}$ |

#### Virucidal against

##### Efficacy For QUATRICIDE PV Second Generation (at 9 ounces per gallon)
##### Efficacy For QUATRICIDE PV-15 Second Generation (at 2.25 ounces per gallon)

This product was evaluated in the presence of 5% serum and 400 ppm hard water with a 10 minute contact time and found to be effective against the following viruses on hard nonporous environmental surfaces.

(Testing is performed per EPA Guidance (DIS/TSS-7). Two separate lots are tested. Inactivation of virus must be demonstrated at all dilutions when no cytotoxicity is observed or at all dilutions above the cytotoxic level when it is observed. The data must demonstrate a 3-log reduction in viral titer for both lots.) (3 lots and 4-Log reduction for Canada).

| | DRIED VIRUS CONTROL | SAMPLE | RESULT | LOG REDUCTION |
|---|---|---|---|---|
| Canine Parvovirus Type 2b, Nike Strain | 7.5 $Log_{10}$ | A | ≤3.5 $Log_{10}$ | ≥4.0 $Log_{10}$ |
| | | B | ≤3.5 $Log_{10}$ | ≥4.0 $Log_{10}$ |

#### Fungicidal against

##### Efficacy For QUATRICIDE PV Second Generation (at 2 ounces per gallon)
##### Efficacy For QUATRICIDE PV-15 Second Generation (at 1/2 ounce per gallon)

This product was evaluated in the presence of 5% serum and 400 ppm hard water with a 10 minute contact time and found to be effective against the following fungi on hard nonporous environmental surfaces.

(Testing is performed per the AOAC fungicidal method (DIS/TSS-6). Two separate lots are tested against Trichophyton mentagrophytes in a suspension test. Killing of all fungal spores in 10 minutes is required.)

| | CARRIER POPULATION | SAMPLE | # CARRIERS | # POSITIVE |
|---|---|---|---|---|
| *Candida albicans* ATCC #10231 | 1.57 X $10^5$ CFU/Carrier | A | 10 | 0/10 |
| | | B | 10 | 0/10 |
| *Trichophyton mentagrophytes* ATCC #9533 | 1.10 X $10^5$ CFU/Carrier | A | 10 | 0/10 |
| | | B | 10 | 0/10 |

#### Mold and Mildew Control

Efficacy For QUATRICIDE PV Second Generation (at 2 ounces per gallon)
Efficacy For QUATRICIDE PV-15 Second Generation (at 1/2 ounce per gallon)

Use this product to control the growth of mold and mildew and their odors on hard, non-porous surfaces. Thoroughly wet all treated surfaces completely. Let air dry. Repeat application weekly or when growth or odor reappears.

| | TILE NUMBER | UNTREATED AFTER 7 DAYS | SAMPLE A AFTER 7 DAYS | SAMPLE B AFTER 7 DAYS |
|---|---|---|---|---|
| Aspergillus niger ATCC #16404 | 1 | Growth 90% | No Growth 0% | No Growth 0% |
| | 2 | Growth 70% | No Growth 0% | No Growth 0% |
| | 3 | Growth 90% | No Growth 0% | No Growth 0% |
| | 4 | Growth 80% | No Growth 0% | No Growth 0% |
| | 5 | Growth 80% | No Growth 0% | No Growth 0% |
| | 6 | Growth 90% | No Growth 0% | No Growth 0% |
| | 7 | Growth 80% | No Growth 0% | No Growth 0% |
| | 8 | Growth 70% | No Growth 0% | No Growth 0% |
| | 9 | Growth 90% | No Growth 0% | No Growth 0% |
| | 10 | Growth 70% | No Growth 0% | No Growth 0% |

#### Non-Food Contact Surface Sanitizer

Add 2 ounces of of Quatricide PV second generation or 1/2 ounce of Quatricide PV-15 second generation to 1 gallon of water to sanitize hard porous and non-porous non-food contact surfaces. Treated surfaces must remain wet for 3 minutes. Then wipe with sponge, mop or cloth or allow to air dry. At this dilution food contact surfaces must be rinsed.

(Testing is performed per EPA Guidance (DIS/TSS-10). Three lots are required, one of which must be > 60 days old. Testing is performed against Staphylococcus aureus and Klebsiella pneumoniae containing 5% organic load. Enterobacter aerogenes may be substituted for Klebsiella pneumoniae. The results must show a reduction of at least 99.9% (3 $Log_{10}$) in the number of each test microorganism over the parallel control count within 5 minutes.)

| | CARRIER POPULATION | SAMPLE | 3 MINUTE KILL CFU/CARRIER | PERCENT KILL |
|---|---|---|---|---|
| *Klebsiella pneumoniae* ATCC 4352 | 6.04 $Log_{10}$ | A (60 Days Old) | >3.56 $Log_{10}$ | >99.9 |
| | | B | >3.56 $Log_{10}$ | >99.9 |
| | | C | >3.56 $Log_{10}$ | >99.9 |
| *Staphylococcus aureus* ATCC #6538 | 6.69 $Log_{10}$ | A (60 Days Old) | >5.21 $Log_{10}$ | >99.9 |
| | | B | >4.82 $Log_{10}$ | >99.9 |
| | | C | >5.21 $Log_{10}$ | >99.9 |

Nothing contained herein grants or extends a license, express or implied, in connection with patents, issued or pending, of the manufacturer or others, The information contained herein is based on the manufacturer's own study and the works of others. The manufacturer makes no warranties, expressed or implied, as to the accuracy, completeness, or adequacy of the information contained herein. The manufacturer shall not be liable (regardless of fault) to the vendee's employees, or anyone for any direct, special or consequential damages arising out of or in connection with the accuracy, completeness, adequacy or furnishing of such information.

### Appendix 14 – Donor Health Screening Questionnaire and Physical Exam

#### *FMT DONOR: Intake History/Physical Exam*

Name………………………………………….. Date of Birth……………………………………………

Phone/Contact…………………………………………….

***Past Medical History:***

Last Physician Visit, for…………………………………………………………………………...

Childhood Illnesses………………………………………. Chickenpox?.....................Zoster?...................

Adult Illnesses……………………………………………………………………………………..

Hospitalizations……………………………………………………………………………………

Surgery……………………………………………… Trauma…………………………………………

Fractures/Injuries/MVAs…………………………………………… Prostheses?………………………..

Headaches?………………………………………………………… Chronic Pain?................................

Psychiatric Hx……………………………………………………………………………………..

Any diarrheal ds/IBS/sensitive stomach/IBD/ulcer……………………………………………………..

Endoscopy Hx……………………………………………………………………………………...

GB/Hepatitis/liver ds……………………renal ds/stones……………………………………………………

Rheum/auto immune ?..........................................Skin disease………………………..…………………

***Immunization/Exposure History:***

Last Td…………………… Other recent…………………………… Any side effects?……………………

Military service?…………………………… Overseas travel?……………………………………………...

………………………………………………………………………………………………………..

Antibiotic Hx: use w/in 6 months ☐………………………………………………………………..........

Transfusions……………………………….. LMP?…………………………………………………….

Fever, COVID-19 symptoms (cough, fever, diarrhea, anhedonia, anosmia, ageusia, and severe fatigue), exposures, prior testing outside this study? ……………………………………………………………………………...

***Additional***

Attempting pregnancy (yes/no) .................. Birth Control Method?……………………………………

Breastfeeding……………………………

Medications……….………………………………………………………………………………....

Allergies…………………………………………………………………………………………...

**ROS:** ☐ Weight loss ☐ Nodes ☐ SOB ☐ Chest pain ☐ DOE ☐ Nausea ☐ Ht burn

☐ RUQ sx ☐ GI-bleed ☐ Nocturia ☐ Blood ☐ Urinary freq

Passed Health Questionnaire: Y☐ N☐ MD *Signature:* ………………………………………………………*Date:* ………………………………

| Family Medical History | mother | father | siblings |
|---|---|---|---|
| DM...................................................................... | ☐ | ☐ | ☐ |
| CA ...................................................................... | ☐ | ☐ | ☐ |
| HTN ...................................................................... | ☐ | ☐ | ☐ |
| Aneurysm ...................................................................... | ☐ | ☐ | ☐ |
| CAD...................................................................... | ☐ | ☐ | ☐ |
| Psych...................................................................... | ☐ | ☐ | ☐ |
| EtOH ...................................................................... | ☐ | ☐ | ☐ |
| IBD………. ...................................................................... | ☐ | ☐ | ☐ |
| Renal Disease or stones ...................................................................... | ☐ | ☐ | ☐ |
| Liver disease or cirrhosis ...................................................................... | ☐ | ☐ | ☐ |
| TB/PPD...................................................................... | ☐ | ☐ | ☐ |
| Migraine Headache……………………………………………………........... | ☐ | ☐ | ☐ |

***Lifestyle***

Coffee………………………………………………………....Vitamins…………………………………………………...

Exercise……………………………………………………….....Tobacco…………………………………………………….

EtOH……………………………………………………….. Drugs…………………………………………………….

Sexual Preference……………………………………………. .Last blood donation……………………………………..

Hx STD/genital ulcers……………………………………... Prior HIV test……………………………………………...

Incarceration hx……………………………………………………………………………………………………...

Diet: Omnivore Vegetarian Vegan Comments……………………………………………………….

Single Y☐ N☐ Married Y☐ N☐ Divorced Y☐ N☐ Children Y☐ N☐

Occupation……………………………………………………………………………………………………...

Living Situation…………………………………………………………………………………………………...

Highest education achieved………………………………………………………………………………………

Additional comments………………………………………………………………………………………..

…………………………………………………………………………………………………………………….

…………………………………………………………………………………………………………………….

…………………………………………………………………………………………………………………….

***Monkeypox Exposure History:***

i. Have you been diagnosed with monkeypox in the last 8 weeks?

ii. Have you had a rash in the last 8 weeks [or felt sick or had a fever, headache, chills, muscle aches, exhaustion, or swollen lymph nodes, or had a new or unexplained rash, which may look like pimples, blisters, or sores]?

iii. You may have been exposed to monkeypox if you answer “yes” to any of the following questions:

   a) Had a sexual partner who was subsequently diagnosed with monkeypox; OR
   b) Had multiple sexual partners in a geographic area with known monkeypox; OR
   c) Direct contact with an individual with infectious rash, scabs, or body fluids; OR
   d) Exposure to respiratory secretions of an individual with monkeypox, such as during prolonged, face-to-face contact, or during intimate physical contact, such as kissing, cuddling, or sex; or sharing of utensils, cups, or straws; OR
   e) Touched objects, fabrics (such as clothing or linens) or surfaces that previously were in contact with the rash or body fluids of someone with monkeypox; OR
   f) Had ungloved contact with a person with monkeypox, been splashed in the eye or oral cavity with saliva of an individual with monkeypox, been within six feet of an infected individual during any procedure that might create aerosols from oral secretions, skin lesions, or dispersal of dried lesion material (e.g., shaking of used bed linens) without wearing an N95 or equivalent respirator and eye protection; OR
   g) Been scratched or bitten by an animal infected with monkeypox or prepared meat or another product from such an animal

iv. Are you at risk of monkeypox exposure? At risk might include a “yes” response to any of the following:

   a) Men, transgender women and non-binary persons assigned male at birth who have sexual contact with men and have had multiple (> 1) or anonymous sexual partners in the past 3 months OR
   b) Women who have had sexual contact with any person listed above in (a) in the past 3 months OR
   c) Sex worker OR
   d) Staff at an establishment where sexual activity occurs (e.g., bathhouses, sex clubs) OR
   e) Persons who attend sex-on-premises venues (e.g., bathhouses, sex clubs) OR
   f) Attended a social gathering with high risk of skin-to-skin contact

Passed History: Y☐ N☐

*Signature: ……………………………………………………………………………Date: ………………………………*

***Physical Exam***

**Height_____**'_____"  **Weight_____**lb Temp: ____**HR________**/min**RR__**/min**BP__**/_______

General…………………………………………………………………………………………….

HEENT…………………………………………………………………………………………

Skin/hair…………………………………………………………………………………………..

Neck………………………………………………………………………………………………

Chest/Back……………………………………………………………………………………...

Heart……………………………………………………………………………………………….

Abdomen………………………………………………………………………………………..

Extremities/vascular……………………………………………………………………………

Neurological……………………………………………………………………………………

Rectal/Guiac……………………………………………………………………………………

**Discussion held with volunteer candidate re:**

- ☐ HIV consent - remind volunteer that HIV positive test results are reported to the State & CDC
- ☐ Opportunity for questions
- ☐ Signed copy of screening consent form given to volunteer
- ☐ Stool collection kit provided
- ☐ Agrees to NP swab collection for COVID-19 evals.
- ☐ Discussion re return of results done

Additional comments…………………………………………………………………………………..

…………………………………………………………………………………………………….

…………………………………………………………………………………………………….

……………………………………………………………………………………………………..

Passed Physical: Y☐ N☐

*Signature: ……………………………………………………………………Date: ………………………………*

### Example Donor History Questionnaire (DHQ) v2.1 with PrEP, PEP, ART

| | Yes | No |
|---|---|---|
| Are you | | |
| 1. Feeling healthy and well today? | ❑ | ❑ |
| 2. Currently taking an antibiotic? | ❑ | ❑ |
| 3. Currently taking any other medication for an infection? | ❑ | ❑ |
| 4. Have you taken any medications on the Medication Deferral List in the time frames indicated? (Review the Medication Deferral List.) | ❑ | ❑ |
| 5. Have you read the educational materials today? | ❑ | ❑ |
| In the past **48 hours,** | | |
| 6. Have you taken aspirin or anything that has aspirin in it? | ❑ | ❑ |
| In the past **8 weeks,** have you | | |
| 7. Donated blood, platelets or plasma? | ❑ | ❑ |
| 8. Had any vaccinations or other shots? | ❑ | ❑ |
| 9. Had contact with someone who was vaccinated for smallpox in the past 8 weeks? | ❑ | ❑ |
| In the past **3 months**, have you | | |
| 10. Had a blood transfusion? | ❑ | ❑ |
| 11. Had a transplant such as organ, tissue, or bone marrow? | ❑ | ❑ |
| 12. Had a graft such as bone or skin? | ❑ | ❑ |
| 13. Come into contact with someone else's blood? | ❑ | ❑ |
| 14. Had an accidental needle-stick? | ❑ | ❑ |
| 15. Had sexual contact with anyone who has ever had HIV/AIDS or has ever had a positive test for the HIV/AIDS virus? | ❑ | ❑ |
| 16. Had sexual contact with a prostitute or anyone else who has ever taken money or drugs or other payment for sex? | ❑ | ❑ |
| 17. Had sexual contact with anyone who has ever used needles to take drugs or steroids, or anything not prescribed by their doctor? | ❑ | ❑ |
| 18. Male donors: Had sexual contact with another male? | ❑ | ❑ |
| 19. Female donors: Had sexual contact with a male who had sexual contact with another male in the past 3 months? | ❑ | ❑ |
| 20. Had a tattoo? | ❑ | ❑ |
| 21. Had ear or body piercing? | ❑ | ❑ |
| 22. Had or been treated for syphilis or gonorrhea? | ❑ | ❑ |
| 23. Used needles to take drugs, steroids, or anything not prescribed by your doctor? | ❑ | ❑ |
| 24. Received money, drugs, or other payment for sex? | ❑ | ❑ |
| In the past **16 weeks,** | | |
| 25. Have you donated a double unit of red cells using an apheresis machine? | ❑ | ❑ |

### Example Donor History Questionnaire (DHQ) v2.1 with PrEP, PEP, ART

| | Yes | No |
|---|---|---|
| In the past **12 months,** have you | | |
| 26. Had sexual contact with a person who has hepatitis? | ❑ | ❑ |
| 27. Lived with a person who has hepatitis? | ❑ | ❑ |
| 28. Been in juvenile detention, lockup, jail, or prison for 72 hours or more consecutively? | ❑ | ❑ |
| In the past **three years,** have you | | |
| 29. Been outside the United States or Canada? | ❑ | ❑ |
| From **1980 through 1996,** | | |
| 30. Did you spend time that adds up to 3 months or more in the United Kingdom countries of England, Northern Ireland, Scotland, Wales, the Isle of Man, the Channel Islands, Gibraltar, or the Falkland Islands? | ❑ | ❑ |
| From **1980 through 2001**, did you | | |
| 31. Spend time that adds up to 5 years or more in France or Ireland? Time spent in Ireland does not include time spent in Northern Ireland which is part of the United Kingdom. | ❑ | ❑ |
| From **1980 to the present**, did you | | |
| 32. Receive a blood transfusion in France, Ireland, England, Northern Ireland, Scotland, Wales, the Isle of Man, the Channel Islands, Gibraltar, or the Falkland Islands? | ❑ | ❑ |
| Have you **EVER** | | |
| 33. Female donors: Been pregnant or are you pregnant now? | ❑ | ❑ |
| 34. Had a positive test for the HIV/AIDS virus? | ❑ | ❑ |
| 35. Had malaria? | ❑ | ❑ |
| 36. Received a dura mater (or brain covering) graft or xenotransplantation product? | ❑ | ❑ |
| 37. Had any type of cancer, including leukemia? | ❑ | ❑ |
| 38. Had any problems with your heart or lungs? | ❑ | ❑ |
| 39. Had a bleeding condition or a blood disease? | ❑ | ❑ |
| 40. Had a positive test result for *Babesia*? | ❑ | ❑ |

| **Use this area for additional questions** | Yes | No |
|---|---|---|
| xx. In the past **3 months,** have you taken any medication by mouth to prevent an HIV infection? | ❑ | ❑ |
| xx. In the past **2 years**, have you received an injection or shot to prevent an HIV infection? | ❑ | ❑ |
| xx. Have you **EVER** taken any medication to treat an HIV infection? | ❑ | ❑ |

***FMT DONOR : Brief Questionnaire***

Name………………………………………….. Date of Birth……………………………………………

Phone/Contact……………………………………………….

***History of Present Illness***

Any recent increased stool frequency, urgency, or bleeding…………………………………..

Any recent rhinorrhea, congestion, or fever……………………………………………………

Antibiotic use within last 4 weeks………………………………………………………..........

**ROS:** ☐ Weight loss ☐ Nodes ☐ SOB ☐ Chest pain ☐ DOE ☐ Nausea ☐ Ht burn

☐ RUQ sx ☐ GI-bleed ☐ Nocturia ☐ Blood ☐ Urinary freq

Passed Health Questionnaire: Yes ☐ No ☐

MD *Signature: ……………………………………Date: ………………………………*

### Appendix 15 – CLIA Certificates

### New York State Department of Health

PFI: 5227

#### Clinical Laboratory Permit

CLIA: 33D0653419

**Mount Sinai Laboratory**
One Gustave L Levy Pl Box 1612
New York NY 10029

Director:
Jeffrey S. Jhang, M.D.

Owner:
Mount Sinai Medical Center

is hereby authorized to perform laboratory procedures at the above location in the following categories in accordance with Article 5, Title V, Section 575 of the Public Health Law. This permit shall become void upon a change in the director, owner or location of the laboratory, and an application for a new permit shall be made to the Department.

*Bacteriology*
*Blood pH and Gases*
*Blood Services*
*Transfusion Service*
*Cellular Immunology*
*Non-Malignant Leukocyte Immunophenotyping*
*Clinical Chemistry*
*Cytokines*
*(for COVID-19 related testing)*
*Cytopathology*
*Gynecological Testing*
*Non-gynecological Testing*

*Cytogenetics*
*(limited to cancer cytogenetics)*
*Diagnostic Immunology*
*Diagnostic Services Serology*
*Endocrinology*
*Genetic Testing*
*Molecular*
*Hematology*
*Histocompatibility*
*Histopathology*
*General*

*Immunohematology*
*Mycobacteriology*
*Mycology*
*Oncology*
*Molecular and Cellular Tumor Markers*
*Soluble Tumor Markers*
*Parasitology*
*Toxicology*
*Clinical Toxicology-Qualitative Testing Only*
*Transplant Monitoring*
*Ther. Sub. Mon./Quant. Tox.*
*Virology*

Renewal
Effective Date: July 1, 2022
Expiration Date: June 30, 2023

Subject to Revocation
Permit Not Transferable

POST CONSPICUOUSLY

Serial: LAP 158874

CENTERS FOR MEDICARE & MEDICAID SERVICES

### CLINICAL LABORATORY IMPROVEMENT AMENDMENTS

*CERTIFICATE OF ACCREDITATION*

**LABORATORY NAME AND ADDRESS**

MAYO CLINIC LABS - ROCHESTER MAIN CAMP
200 FIRST STREET SW
ROCHESTER, MN 55905-0002

**CLIA ID NUMBER**

24D0404292

**EFFECTIVE DATE**

02/28/2021

**LABORATORY DIRECTOR**

WILLIAM G MORICE M.D.

**EXPIRATION DATE**

02/27/2023

Pursuant to Section 353 of the Public Health Services Act (42 U.S.C. 263a) as revised by the Clinical Laboratory Improvement Amendments (CLIA), the above named laboratory located at the address shown hereon (and other approved locations) may accept human specimens for the purposes of performing laboratory examinations or procedures.

This certificate shall be valid until the expiration date above, but is subject to revocation, suspension, limitation, or other sanctions for violation of the Act or the regulations promulgated thereunder.

CMS
CENTERS FOR MEDICARE & MEDICAID SERVICES

Regina S. Van Brakle
Regina S. Van Brakle, Acting Director
Division of Laboratory Services
Survey and Certification Group
Center for Clinical Standards and Quality

735 Certs2_020221

**If you currently hold a Certificate of Compliance or Certificate of Accreditation, below is a list of the laboratory specialties/subspecialties you are certified to perform and their effective date:**

| LAB CERTIFICATION (CODE) | EFFECTIVE DATE |
|---|---|
| HISTOCOMPATIBILTY (010) | 01/31/2003 |
| BACTERIOLOGY (110) | 10/13/1995 |
| MYCOBACTERIOLOGY (115) | 10/13/1995 |
| MYCOLOGY (120) | 10/13/1995 |
| PARASITOLOGY (130) | 10/13/1995 |
| VIROLOGY (140) | 10/13/1995 |
| GENERAL IMMUNOLOGY (220) | 10/13/1995 |
| ROUTINE CHEMISTRY (310) | 10/13/1995 |
| URINALYSIS (320) | 10/13/1995 |
| ENDOCRINOLOGY (330) | 10/13/1995 |
| TOXICOLOGY (340) | 10/13/1995 |
| HEMATOLOGY (400) | 10/13/1995 |
| ABO & RH GROUP (510) | 10/13/1995 |

| LAB CERTIFICATION (CODE) | EFFECTIVE DATE |
|---|---|
| ANTIBODY TRANSFUSION (520) | 10/13/1995 |
| ANTIBODY NON-TRANSFUSION (530) | 10/13/1995 |
| ANTIBODY IDENTIFICATION (540) | 10/13/1995 |
| COMPATIBILITY TESTING (550) | 10/13/1995 |
| HISTOPATHOLOGY (610) | 10/13/1995 |
| ORAL PATHOLOGY (620) | 10/13/1995 |
| CYTOLOGY (630) | 06/13/2003 |
| RADIOBIOASSAY (800) | 02/09/2011 |
| CYTOGENETICS (900) | 10/13/1995 |

FOR MORE INFORMATION ABOUT CLIA, VISIT OUR WEBSITE AT WWW.CMS.GOV/CLIA OR CONTACT YOUR LOCAL STATE AGENCY. PLEASE SEE THE REVERSE FOR YOUR STATE AGENCY'S ADDRESS AND PHONE NUMBER.
PLEASE CONTACT YOUR STATE AGENCY FOR ANY CHANGES TO YOUR CURRENT CERTIFICATE.

### Appendix 16 – NIH Adverse Event and Serious Adverse Event Guidelines

#### *NIA Adverse Event and Serious Adverse Event Guidelines*

##### BACKGROUND

This guideline describes the requirements and processes for reporting adverse events and unanticipated problems to the National Institute on Aging (NIA). It incorporates guidelines provided by the Office of Human Research Protections (OHRP) of the Department of Health and Human Services (DHHS) and describes Food and Drug Administration (FDA) reporting requirements. NIH is obligated to ensure that researchers comply with their approved reporting procedures. Clinical trial investigators funded by NIA are obligated under federal regulations to appropriately inform the Institute of adverse events and unanticipated problems, and NIA is required to ensure that the appropriate procedures are in place to support this reporting.

##### DEFINITIONS

Definitions are from the January 2007 OHRP *Guidance on Reviewing and Reporting Unanticipated Problems Involving Risks to Subjects or Others and Adverse Events, OHRP Guidance*.

**Adverse Event (AE):**

Any untoward or unfavorable medical occurrence in a human study participant, including any abnormal sign (e.g. abnormal physical exam or laboratory finding), symptom, or disease, temporally associated with the participants' involvement in the research, whether or not considered related to participation in the research.

**Serious Adverse Event (SAE):**

Any adverse event that:

- Results in death
- Is life threatening, or places the participant at immediate risk of death from the event as it occurred
- Requires or prolongs hospitalization
- Causes persistent or significant disability or incapacity
- Results in congenital anomalies or birth defects
- Is another condition which investigators judge to represent significant hazards

**Unanticipated Problem (UP):**

Defined by DHHS 45 CFR part 46 as any incident, experience, or outcome that meets all of the following criteria:

- unexpected, in terms of nature, severity, or frequency, given (a) the research procedures that are described in the protocol-related documents, such as the IRB-approved research protocol and informed consent document; and (b) the characteristics of the study population;
- related or possibly related to participation in the research (in this guidance document, possibly related means there is a reasonable possibility that the incident, experience, or outcome may have been caused by the procedures involved in the research);
- suggests that the research places participants or others at a greater risk of harm (including physical, psychological, economic, or social harm) than was previously known or recognized.

**Adverse Events versus Unanticipated Problems**

- The vast majority of adverse events occurring in human subjects are not unanticipated problems.
- A small proportion of adverse events are unanticipated problems.
- Unanticipated problems include other incidents, experiences, and outcomes that are not adverse events.

#### RESPONSIBILITIES

Both the NIA and the investigators it funds have responsibilities with respect to safety reporting.

##### Investigators

Investigators conducting clinical research are responsible for:

- Assurance that their protocols are conducted in compliance with these guidelines.
- Submission of IRB-approved protocol to the NIA program office and to the Data and Safety Monitoring Board (DSMB), as applicable.

  A Data and Safety Monitoring Plan (DSMP) that is commensurate with the study risk and reflects this guideline should also be submitted either as part of the application or a separate document.

  - The DSMP should describe collection and reporting plans for adverse events, serious adverse events, and unanticipated problems commensurate with nature and complexity of the study.
  - The DSMP should include recipients of Serious Adverse Event and Unanticipated Problem reports (e.g. Institutional Review Boards (IRBs), DSMB or Safety Officer, and NIA).

The DSMB should approve both the protocol and the DSMP.

- Adherence to the DSMP with respect to timely submission of adverse events, serious adverse events, and unanticipated problems.

**NIA**

NIA program staff members are responsible for providing:

- Assistance to extramural investigators in understanding and applying adverse event and serious adverse event guidelines and for ensuring compliance with OHRP guidance in all NIA funded clinical research.
- Oversight of these guidelines, which includes periodic review and revision as relevant rules and regulations change.
- Assurance that the DSMP addresses reporting of adverse events, serious adverse events, and unanticipated problems.
- Verification that all corrective action plans have been adequately implemented.
- Assurance that the study has an independent safety monitoring body commensurate with study risk (DSMB or Safety Officer).
- Ongoing oversight of the safety reporting process to assure that potential safety issues are addressed.

#### REPORTING PROCESSES

Adverse Events, Serious Adverse Events, and Unanticipated Problems have specific reporting procedures.

##### Adverse Event Reporting

All AEs are collected on an Adverse Event Form, either in paper or electronic format. A sample AE Form is shown in Adverse Event Form. All AEs experienced by the participant during the time frame specified in the protocol (e.g., from the start of intervention through the end of the study) are to be reported, as outlined in the protocol. The reporting requirements to the IRB, to the NIA and to the FDA (in case of drug and device studies) may differ and must be complied with.

Please note that the AE form contains a column to indicate whether the event is SERIOUS. Thus, SAEs are a subset of the reported AEs.

Routine reporting of AEs is described in the DSMP and may be monthly or quarterly as determined with the NIA staff and /or the DSMB/Safety Officer.

##### Serious Adverse Event Reporting

When SAEs occur that are unanticipated (i.e., not listed in the Data and Safety Monitoring Plan) and that are related to the intervention, they should be reported to NIA

Program Officer and to the DSMB Chair (or a Safety Officer, for studies without the DSMB) or to the designated DSMB member if a DSMB is established within 48 hours of study's knowledge of SAE. The summary of all other SAEs should be reported to NIA Program Officer and to the DSMB (or a Safety Officer) quarterly, unless otherwise requested by the DSMB or a Safety Officer. Expected SAEs should be listed in the Data and Safety Monitoring Plan. All deaths require expedited reporting (usually within 24 hours of study's knowledge of death). The report of death should be submitted to NIA Program Officer and to the DSMB Chair (or a Safety Officer, for studies without the DSMB) or to the designated DSMB member if a DSMB is established.

The expedited report should be followed by a detailed, written SAE report as soon as possible. Follow up information may be required and asked for by the independent safety monitoring body directly, or through the NIA or its representative. A sample of the SAE reporting form used for NIH Intramural Programs is shown in Serious Adverse Event Form.

##### Unanticipated Problem Reporting

Investigator institutions must have written procedures for ensuring prompt reporting to the IRB and NIA, and others as appropriate, of any Unanticipated Problem involving risks to study participants or others (45 CFR 46.103(b)(5)). The Unanticipated Problems reporting procedures must include a corrective plan and measures to prevent reoccurrence. It is recommended that such events be reported within 48 hours to NIA unless they are also SAEs.

Reports of Unanticipated Problems, as defined above, should be forwarded to OHRP using, typically within two weeks of the event.

The flow chart in AE/SAE Process Flow provides an algorithm of the reporting process.

#### CLASSIFYING ADVERSE EVENTS

Adequate review, assessment, and monitoring of adverse events require that they be classified as to ***severity***, ***expectedness***, and potential ***relatedness*** to the study intervention. Study protocols must include a description of how adverse events will be classified in these terms. These classifications determine the reporting requirements.

##### Severity

Classifications often include the following:

- ***Mild:*** Awareness of signs or symptoms, but easily tolerated and are of minor irritant type causing no loss of time from normal activities. Symptoms do not require therapy or a medical evaluation; signs and symptoms are transient.
- ***Moderate:*** Events introduce a low level of inconvenience or concern to the participant and may interfere with daily activities, but are usually improved by

simple therapeutic measures; moderate experiences may cause some interference with functioning

- ***<u>Severe</u>:*** Events interrupt the participant's normal daily activities and generally require systemic drug therapy or other treatment; they are usually incapacitating

Severity is not synonymous with seriousness. A ***severe*** rash is not likely to be an ***SAE.*** Likewise, a ***severe*** headache is not necessarily an ***SAE.*** However, ***mild*** chest pain may result in a day's hospitalization and thus is an ***SAE.***

**Expectedness**

AEs must be assessed as to whether they were expected to occur or unexpected, meaning not anticipated based on current knowledge found in the protocol, investigator brochure, product insert, or label. Categories are:

- ***<u>Unexpected</u> -*** nature or severity of the event is not consistent with information about the condition under study or intervention in the protocol, consent form, product brochure, or investigator brochure.

- ***<u>Expected</u> -*** event is known to be associated with the intervention or condition under study.

**Relatedness**

The potential event relationship to the study intervention and/or participation is assessed by the site investigator. A comprehensive scale in common use to categorize an event is:

- *Definitely Related*: The adverse event is clearly related to the investigational agent/procedure – i.e. an event that follows a reasonable temporal sequence from administration of the study intervention, follows a known or expected response pattern to the suspected intervention, that is confirmed by improvement on stopping and reappearance of the event on repeated exposure and that could not be reasonably explained by the known characteristics of the subject's clinical state.
- *Possibly Related:* An adverse event that follows a reasonable temporal sequence from administration of the study intervention follows a known or expected response pattern to the suspected intervention, but that could readily have been produced by a number of other factors.
- *Not Related:* The adverse event is clearly not related to the investigational agent/procedure - i.e. another cause of the event is most plausible; and/or a clinically plausible temporal sequence is inconsistent with the onset of the event and the study intervention and/or a causal relationship is considered biologically implausible.

**SAFETY REPORTING REQUIREMENTS FOR STUDIES INVOLVING INVESTIGATIONAL NEW DRUGS (IND)**

The FDA amended its regulations governing IND safety reporting for human drug products (21 CFR 312 and 320). The following modified definitions and reporting requirements apply to research subject to IND applications:

**Definitions:**

**Sponsor:**

Sponsor means a person or institution that takes responsibility for and initiates a clinical investigation. The sponsor does not actually conduct the investigation unless the sponsor is a Sponsor-Investigator.

**Sponsor-Investigator:**

Sponsor-Investigator is an individual who both initiates and conducts an investigation and under whose immediate direction the investigational drug is administered or dispensed. The requirements applicable to Sponsor-Investigator include both those applicable to an investigator and a sponsor.

**Adverse event:**

Any untoward or unfavorable medical occurrence in a human study participant, including any abnormal sign (e.g. abnormal physical exam or laboratory finding), symptom, or disease, temporally associated with the participants' involvement in the research, whether or not considered related to participation in the research.

**Adverse reaction:**

Any adverse event caused by the drug.

**Suspected adverse reaction:**

Any adverse event for which there is a reasonable possibility that the drug caused the adverse event. "Reasonable possibility" means there is evidence to suggest a causal relationship between the drug and the adverse event.

***Serious* adverse event or *serious* suspected adverse reaction:**

An adverse event or suspected adverse reaction is considered "serious" if, in the view of either the investigator or sponsor, it results in any of the following outcomes:

- Death
- A life-threatening adverse event
- Inpatient hospitalization, or prolonged of existing hospitalization

- A persistent or significant incapacity or substantial disruption of the ability to conduct normal life functions
- A congenital anomaly/birth defect
- Important medical events that may not result in death, be life threatening, or require hospitalization may be considered serious, when based upon appropriate medical judgment, they may jeopardize the patient or subject and may require medical or surgical intervention to prevent one of the outcomes listed in this definition.

**_Life-threatening_ adverse event or _life-threatening_ suspected adverse reaction:**

An adverse event or suspected adverse reaction is considered “life-threatening” if, in the view of the investigator, its occurrence places the patient or subject at immediate risk of death. It does not include an adverse event or suspected adverse reaction that, had it occurred in a more severe form, might have caused death.

**_Unexpected_ adverse event or _unexpected_ suspected adverse reaction:**

An adverse event or suspected adverse reaction is considered “unexpected” if it is not listed in the investigator brochure (IB); is not listed at the specificity or severity that has been observed; or if IB is not required or not available, is not consistent with the risk information described in the protocol or safety monitoring plan.

#### IND Safety Reporting

An overview of the Safety Reporting requirements are described below and in the flow diagram, as shown in Figure 1.

**Fatal or life-threatening serious unexpected suspected adverse reactions (SUSARs) reports:**

The sponsor (or sponsor-investigator) must notify the FDA of any SUSARs to the study drug as soon as possible but *no later than 7 calendar days* after the initial receipt of the information.

**Other reporting requirements:**

The sponsor (or sponsor-investigator) must also notify the FDA and all participating Principal Investigators (in multi-site studies) in an IND safety report about potential serious risks, from clinical trials and any other source, *no later than 15 days* in the following cases:

- **SUSARs that are not fatal or life-threatening**. Any suspected adverse reaction that is *both serious and unexpected* must be reported. The report must proceed only if there is evidence to suggest a causal relationship between the drug and the adverse event.
- **Finding from other studies**. Any findings from epidemiological studies, pooled analysis of multiple studies, or clinical studies (other than those reported under serious and unexpected adverse reactions above) must be reported whether or not conducted under an IND or whether or not conducted by the investigator, that suggest a significant risk in humans exposed to the drug.
- **Findings from animal or in vitro testing**. Any finding from animal or in vitro testing whether or not conducted by the investigator that suggest a significant risk in humans exposed to the drug such as reports of mutagenicity, teratogenicity, or carcinogenesis or reports of organ toxicity, must be reported.
- **Increased rate of occurrence of serious suspected adverse reactions.** Any clinically important increase in the rate of serious suspected adverse reactions over that listed in the protocol or IB must also be reported.
- **SUSARs that are part of study endpoints.** Study endpoints (e.g. mortality or major morbidity) must be reported to the FDA as described in the protocol and ordinarily would not be reported as IND safety reports. However, if a serious and unexpected adverse event occurs for which there is evidence suggesting a causal relationship between the drug and the event (e.g., death from anaphylaxis), the event must be reported under 21 CFR 312.32 as a serious and unexpected suspected adverse reaction even if it is a component of the study endpoint (e.g., all-cause mortality).

Note: The most frequent and most serious adverse experiences that are not serious unexpected suspected adverse reactions to the study drug have to be summarized and reported to the FDA in the annual IND report.

**Figure 1: FDA IND Expedited Safety Reporting Requirements for Individual Case Safety Reports Flow**

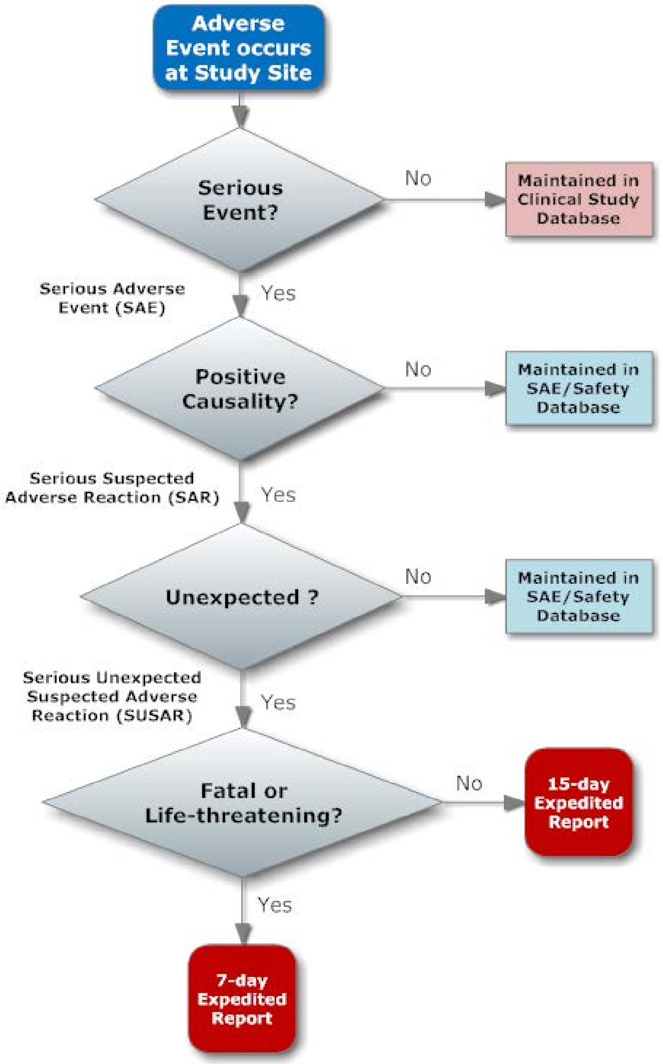

#### Investigator Reports:

An investigator (if he/she is not a sponsor-investigator) must report to the sponsor any *serious* adverse event within 24 hours of investigator learning about the event, whether or not considered drug related, including those listed in the protocol or investigator brochure and must include an assessment of whether there is a reasonable possibility that the drug caused the event.

The investigator must also record *nonserious* adverse events and report them to the sponsor according to the timetable for reporting specified in the protocol.

**References**

NIA Implementation of Policies for Human Intervention Studies

January 2007 OHRP Guidance *on Reviewing and Reporting Unanticipated Problems Involving Risks to Subjects or Others and Adverse Events,* http://www.hhs.gov/ohrp/policy/advevntguid.html

21 CFR 312.50, General Responsibilities of the Sponsor, http://www.accessdata.fda.gov/scripts/cdrh/cfdocs/cfcfr/CFRSearch.cfm?fr=312.50

21 CFR 312.32, IND Safety Reports, http://www.accessdata.fda.gov/scripts/cdrh/cfdocs/cfcfr/CFRSearch.cfm?fr=312.32

ICH E6 5.0 Sponsor, Good Clinical Practices, http://www.ich.org/fileadmin/Public_Web_Site/ICH_Products/Guidelines/Efficacy/E6_R1/Step4/E6_R1__Guideline.pdf
